# Education/training for health workers/students on inclusive and gender-affirmative care for trans and gender-diverse people: a systematic review

**DOI:** 10.64898/2026.06.04.26354880

**Authors:** Jun Xia, Zheng Zhu, Guowen Zhang, Quan Shen, Esther Su, Jan Schoones, Jon Arcelus, Tiantian Hu, Mengqi Xu, Xiaoxin Zhang, Zhan Zhao, Zheng Ye, Xiaomei Yao

**Author notes:** **Correspondence** Jun Xia, Nottingham Ningbo GRADE Centre, School of Economics, University of Nottingham Ningbo China, Ningbo, China.; telephone: 86+18355159105, Zheng Zhu, Fudan University School of Nursing, Fudan University, Shanghai, China.; Xiaomei Yao, Department of Health Research Methods, Evidence and Impact; Department of Oncology, McMaster University, Canada. **Co-first authors:** Jun Xia and Zheng Zhu had full access to all of the data in the study and take responsibility for the integrity of the data and the accuracy of the data analysis. Jun Xia and Zheng Zhu contributed equally as co–first authors.

## Abstract

**Introduction:** Trans and gender-diverse (TGD) individuals often face stigma and discrimination in healthcare, hindering access to gender-affirming care. Training healthcare workers on TGD health aims to foster inclusive and affirming care practices. This review aimed to evaluate the effectiveness of TGD health training programs for healthcare workers.

**Methods:** This systematic review followed the PRISMA guidelines and was registered with PROSPERO (CRD42023443288). We searched 13 databases for studies up to March 2024, with no language/geographic restrictions. Ten reviewers screened studies in pairs, resolving discrepancies via discussion or third-reviewer input. We included randomized/non-randomized comparative and before-after studies for quantitative analysis (mean difference [MD] or standardized mean difference [SMD] with 95% CIs), and qualitative/mixed-methods studies for thematic synthesis. Evidence certainty was assessed using GRADE (quantitative) and GRADE-CERQual (qualitative). Outcomes included knowledge, attitudes, skills, discrimination, competence, comfort, TGD quality of life, and stakeholder preferences.

**Results:** From 20,188 records, 85 studies were included. Training appears to have improved healthcare workers’ knowledge (SMD=1.08, 95% CI 0.78-1.39), attitudes (SMD=0.22, 95% CI 0.05-0.39), skills (SMD=0.96, 95% CI 0.56-1.37), competence (SMD=0.55, 95% CI 0.29-0.81), and comfort (SMD=0.69, 95% CI 0.17-1.21). Qualitative analysis of 130 findings identified 18 categories and four key themes on intervention design and impact.

**Conclusions:** TGD training programs may enhance health workers’ knowledge, attitudes, skills, competence, and comfort. Well-structured, interactive, and inclusive programs showed promise, but evidence certainty was low with limited follow-up. Further high-quality research is needed to confirm these findings.

## 1 Introduction

Trans and gender-diverse (TGD) people often face significant stigma and discrimination, hindering their access to and utilization of comprehensive, gender-affirming healthcare.^1^ Education for health workers can improve gender sensitivity and their understanding of gender identity’s multifaceted aspects. Yet, evidence evaluating this impact is limited. Previous reviews, often focused on mixed LGBTQ groups, fail to isolate the effects specific to transgender and gender-diverse (TGD) individuals.^2–4^ The effectiveness of different interventions, particularly how specific features increase efficacy by aligning with health workers’ values, remain insufficiently explored.

To address this gap, we conducted a comprehensive review to: 1) quantitatively assess intervention effectiveness on nine key outcomes for gender-inclusive and affirming care (GAC), and 2) qualitatively explore health workers’ values and preferences regarding these interventions.

## 2 Methods

### 2.1 Search strategy and selection criteria

This comprehensive systematic review was conducted following the Preferred Reporting Items for Systematic Reviews and Meta-Analyses (PRISMA) guidelines.^5^ It was registered with PROSPERO (CRD42023443288).

An information specialist (JS) designed and ran the search using synonyms and MeSH terms for three concepts: “health workers” “education” and “trans and gender-diverse people”. The strategy (Appendix 1) covered 13 major databases from inception until 7^th^ March 2024 without date or language restriction: PubMed, MEDLINE (Ovid), Embase (Ovid), Emcare (Ovid), PsycINFO (EBSCOhost), CENTRAL (Cochrane Library), Web of Science, Social Services Abstracts (ProQuest), Sociological Abstracts (ProQuest), Academic Search Premier (EBSCOhost), Psychology and Behavioural Sciences Collection (EBSCOhost), ERIC (EBSCO) and LILACS. We also hand-searched references and tracked citations of included studies.

We included randomised controlled trials (RCTs), non-randomised comparative studies (NRS), and uncontrolled before-and-after studies (UBA) that investigated education or training interventions on inclusivity and GAC, aimed at health workers/ students. Health workers are defined as all people primarily engaged in activities designed to enhance health^6^ (e.g., health services receptionists, nurses, doctors, security personnel, and others) and included both pre-service and in-service health students and workers. Studies from any geographic location or healthcare setting were eligible for inclusion. However, we excluded studies that exclusively focused on GAC in children or adolescents. Interventions could be facilitated or self-directed.

We complemented the quantitative review with a qualitative synthesis exploring values and preferences, including qualitative studies (e.g., phenomenology, grounded theory) and mixed-methods studies containing direct participant quotations from any setting.

### 2.2 Study outcomes

We focused on ten critical outcomes of interest in this review. These included: 1) access to health services; 2) violence experienced by TGD individuals in healthcare settings; 3) changes in the competence of health workers in providing inclusive care and GAC, including knowledge, attitudes, skills, and behaviour (e.g., discrimination); 4) stigma and discrimination experienced by TGD individuals in healthcare settings; 5) changes in health outcomes for TGD individuals; 6) changes in the comfort of health workers in delivering services TGD people; 7) changes in the comfort of recipients of care; 8) access to and utilisation of healthcare services by TGD people; and 9) changes in the quality of life of TGD people following the target intervention; 10) values and preferences among stakeholders.

### 2.3 Data extraction, management, and analysis

All references were exported to Endnote (Version X9; Philadelphia, USA)^7^ for deduplication. Rayyan,^8^ a free web-based tool, was used for the initial screening of titles and abstracts. Ten reviewers (JX, ZY, ZZ, YX, ZZ, QS, GWZ, MQX, XXZ, TTH) worked in duplicate to screen titles/abstracts and assess full texts of potentially eligible studies, recording exclusion reasons. Disagreements were resolved by consensus or third-party adjudication. Data extraction was similarly performed in duplicate using a pilot-tested, standardised form.

For continuous outcomes in the quantitative review, we calculated mean differences (MD) or standardized mean differences (SMD) with 95% CIs, as appropriate. When studies reported multiple subscales, we aggregated them into single pre- and post-intervention values for meta-analysis using a random-effects model.

In the qualitative review, thematic synthesis on values and preferences was conducted following the methodology proposed by Thomas and Harden.^9^ Following line-by-line coding, initial codes were grouped into descriptive themes. Axial coding further delineated subcategories within each theme. Through discussion, two reviewers co-constructed the themes and developed a hypothesized model of their relationships, supported by analytical and reflexive memos.

### 2.4 Assessment of risk of bias and evidence certainty

The risk of bias for the quantitative review was assessed by one reviewer and verified by a second reviewer, with discrepancies resolved through discussion with additional reviewers as needed. The Cochrane Risk of Bias 2 (RoB2) Tool was used for RCTs (https://www.riskofbias.info/welcome/rob-2-0-tool),^10^ and the Risk of Bias in Non-randomized Studies of Interventions (ROBINS-I) was used for NRS and UBA.^11^ For qualitative studies, we applied the Critical Appraisal Skills Programme (CASP) checklist.^12^

The certainty of evidence in the quantitative review was evaluated using the Grading of Recommendations, Assessment, Development, and Evaluations (GRADE) approach.^11,13^ We adopted Cohen’s thresholds of effect size to assess imprecision for continuous variables (i.e., 0.2, 0.5, and 0.8 indicating small, moderate, and large thresholds).^14^ Confidence intervals crossing one or two thresholds led to a one- or two-level downgrade, respectively. We used GRADEPro to create an Evidence Profile for each outcome with available data (https://www.gradepro.org/).^15^

For the qualitative review, we used the GRADE-Confidence in Evidence from Reviews of Qualitative research (GRADE-CERQual) approach,^16,17^ and GRADE-CERQual Evidence Profile was presented within a Summary of Qualitative Findings (SoQF) table.^17^ The CERQual assessment was primarily undertaken by two reviewers (ZZ, GWZ) and final decisions were discussed amongst the entire review team. Confidence ratings for each finding were categorised from “high” to “very low”, based on the four components of the CERQual framework.

## 3 Results

### 3.1 Study selection

Initial search yielded 20,188 records; 85 studies met criteria (70 quantitative, 30 qualitative), including 17 mixed-methods studies (Figure 1).

**Figure 1.**
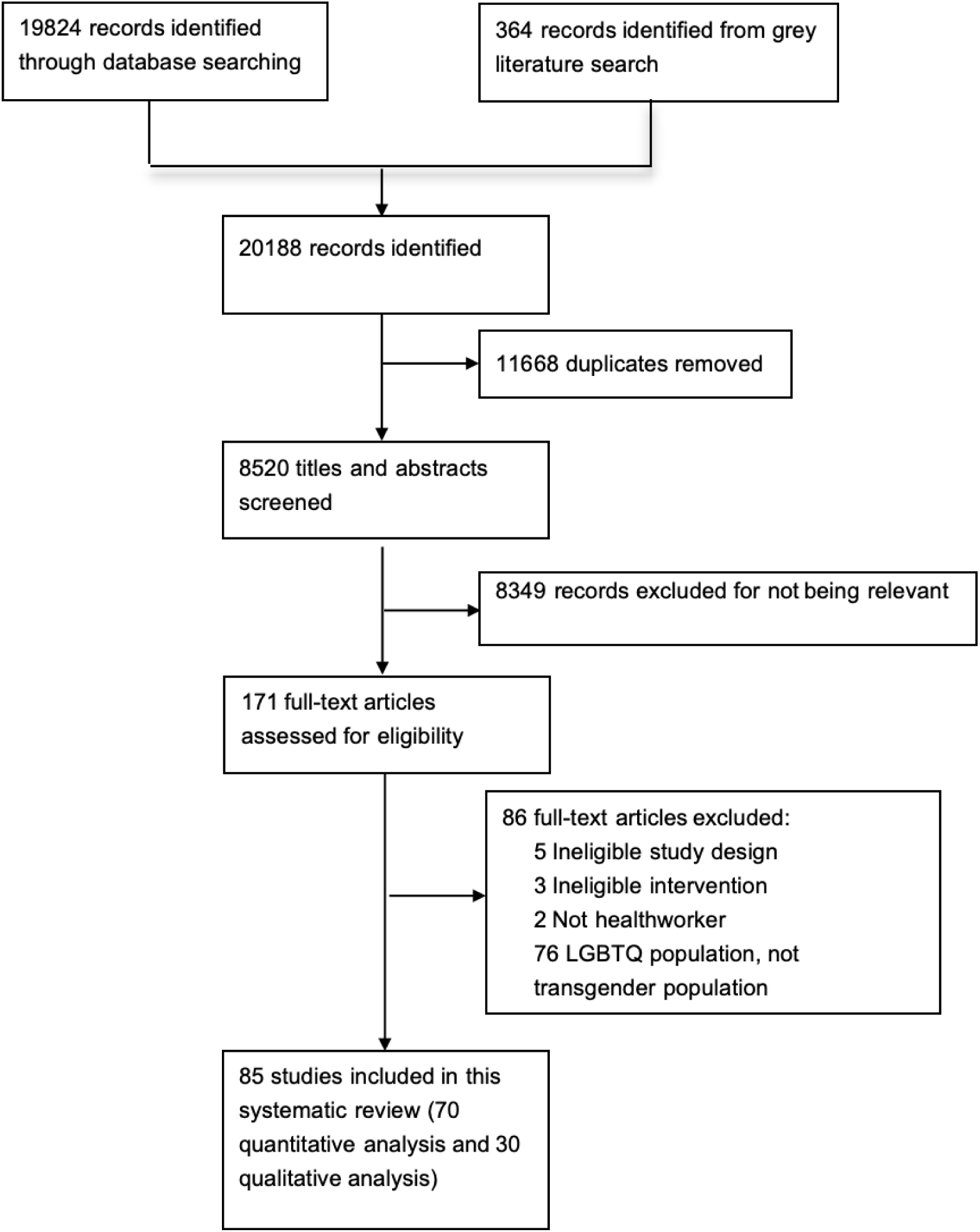
PRISMA Flowchart.

### 3.2 Characteristics of included studies

A quantitative synthesis was performed on 70 studies (Appendix 2), which were predominantly from the USA and included three RCTs,^18–20^ two NRS,^21,22^ and 65 UBAs. These studies evaluated outcomes such as knowledge, skills, and attitudes. Additionally, a qualitative synthesis included 30 studies (Appendix 3), with 17 mixed-methods studies contributing to both analyses.

Of the 30 qualitative studies(Appendix 3), 19 used a concurrent mixed-methods design^23–41^ while 11 studies used a qualitative design.^42–52^ Of the 30 studies, 24 used multiple education and training approaches.^23–40,44,45,47,48,50,52^ Nine included lectures^23,24,32,39,40,44,45,47,52^ and 17 involved interactive sessions.^23,24,26–29,32,33,35–37,39,44,45,47,50,52^ Five included training recipients from multiple disciplines^28,37–39,43^ and 6 involved both students and health professionals.^28,35,37,41,49,50^.

### 3.3 Risk of bias and evidence certainty

The evidence certainty was “Very Low” per GRADE assessment, as all studies had high risk of bias (RCTs), serious risk (comparative), or serious/critical risk (UBAs), due to inconsistency, indirectness, imprecision, and publication bias (Appendices 4-5).

Of the 30 qualitative studies, 24 had significant methodological limitations, while only 7 raised no or minor concerns ^40,42–44,50–52^. Seven studies had moderate concerns ^24,28,38,45,46,48,49^, and 16 had serious concerns ^23,25–27,29–37,39,41,47^. According to the GRADE-CERQual approach, confidence in all findings was rated as “Low” or “Very Low” (Appendices 4 and 6).

### 3.4 Findings

#### 3.4.1 Knowledge outcome

Of the 43 studies measuring knowledge, one RCT found a trivial improvement, while two NRSs reported moderate gains (Appendix 5: section 1). A meta-analysis of 18

UBAs (n≈1,100) showed a large effect (SMD 1.08, 95% CI 0.78-1.39) (Figure 2), with consistent results across subgroups (Appendix 7: Figures 3-6). The remaining 22 UBAs, though not pooled, all reported knowledge improvements.

**Figure 2.**
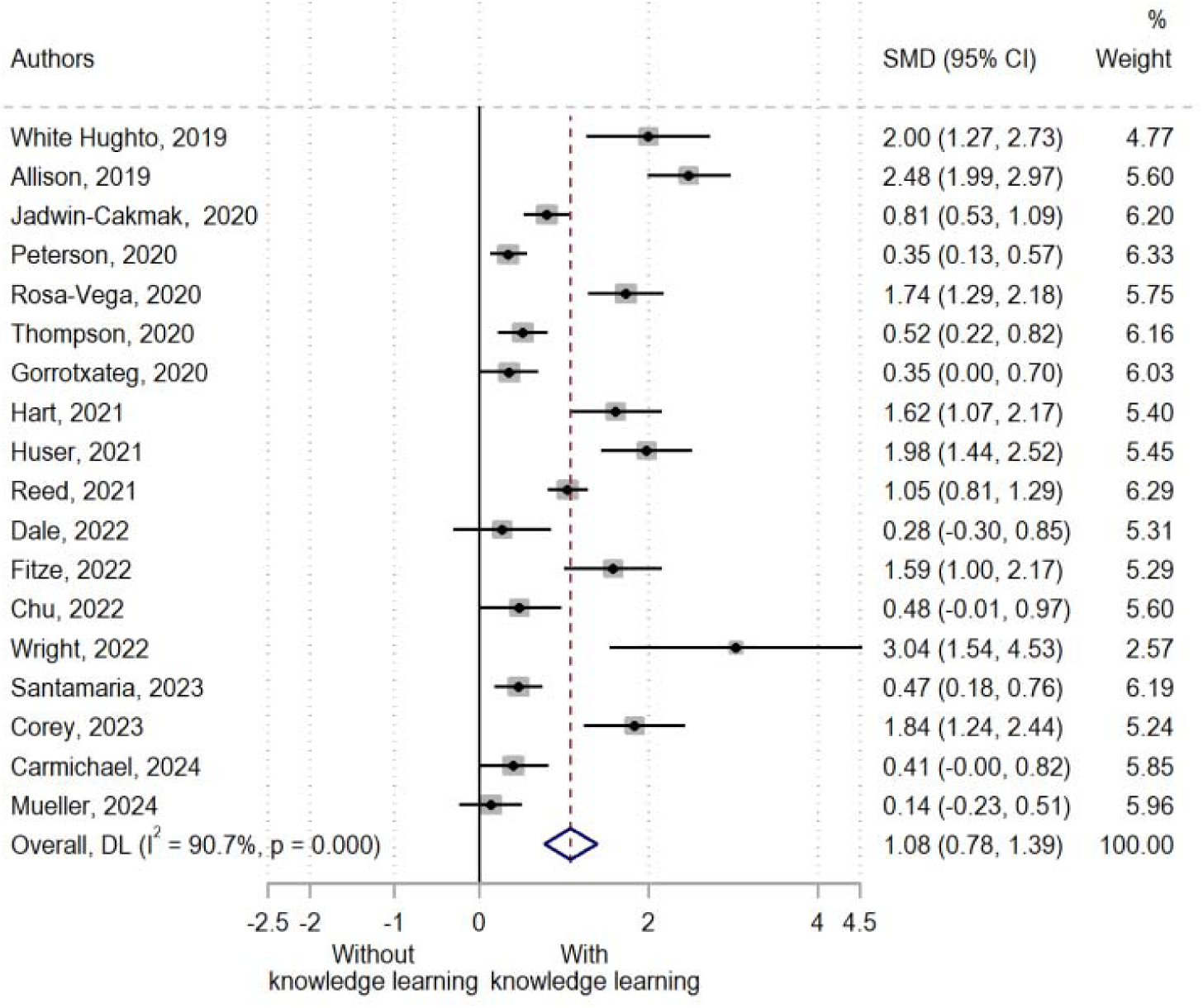
Change in competence: knowledge (main analysis, higher score = better outcome)

**Figure 3.**
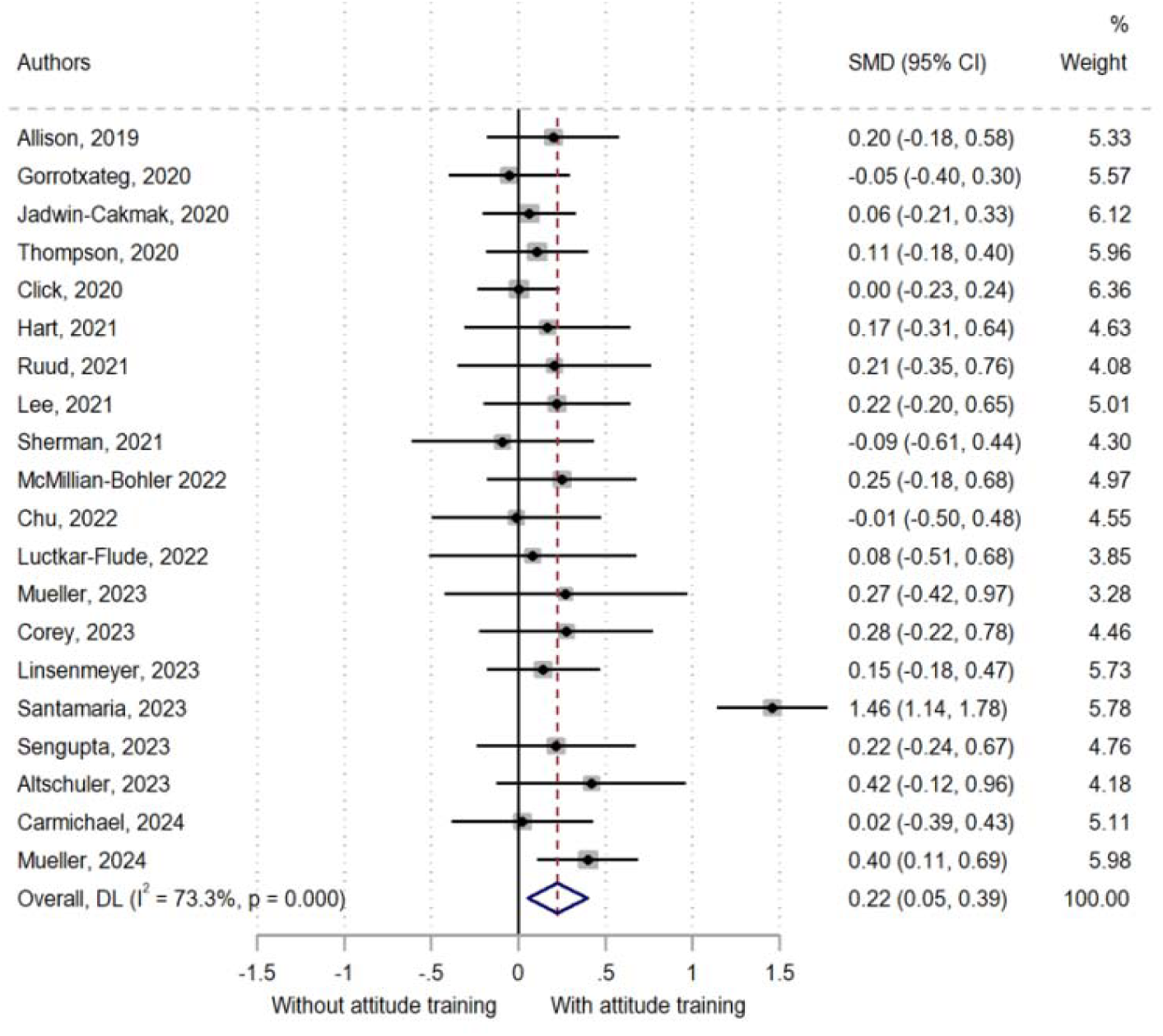
Change in competence: attitude (higher score = better outcome)

#### 3.4.2 Attitude outcome

Of 35 studies on attitude change, one NRS^22^ showed a trivial improvement (SMD 0.17, 95% CI -0.09 to 0.43) (Appendix 5: section 2). A meta-analysis of 20 UBAS,^1,25,27,28,33,35–37,40,53–77^ indicated a slight overall improvement (SMD 0.22, 95% CI 0.05–0.39) (Figure 3), with subgroup analyses revealing trivial differences across interventions and settings (Appendix 7: Figures 7-10).

#### 3.4.3 Discrimination outcome

Two UBAs reported changes in discrimination (Appendix 5: section 3).^74,78^ The pooled summary effect indicated no clear difference between groups (SMD -0.31, 95% CI -0.68 to 0.06) before and after the intervention (Appendix 7: Figure 11).

#### 3.4.4 Skills outcome

Two comparative studies^21,22^ and 16 UBAs reported changes in skills (Appendix 5: section 4).^26,33,37,40,53,62,70–73,76,79–83^ A meta-analysis of 9 UBAs showed large improvements (SMD 0.96, 95% CI 0.56–1.37) (Appendix 7: Figure 12), consistent with comparative studies and non-pooled data. Subgroup analysis indicated mixed-learning approaches (SMD 1.75) may be more effective than self-directed or facilitated-only learning (SMD 0.47) (Appendix 7: Figure 13).

#### 3.4.5 Competence outcome

Two RCTs^18,19^ and 31 UBAs^1,24,26,31–34,38,47,53,54,58–61,63,64,66,71,76,77,79,83–91^ assessed competence improvements. Both RCTs favored the intervention (Appendix 5: section 5). Meta-analysis of 16 UBAs (n=909–1,120) demonstrated moderate overall improvement (SMD 0.55, 95% CI 0.29–0.81). Subgroup analyses revealed larger effects in studies with bigger sample sizes (SMD 0.64) compared to smaller studies (SMD 0.43), while other subgroup differences were trivial (Appendix 7: Figure 14-18).

#### 3.4.6 Comfort outcome

Of 19 UBAs measuring comfort^1,24,33,37,56,57,63,64,66,67,72,73,81,92–97^ 12 lacked sufficient data for pooling. The remaining 7 studies (N=379-420) demonstrated moderate improvement (SMD 0.69, 95% CI 0.17-1.20) (Appendix 5: section 6 and Appendix 7: Figure 19).

No studies reported data on TGD health outcomes, care access, violence, or healthcare stigma.

#### 3.4.7 Values and preferences

A synthesis of 30 studies yielded 130 findings, which were consolidated into 18 categories and ultimately four overarching synthesised findings. These synthesised findings all centered on training aspects within healthcare settings. The first emphasises the importance of structure and content in training, specifically focusing on the pedagogical preferences of health workers and students regarding inclusive and gender-affirming care. Table 1 shows the summary of finding 1 and categories in more detail. The second synthesised finding highlights the multifaceted impacts of such training, with professionals and students acknowledging the diverse benefits and values of participating in these programs. The third synthesised finding explores the various factors that influence the successful implementation of this training. Lastly, the fourth synthesised finding presents suggestions from health workers on how to effectively design and conduct training interventions to promote inclusive and GAC, reflecting their insights and experiences in the field. Appendix 6 (Tables 2-4) shows the summary of findings 2-4 and categories in more detail.

**Table 1.**
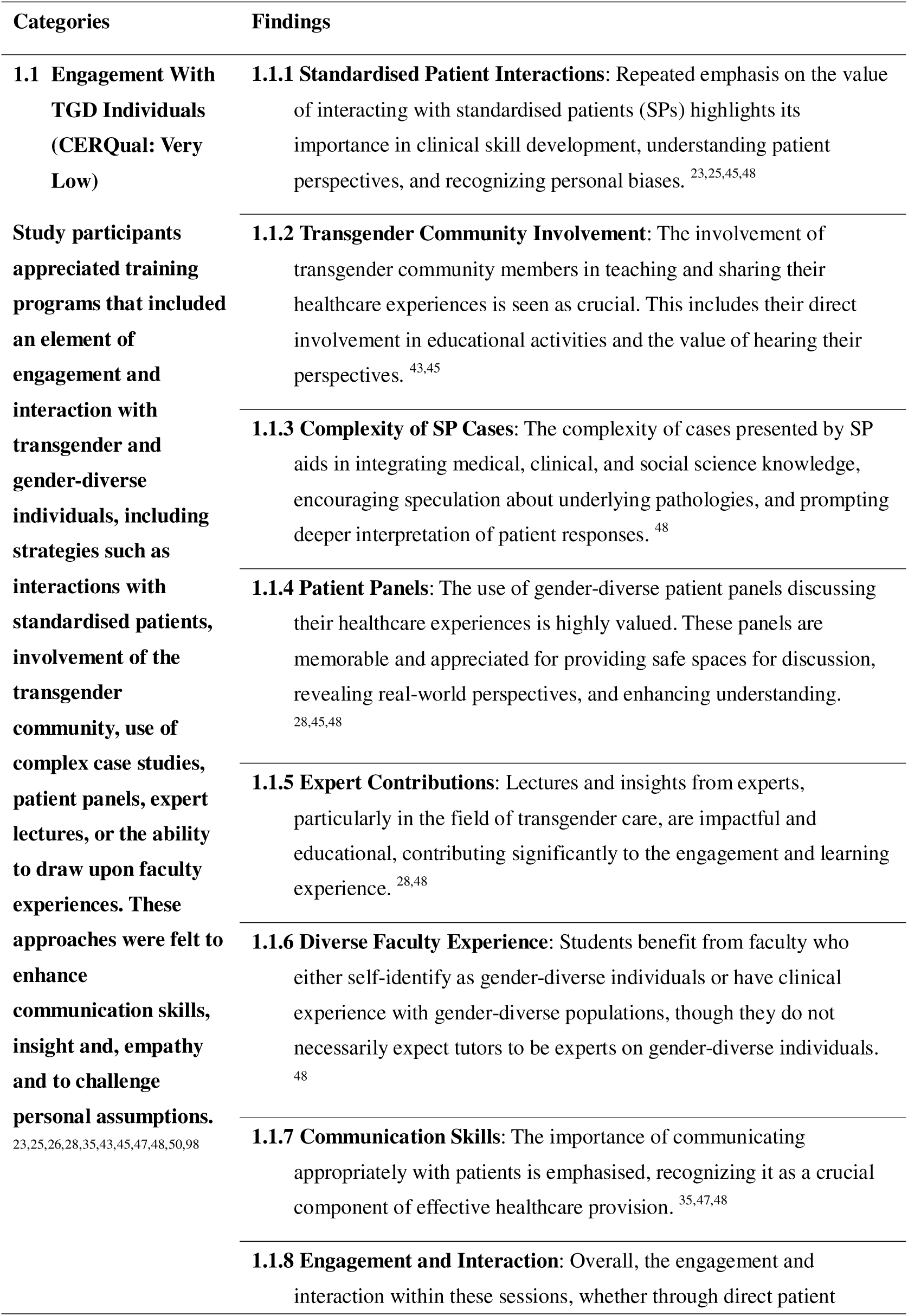
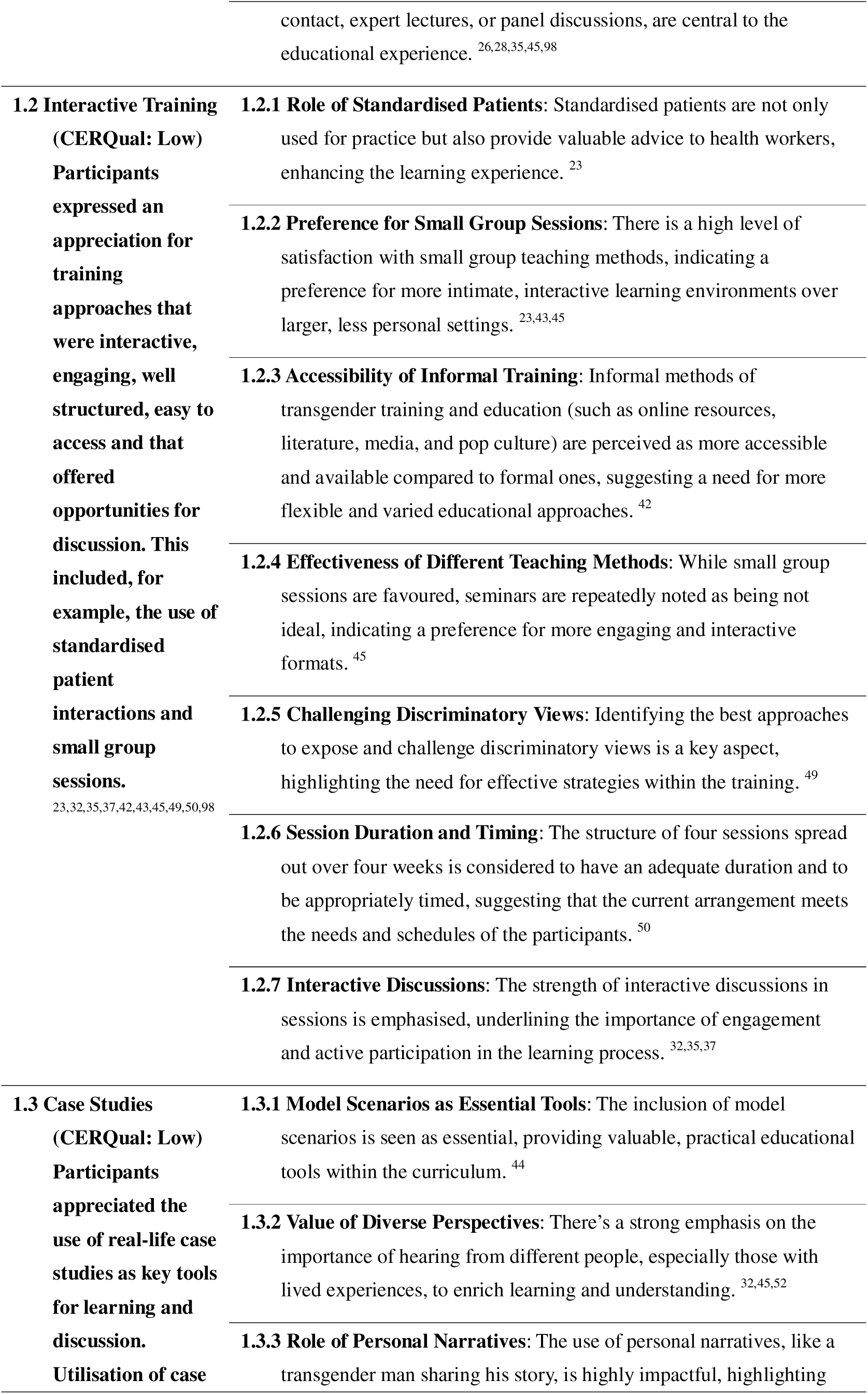
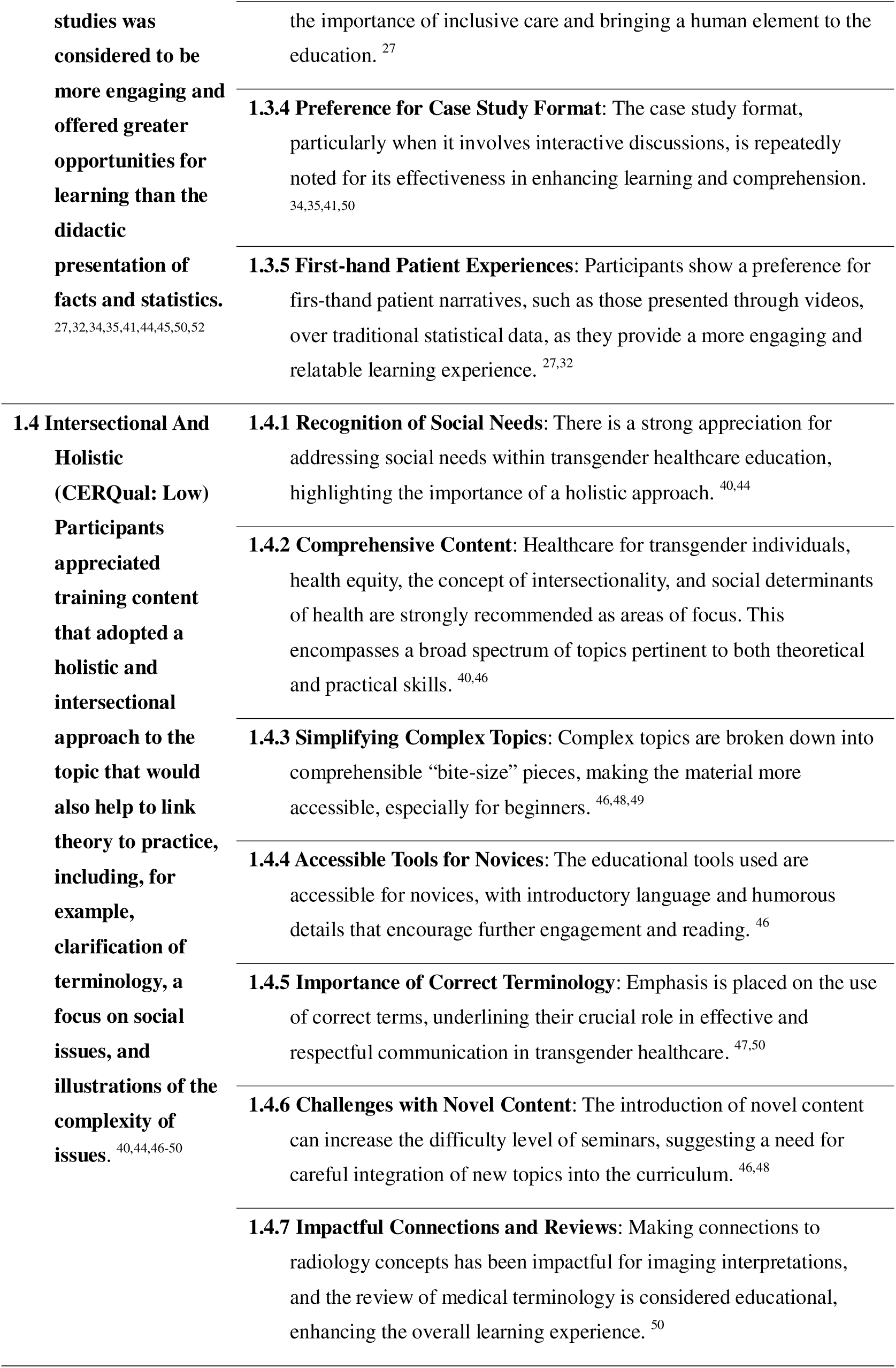
Categories and findings for synthesised finding 1: Training structure and content. Pedagogical preferences of health workers and students regarding training on inclusive and gender-affirming care.

| Categories | Findings |
| --- | --- |
| **1.1 Engagement With TGD Individuals (CERQual: Very Low)**<br>**Study participants appreciated training programs that included an element of engagement and interaction with transgender and gender-diverse individuals, including strategies such as interactions with standardised patients, involvement of the transgender community, use of complex case studies, patient panels, expert lectures, or the ability to draw upon faculty experiences. These approaches were felt to enhance communication skills, insight and, empathy and to challenge personal assumptions.**<br>23,25,26,28,35,43,45,47,48,50,98 | **1.1.1 Standardised Patient Interactions:** Repeated emphasis on the value of interacting with standardised patients (SPs) highlights its importance in clinical skill development, understanding patient perspectives, and recognizing personal biases.<sup>23,25,45,48</sup><br>**1.1.2 Transgender Community Involvement:** The involvement of transgender community members in teaching and sharing their healthcare experiences is seen as crucial. This includes their direct involvement in educational activities and the value of hearing their perspectives.<sup>43,45</sup><br>**1.1.3 Complexity of SP Cases:** The complexity of cases presented by SP aids in integrating medical, clinical, and social science knowledge, encouraging speculation about underlying pathologies, and prompting deeper interpretation of patient responses.<sup>48</sup><br>**1.1.4 Patient Panels:** The use of gender-diverse patient panels discussing their healthcare experiences is highly valued. These panels are memorable and appreciated for providing safe spaces for discussion, revealing real-world perspectives, and enhancing understanding.<sup>28,45,48</sup><br>**1.1.5 Expert Contributions:** Lectures and insights from experts, particularly in the field of transgender care, are impactful and educational, contributing significantly to the engagement and learning experience.<sup>28,48</sup><br>**1.1.6 Diverse Faculty Experience:** Students benefit from faculty who either self-identify as gender-diverse individuals or have clinical experience with gender-diverse populations, though they do not necessarily expect tutors to be experts on gender-diverse individuals.<sup>48</sup><br>**1.1.7 Communication Skills:** The importance of communicating appropriately with patients is emphasised, recognizing it as a crucial component of effective healthcare provision.<sup>35,47,48</sup><br>**1.1.8 Engagement and Interaction:** Overall, the engagement and interaction within these sessions, whether through direct patient |
|  | contact, expert lectures, or panel discussions, are central to the educational experience. <sup>26,28,35,45,98</sup> |
| **1.2 Interactive Training (CERQual: Low)**<br>**Participants expressed an appreciation for training approaches that were interactive, engaging, well structured, easy to access and that offered opportunities for discussion. This included, for example, the use of standardised patient interactions and small group sessions.**<br>23,32,35,37,42,43,45,49,50,98 | **1.2.1 Role of Standardised Patients:** Standardised patients are not only used for practice but also provide valuable advice to health workers, enhancing the learning experience. <sup>23</sup><br>**1.2.2 Preference for Small Group Sessions:** There is a high level of satisfaction with small group teaching methods, indicating a preference for more intimate, interactive learning environments over larger, less personal settings. <sup>23,43,45</sup><br>**1.2.3 Accessibility of Informal Training:** Informal methods of transgender training and education (such as online resources, literature, media, and pop culture) are perceived as more accessible and available compared to formal ones, suggesting a need for more flexible and varied educational approaches. <sup>42</sup><br>**1.2.4 Effectiveness of Different Teaching Methods:** While small group sessions are favoured, seminars are repeatedly noted as being not ideal, indicating a preference for more engaging and interactive formats. <sup>45</sup><br>**1.2.5 Challenging Discriminatory Views:** Identifying the best approaches to expose and challenge discriminatory views is a key aspect, highlighting the need for effective strategies within the training. <sup>49</sup><br>**1.2.6 Session Duration and Timing:** The structure of four sessions spread out over four weeks is considered to have an adequate duration and to be appropriately timed, suggesting that the current arrangement meets the needs and schedules of the participants. <sup>50</sup><br>**1.2.7 Interactive Discussions:** The strength of interactive discussions in sessions is emphasised, underlining the importance of engagement and active participation in the learning process. <sup>32,35,37</sup> |
| **1.3 Case Studies (CERQual: Low)**<br>**Participants appreciated the use of real-life case studies as key tools for learning and discussion.**<br>**Utilisation of case** | **1.3.1 Model Scenarios as Essential Tools:** The inclusion of model scenarios is seen as essential, providing valuable, practical educational tools within the curriculum. <sup>44</sup><br>**1.3.2 Value of Diverse Perspectives:** There's a strong emphasis on the importance of hearing from different people, especially those with lived experiences, to enrich learning and understanding. <sup>32,45,52</sup><br>**1.3.3 Role of Personal Narratives:** The use of personal narratives, like a transgender man sharing his story, is highly impactful, highlighting |
| studies was considered to be more engaging and offered greater opportunities for learning than the didactic presentation of facts and statistics.<br>27,32,34,35,41,44,45,50,52 | the importance of inclusive care and bringing a human element to the education. <sup>27</sup><br>**1.3.4 Preference for Case Study Format:** The case study format, particularly when it involves interactive discussions, is repeatedly noted for its effectiveness in enhancing learning and comprehension. <sup>34,35,41,50</sup><br>**1.3.5 First-hand Patient Experiences:** Participants show a preference for first-hand patient narratives, such as those presented through videos, over traditional statistical data, as they provide a more engaging and relatable learning experience. <sup>27,32</sup> |
| **1.4 Intersectional And Holistic (CERQual: Low) Participants appreciated training content that adopted a holistic and intersectional approach to the topic that would also help to link theory to practice, including, for example, clarification of terminology, a focus on social issues, and illustrations of the complexity of issues.**<br>40,44,46-50 | **1.4.1 Recognition of Social Needs:** There is a strong appreciation for addressing social needs within transgender healthcare education, highlighting the importance of a holistic approach. <sup>40,44</sup><br>**1.4.2 Comprehensive Content:** Healthcare for transgender individuals, health equity, the concept of intersectionality, and social determinants of health are strongly recommended as areas of focus. This encompasses a broad spectrum of topics pertinent to both theoretical and practical skills. <sup>40,46</sup><br>**1.4.3 Simplifying Complex Topics:** Complex topics are broken down into comprehensible “bite-size” pieces, making the material more accessible, especially for beginners. <sup>46,48,49</sup><br>**1.4.4 Accessible Tools for Novices:** The educational tools used are accessible for novices, with introductory language and humorous details that encourage further engagement and reading. <sup>46</sup><br>**1.4.5 Importance of Correct Terminology:** Emphasis is placed on the use of correct terms, underlining their crucial role in effective and respectful communication in transgender healthcare. <sup>47,50</sup><br>**1.4.6 Challenges with Novel Content:** The introduction of novel content can increase the difficulty level of seminars, suggesting a need for careful integration of new topics into the curriculum. <sup>46,48</sup><br>**1.4.7 Impactful Connections and Reviews:** Making connections to radiology concepts has been impactful for imaging interpretations, and the review of medical terminology is considered educational, enhancing the overall learning experience. <sup>50</sup> |

## 4 Discussion

A synthesis of 85 studies on intervention effectiveness and stakeholder preferences identified interactive methods—including standardized patient interactions, group discussions, and role-playing—as most effective. These approaches promote active engagement with transgender healthcare scenarios, enabling learners to practice empathetic communication and develop practical, clinically applicable skills. Integrating lived experiences through personal narratives or case studies further enhances relatability and person-centered understanding, challenging biases and fostering genuine, respectful care. Prioritizing such experiential learning, including direct TGD community engagement, can significantly strengthen the application of gender-affirming principles and improve training outcomes.

Based on an analysis of values and preferences, both experts and participants expressed concern about the limitations of single-session training. They emphasized the value of longitudinal or repeated training, which allows healthcare workers and students to revisit complex topics, internalize gender-affirming principles, and progressively strengthen practical skills. Such an approach supports deeper and more lasting understanding, offers opportunities to address evolving challenges, and adapts to new evidence or social norms.

However, this review found that most current training consists of short-term, one-off sessions—such as single lectures—without structured follow-up or reinforcement. This raises concerns about the long-term durability of attitude and behavior change toward TGD individuals. Moreover, the evidence base lacks studies specifically evaluating the long-term effects of repeated training, underscoring a critical gap in both research and program design. The absence of longitudinal reinforcement not only challenges the sustainability of outcomes but also points to a key area for future research.

Finally, holistic programs that integrate clinical content with social determinants of health and equity provide a more comprehensive perspective. By addressing systemic issues, they prepare practitioners to navigate the complex realities TGD individuals face, fostering more inclusive care. Together, practical engagement, sustained learning, and comprehensive content form the foundation of effective TGD education.

### 4.1 Strengths and weaknesses

This review demonstrates several methodological strengths. The study was conducted and reported in accordance with rigorous PRISMA guidelines and was prospectively registered, ensuring transparency. The search strategy was exceptionally comprehensive, encompassing 13 major databases without language or geographic restrictions, thereby minimizing selection bias and enhancing the global relevance of the findings. The employment of a dual-reviewer process for screening, selection, and data extraction, with discrepancies resolved through consensus or third-party adjudication, significantly bolstered the reliability of the study inclusion and data collection. A key strength is the integrated mixed-methods approach, which combined a quantitative meta-analysis of intervention effectiveness across six key outcomes with a qualitative thematic synthesis of stakeholder values and preferences. This dual perspective provides a more nuanced understanding of both the measurable impact and the contextual factors influencing training programs for transgender and gender-diverse healthcare.

This review has several notable limitations that should be considered when interpreting the findings. The evidence base is notably constrained, as data were available for only six outcomes, all of which were downgraded due to a substantial risk of bias, inconsistency, and imprecise confidence intervals, thereby reducing the overall certainty of the evidence. Furthermore, the generalizability of the results is limited, as most included studies were conducted in high-income countries and focused on pre-service students, whose responses may not translate to practicing healthcare workers in diverse global contexts. Critical gaps remain, including a lack of focus on non-clinical staff and on the specific complexities of gender-affirming care. Finally, the short duration of interventions and the absence of long-term follow-up assessments compromise the robustness of conclusions regarding the sustained impact of the training in real-world settings.

## 5 Conclusion

This study is the first comprehensive systematic review to assess the impact of education and training interventions aimed at health workers and students to promote inclusive care and GAC to TGD people. We found that, compared to the baseline (i.e., no targeted education and training), educational interventions generally tended to improve knowledge, attitudes, skill, general competence, discrimination, and comfort of health workers in providing care and services for TGD people. However, the certainty of evidence was low, limiting the strength of conclusions.^99^ Further, although there were some positive trends, due to the lack of longitudinal assessments, and patient-reported outcomes, we remain uncertain whether the observed changes represent a clinically meaningful improvement in the long term. Future studies are necessary to validate these findings and clarify their practical implications.

## Acknowledgements

**Contributors:** we extend sincere gratitude to Dr. Maeve Brito de Mello, Siobhan Fitzpatrick, Dr. Anna Coates, and Dr. Karel Blondeel for their technical guidance and coordination. We would like to thank Professor Catrin Evans for her input on GRADE-CERQual assessment. We also thank Dr. Nandi Siegfried for her critical methodological insights, which ensured rigor and adherence to best practices, and Dr. Asa Radix for refining the review’s clinical relevance. Their collective expertise greatly strengthened the credibility and accuracy of this work.

## Prior presentations

Not applicable.

## Ethics statement

Not applicable.

## Data availability statements

The datasets during and/or analyzed during the current study available from the corresponding author on reasonable request.

## Author Contributions

JX, XY and ZZ conceived and designed this review. JX, ZY, ZZ, and XY performed initial pilot screening. JX, XY, ZY, ZZ, TTH, GWZ, MQX, QS, XXZ and ZZ screened and reviewed full texts for eligibility, extracted the data and completed the quality assessment. JS performed literature search and reference management. JX, XY, ZY, ZZ, resolved any conflicts in quality assessment. ZY, JX, ZZ, QS, GWZ, ES prepared the tables and figures and conducted the data analysis. JX, ZZ, ZY, XY, ES conducted data interpretation and drafted the first draft of the manuscript. All authors approved the project plan and reviewed and revised the final manuscript before submission.

## Appendix 1: Search strategy

### Electronic databases search strategy

**Review question:** Do education and/or training interventions on inclusive and/or gender-affirming care for health workers and students lead to inclusive and gender-affirmative care for trans and gender-diverse people?

**Initial search date**: 30/05/2023

**Search results summary**: 18892 references sourced from the databases below

- PubMed: 2694
- MEDLINE (OVID): 2773
- Embase (OVID): 2753
- Web of Science: 2283
- Cochrane CENTRAL: 72
- Emcare (OVID): 1798
- PsycINFO (EBSCOhost): 2464
- Social Services Abstracts (ProQuest): 512
- Sociological Abstracts (ProQuest): 859
- Academic Search Premier (EBSCOhost): 1904
- Psychology and Behavioral Sciences Collection (EBSCOhost): 308
- ERIC (ProQuest): 420
- LILACS: 52

**Update search date:** 07/03/2024

**Search results summary**: 8145 references, from which 932 new and unique, sourced from the databases below

- PubMed: 3140 - 386 new&unique
- MEDLINE (OVID): 3205 - 13 new&unique
- Embase (OVID): 3046 - 110 new&unique
- Web of Science: 2714 - 117 new&unique
- Cochrane CENTRAL: 82 - 6 new&unique
- Emcare (OVID): 2101 - 43 new&unique
- PsycINFO (EBSCOhost): 2680 - 85 new&unique
- Social Services Abstracts (ProQuest): 564 - 21 new&unique
- Sociological Abstracts (ProQuest): 507 - 27 new&unique
- Academic Search Premier (EBSCOhost): 2154 - 90 new&unique
- Psychology and Behavioral Sciences Collection (EBSCOhost): 341 - 0 new&unique
- ERIC (ProQuest): 479 - 28 new&unique
- LILACS: 60 - 6 new&unique

Three subject components:

- Health Workers
- Education
- Trans and gender-diverse people

#### PubMed

http://www.ncbi.nlm.nih.gov/pubmed?otool=leiden

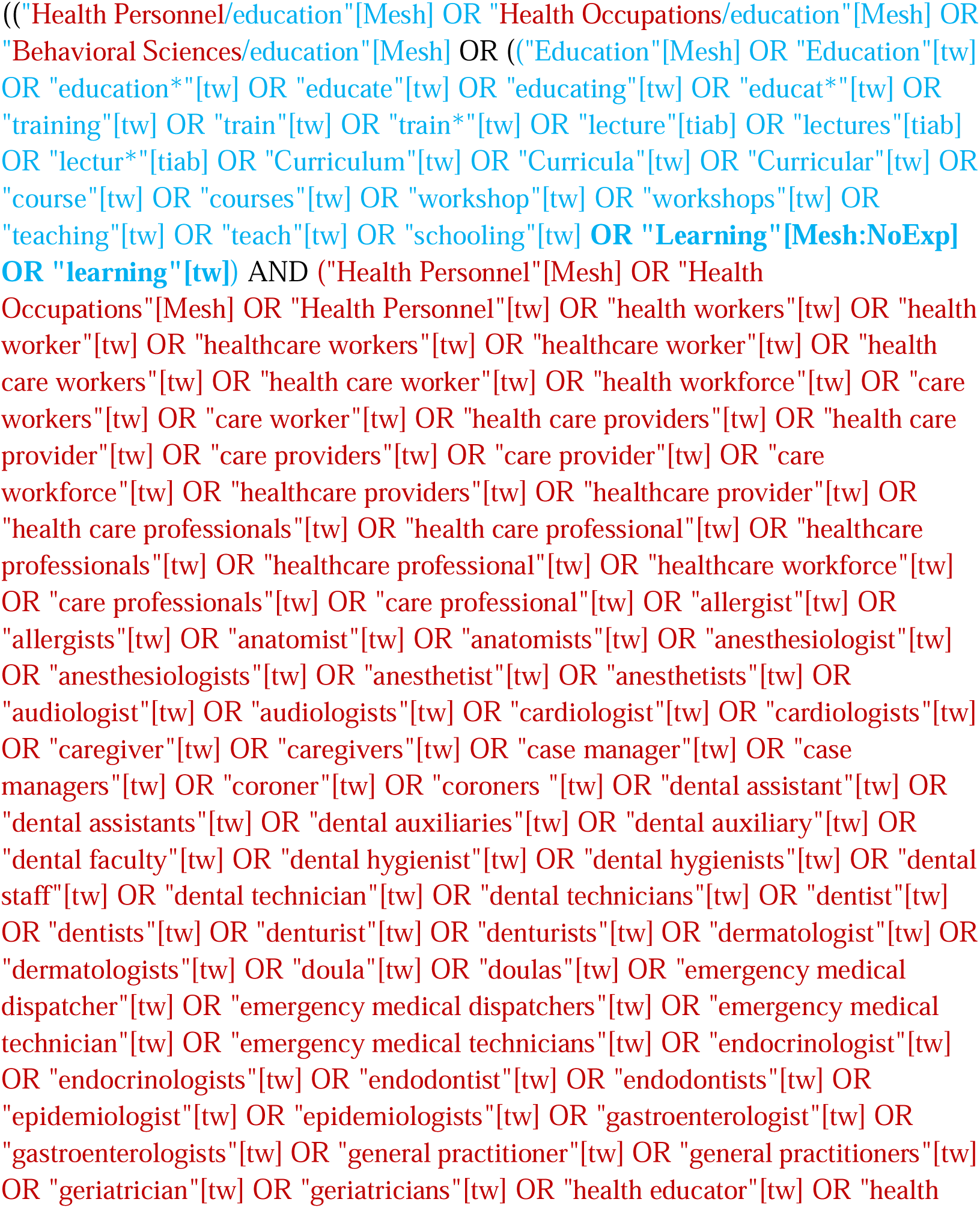
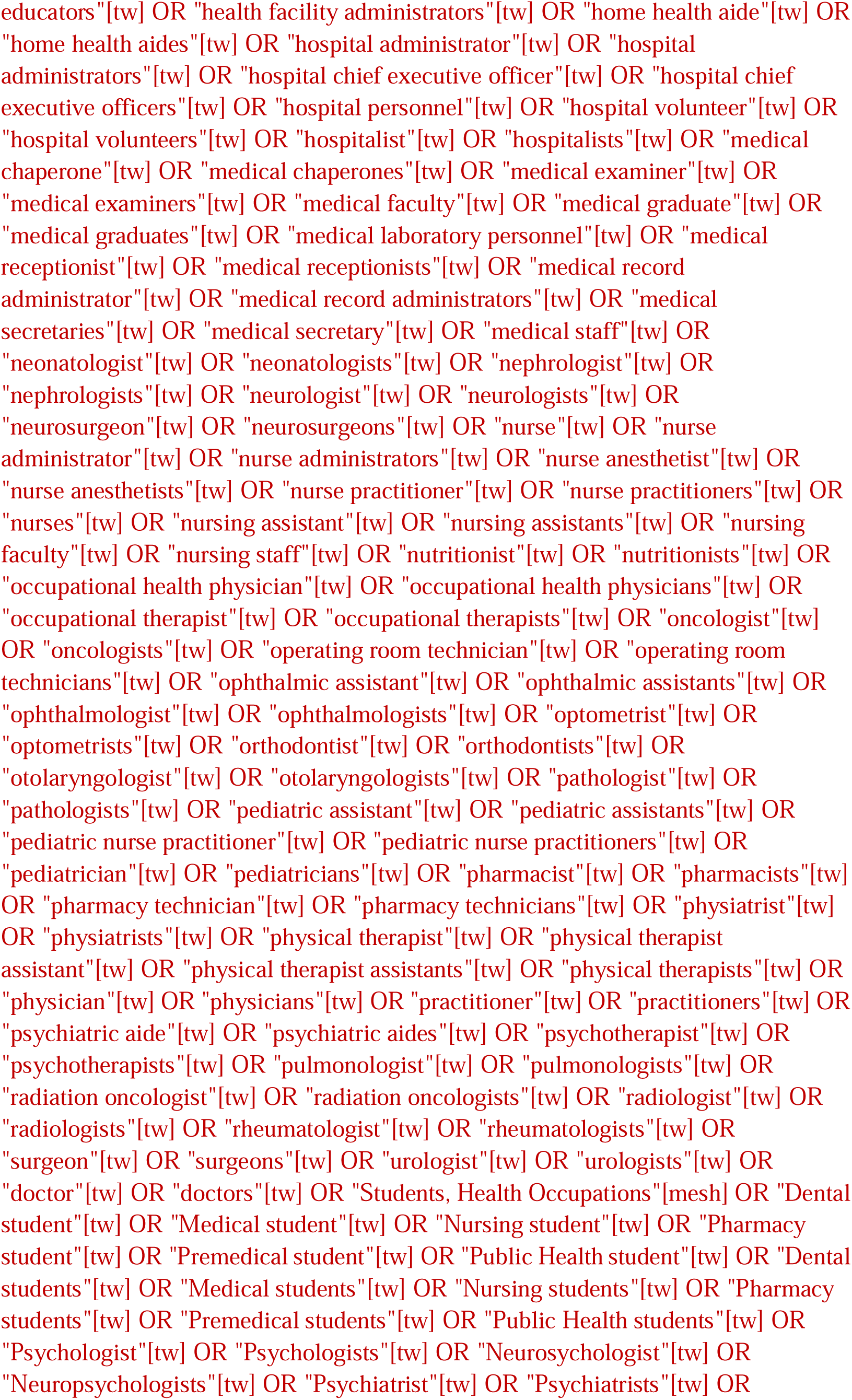
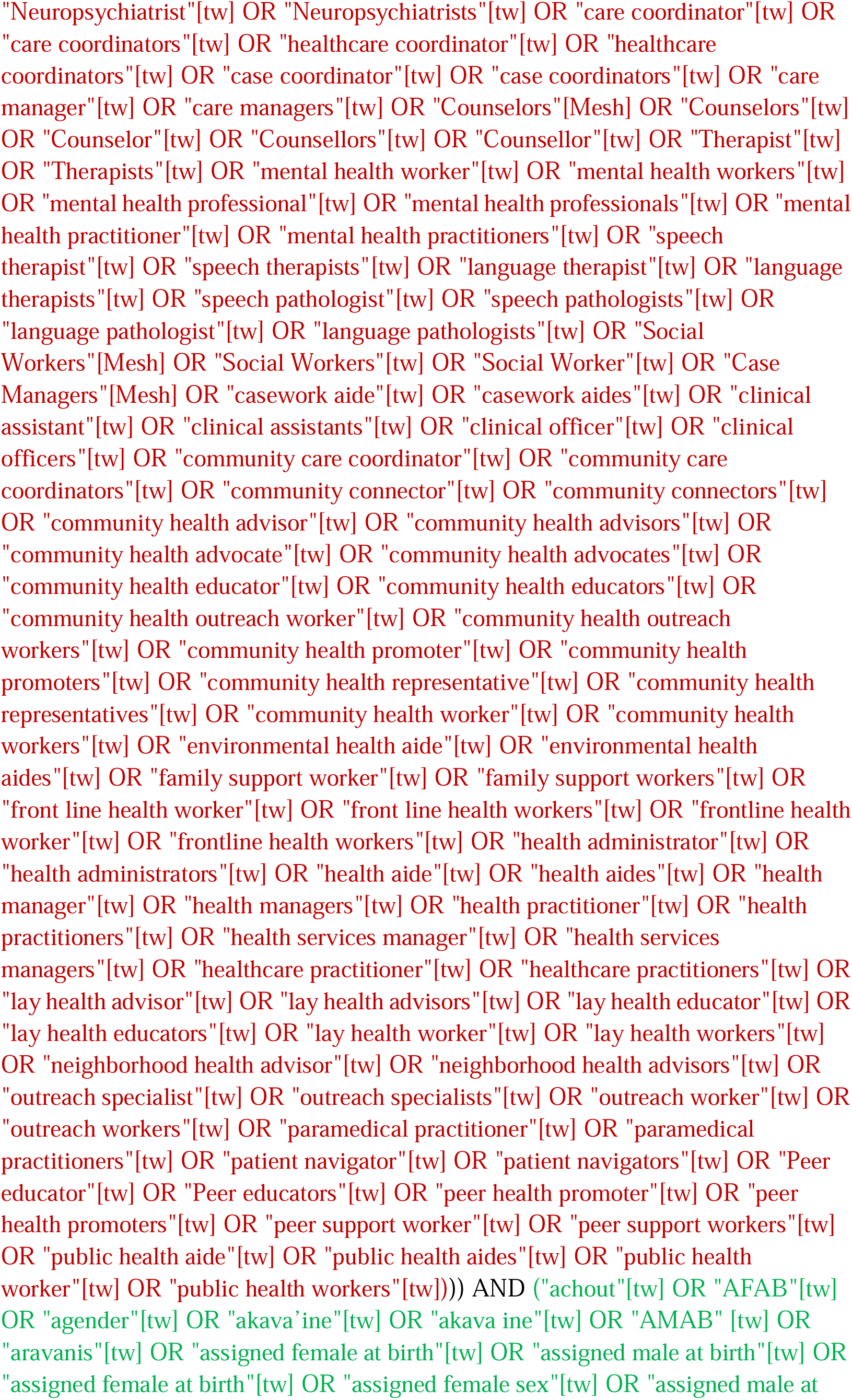
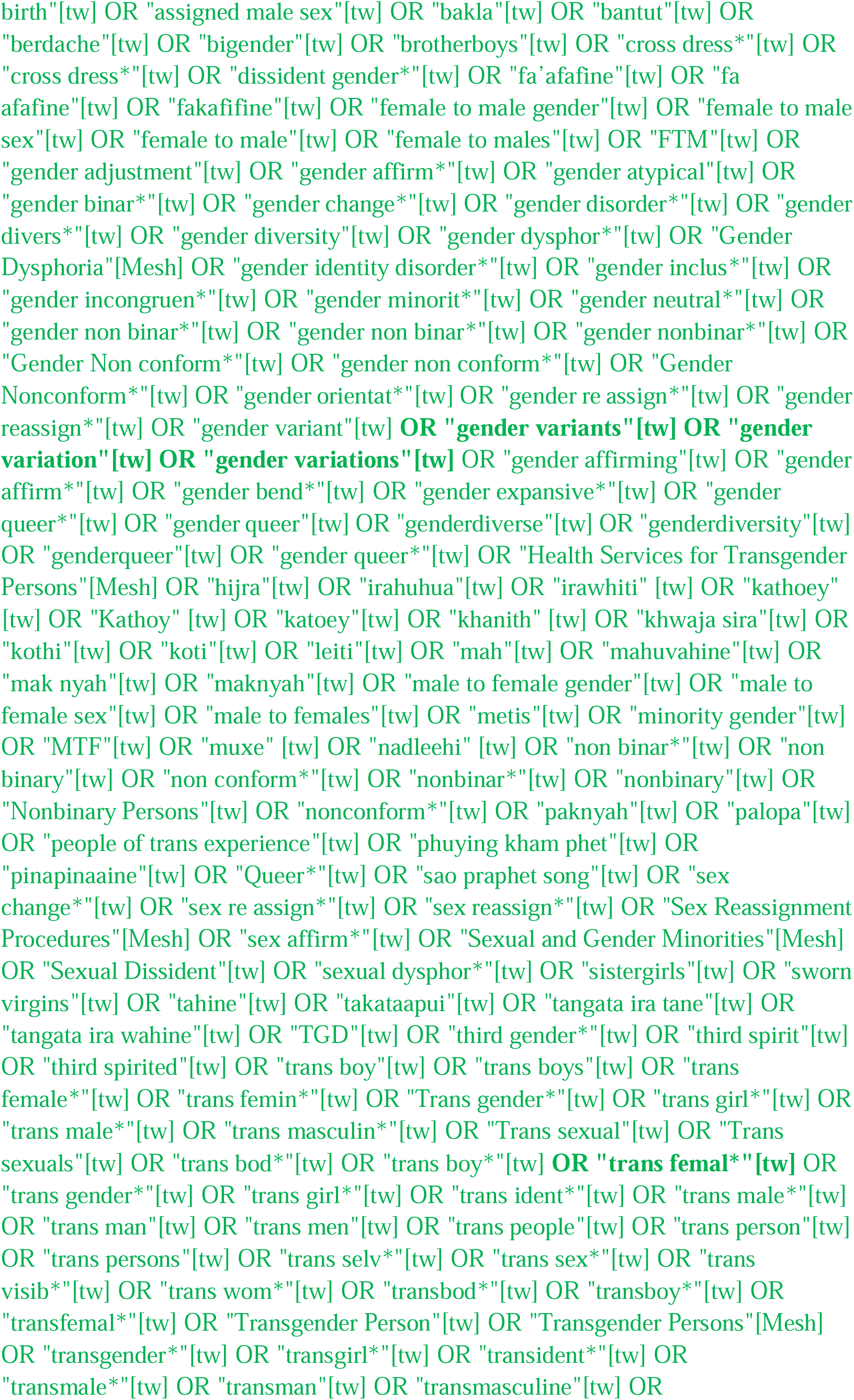
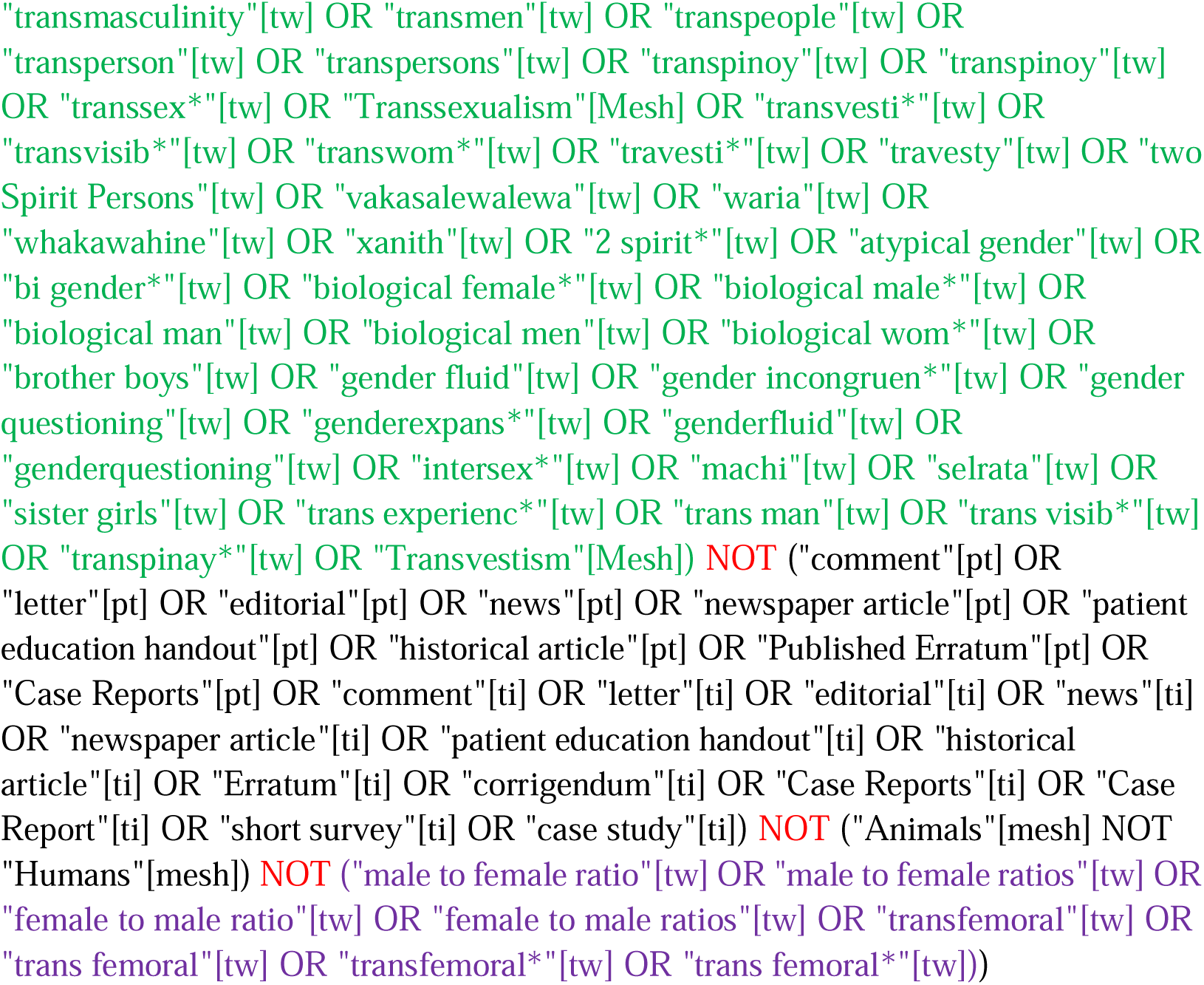

#### MEDLINE via OVID

http://gateway.ovid.com/ovidweb.cgi?T=JS&MODE=ovid&NEWS=n&PAGE=main&D=medall

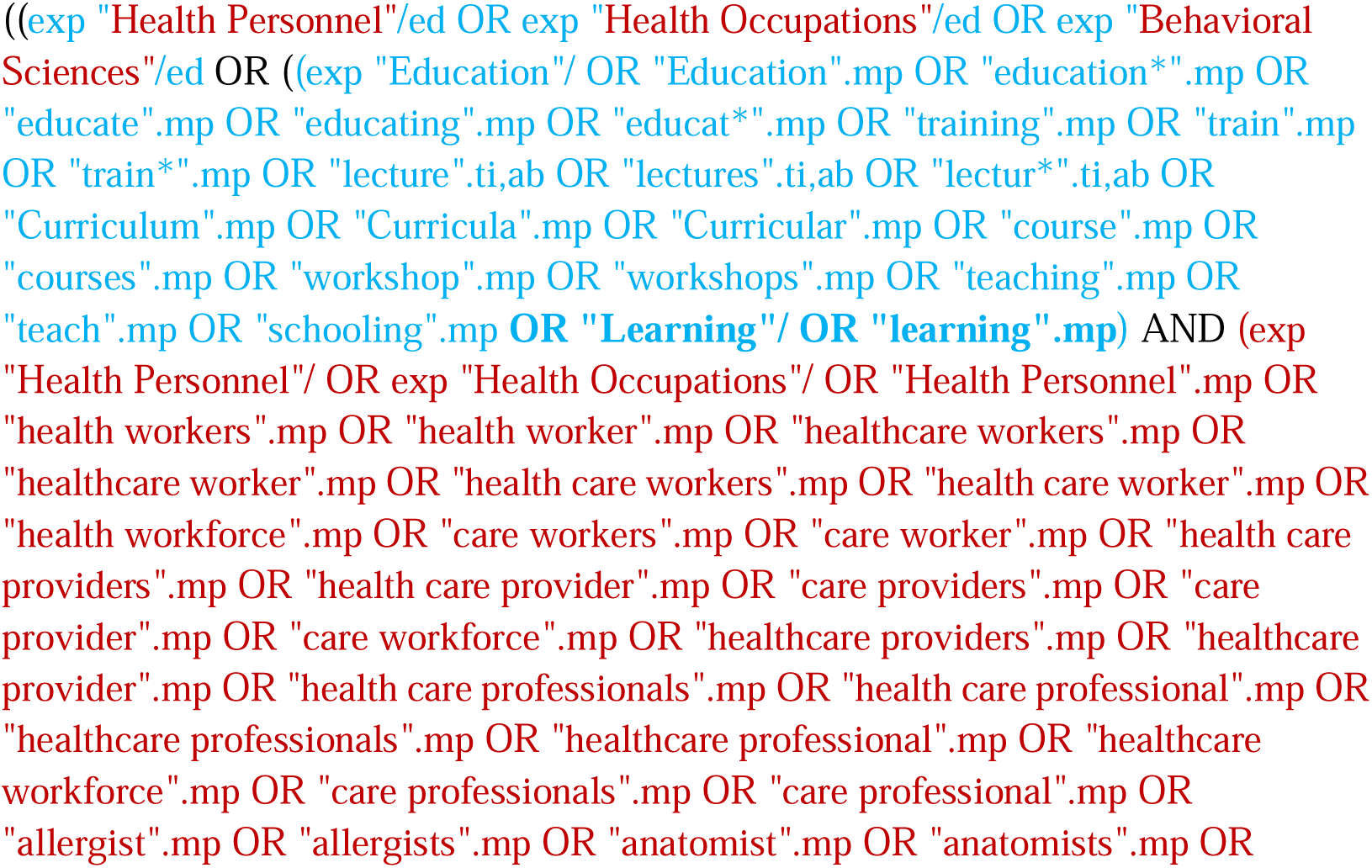
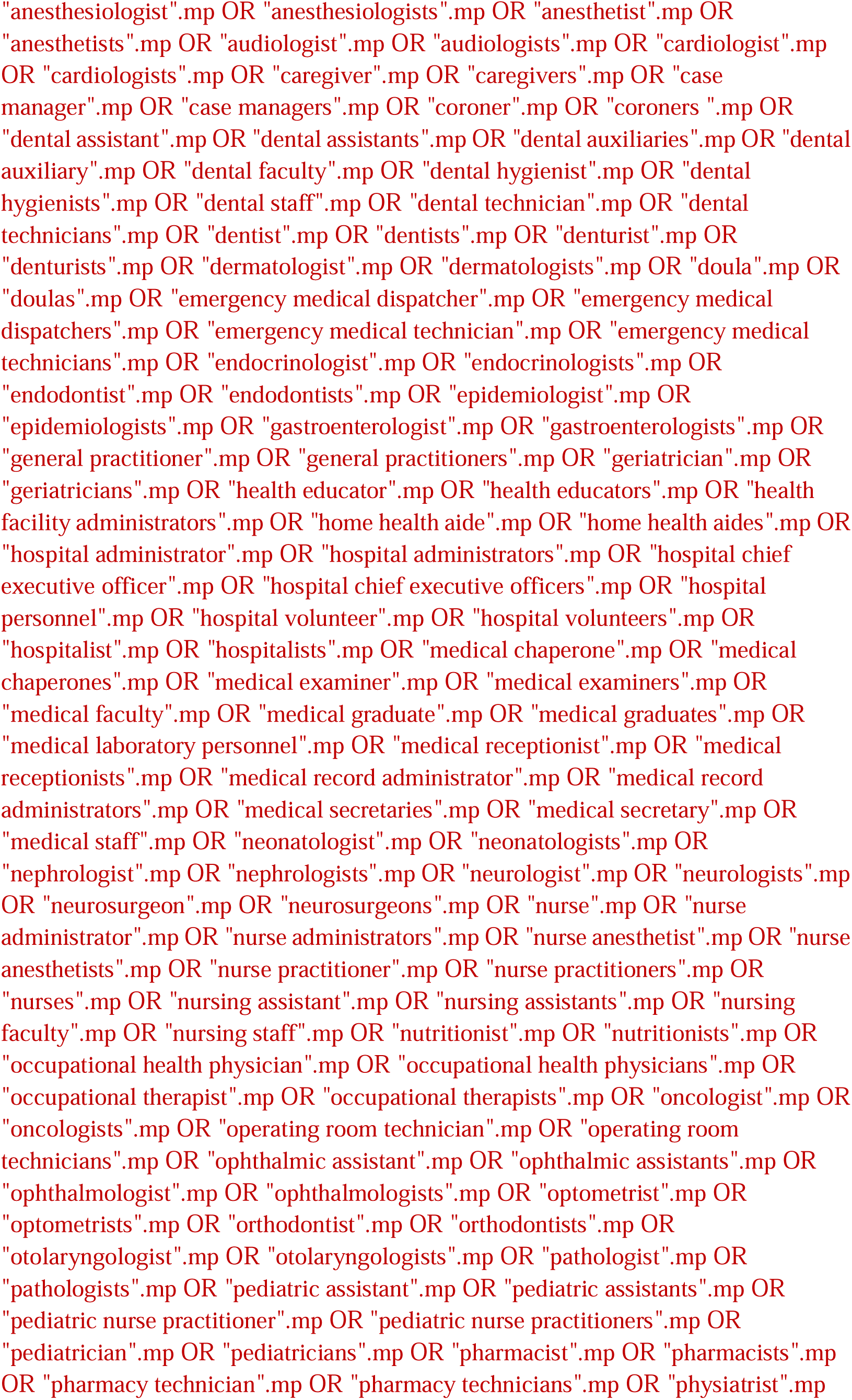
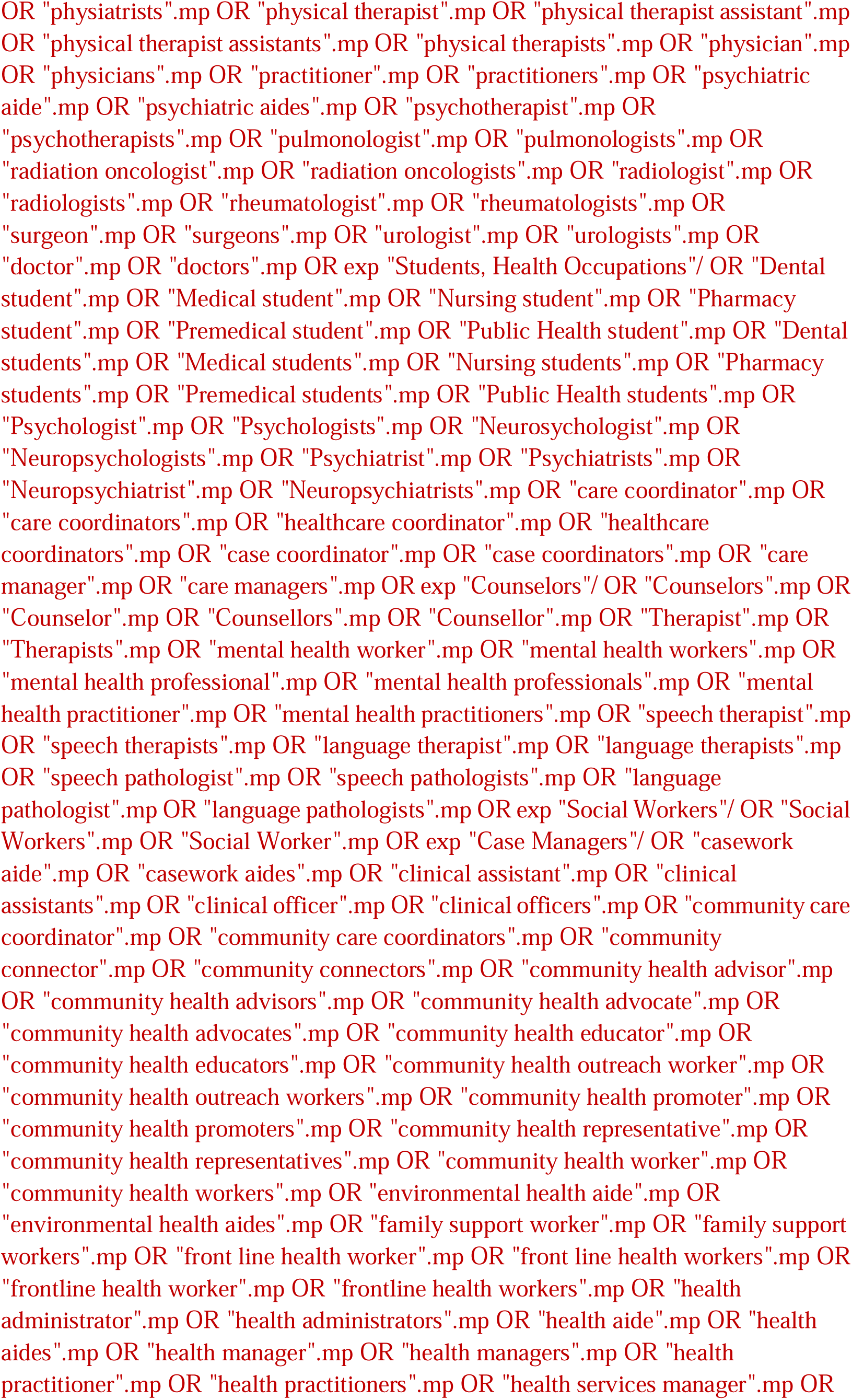
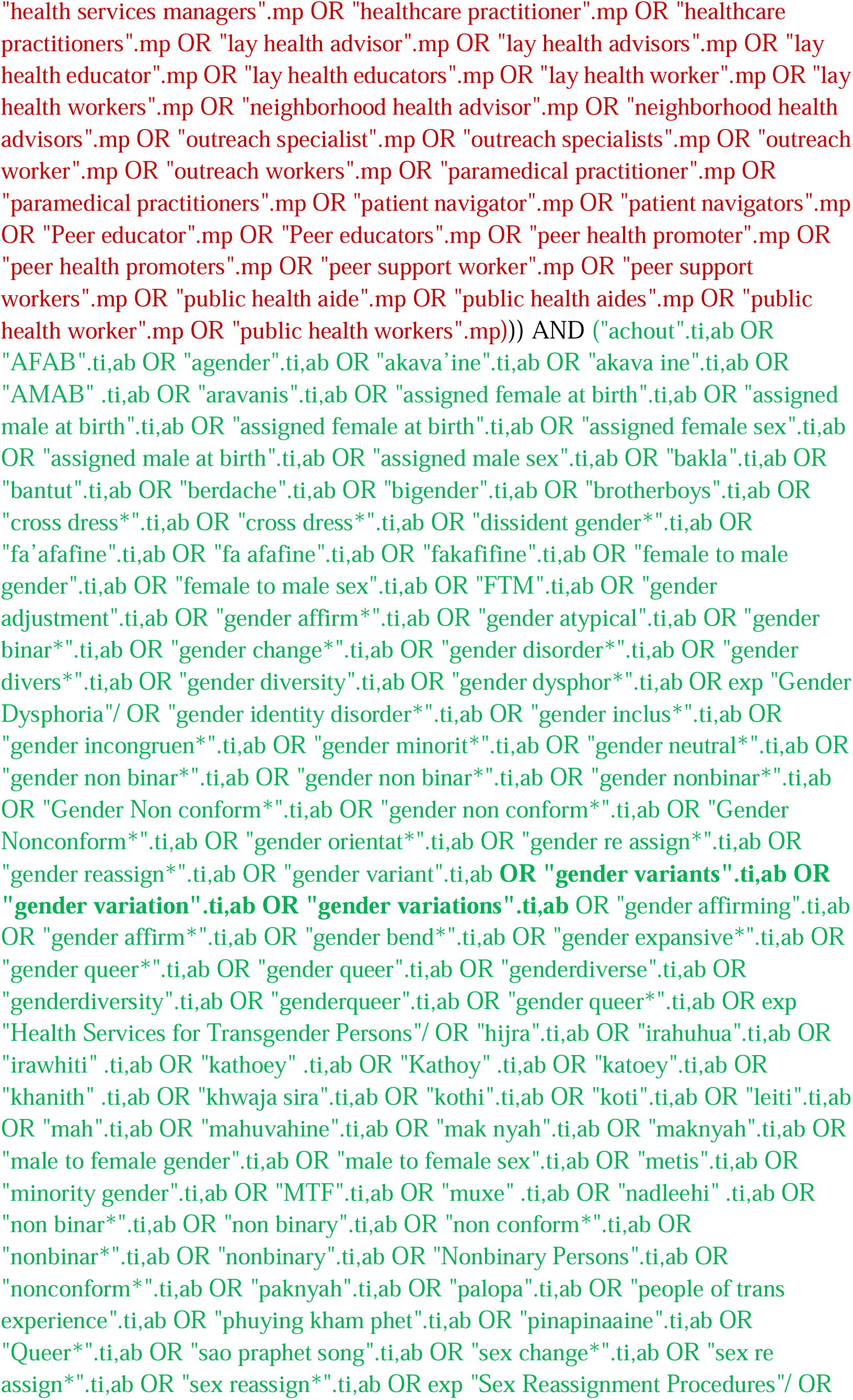
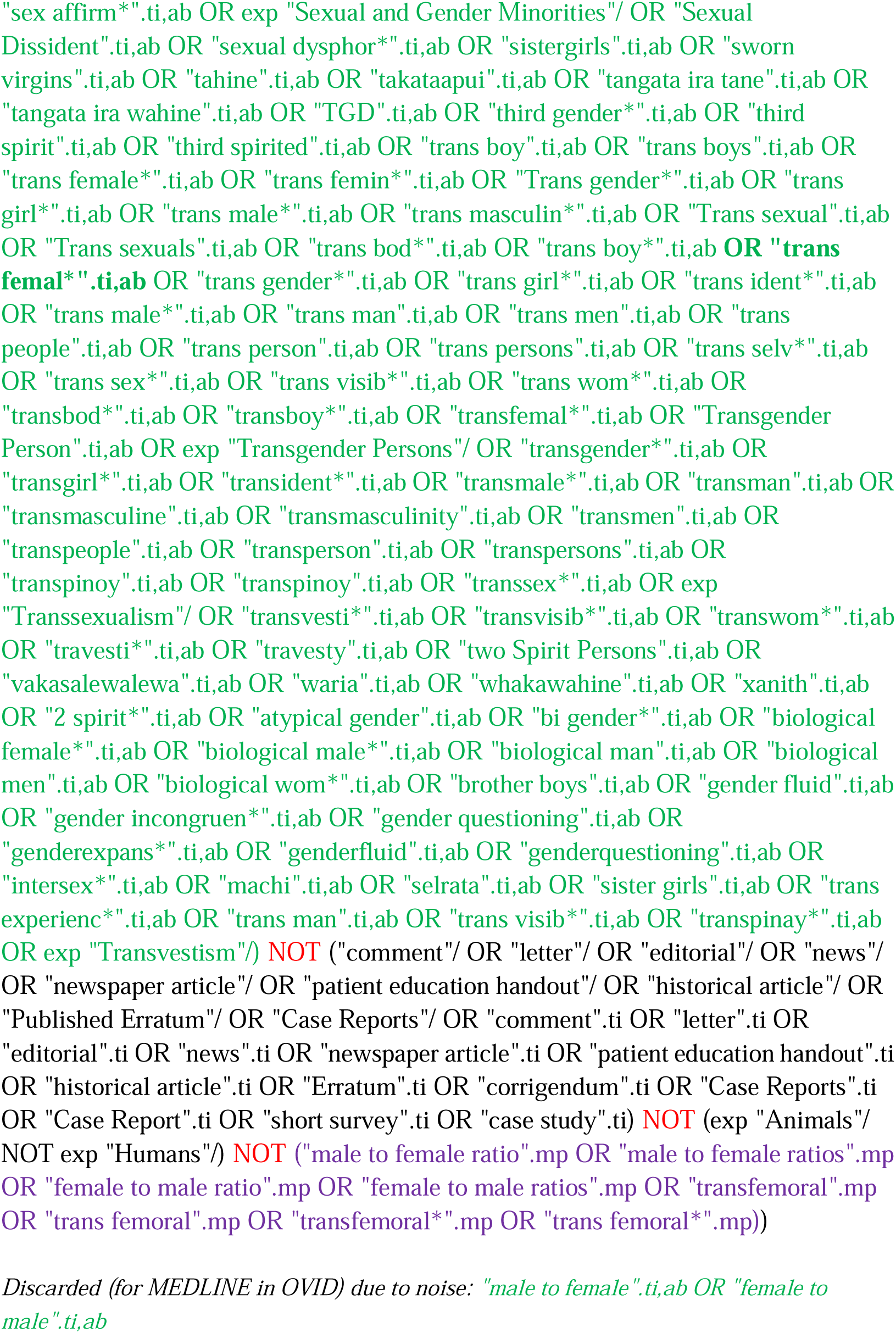

#### Embase

http://ovidsp.ovid.com/ovidweb.cgi?T=JS&PAGE=main&MODE=ovid&D=oemezd

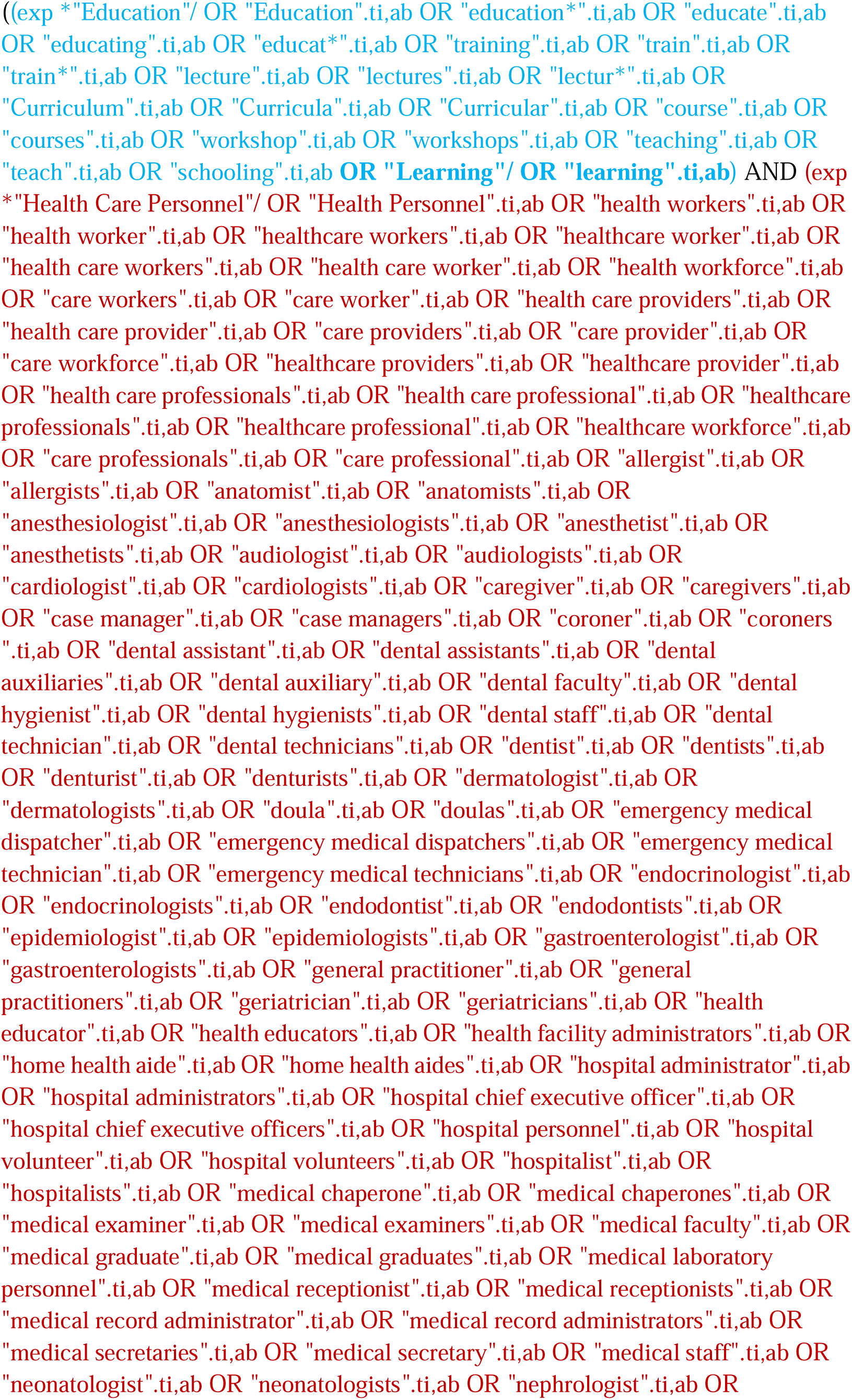
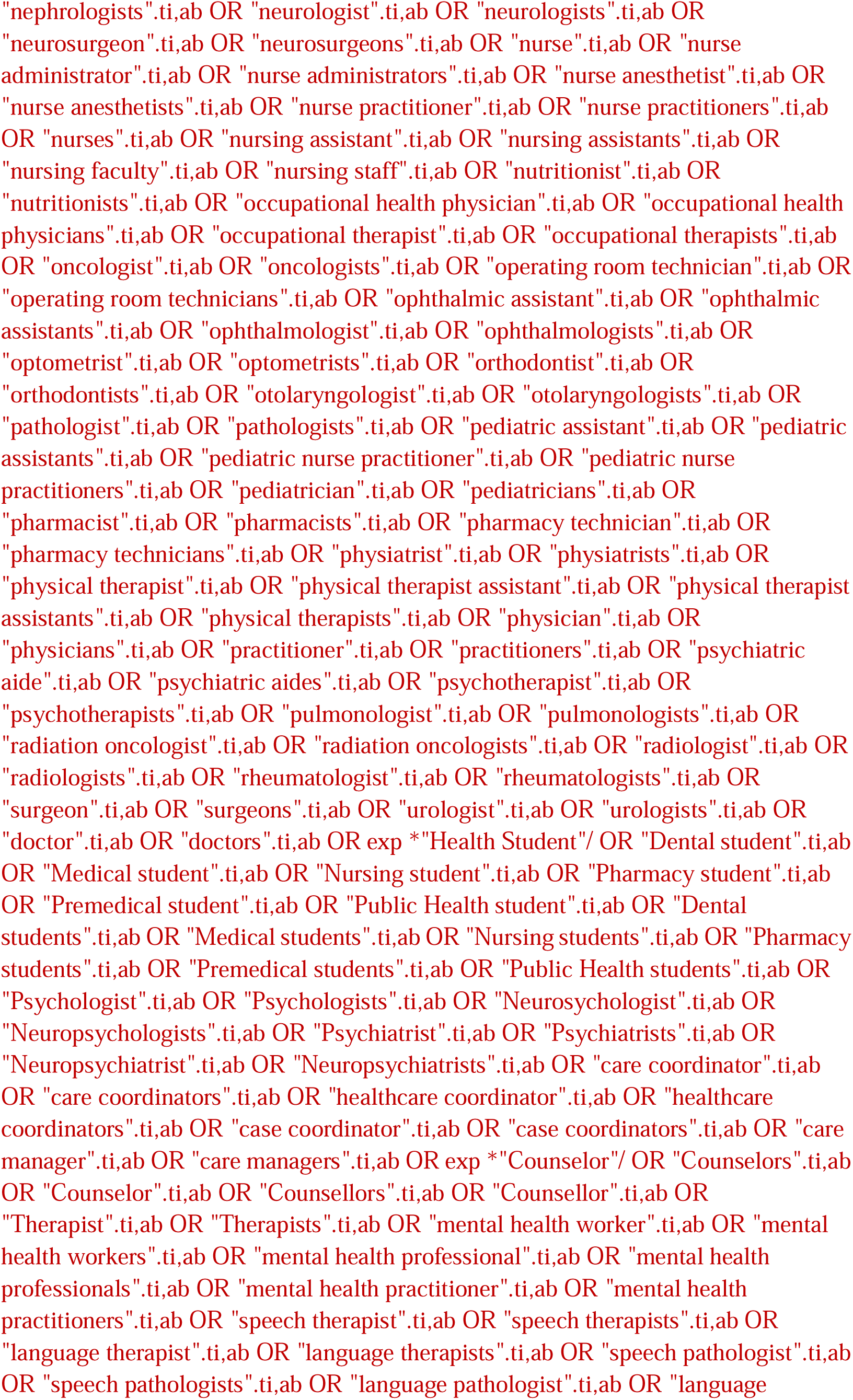
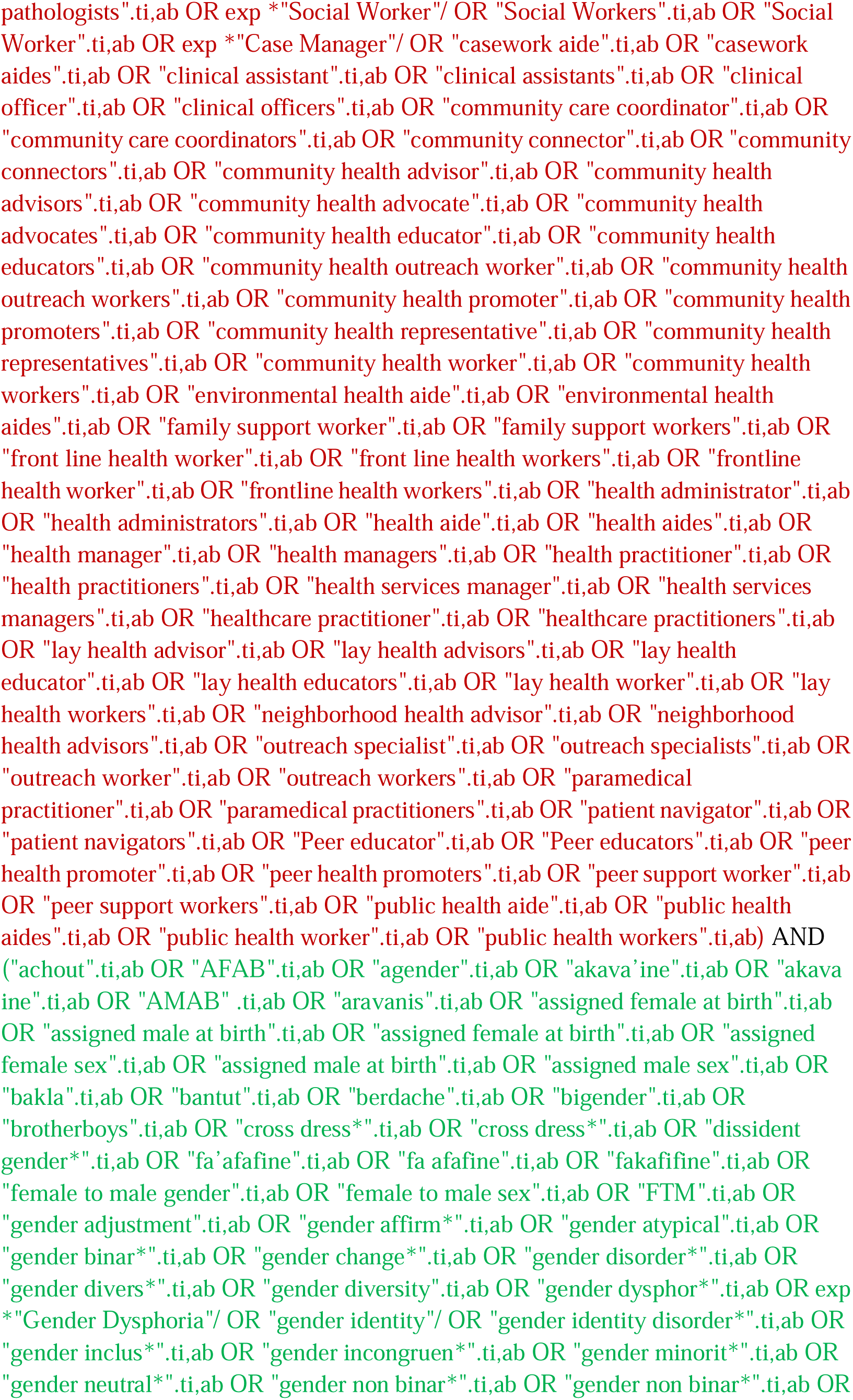
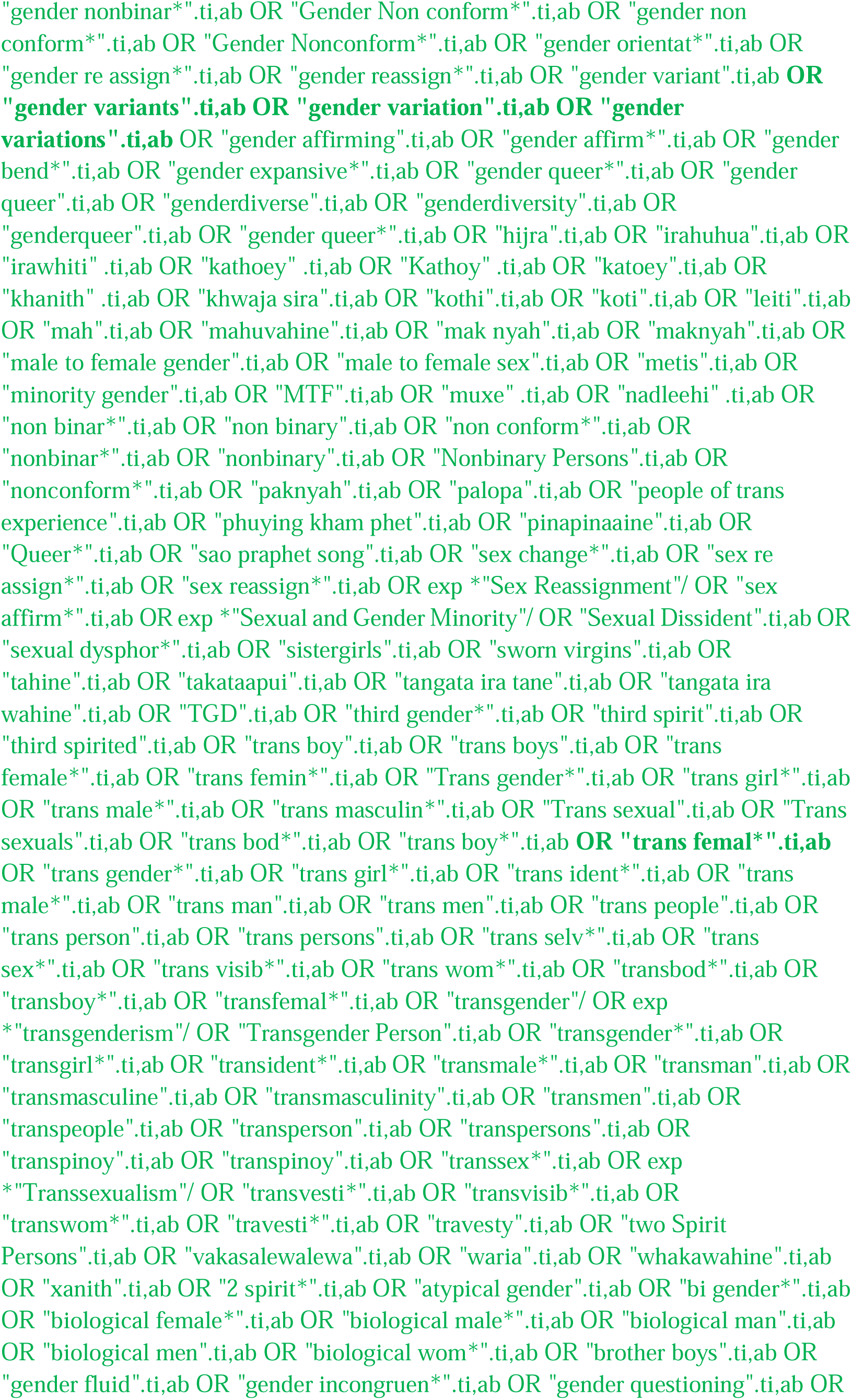
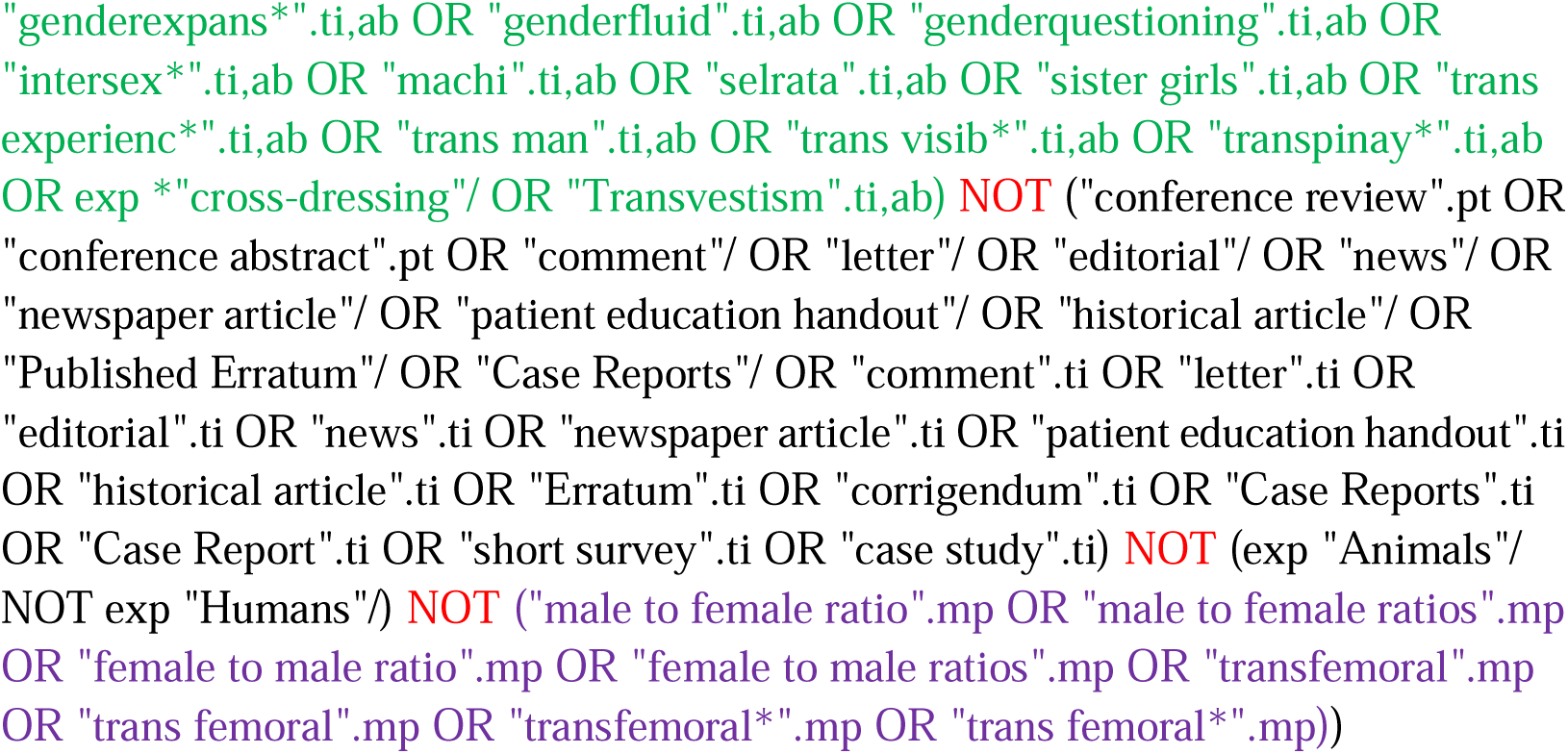

#### Web of Science

http://isiknowledge.com/wos

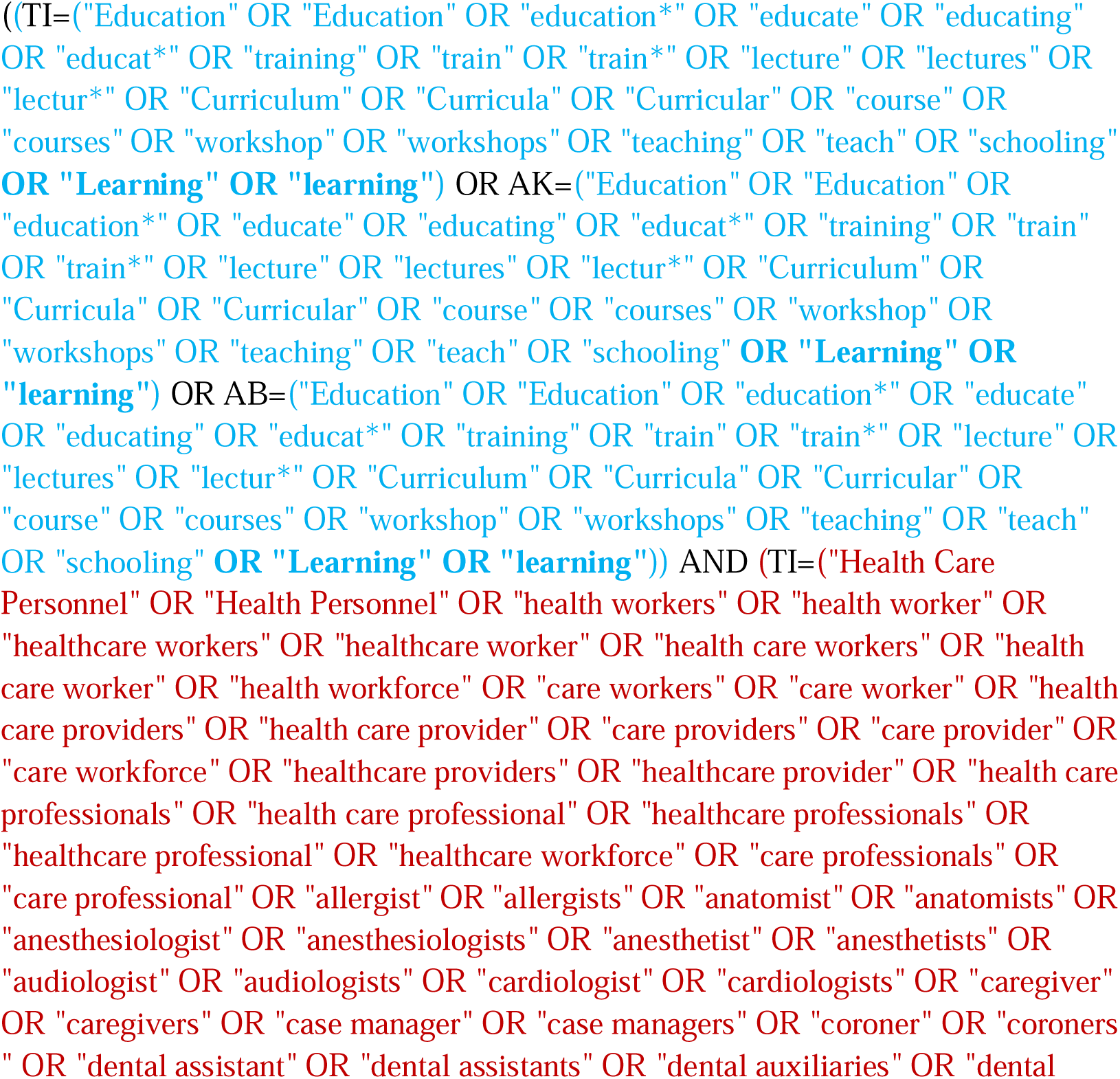
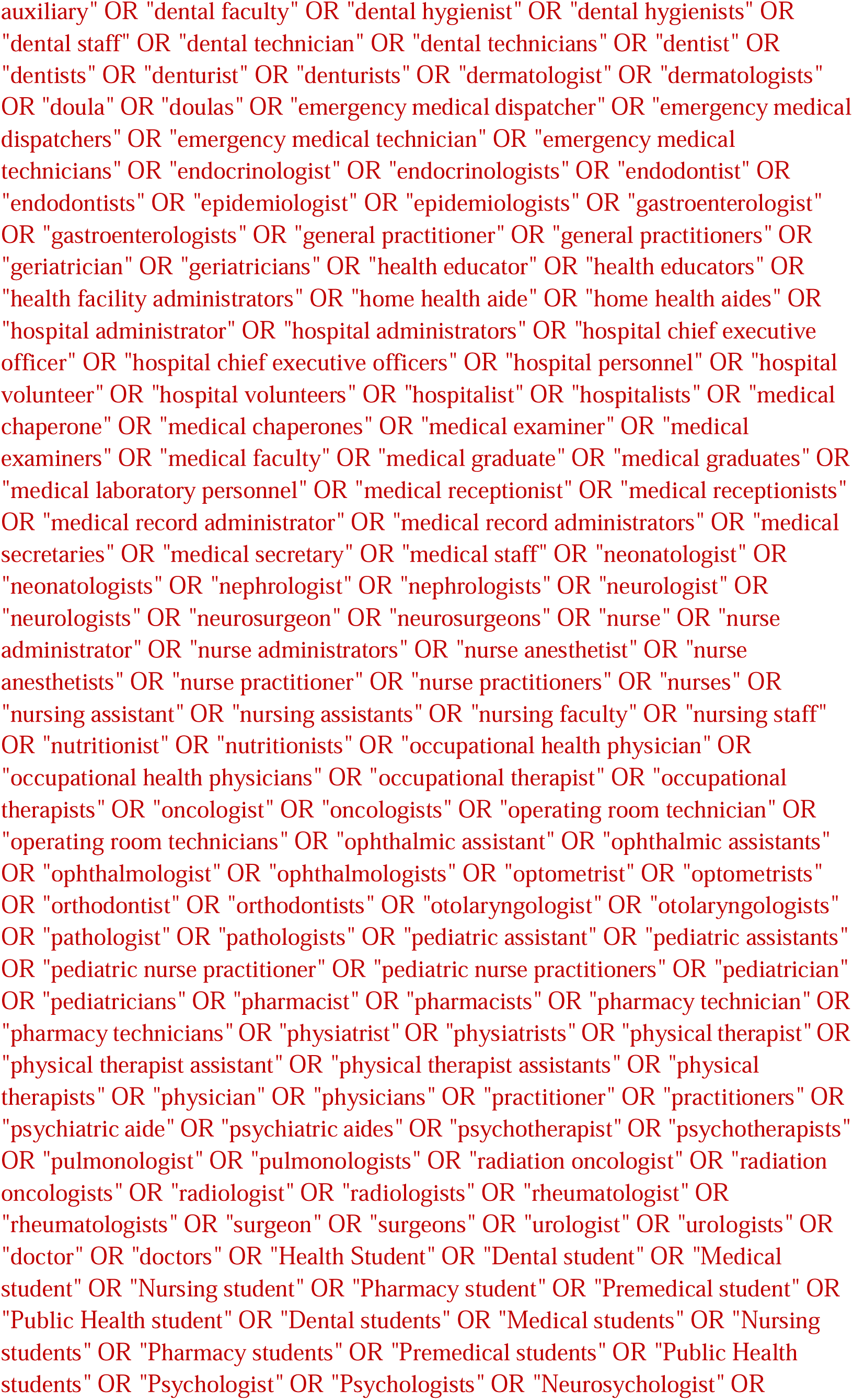
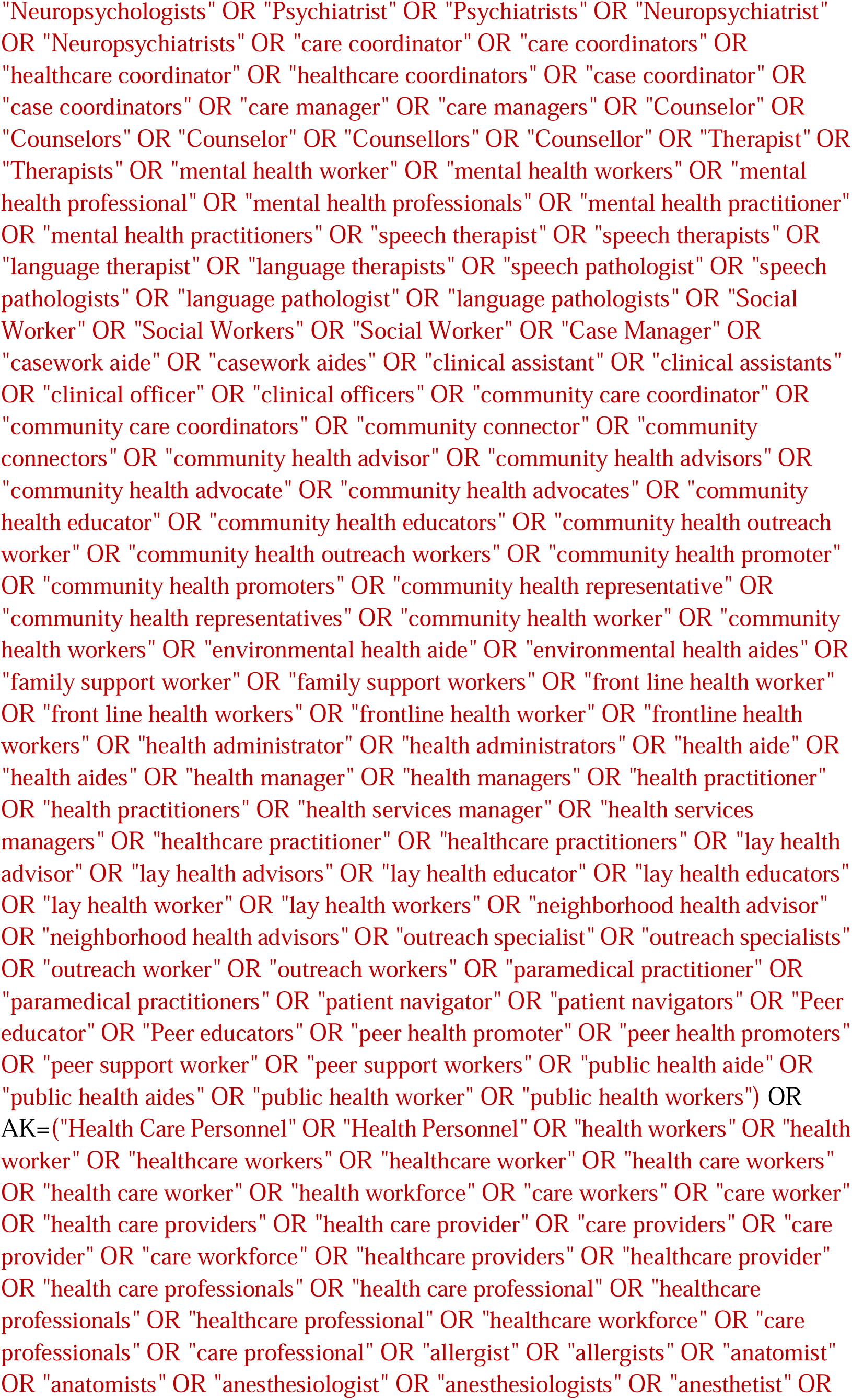
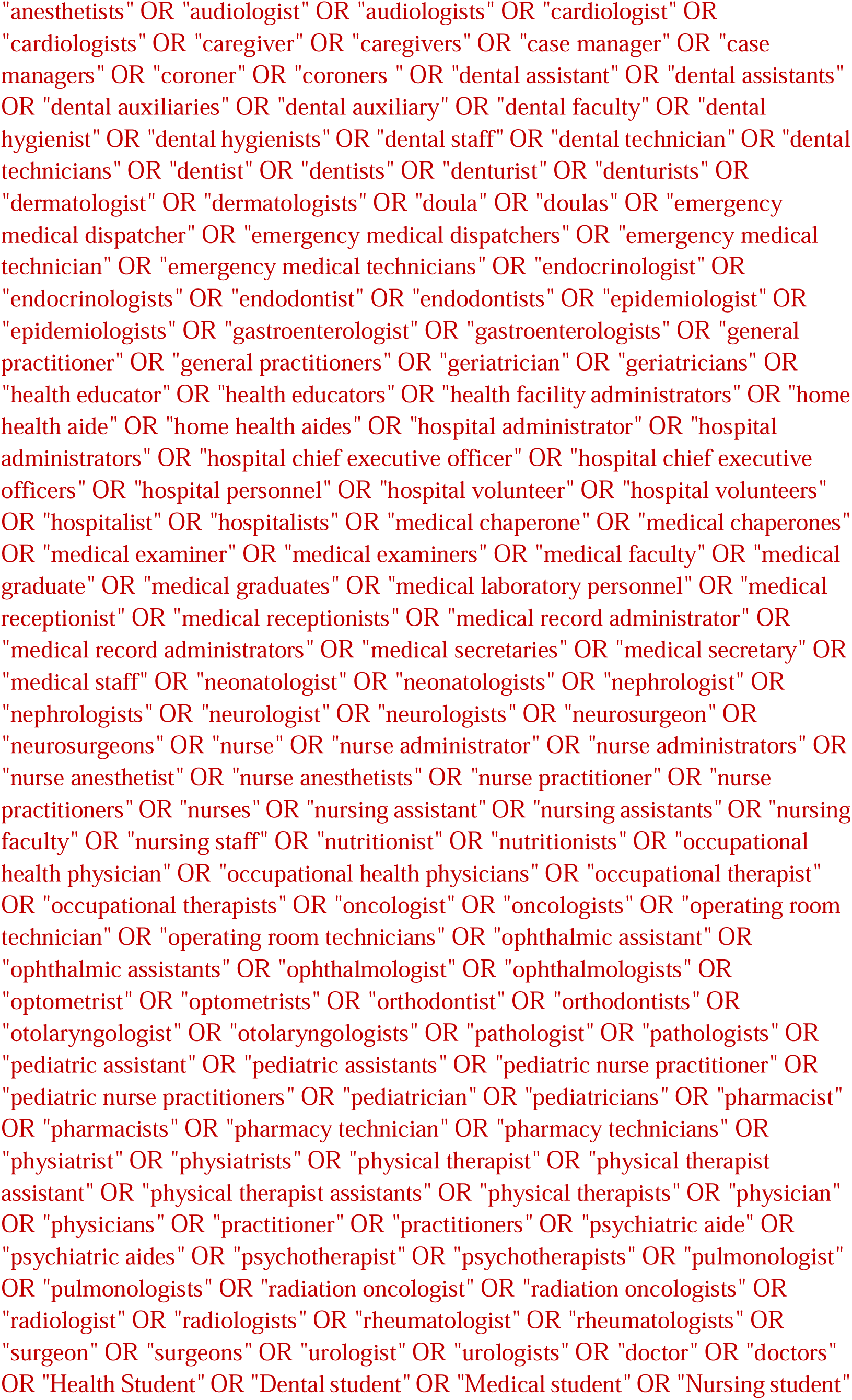
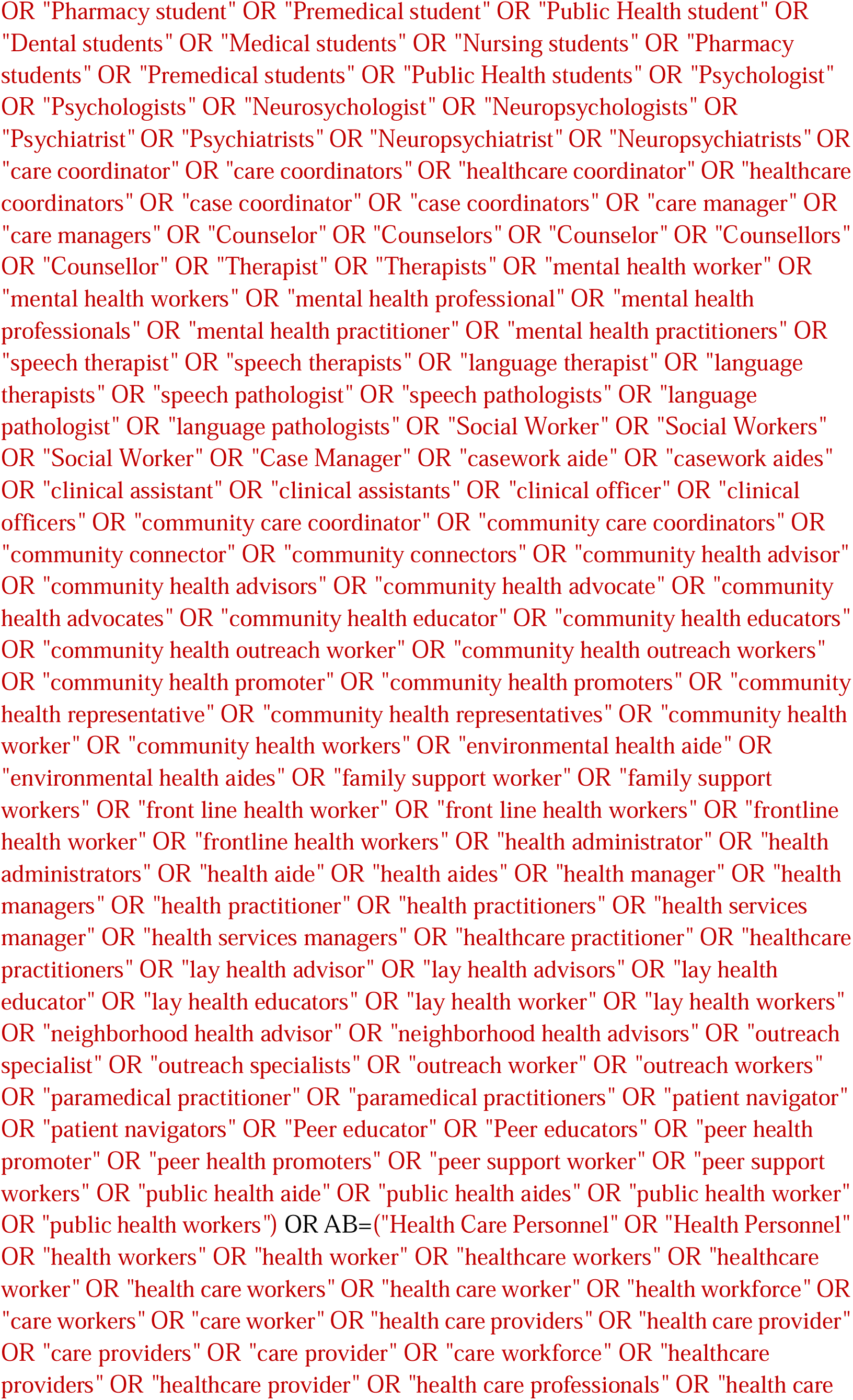
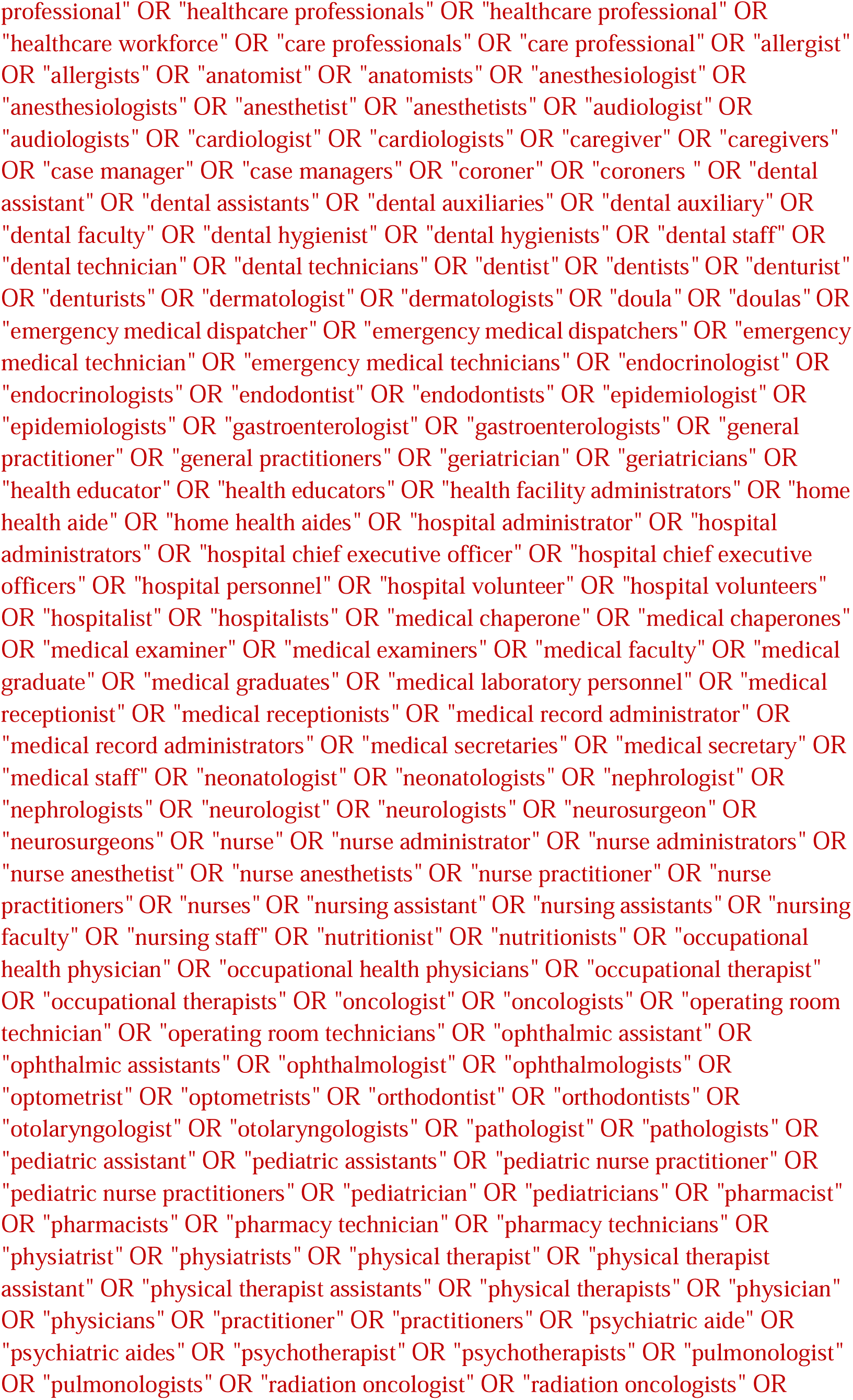
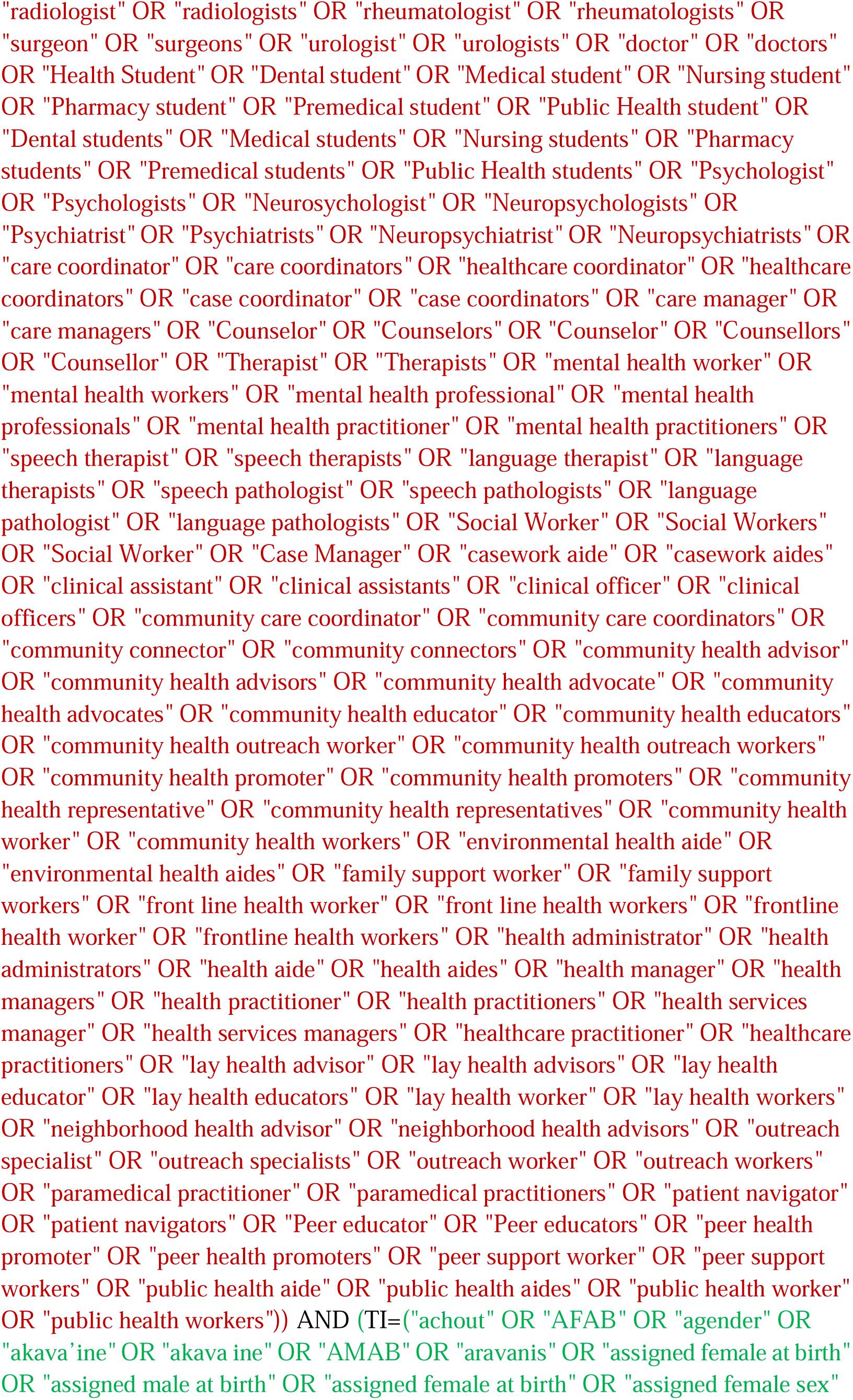
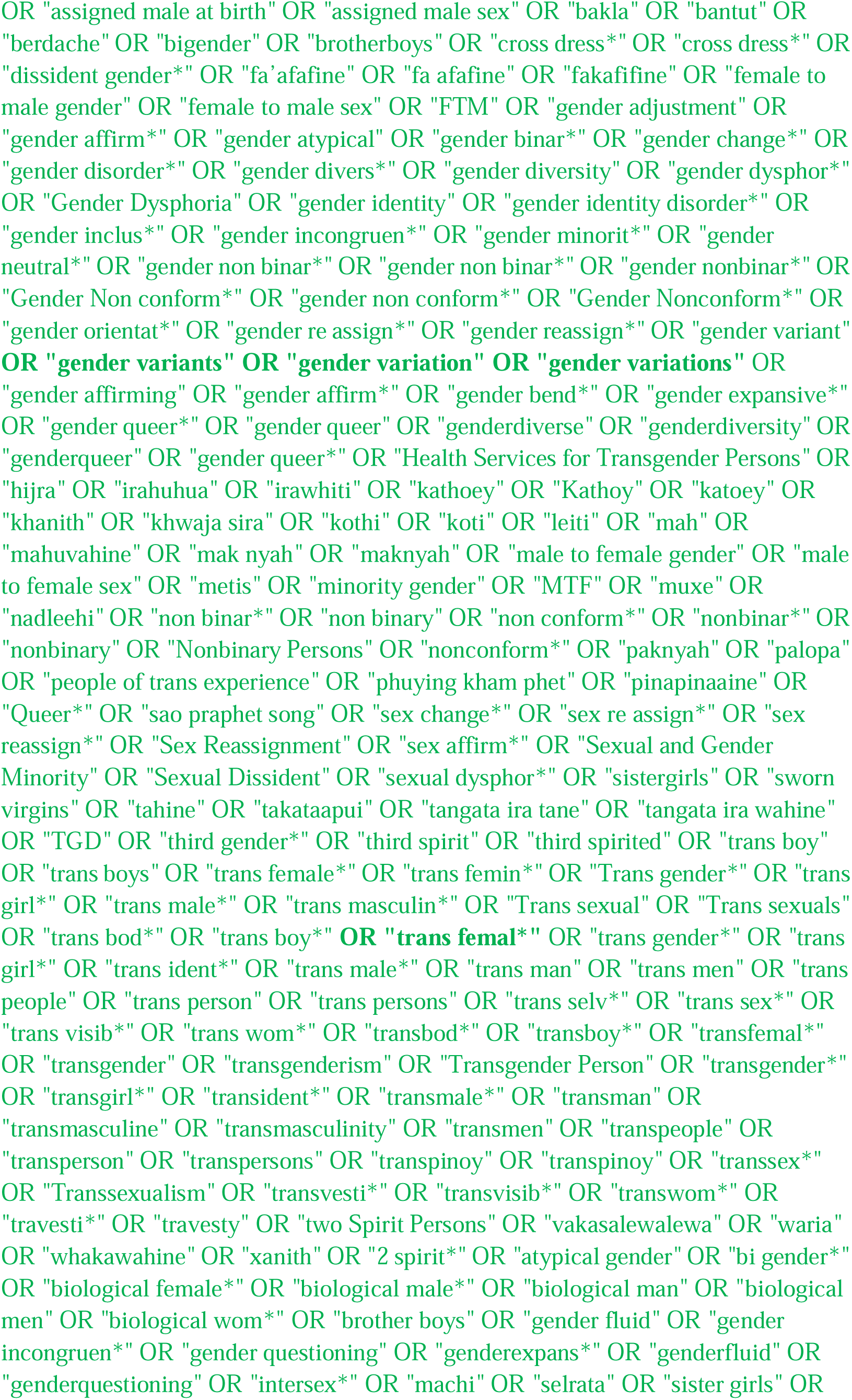
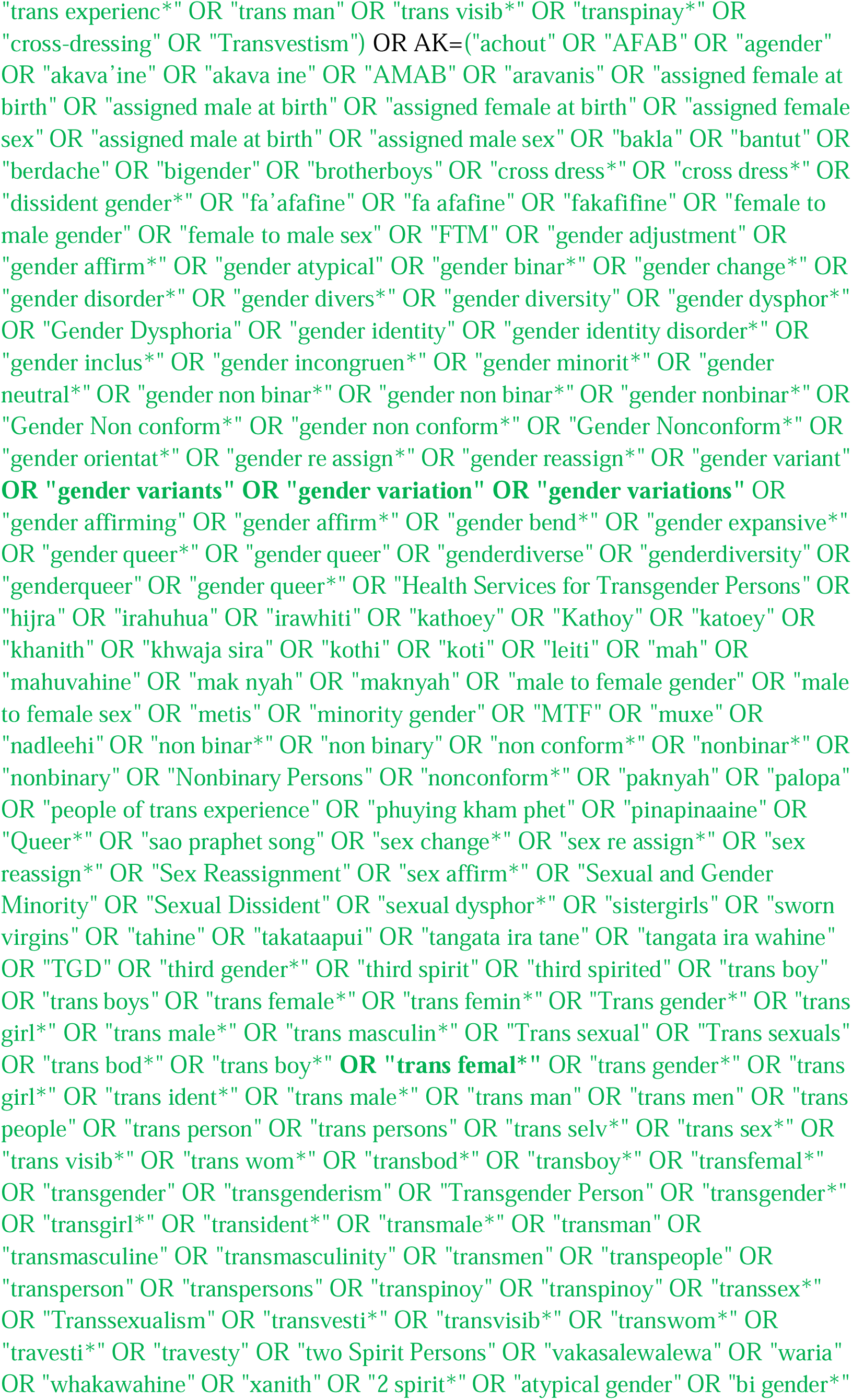
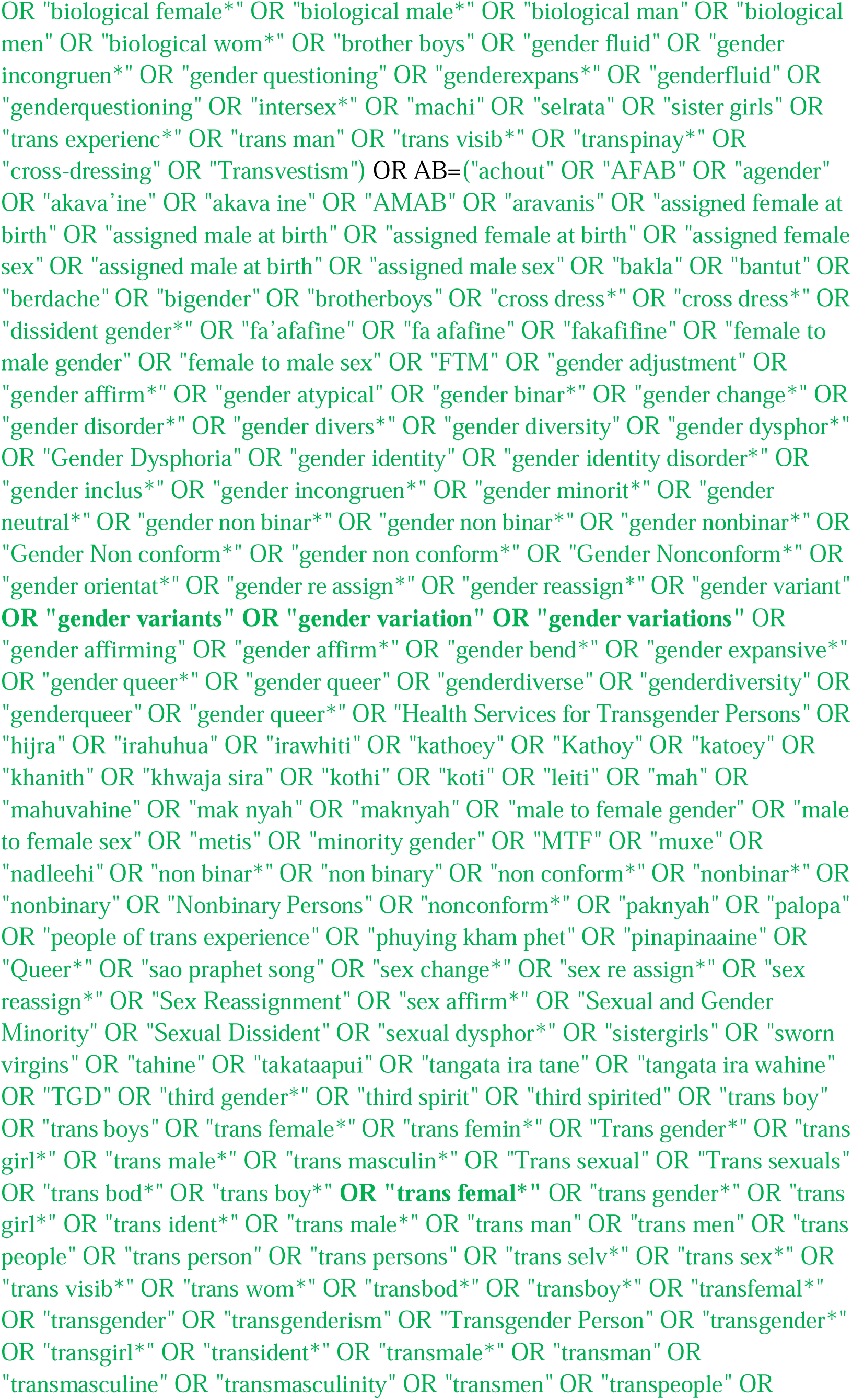
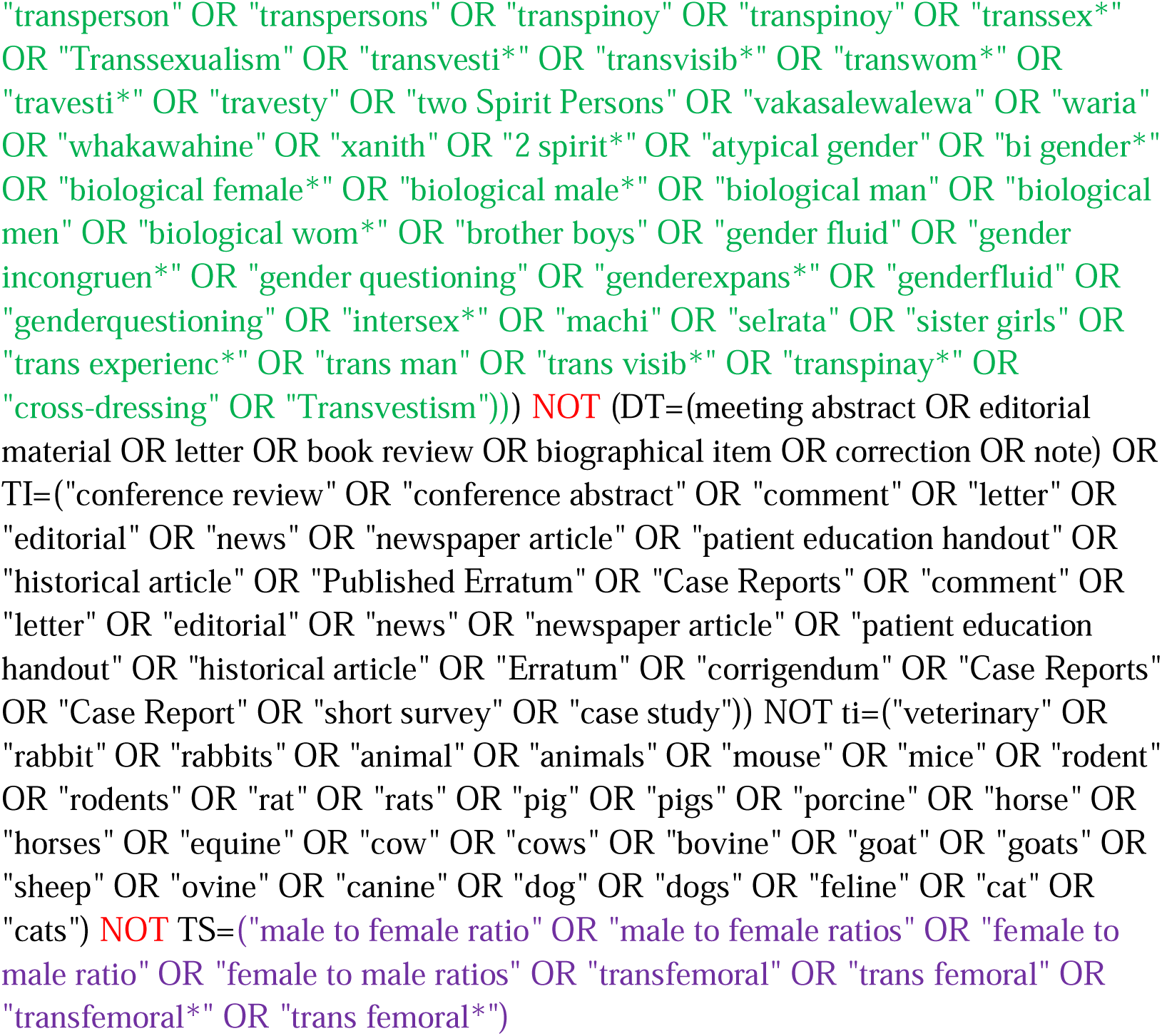

#### Cochrane CENTRAL

https://www.cochranelibrary.com/advanced-search/search-manager

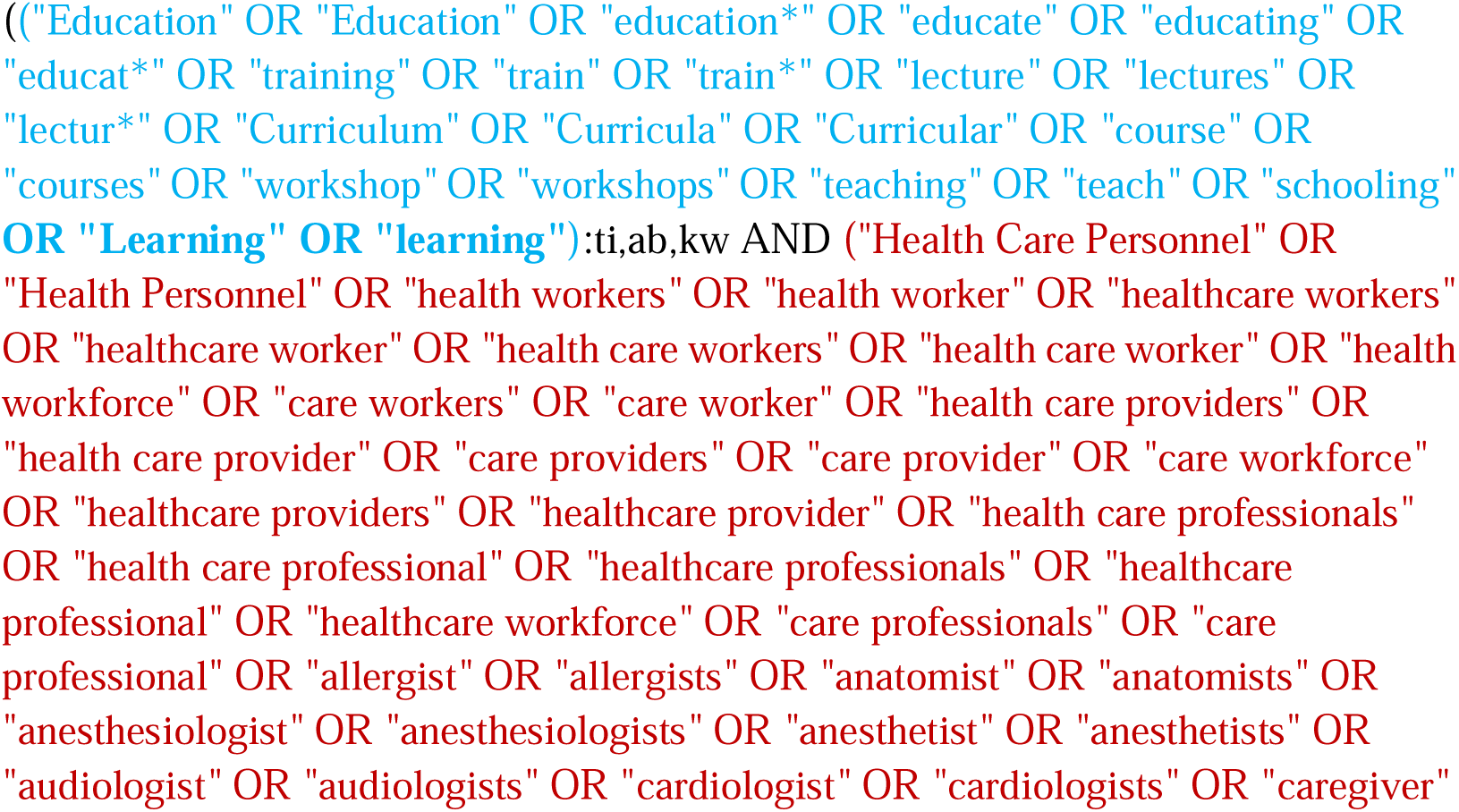
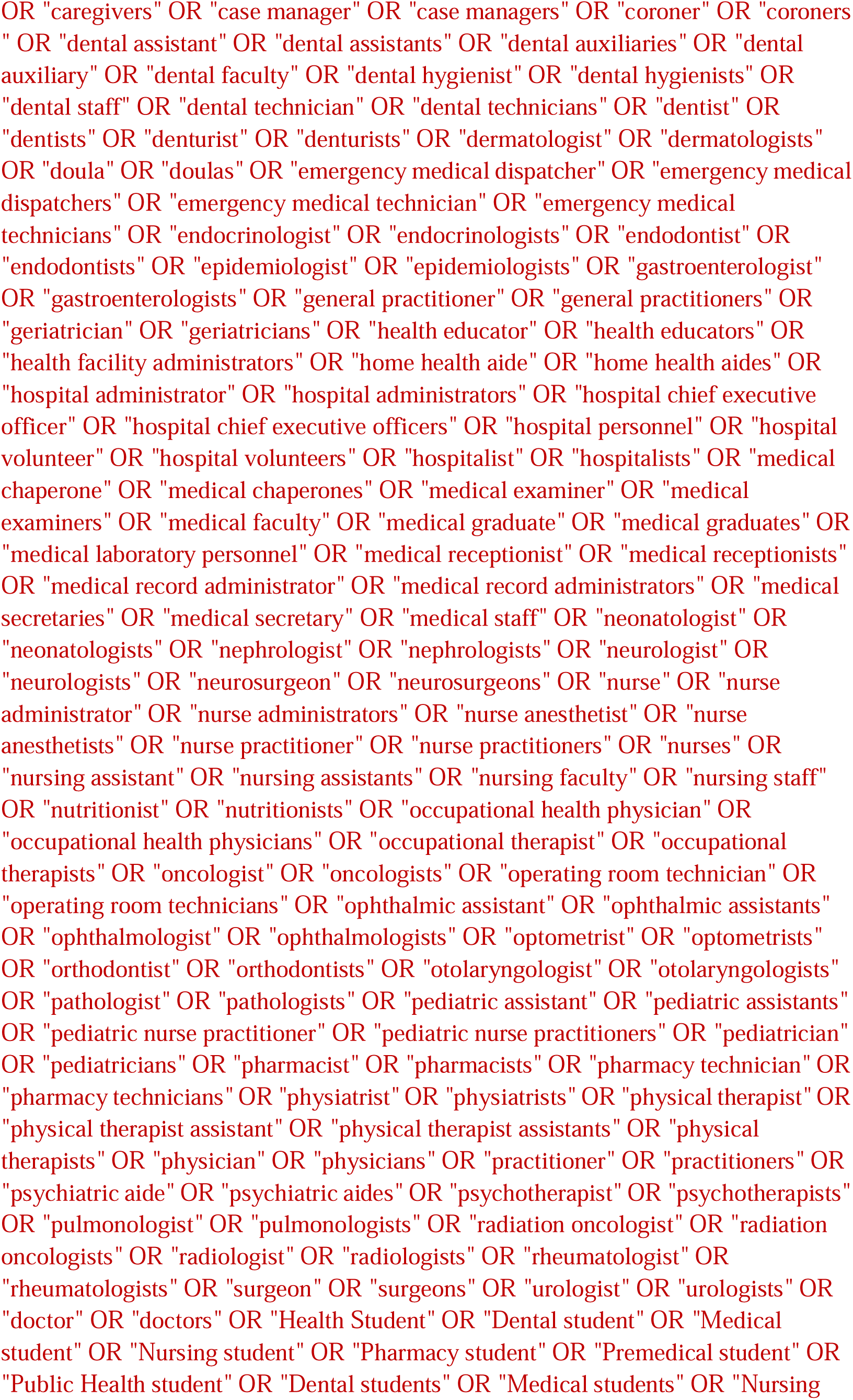
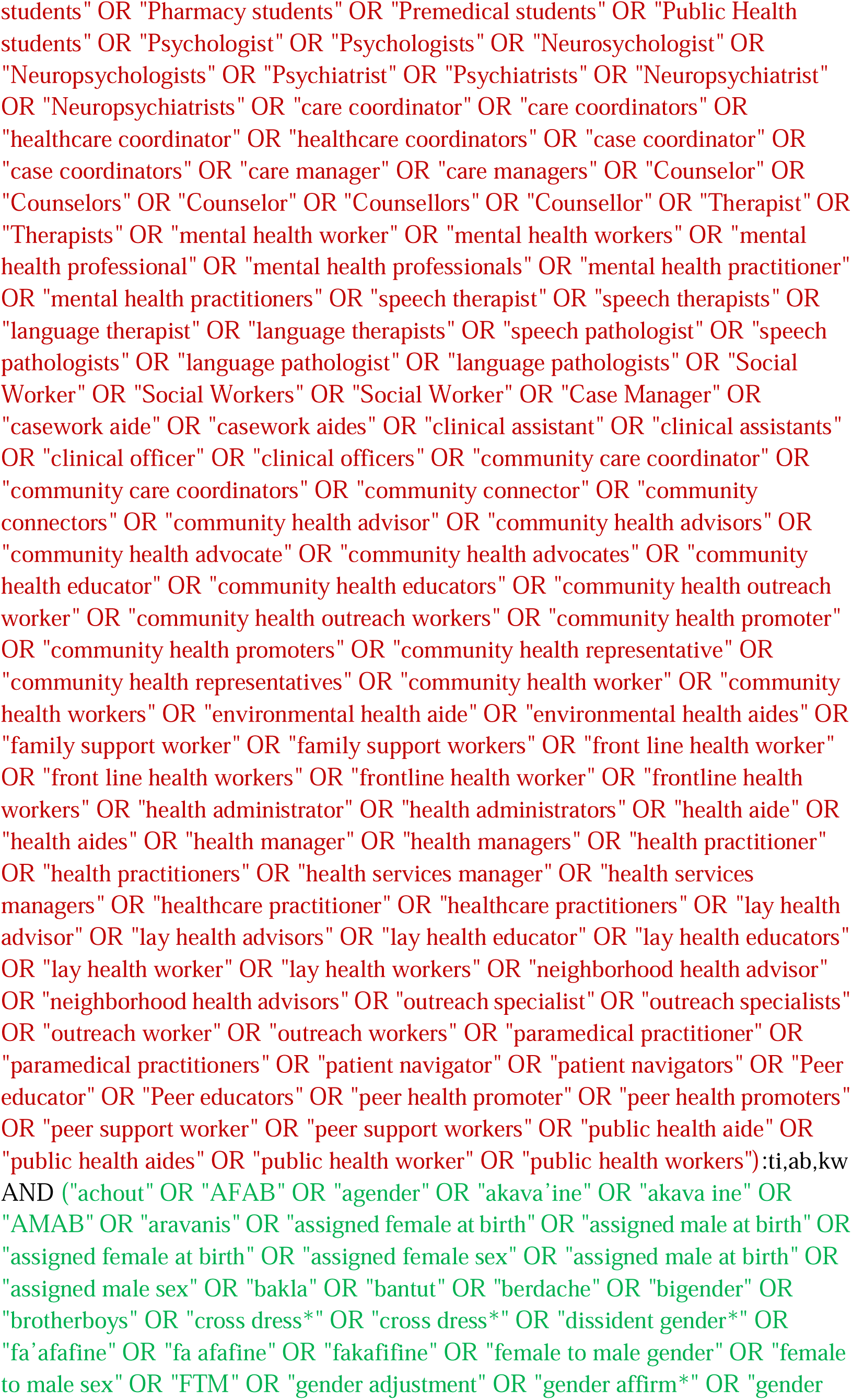
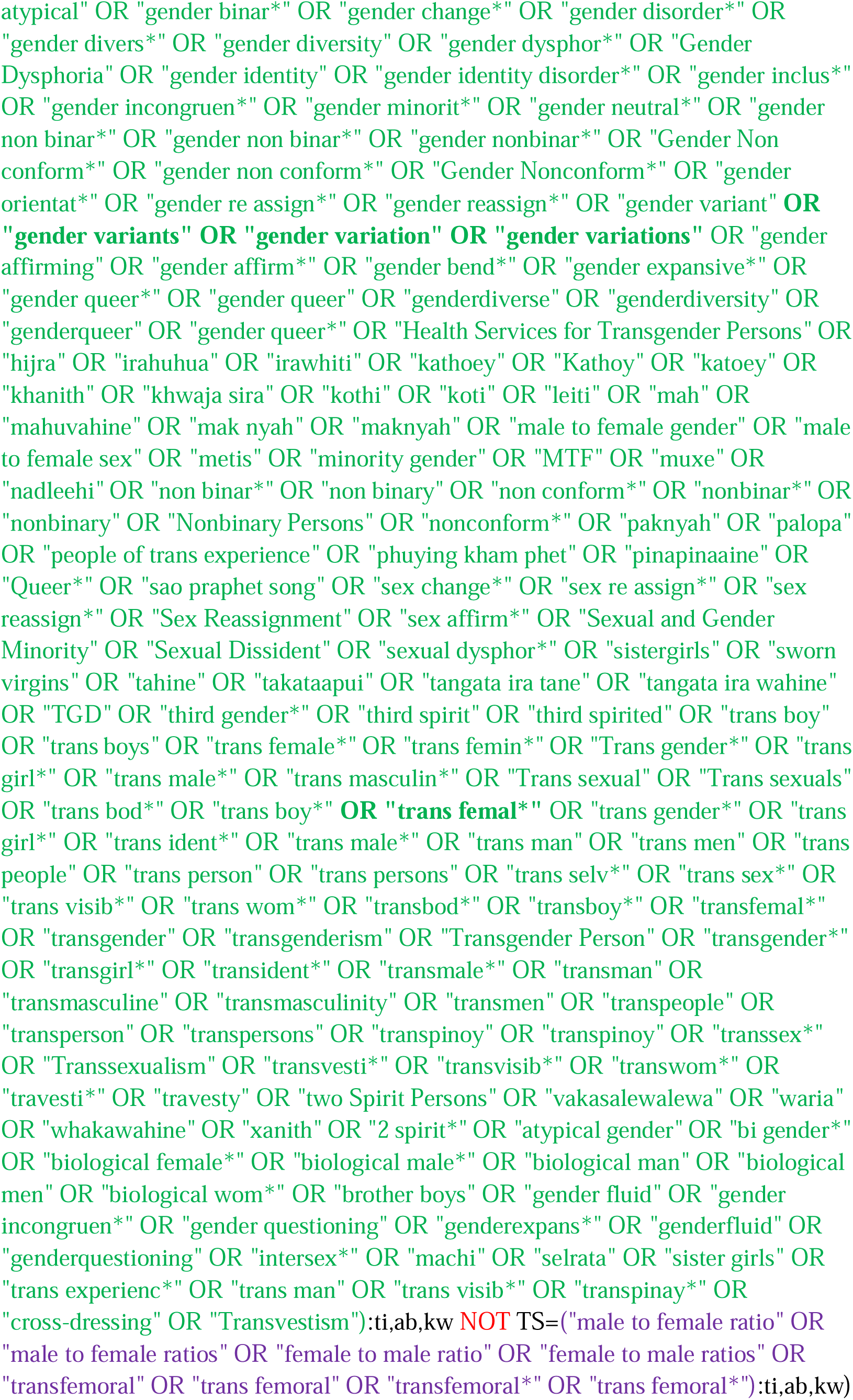

NOT (conference abstract OR meeting abstract OR conference proceeding OR conference proceedings):pt

#### Emcare (OVID)

http://ovidsp.ovid.com/ovidweb.cgi?T=JS&NEWS=n&CSC=Y&PAGE=main&D=emcr

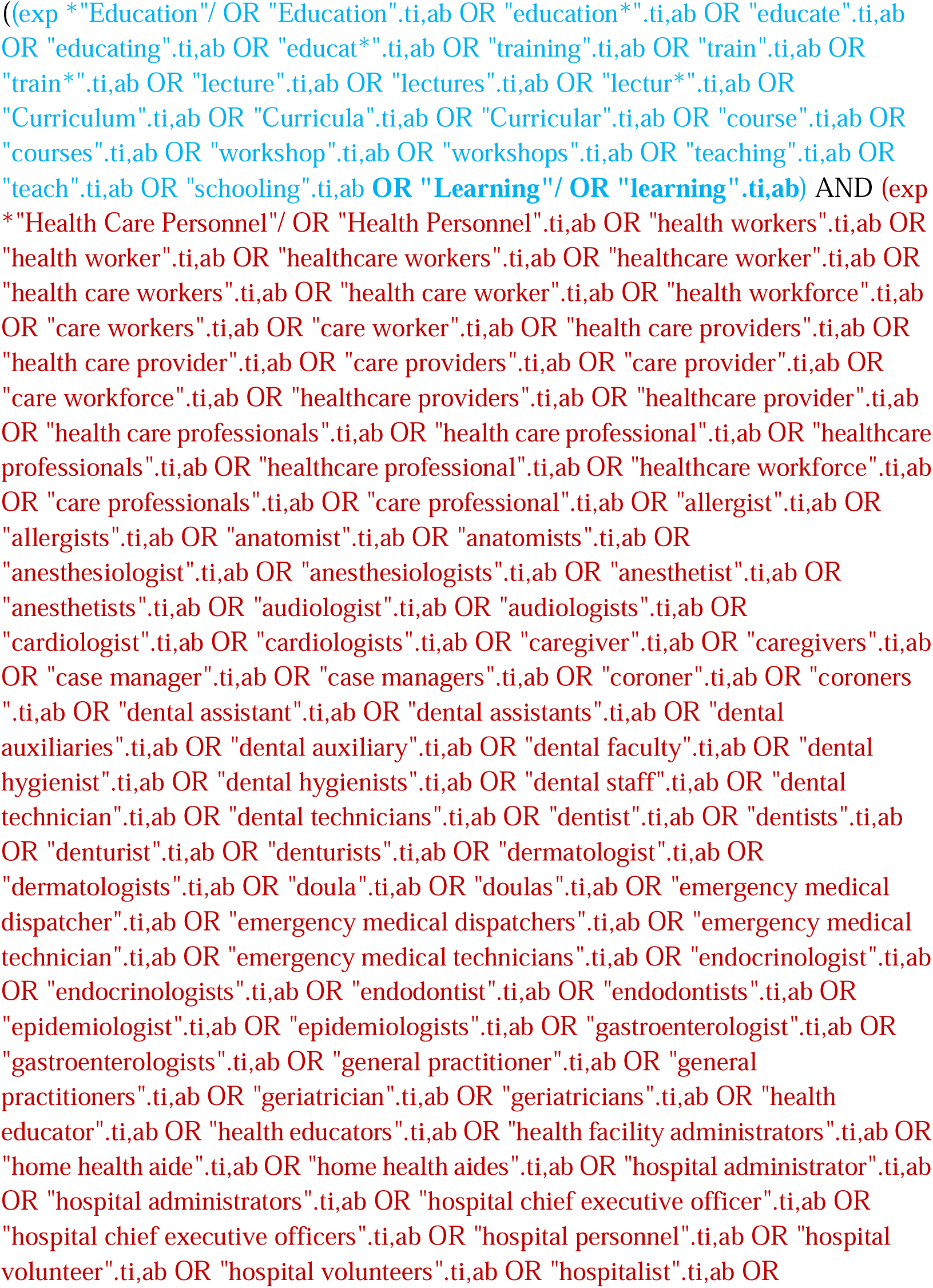
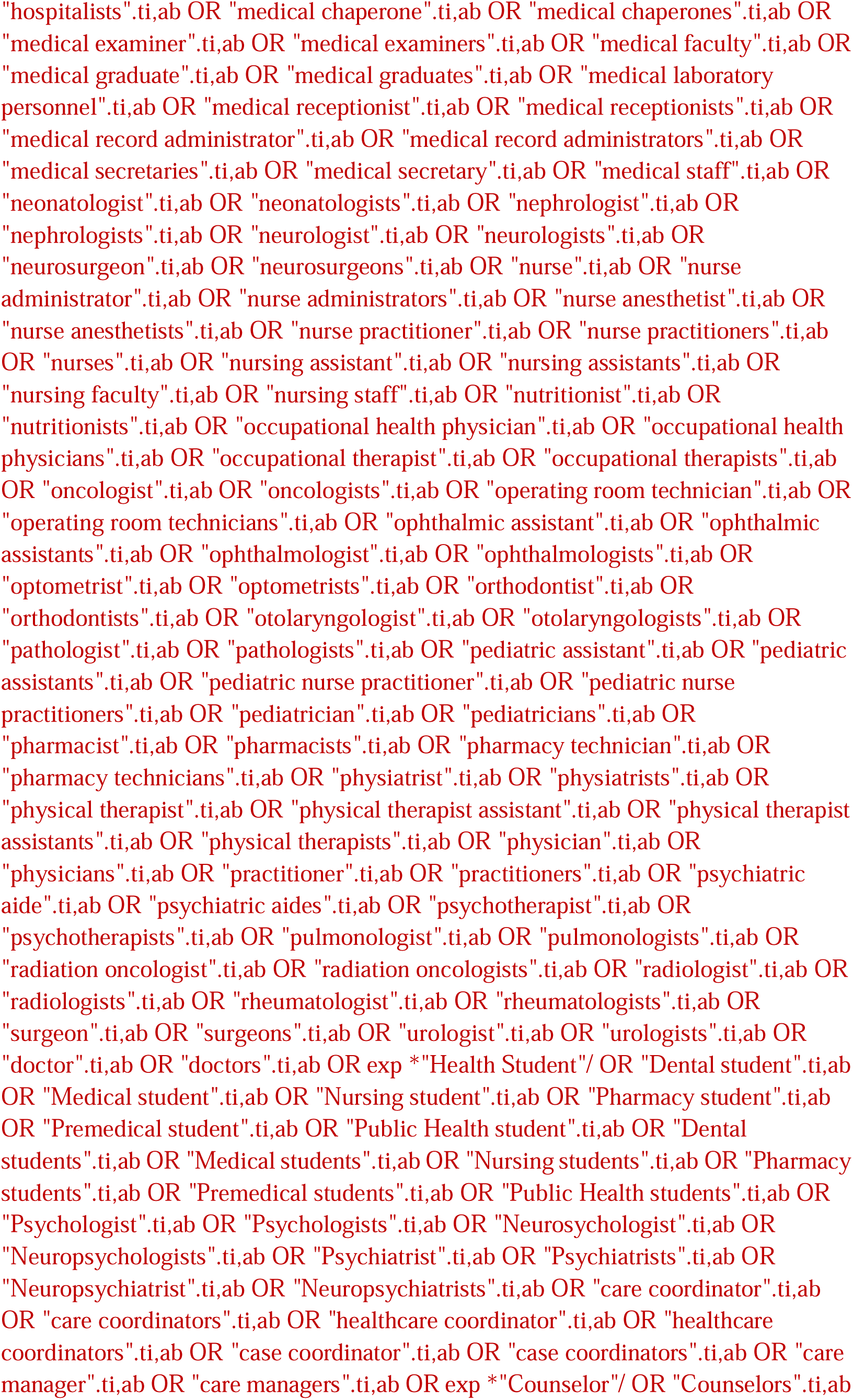
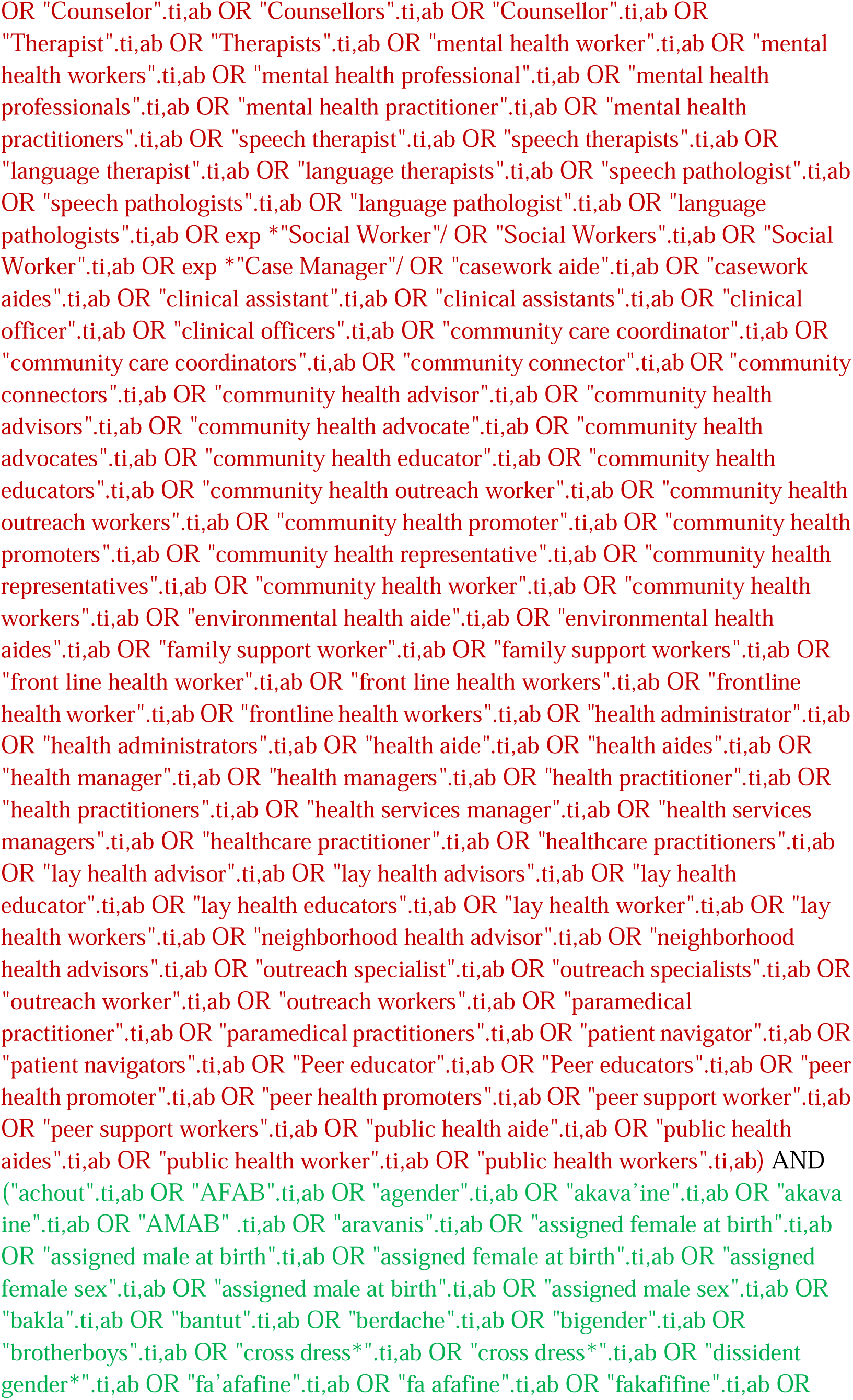
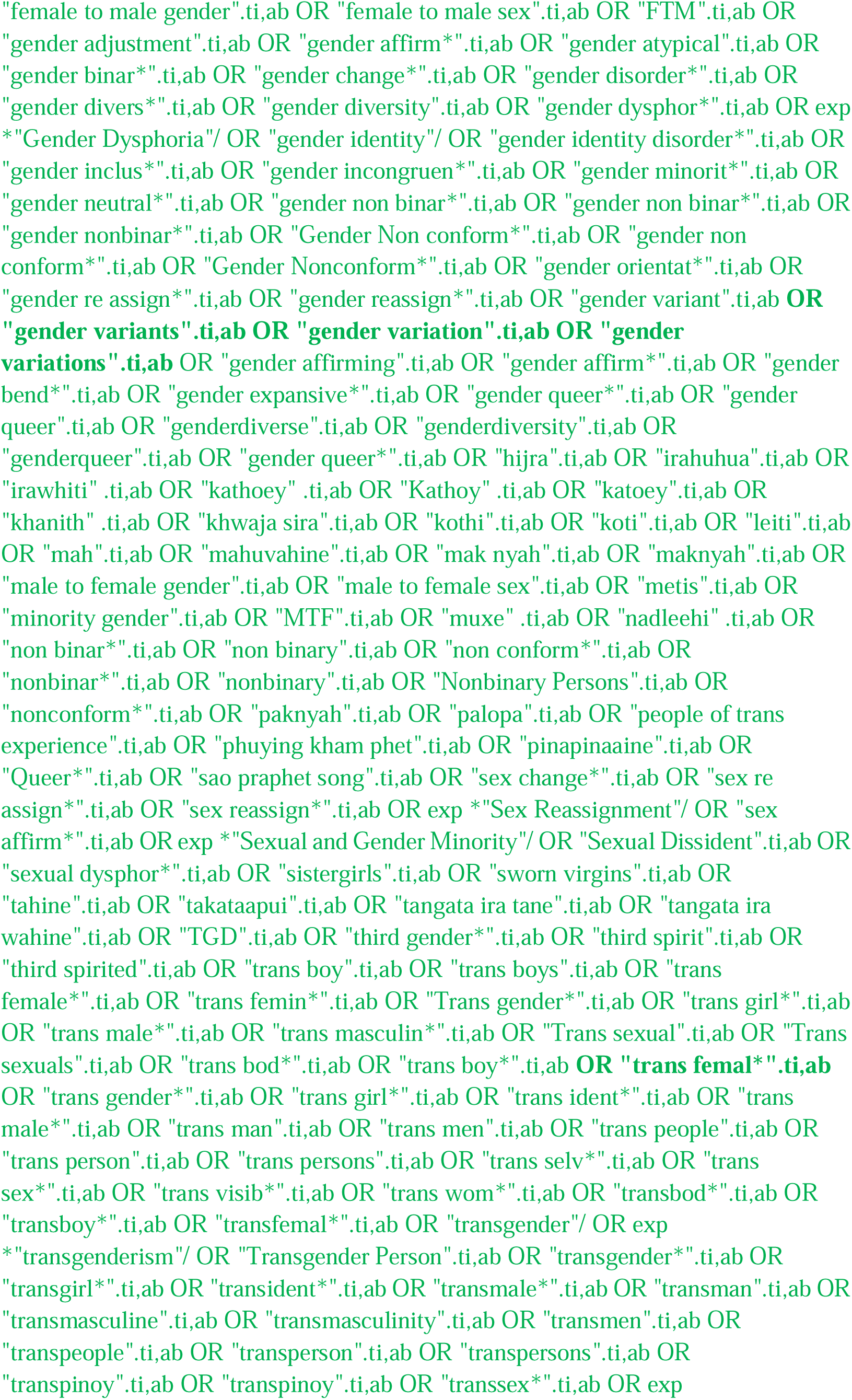
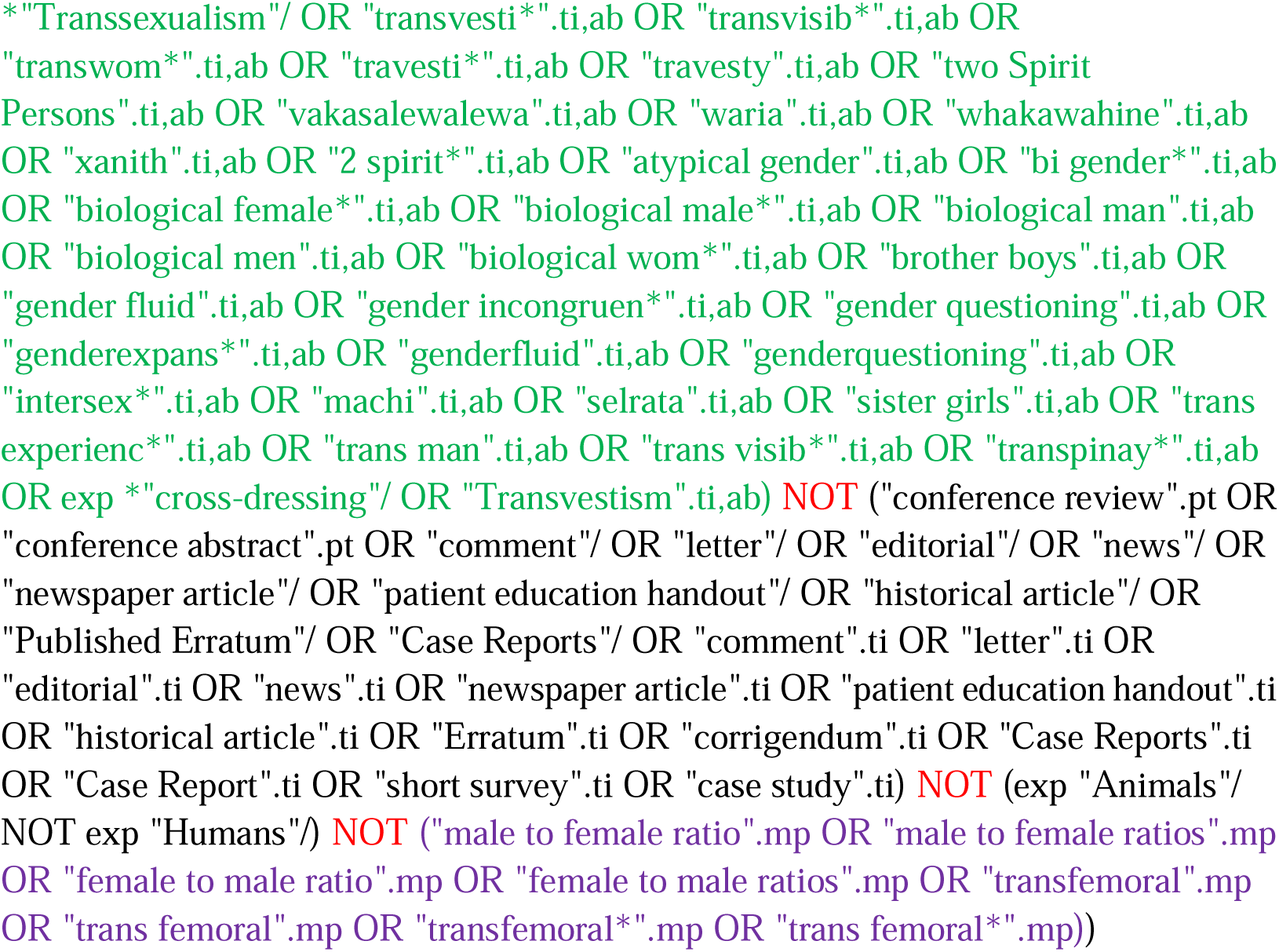

#### PsycINFO (EBSCOhost)

http://search.EBSCOhost.com/login.aspx?authtype=ip,uid&profile=lumc&defaultdb=psyh

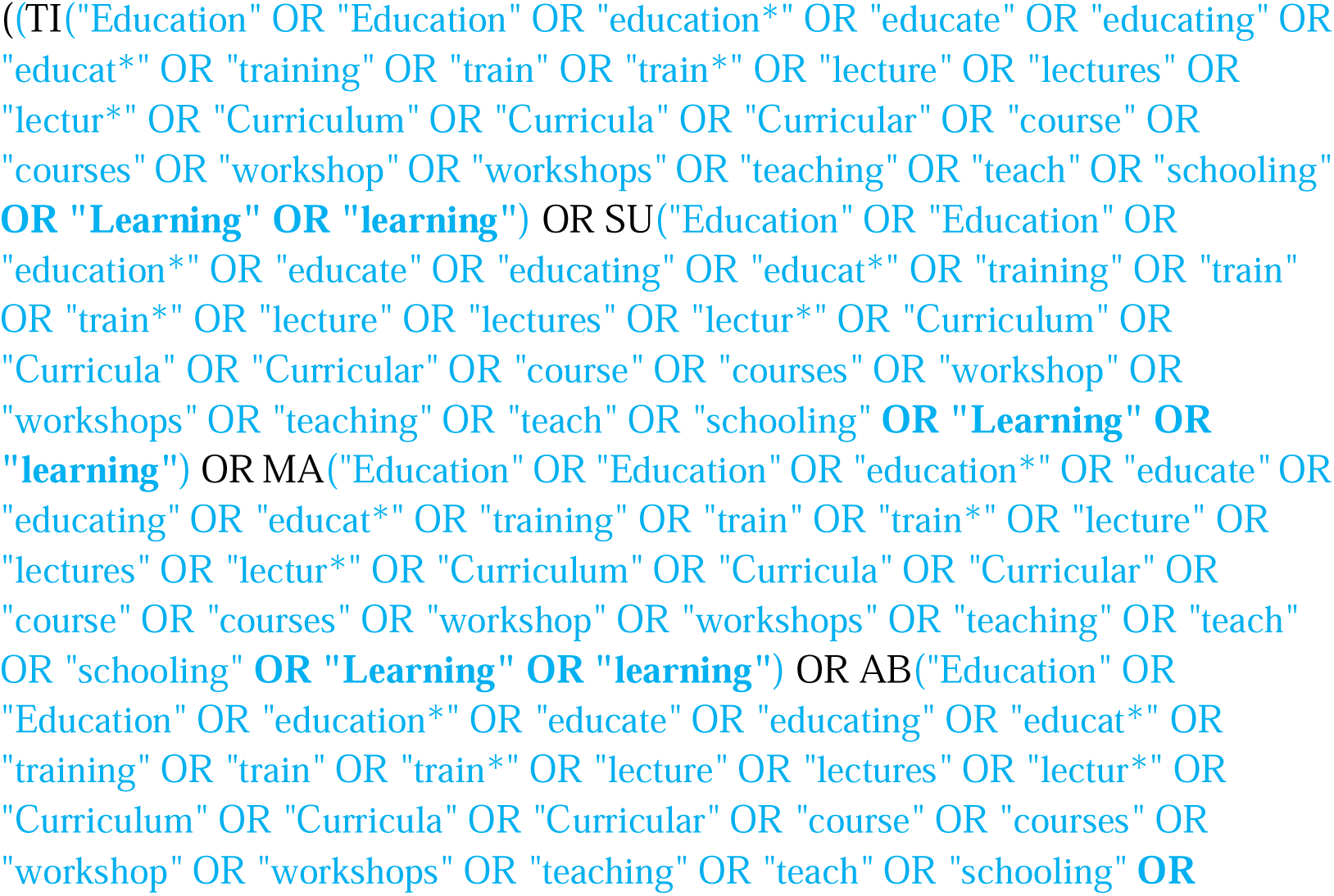
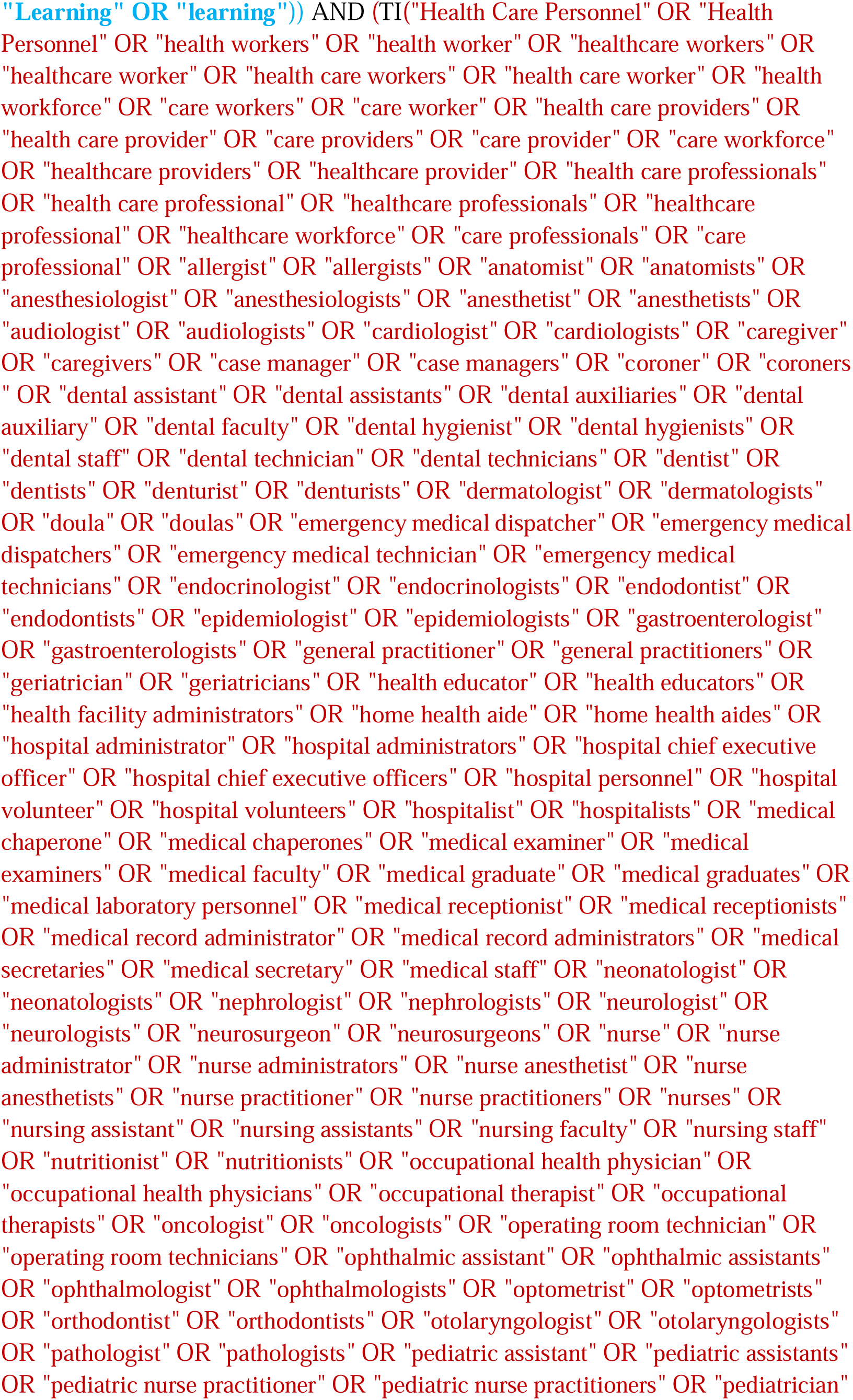
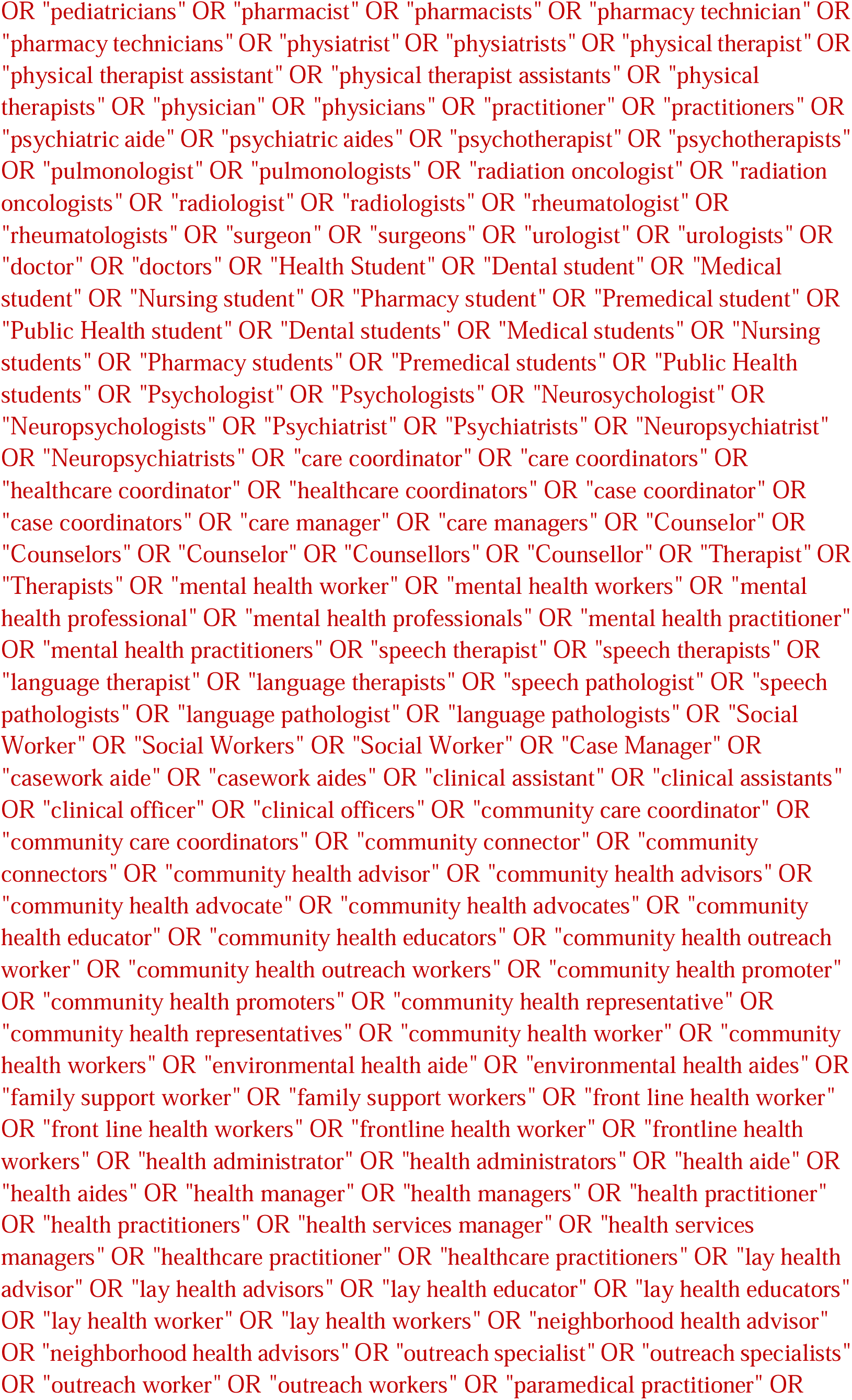
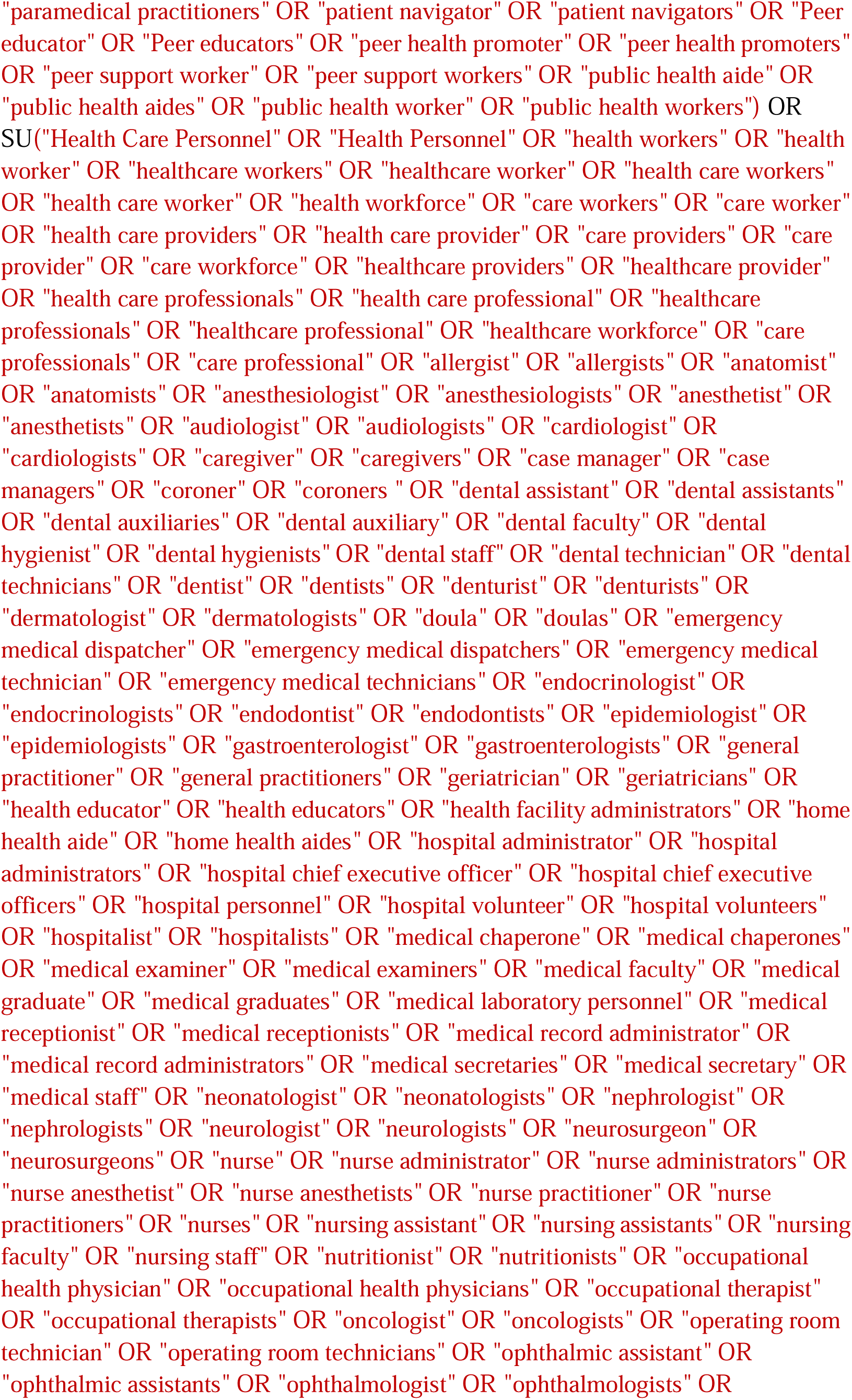
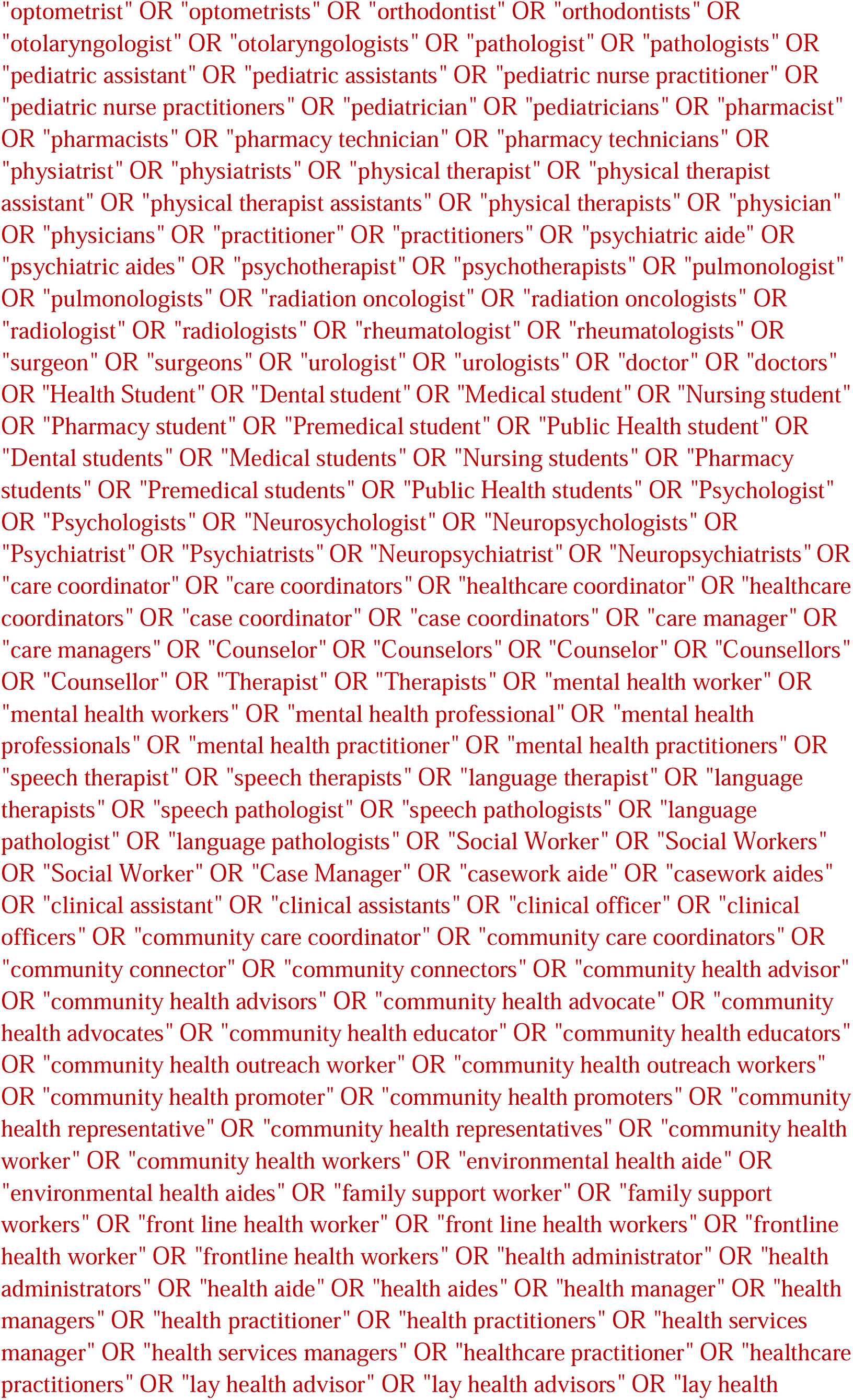
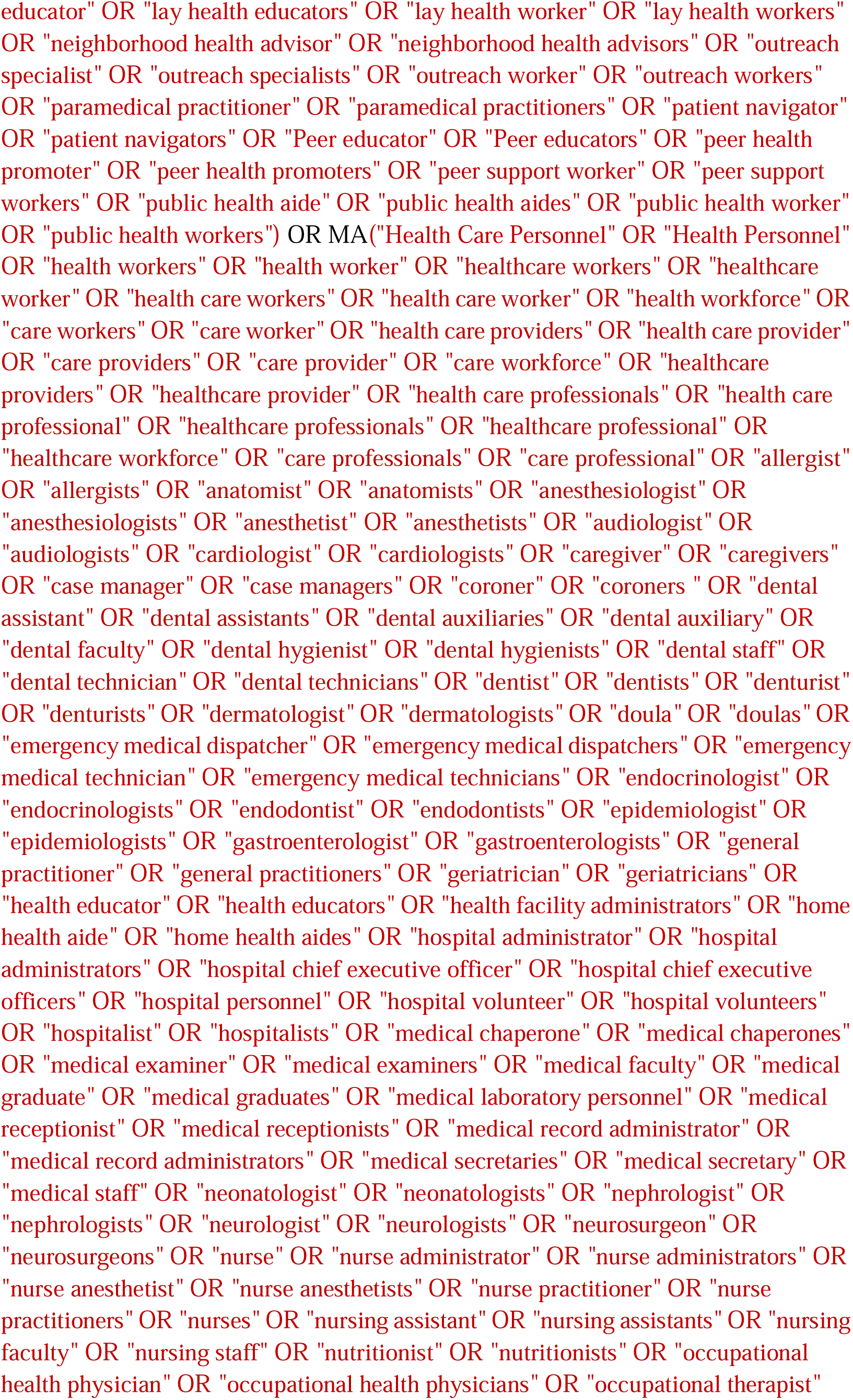
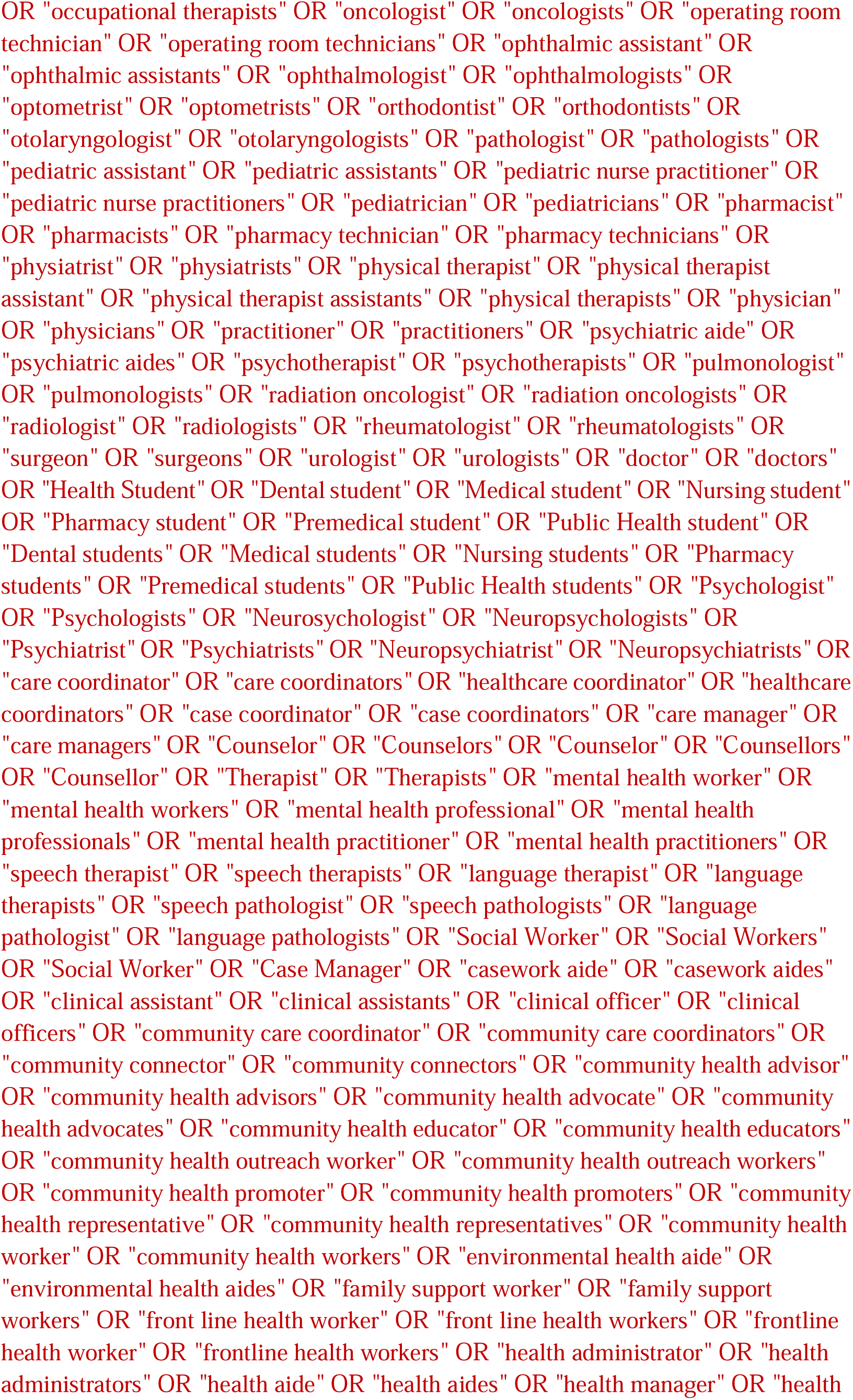
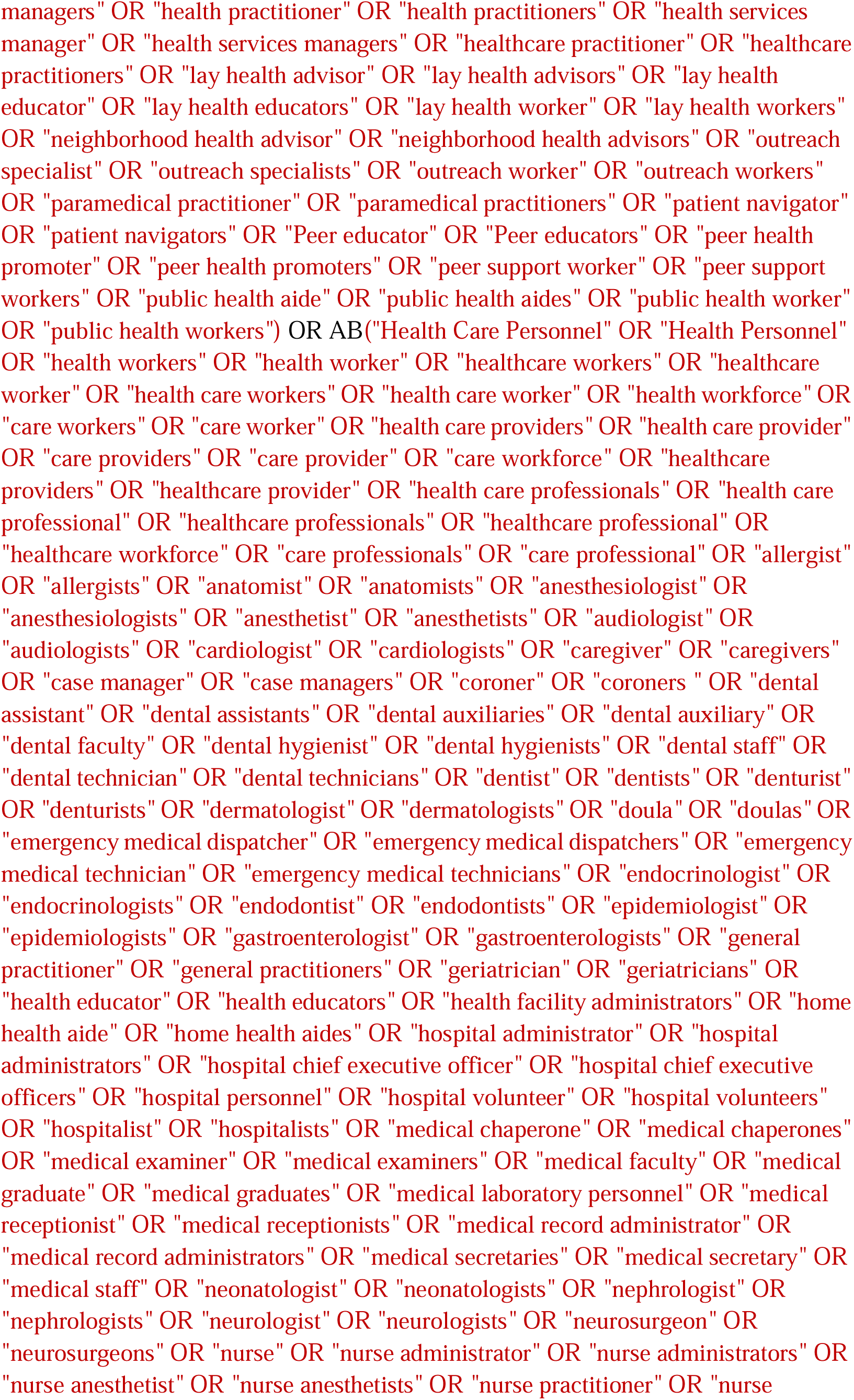
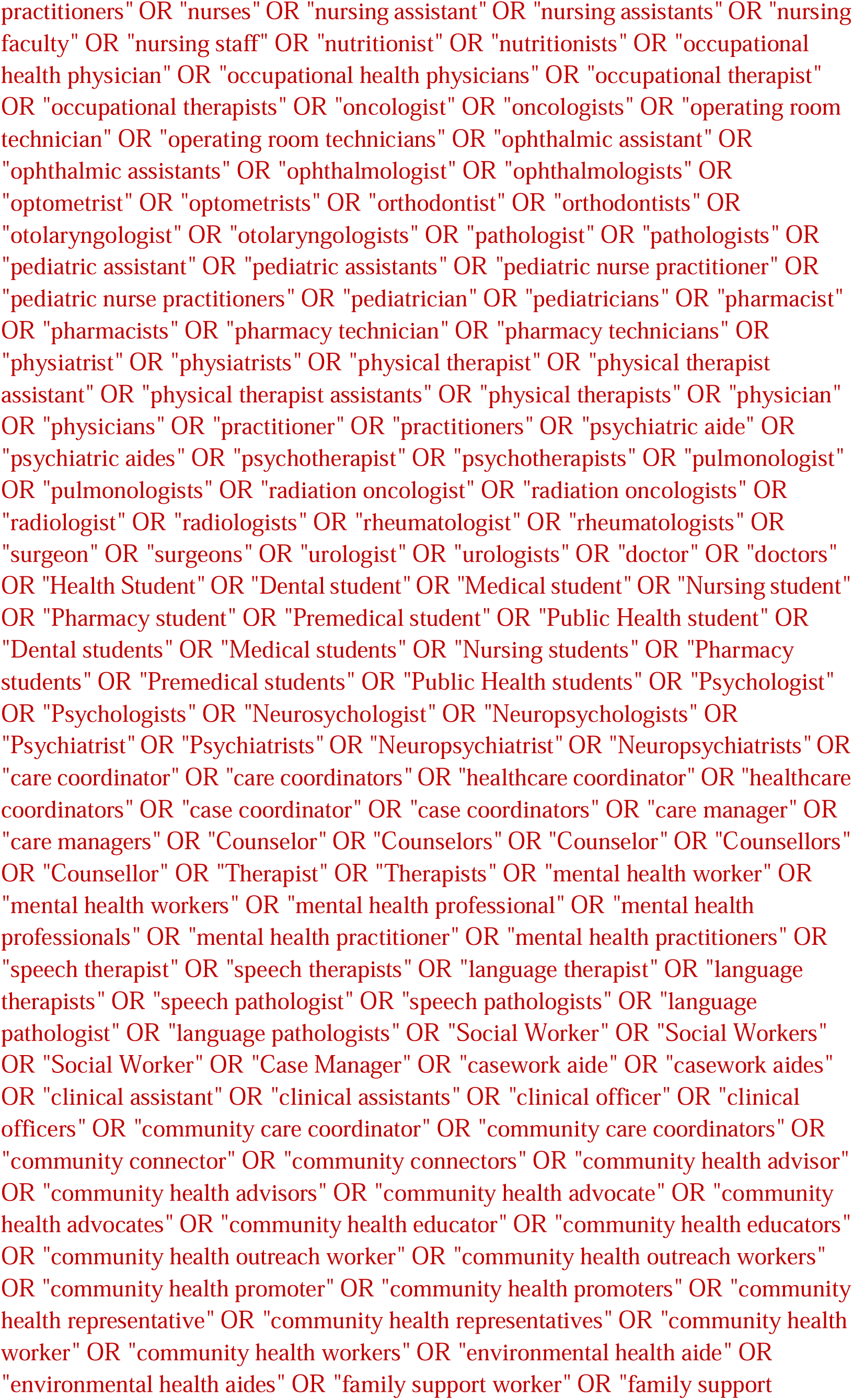
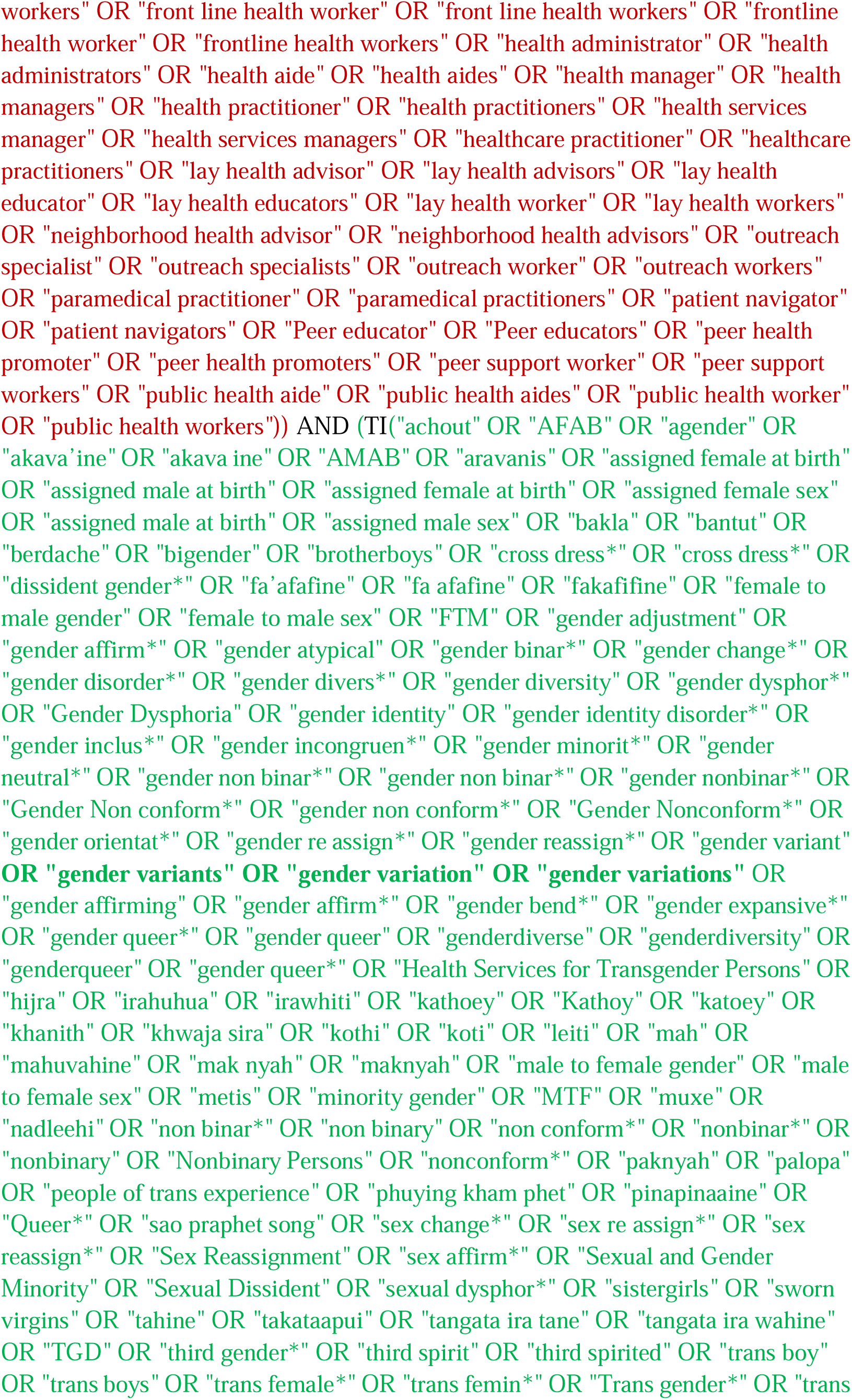
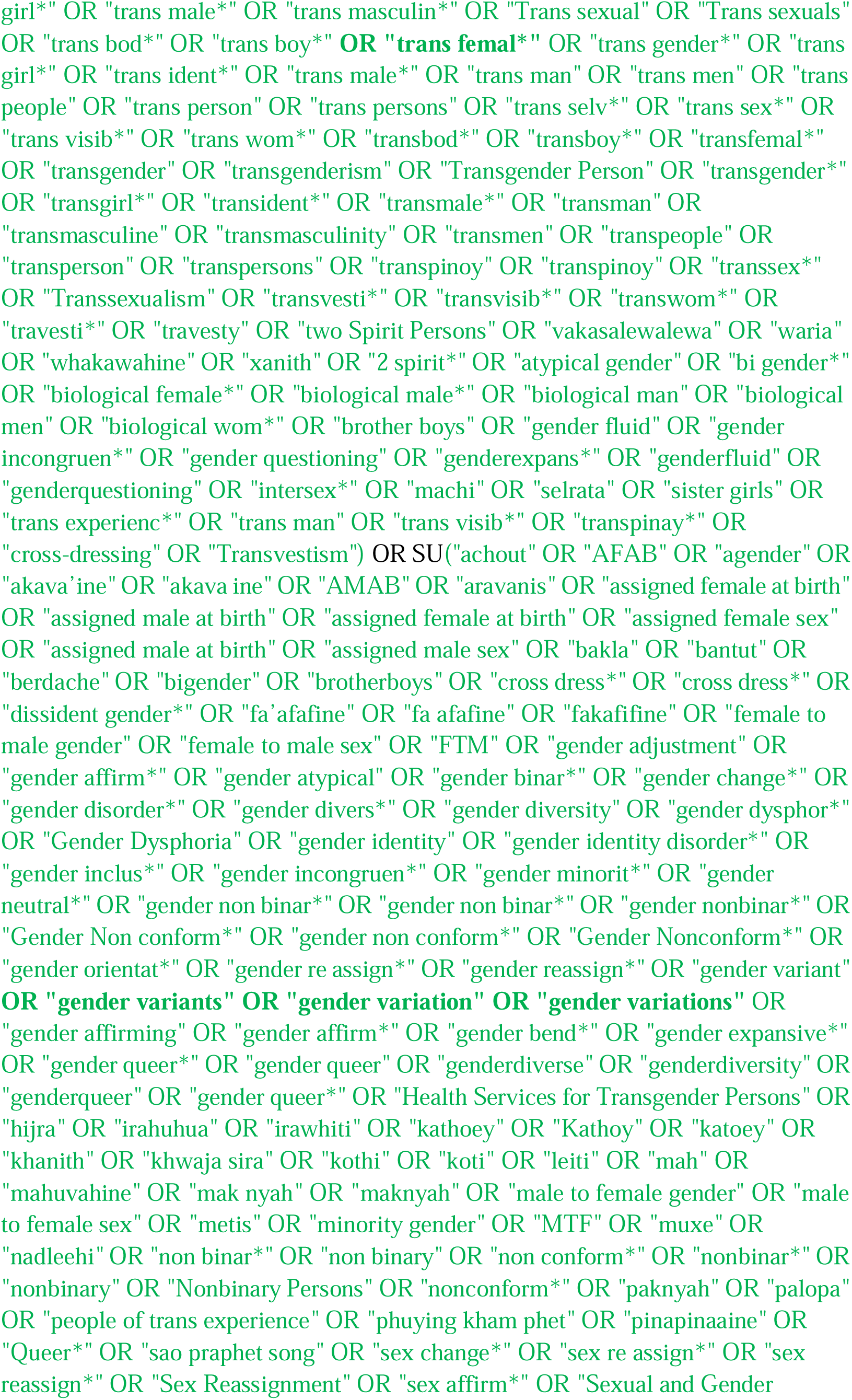
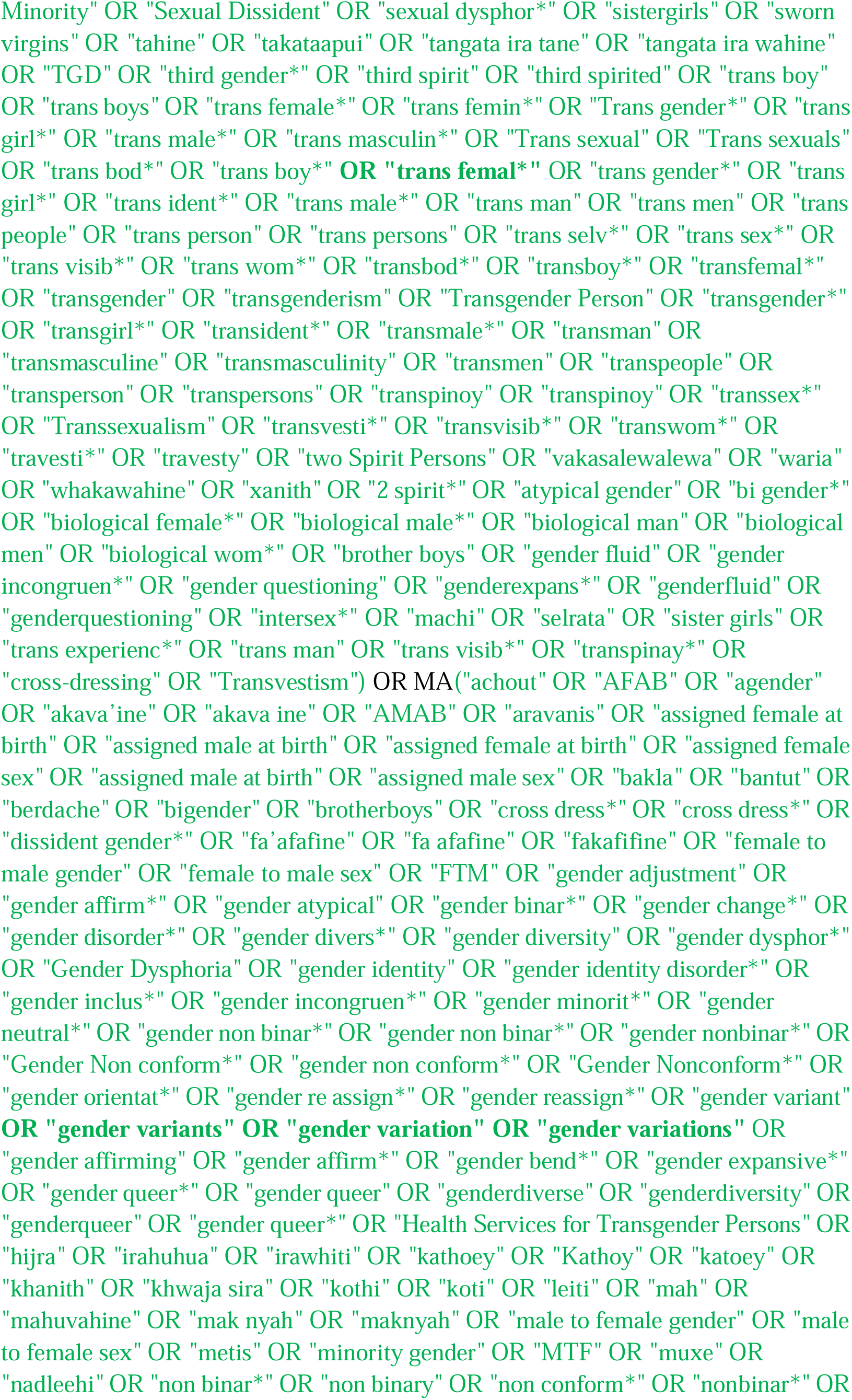
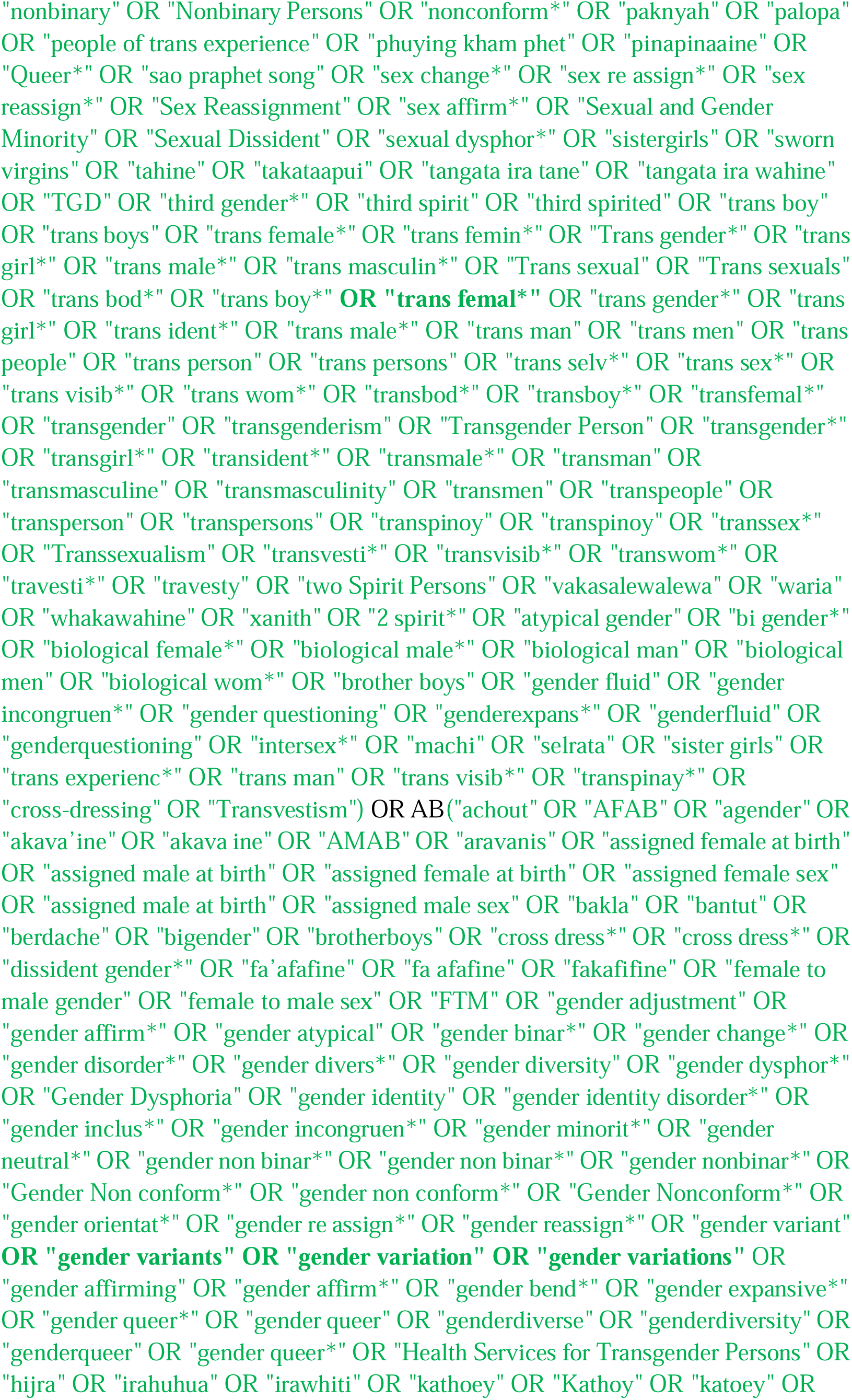
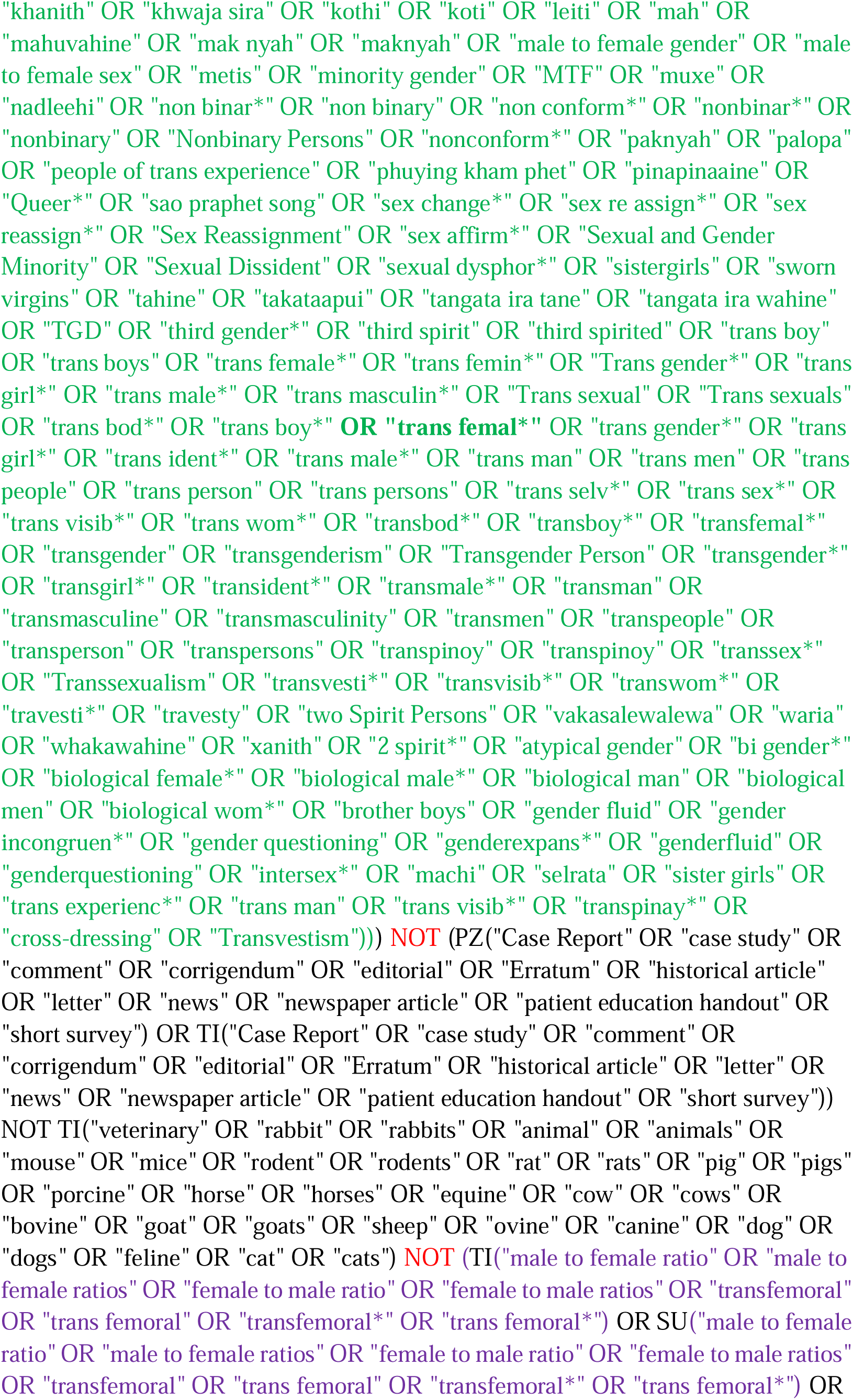
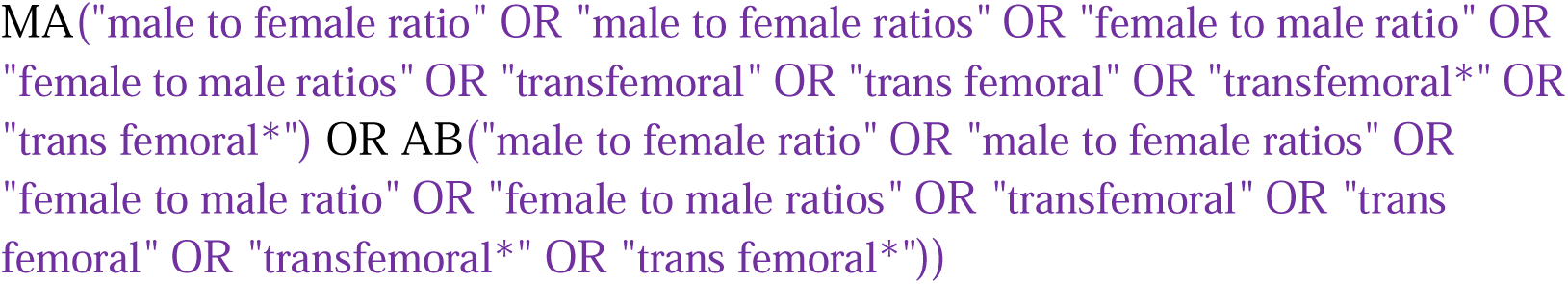

#### Social Services Abstracts

https://search.proquest.com/socialservices?accountid=12045

http://databases.library.leiden.edu/?bibid=990027333030302711&redirect=true

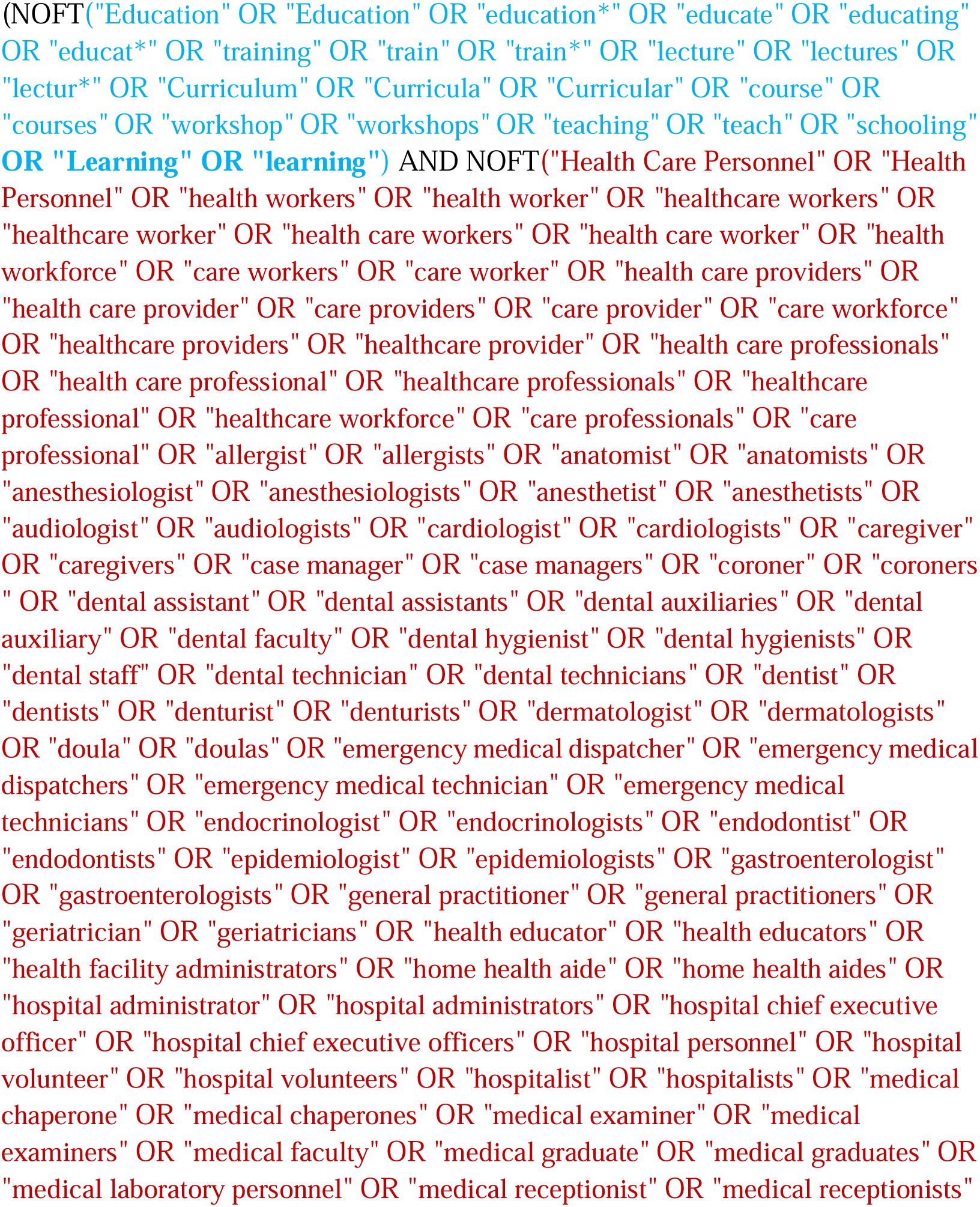
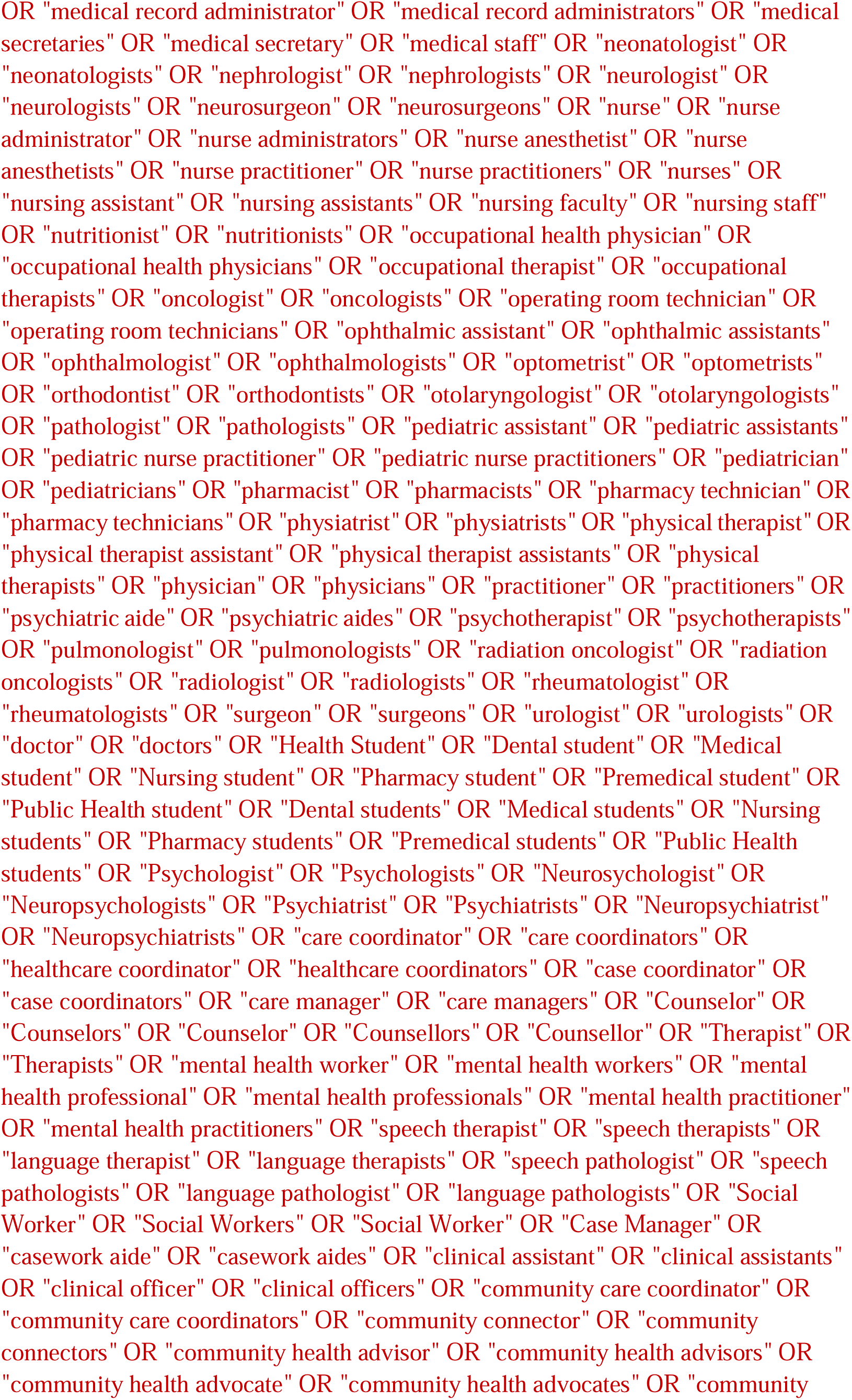
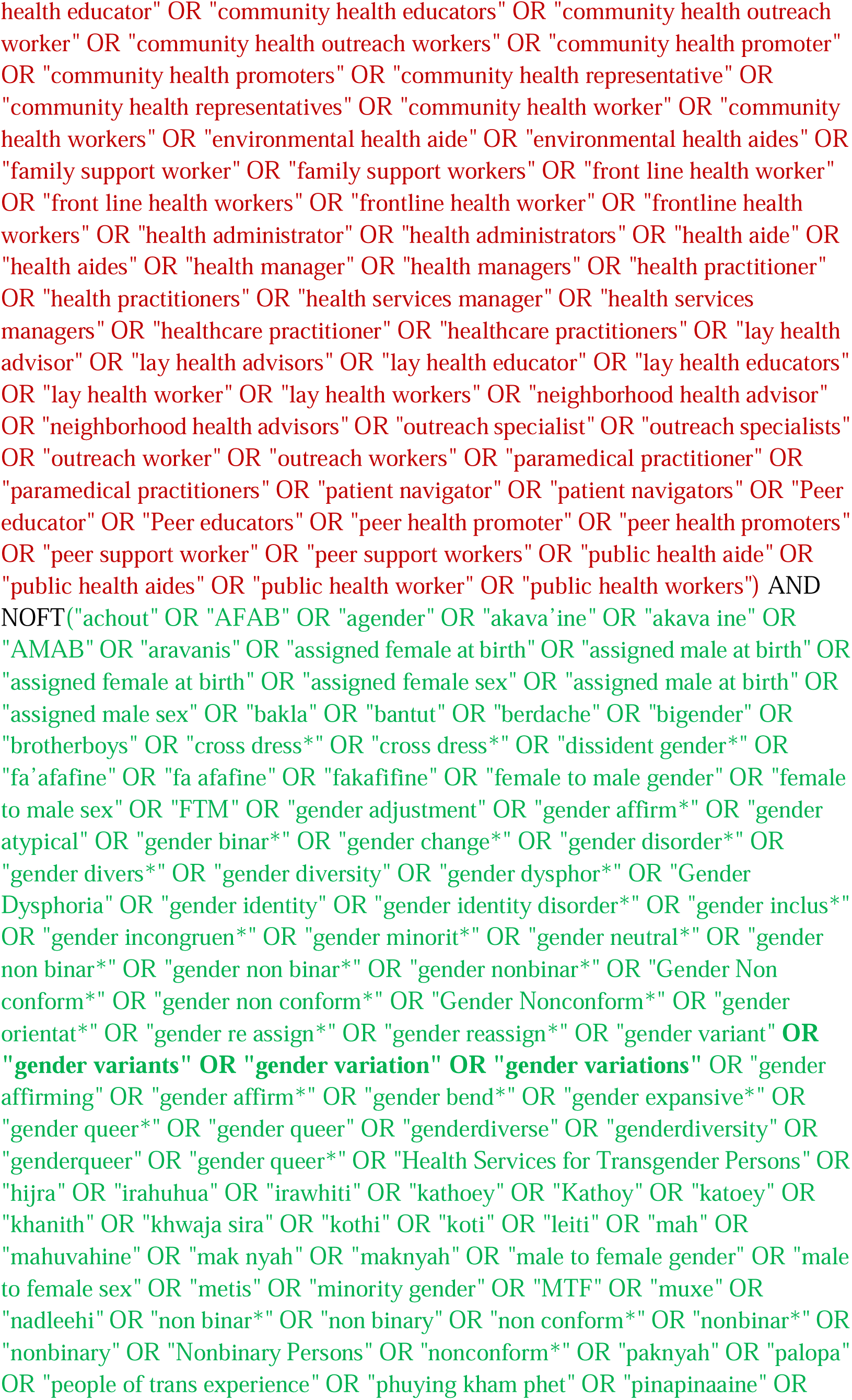
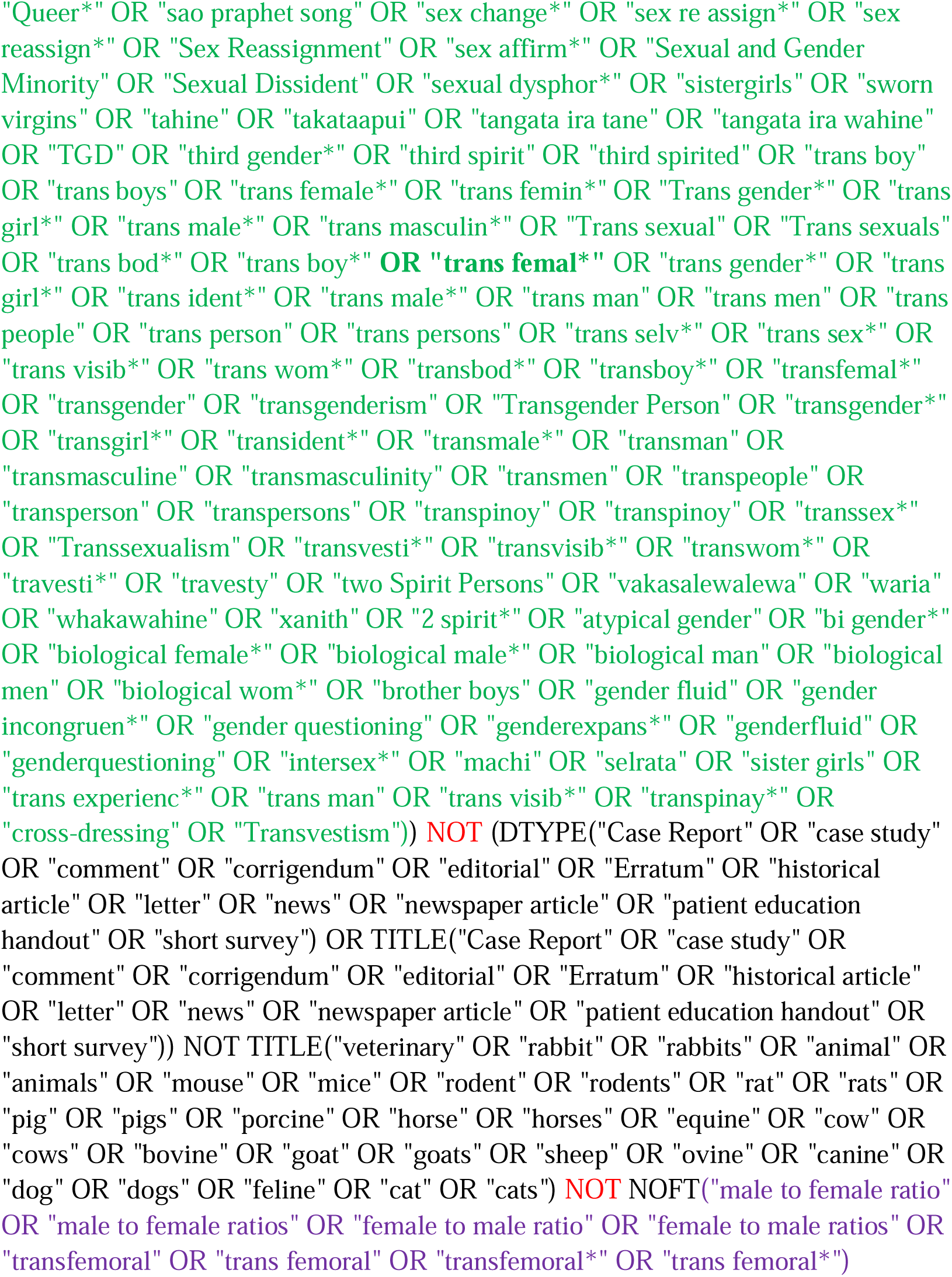

#### Sociological Abstracts

https://search.proquest.com/socabs/index

http://databases.library.leiden.edu/?bibid=990024621960302711&redirect=true

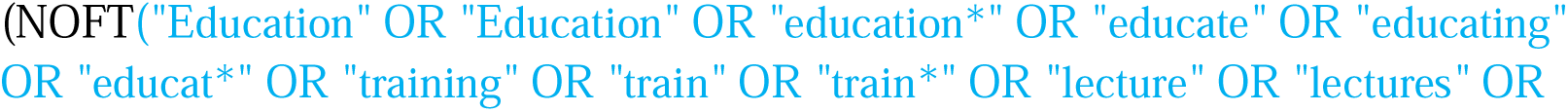
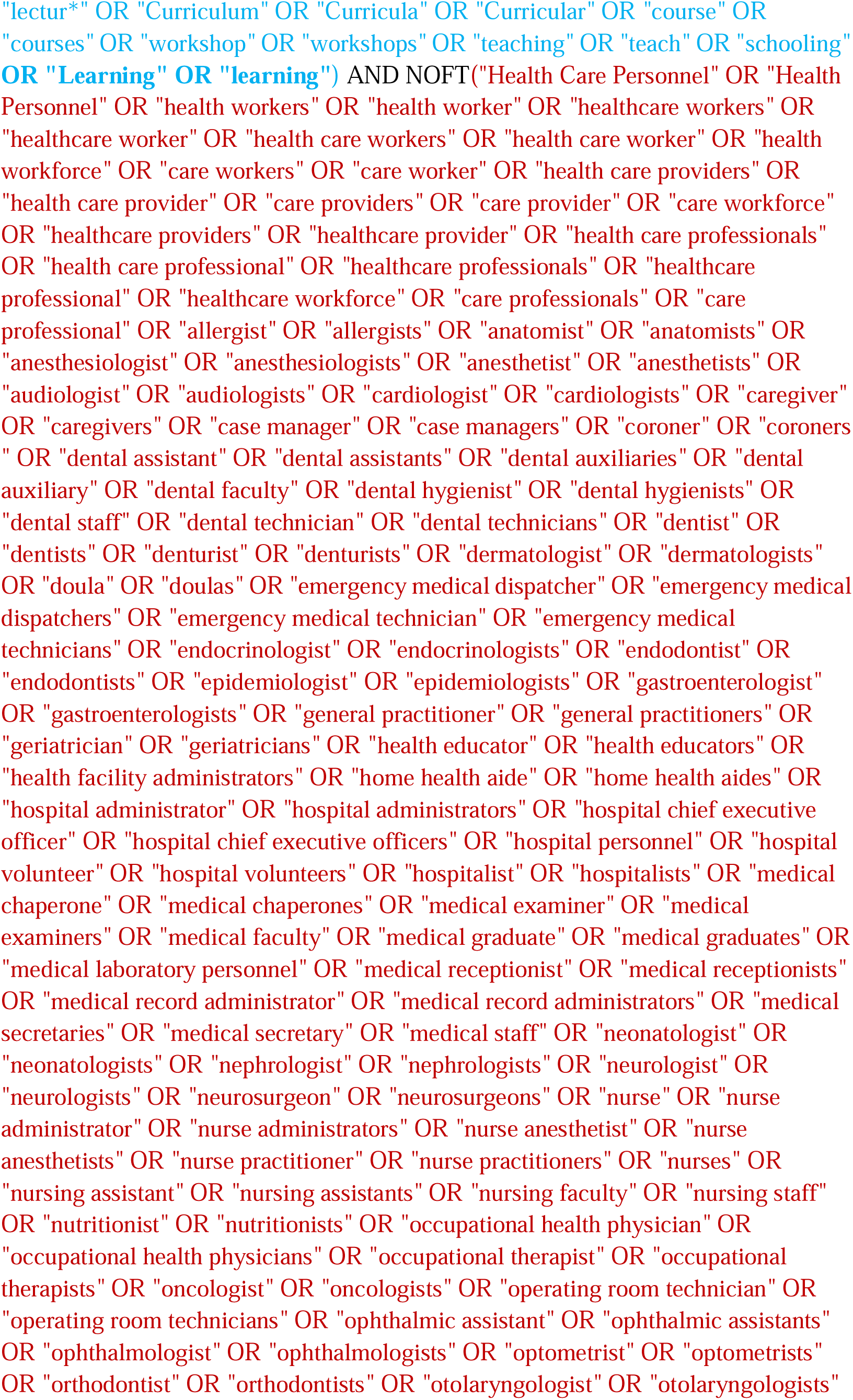
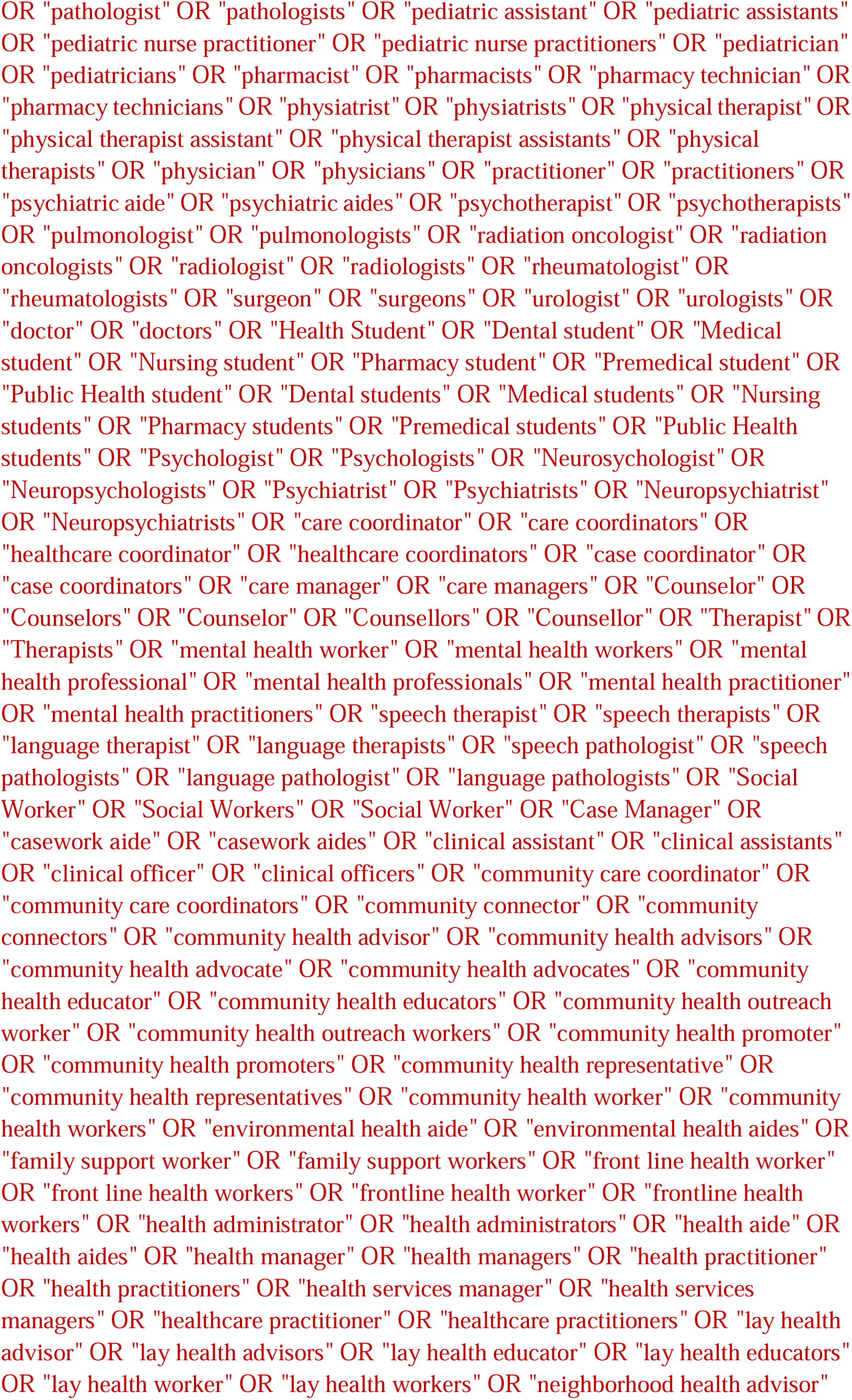
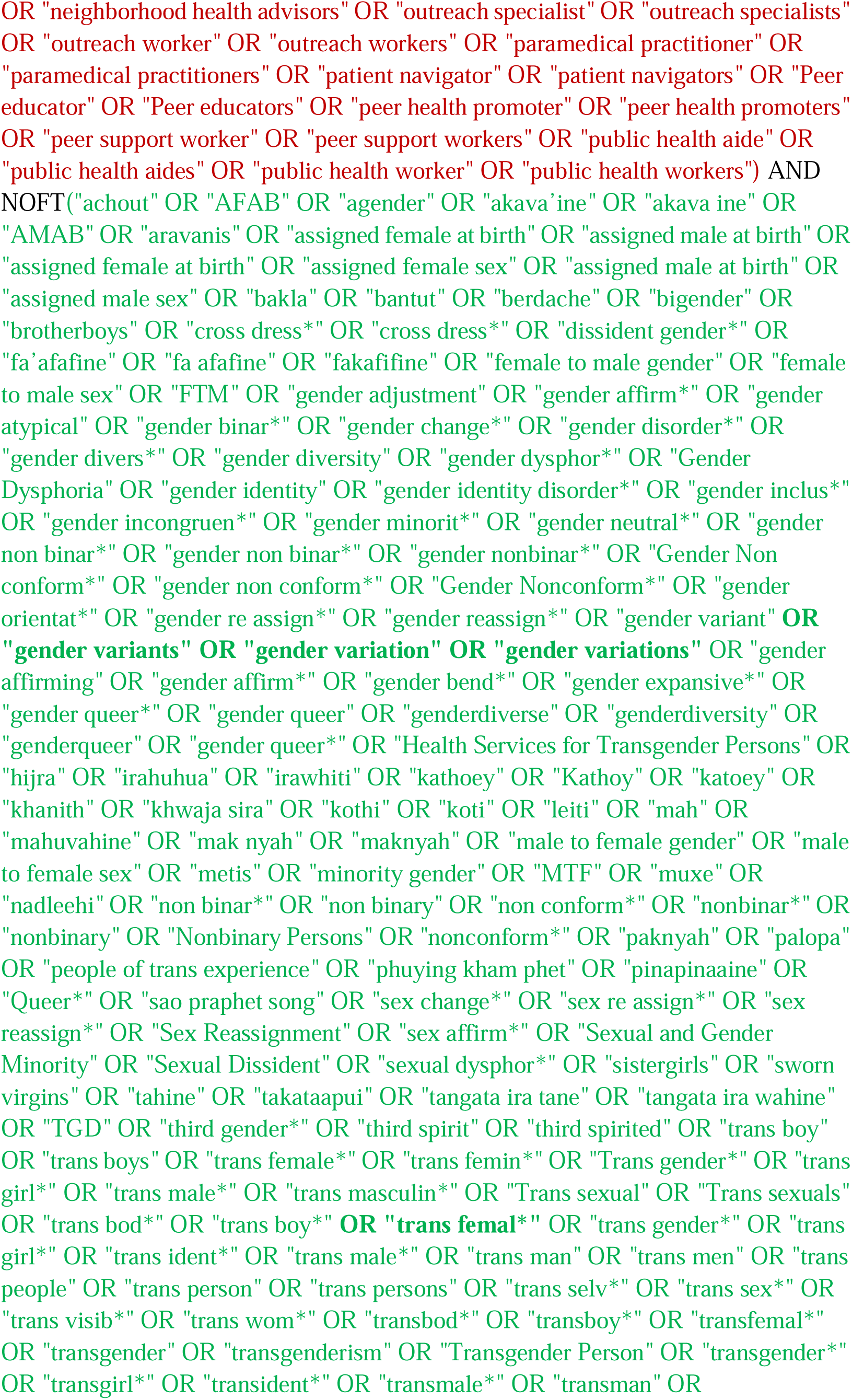
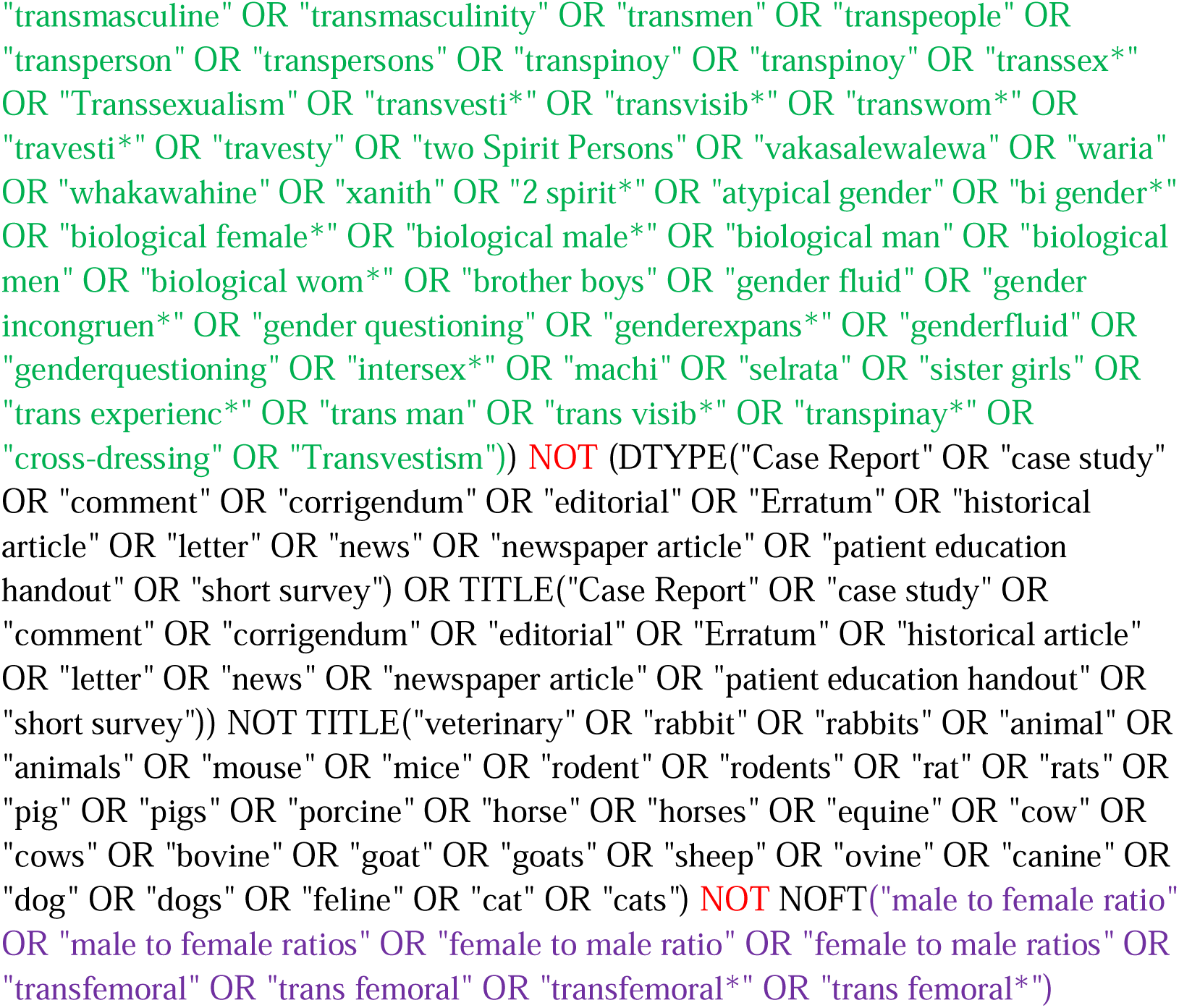

#### Academic Search Premier (EBSCOhost)

http://search.EBSCOhost.com/login.aspx?authtype=ip,uid&profile=lumc&defaultdb=aph

Limit to Academic Journals

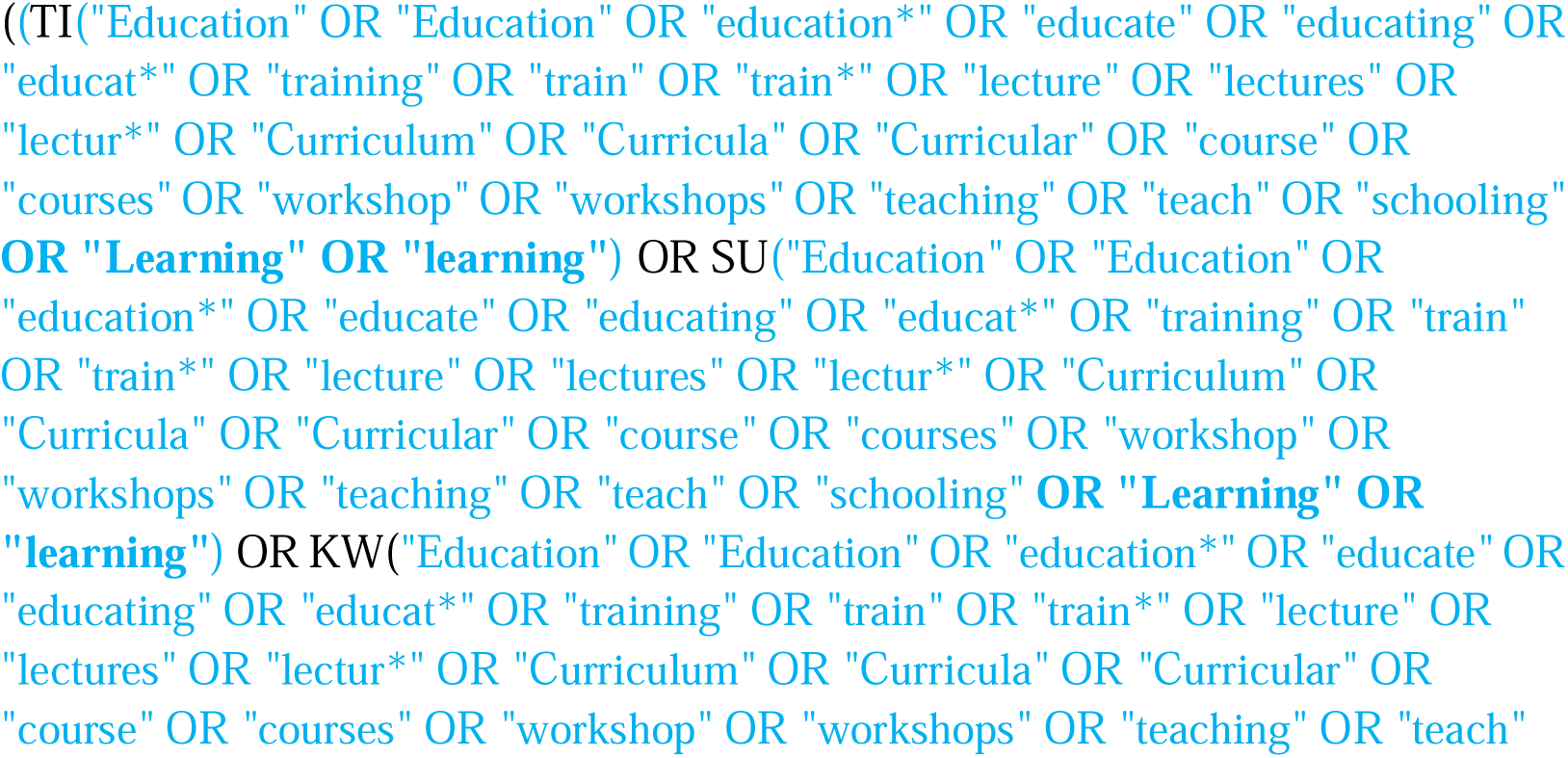
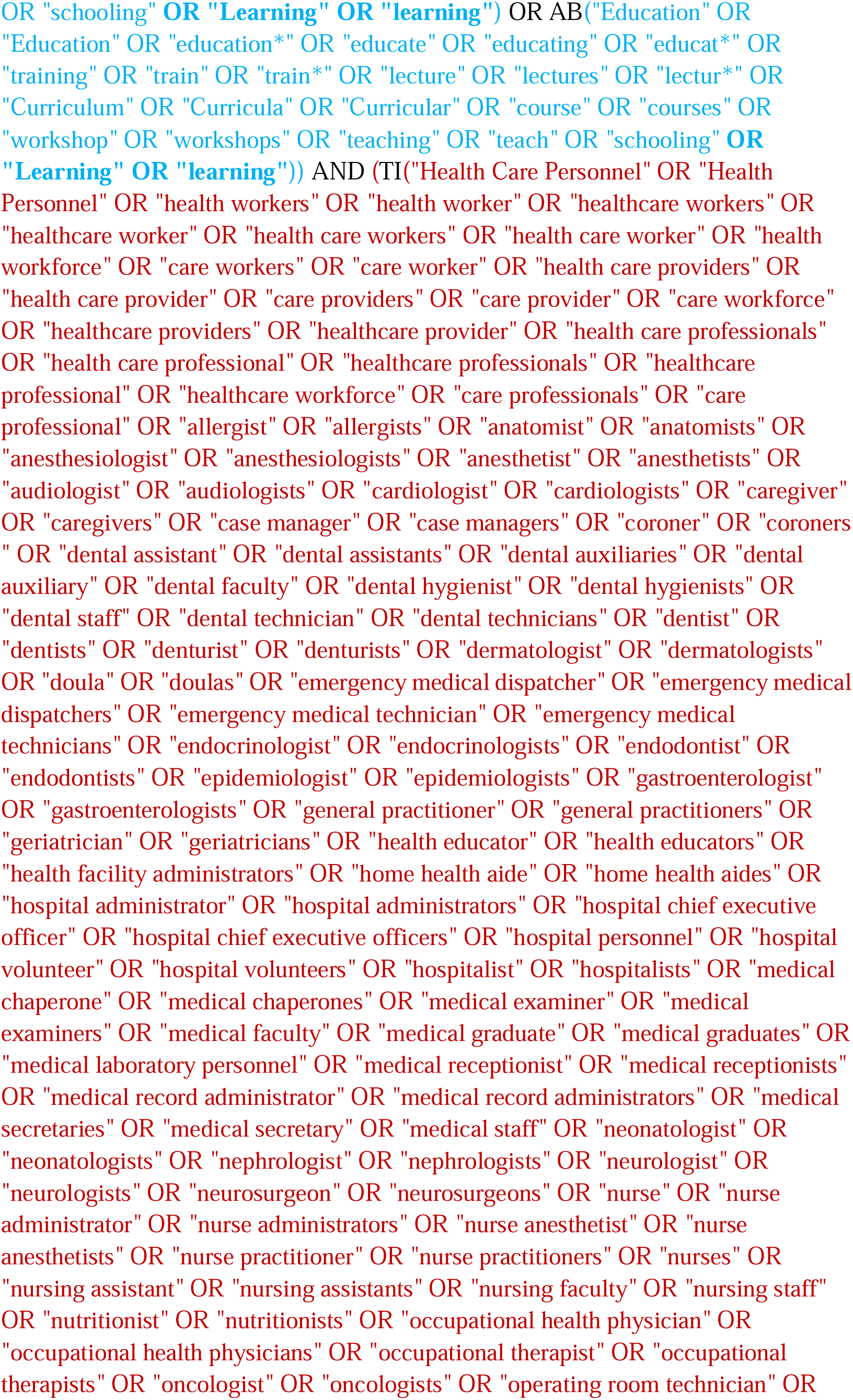
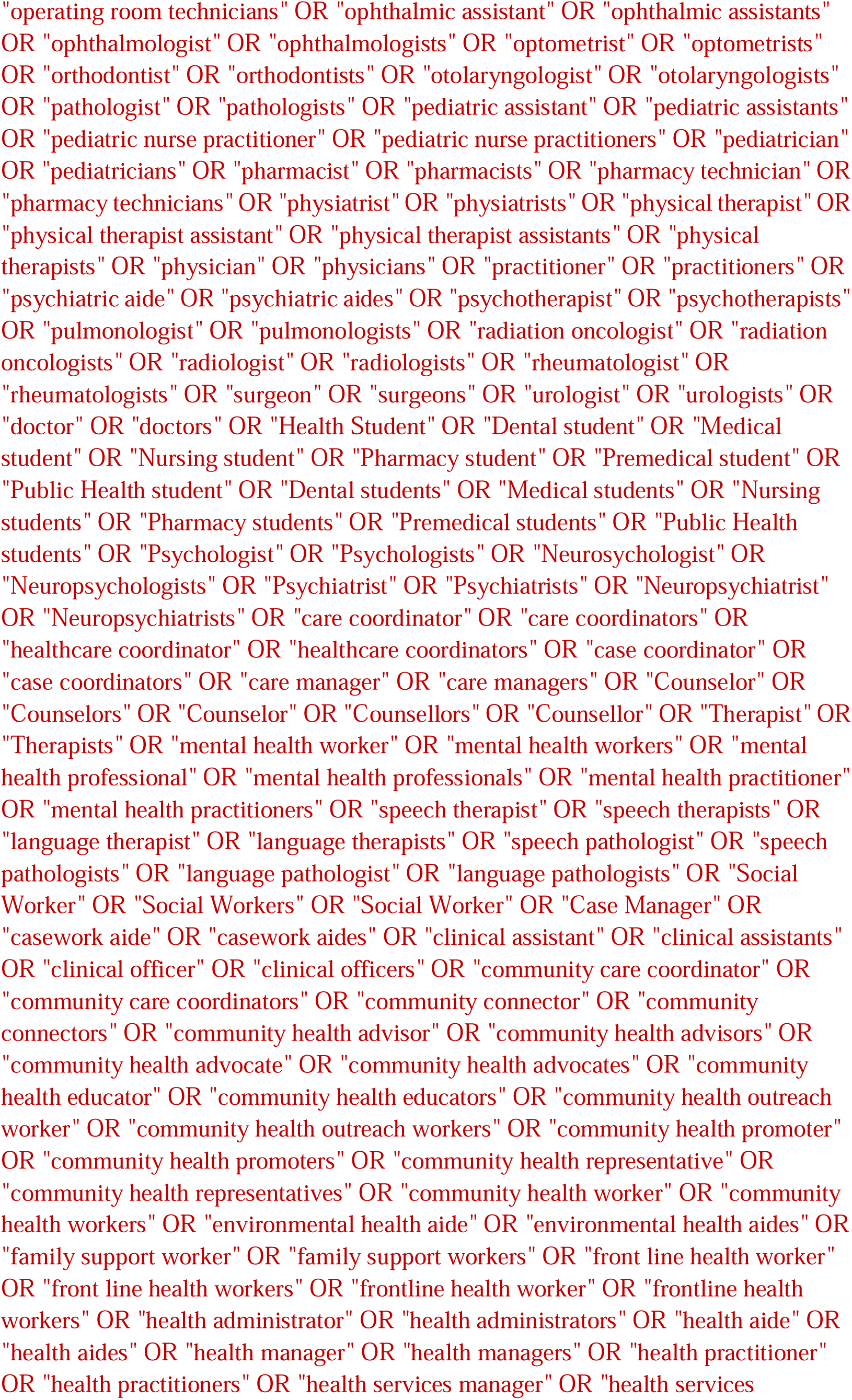
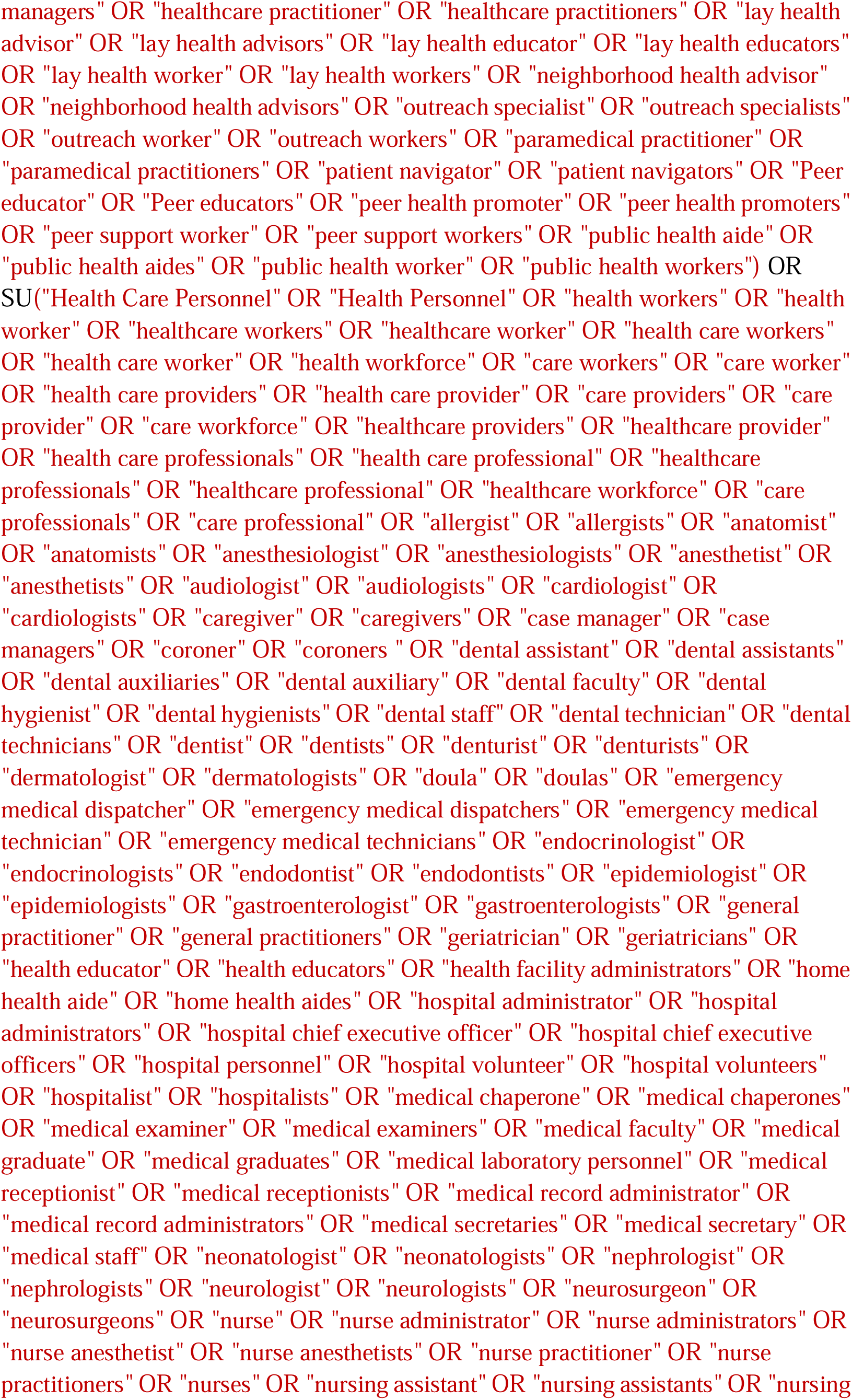
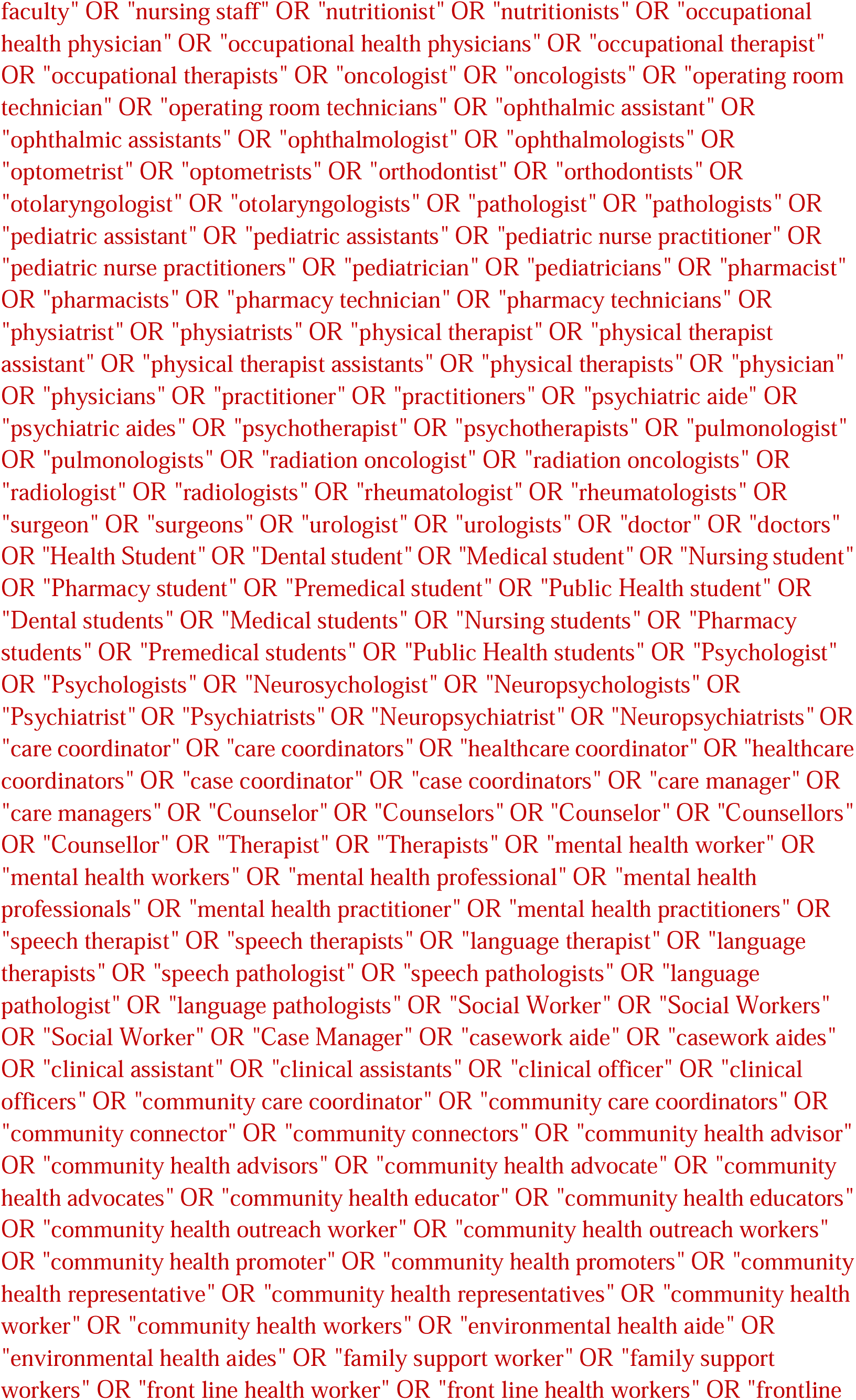
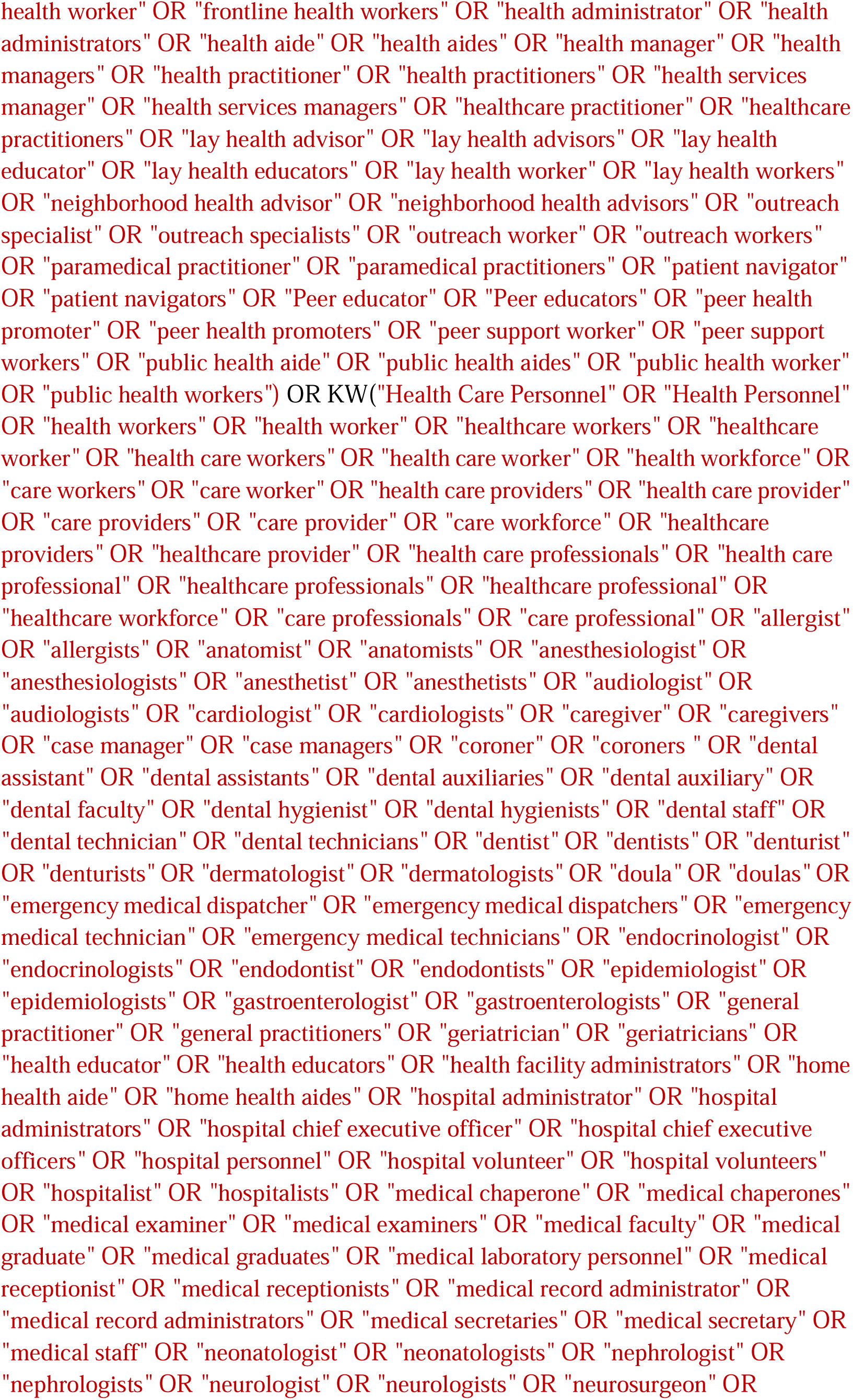
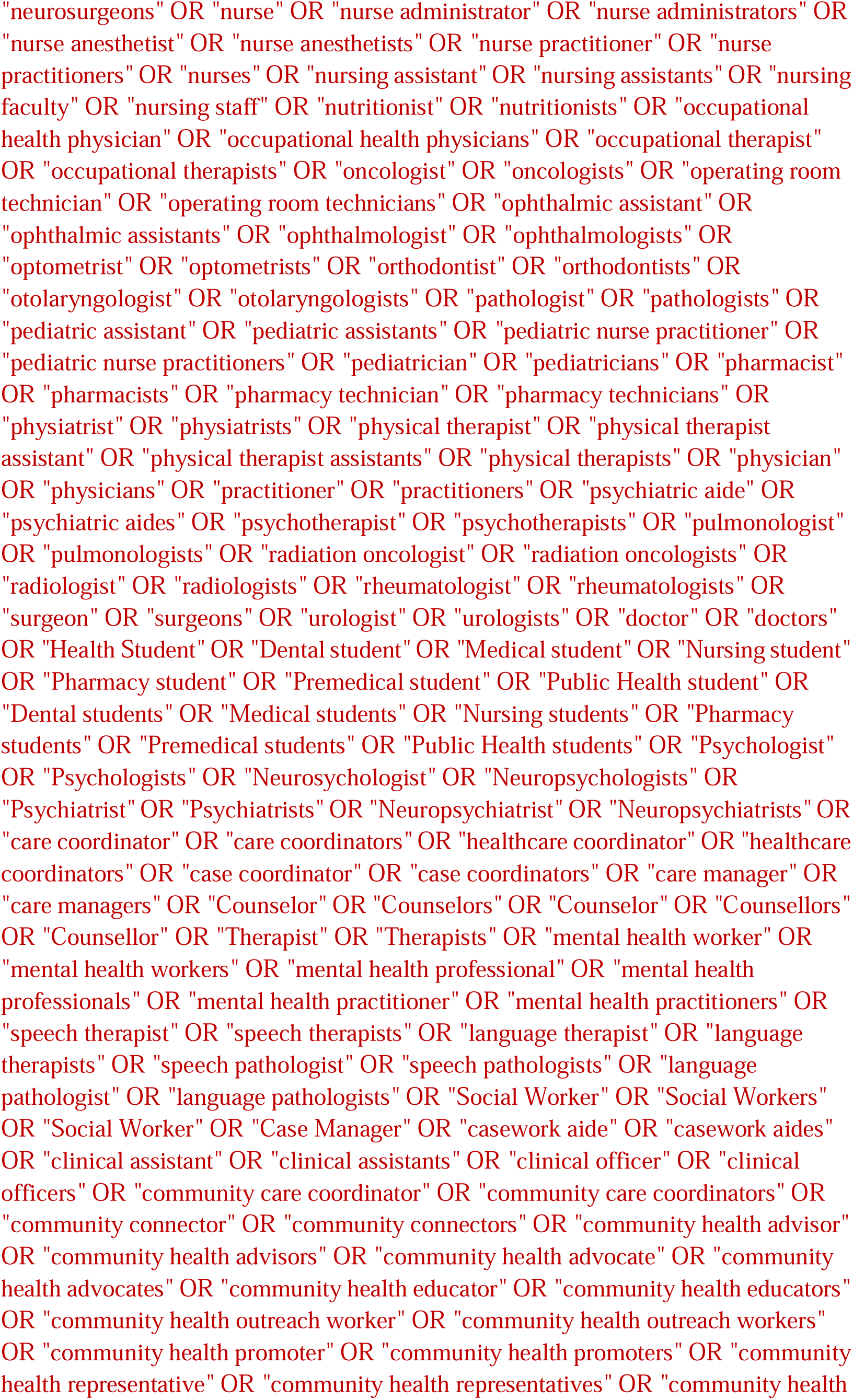
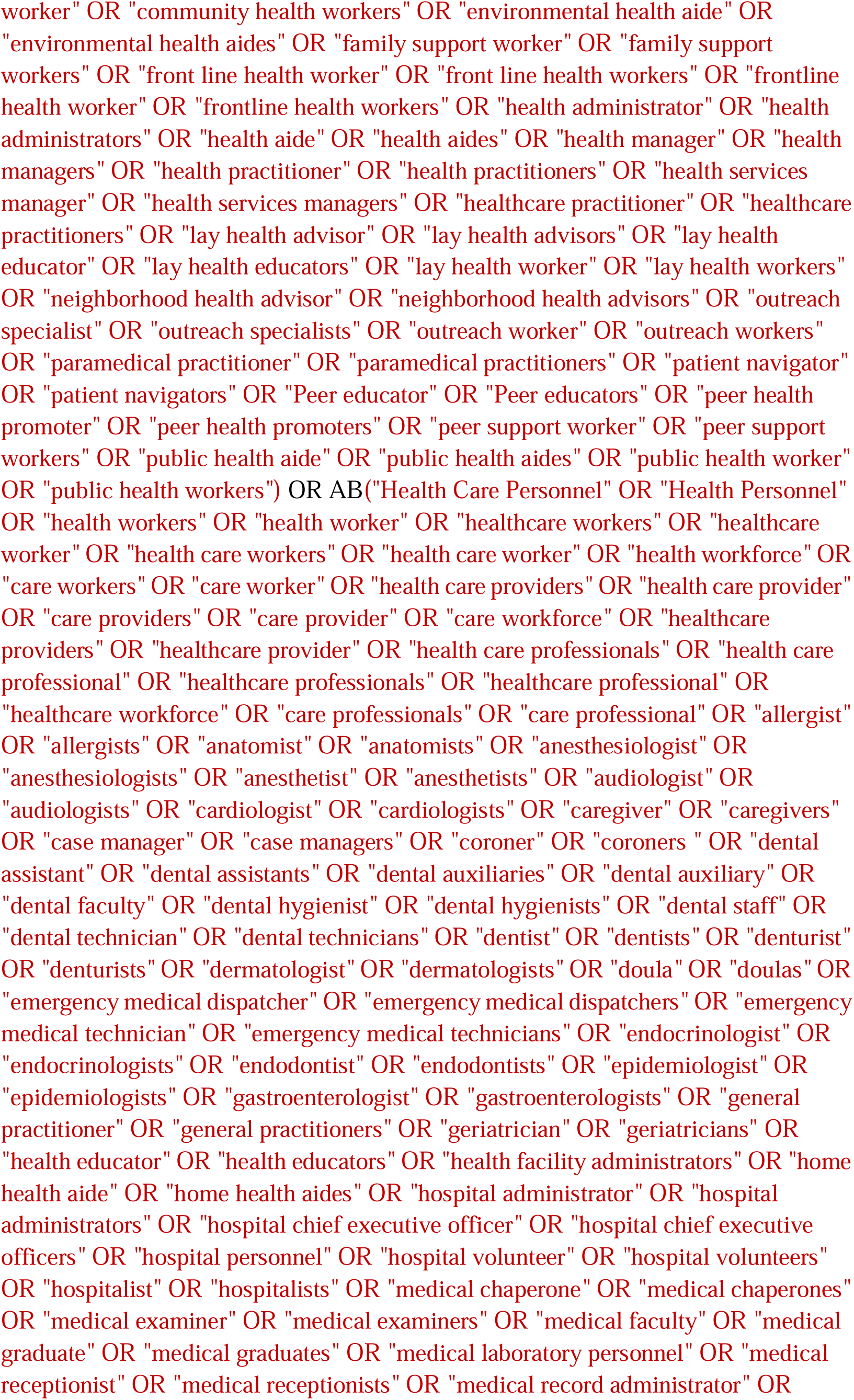
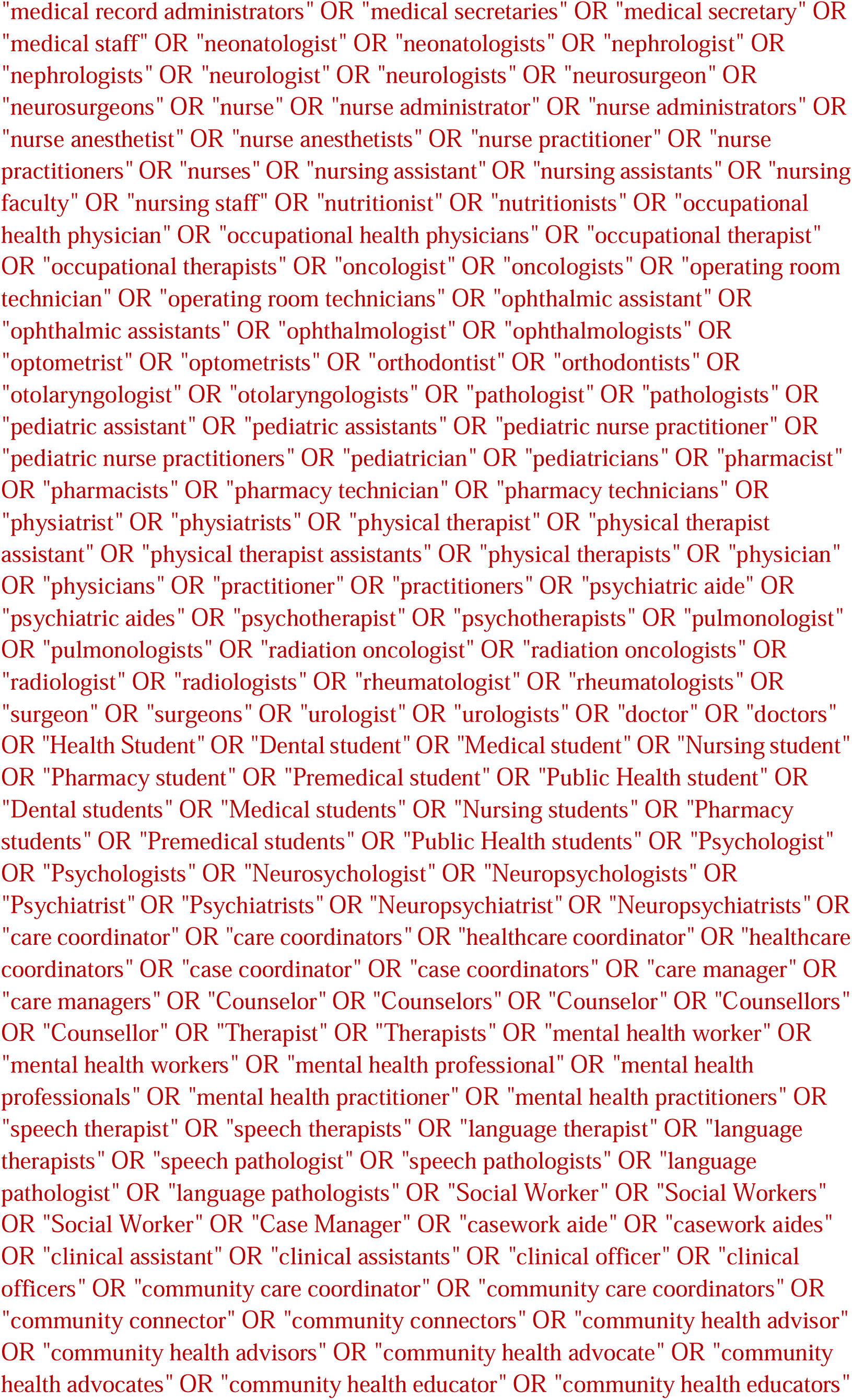
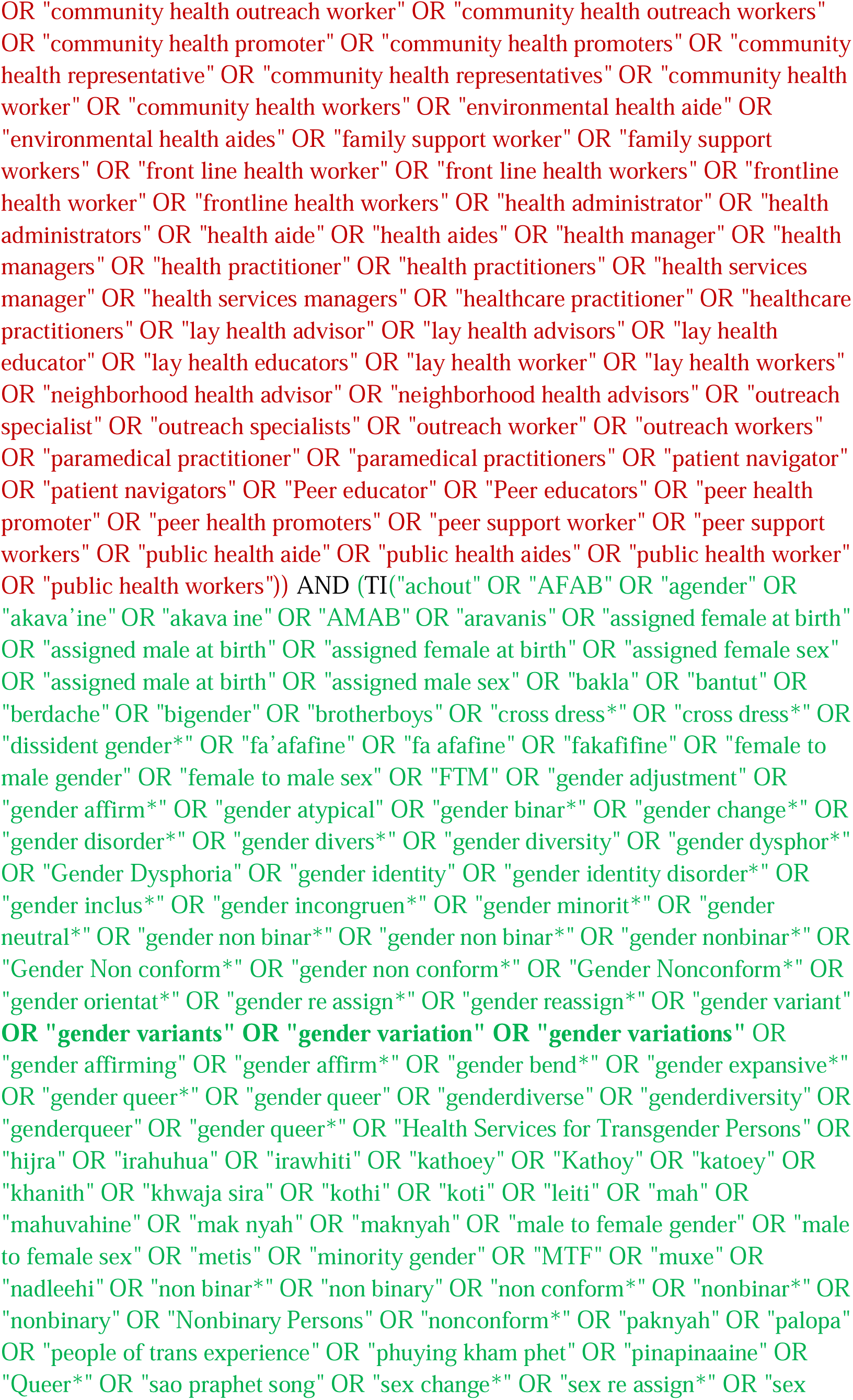
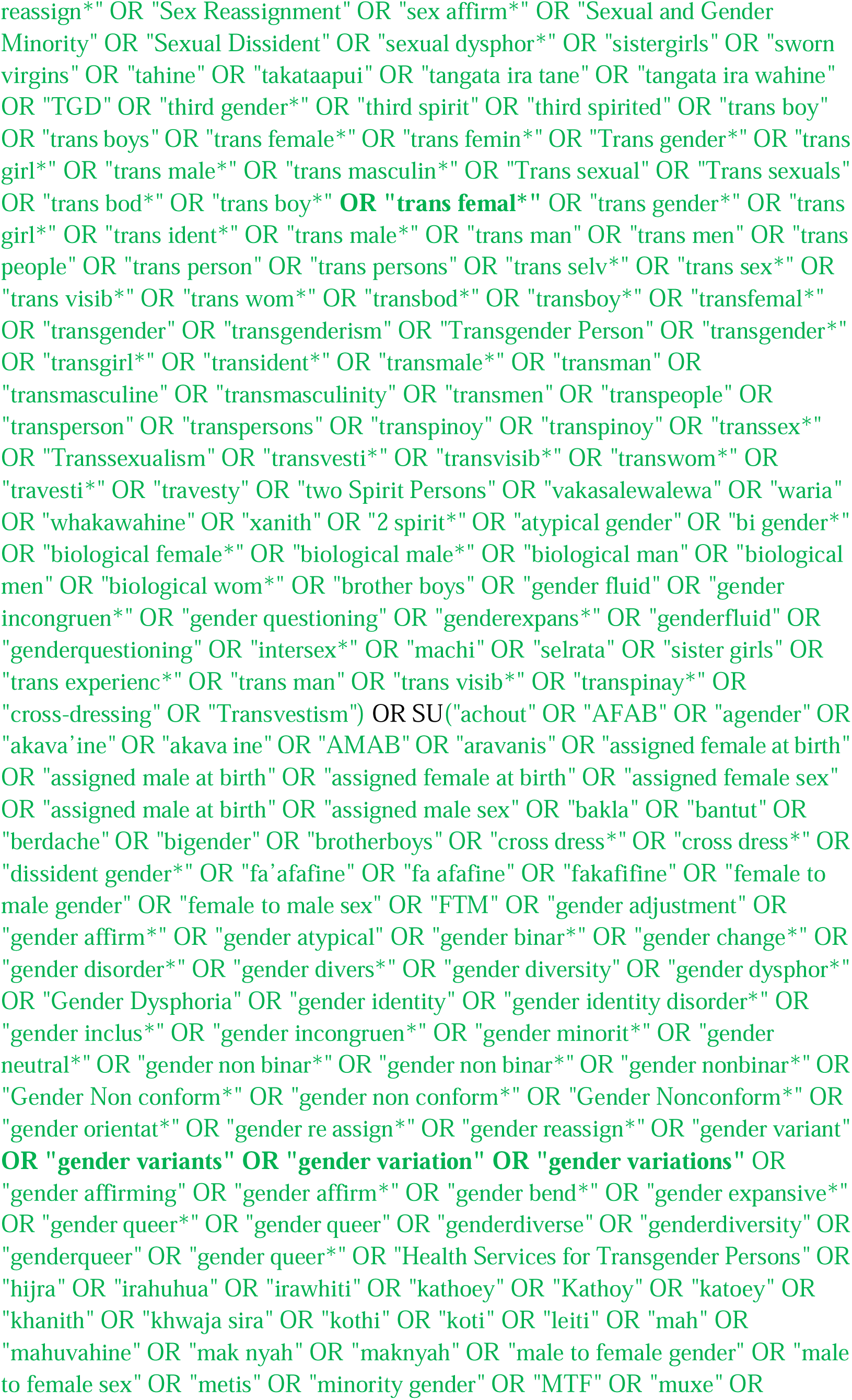
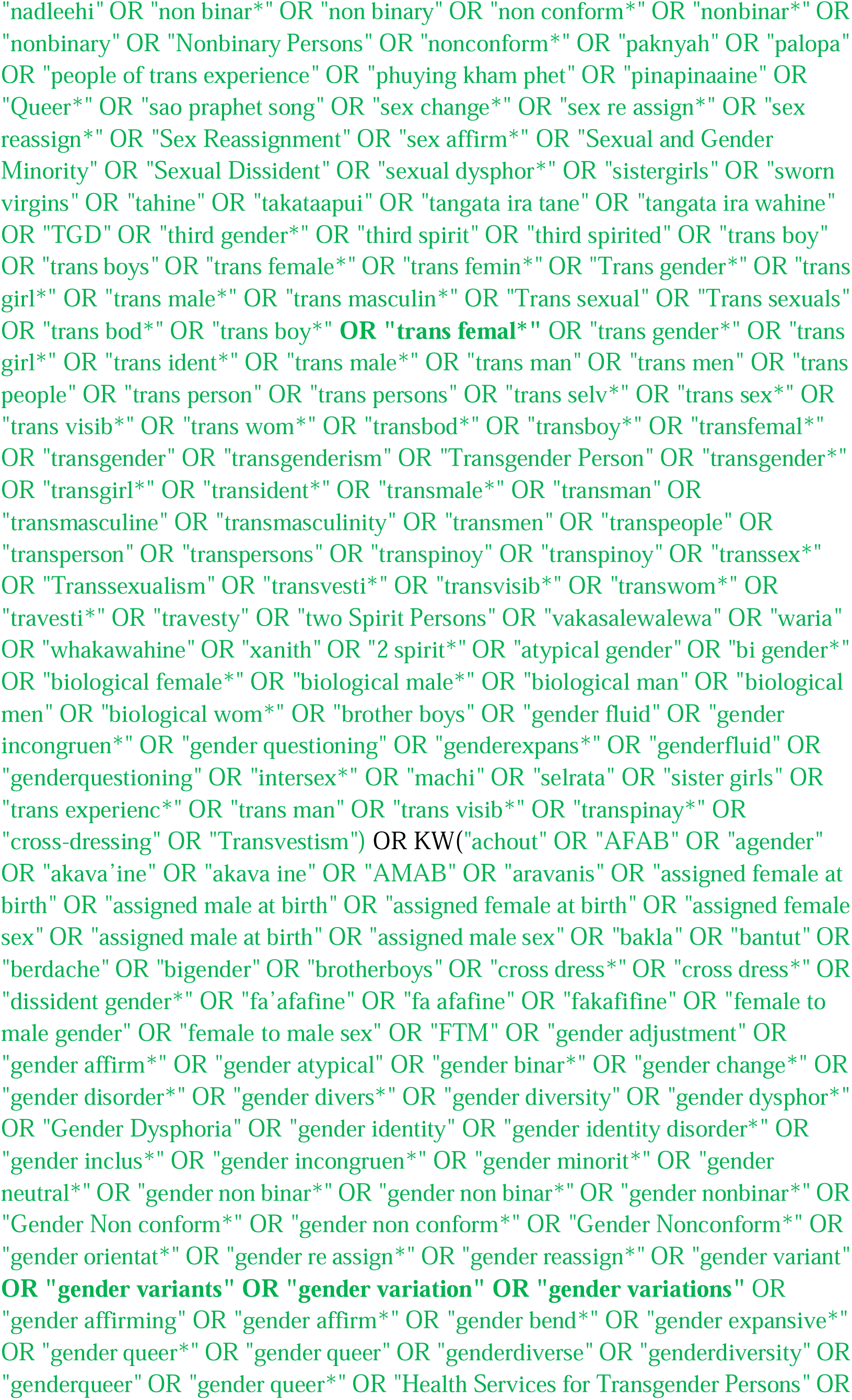
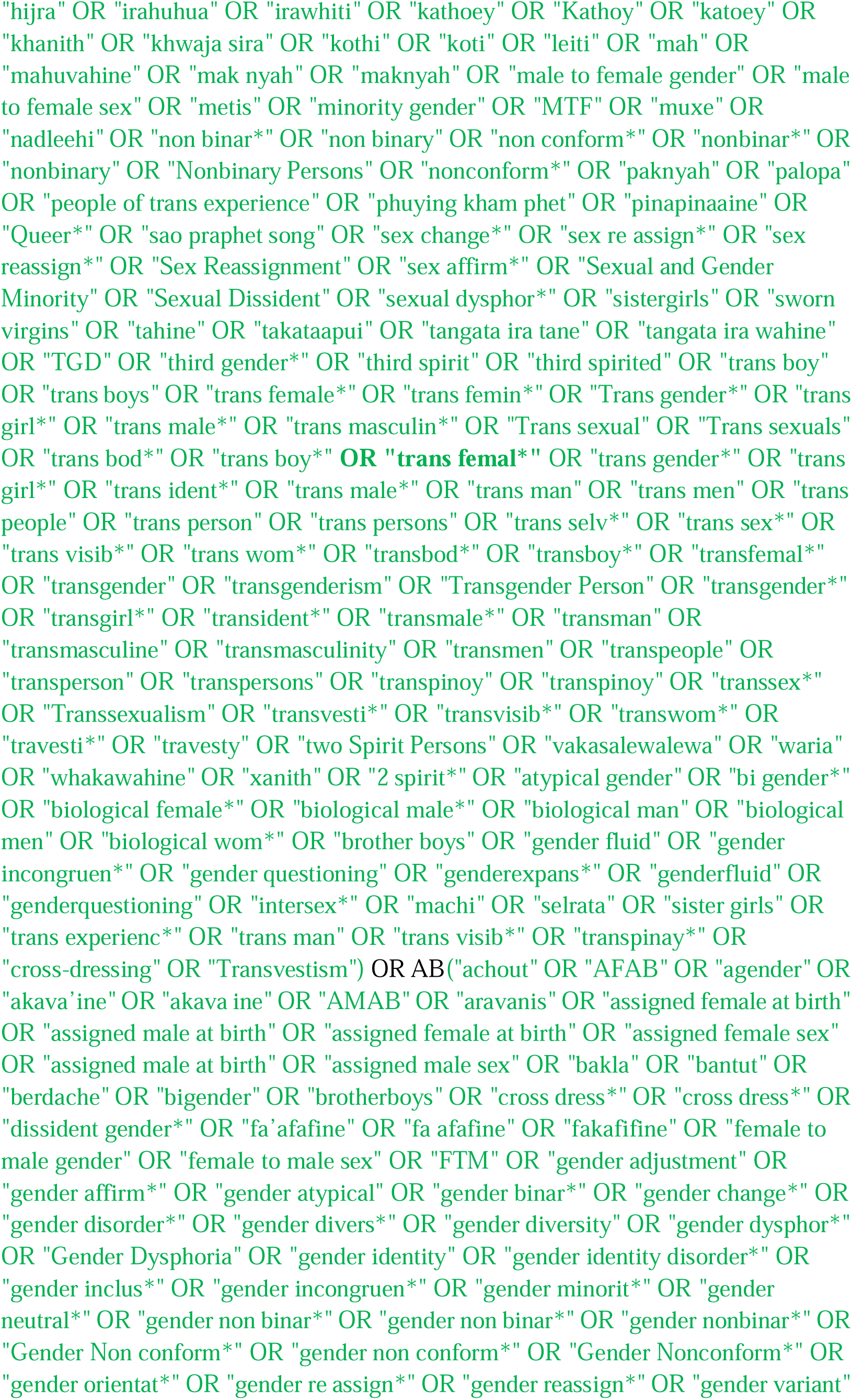
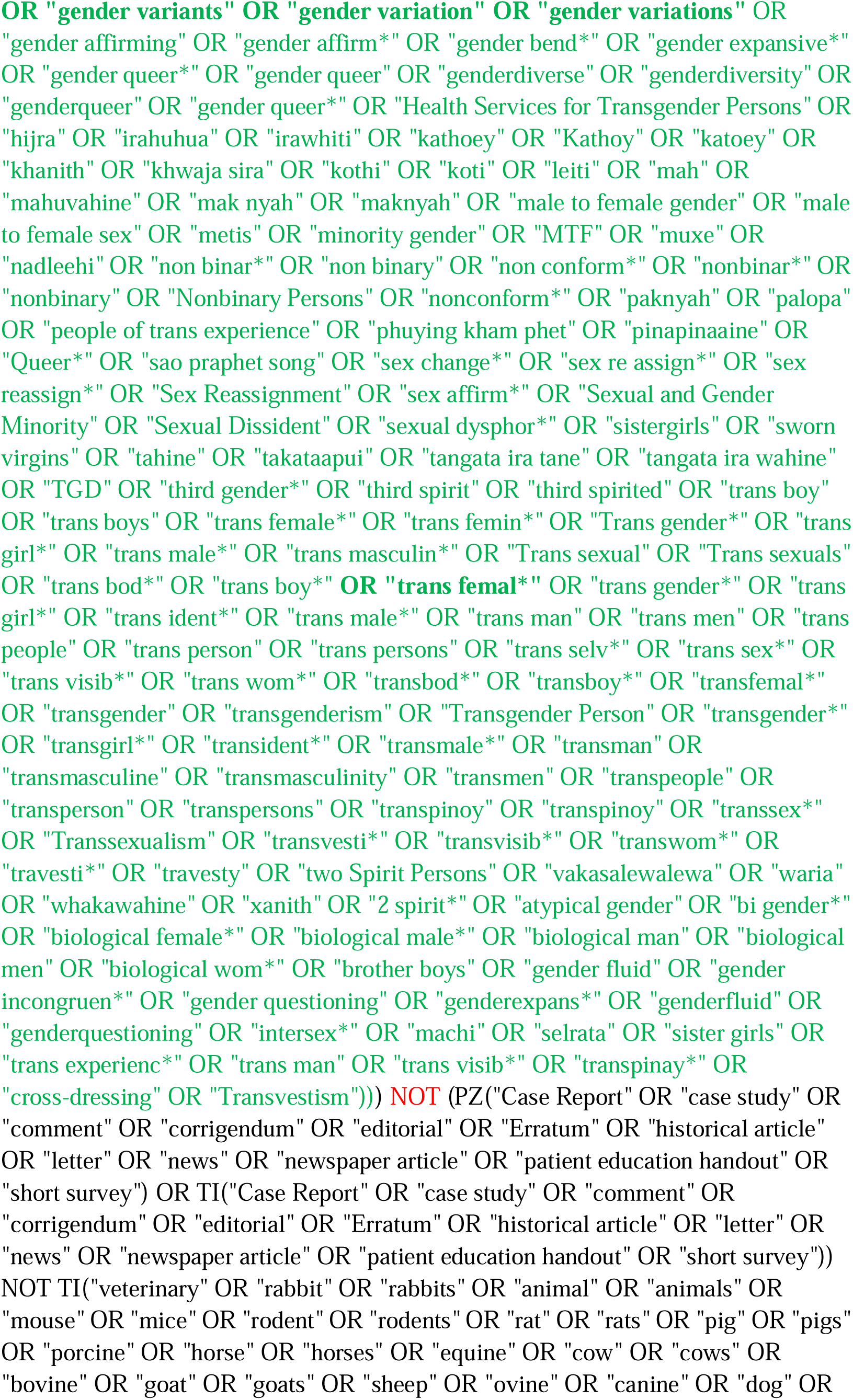
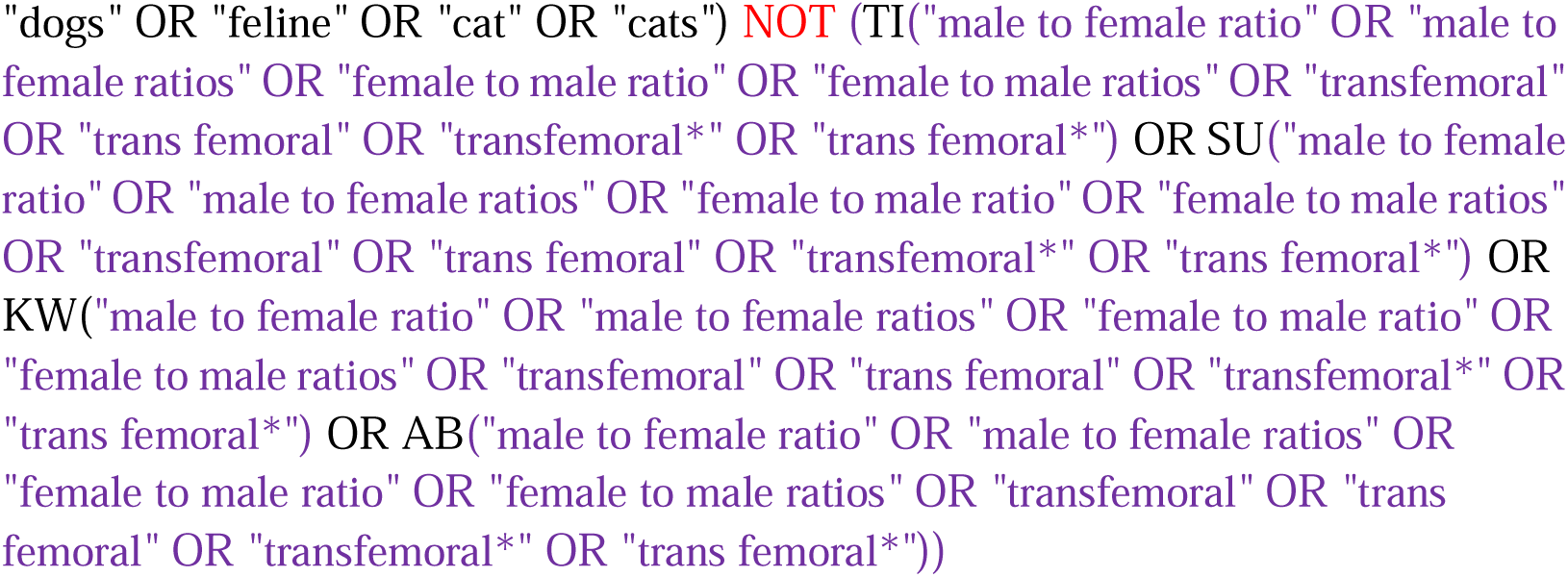

#### Psychology and Behavioral Sciences Collection (EBSCOhost)

http://search.EBSCOhost.com/login.aspx?authtype=ip,uid&profile=lumc&defaultdb=aph

Limit to Academic Journals

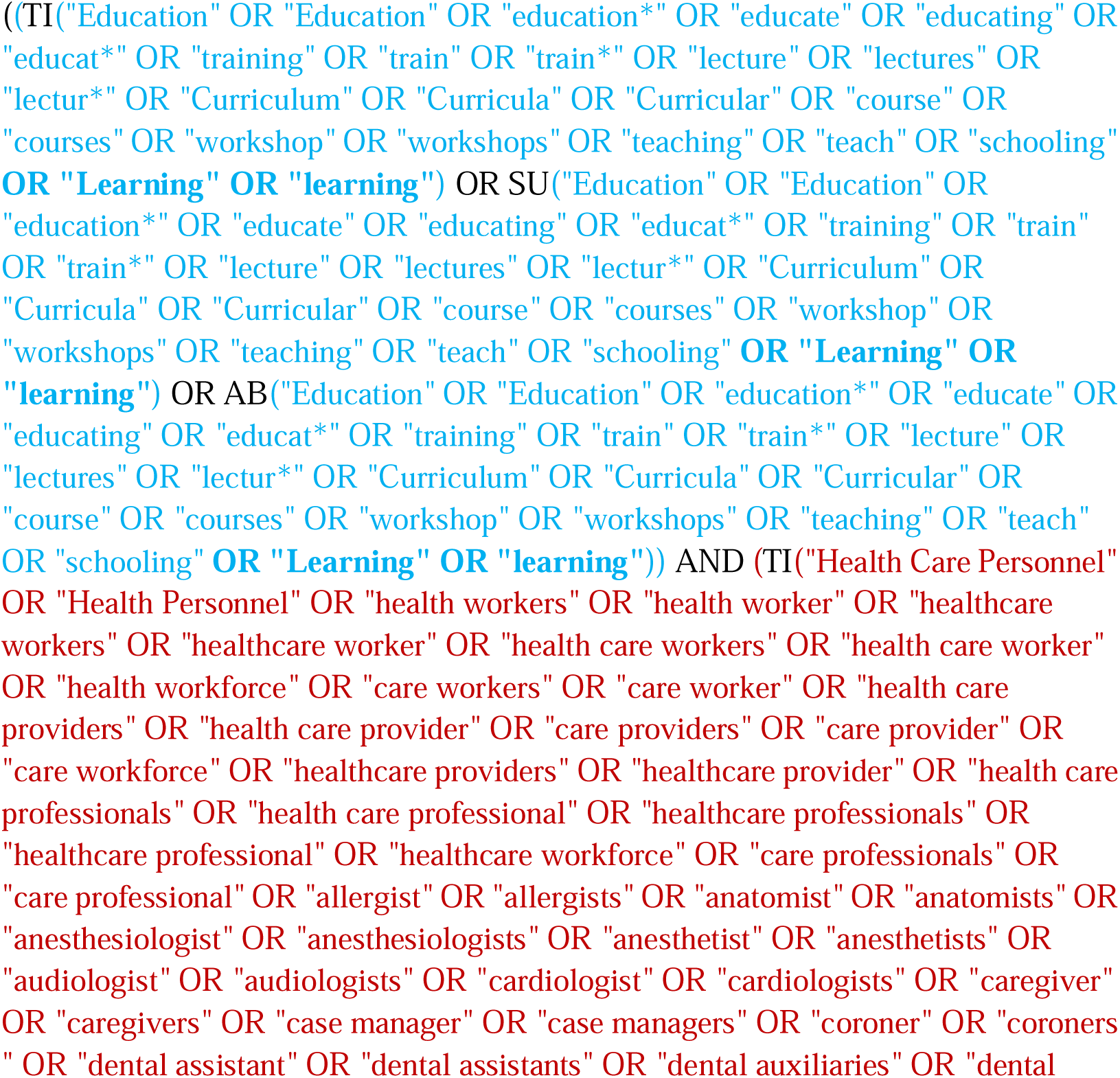
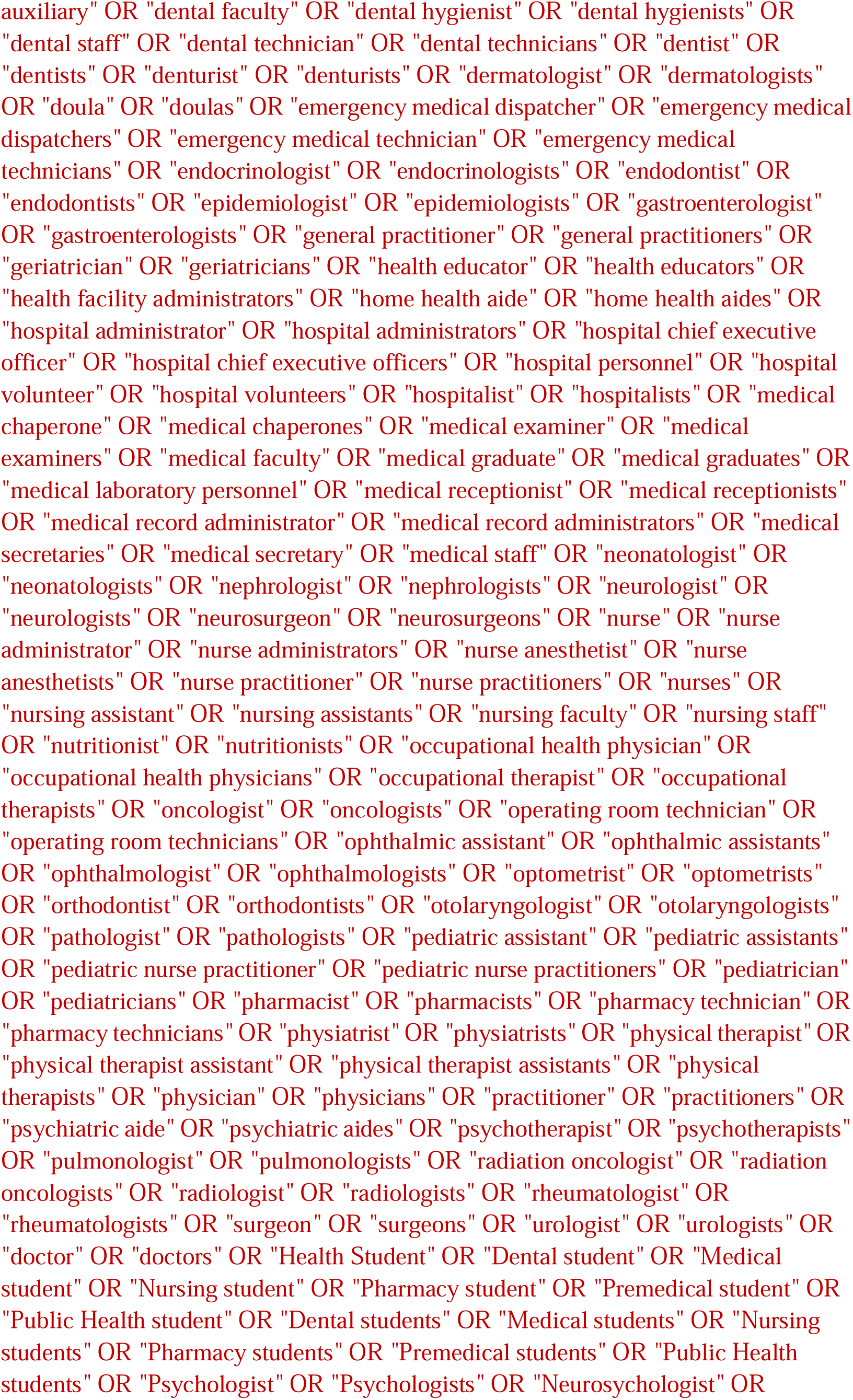
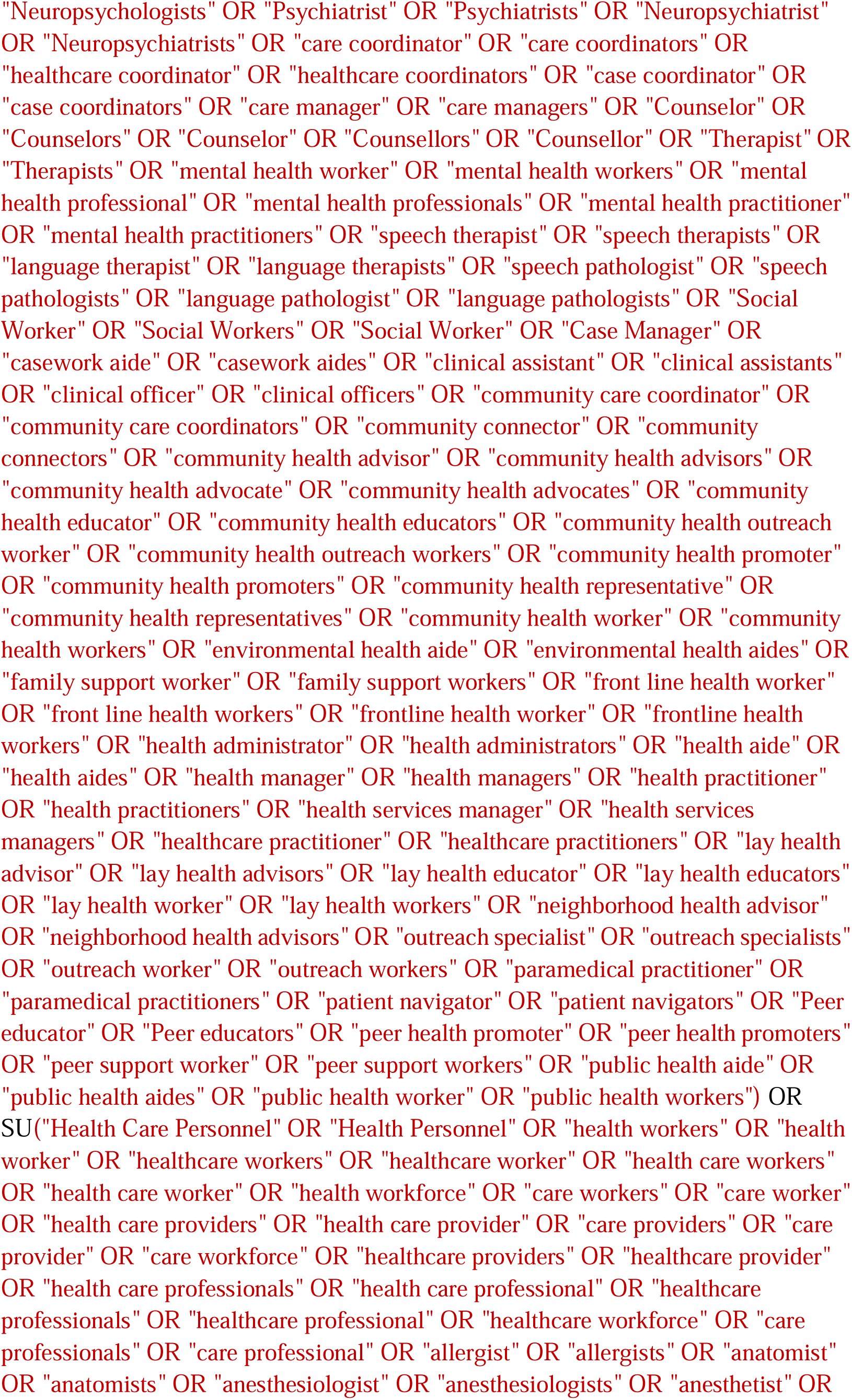
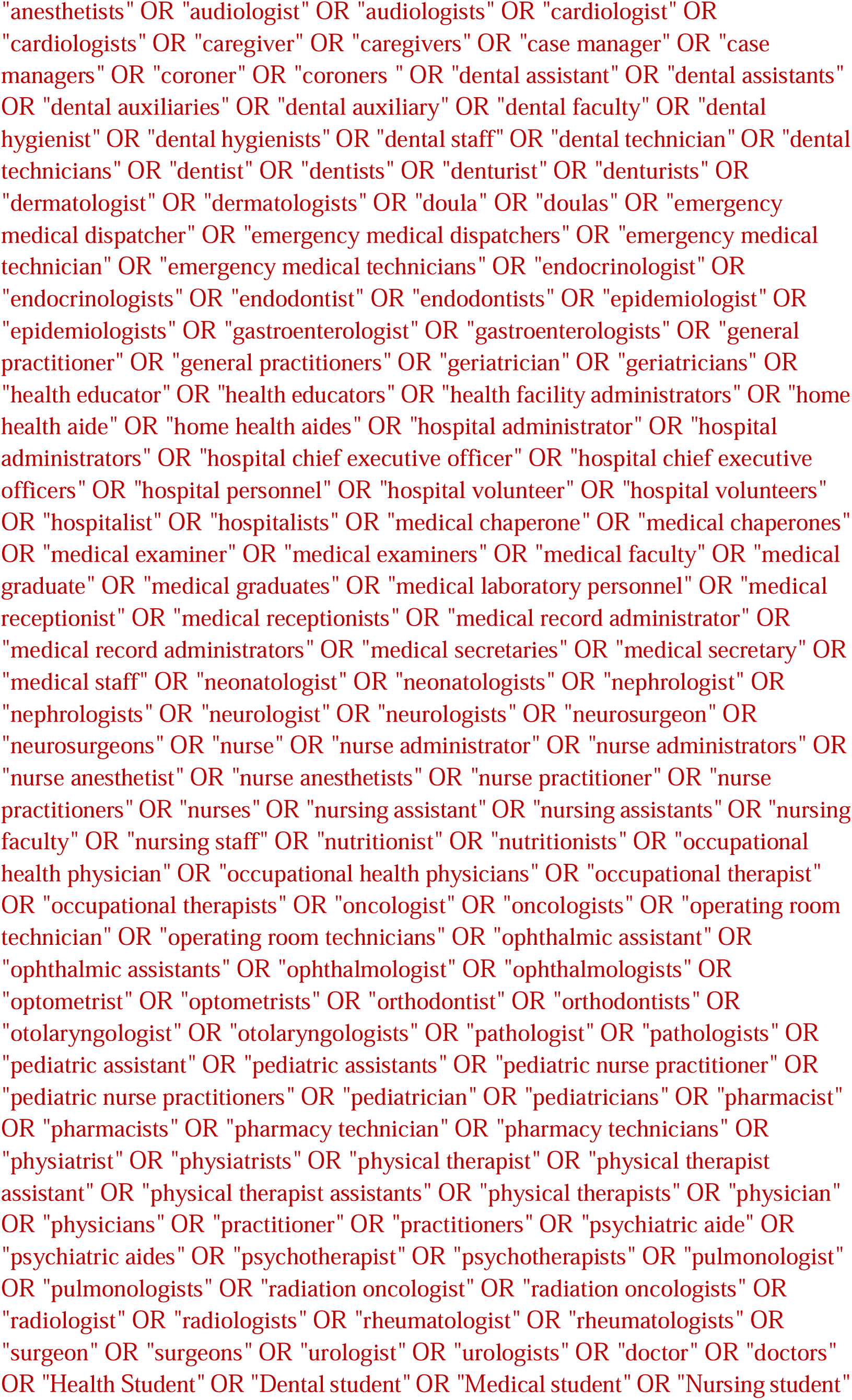
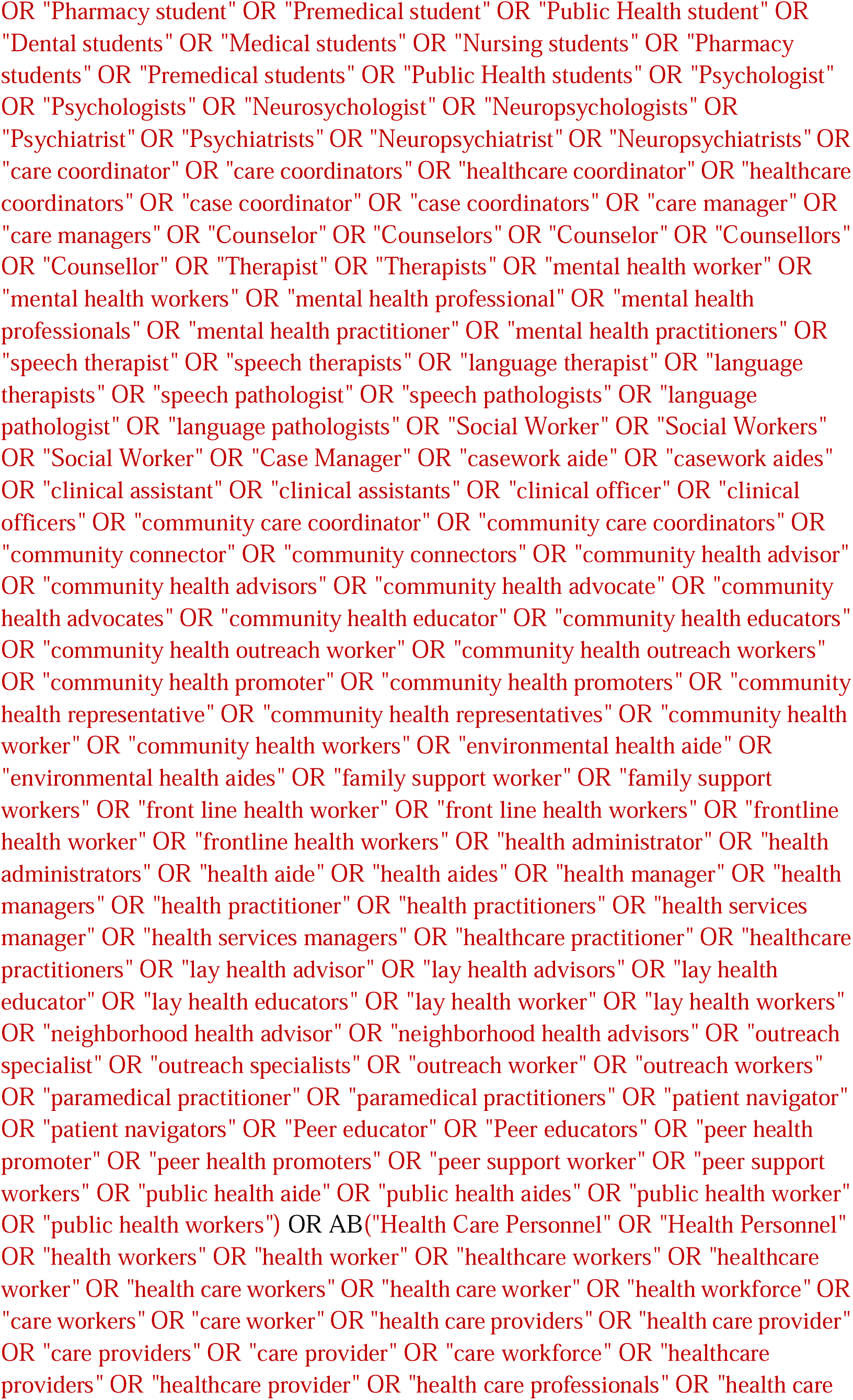
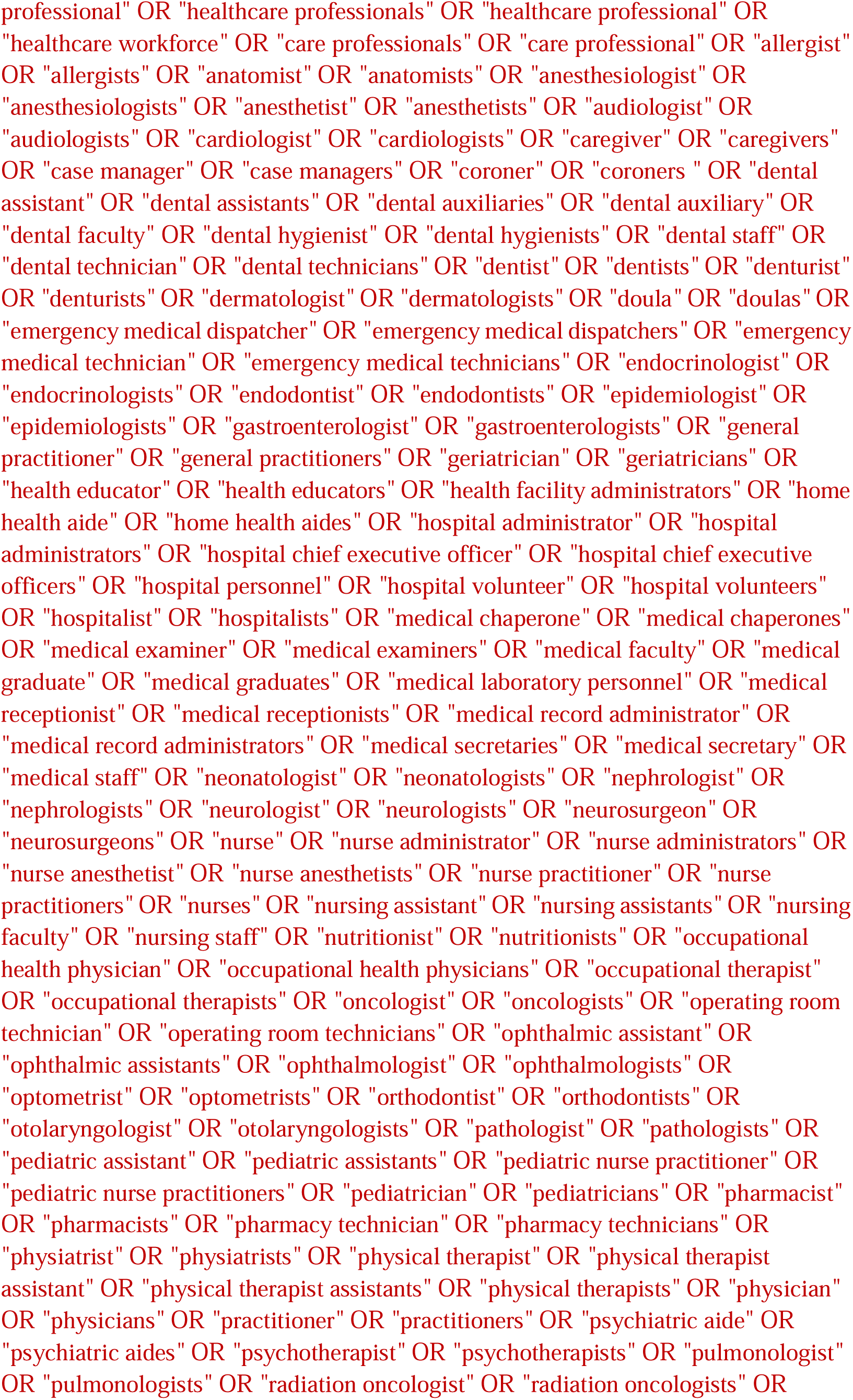
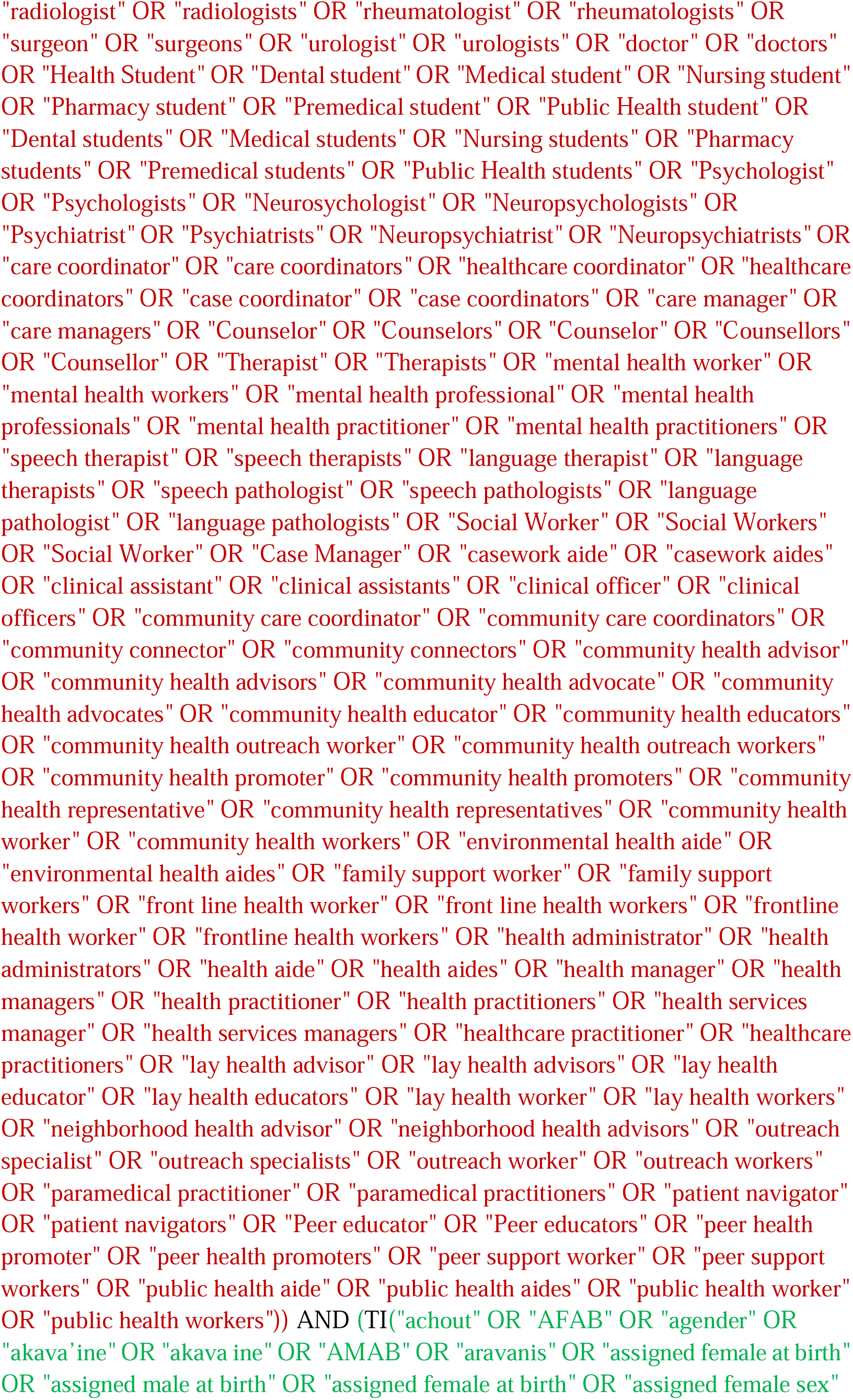
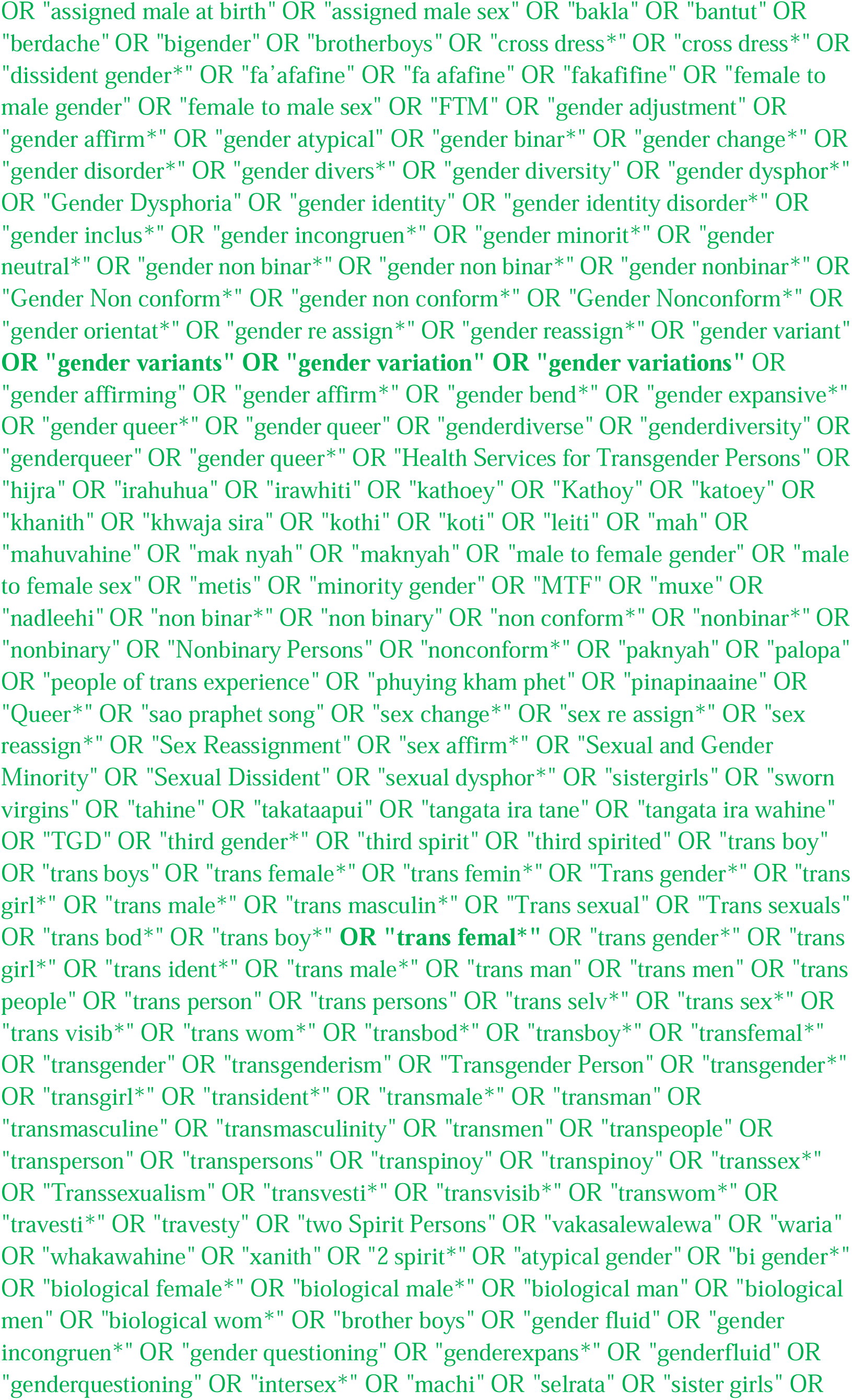
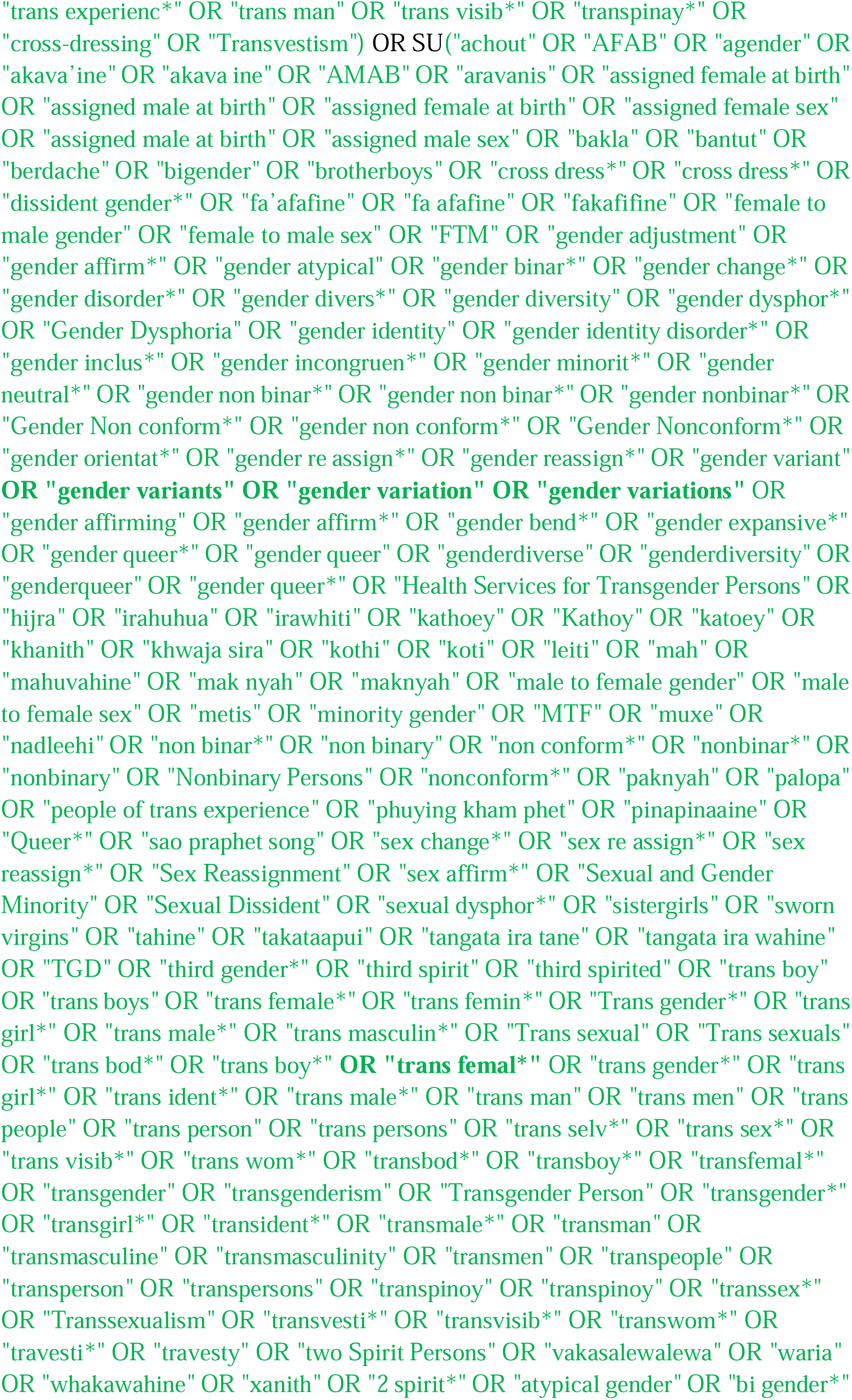
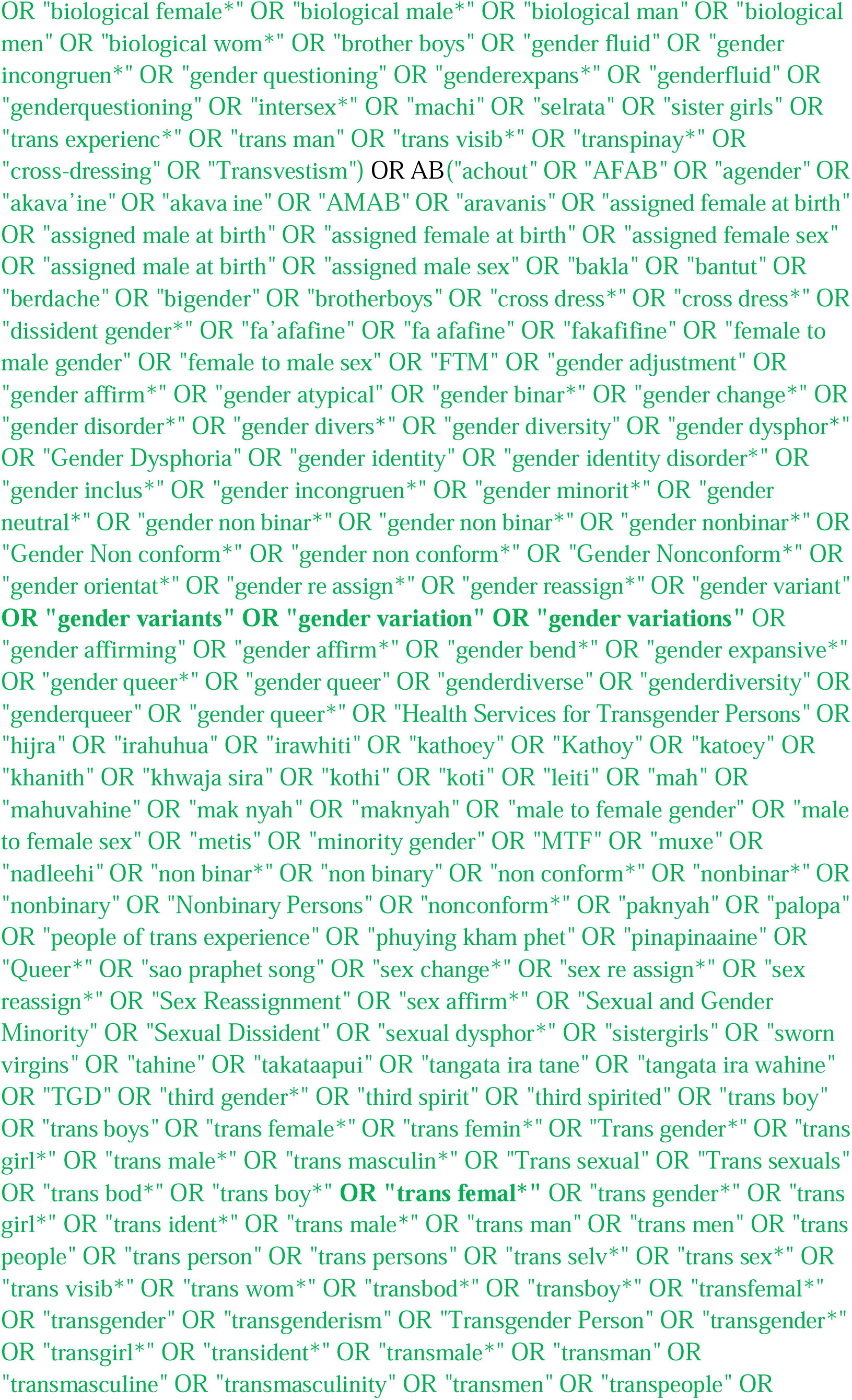
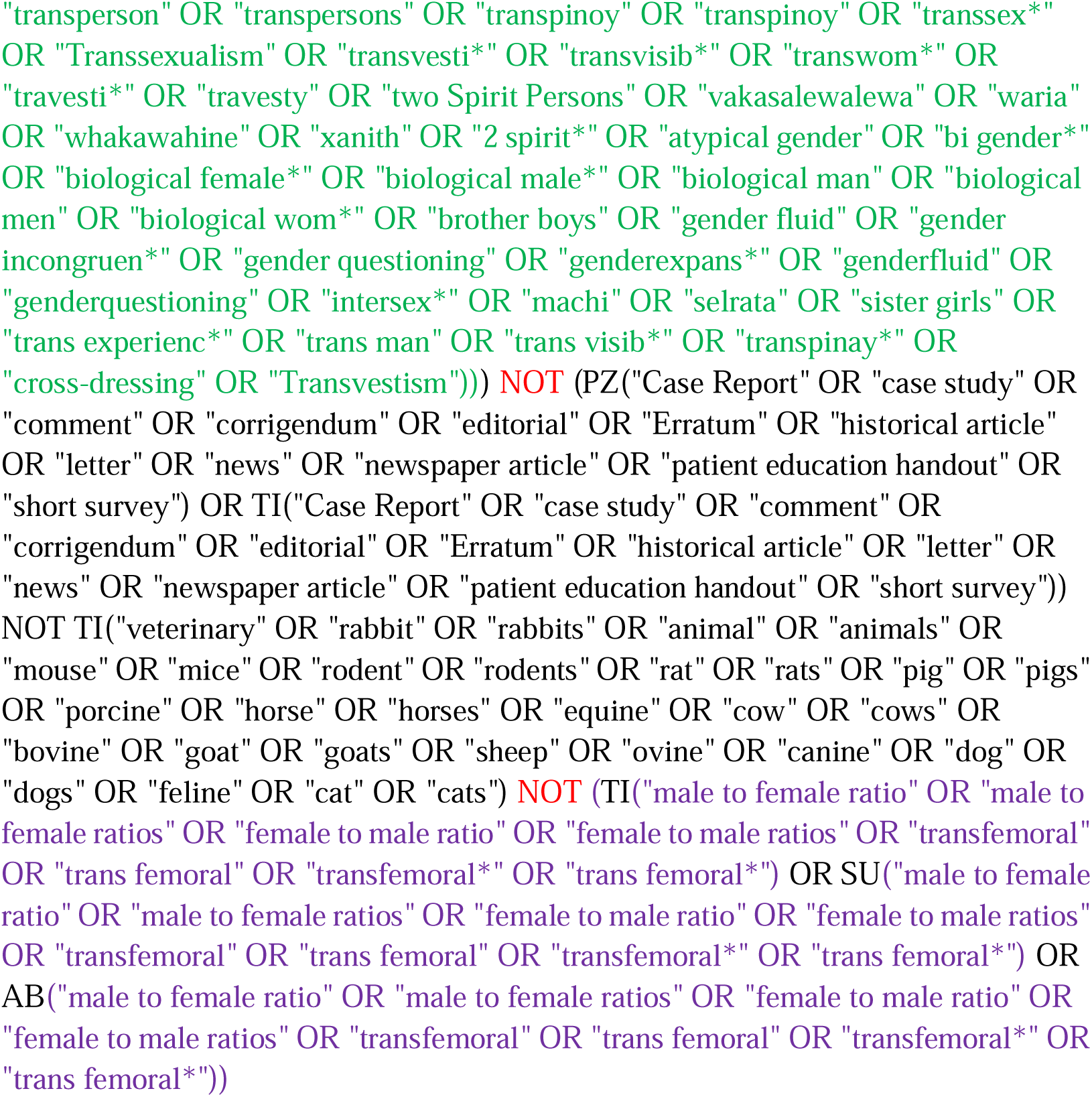

#### ERIC (EBSCOhost)

Via Ebsco: http://databases.library.leiden.edu/?bibid=990024848320302711&redirect=true

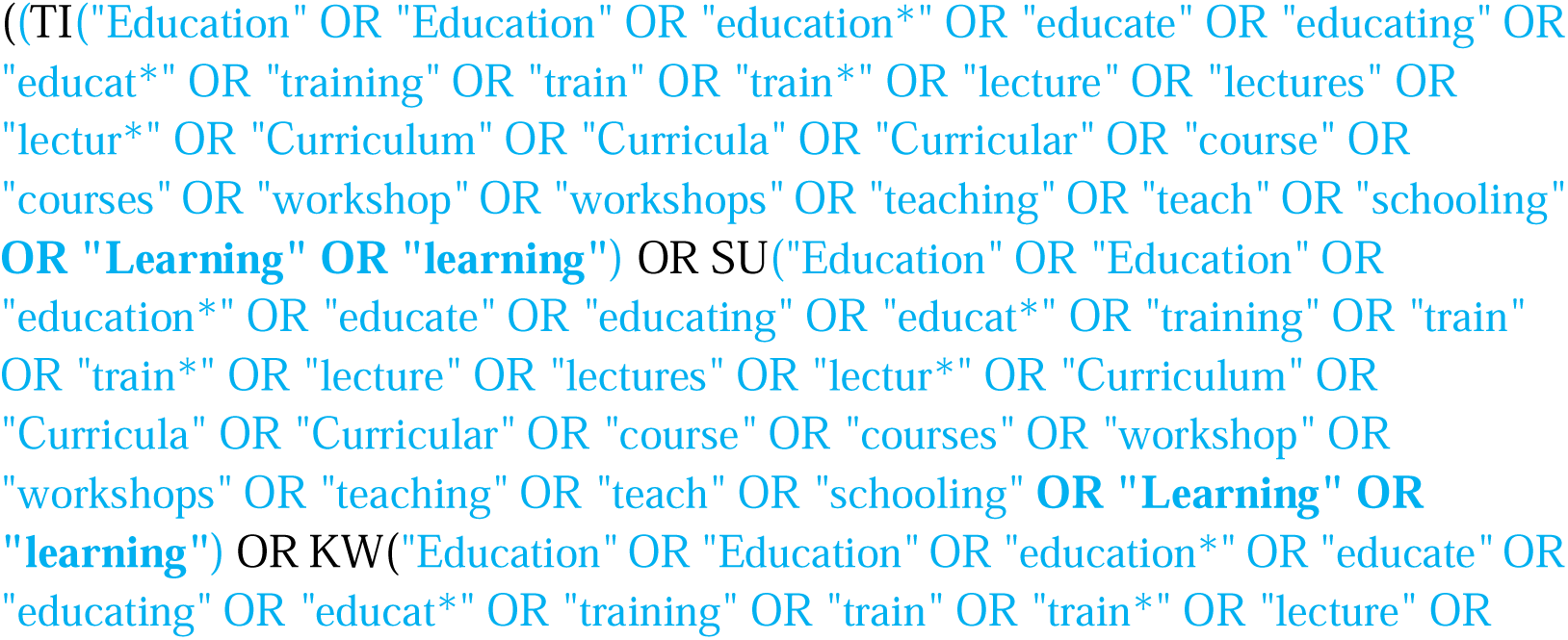
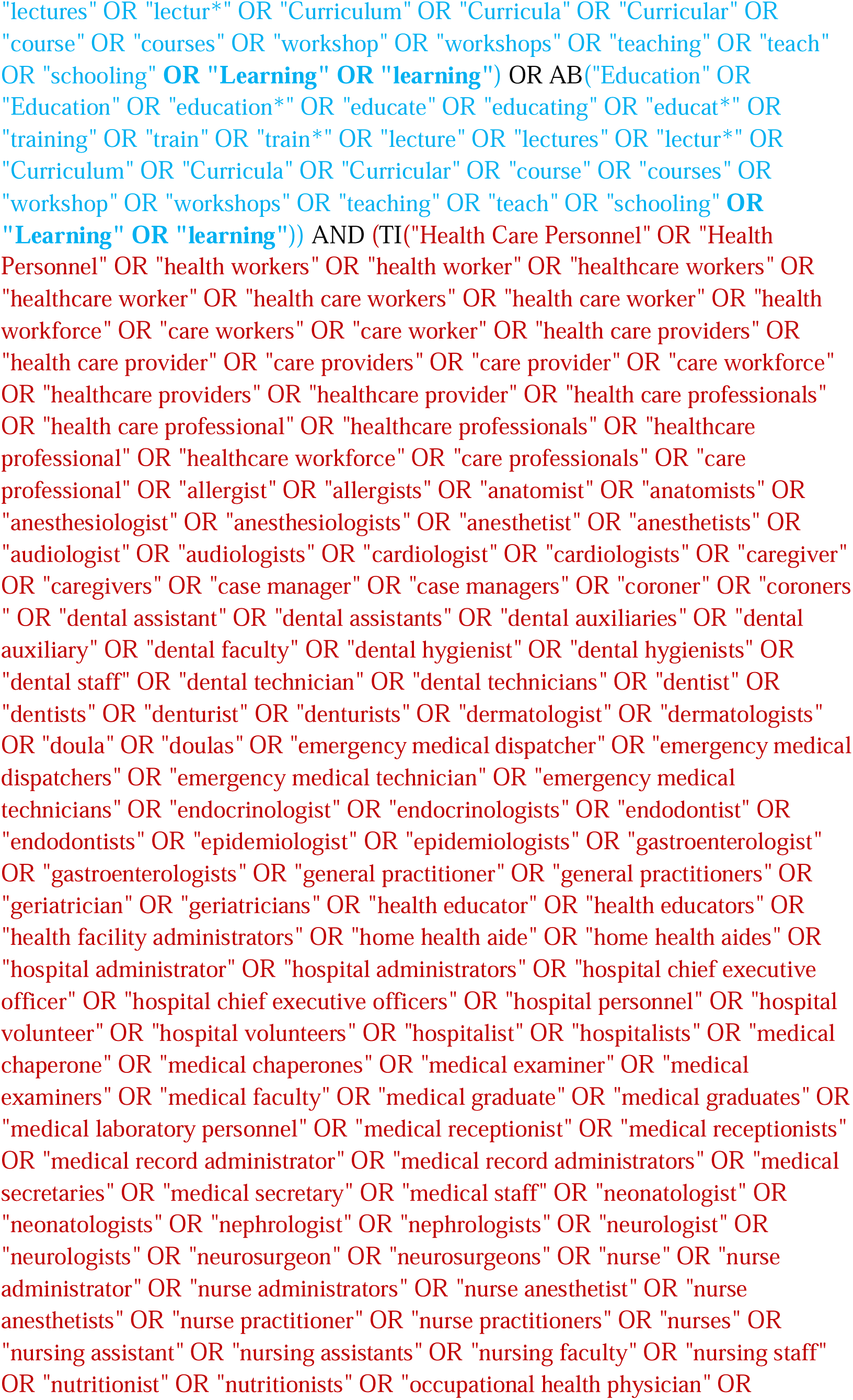
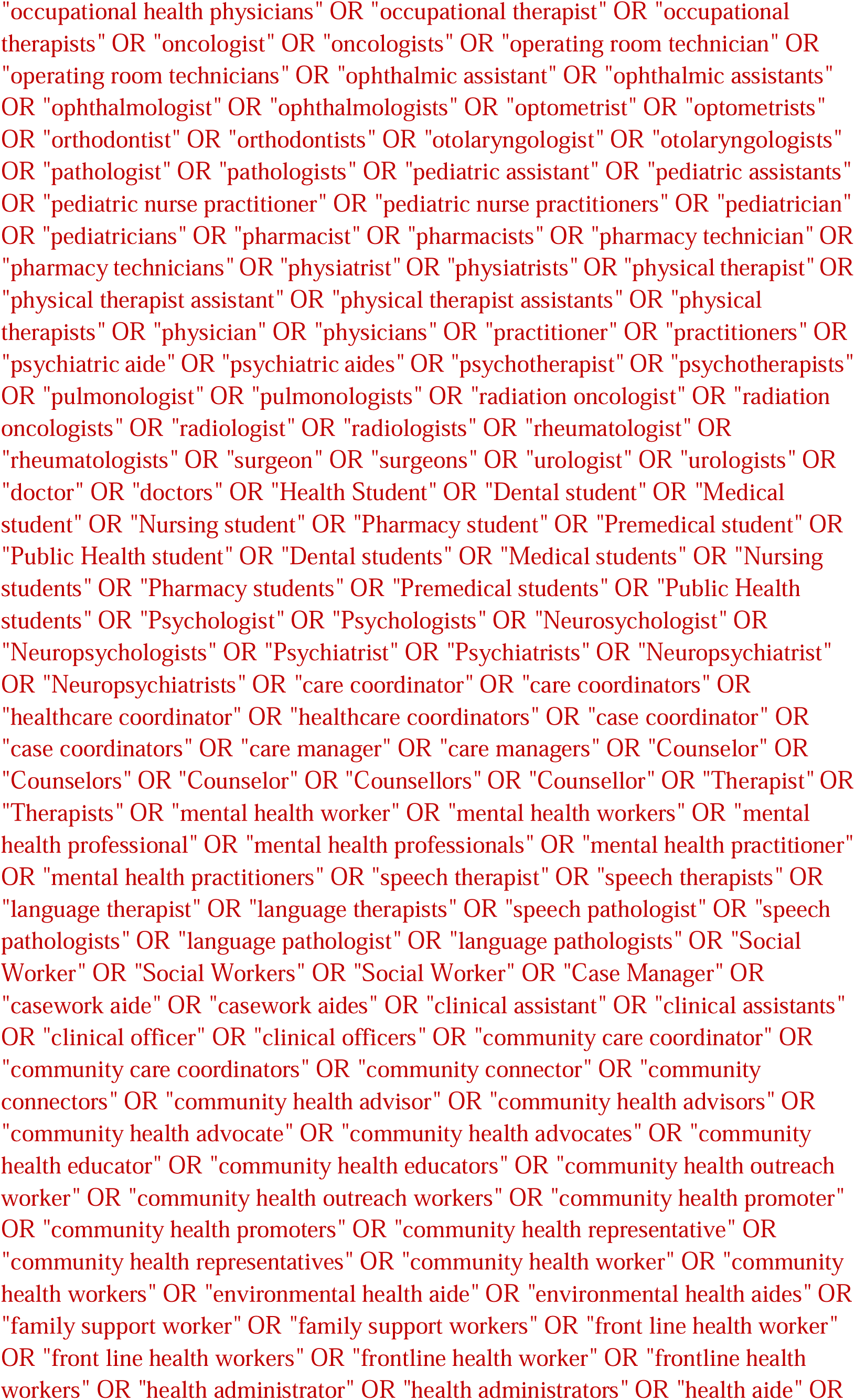
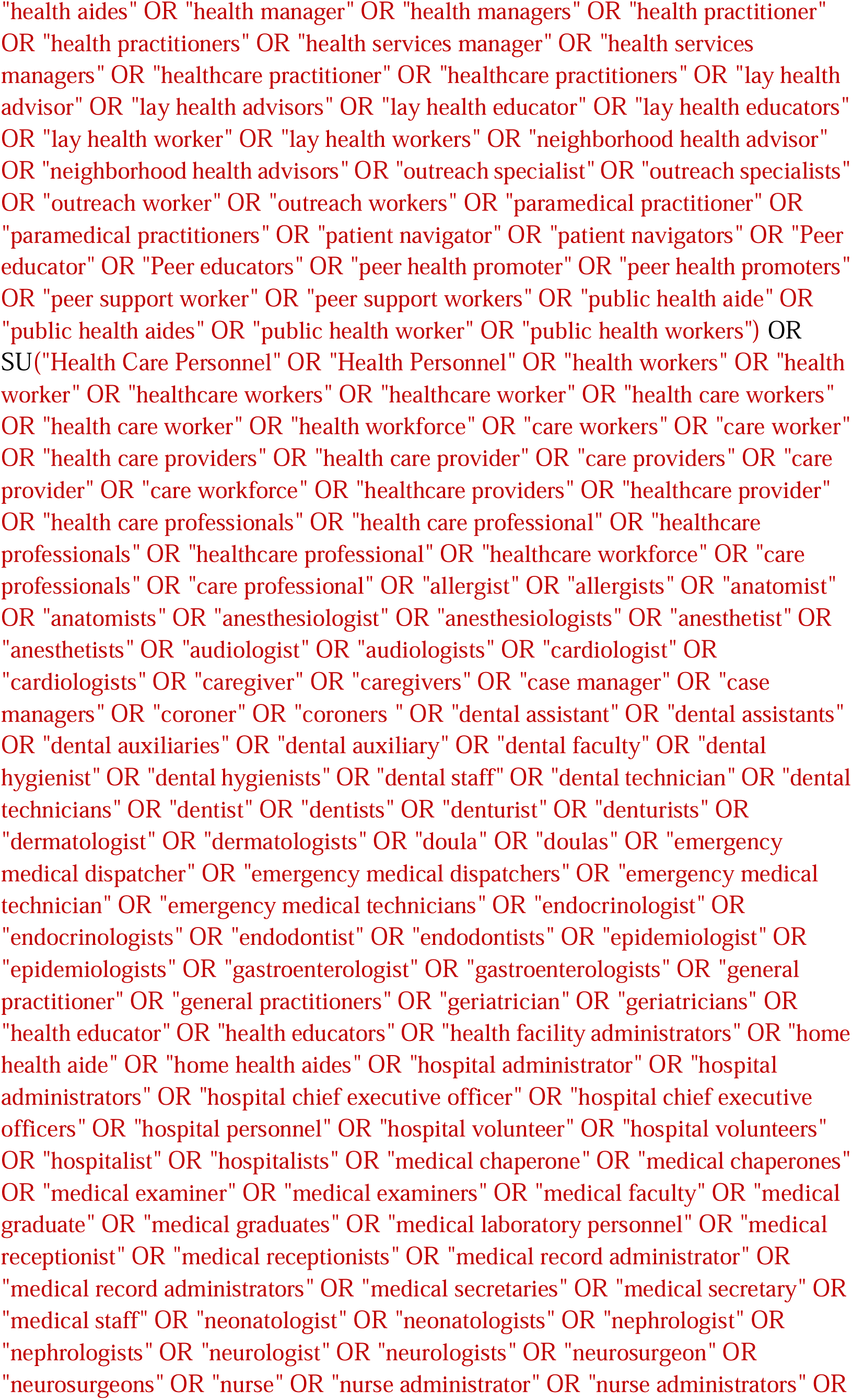
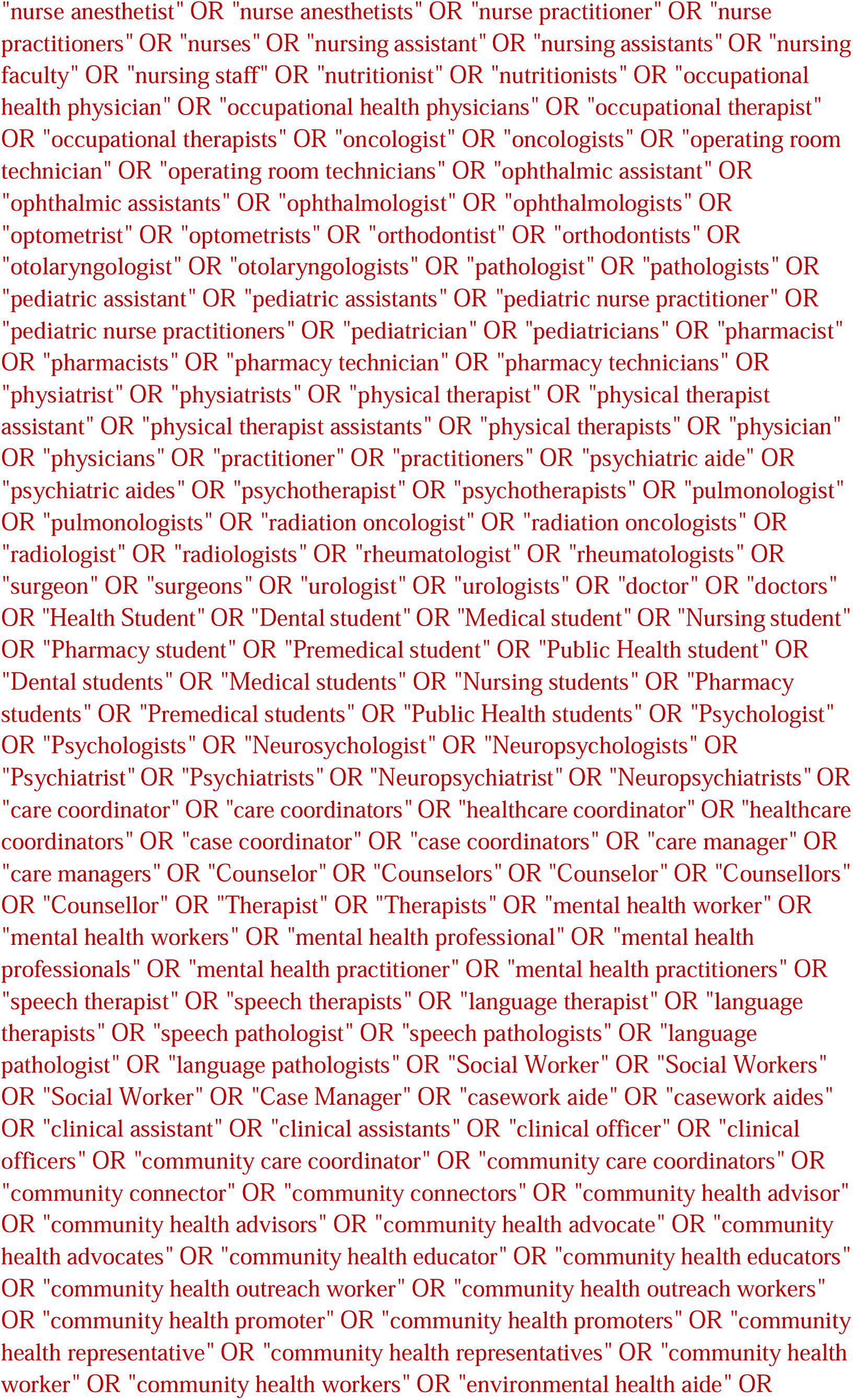
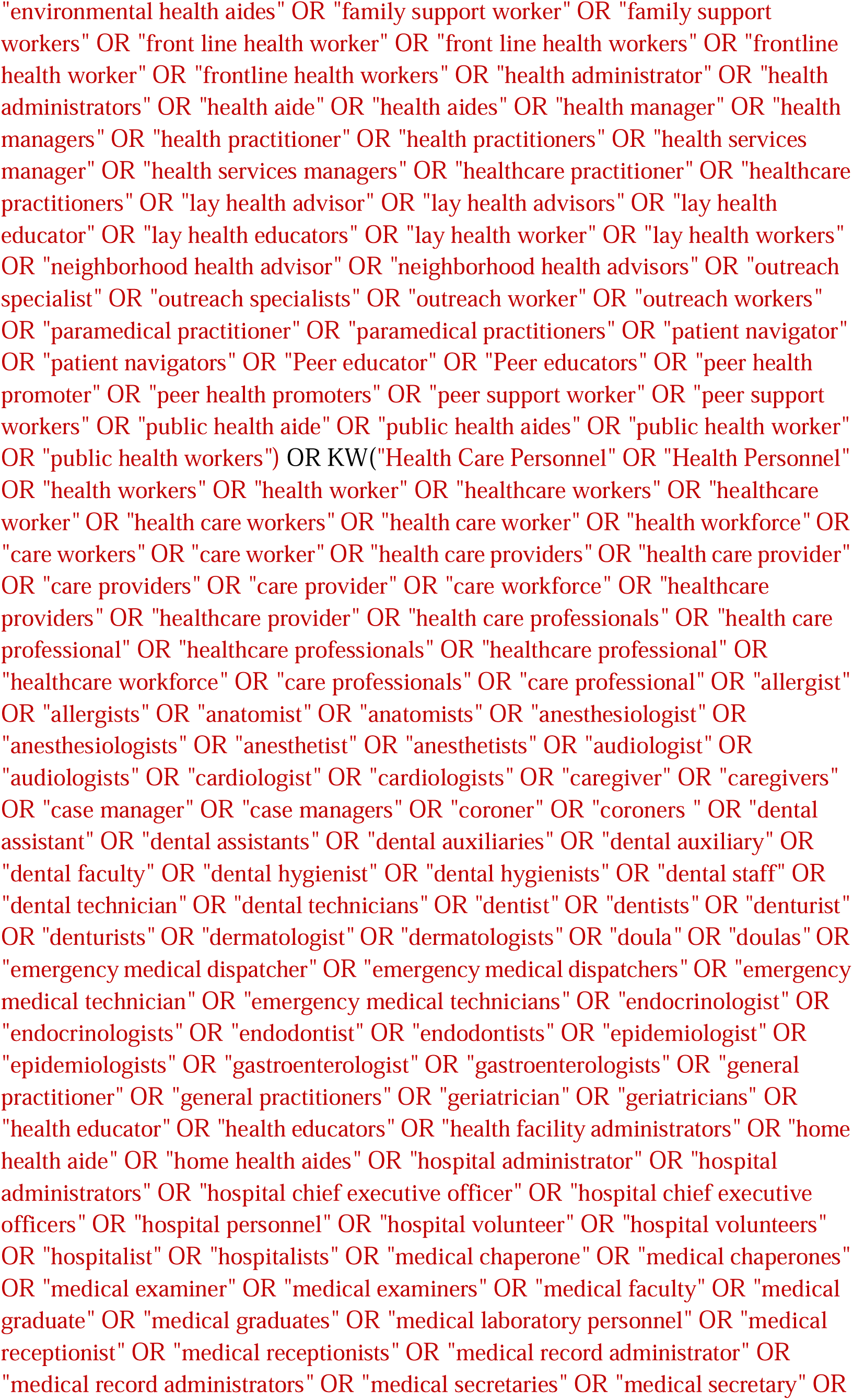
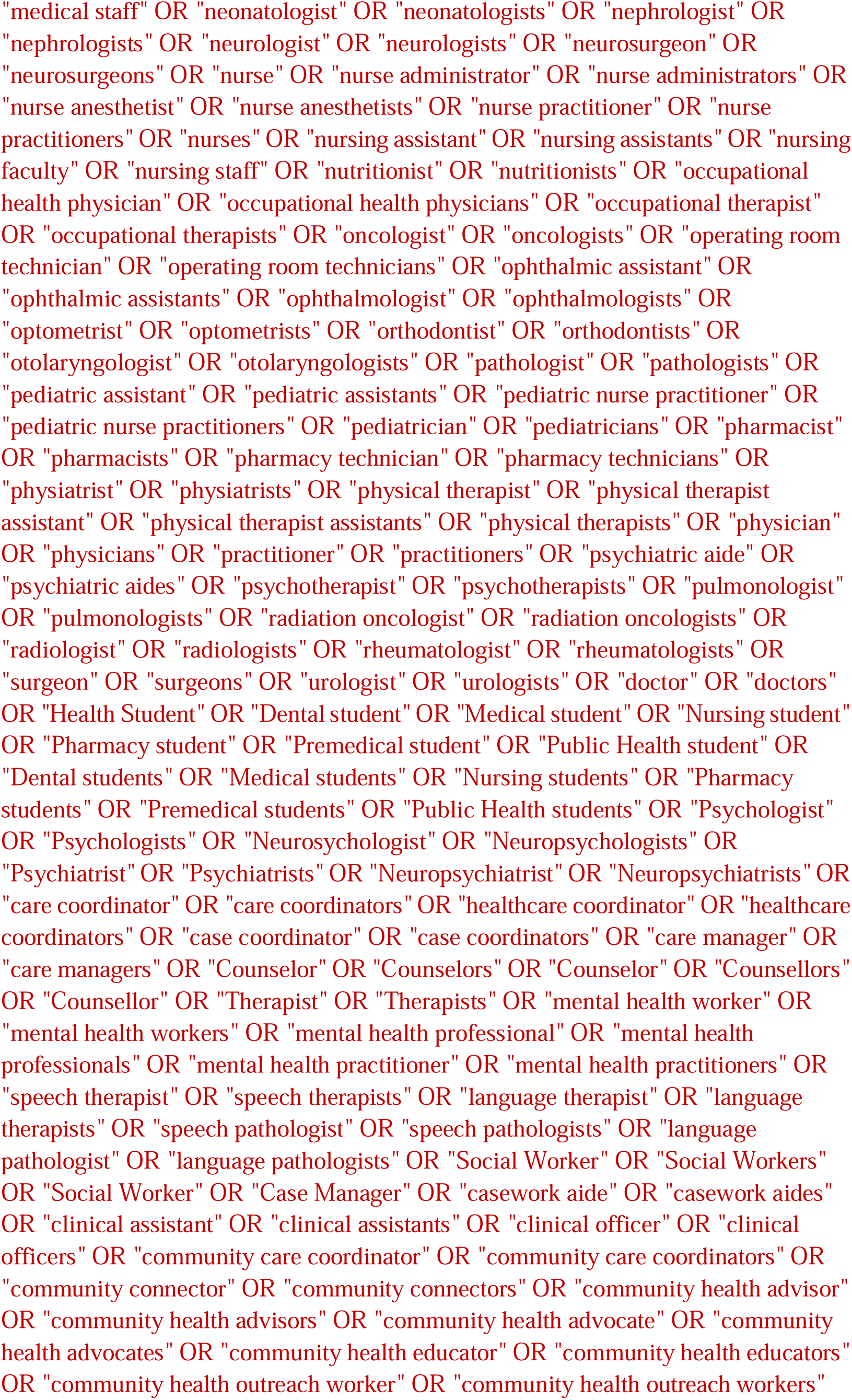
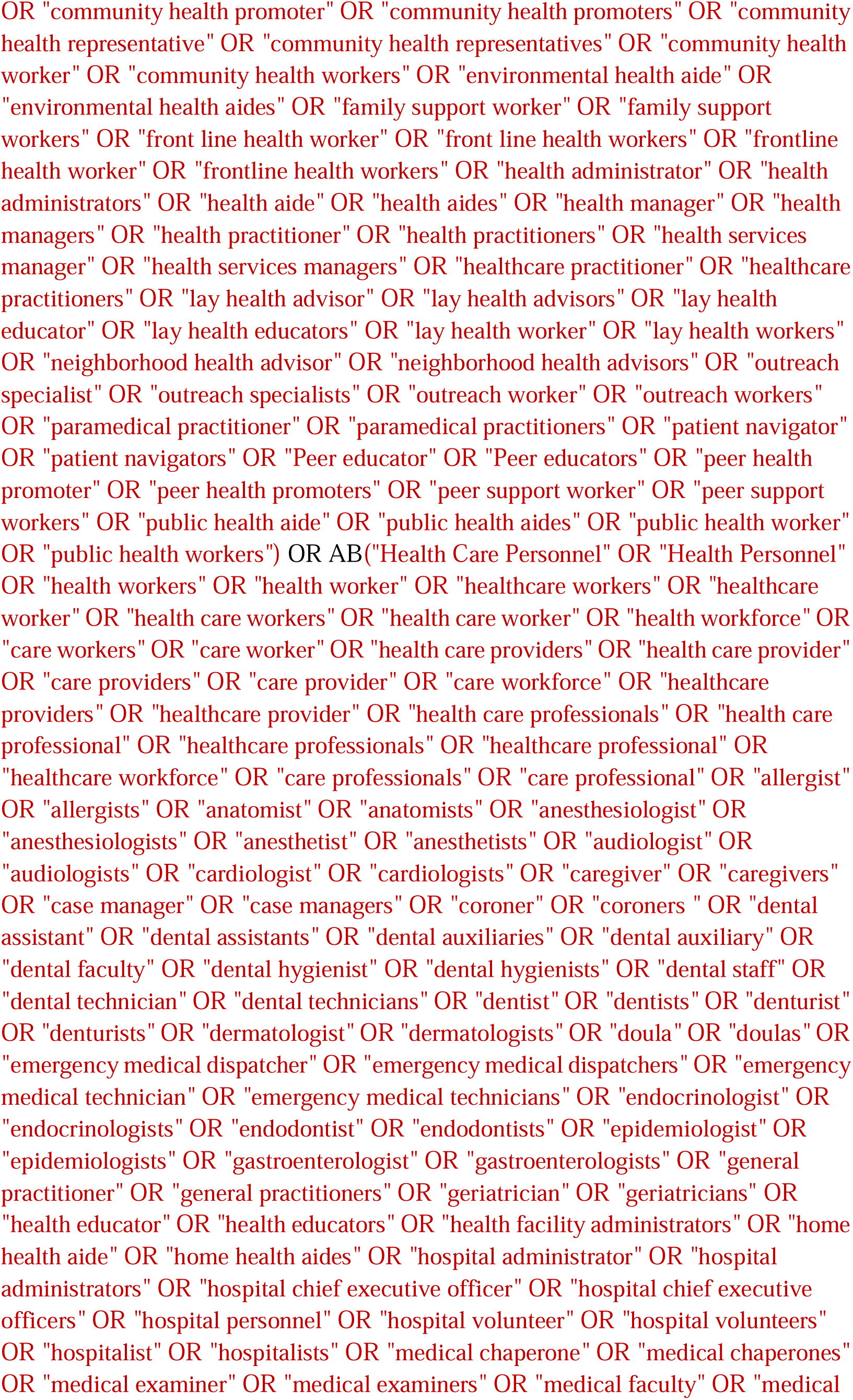
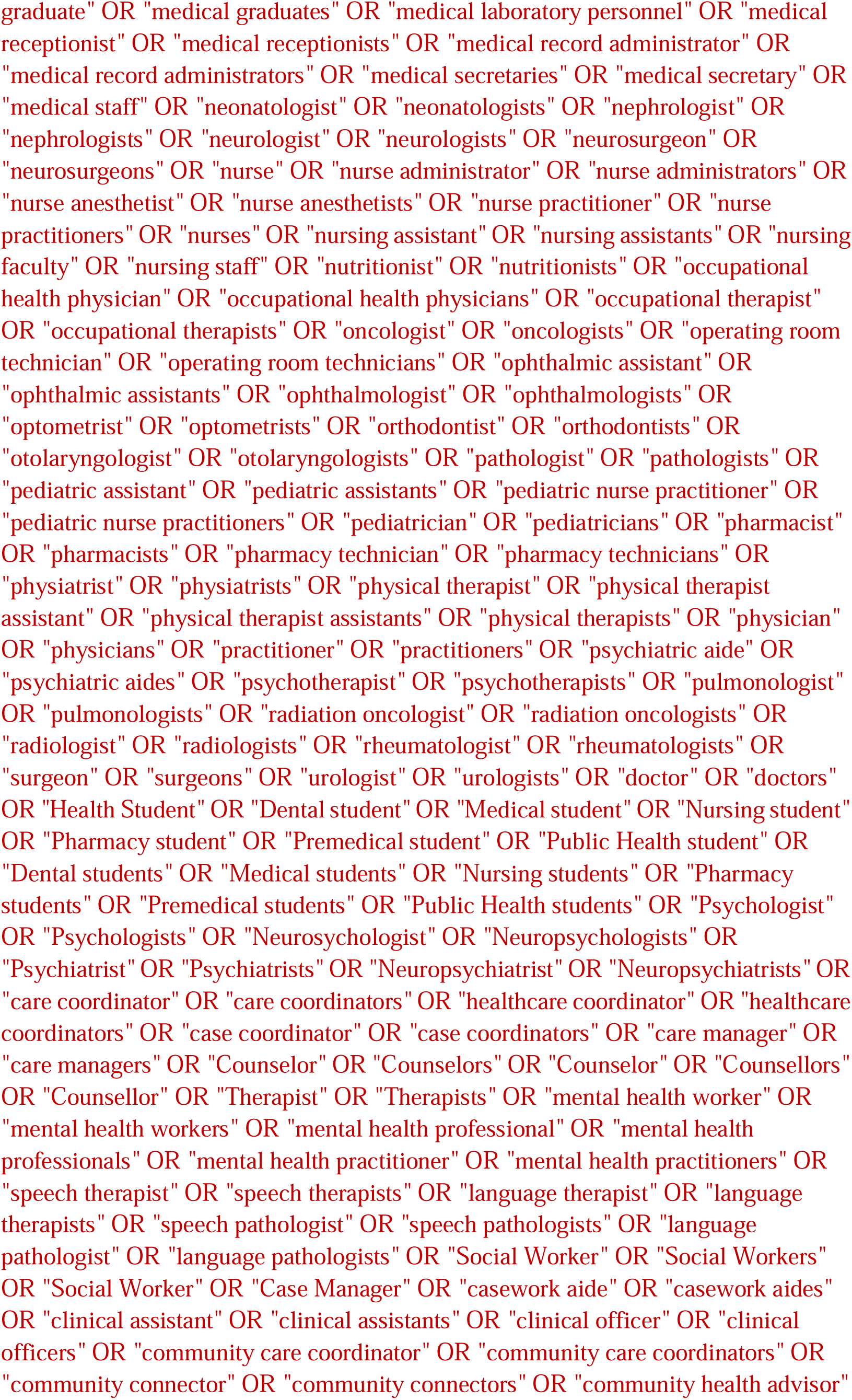
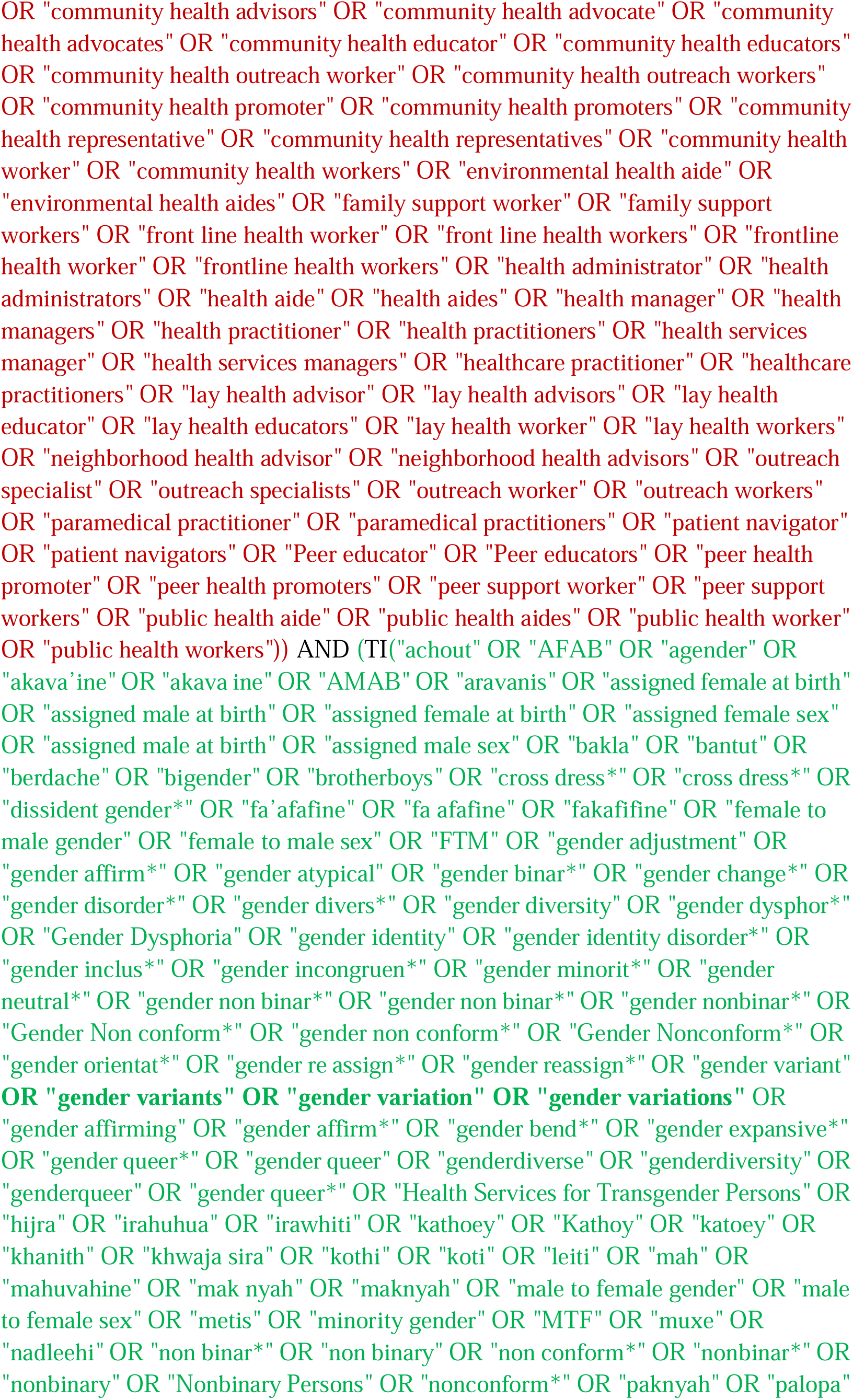
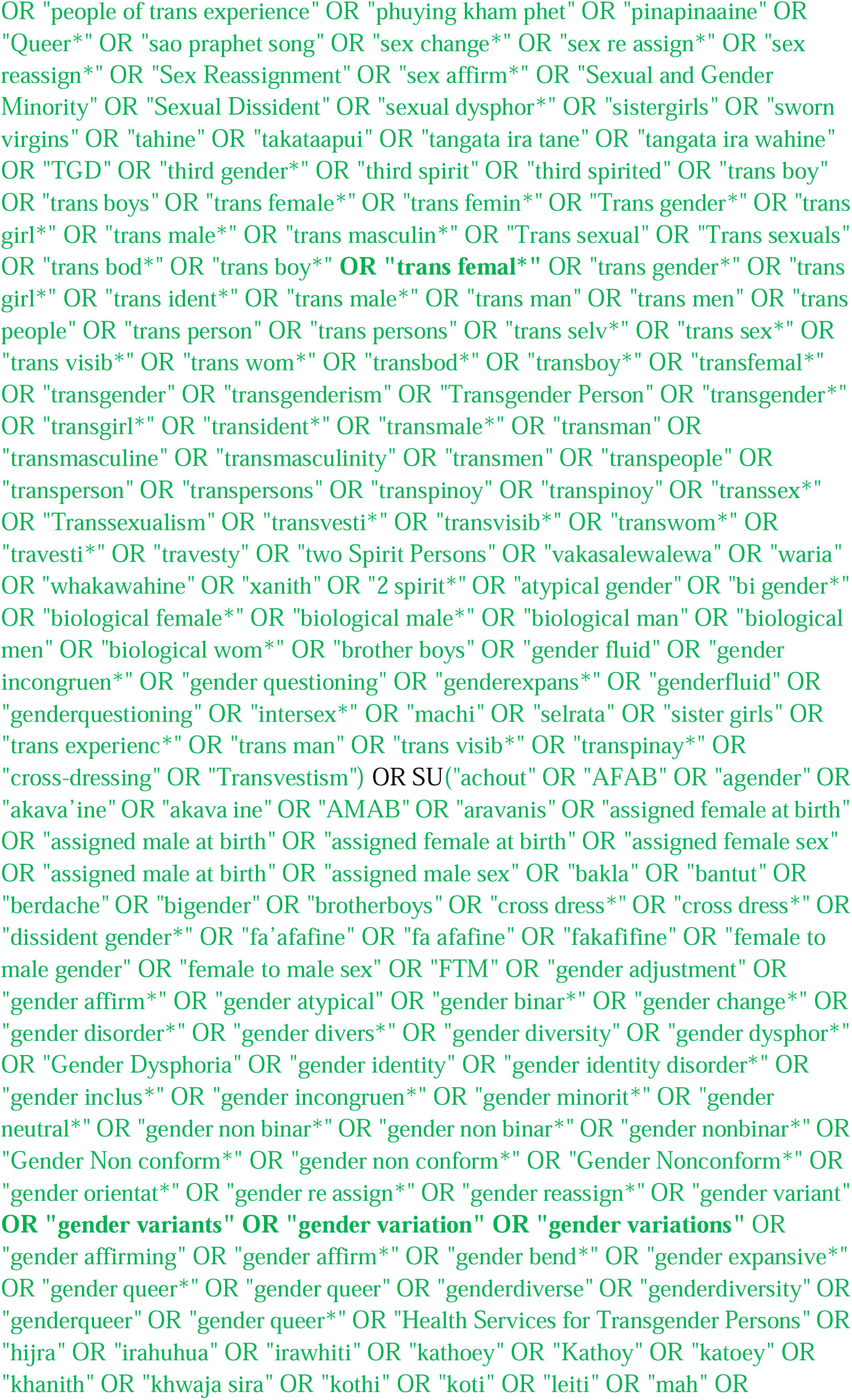
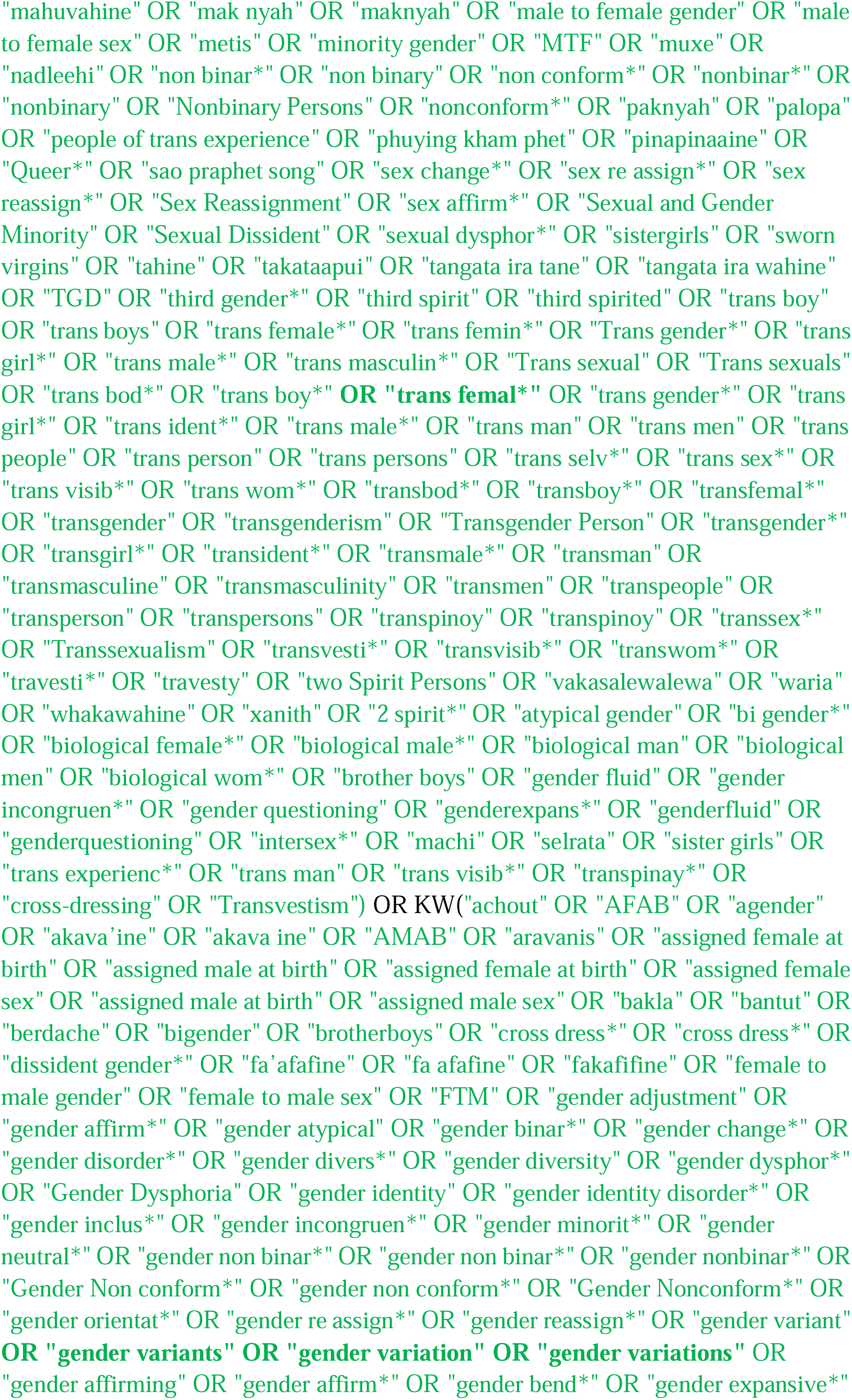
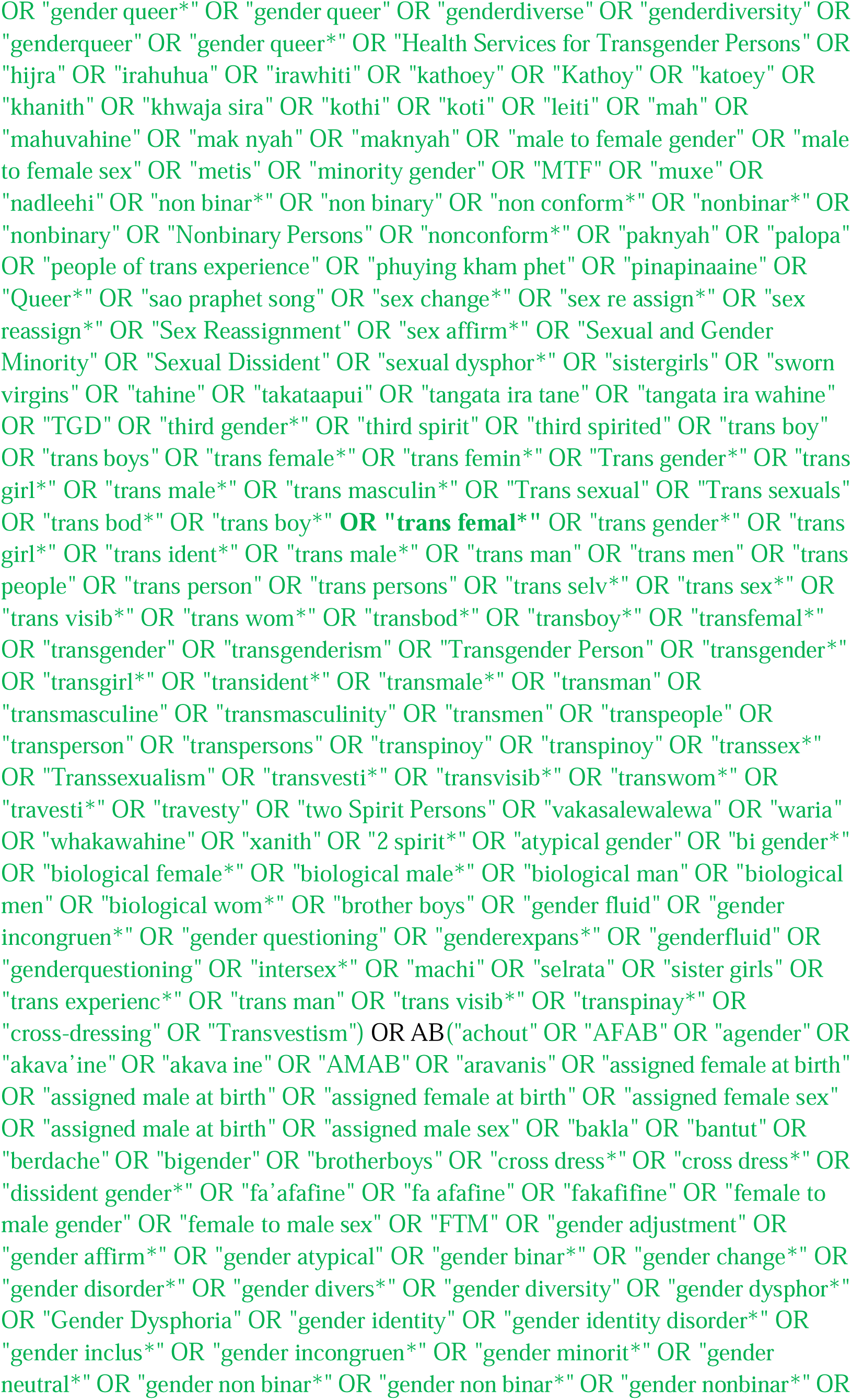
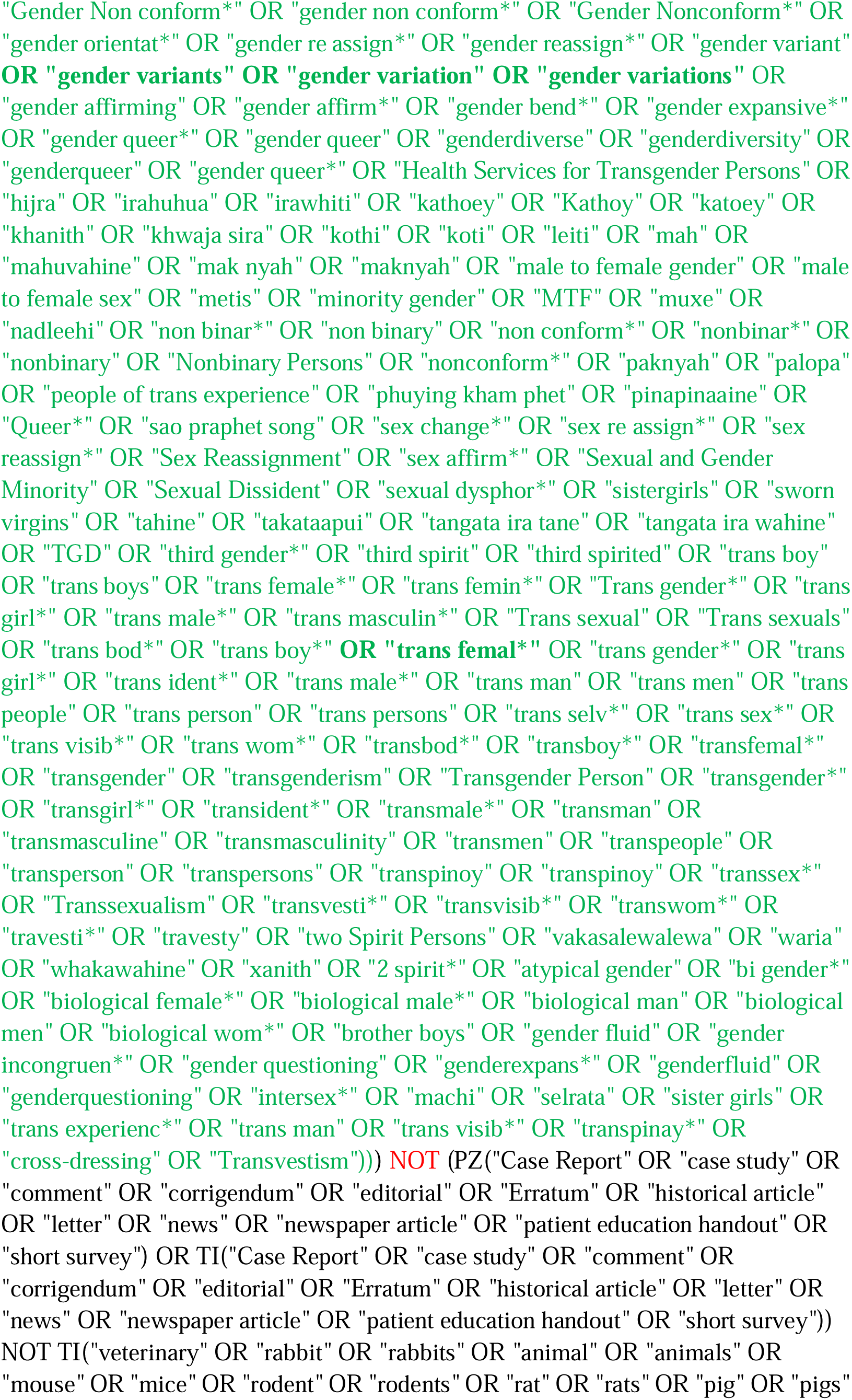
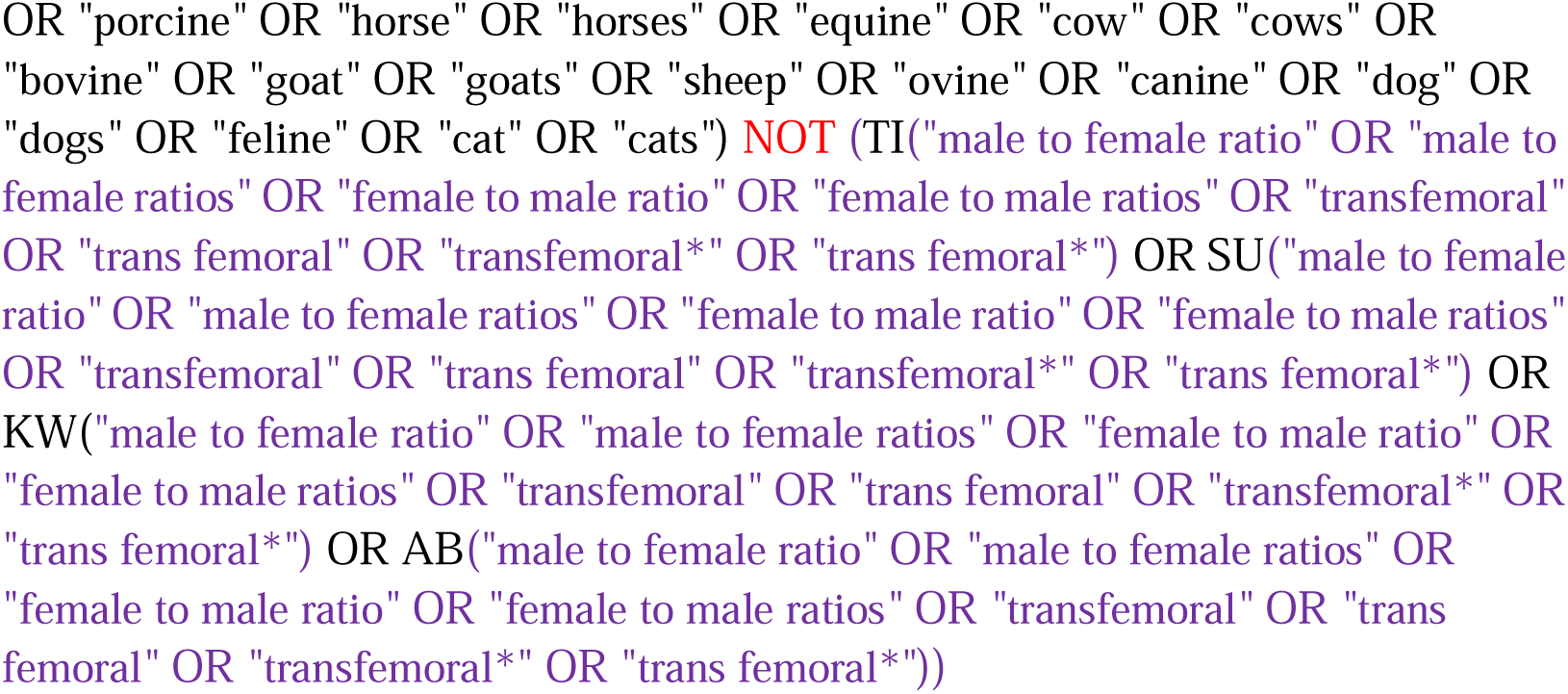

#### LILACS

https://lilacs.bvsalud.org/en/

Simplified version - three search strings

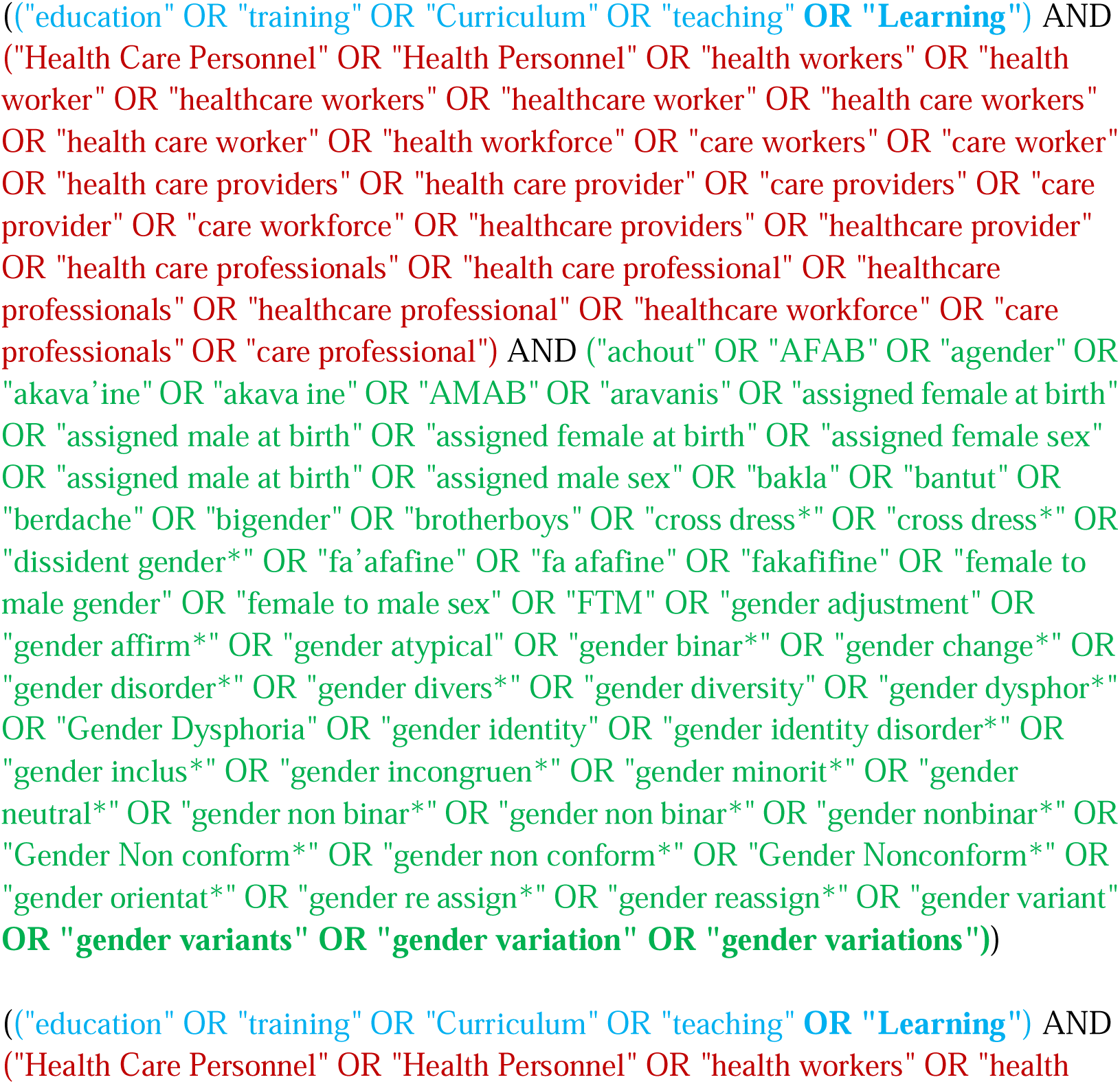
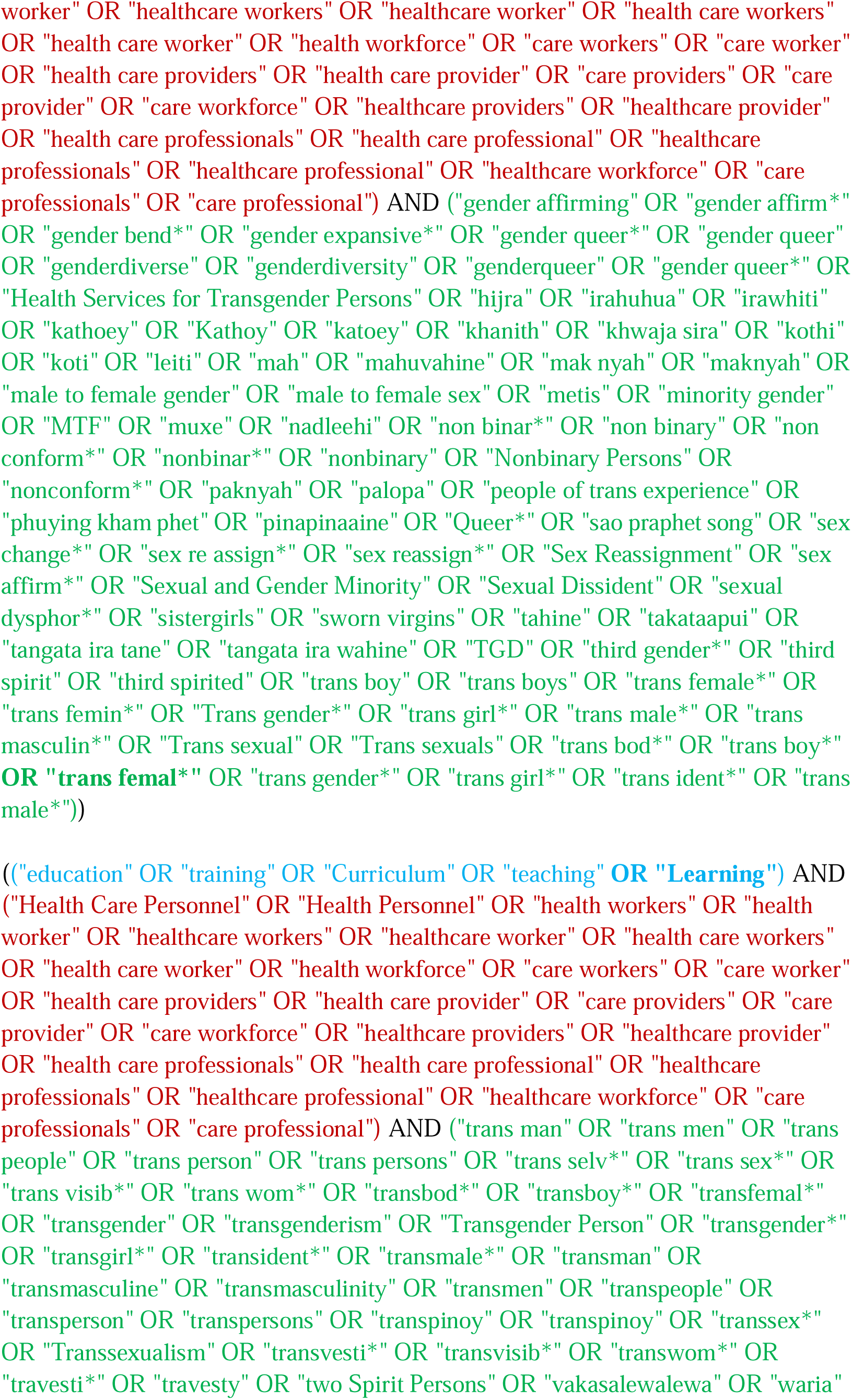
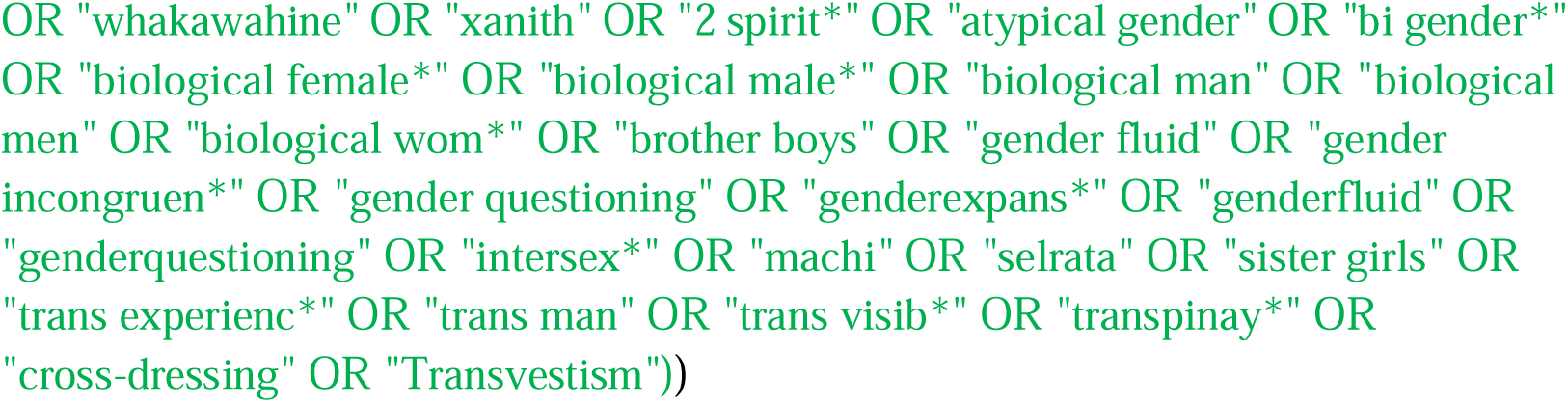

### Grey literature: conference or meeting abstracts search strategy

**Healthcare question:** What are effective education and or training interventions for health workers to provide gender-affirming and or inclusive care for trans and gender-diverse people

WHO education for health workers in relation to trans and gender-diverse people

**Search date**: 22/12/2023

**Search results summary**: 301 references, sourced from:

- Embase (OVID): 309 - 298 unique
- Web of Science: 0
- Cochrane CENTRAL: 24 - 3 unique

**Other notes**

Three subject components:

- Health Workers
- Education
- Trans and gender-diverse people

Limit to RCTs

**Search strategy**

#### Embase

http://ovidsp.ovid.com/ovidweb.cgi?T=JS&PAGE=main&MODE=ovid&D=oemezd

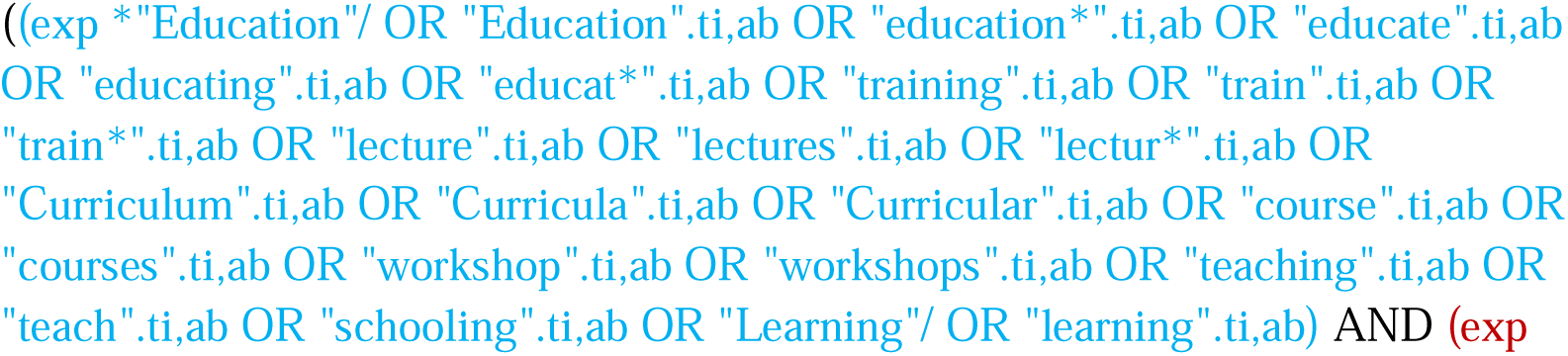
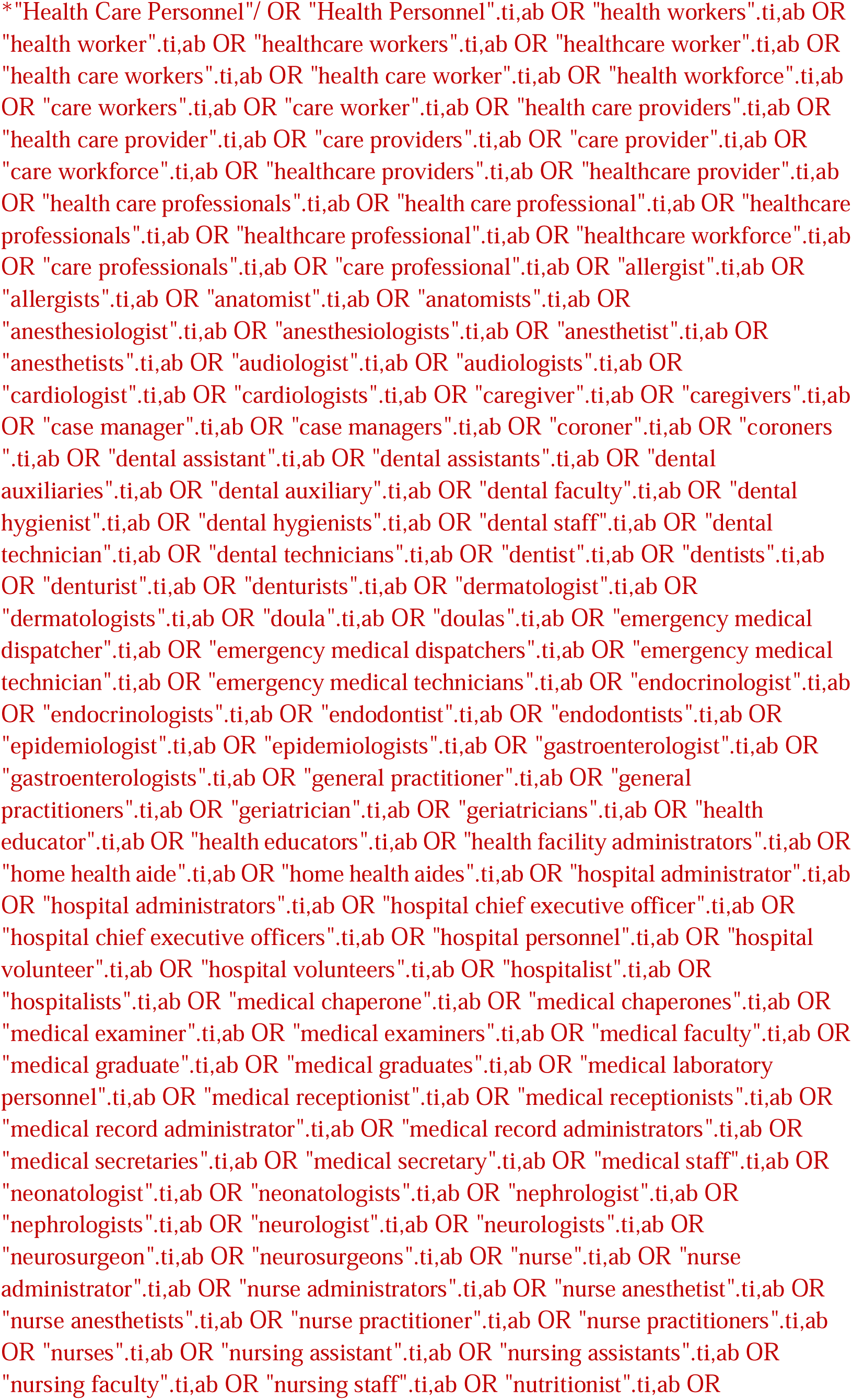
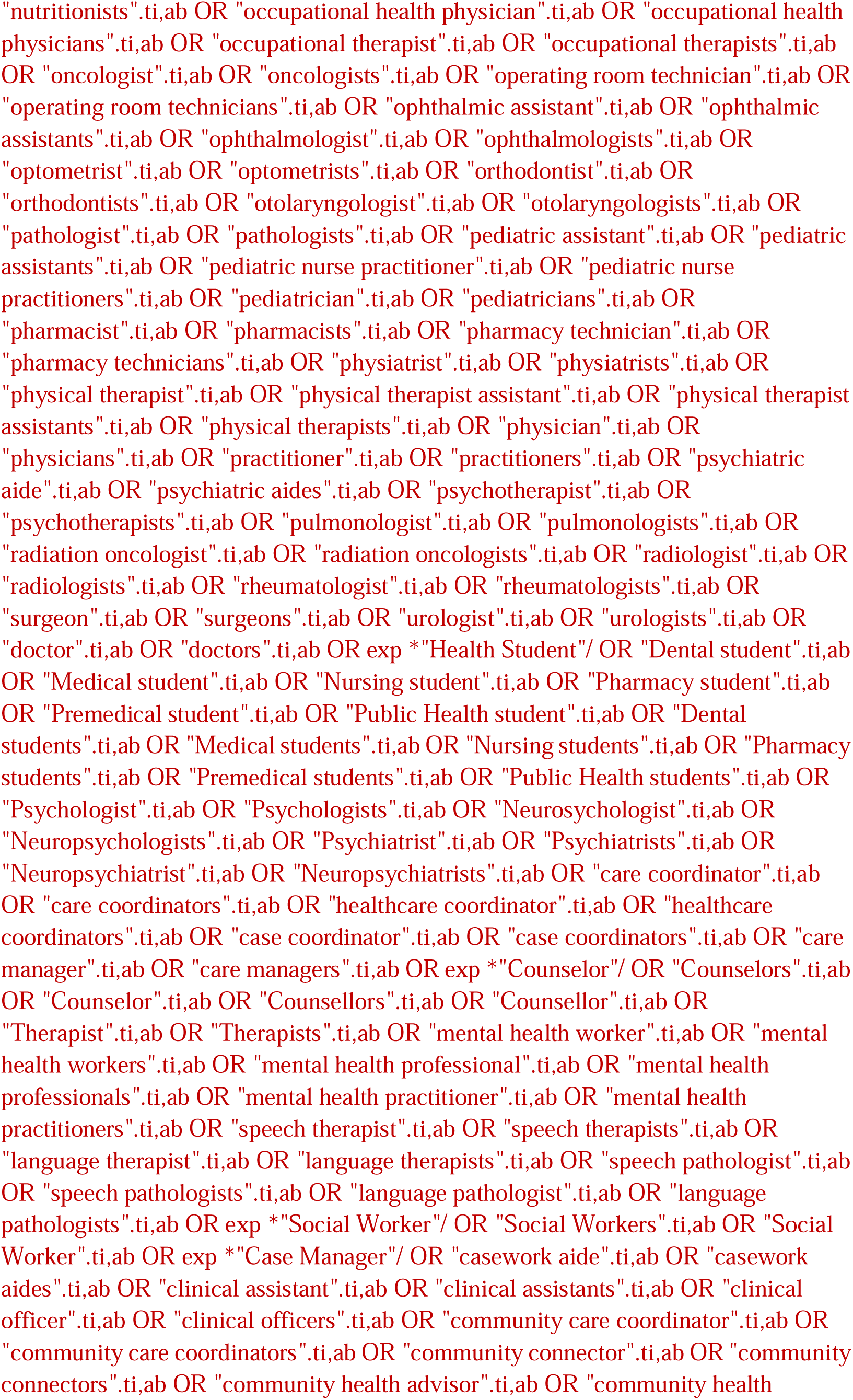
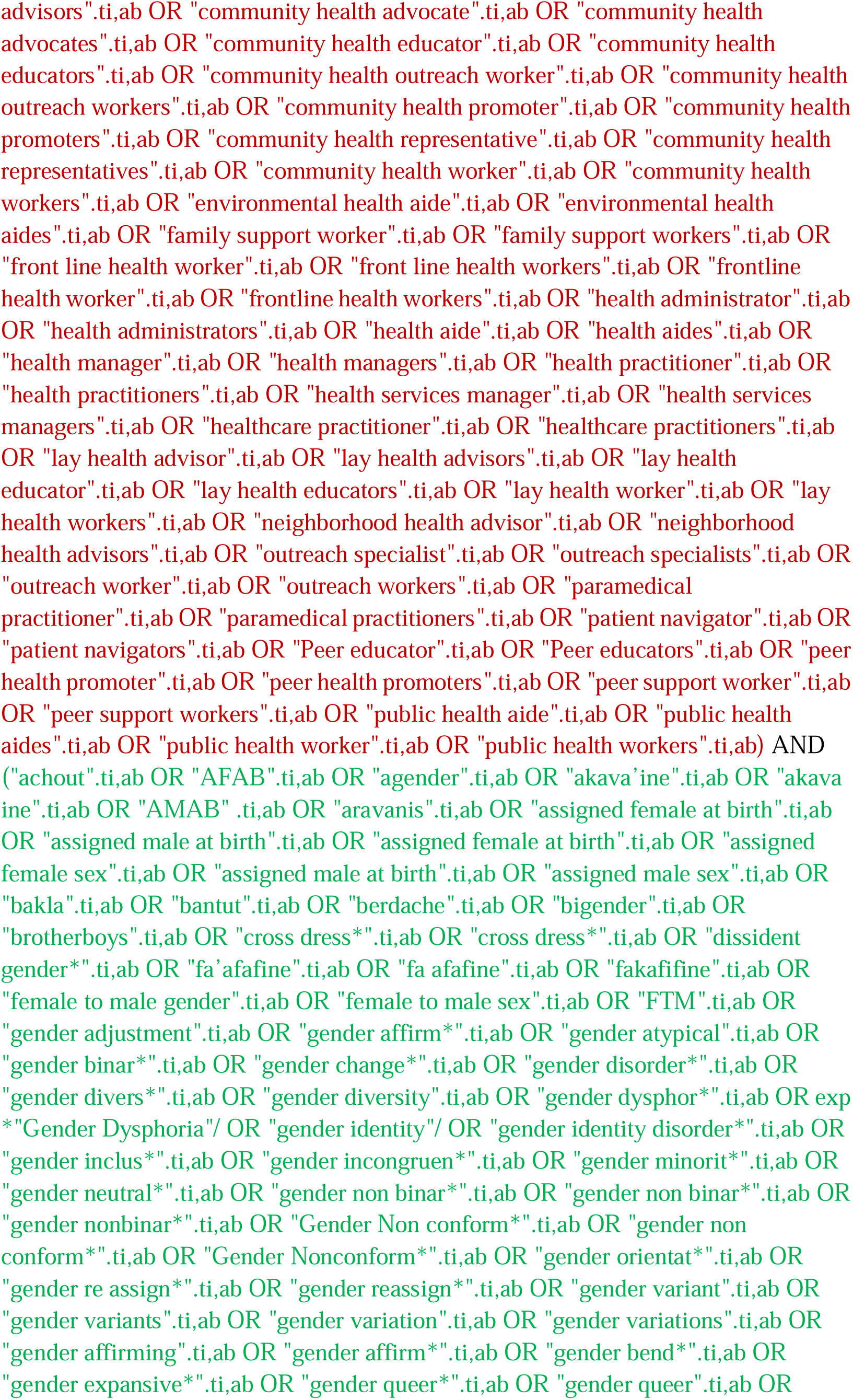
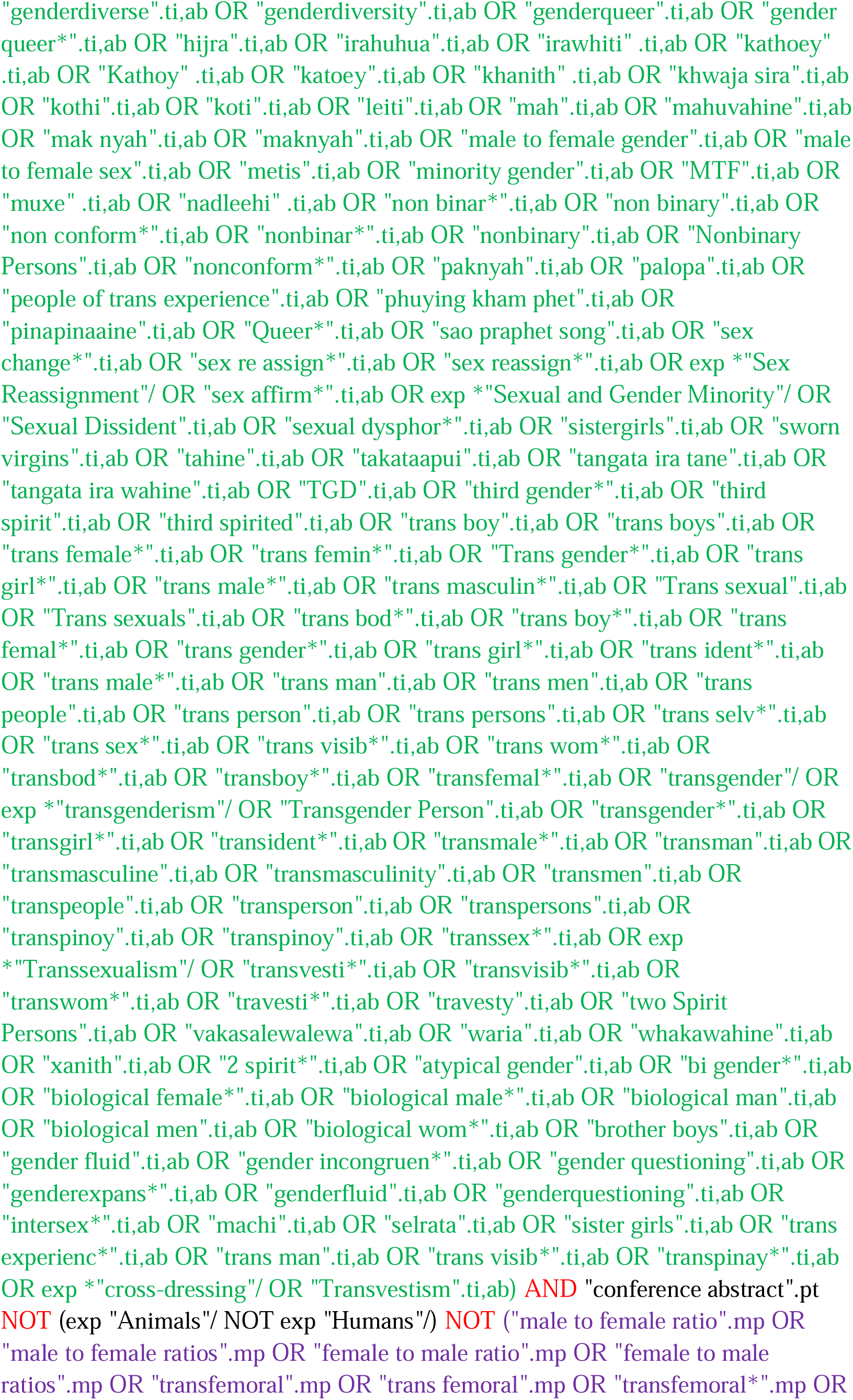
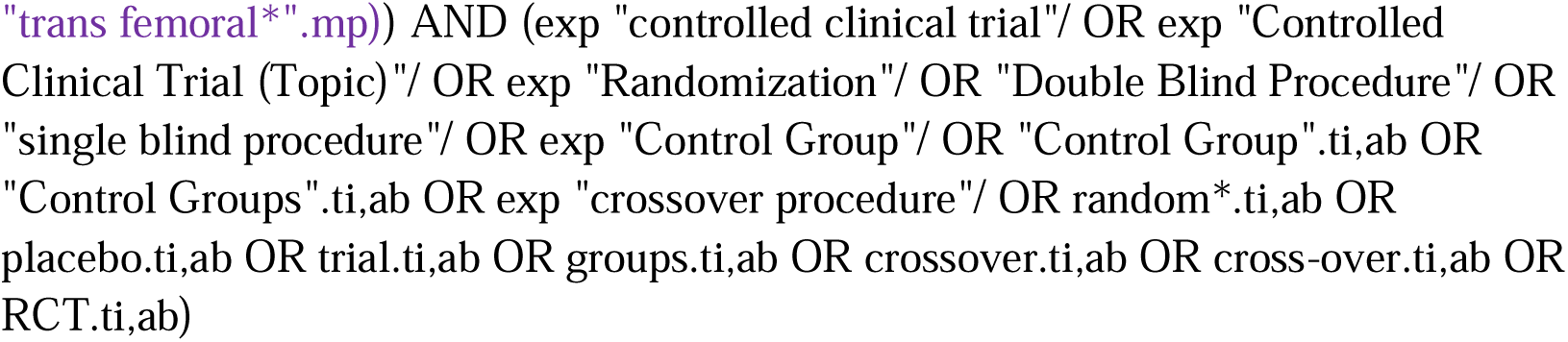

#### Web of Science

http://isiknowledge.com/wos

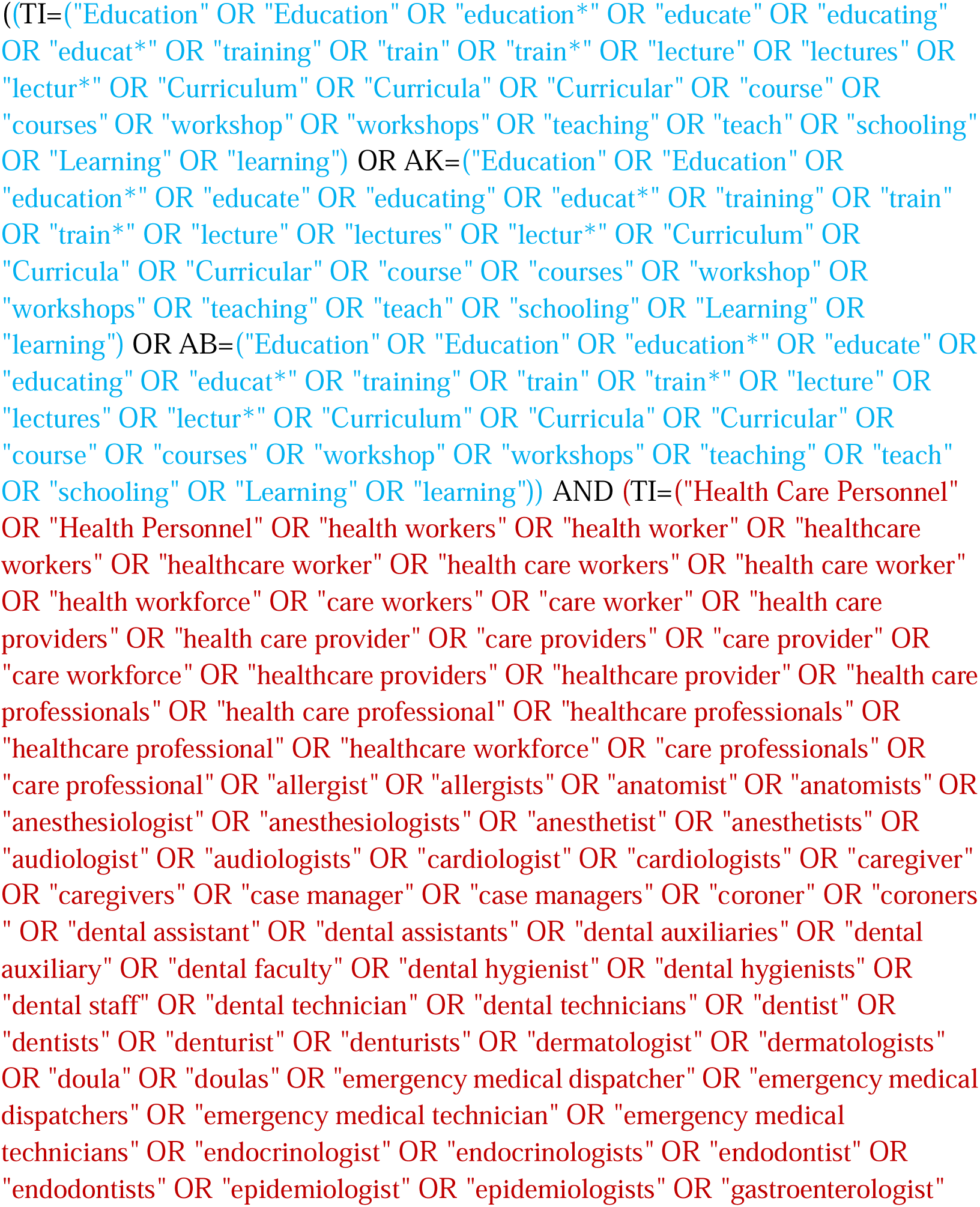
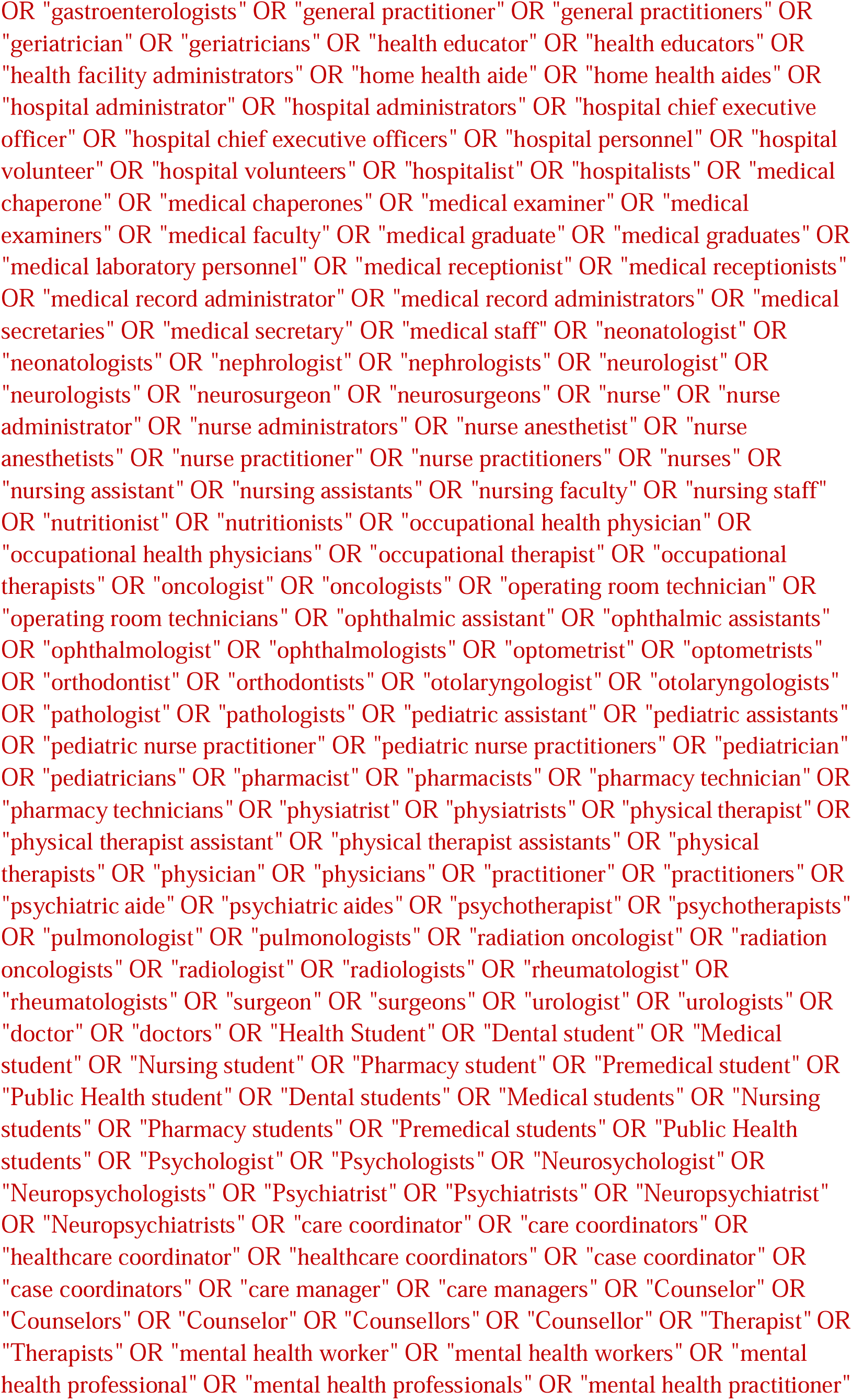
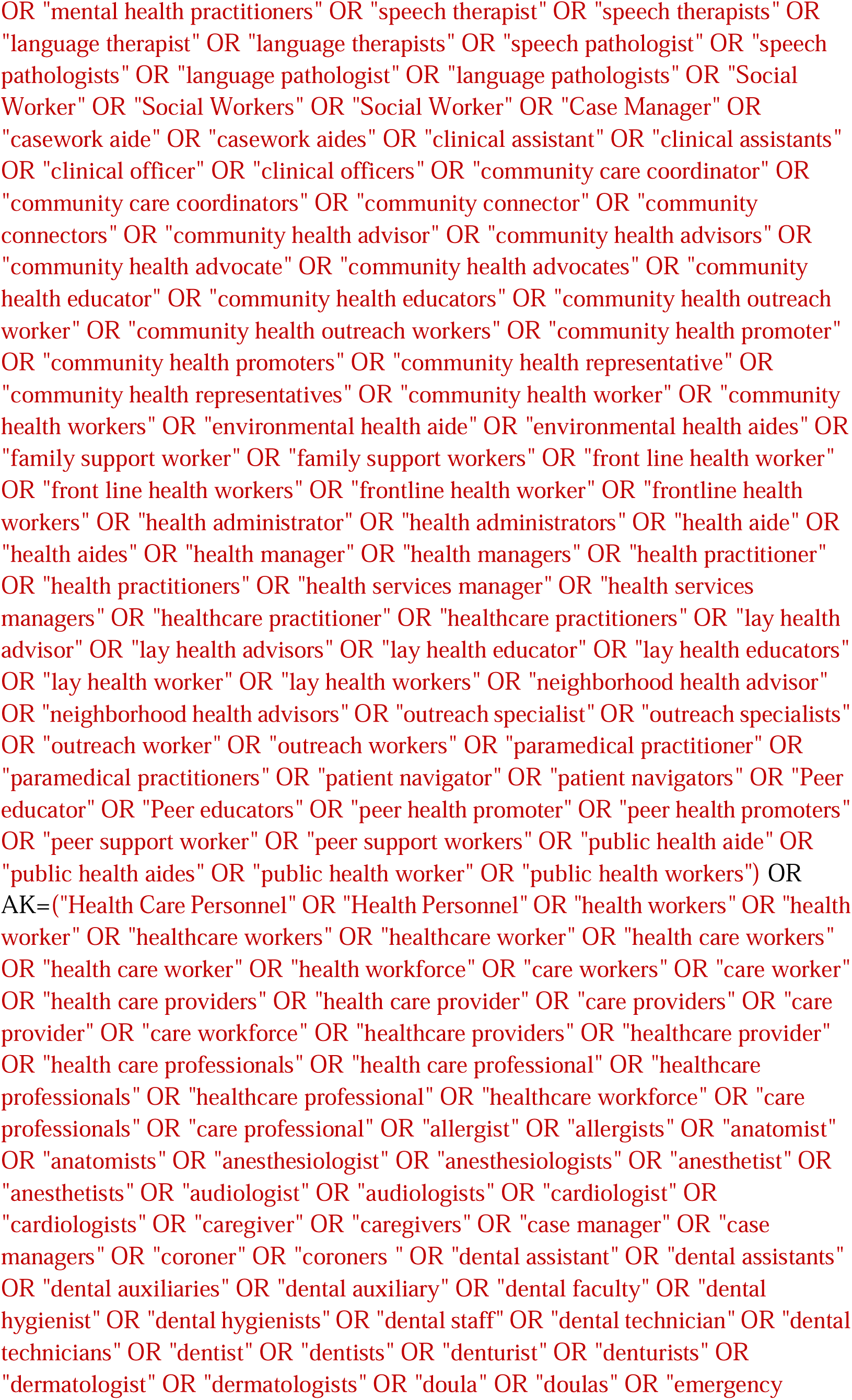
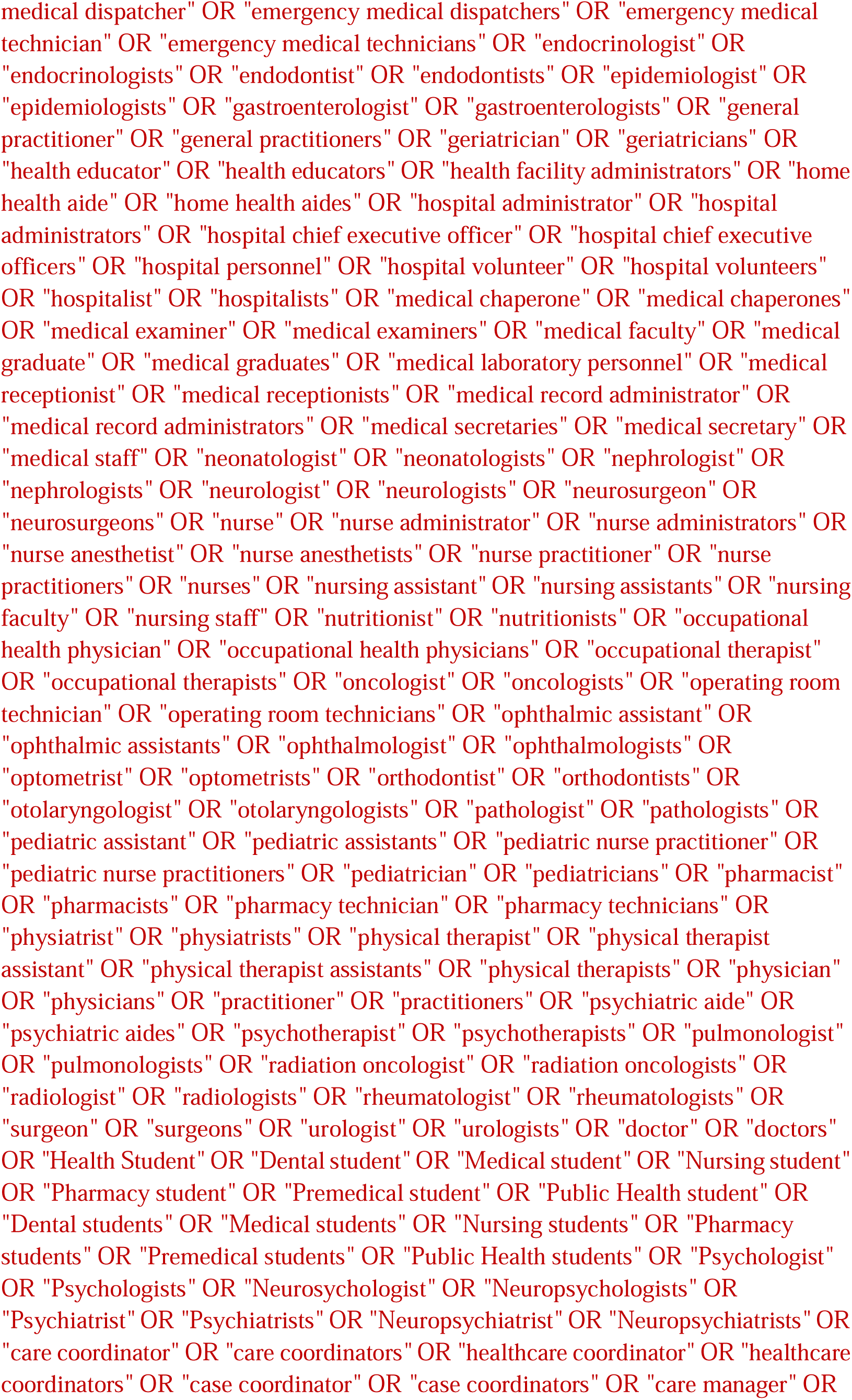
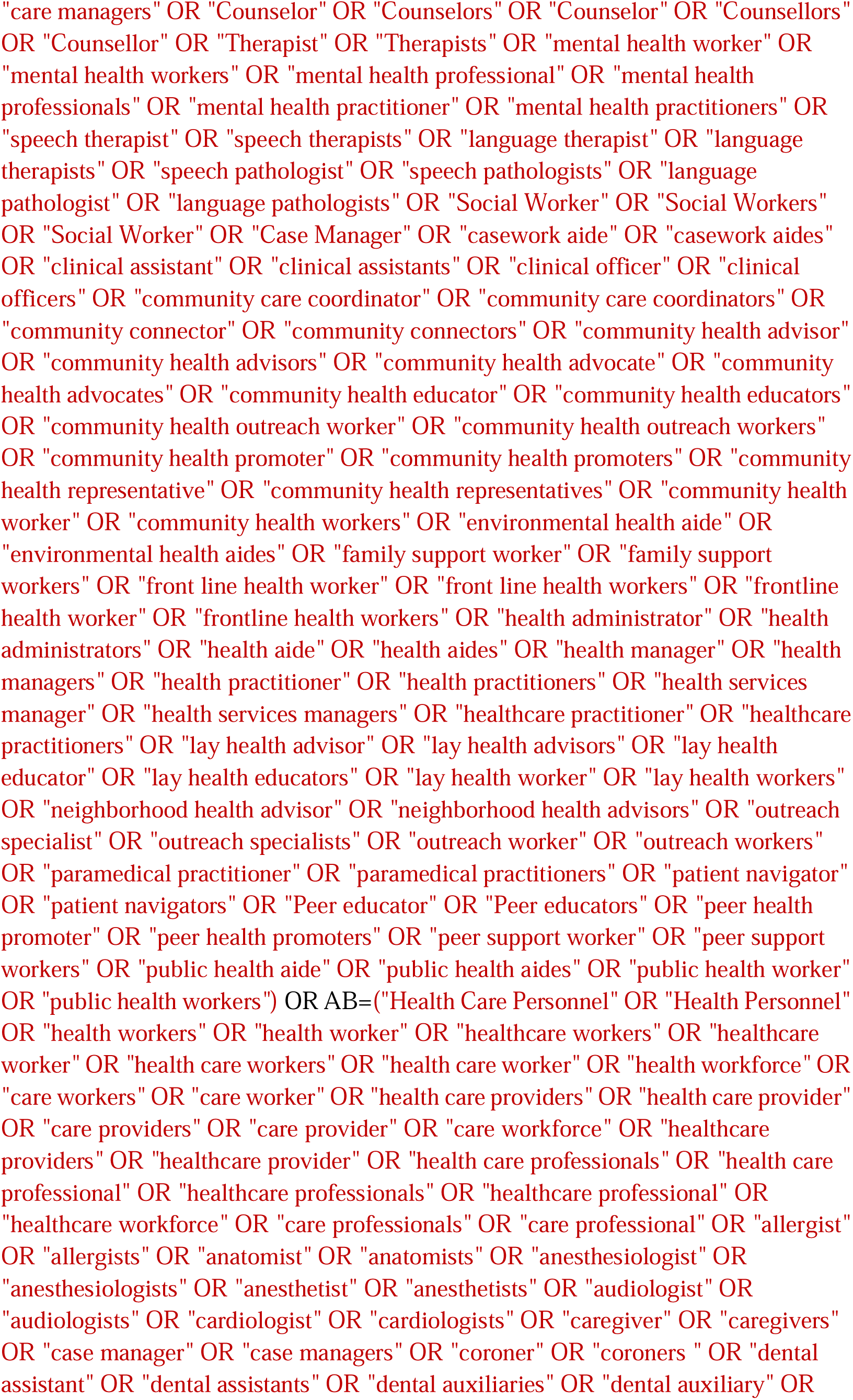
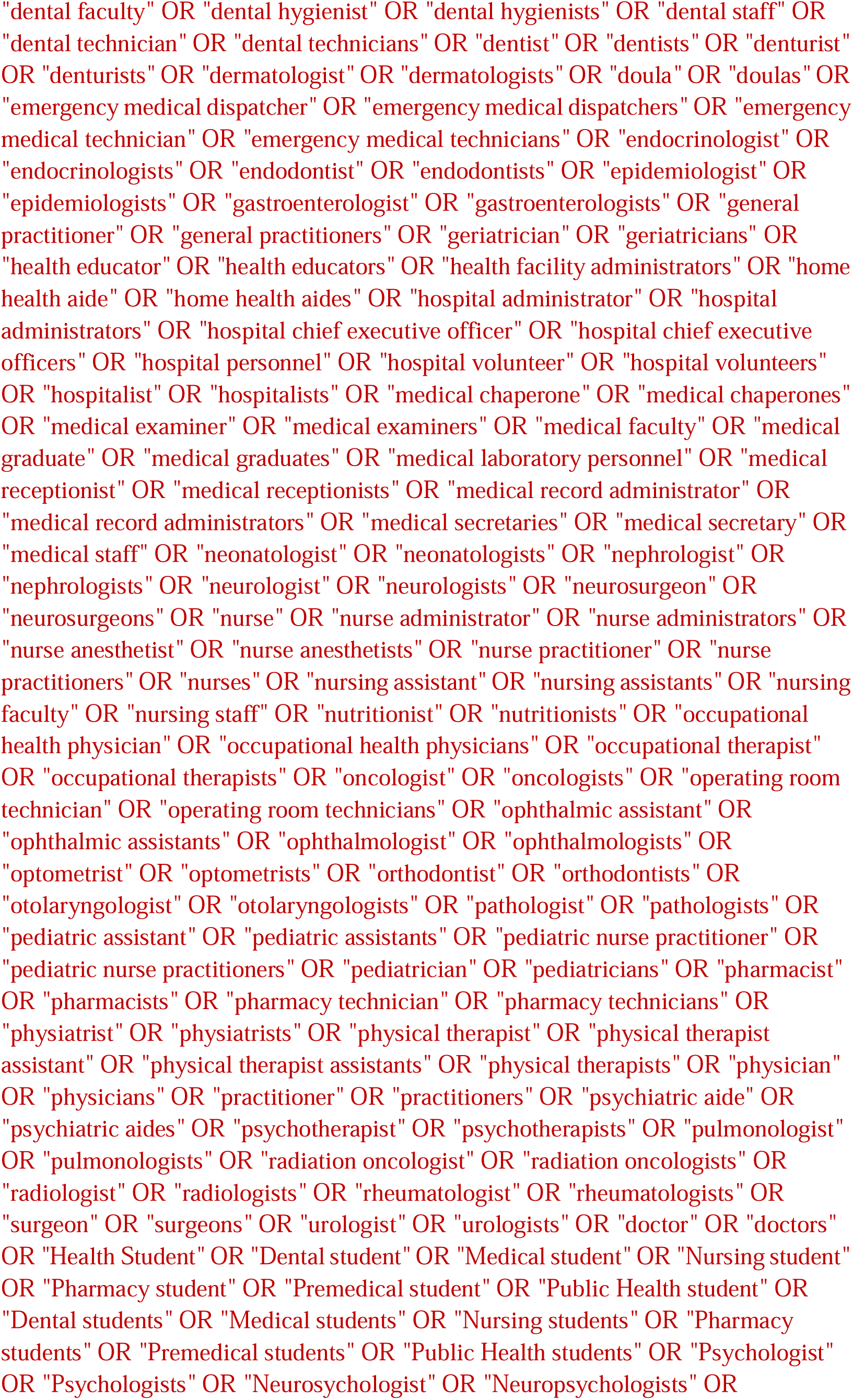
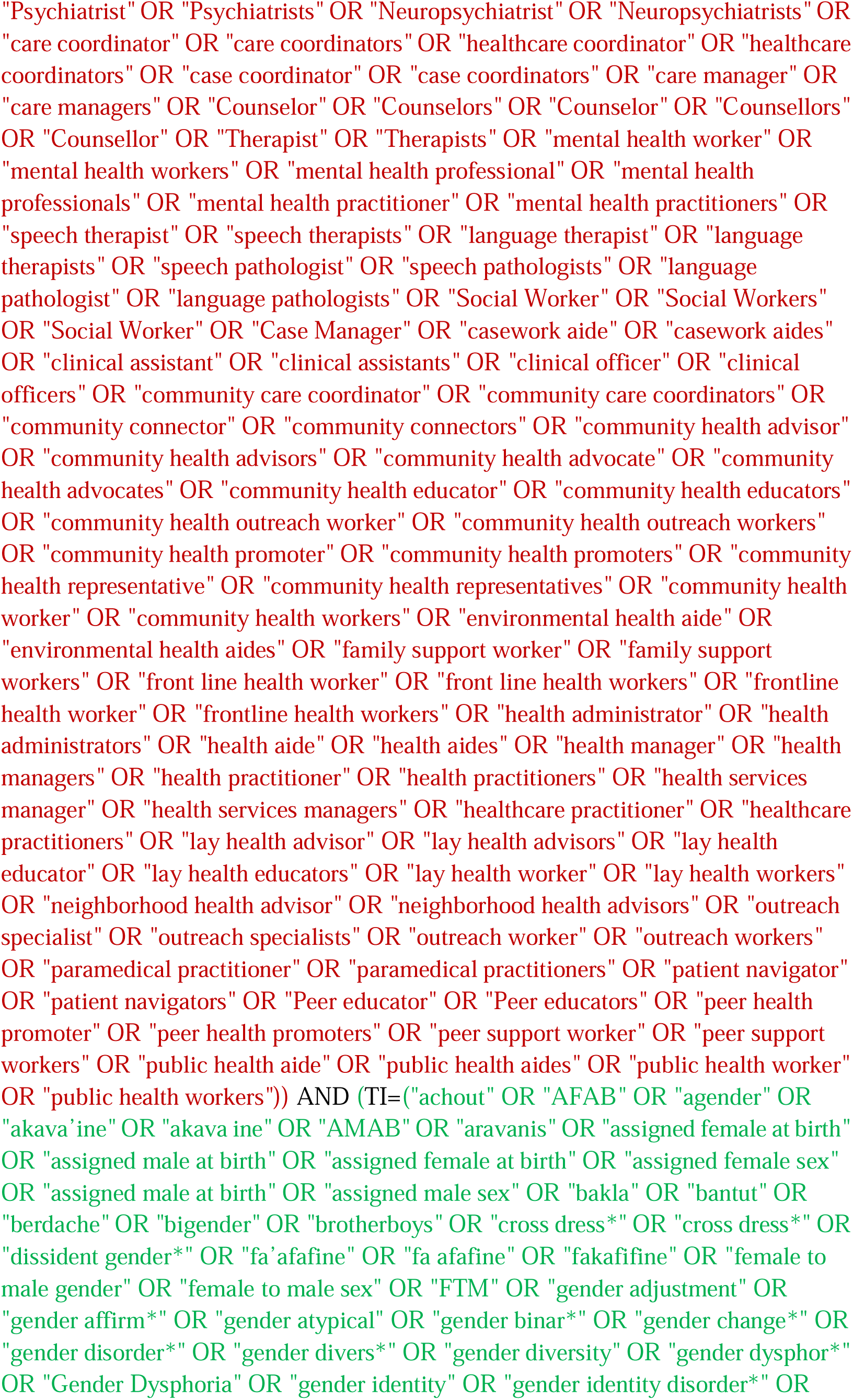
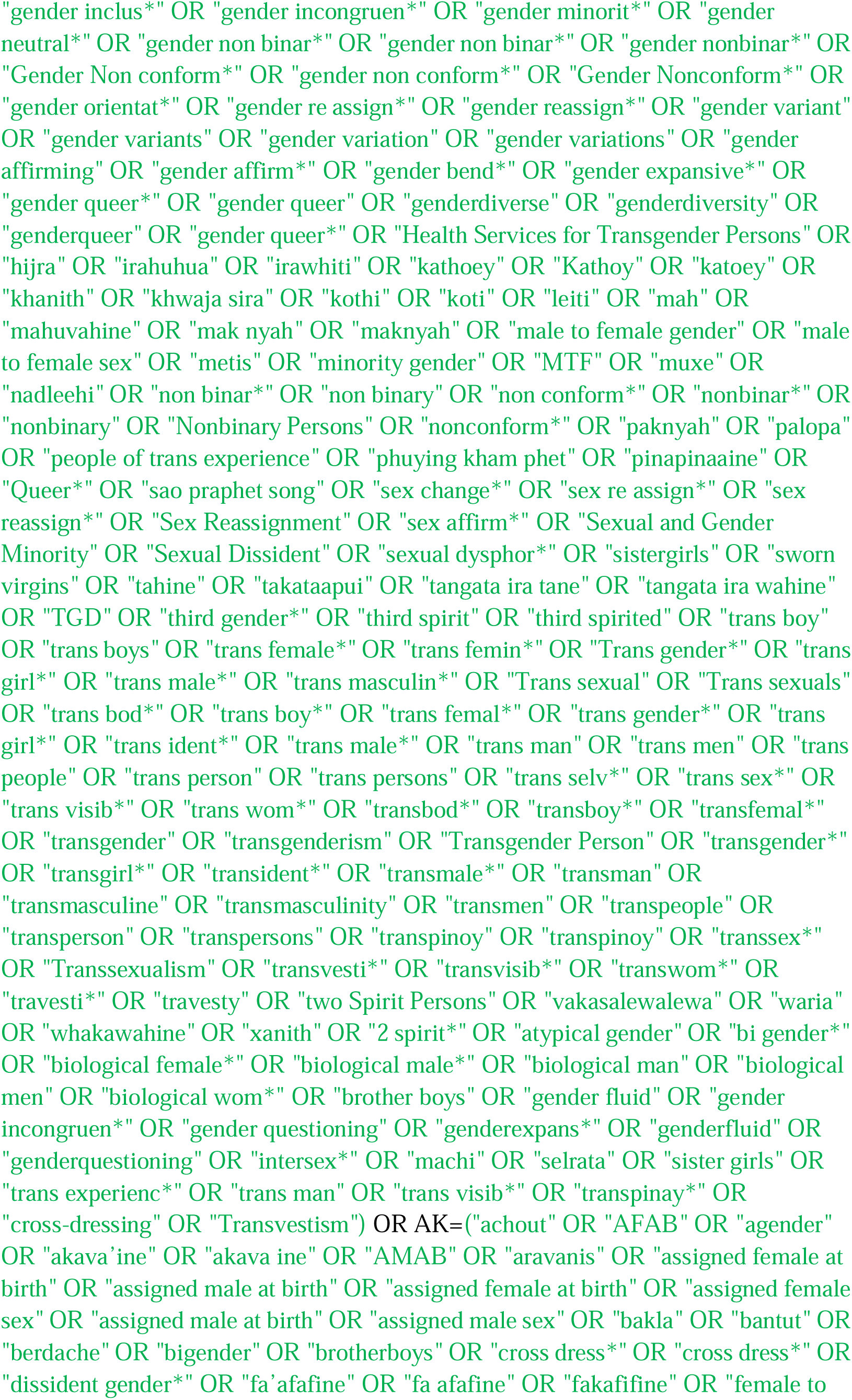
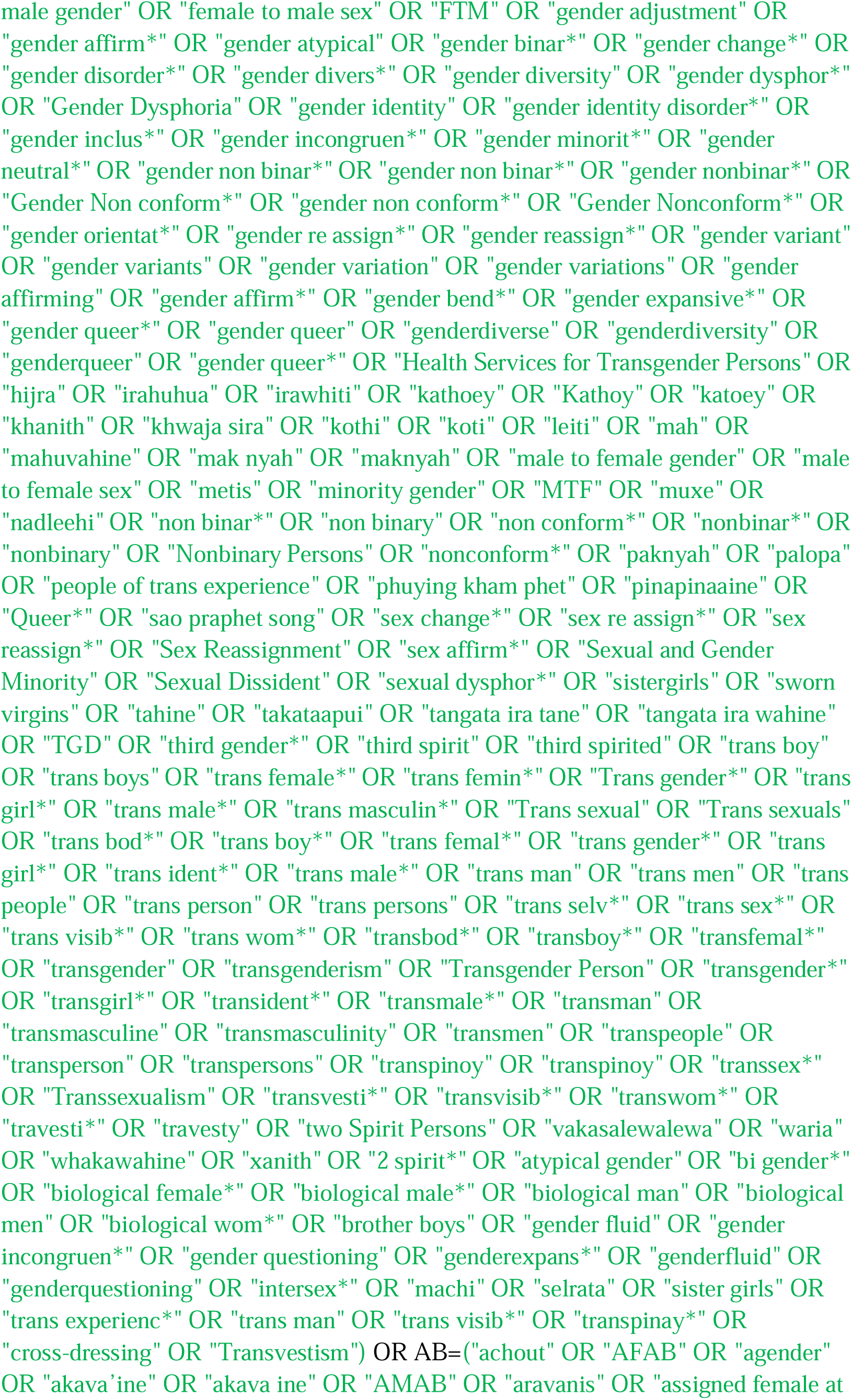
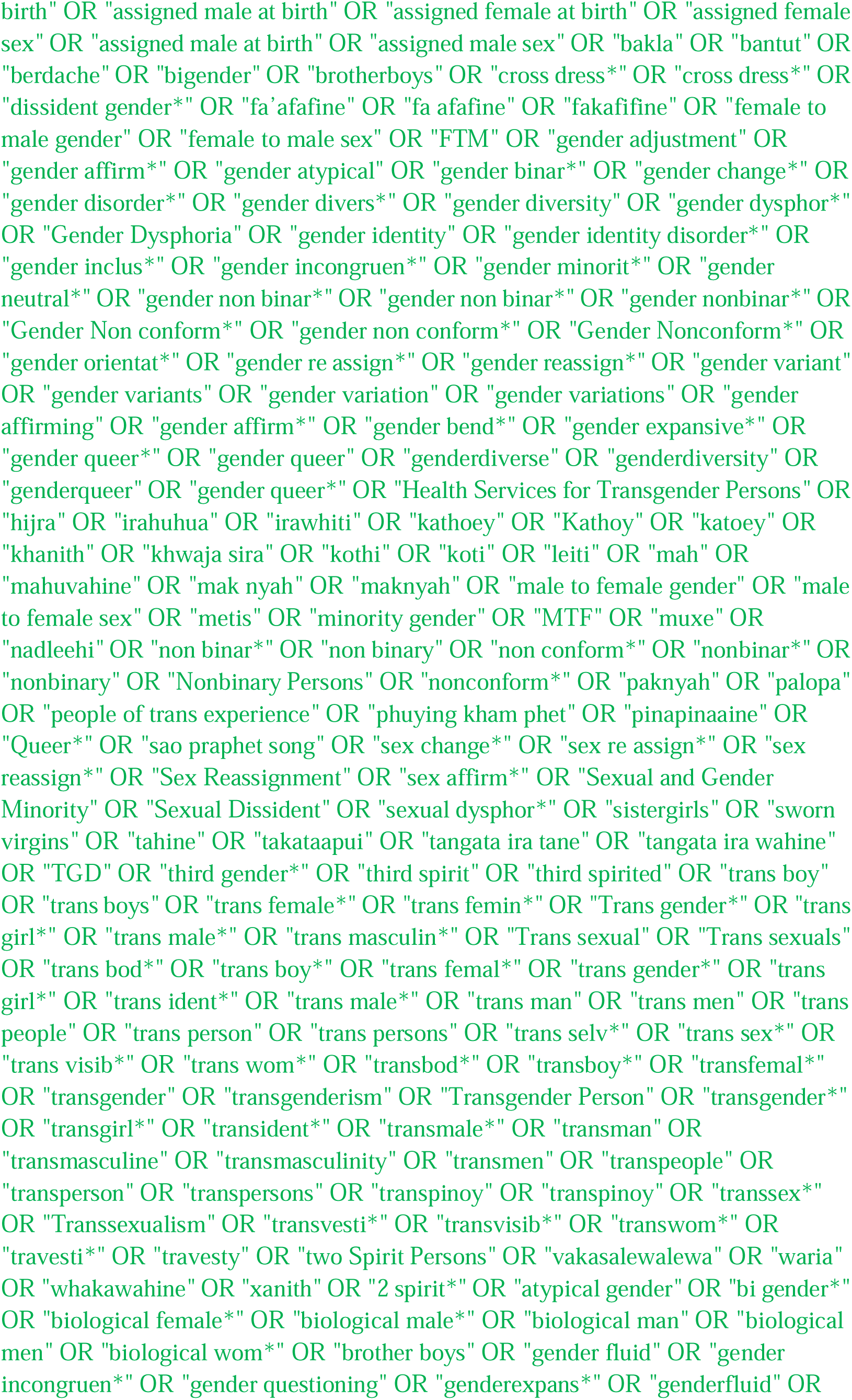
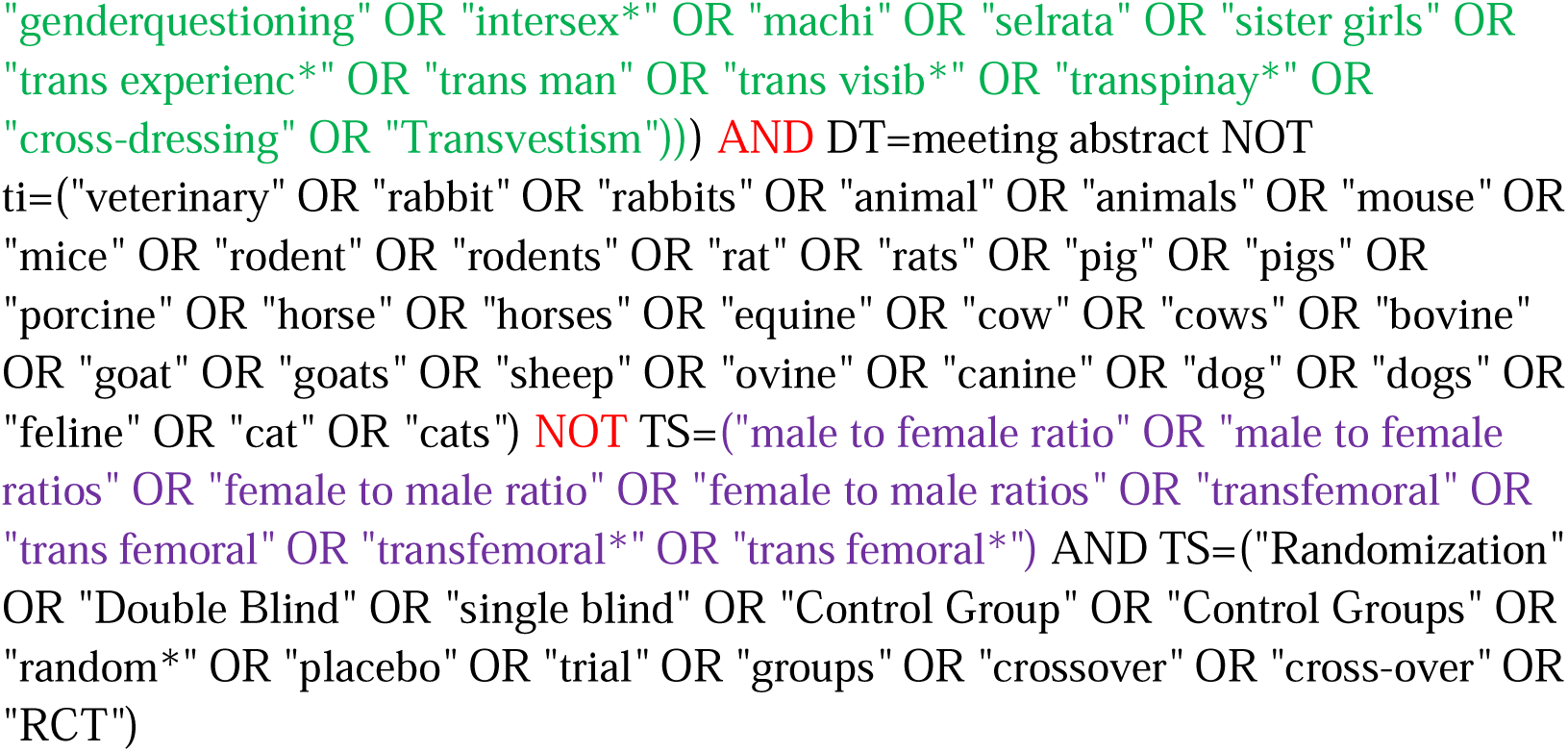

#### Cochrane CENTRAL

https://www.cochranelibrary.com/advanced-search/search-manager

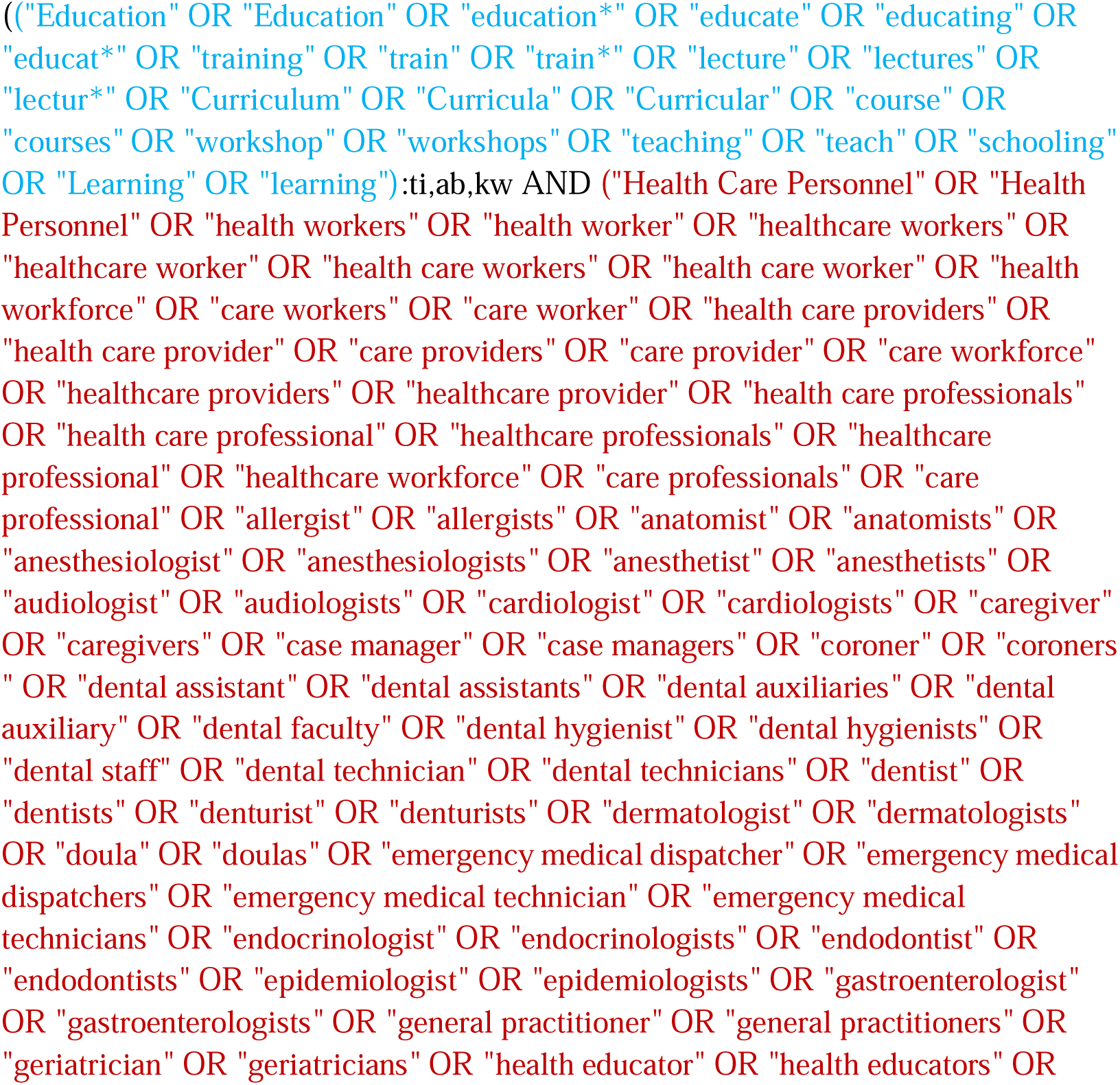
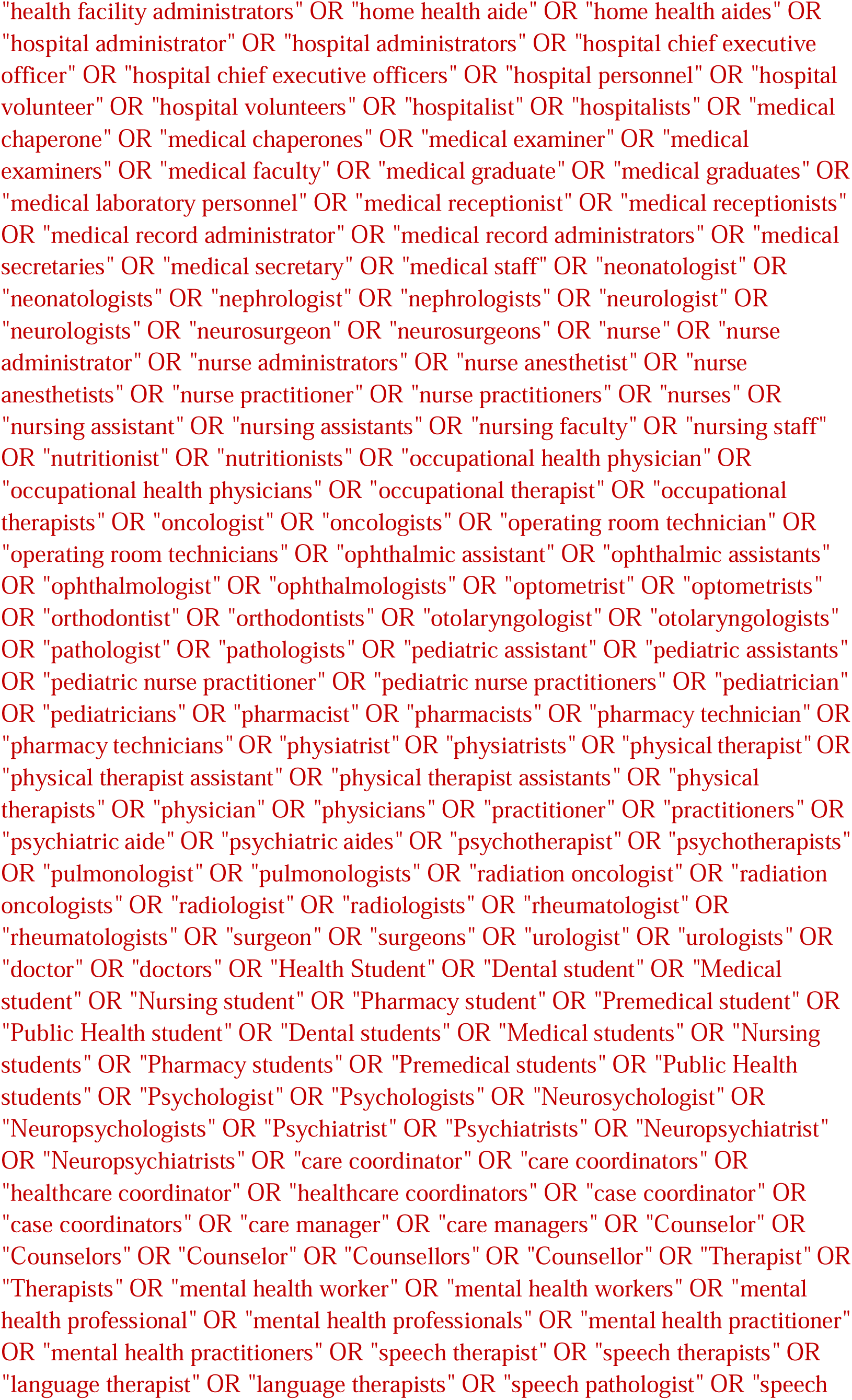
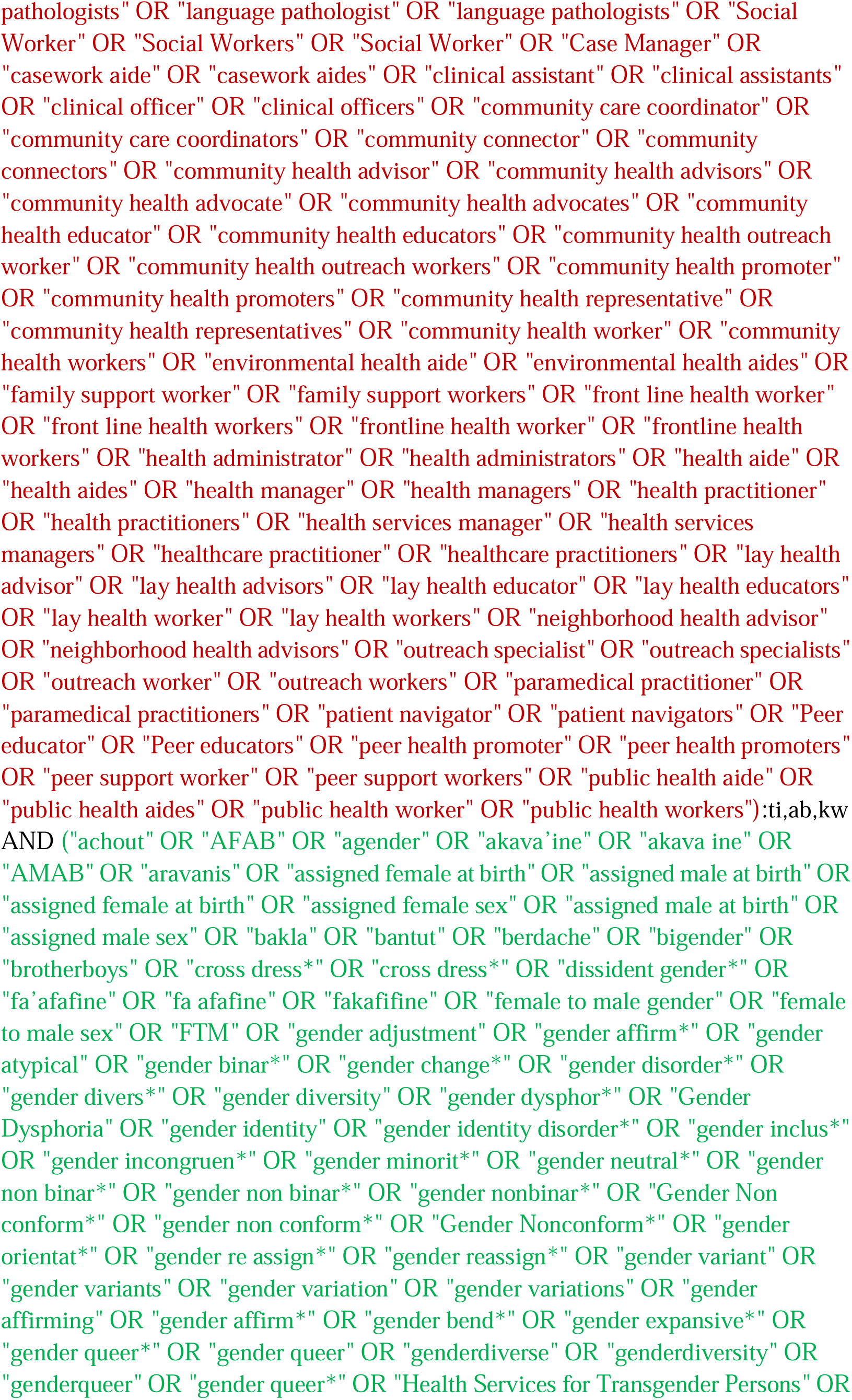
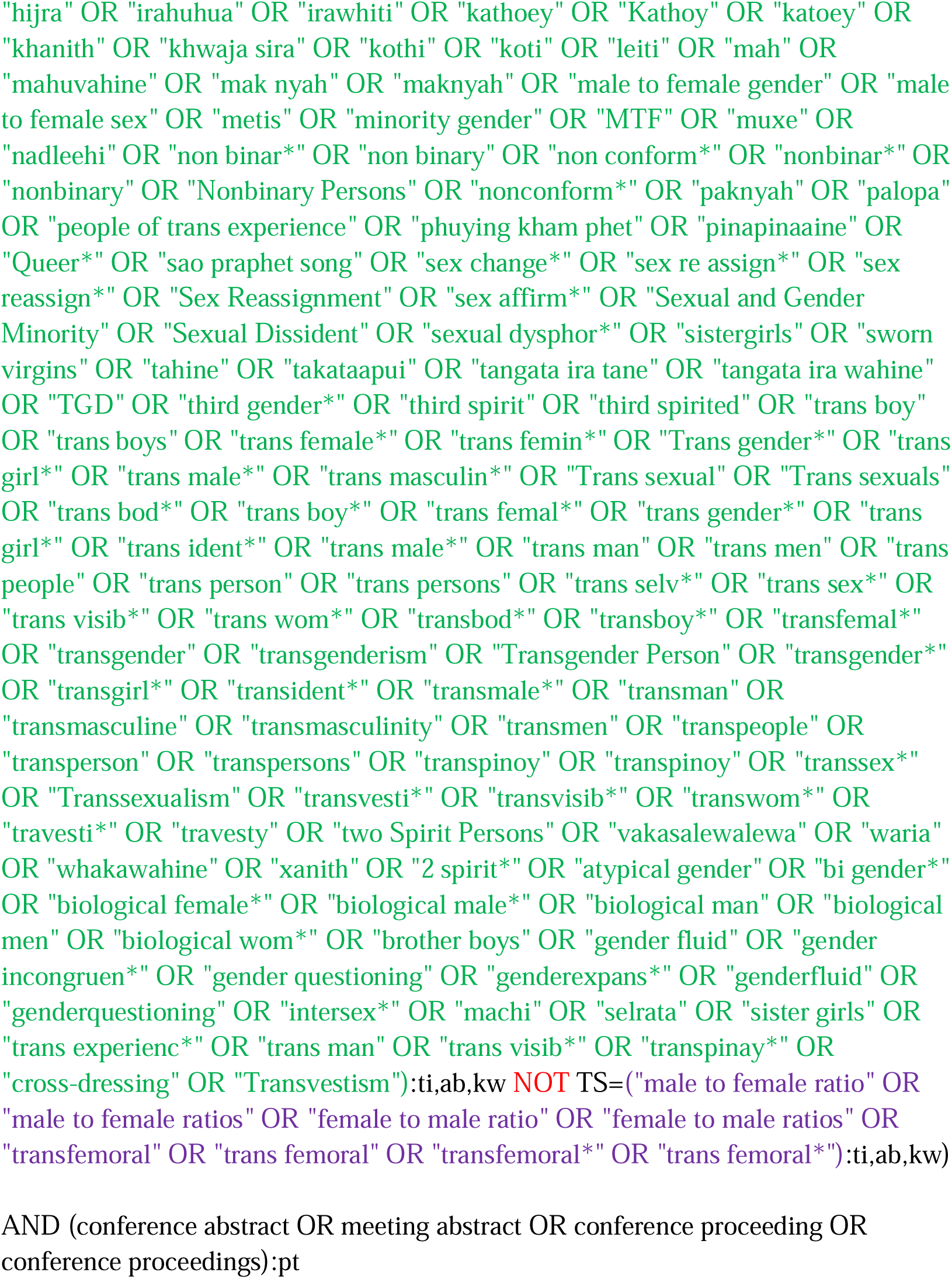

### 2024 Update Search

March 7, 2024: 8145 references, from which 932 new, sourced from:

- PubMed: 3140 - 386 new&unique
- MEDLINE (OVID): 3205 - 13 new&unique
- Embase (OVID): 3046 - 110 new&unique
- Web of Science: 2714 - 117 new&unique
- Cochrane CENTRAL: 82 - 6 new&unique
- Emcare (OVID): 2101 - 43 new&unique
- PsycINFO (EBSCOhost): 2680 - 85 new&unique
- Social Services Abstracts (ProQuest): 564 - 21 new&unique
- Sociological Abstracts (ProQuest): 507 - 27 new&unique
- Academic Search Premier (EBSCOhost): 2154 - 90 new&unique
- Psychology and Behavioral Sciences Collection (EBSCOhost): 341 - 0 new&unique
- ERIC (ProQuest): 479 - 28 new&unique
- LILACS: 60 - 6 new&unique

#### PubMed

http://www.ncbi.nlm.nih.gov/pubmed?otool=leiden

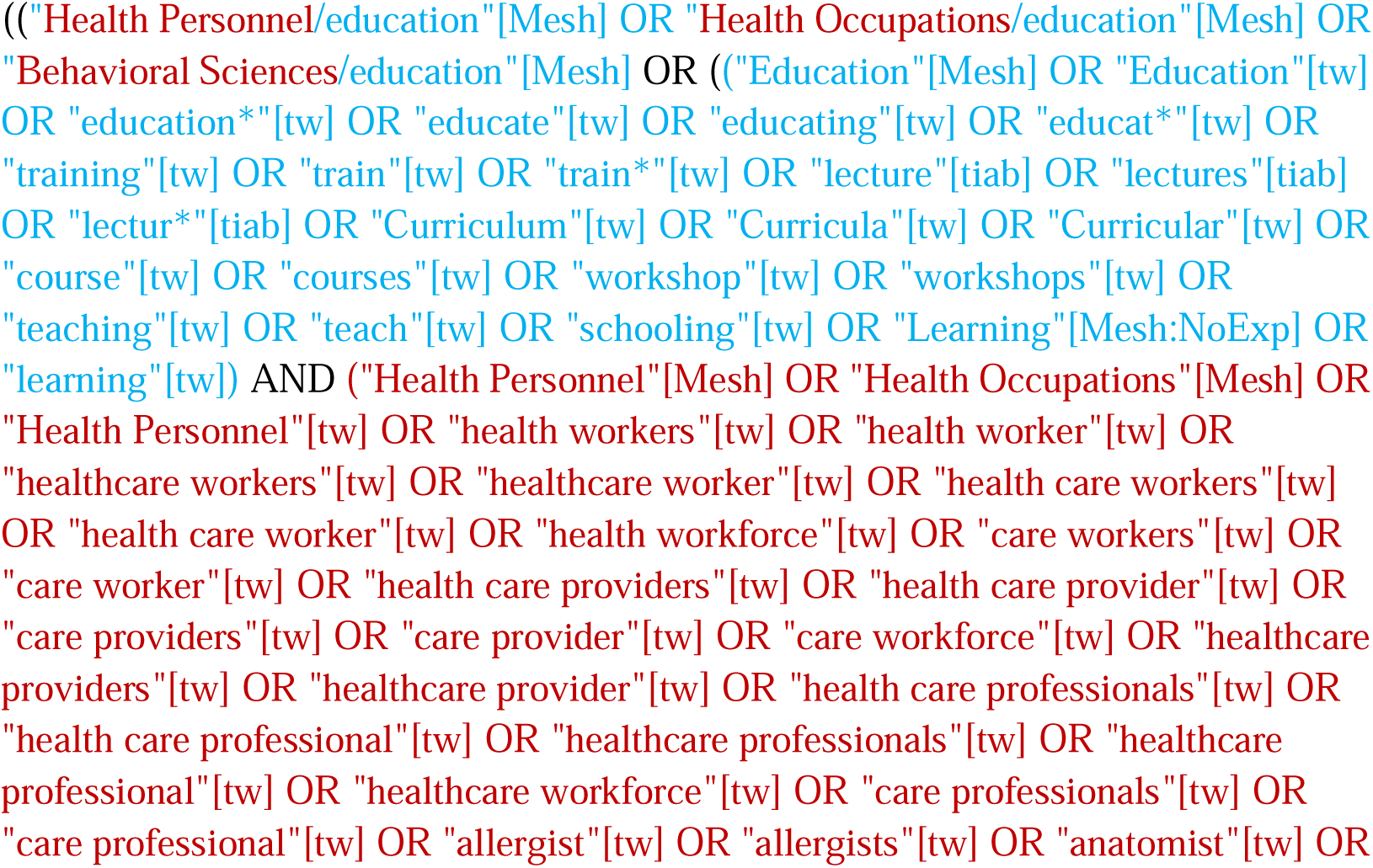
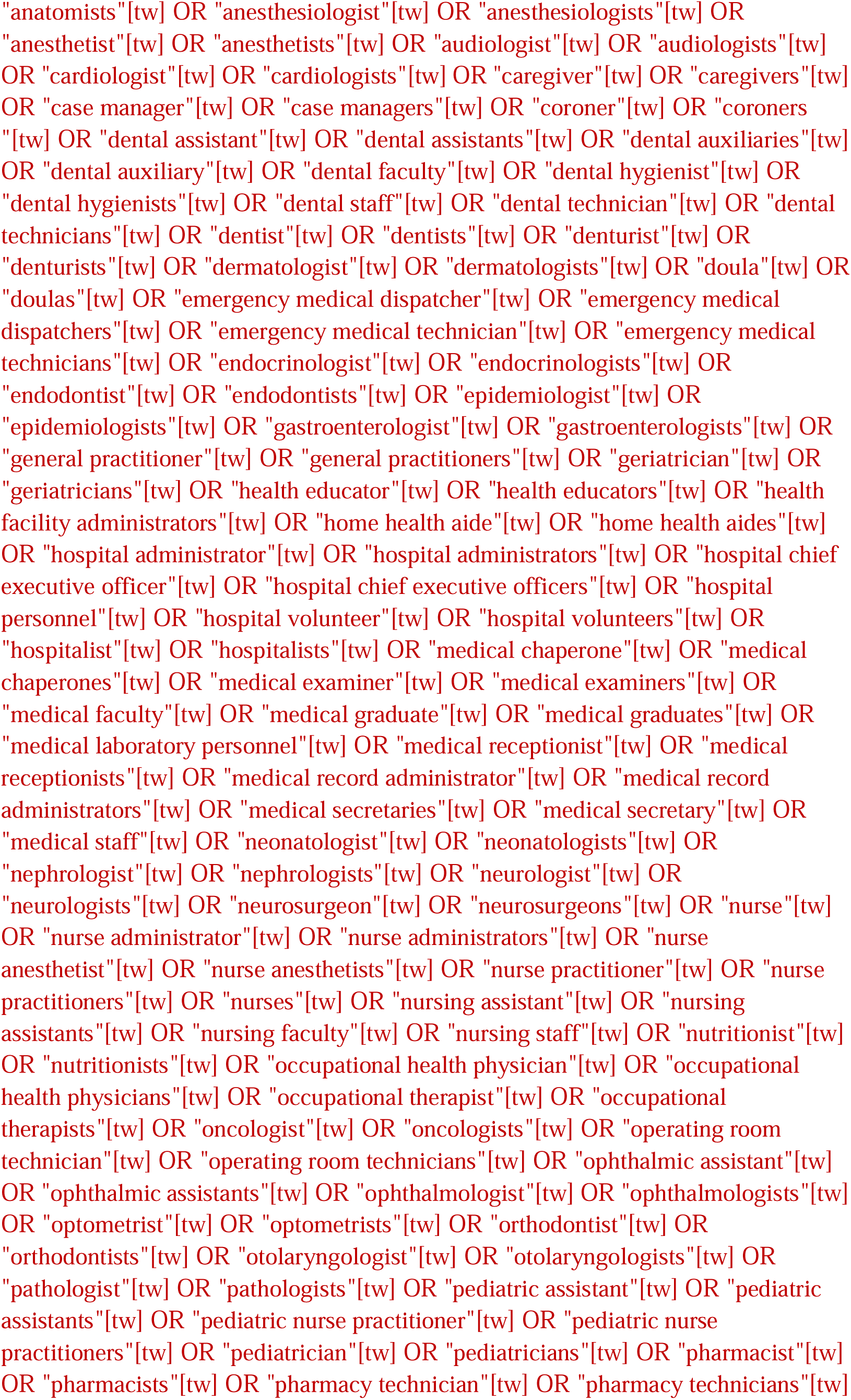
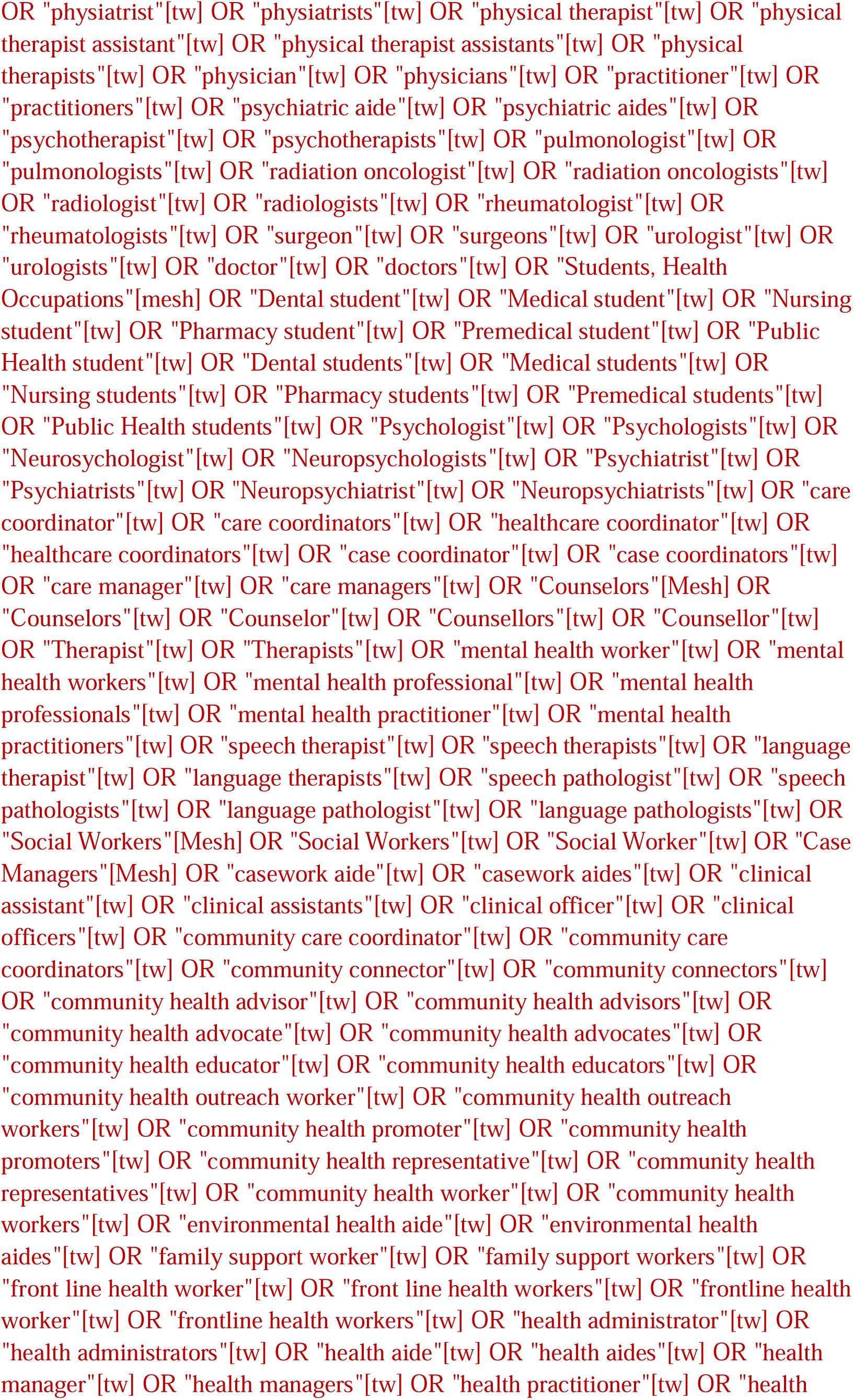
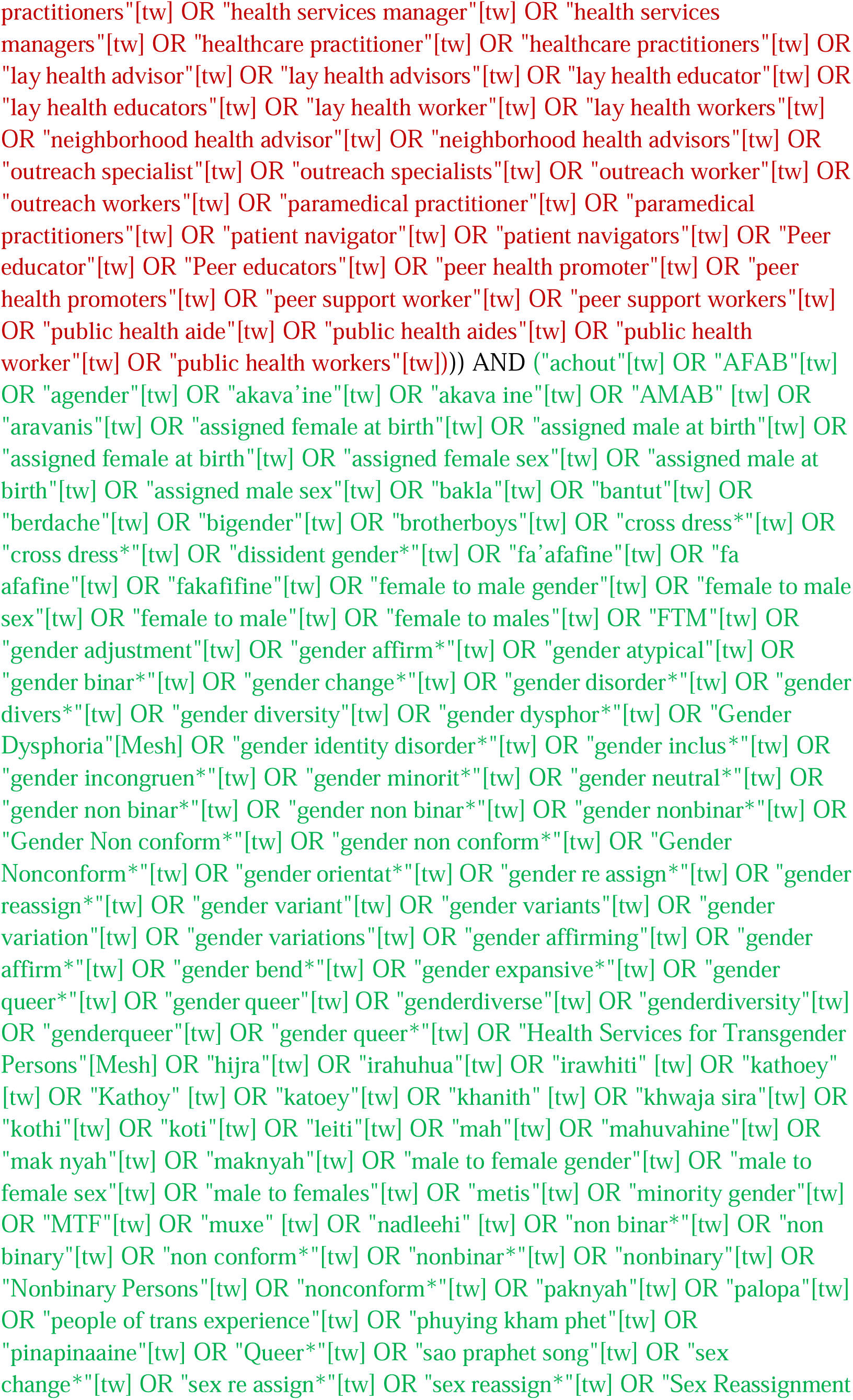
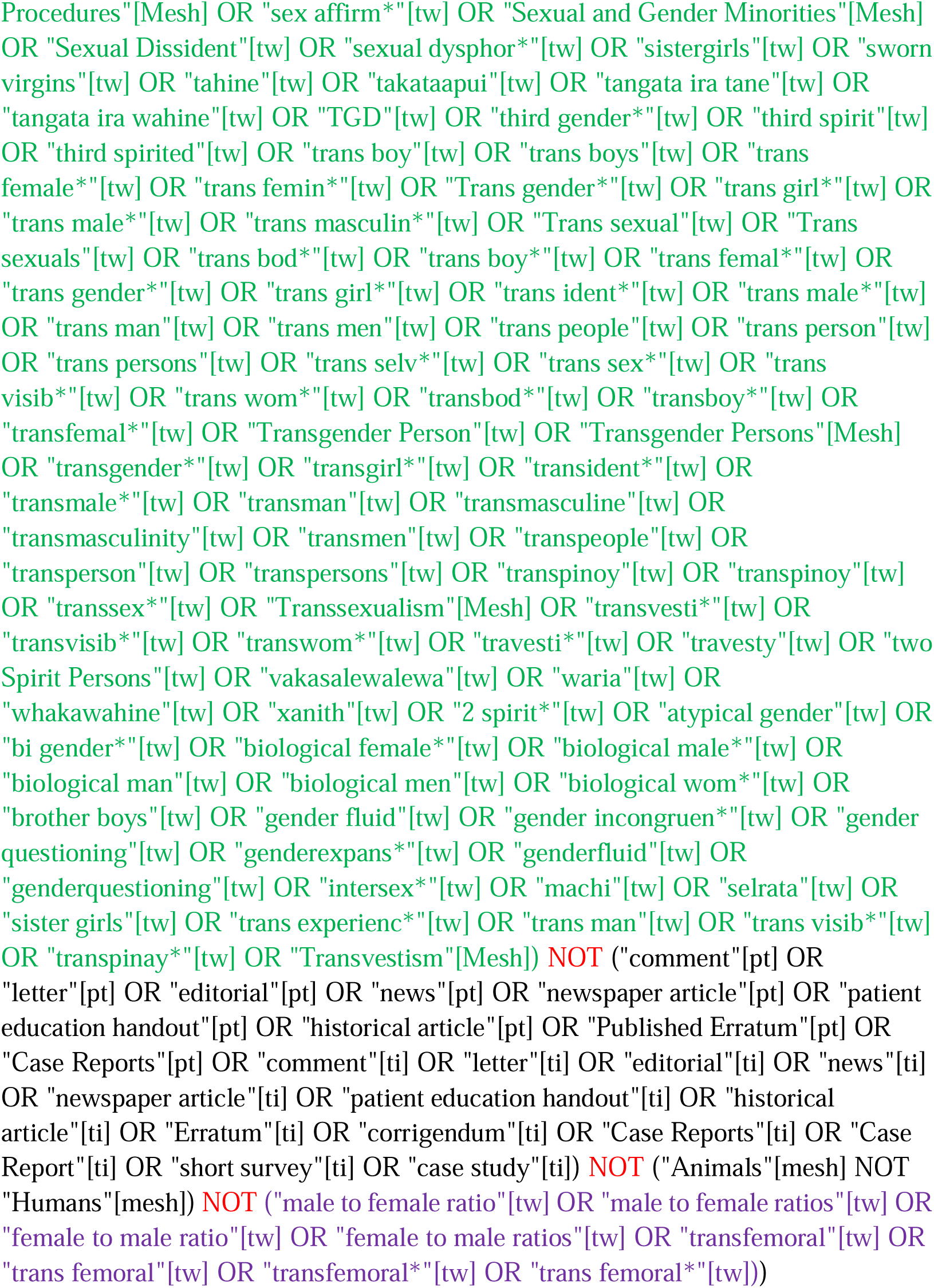

#### MEDLINE via OVID

http://gateway.ovid.com/ovidweb.cgi?T=JS&MODE=ovid&NEWS=n&PAGE=main&D=medall

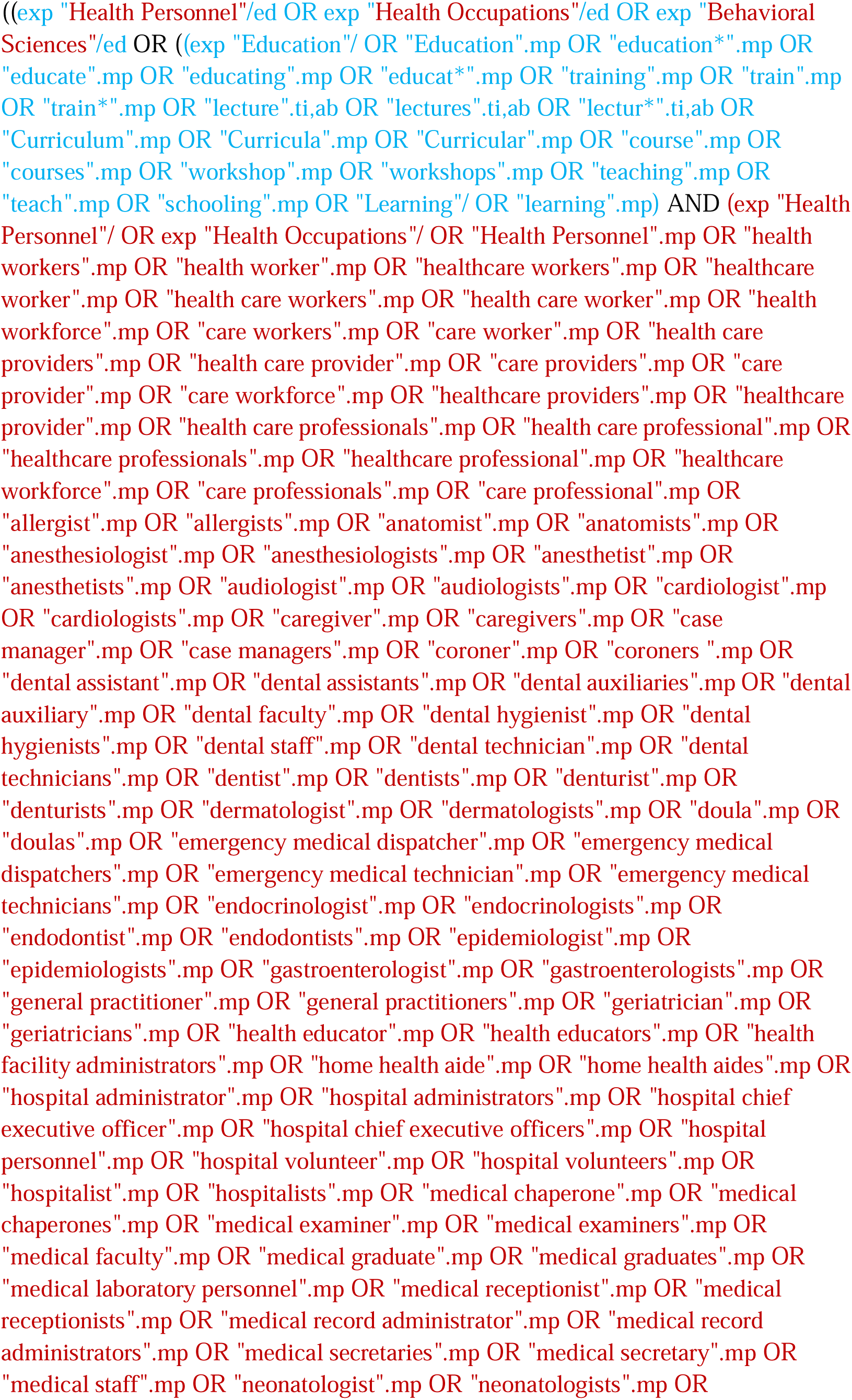
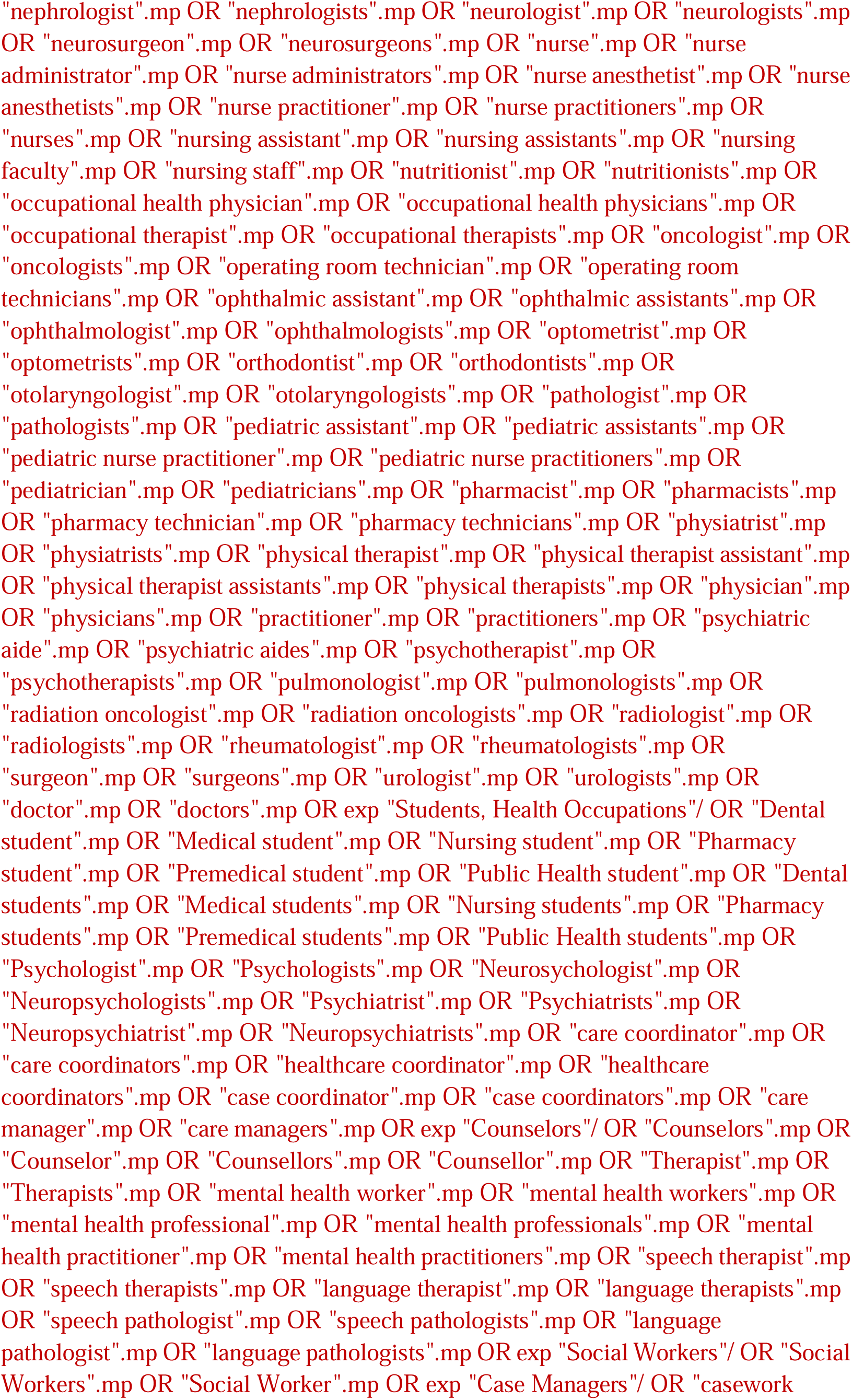
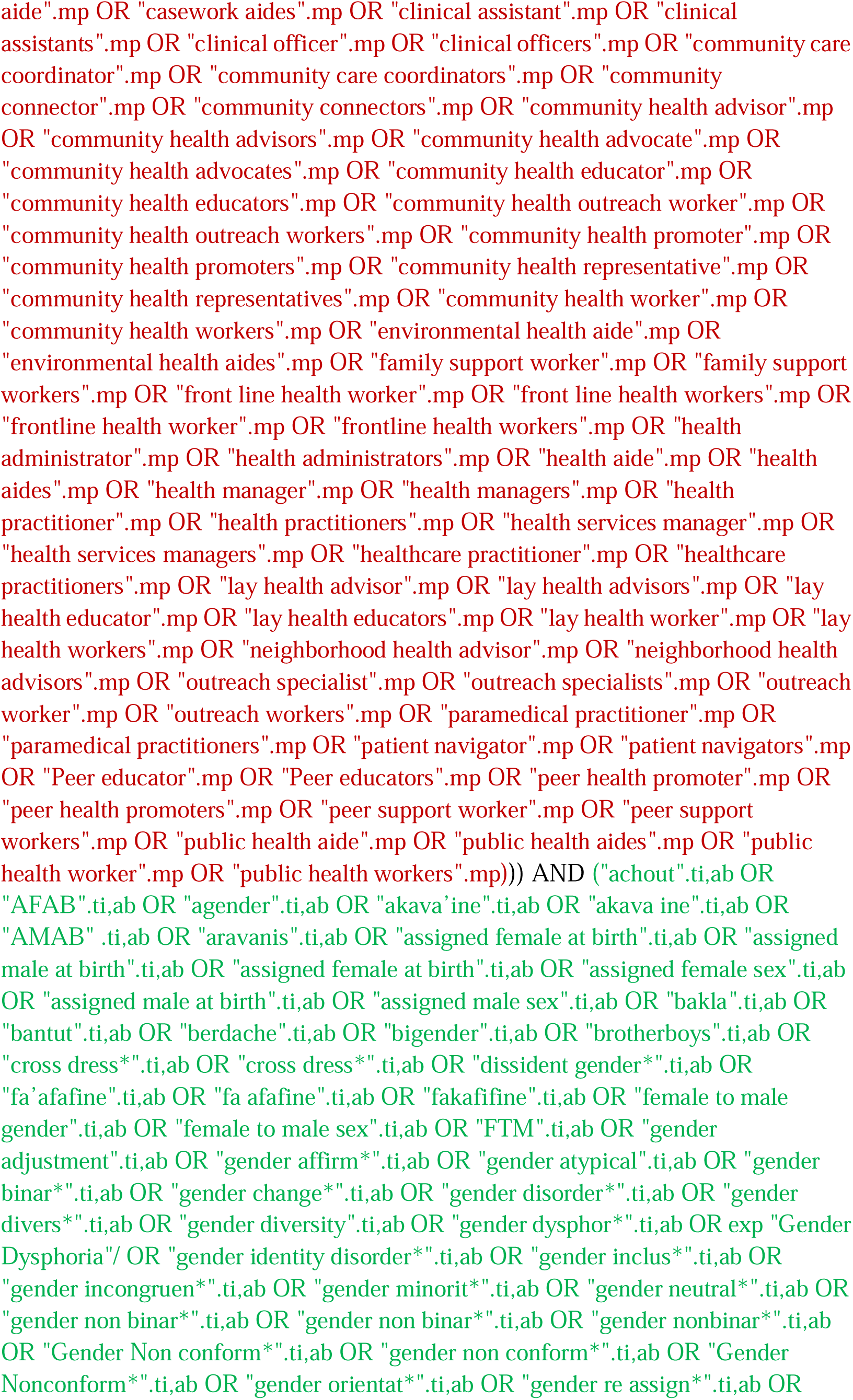
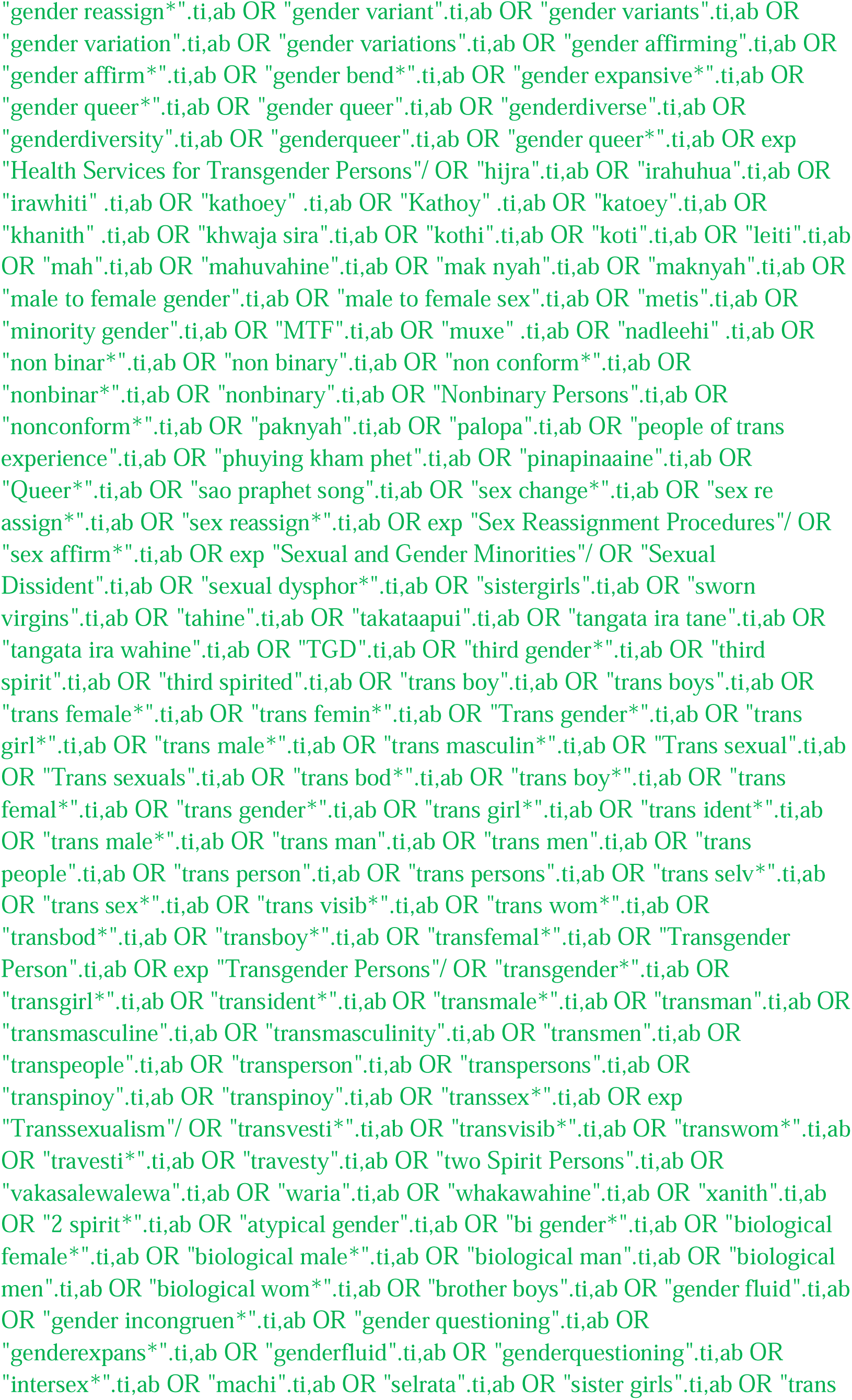
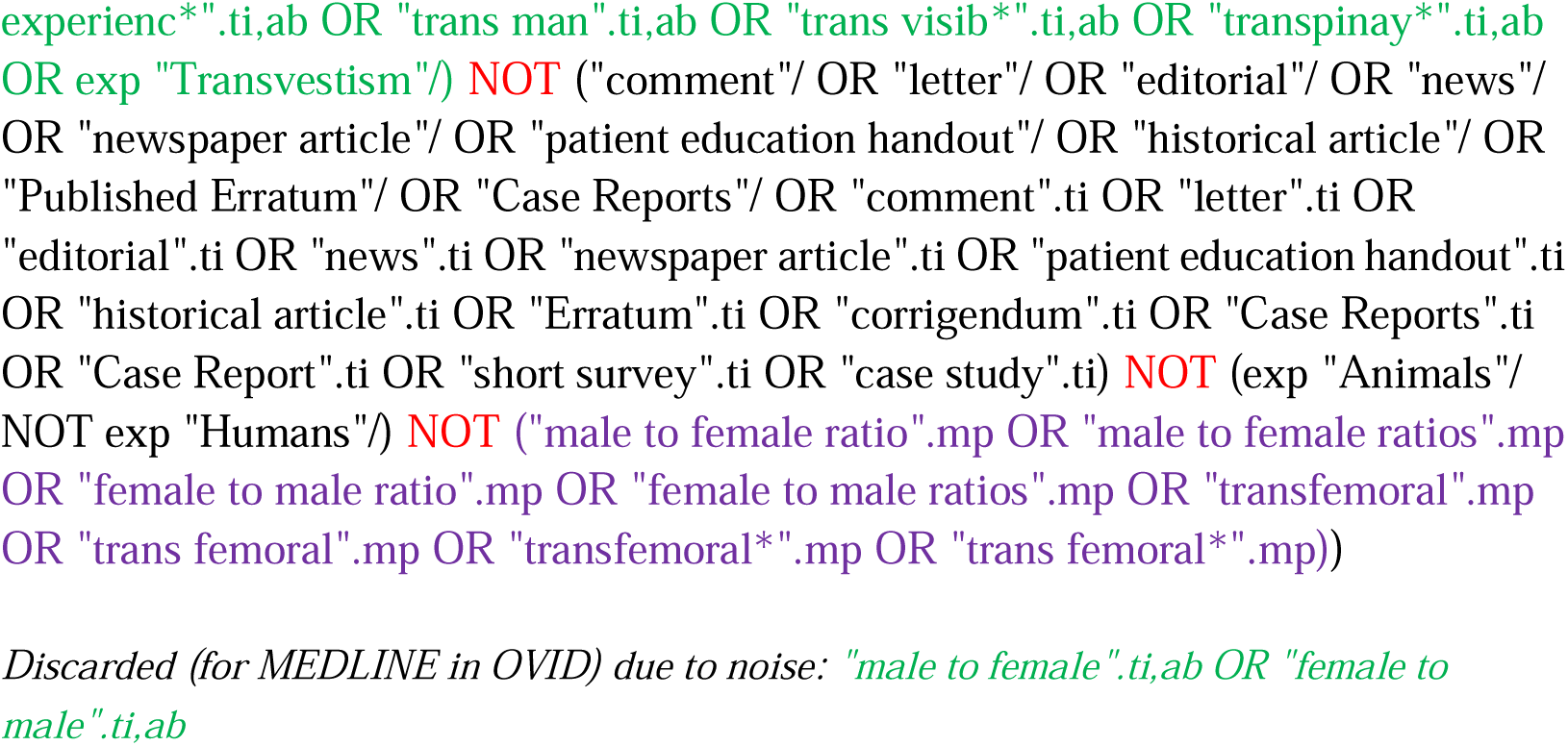

#### Embase

http://ovidsp.ovid.com/ovidweb.cgi?T=JS&PAGE=main&MODE=ovid&D=oemezd

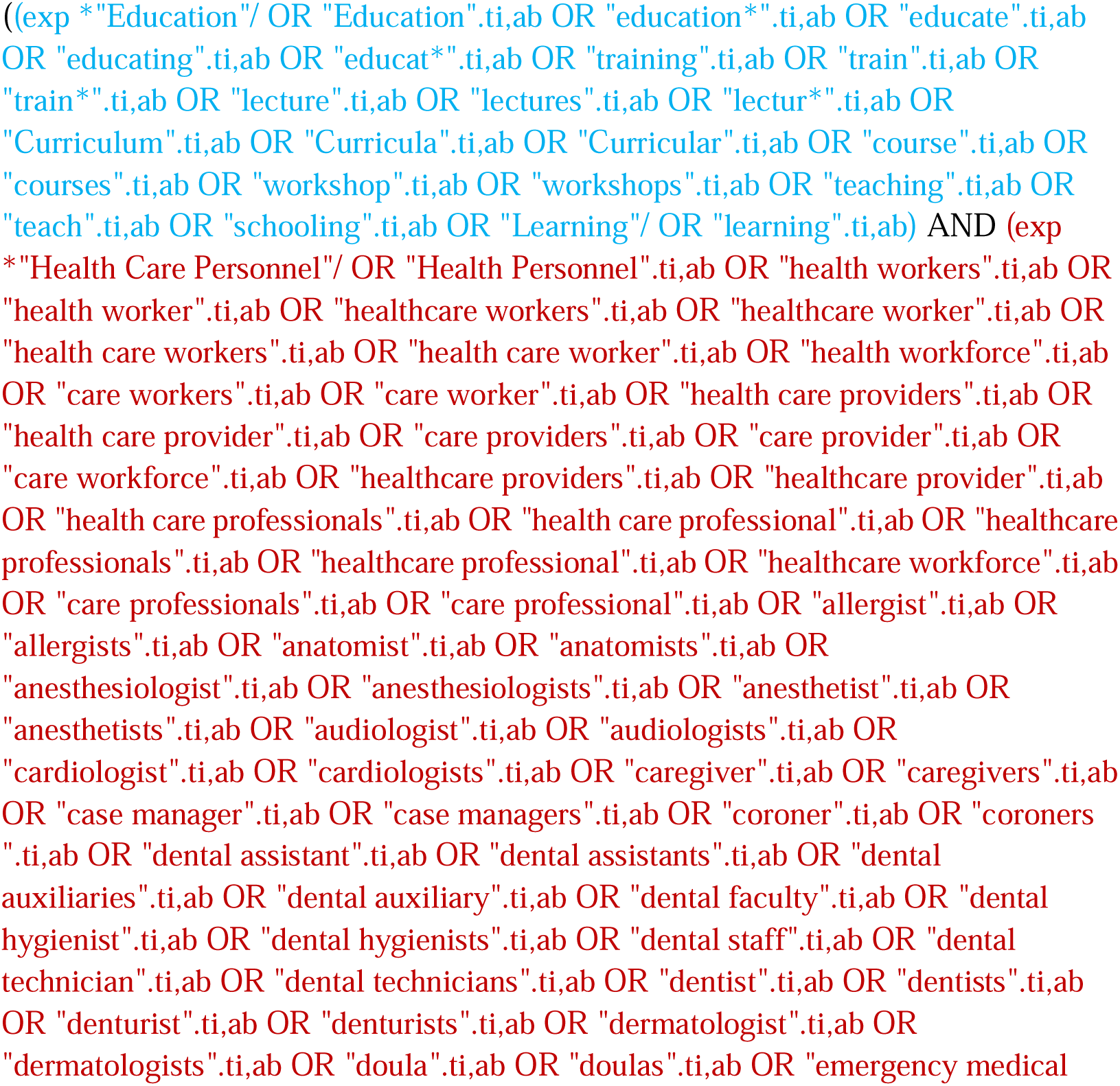
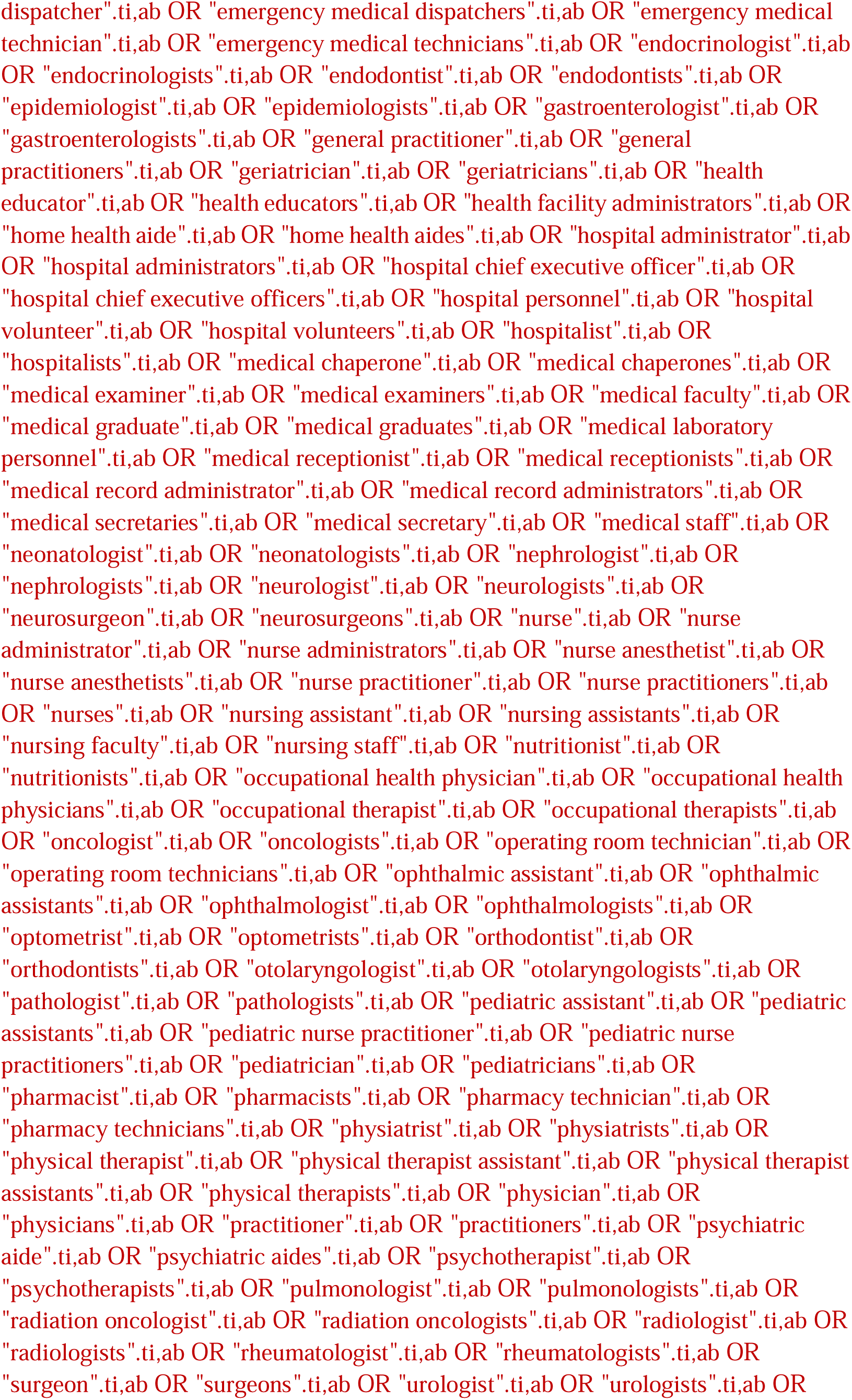
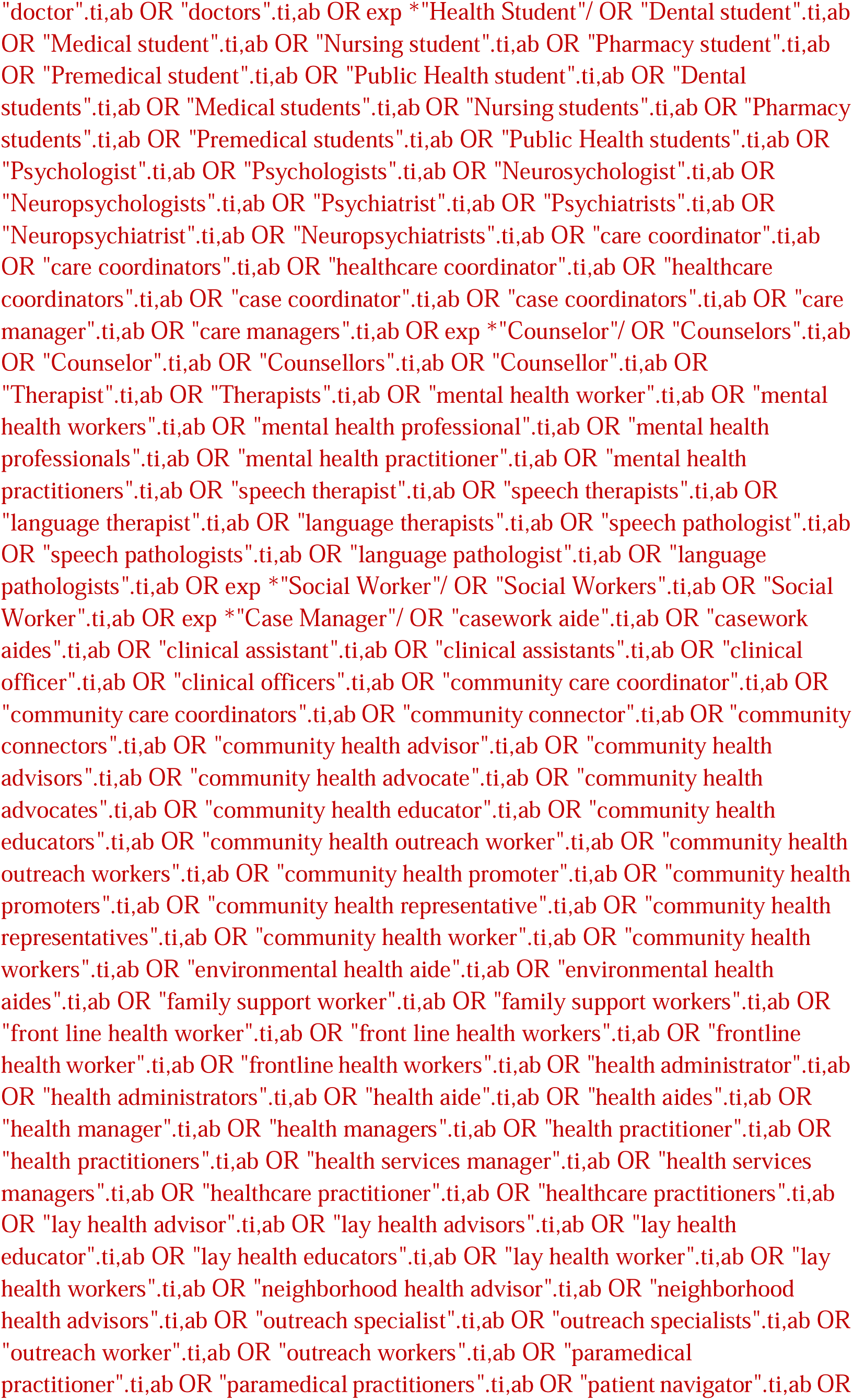
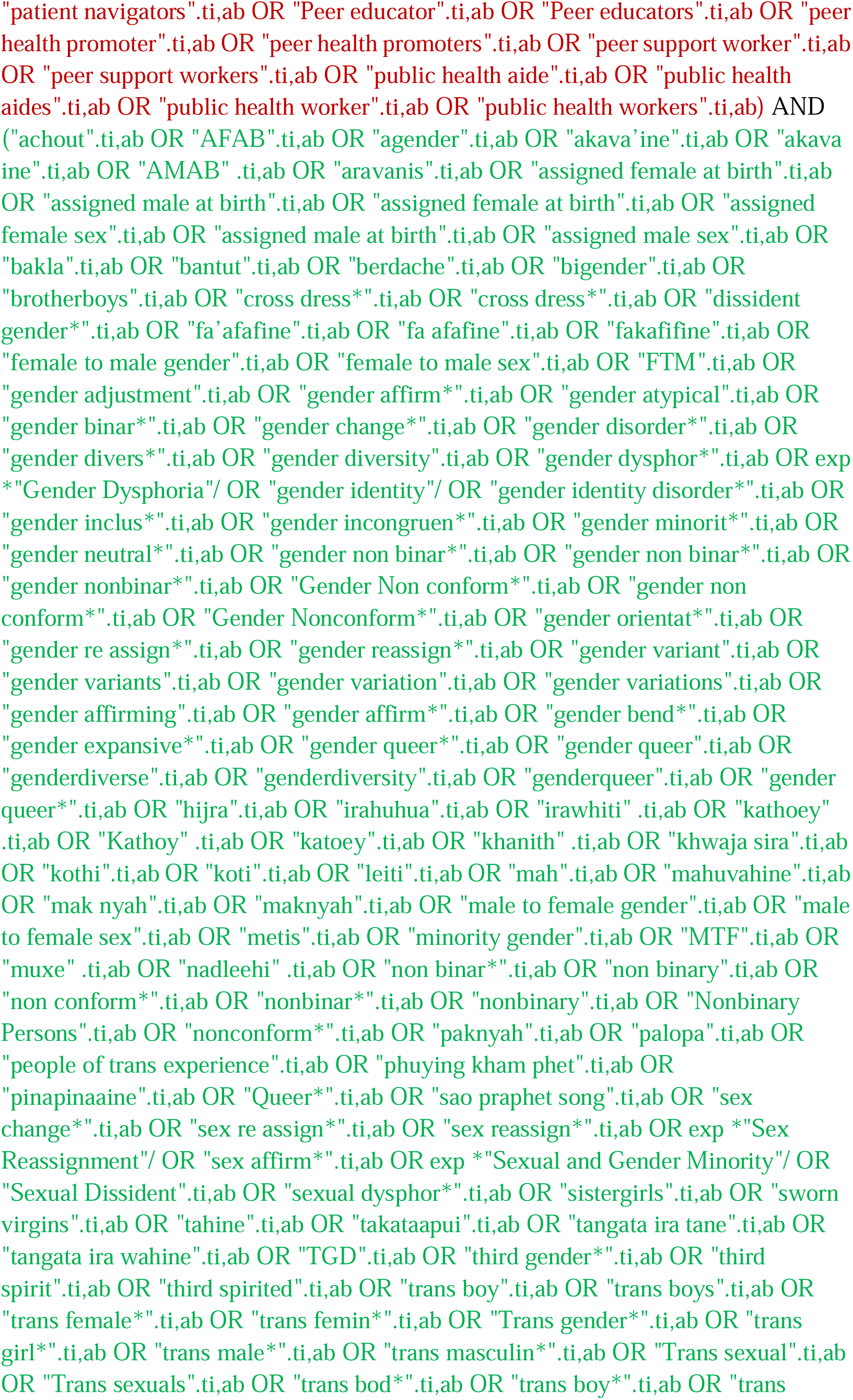
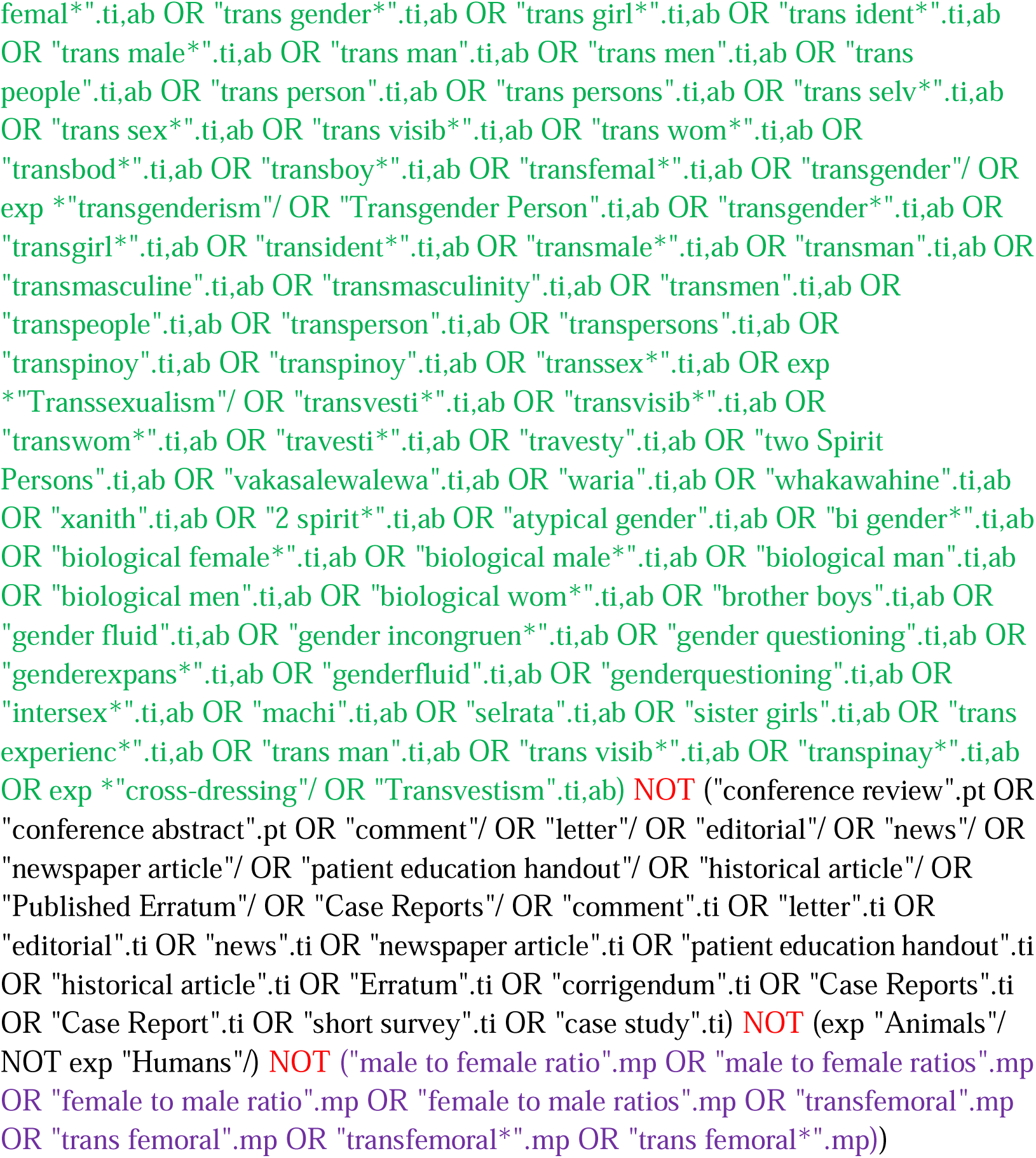

#### Web of Science

http://isiknowledge.com/wos

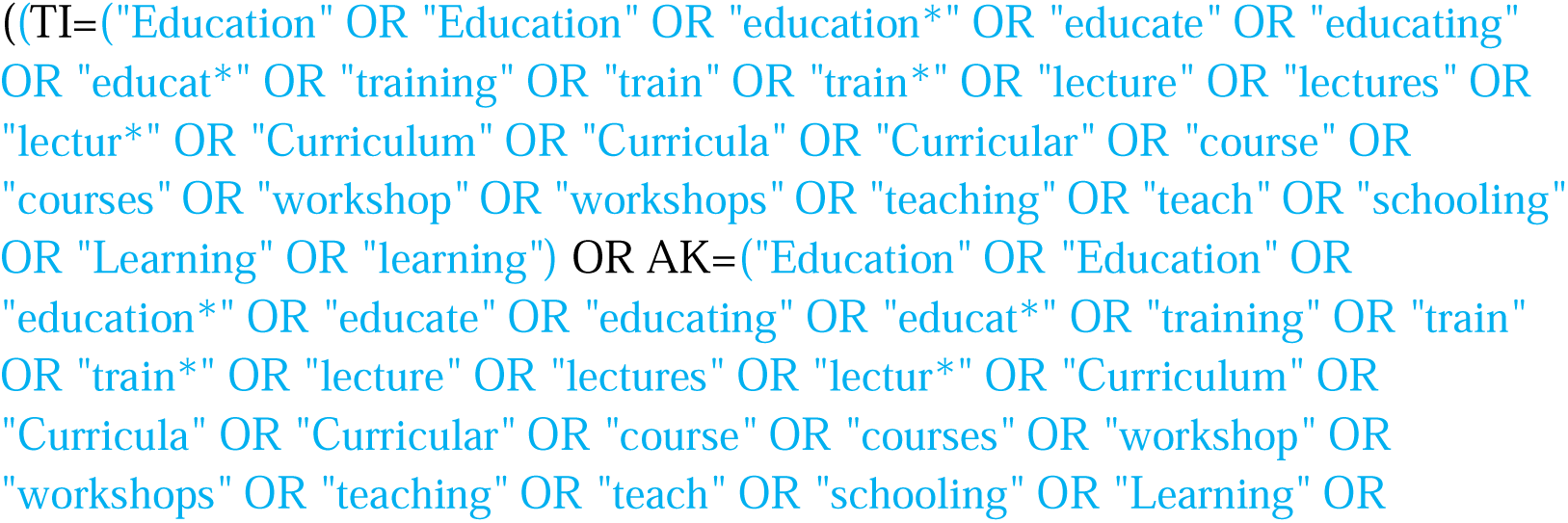
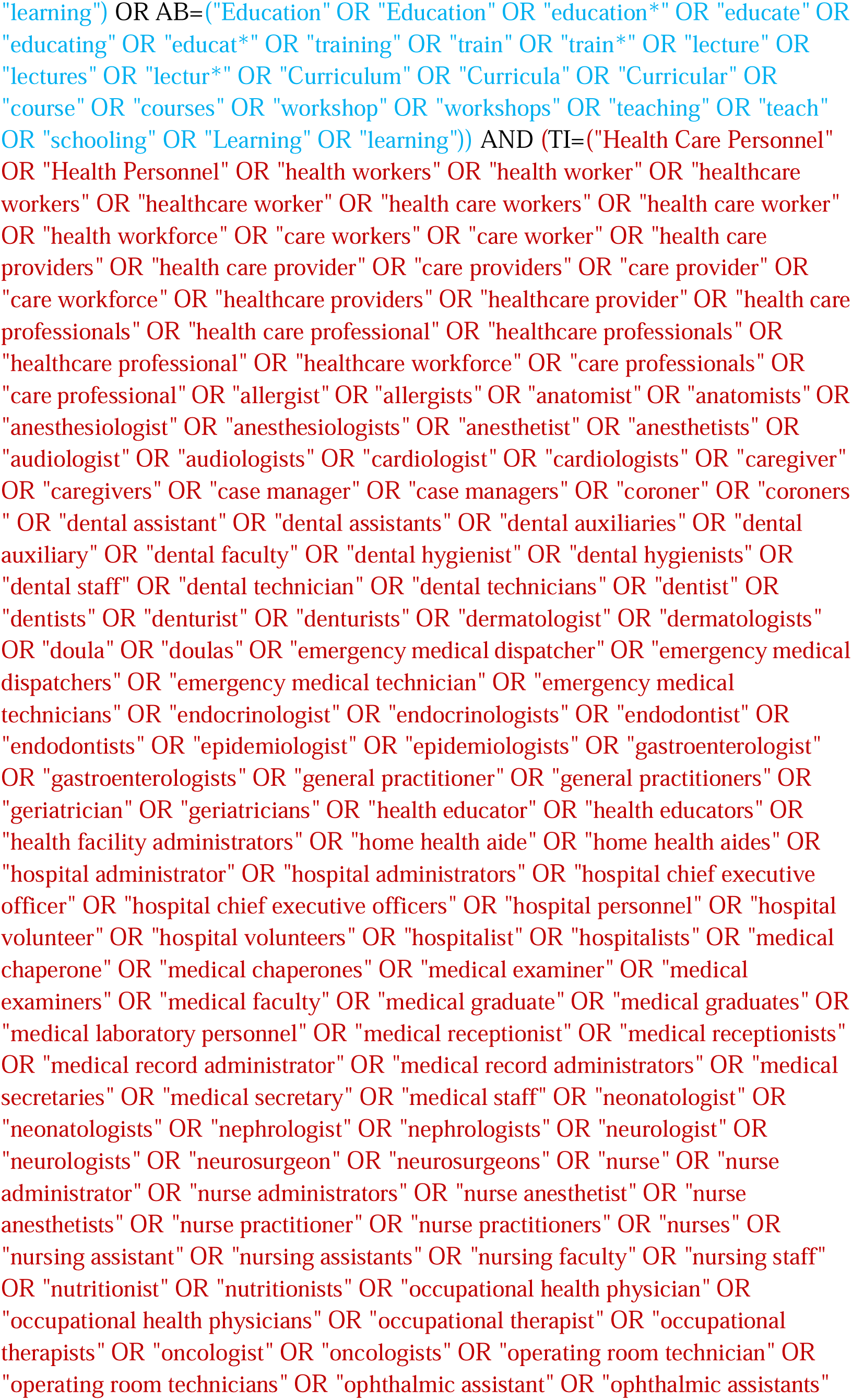
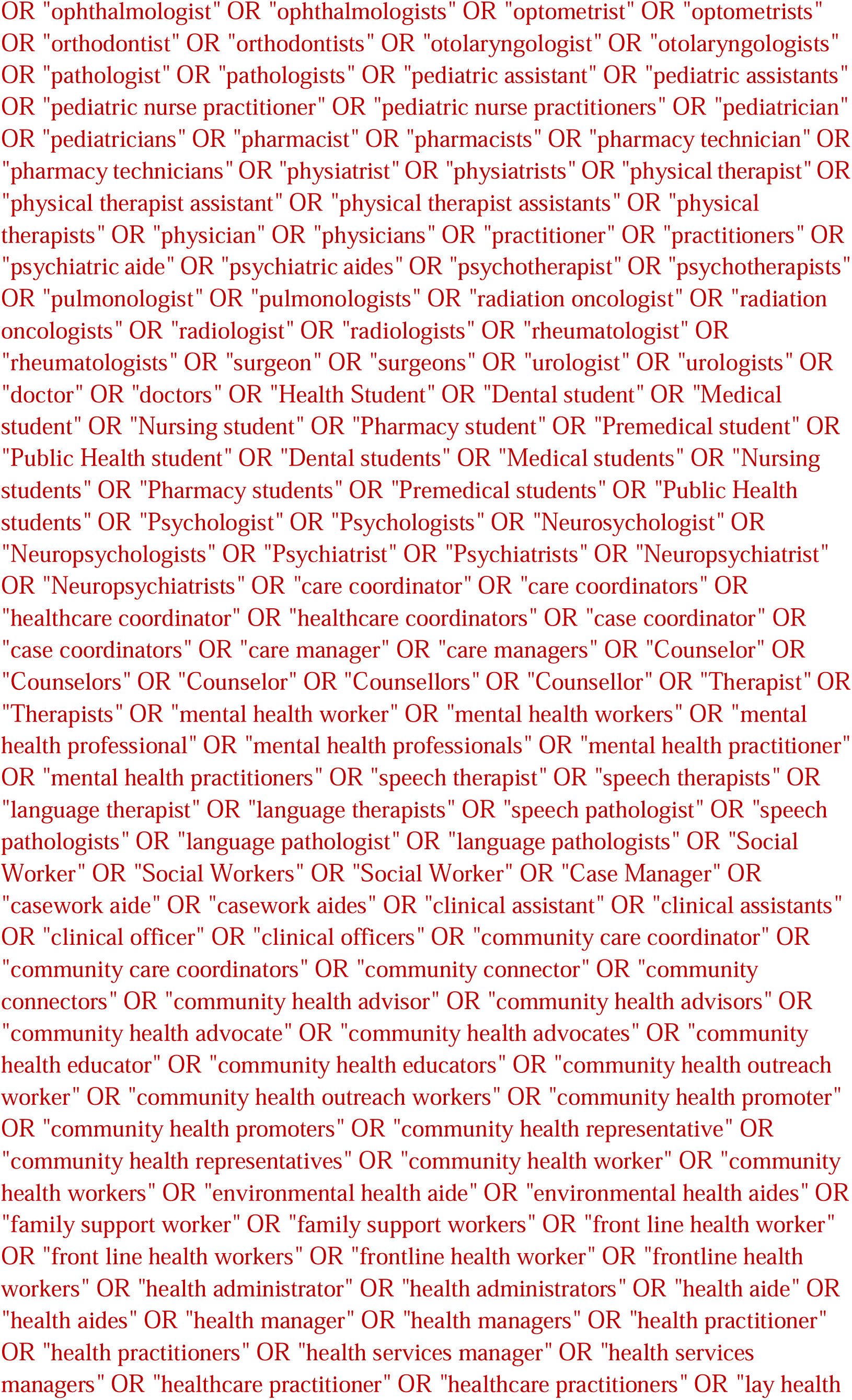
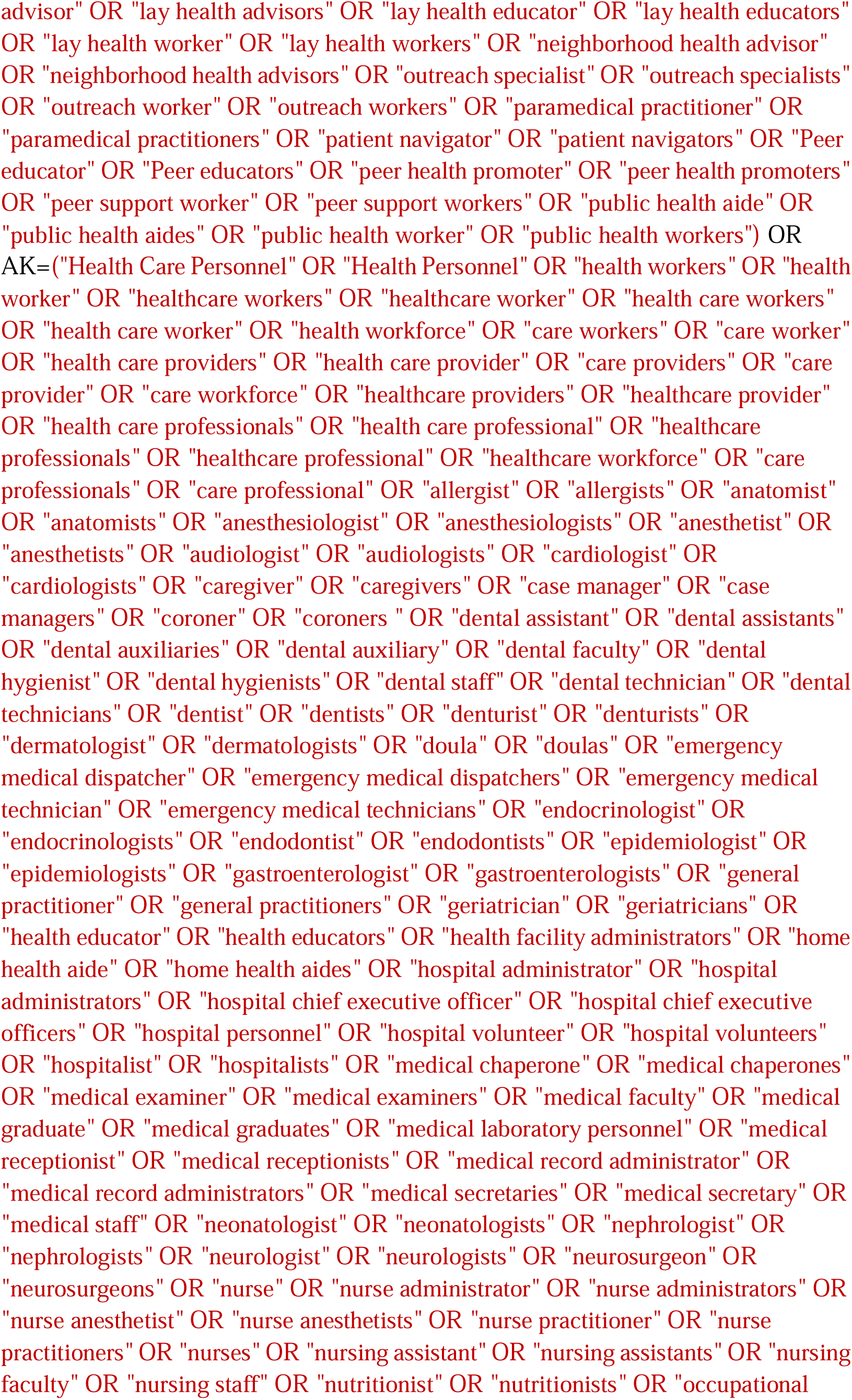
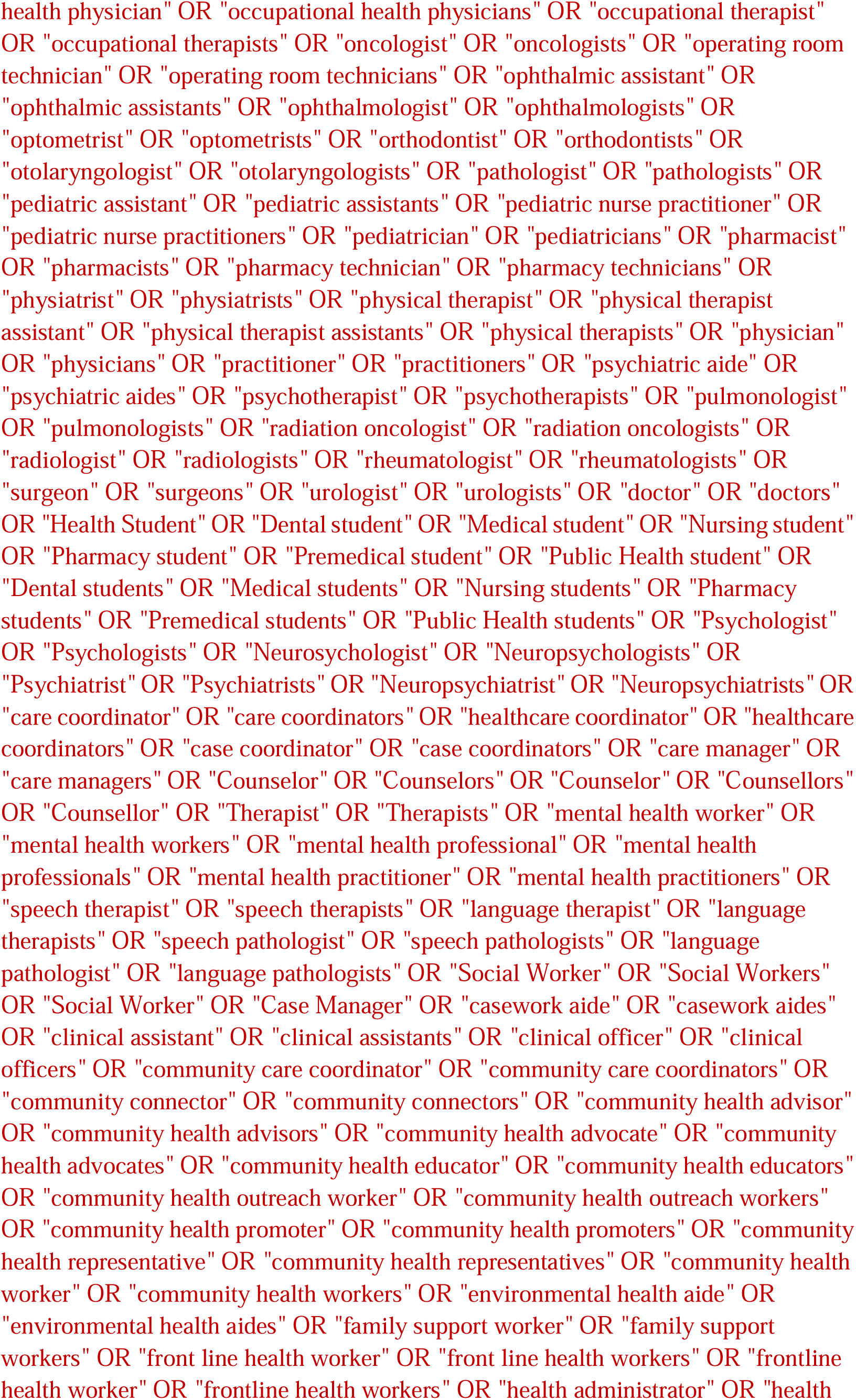
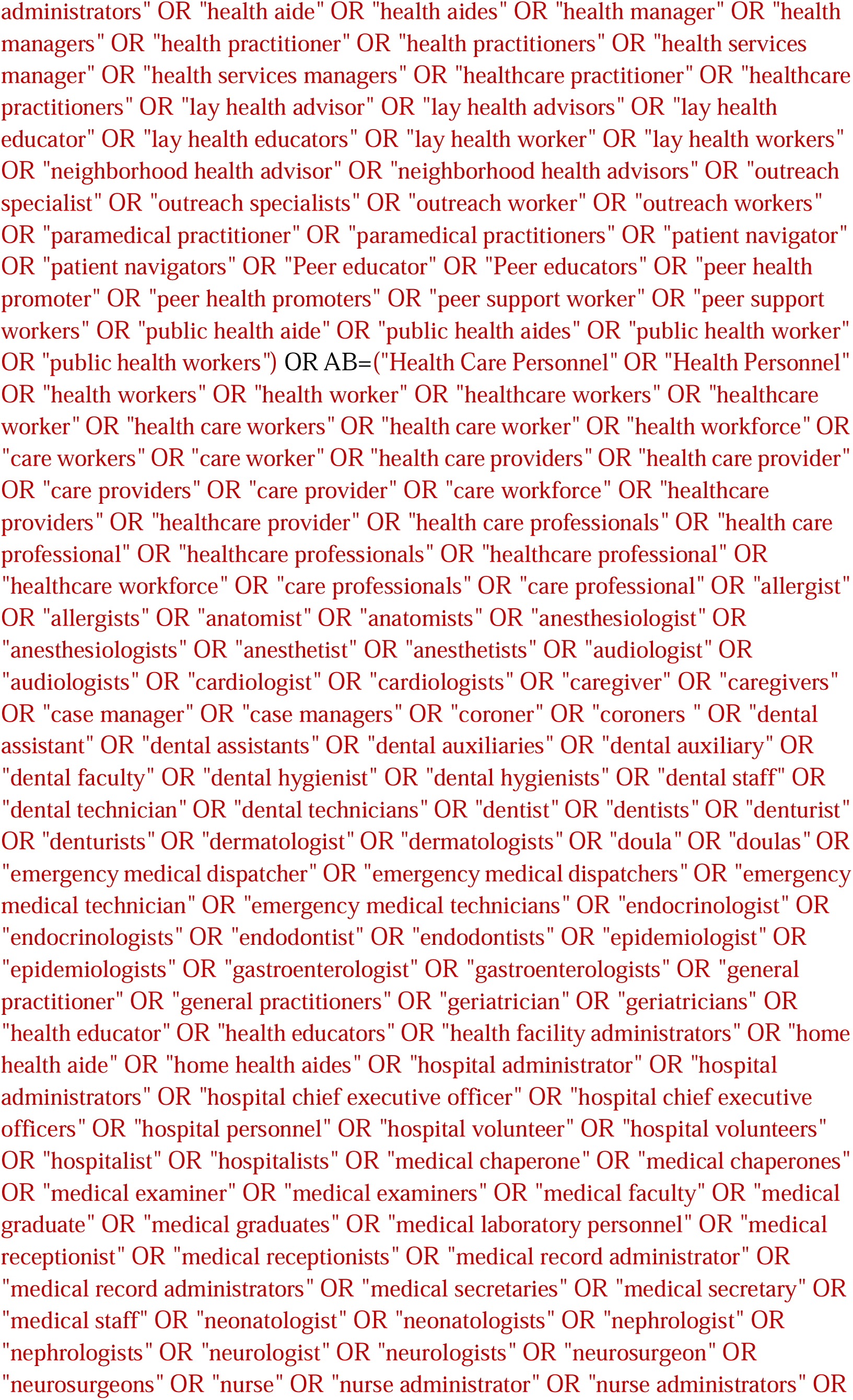
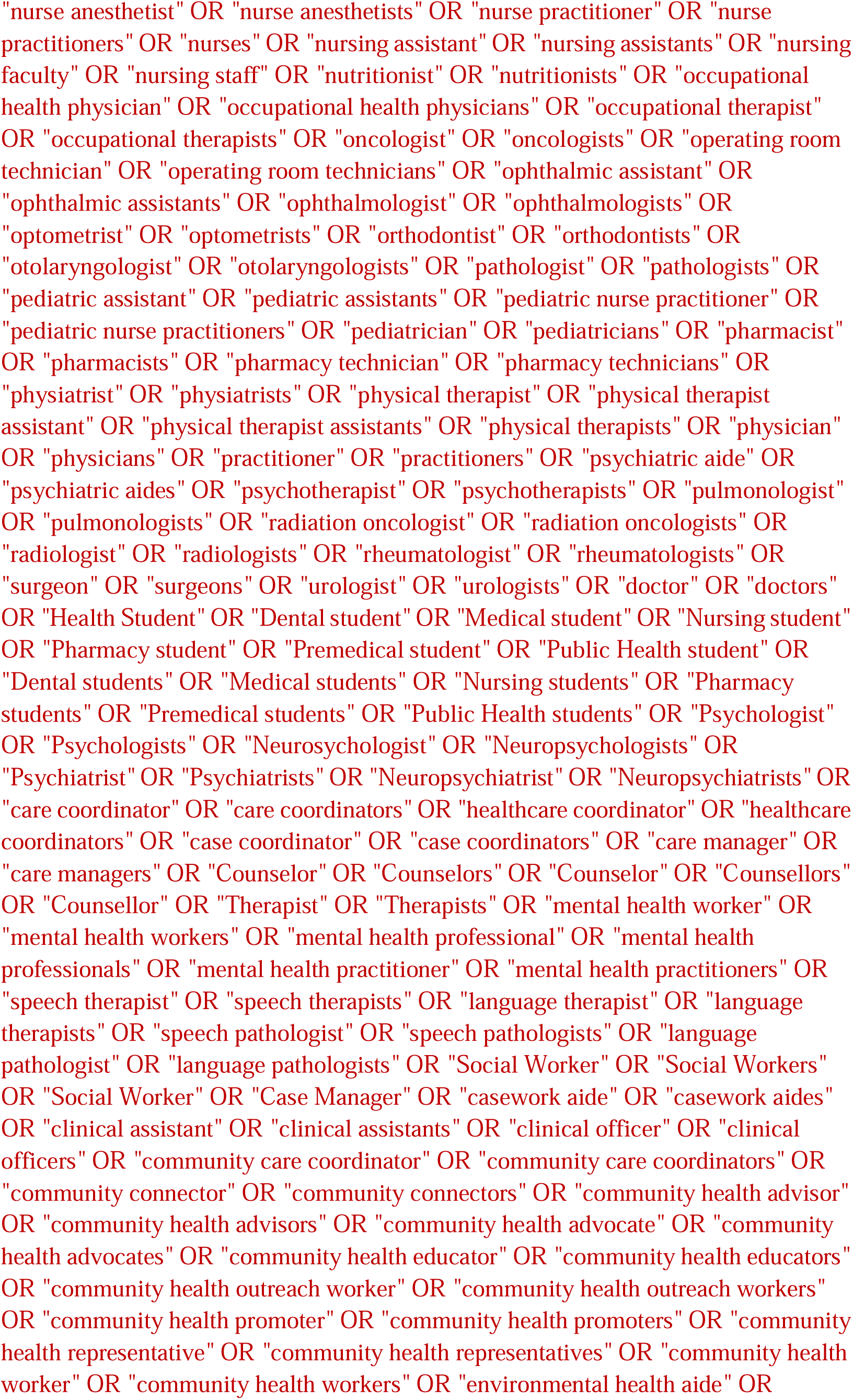
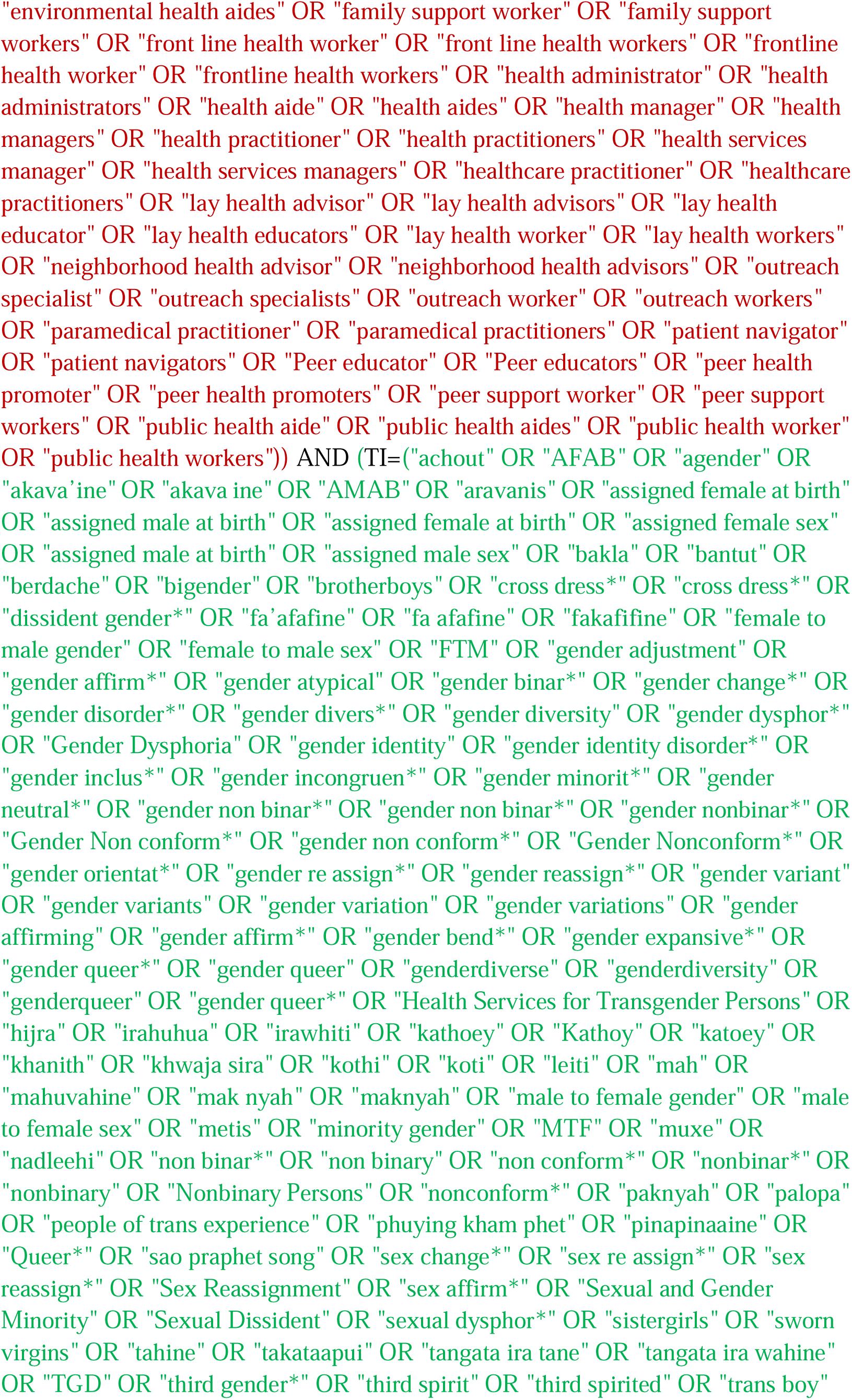
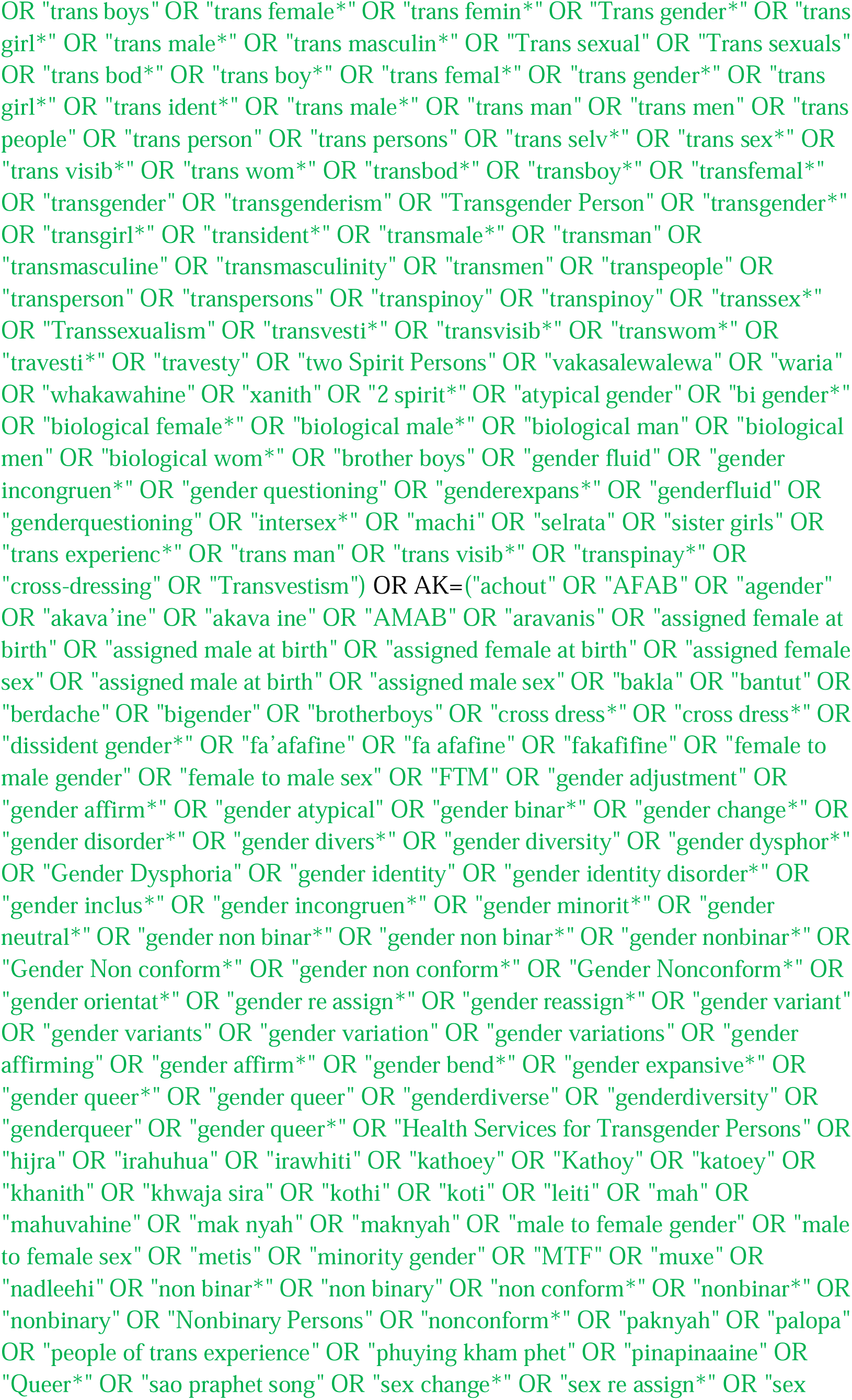
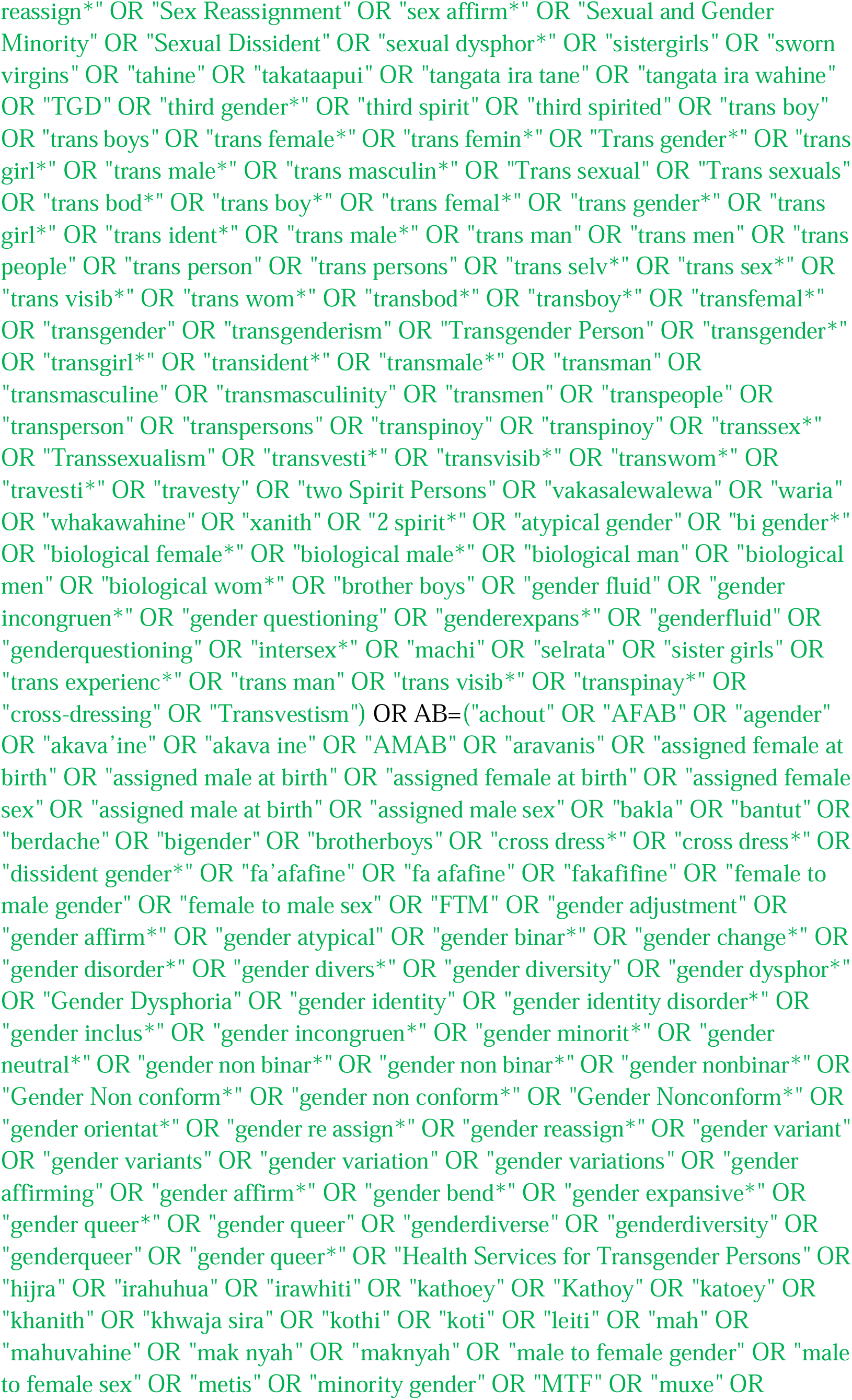
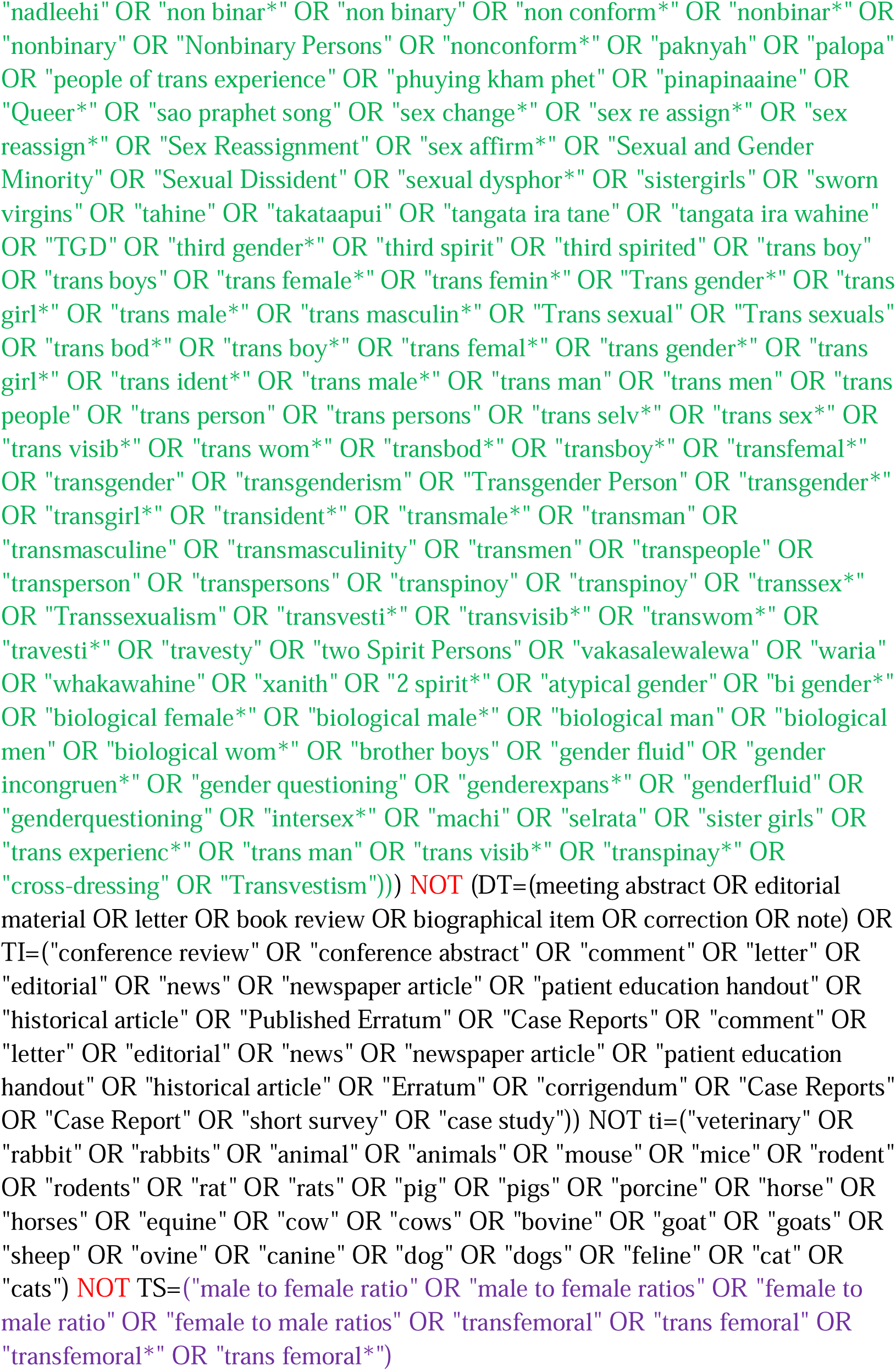

#### Cochrane CENTRAL

https://www.cochranelibrary.com/advanced-search/search-manager

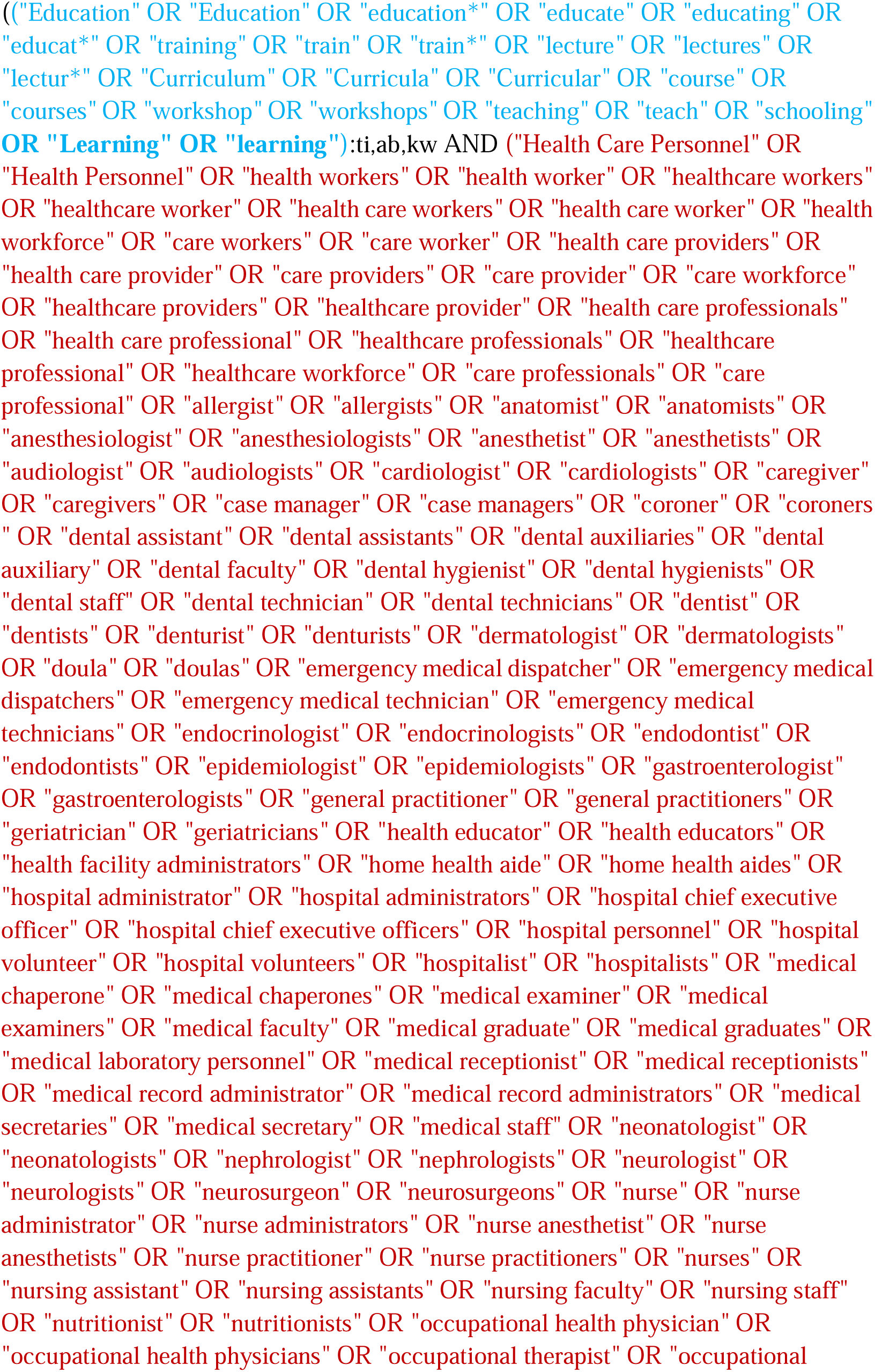
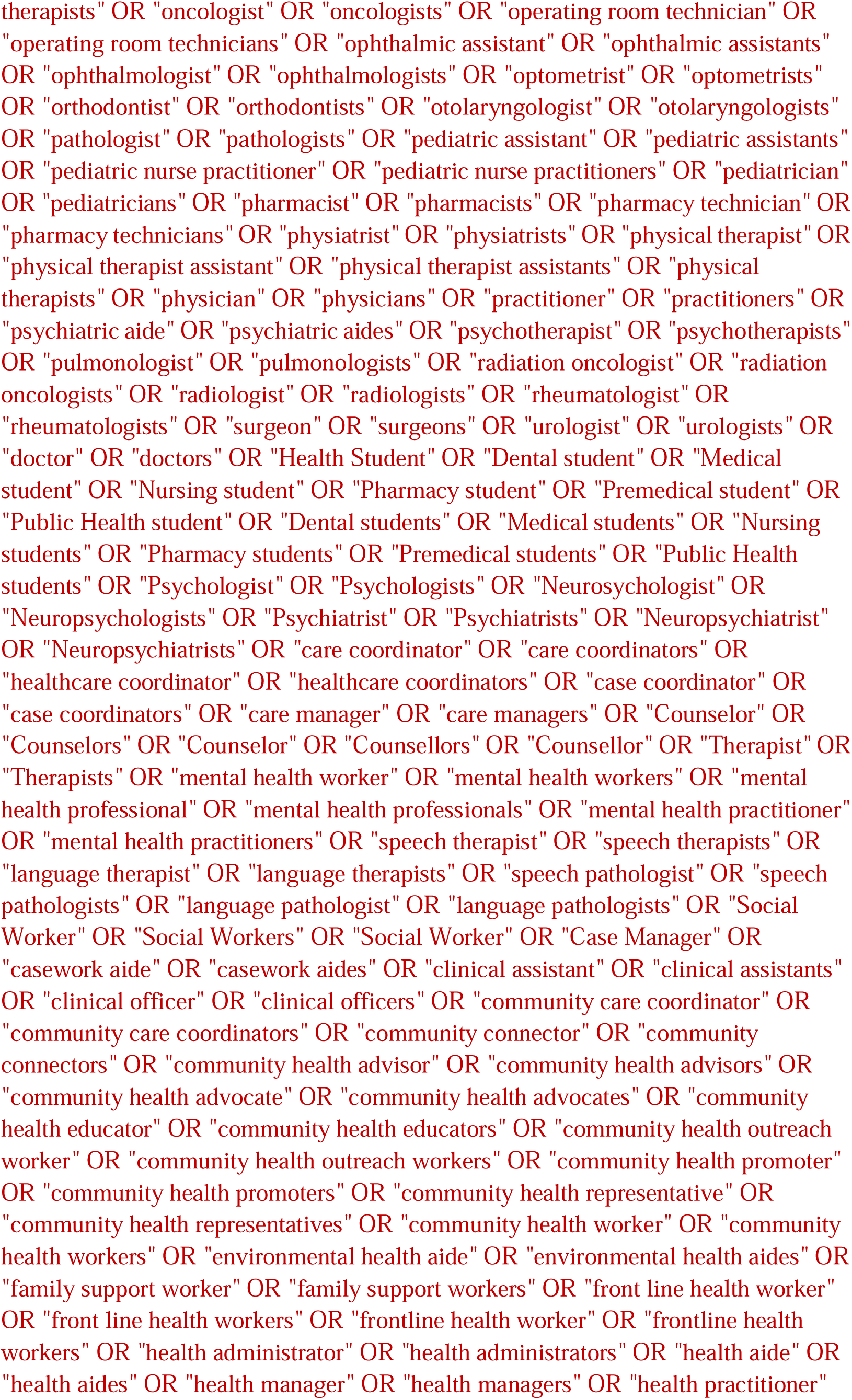
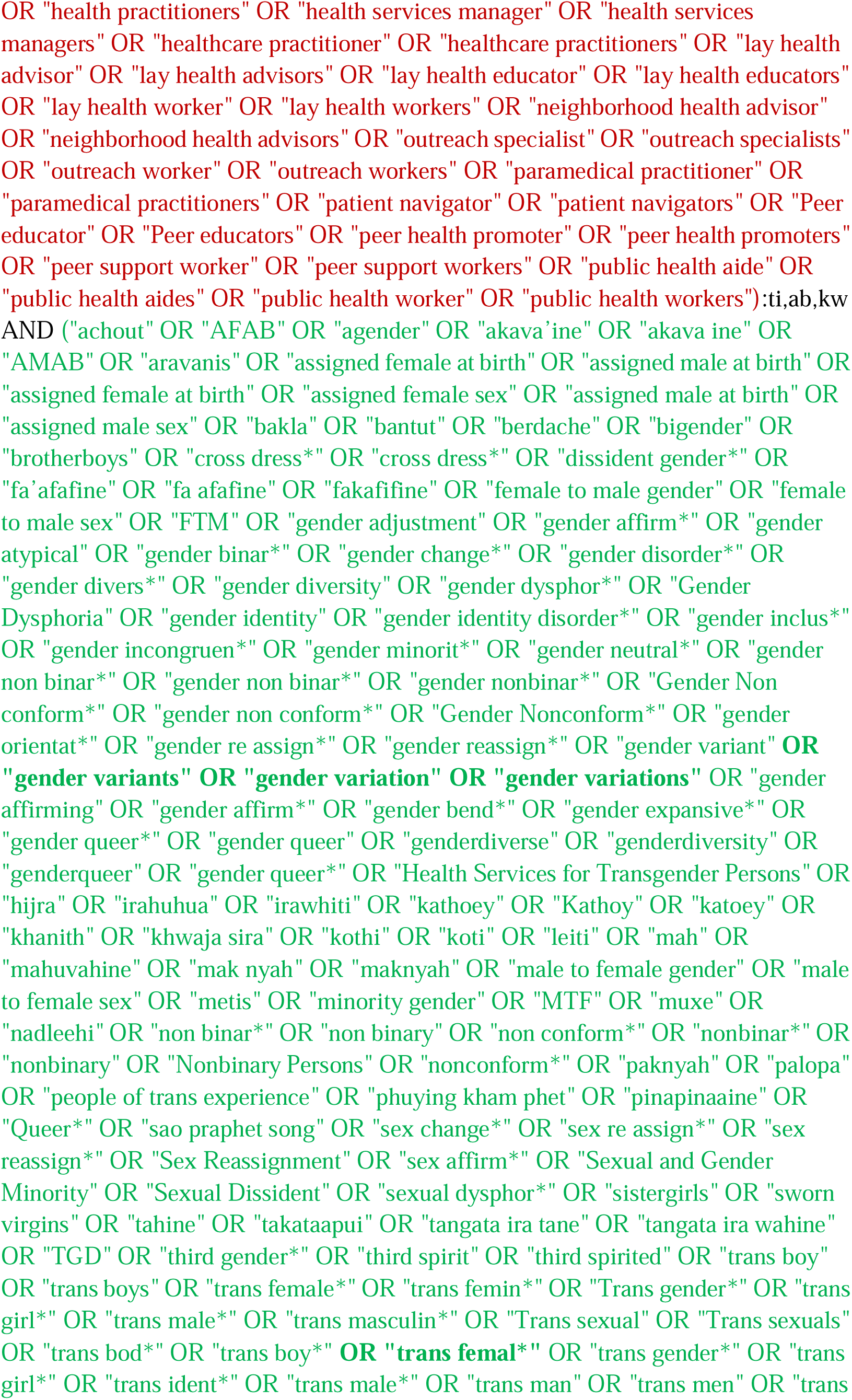
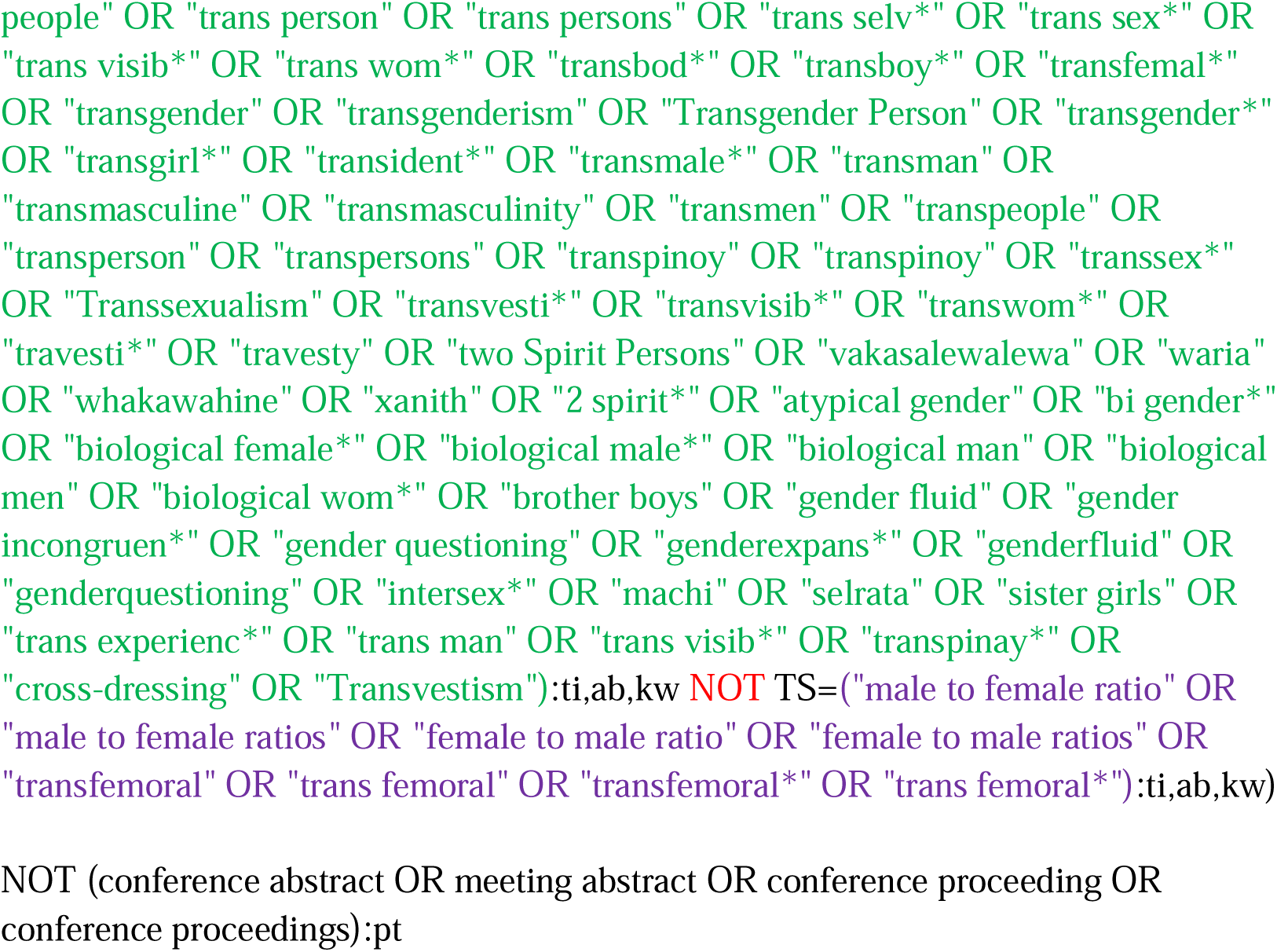

#### Emcare (OVID)

http://ovidsp.ovid.com/ovidweb.cgi?T=JS&NEWS=n&CSC=Y&PAGE=main&D=emcr

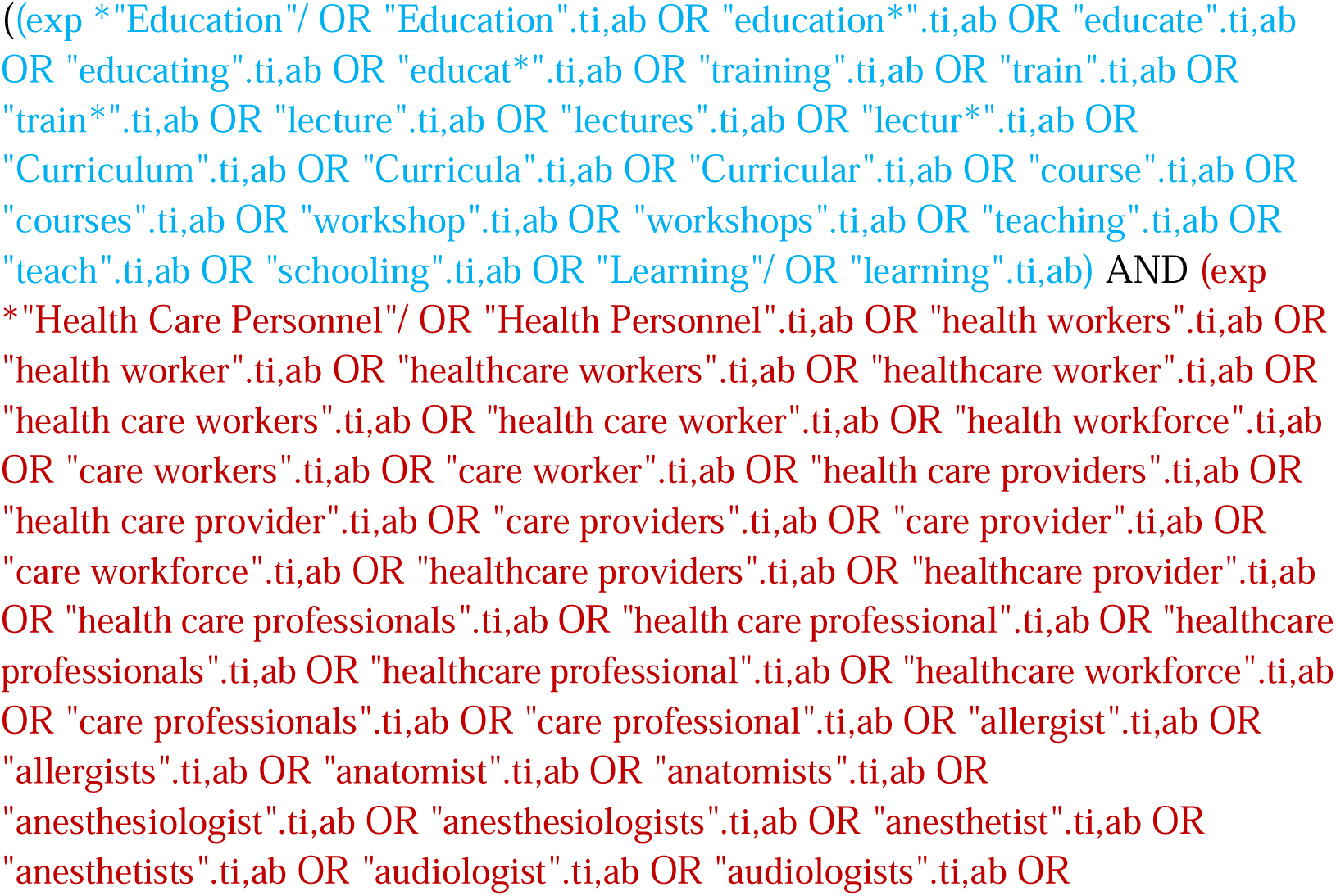
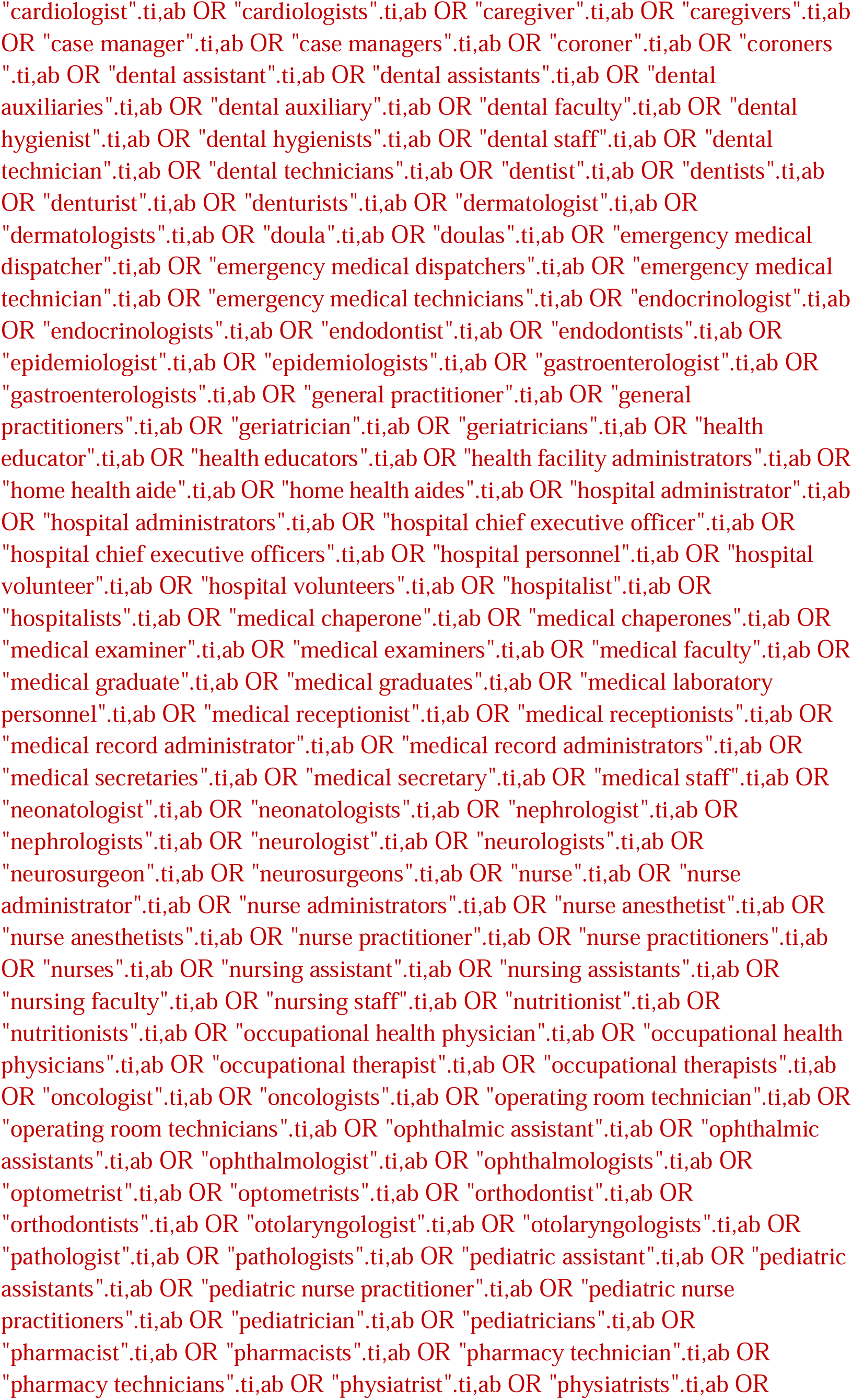
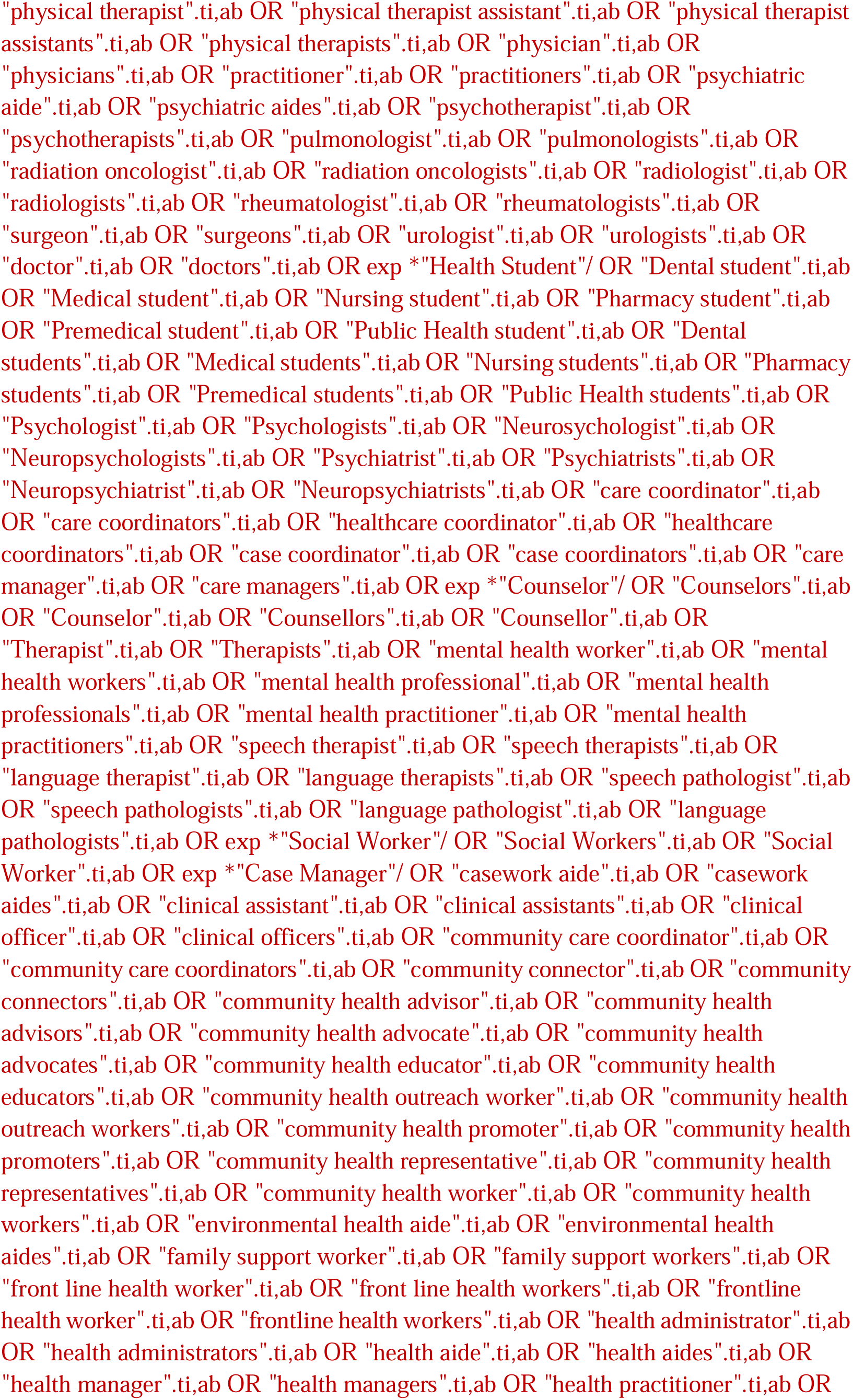
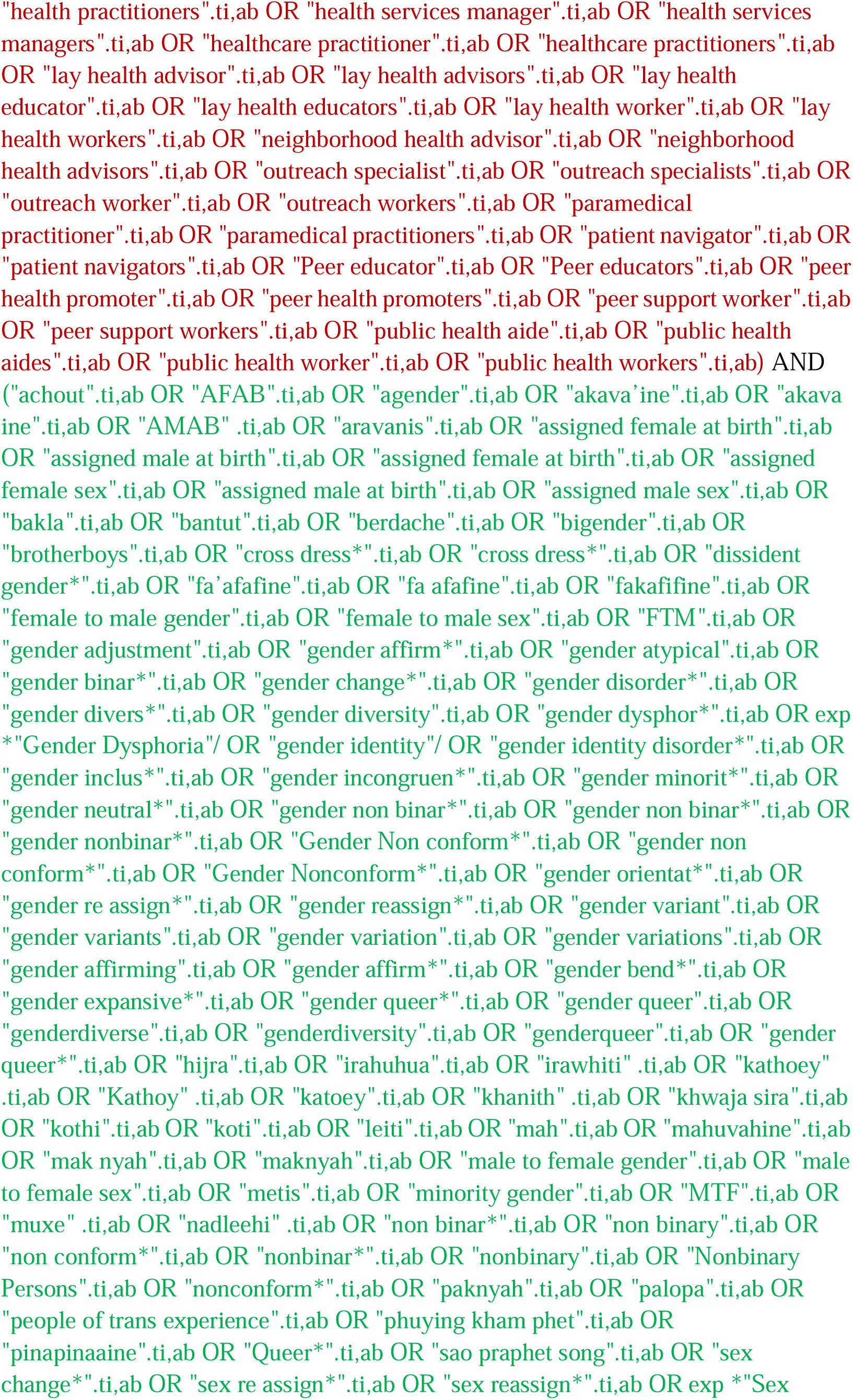
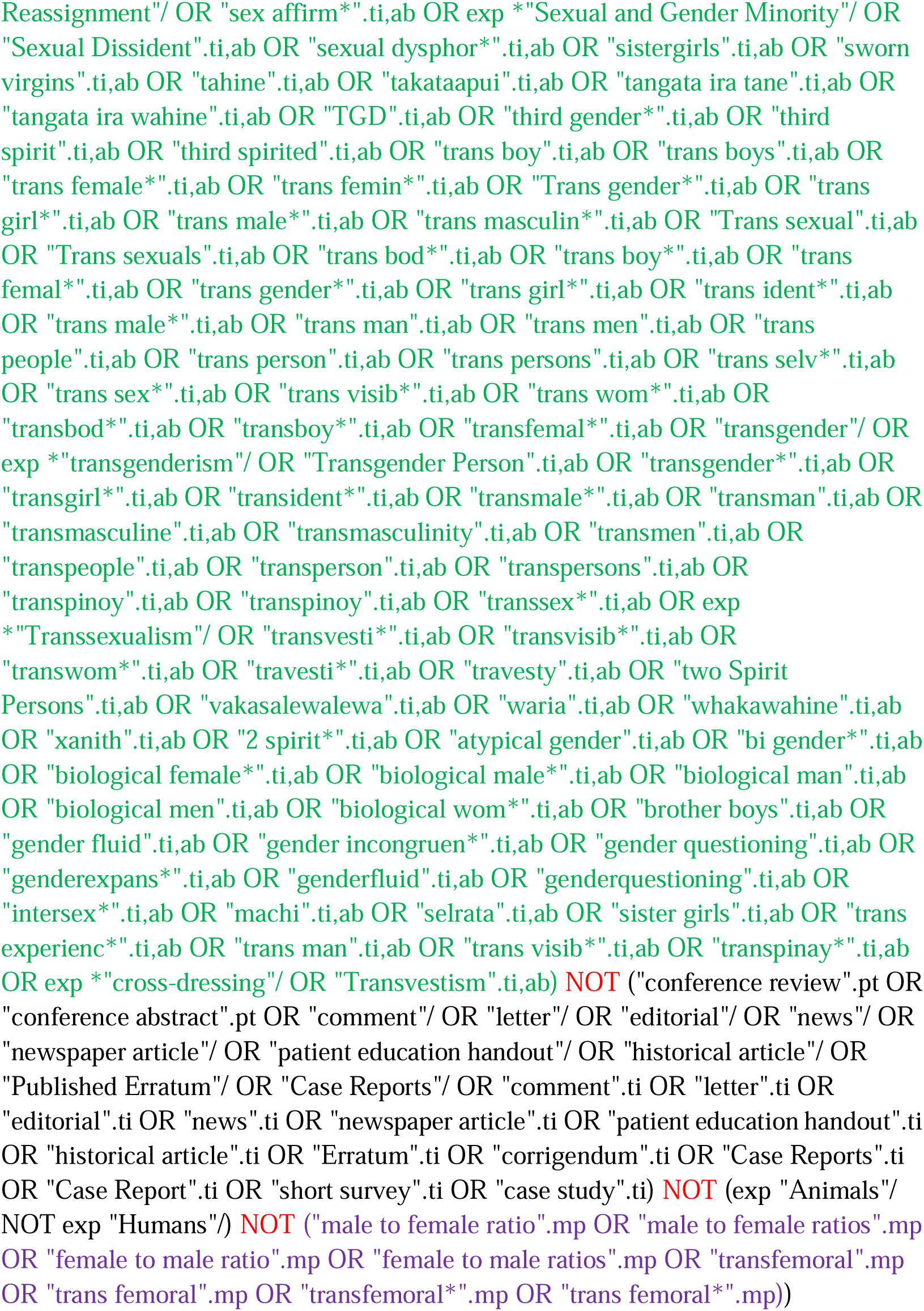

#### PsycINFO (EBSCOhost)

http://search.EBSCOhost.com/login.aspx?authtype=ip,uid&profile=lumc&defaultdb=psyh

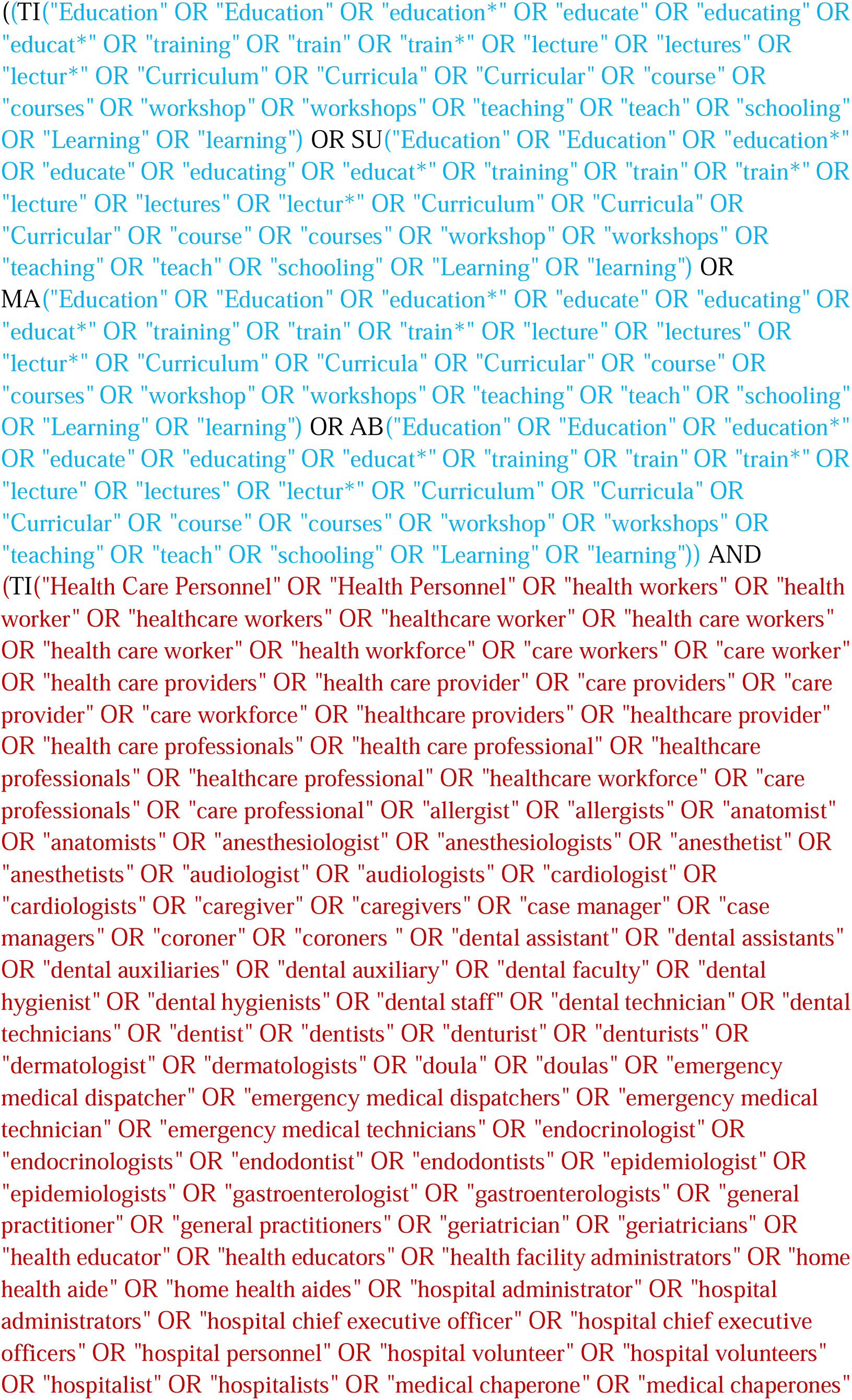
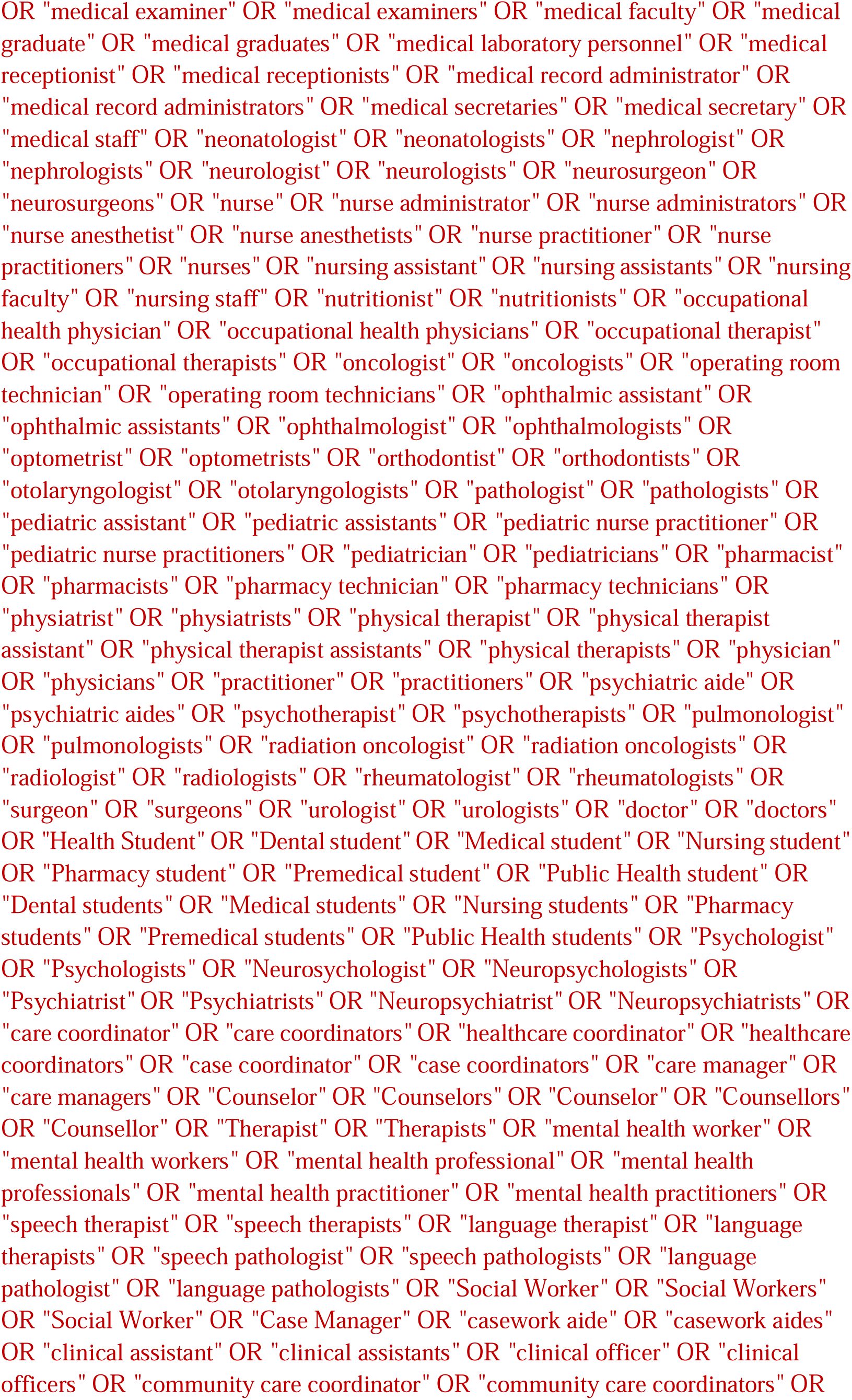
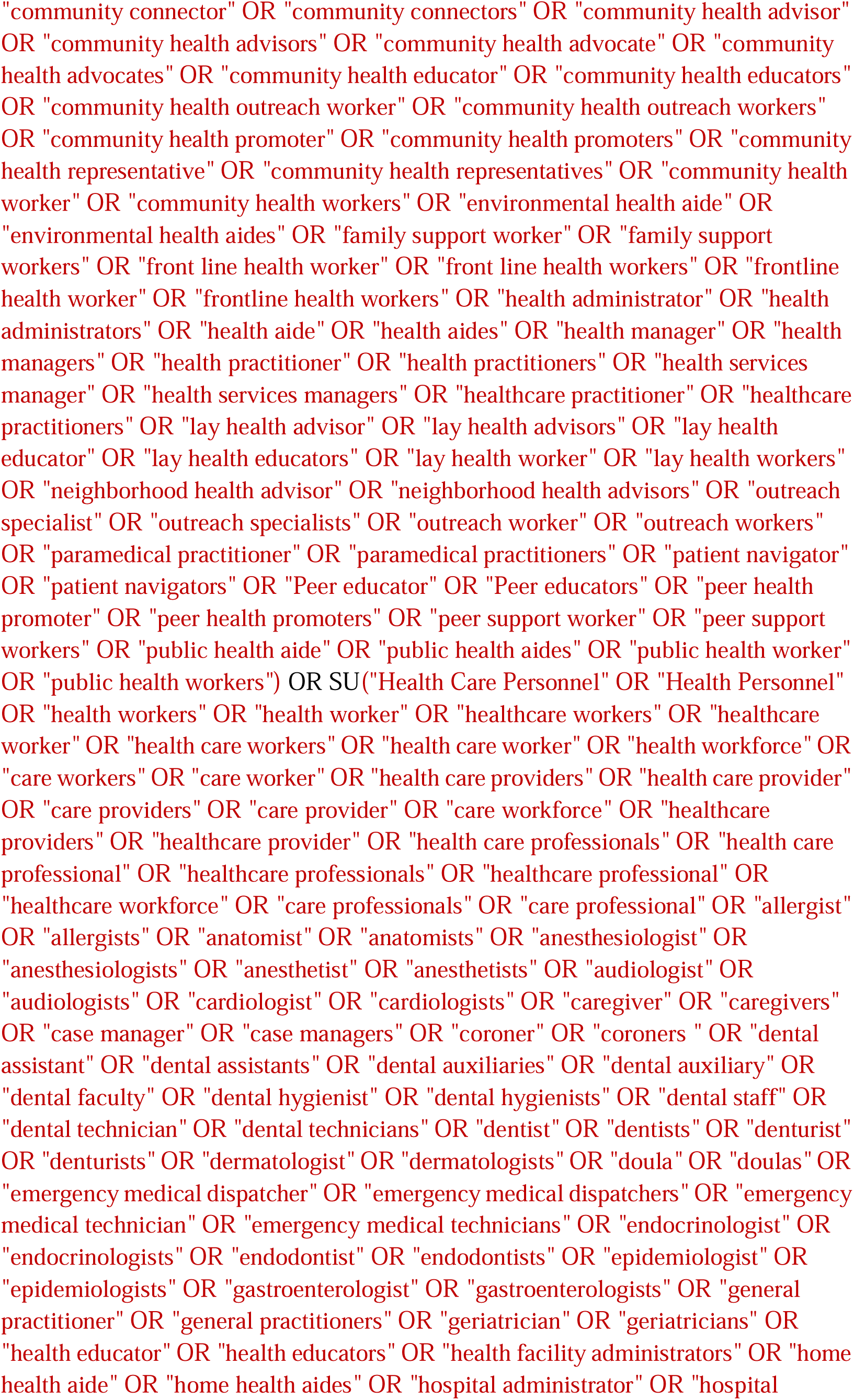
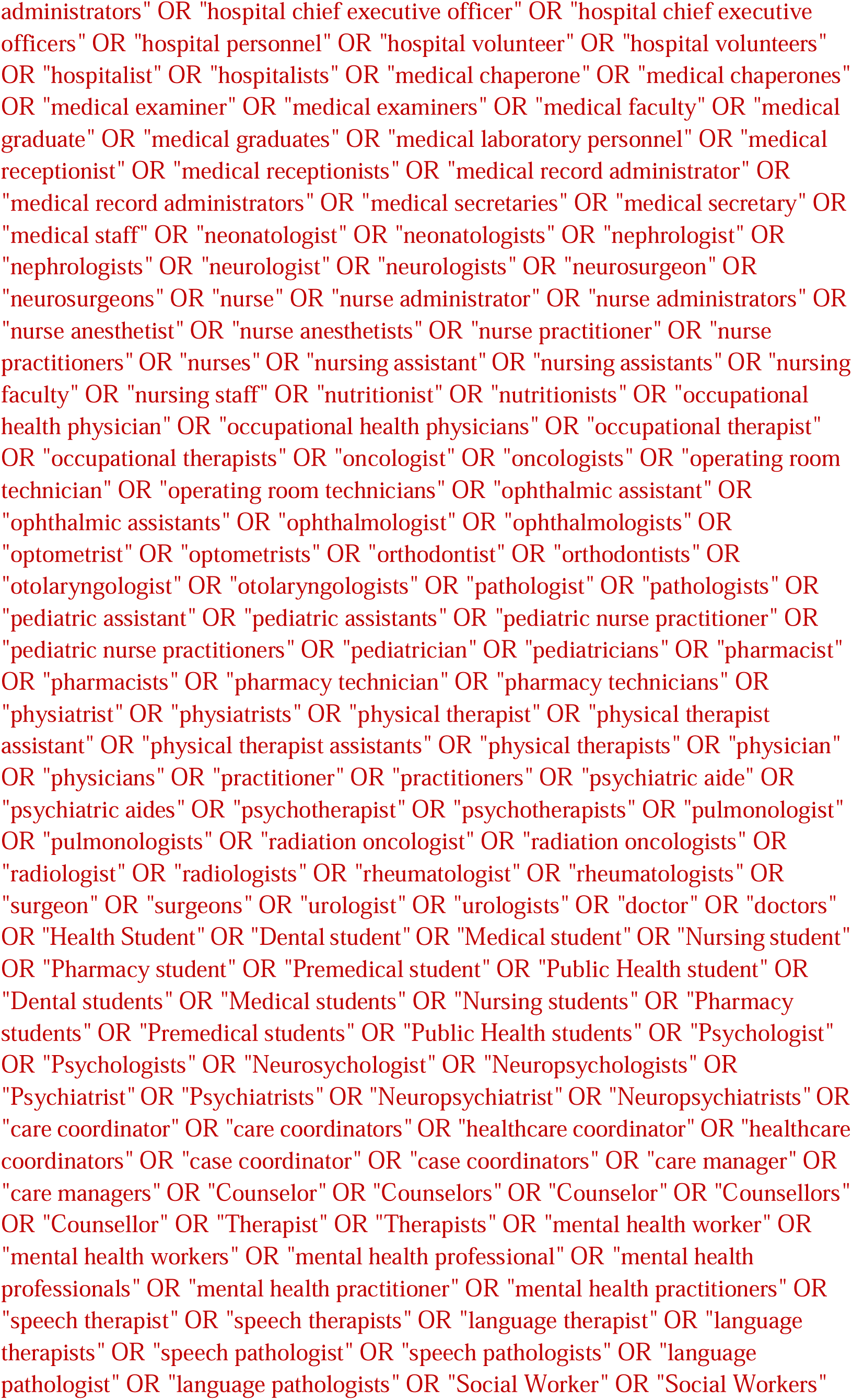
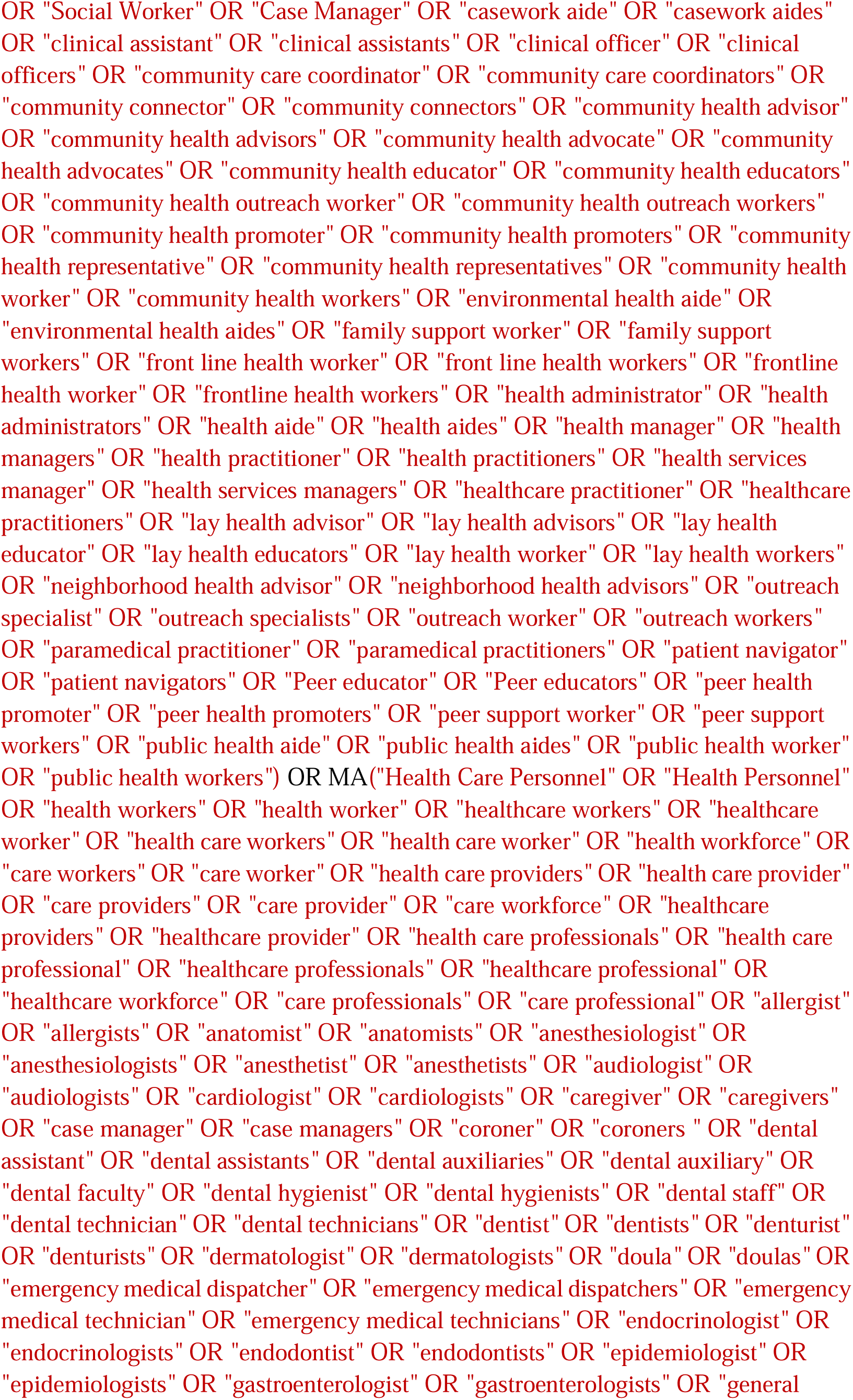
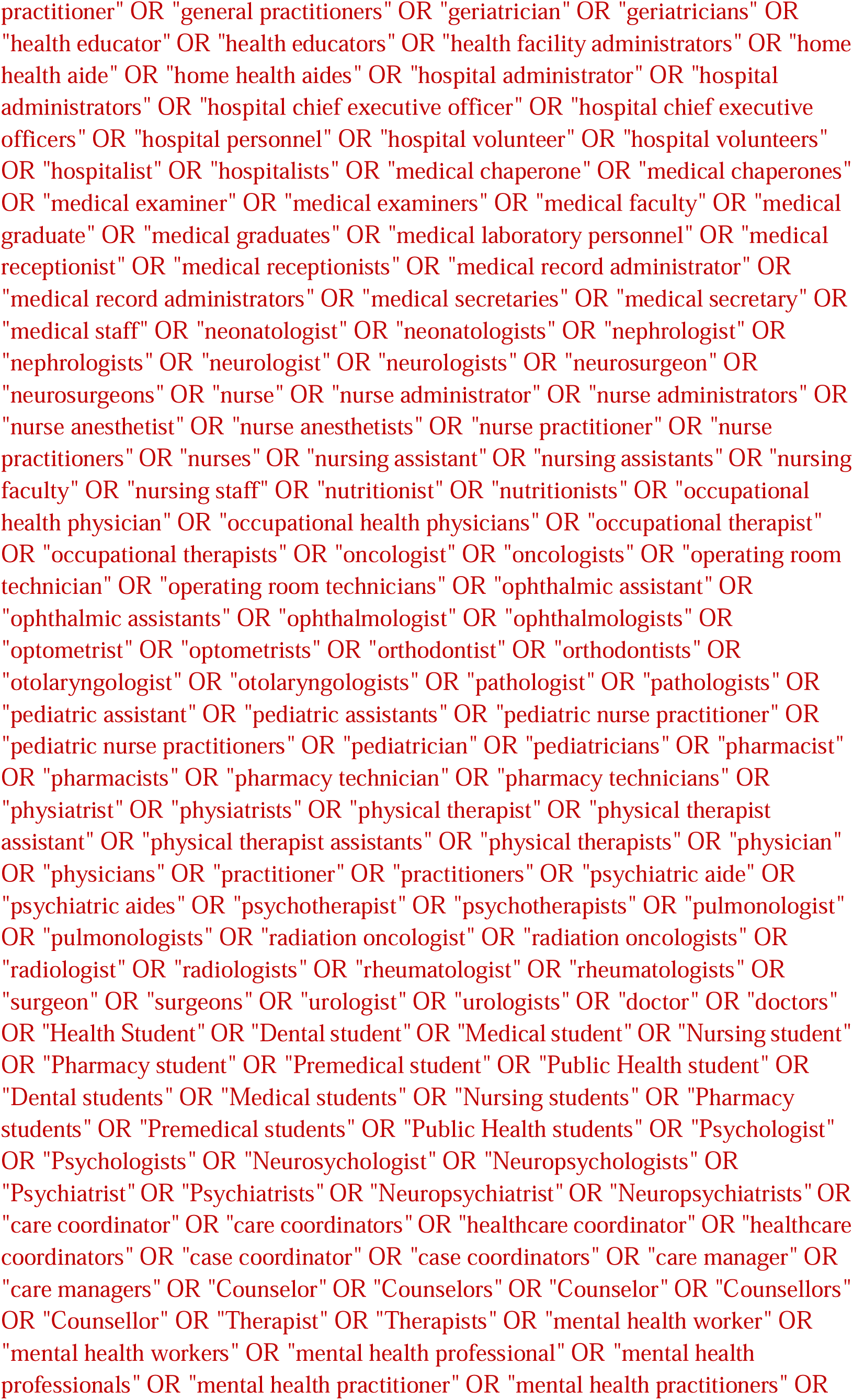
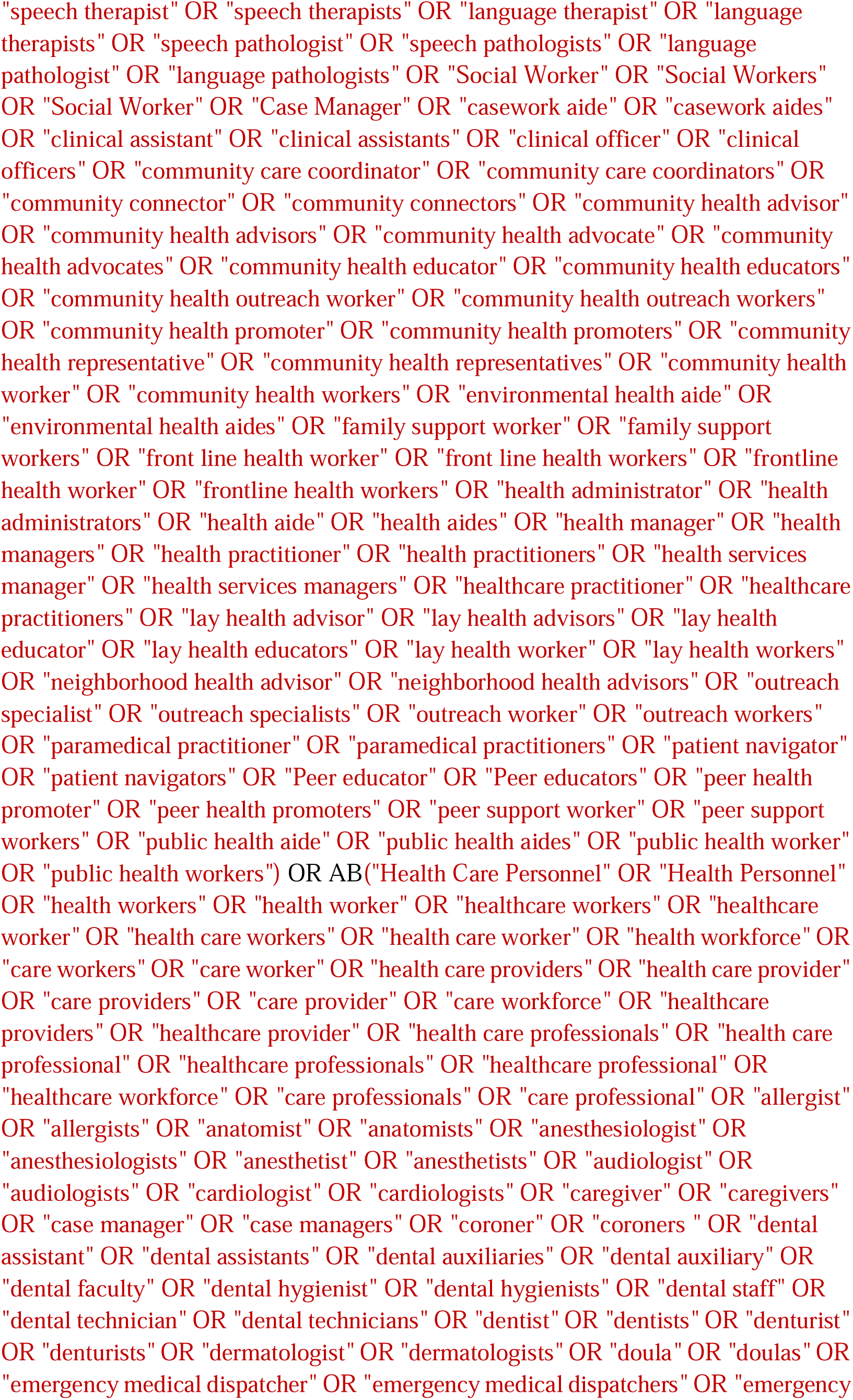
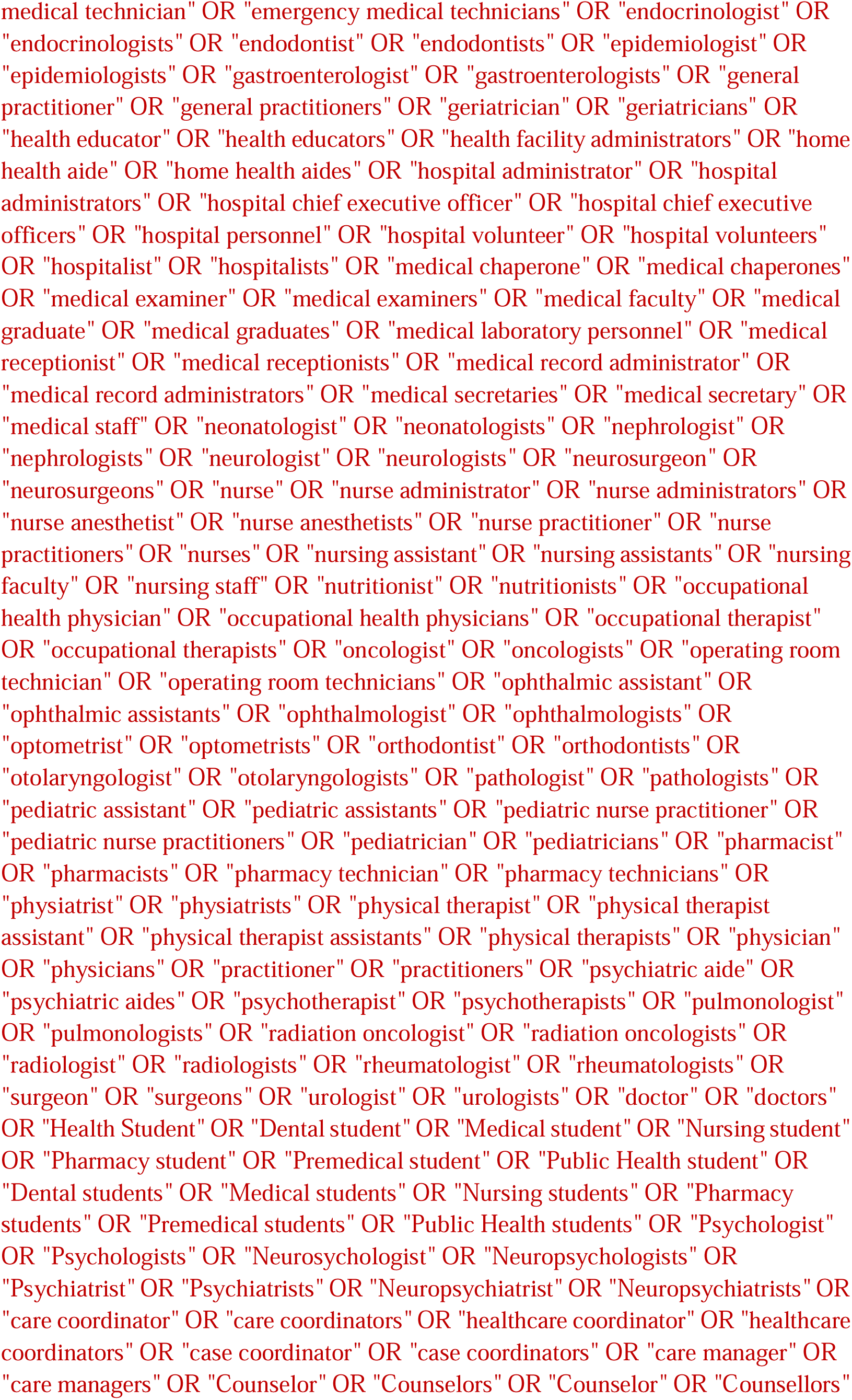
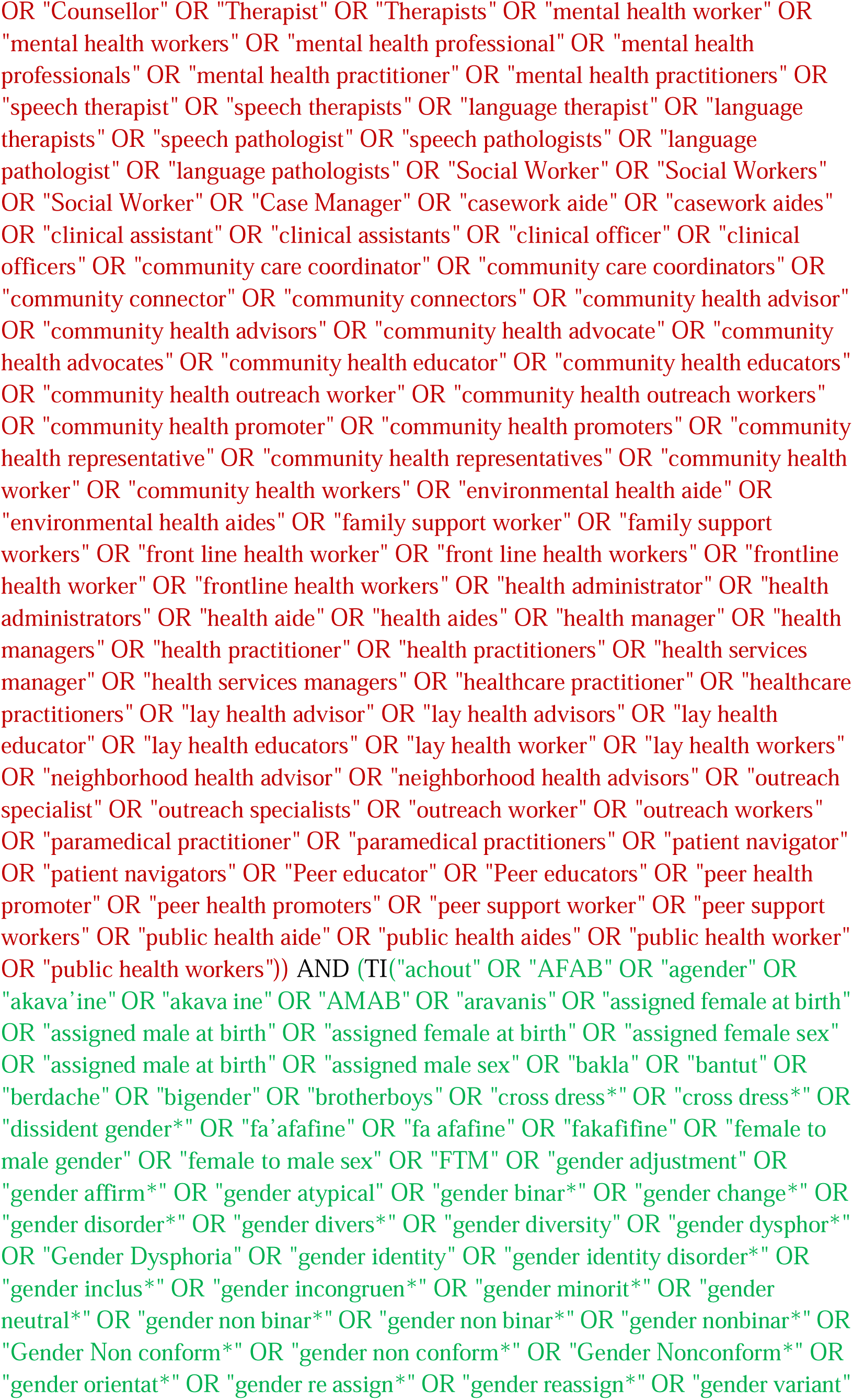
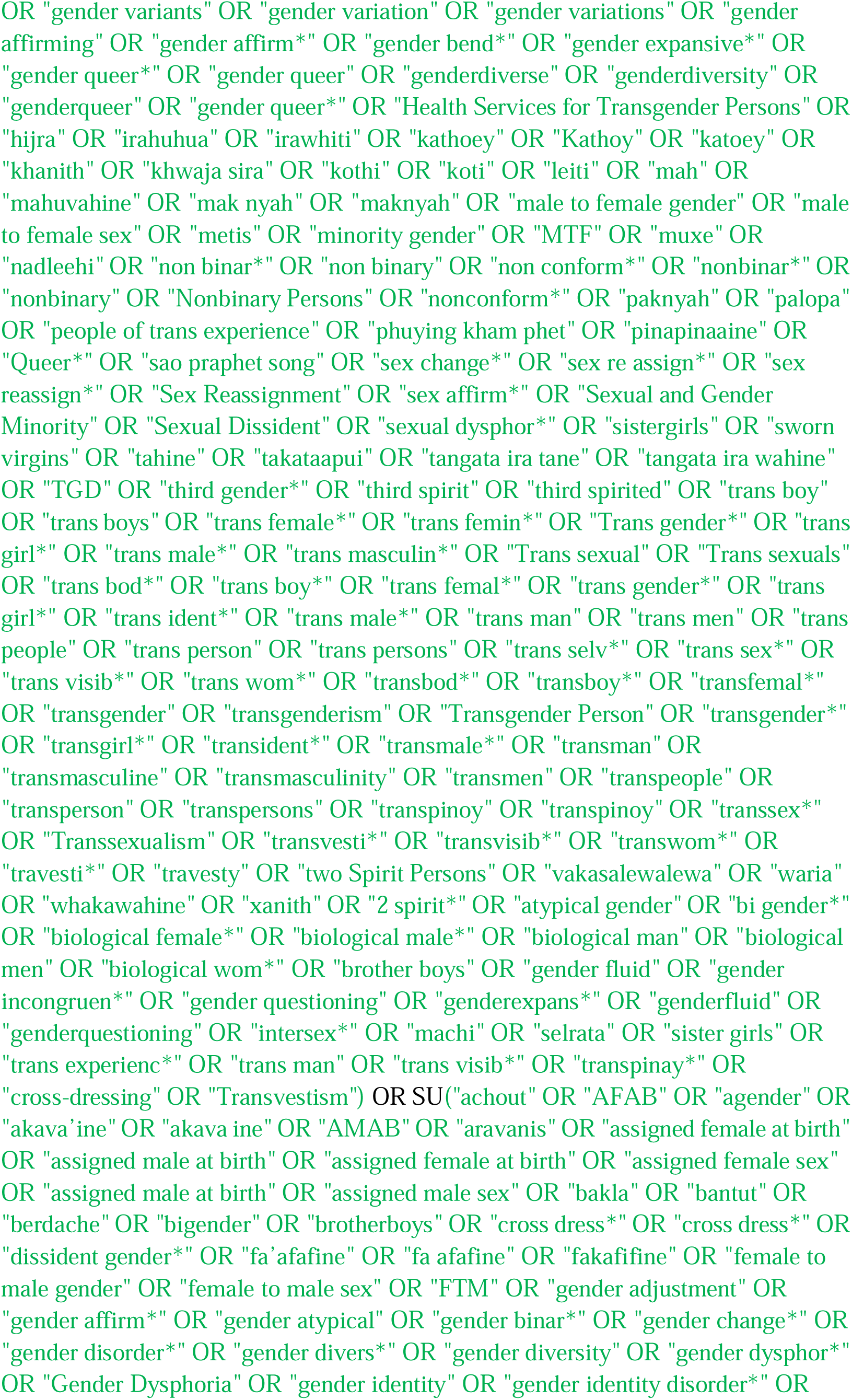
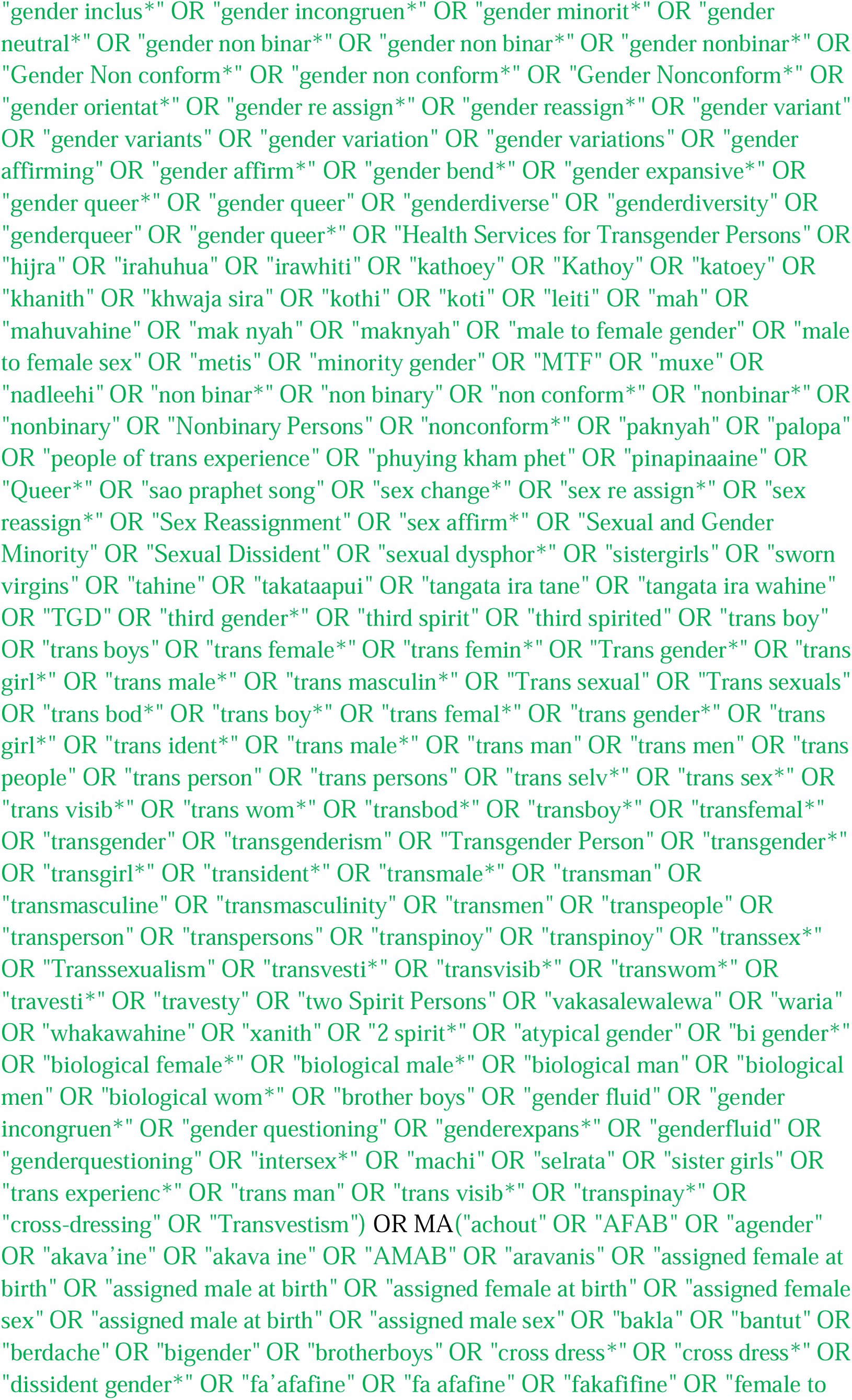
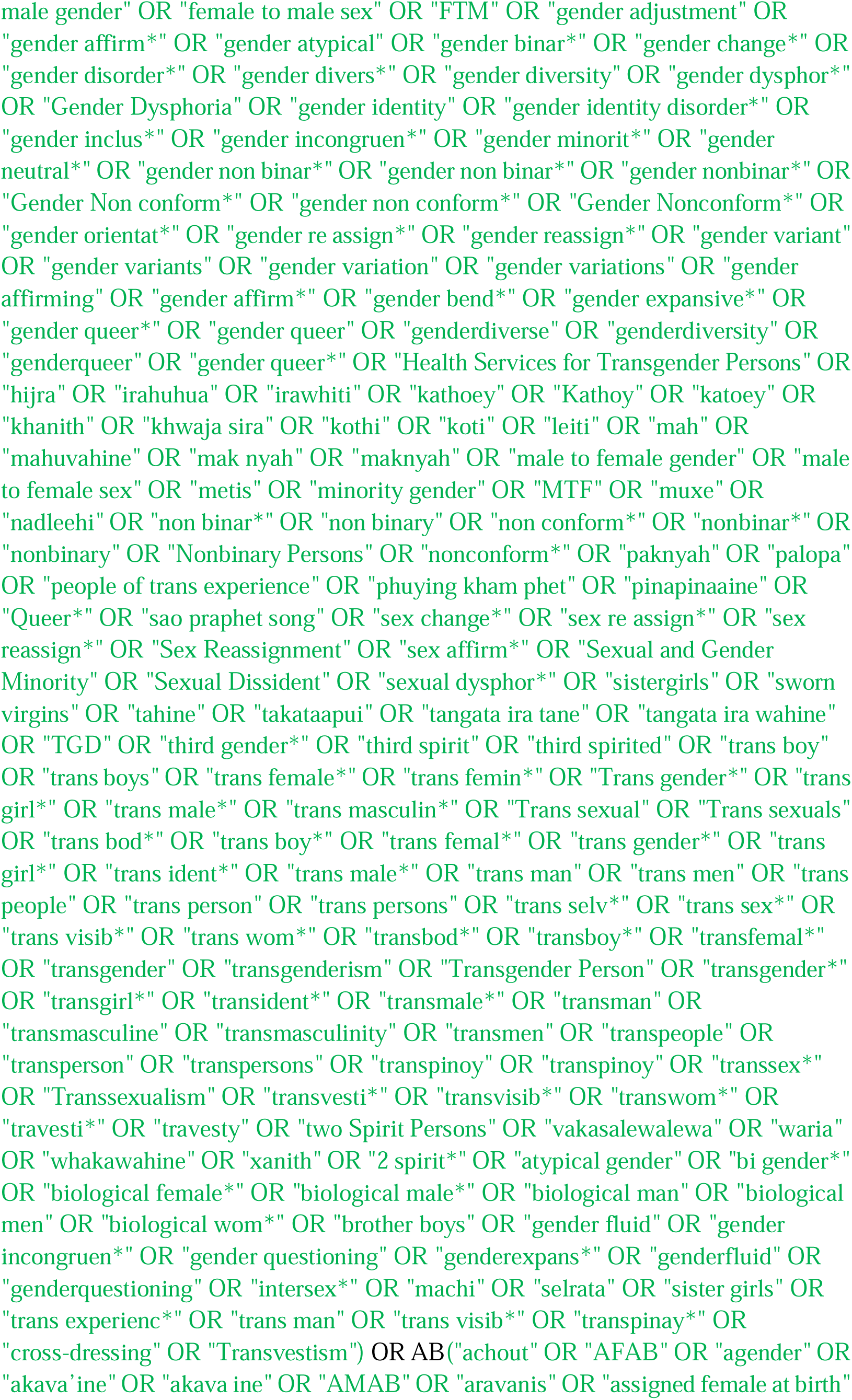
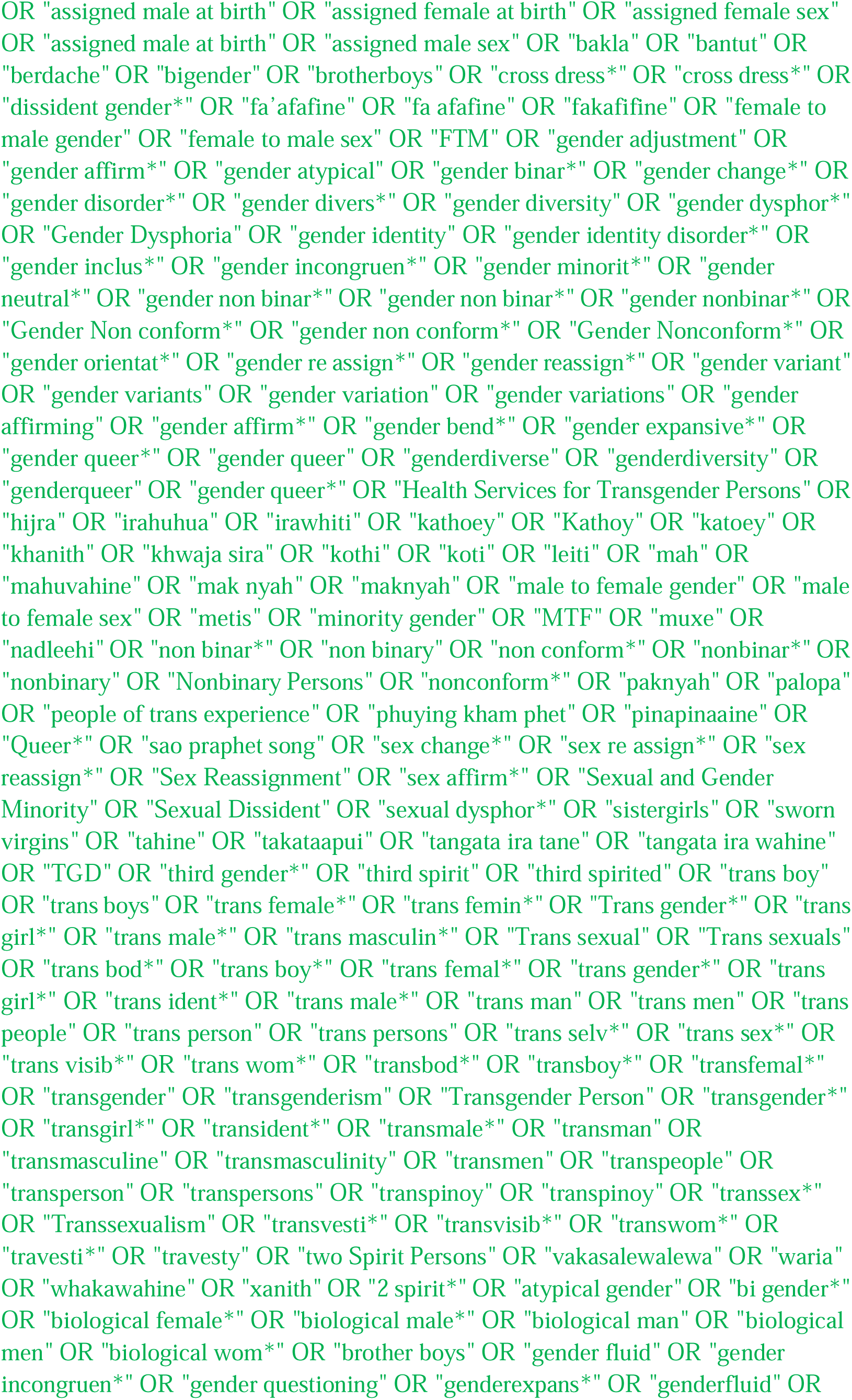
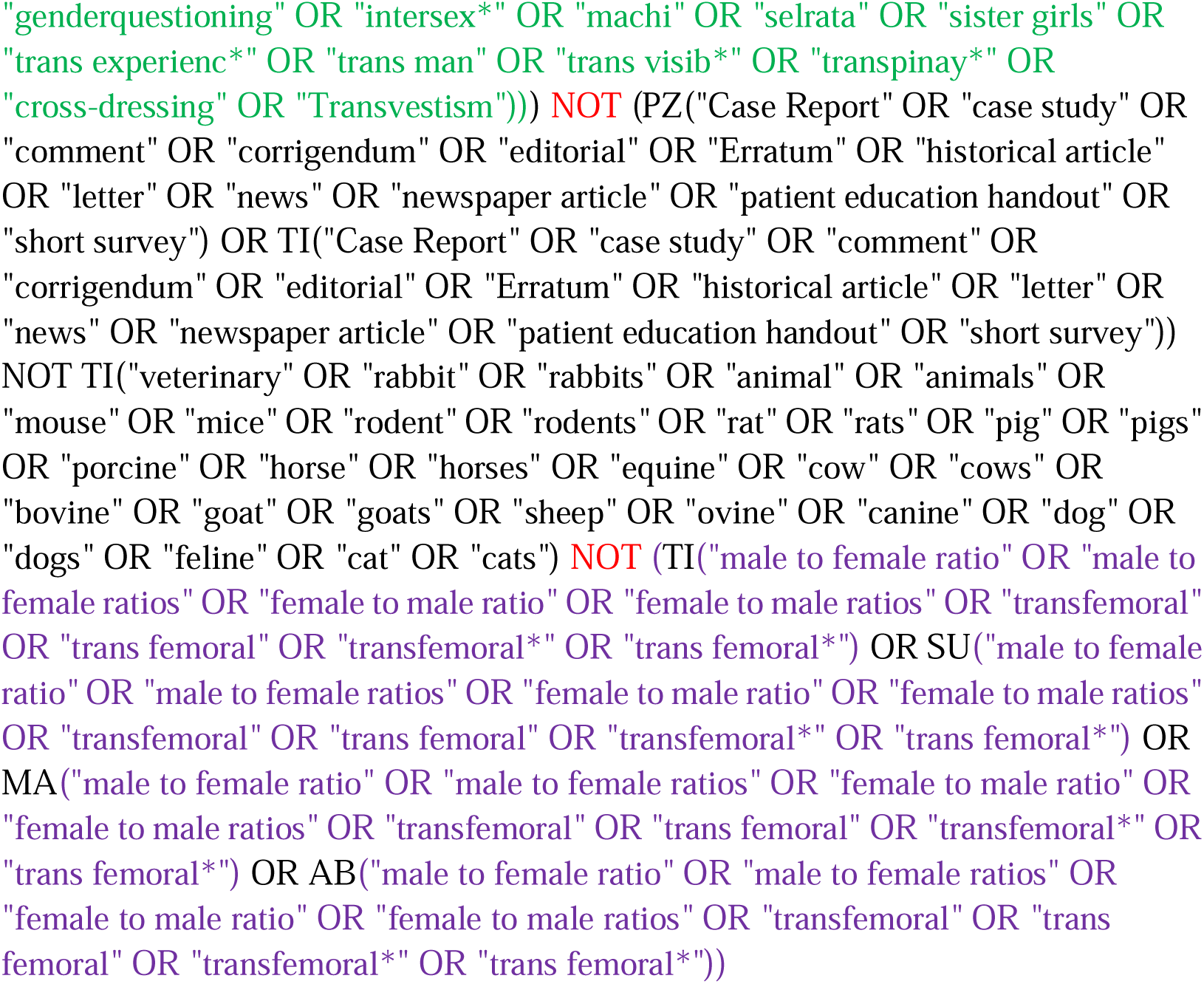

#### Social Services Abstracts

https://search.proquest.com/socialservices?accountid=12045

http://databases.library.leiden.edu/?bibid=990027333030302711&redirect=true

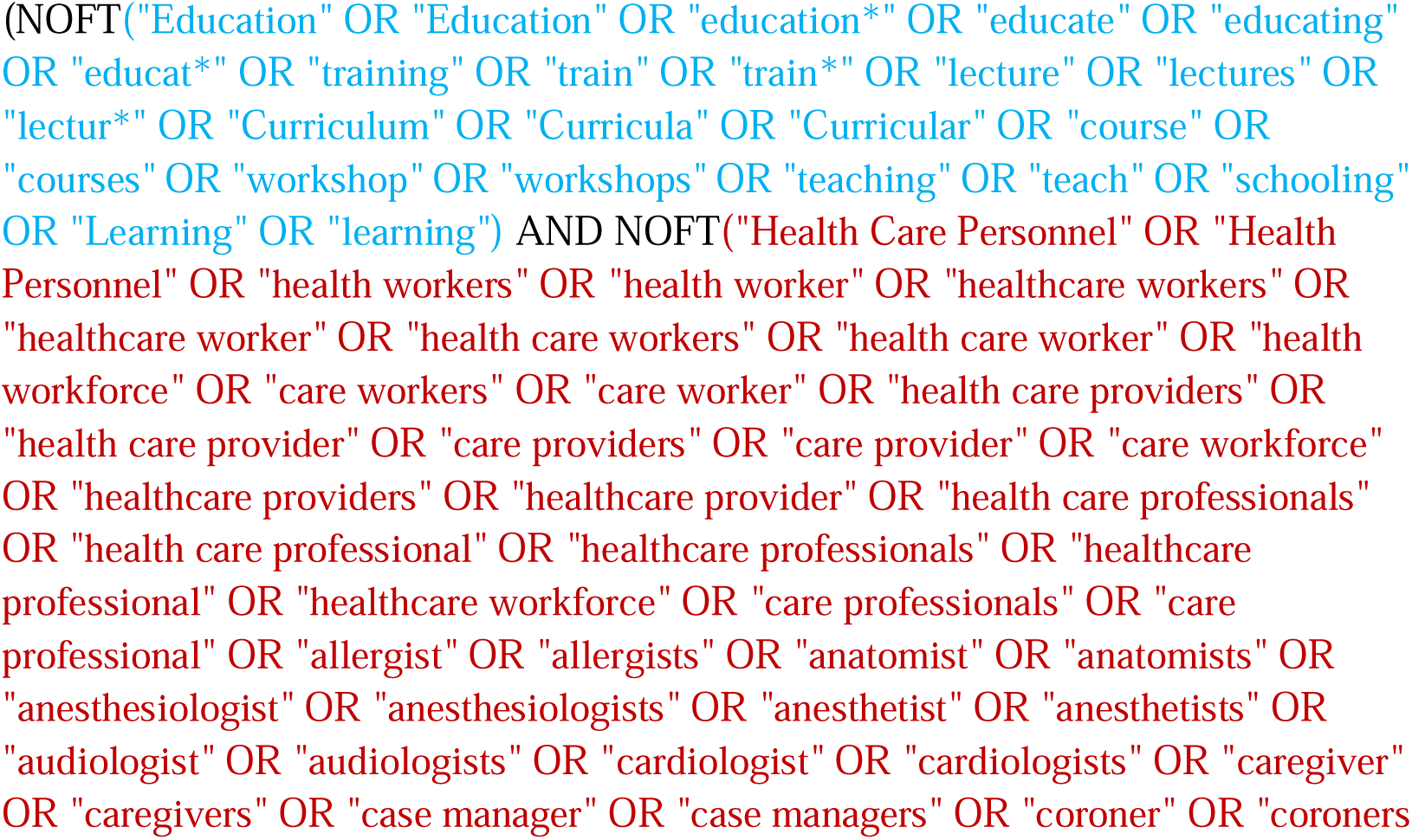
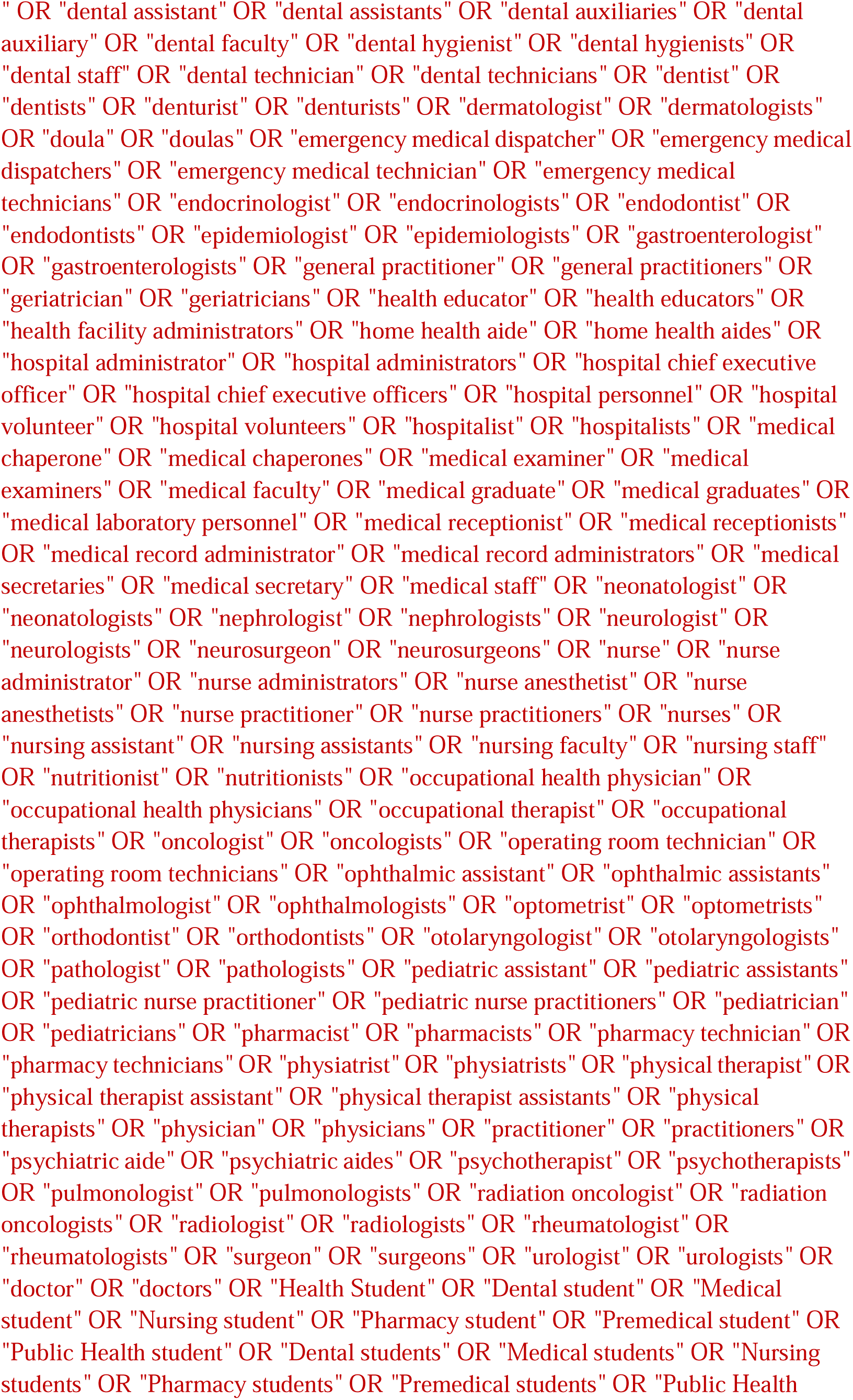
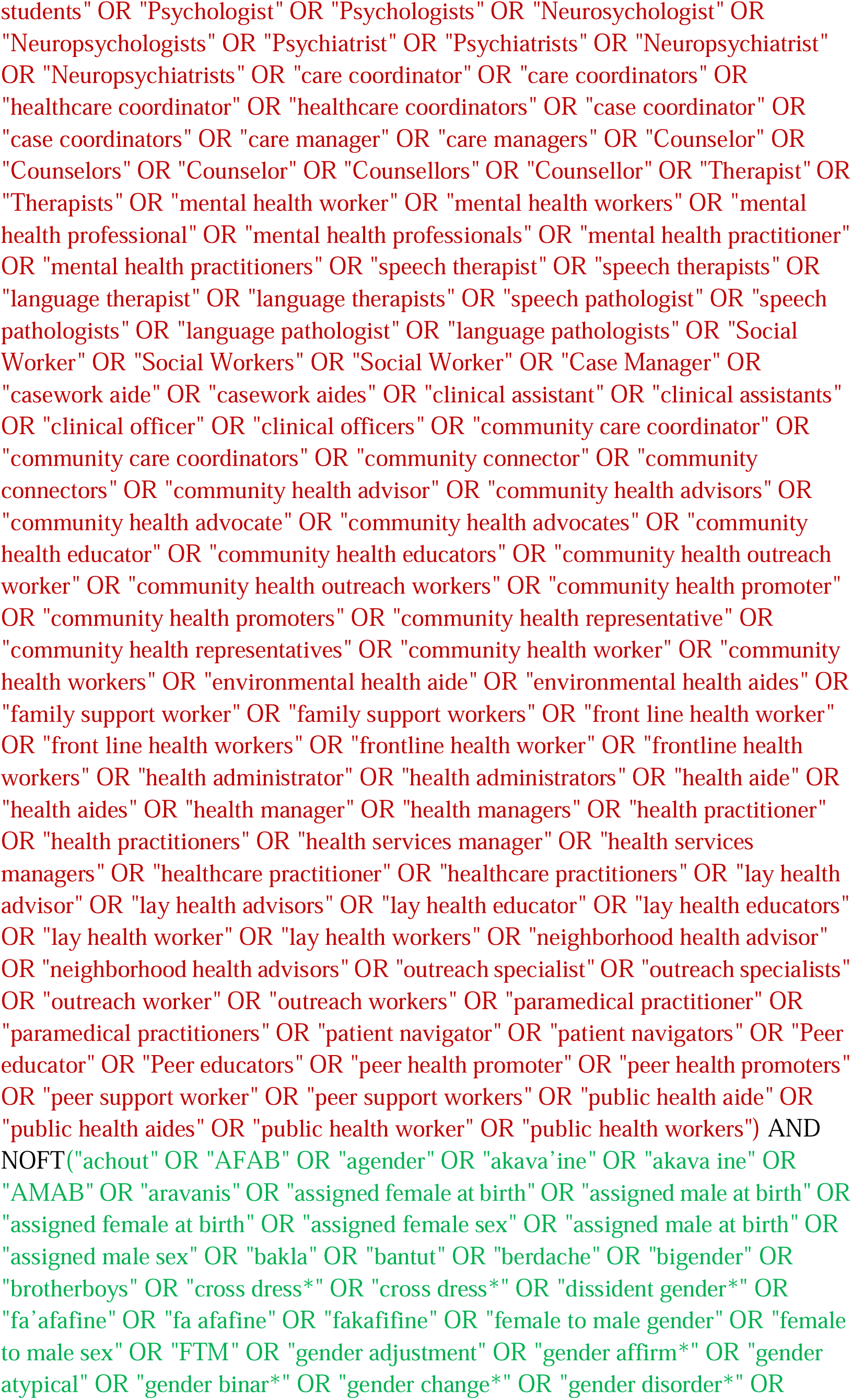
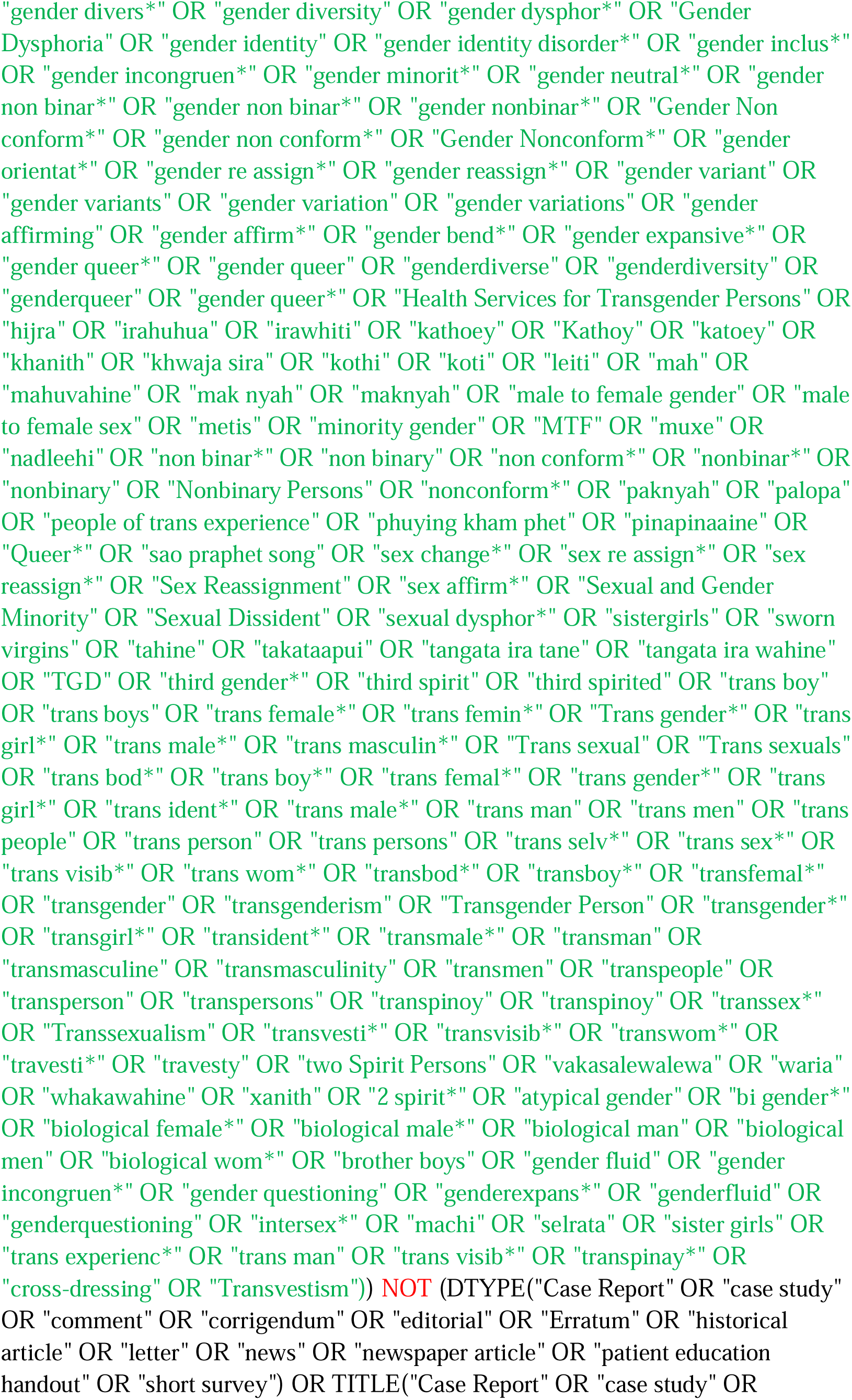
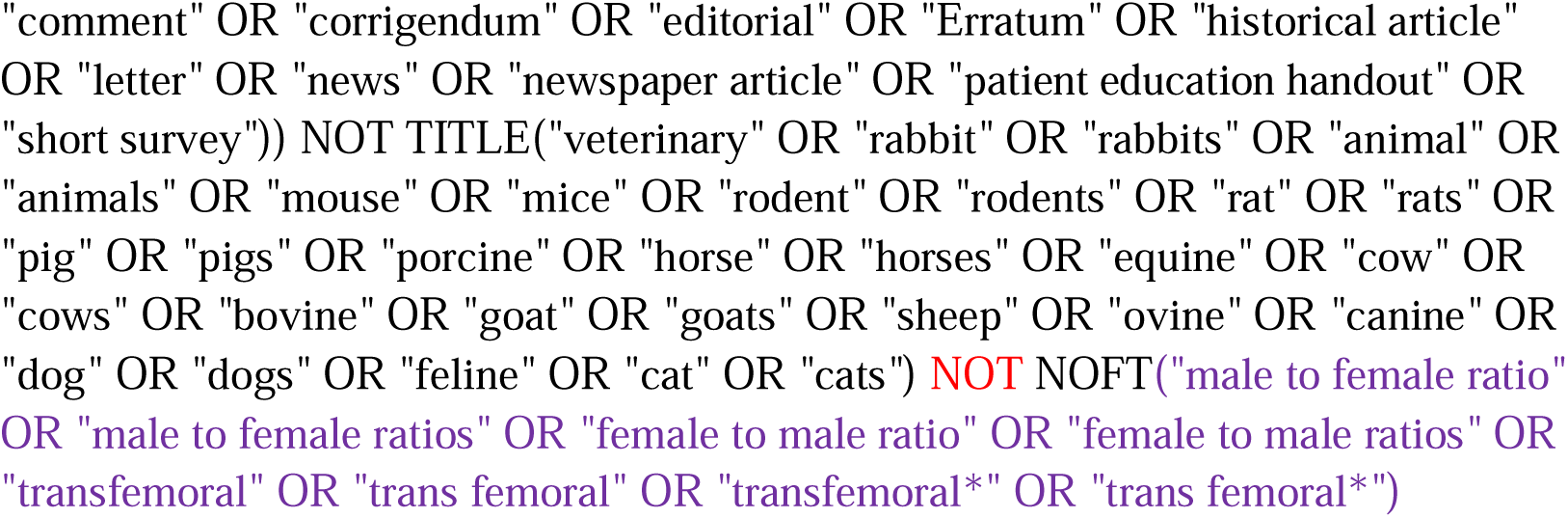

#### Sociological Abstracts

https://search.proquest.com/socabs/index

http://databases.library.leiden.edu/?bibid=990024621960302711&redirect=true

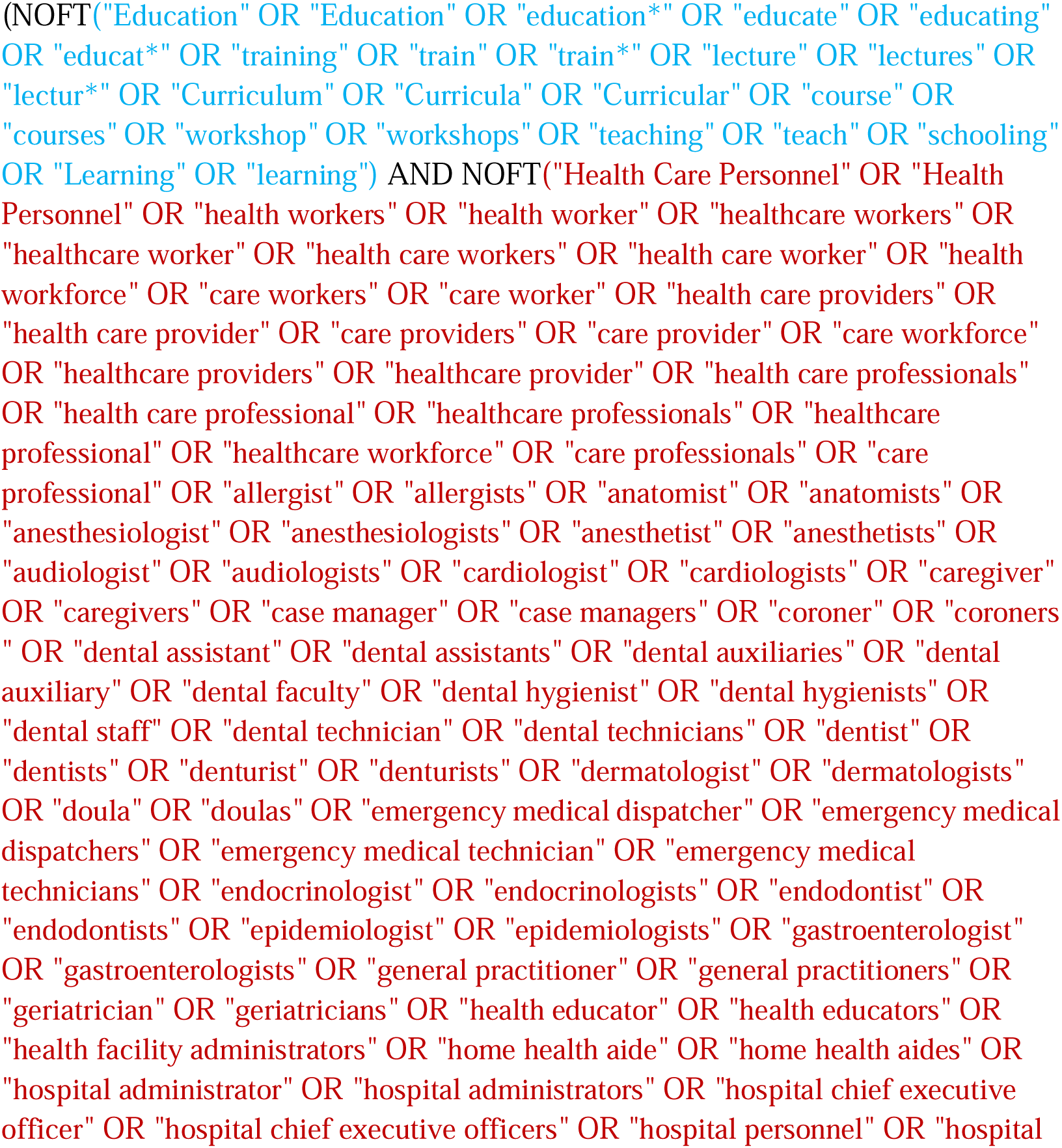
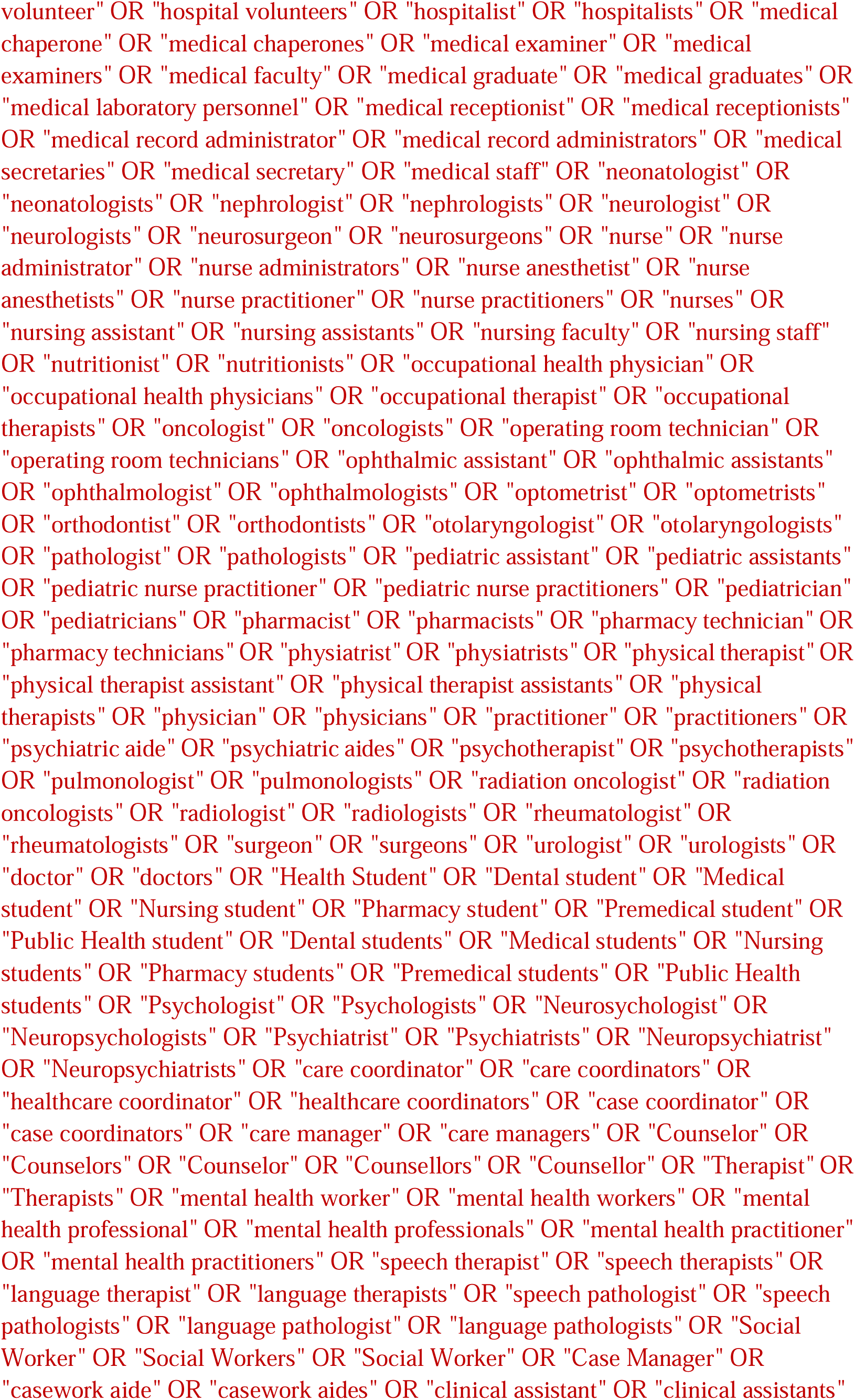
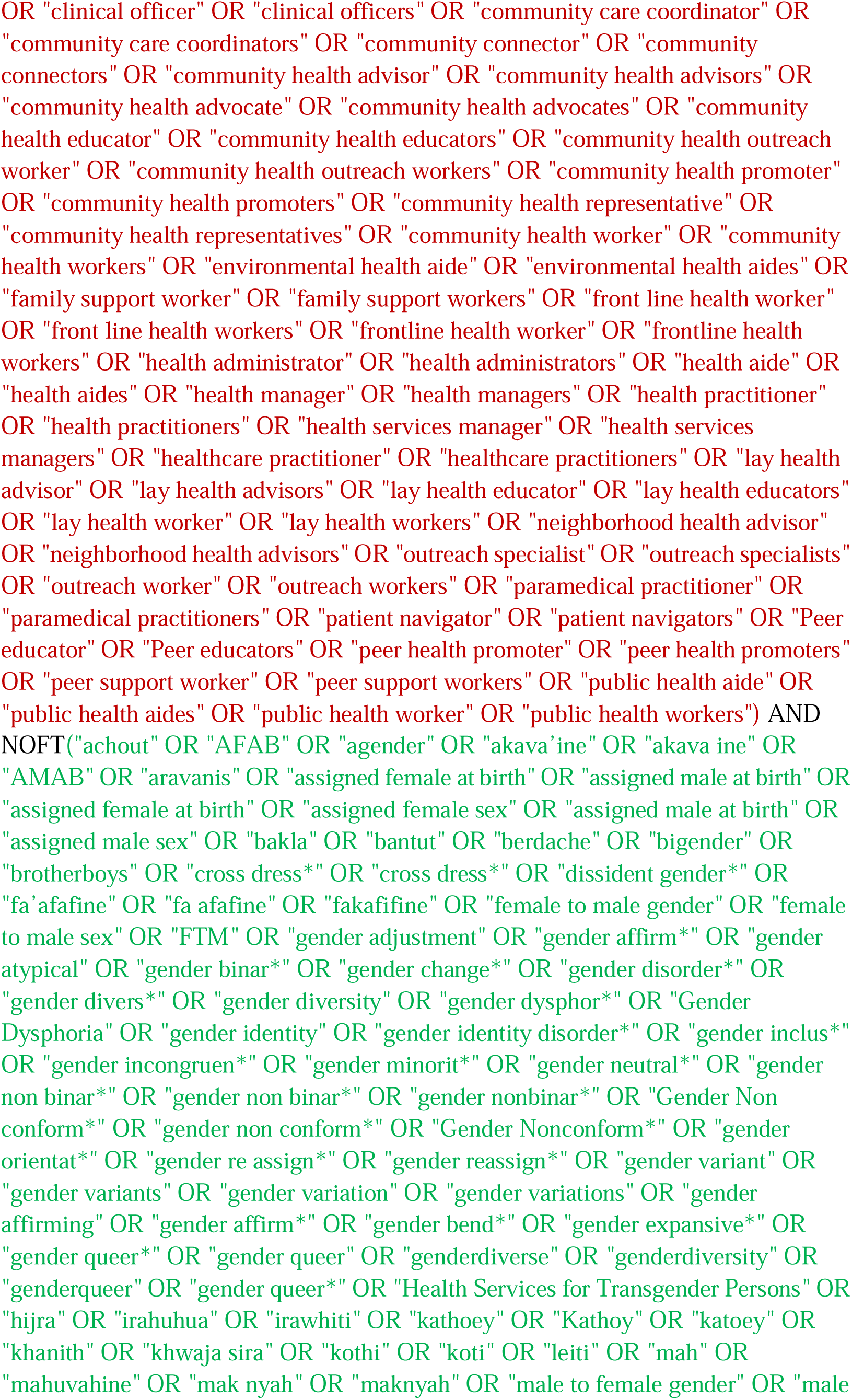
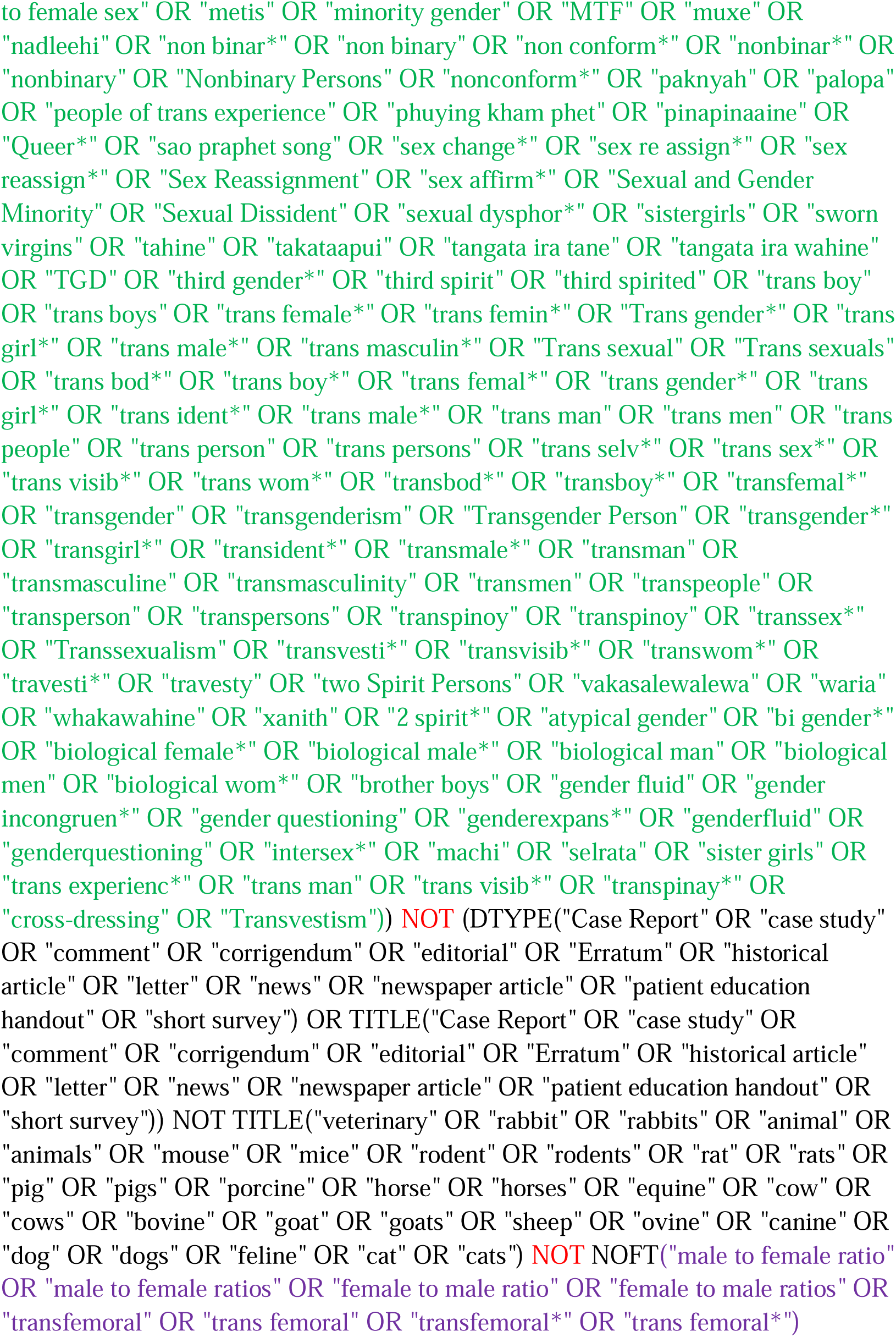

#### Academic Search Premier (EBSCOhost)

http://search.EBSCOhost.com/login.aspx?authtype=ip,uid&profile=lumc&defaultdb=aph

Limit to Academic Journals

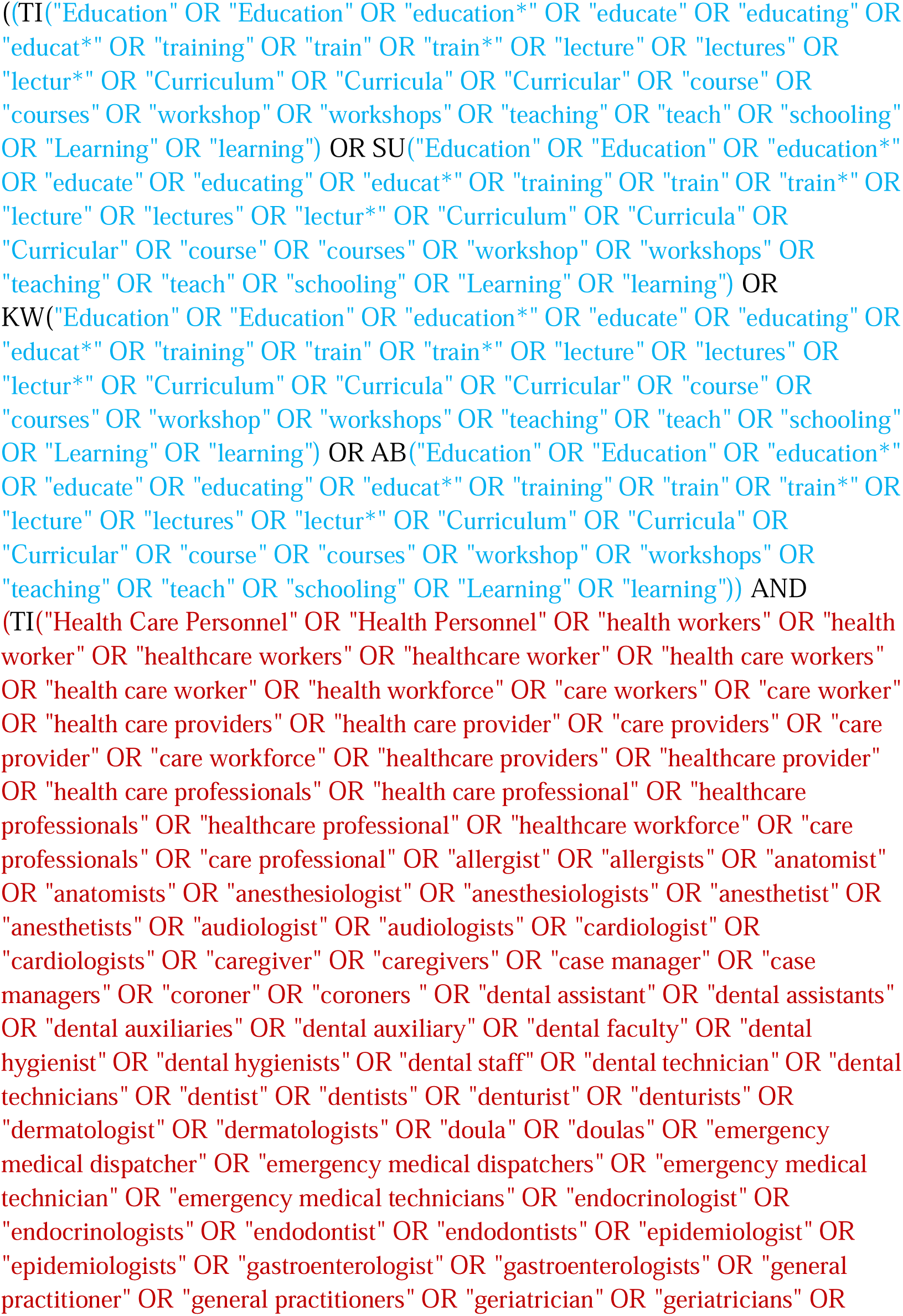
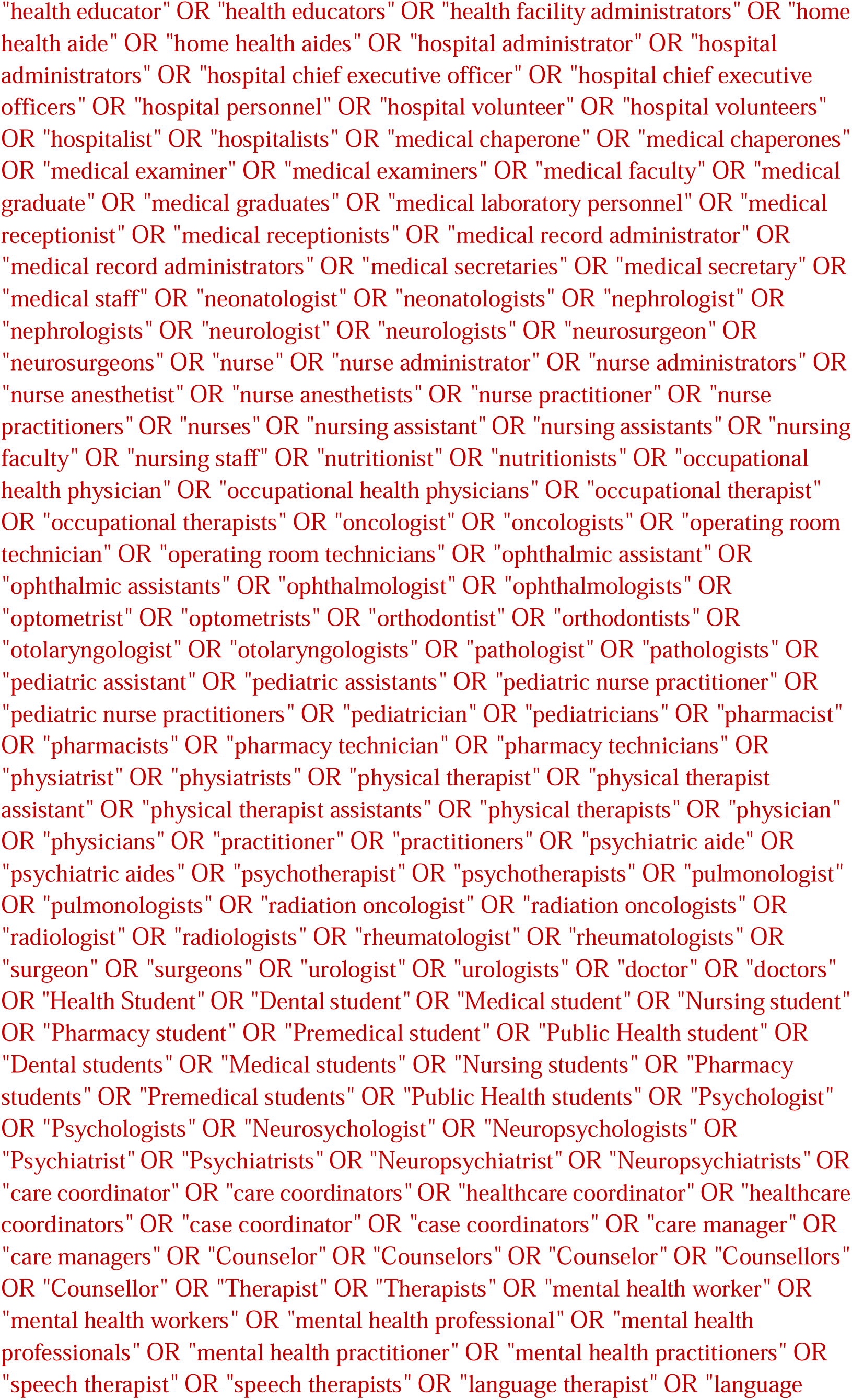
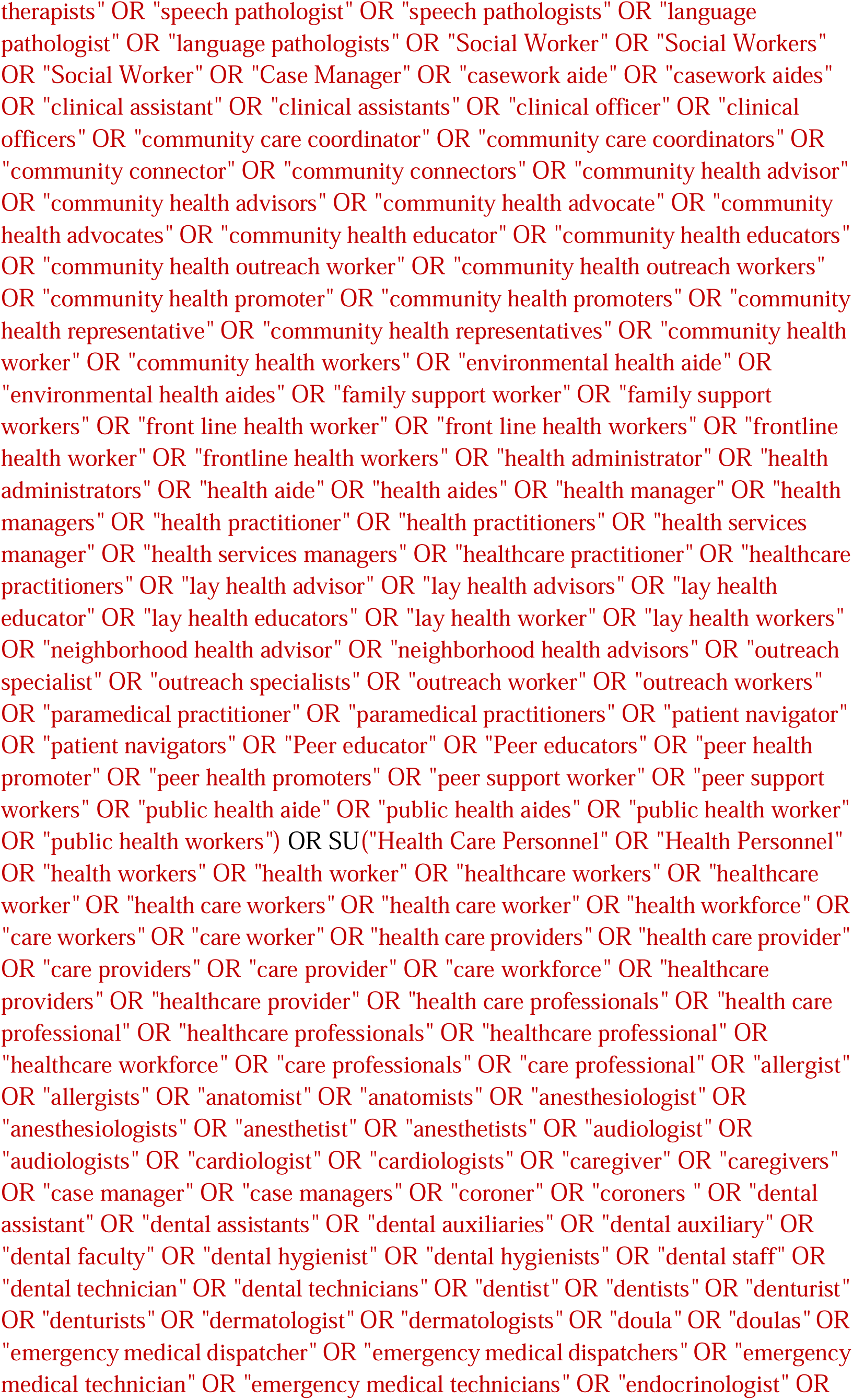
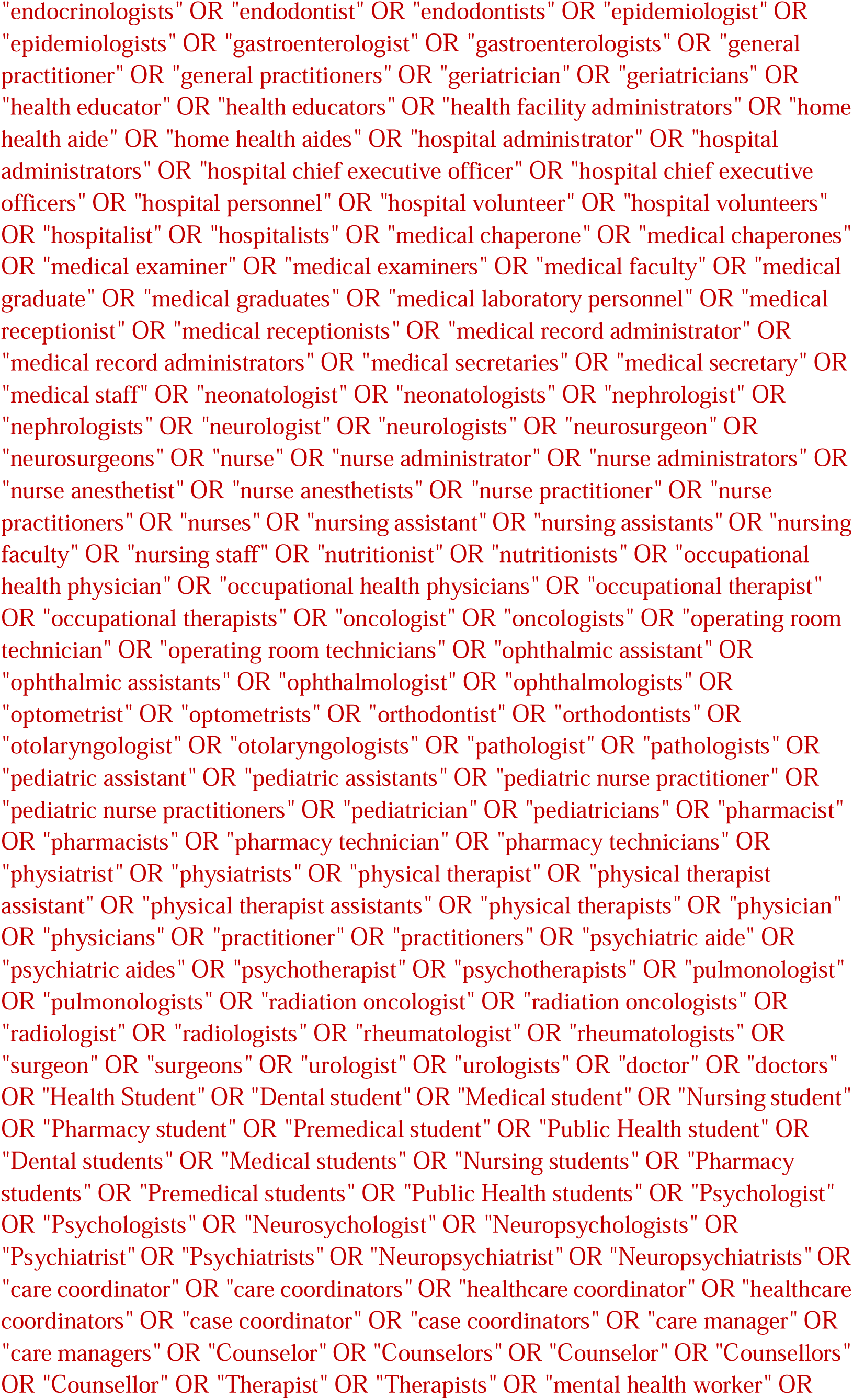
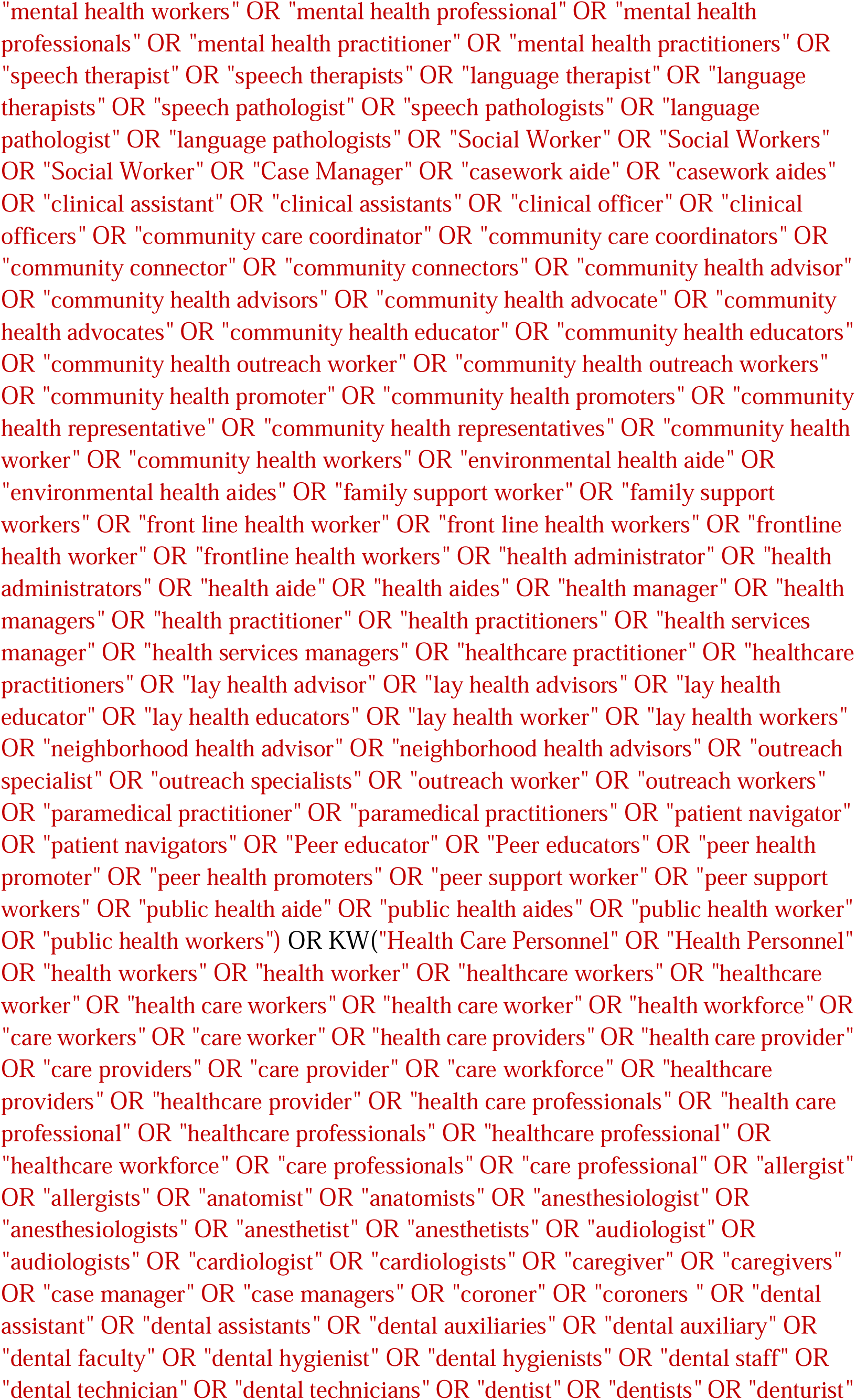
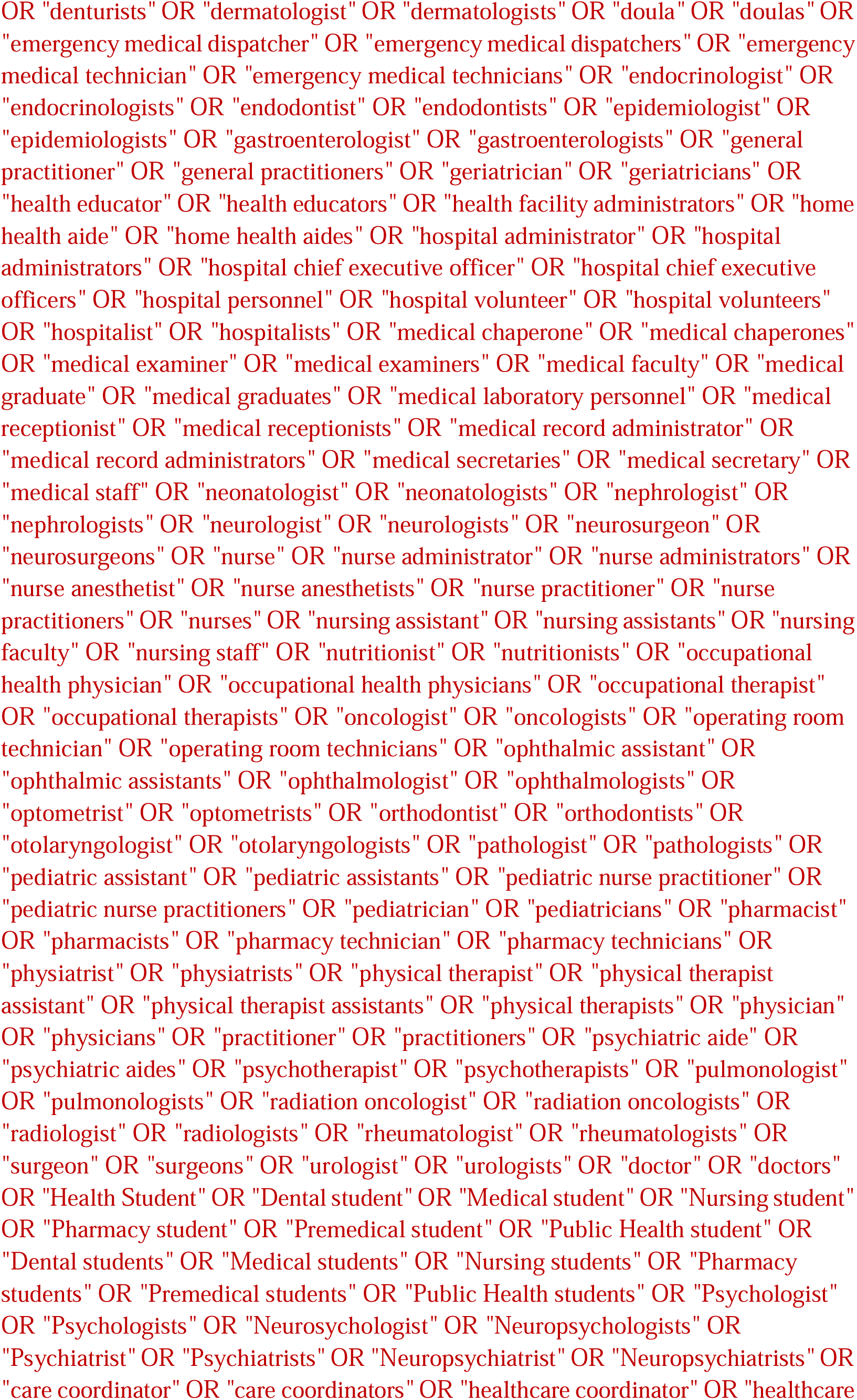
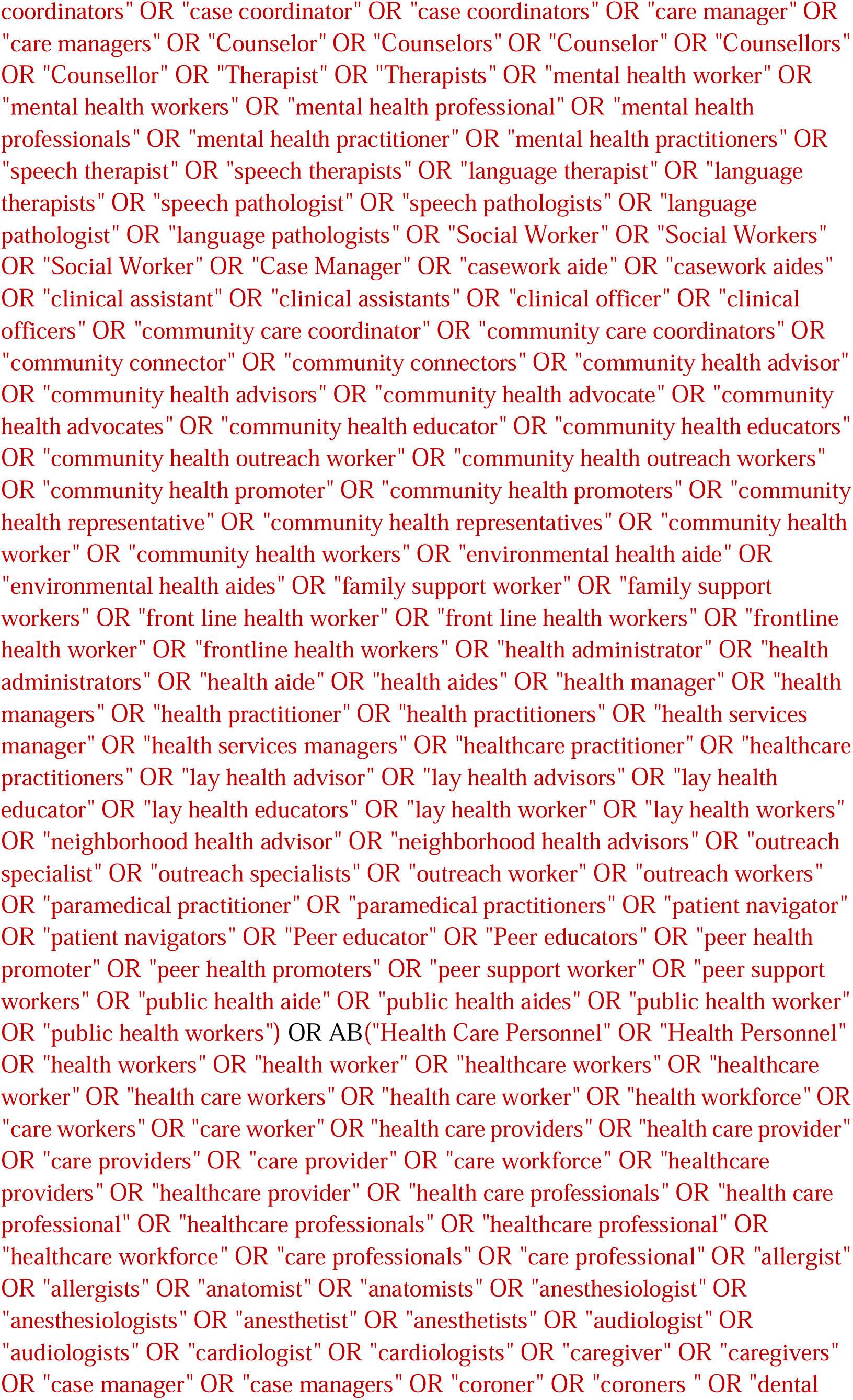
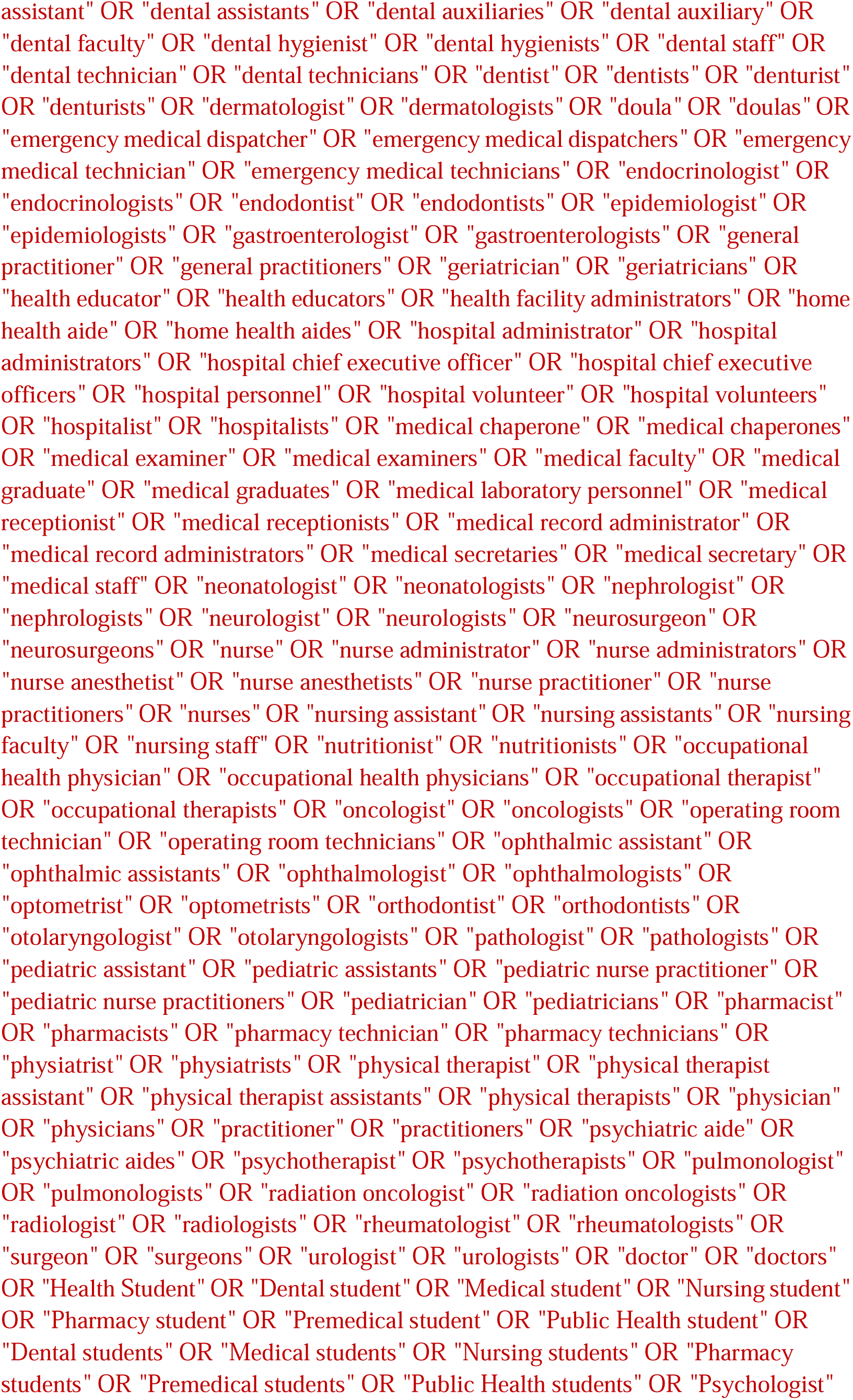
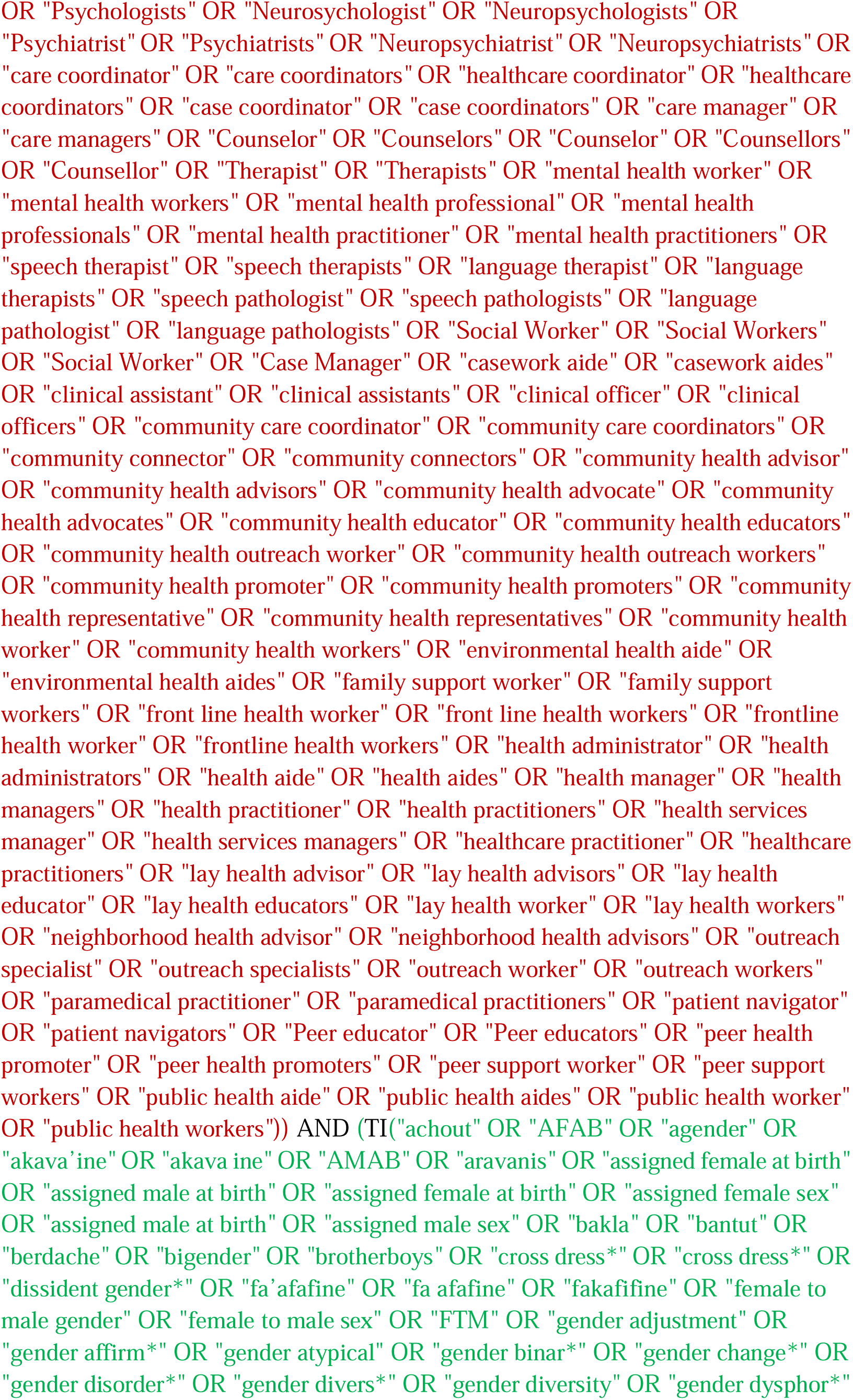
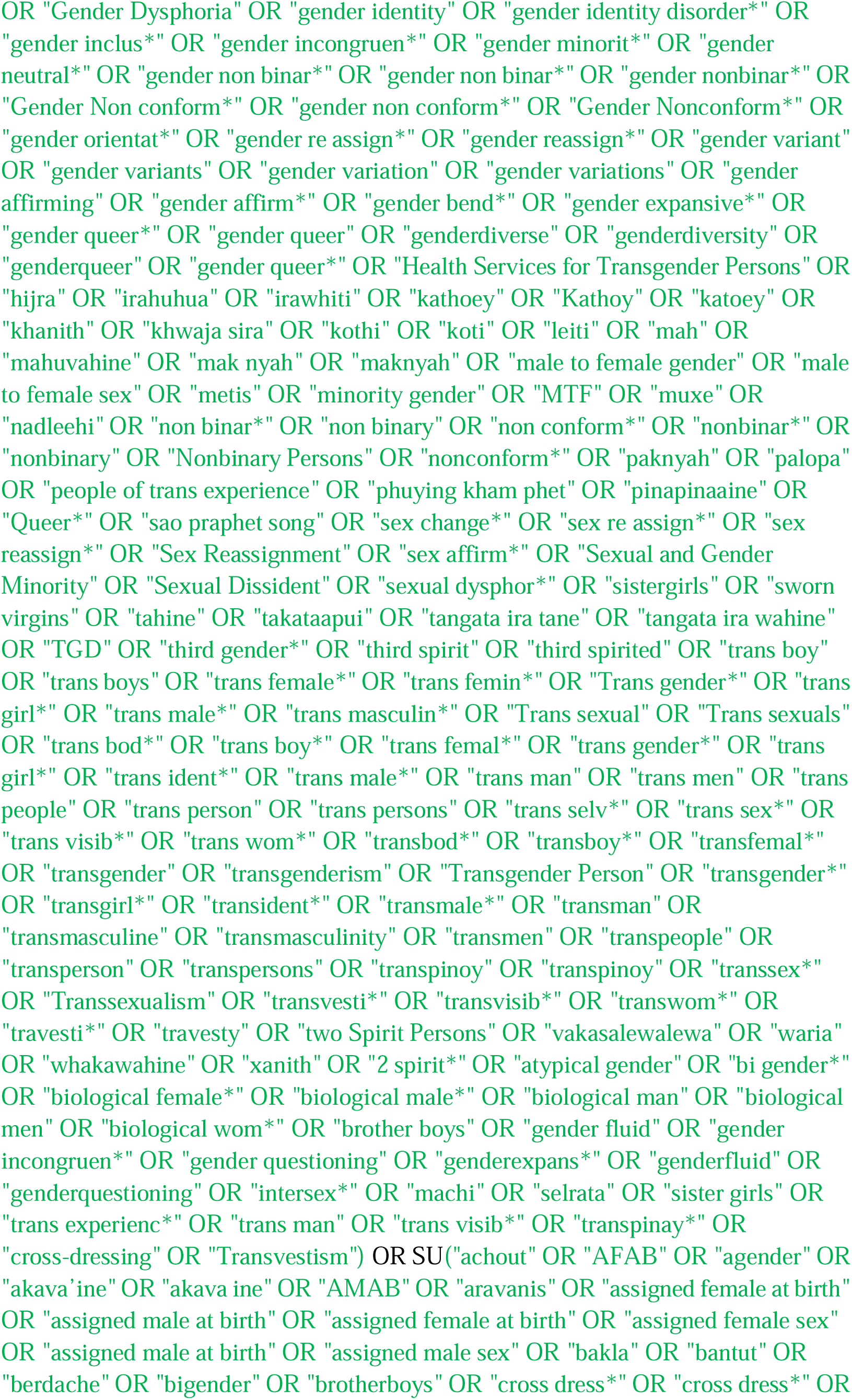
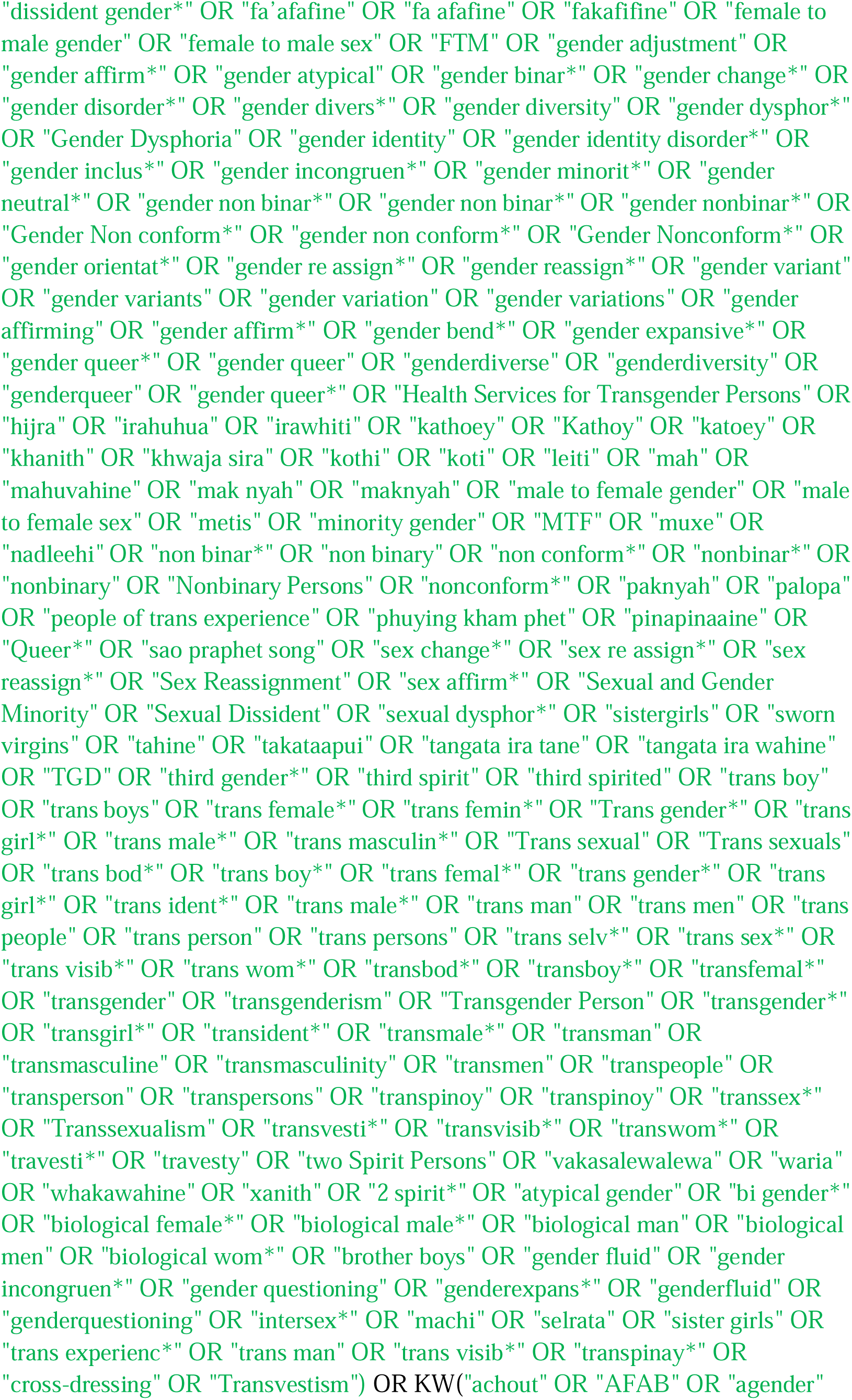
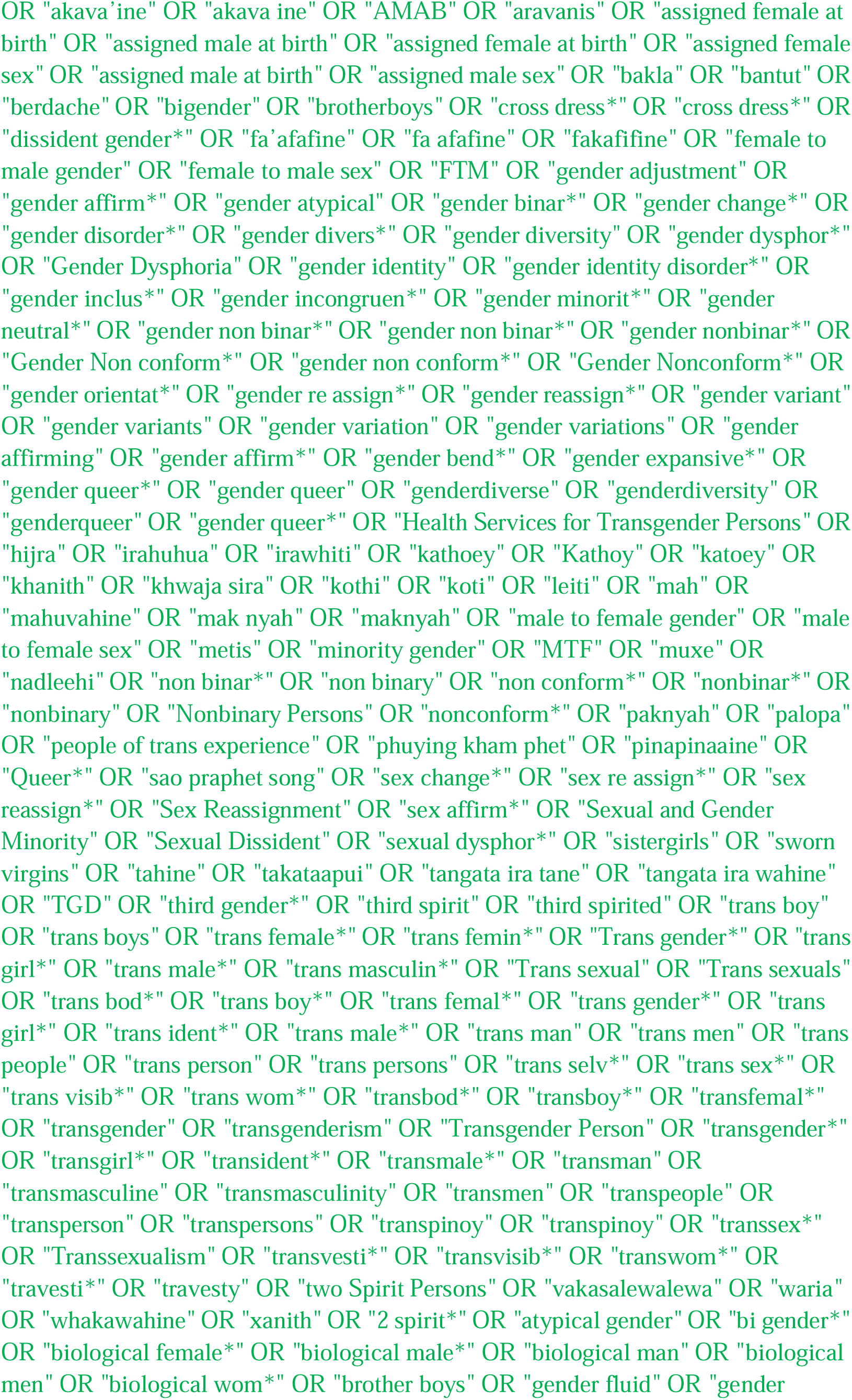
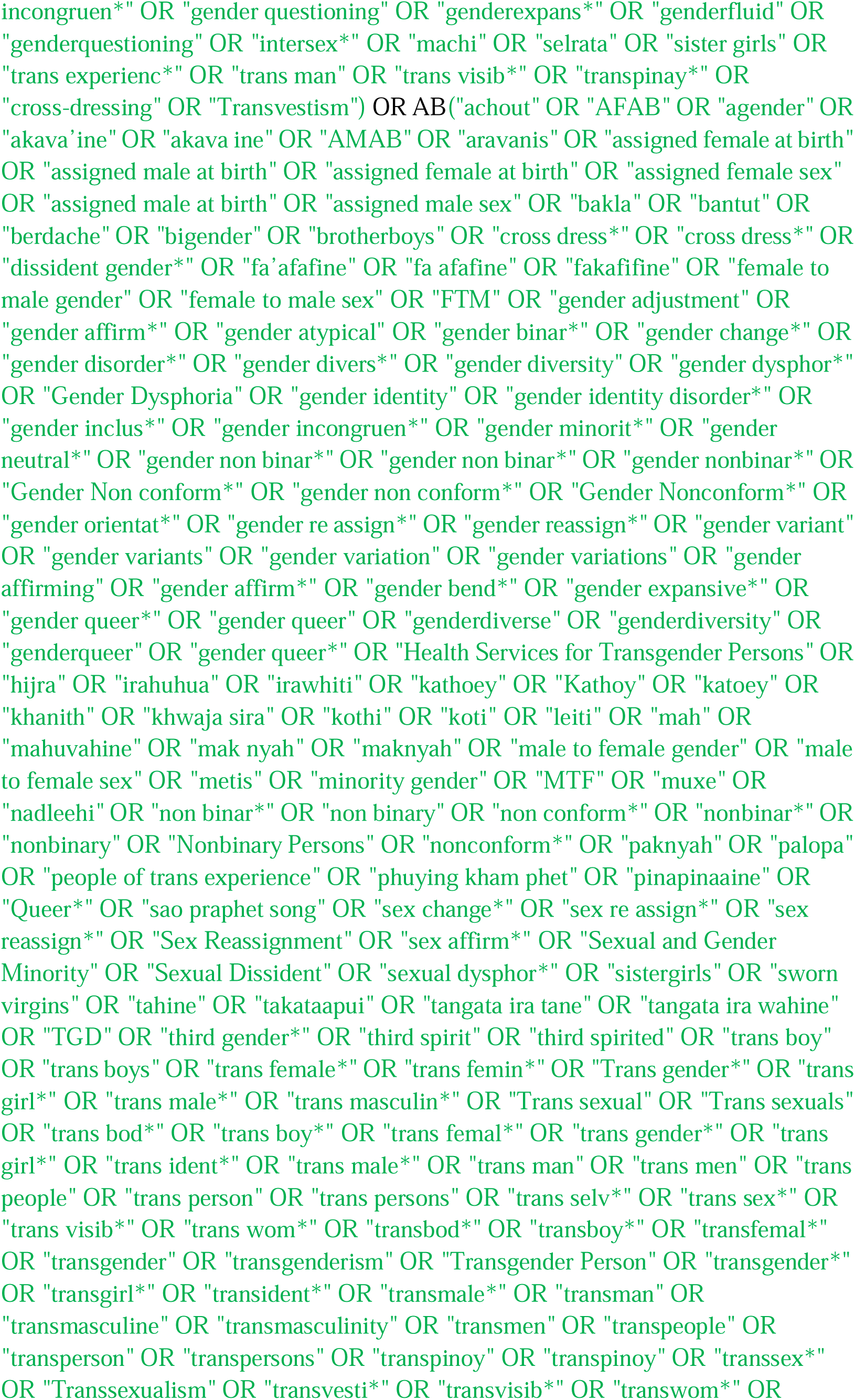
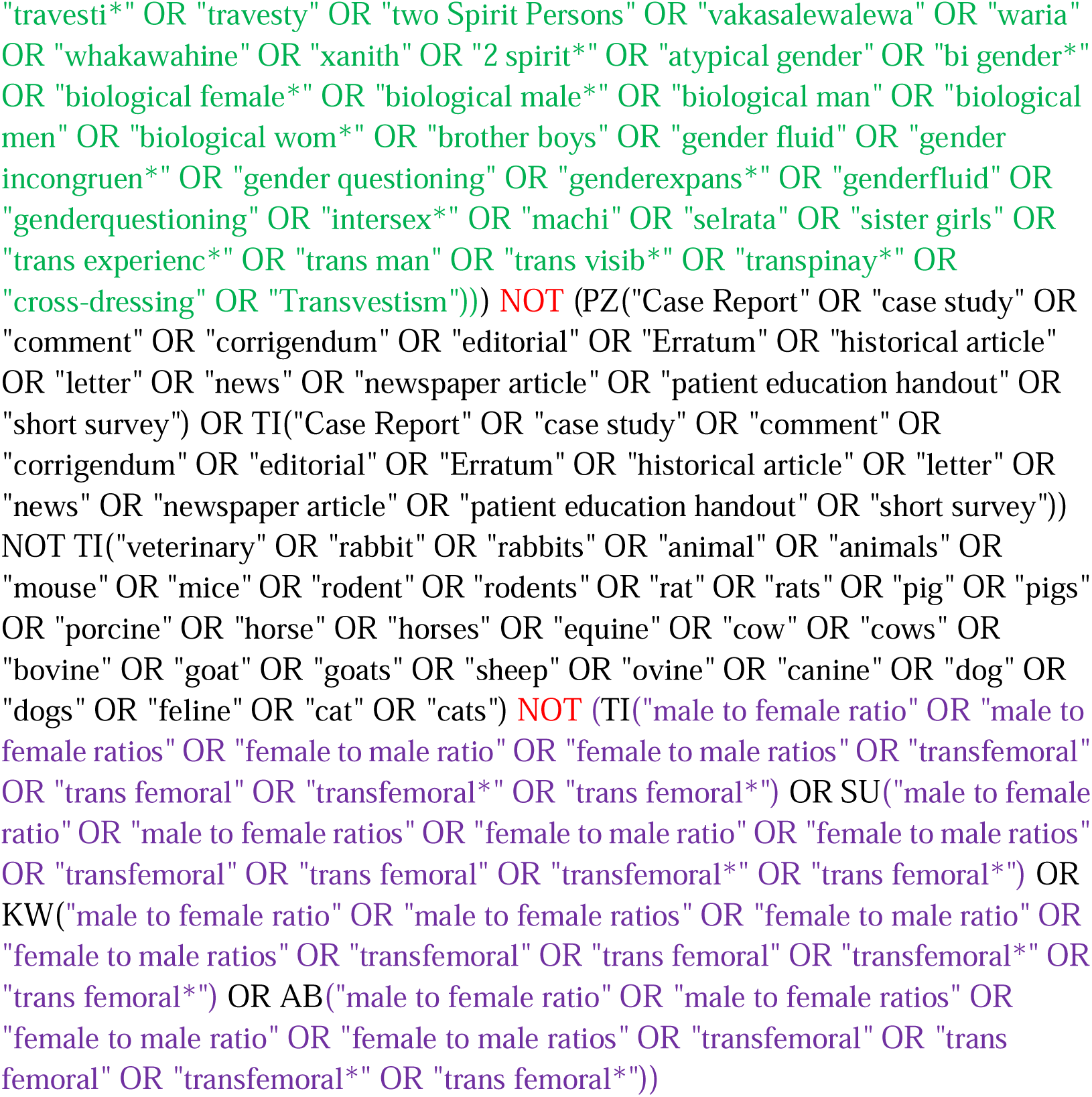

#### Psychology and Behavioral Sciences Collection (EBSCOhost)

http://search.EBSCOhost.com/login.aspx?authtype=ip,uid&profile=lumc&defaultdb=aph

Limit to Academic Journals

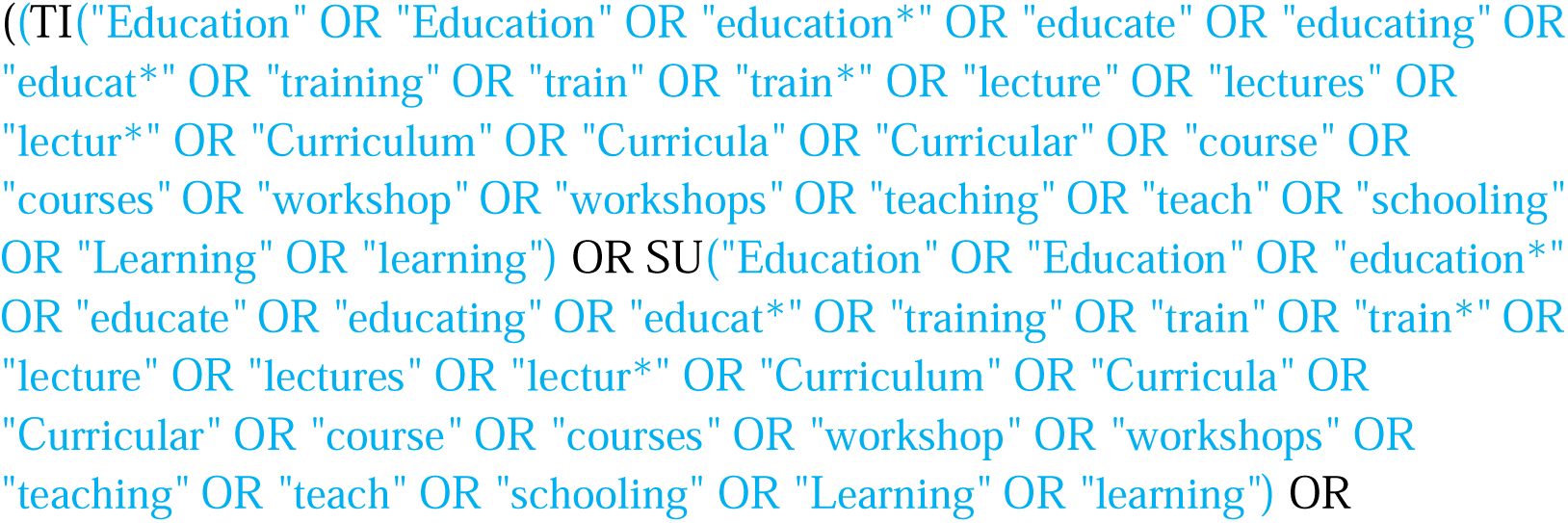
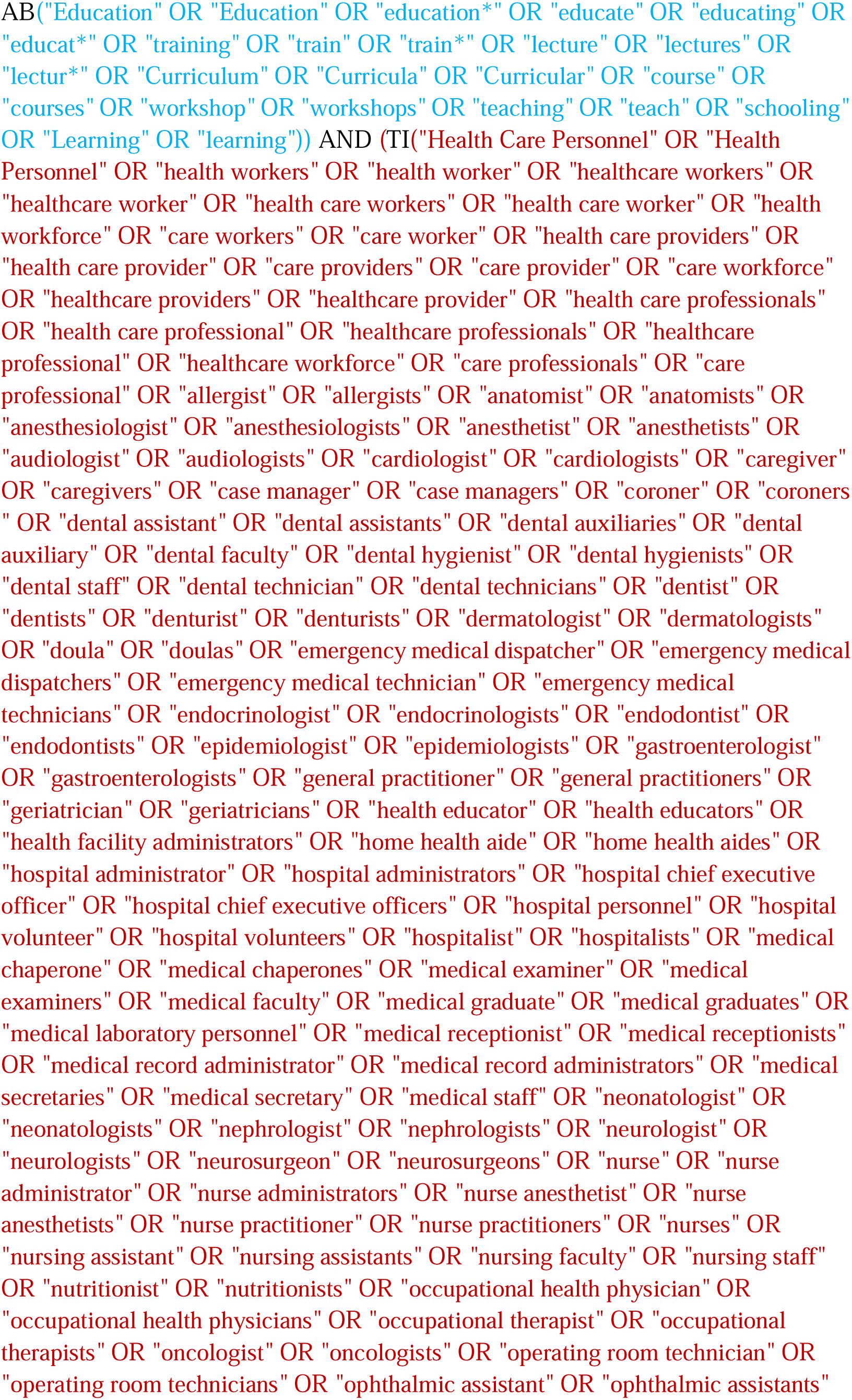
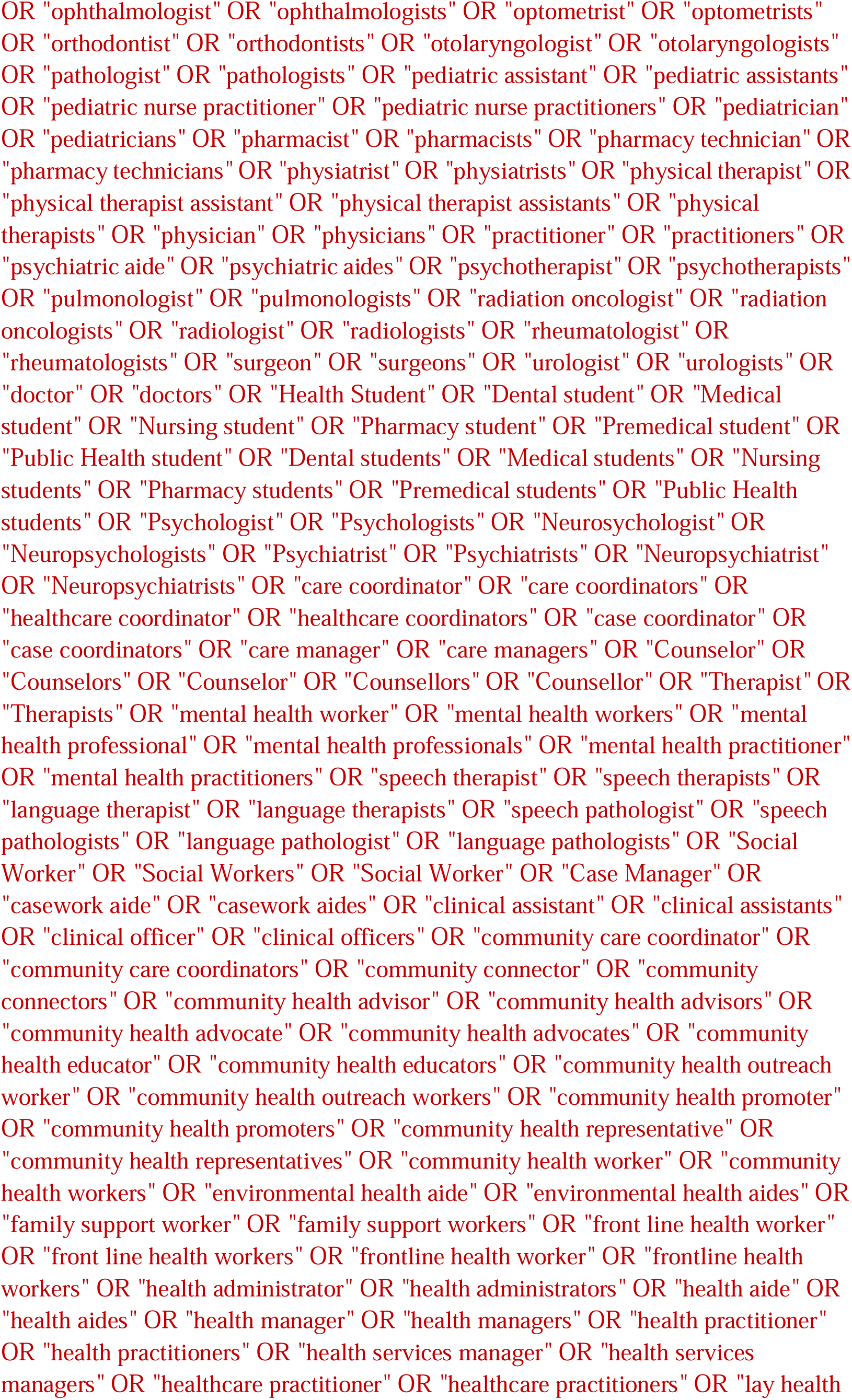
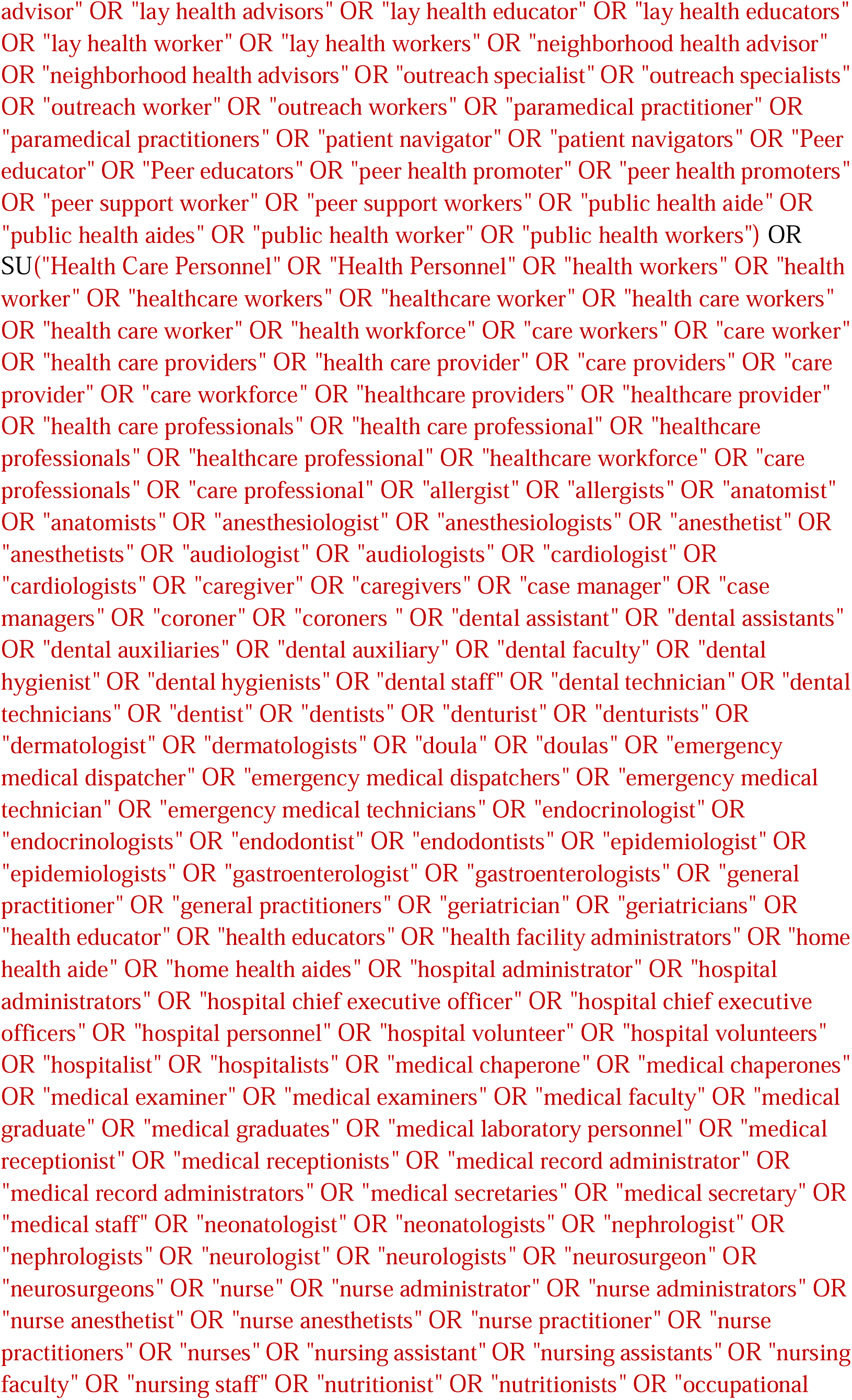
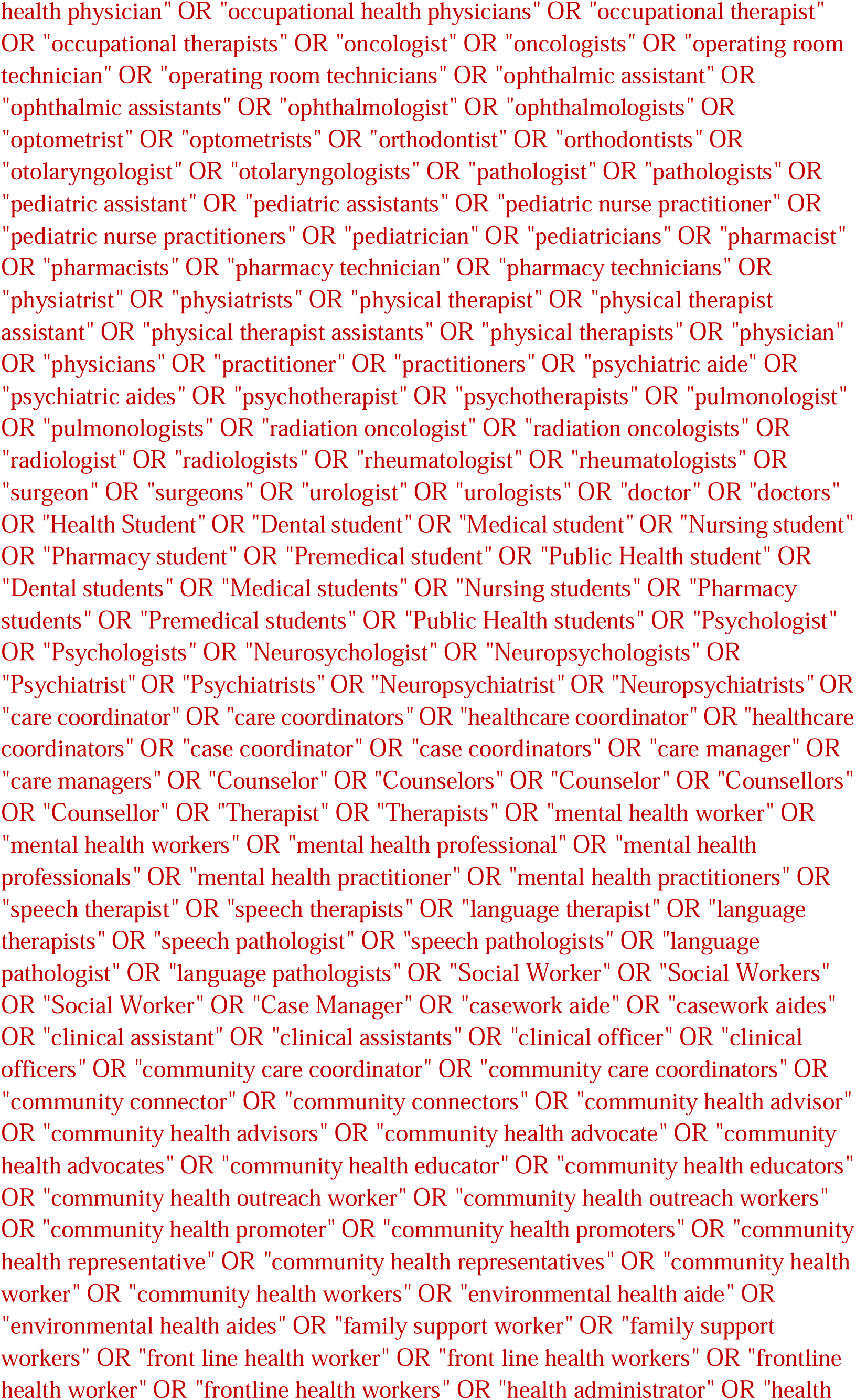
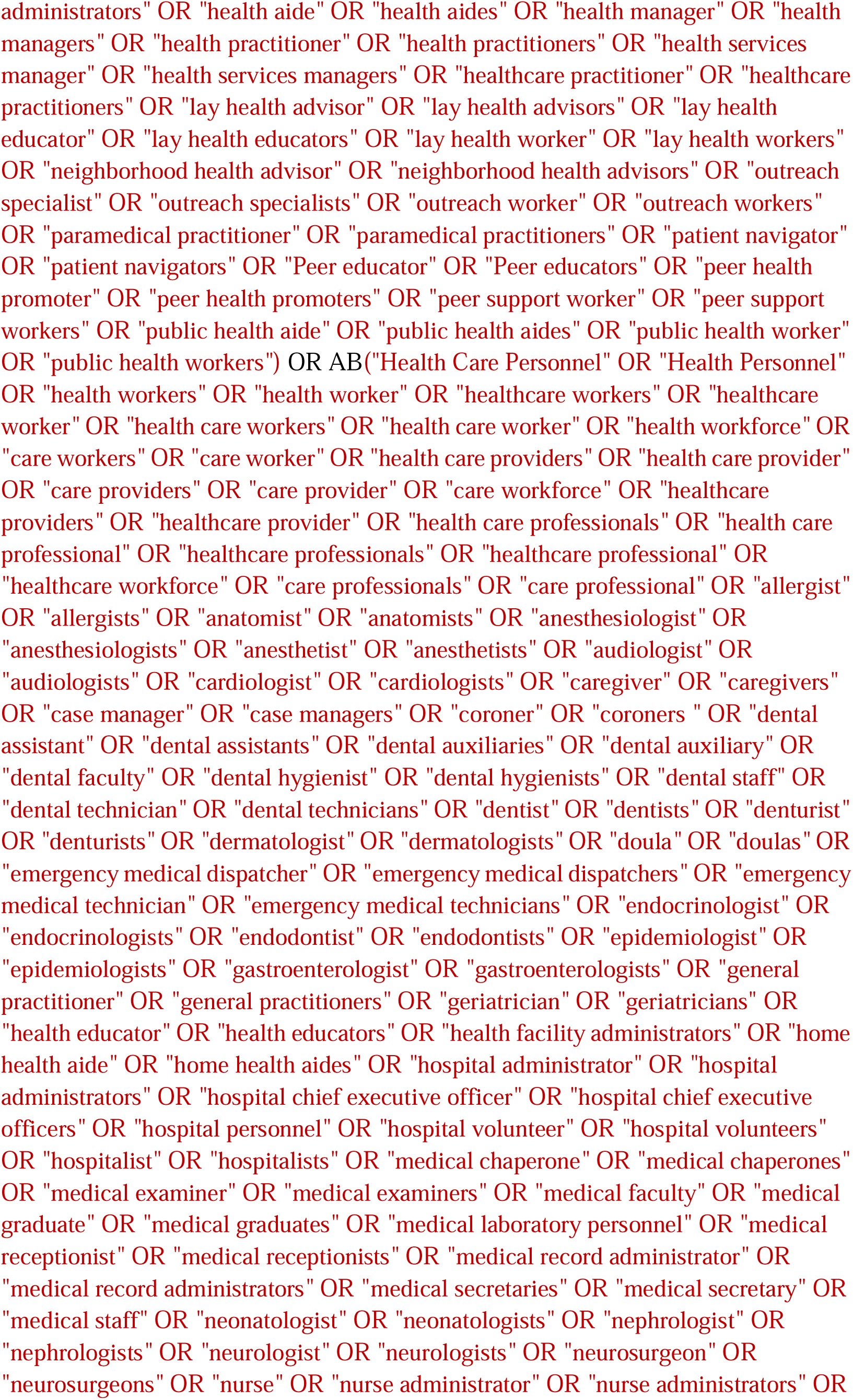
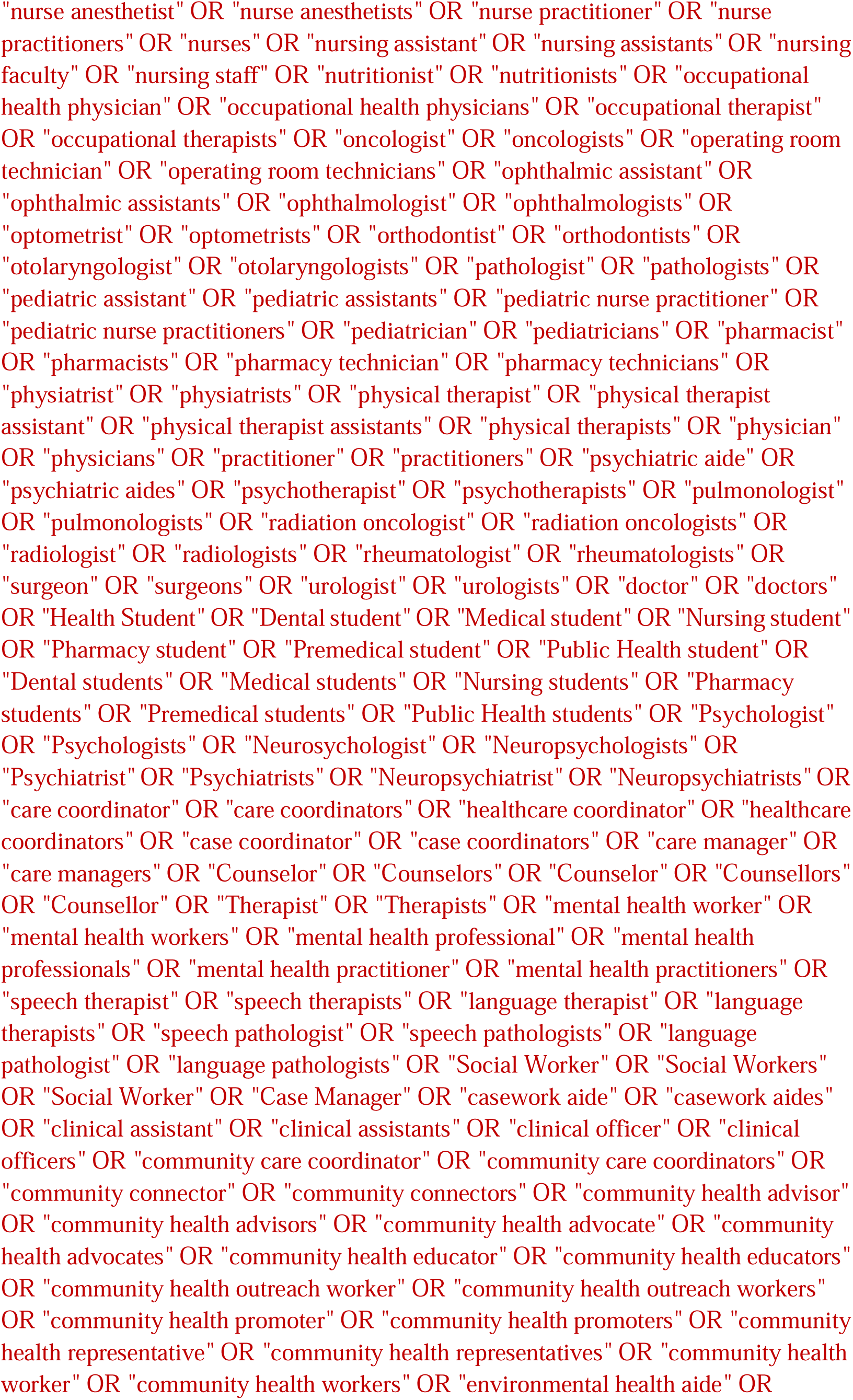
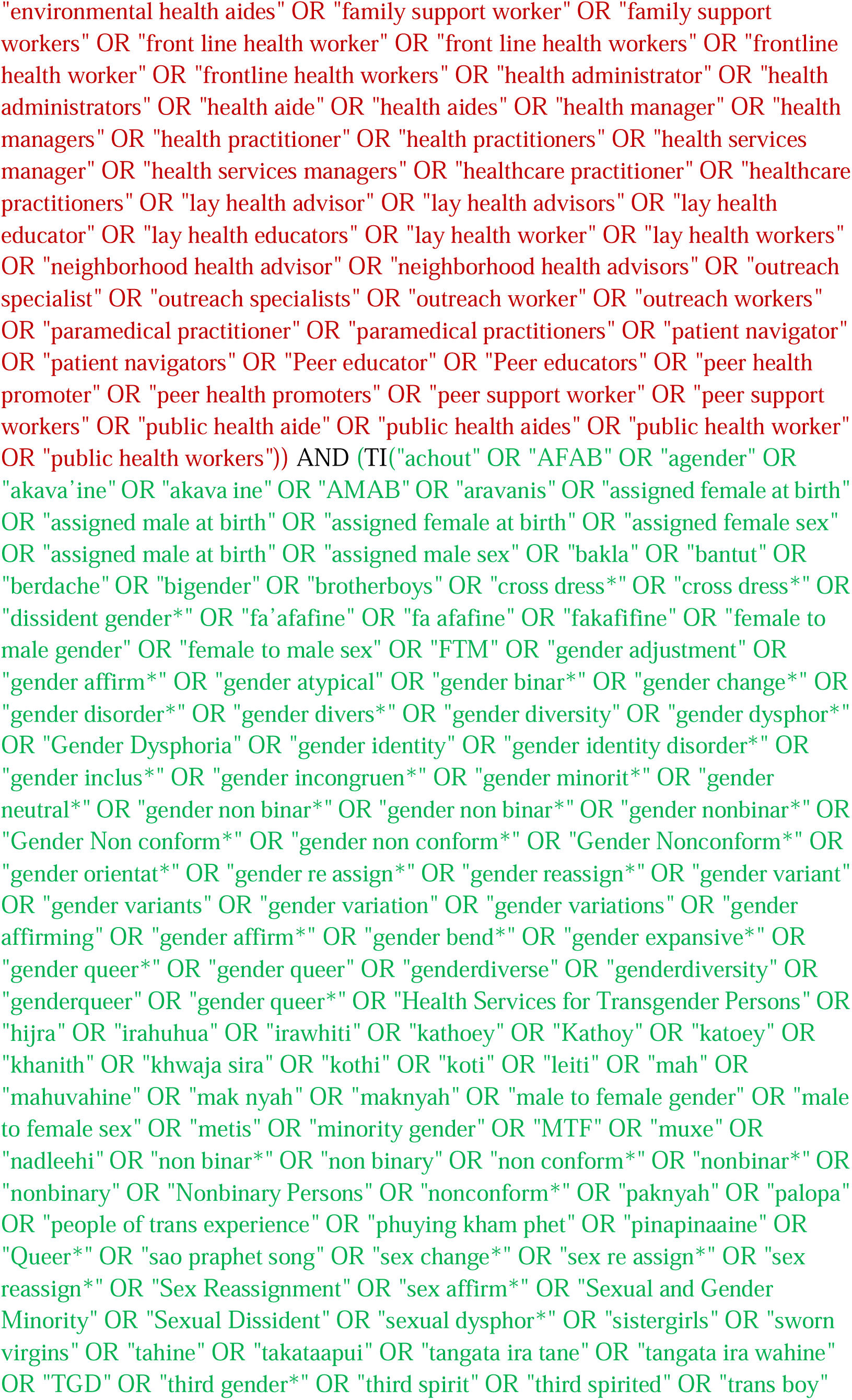
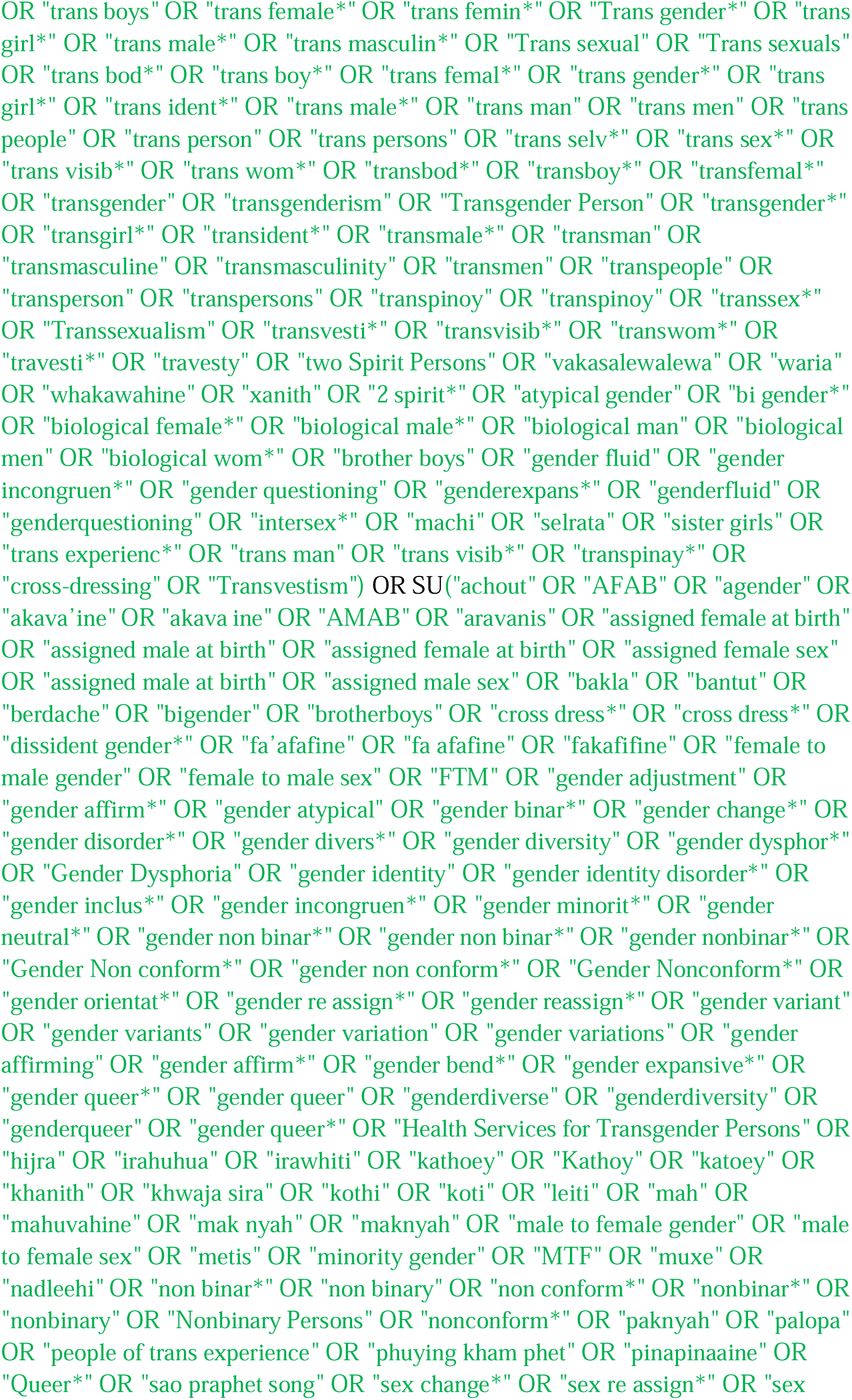
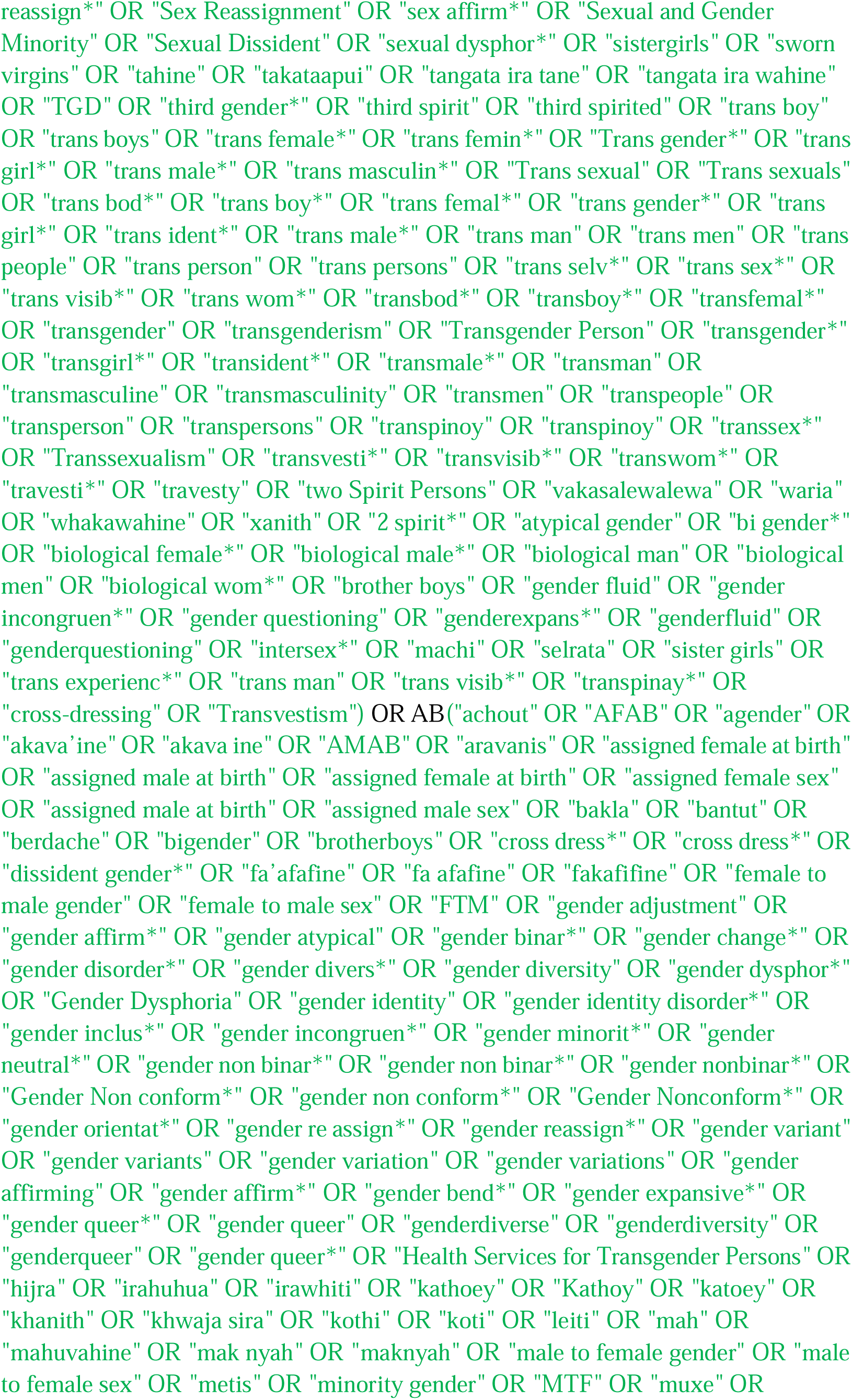
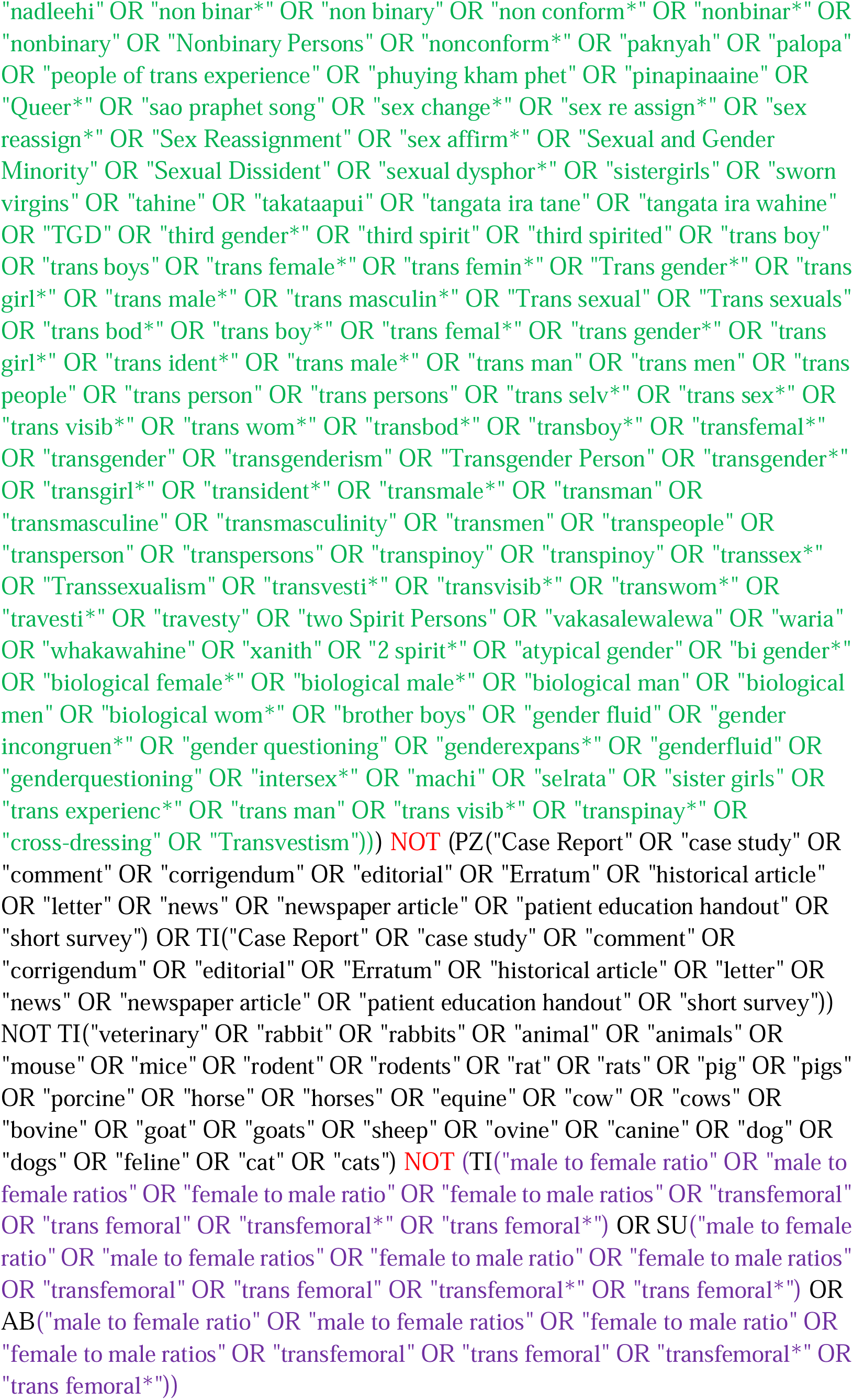

#### ERIC (EBSCOhost)

Via Ebsco: http://databases.library.leiden.edu/?bibid=990024848320302711&redirect=true

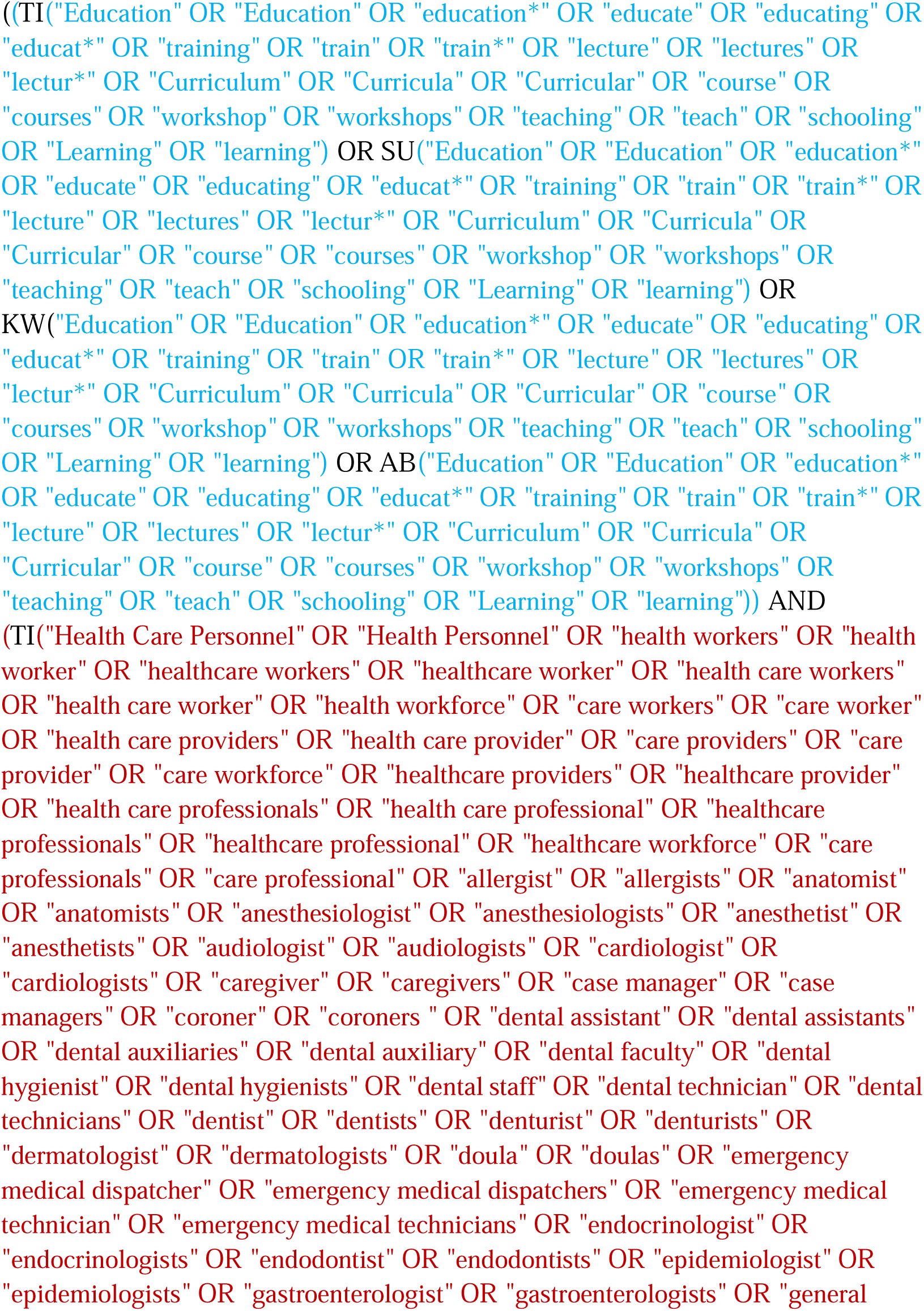
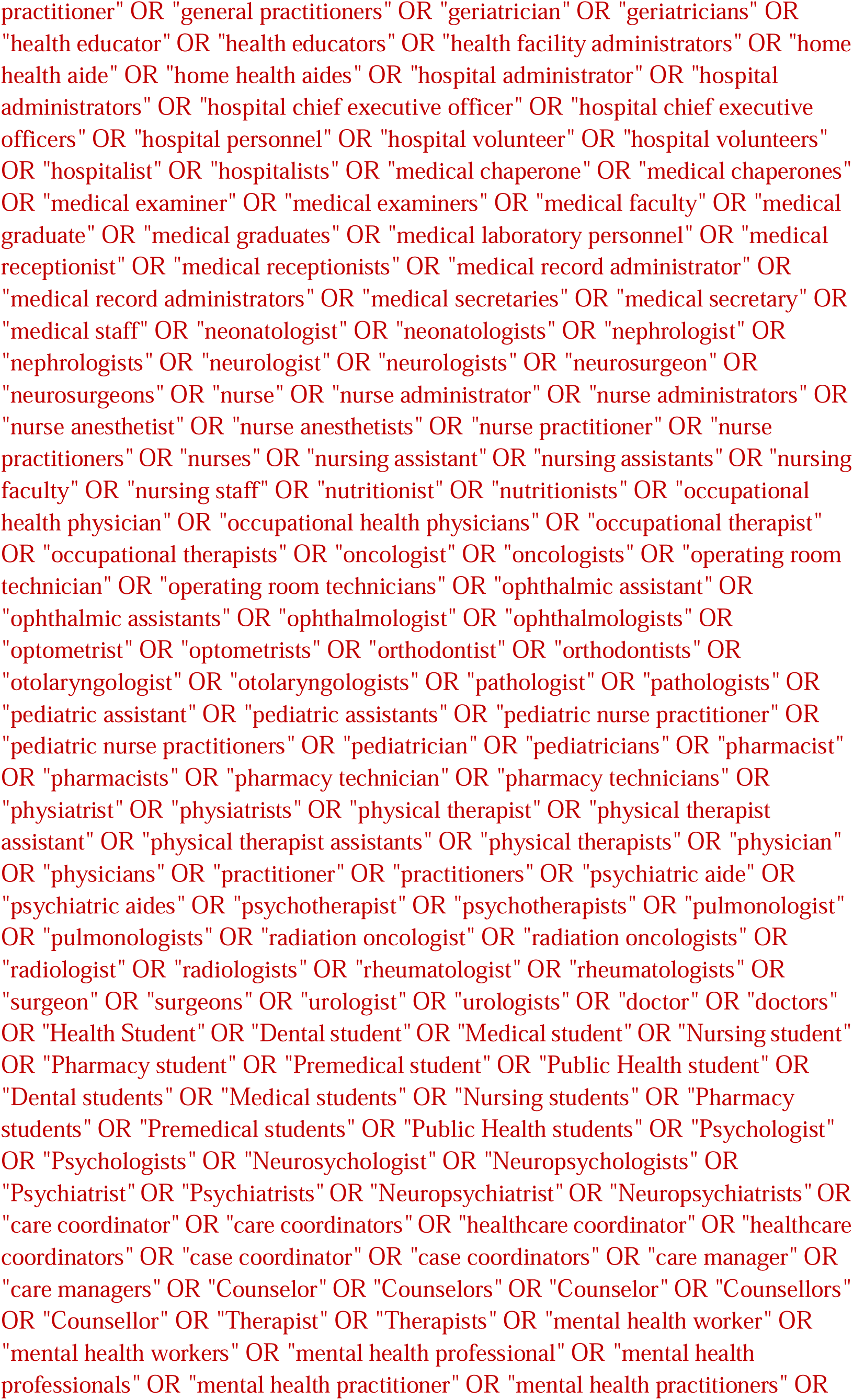
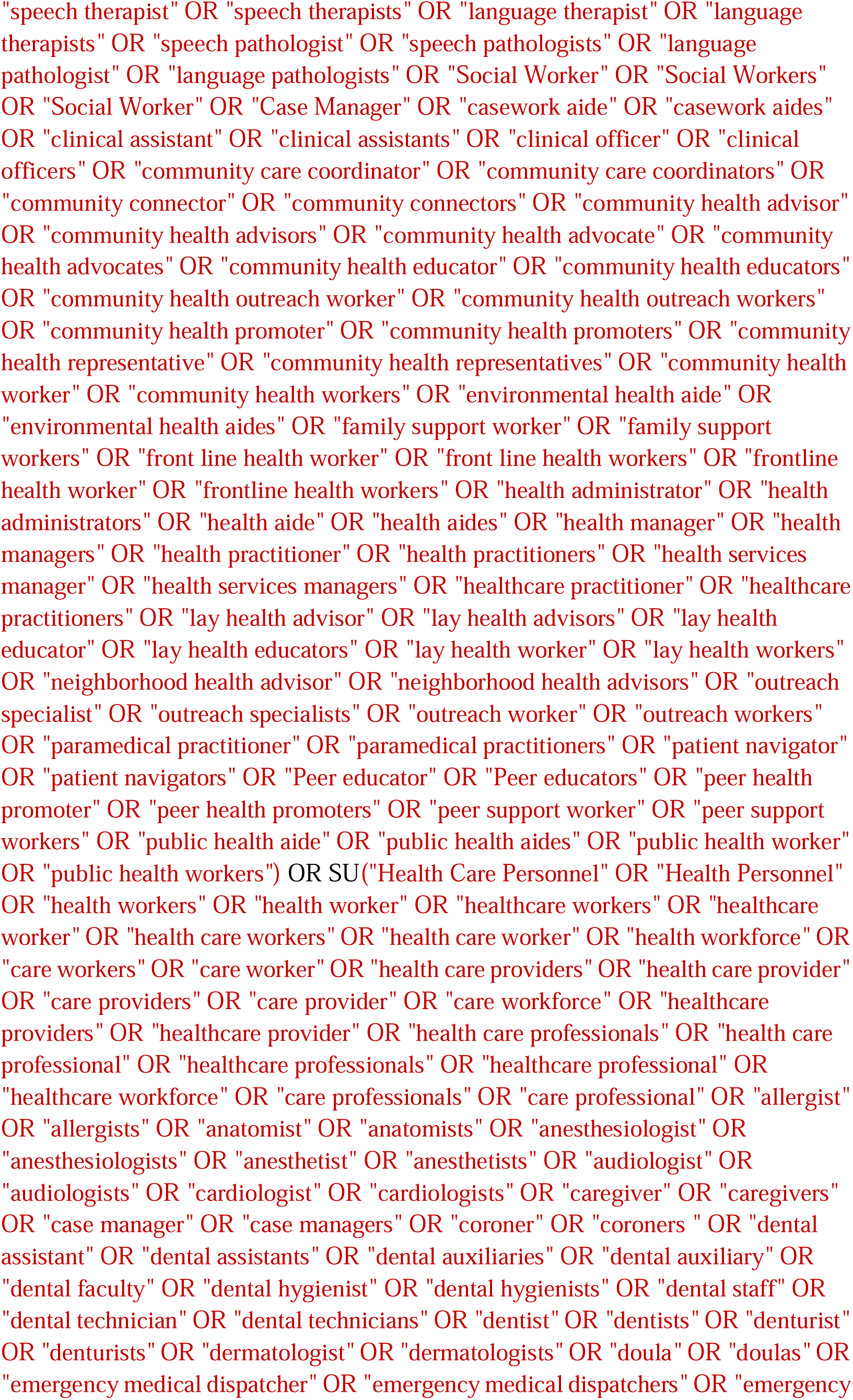
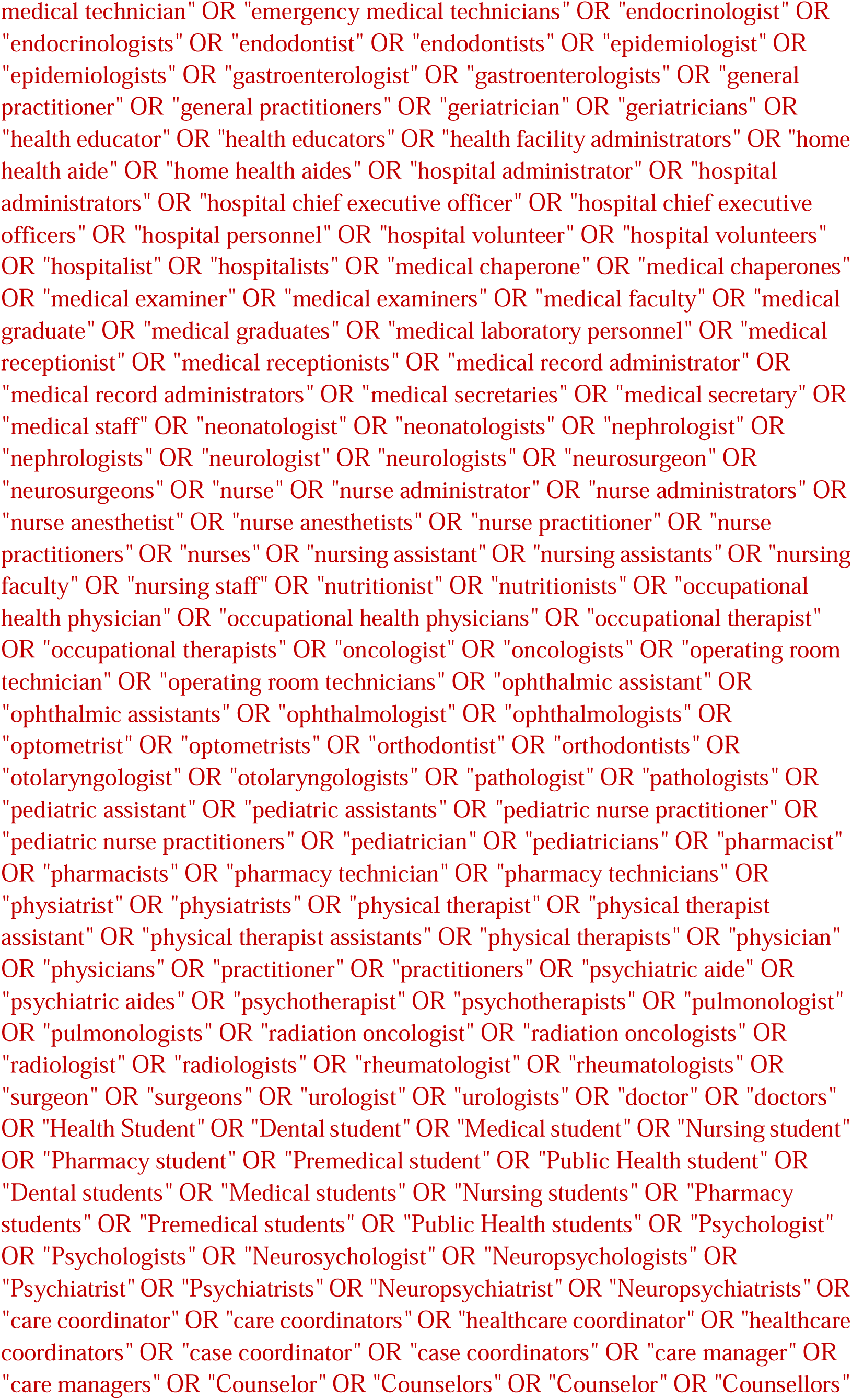
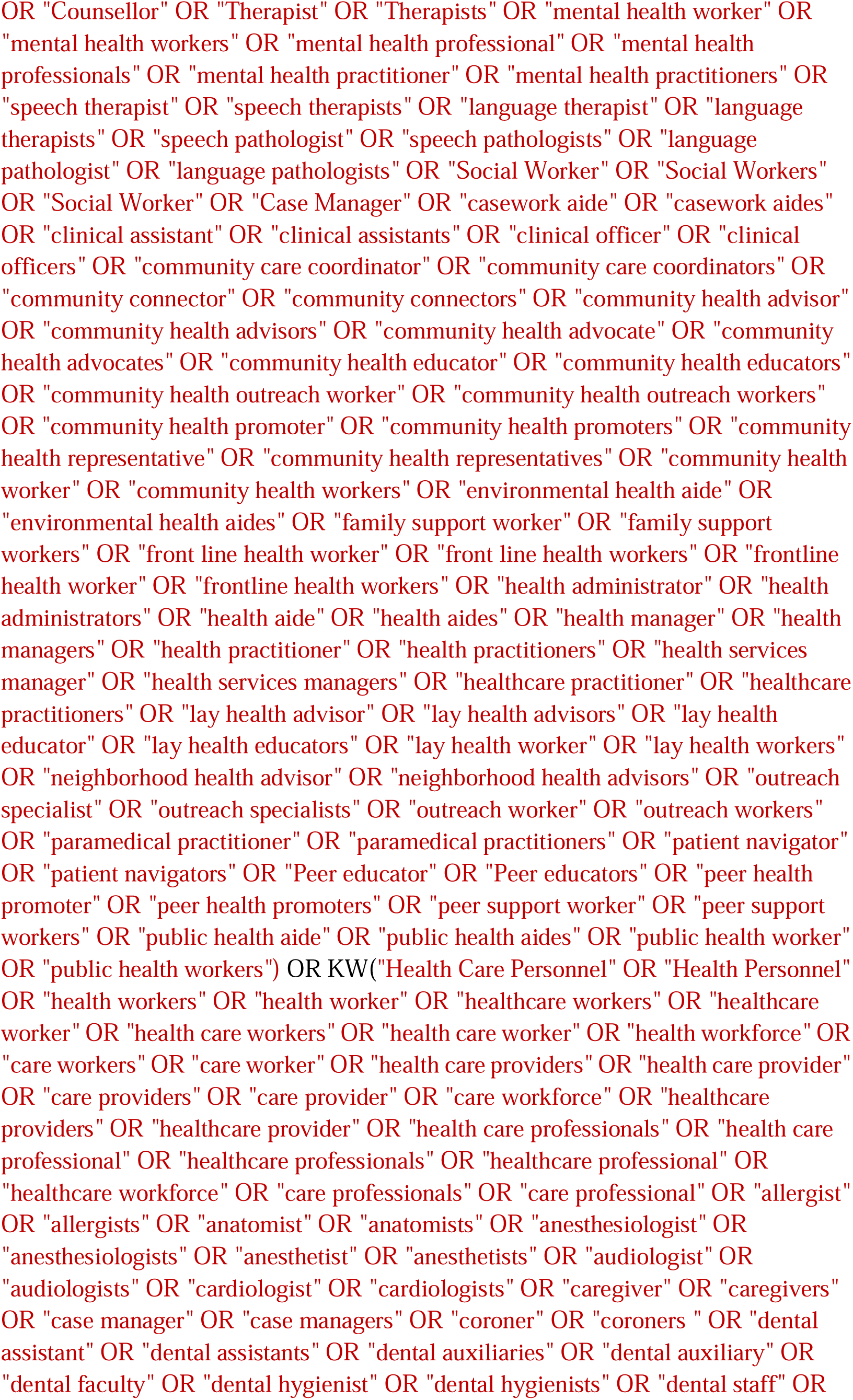
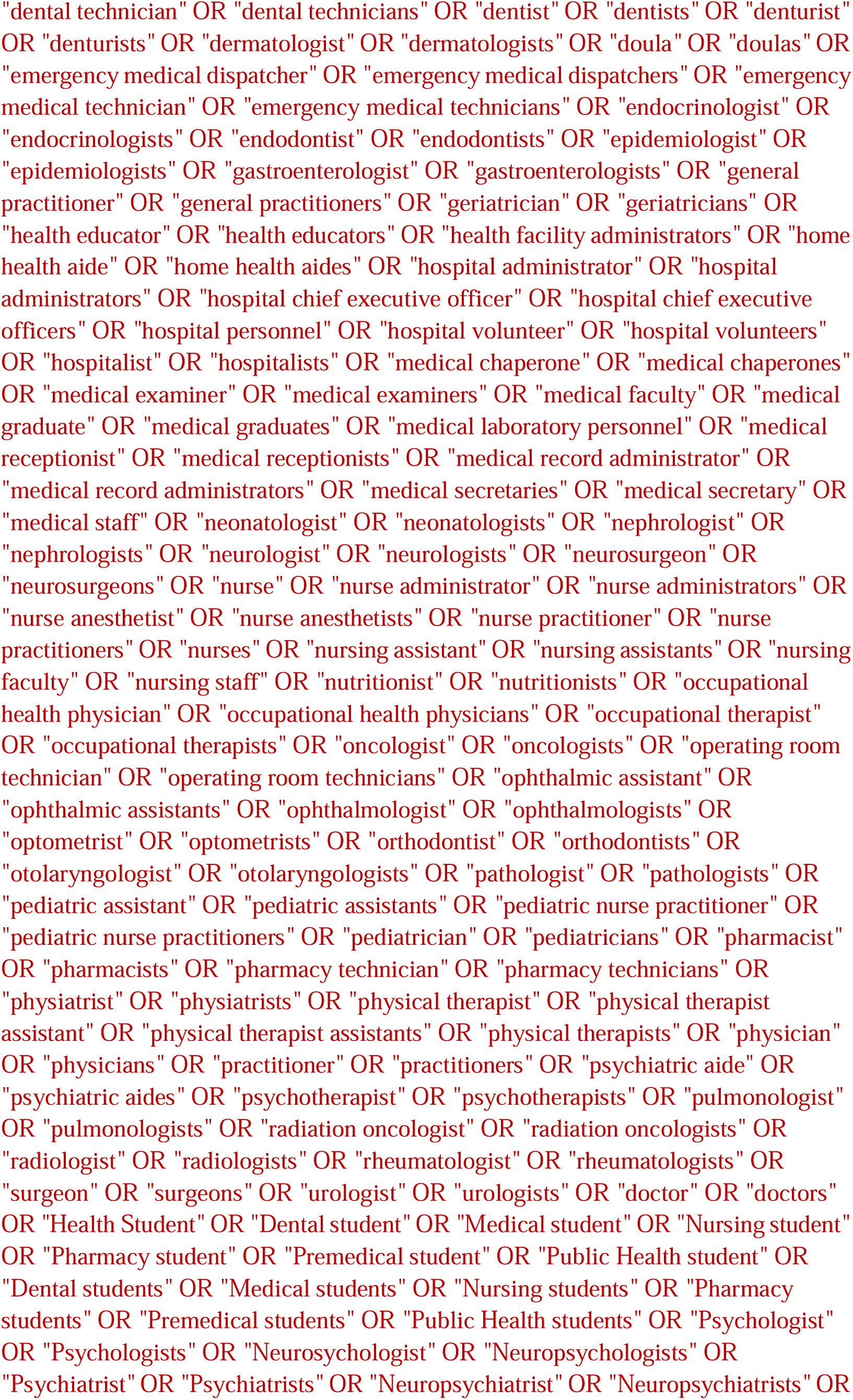
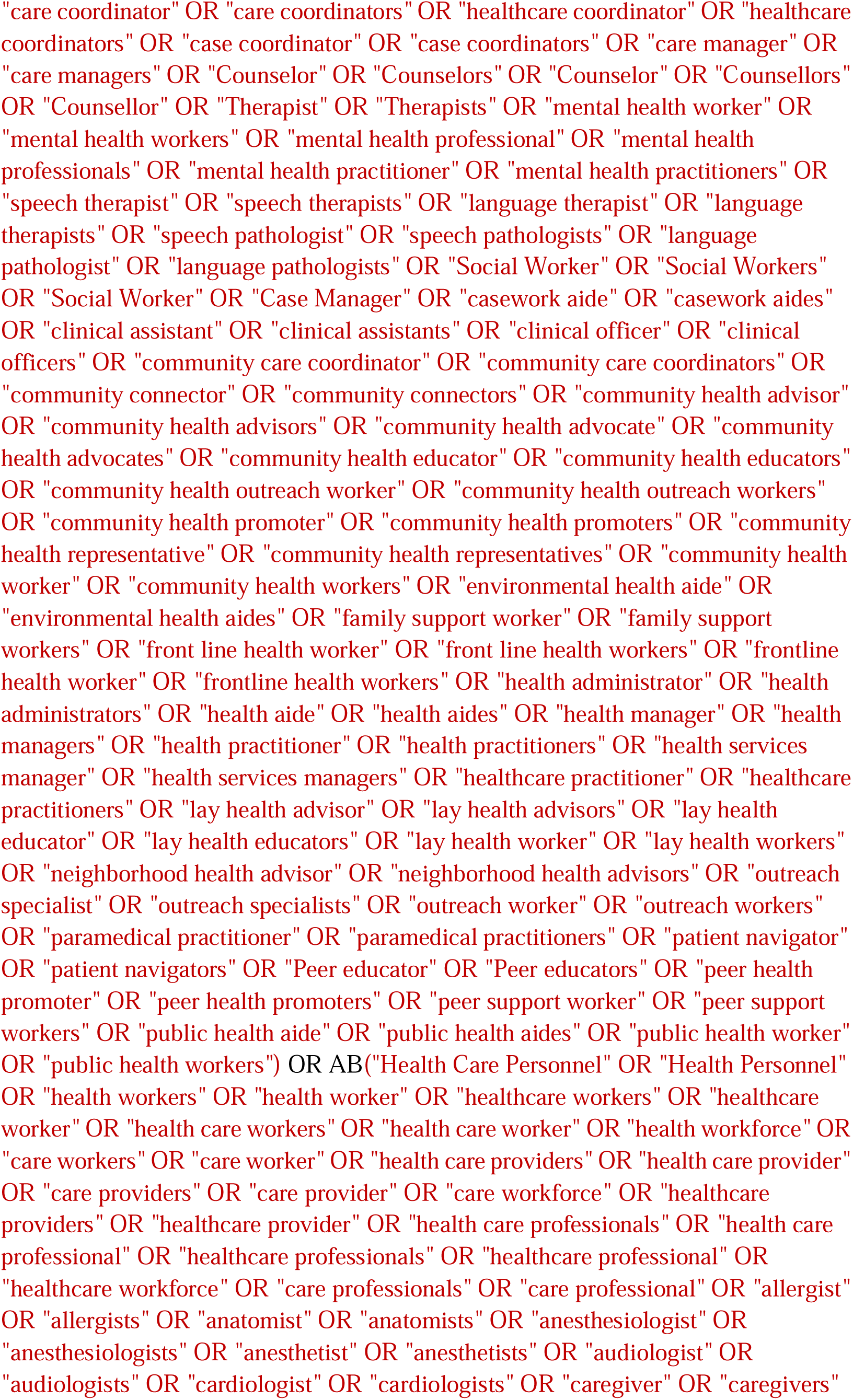
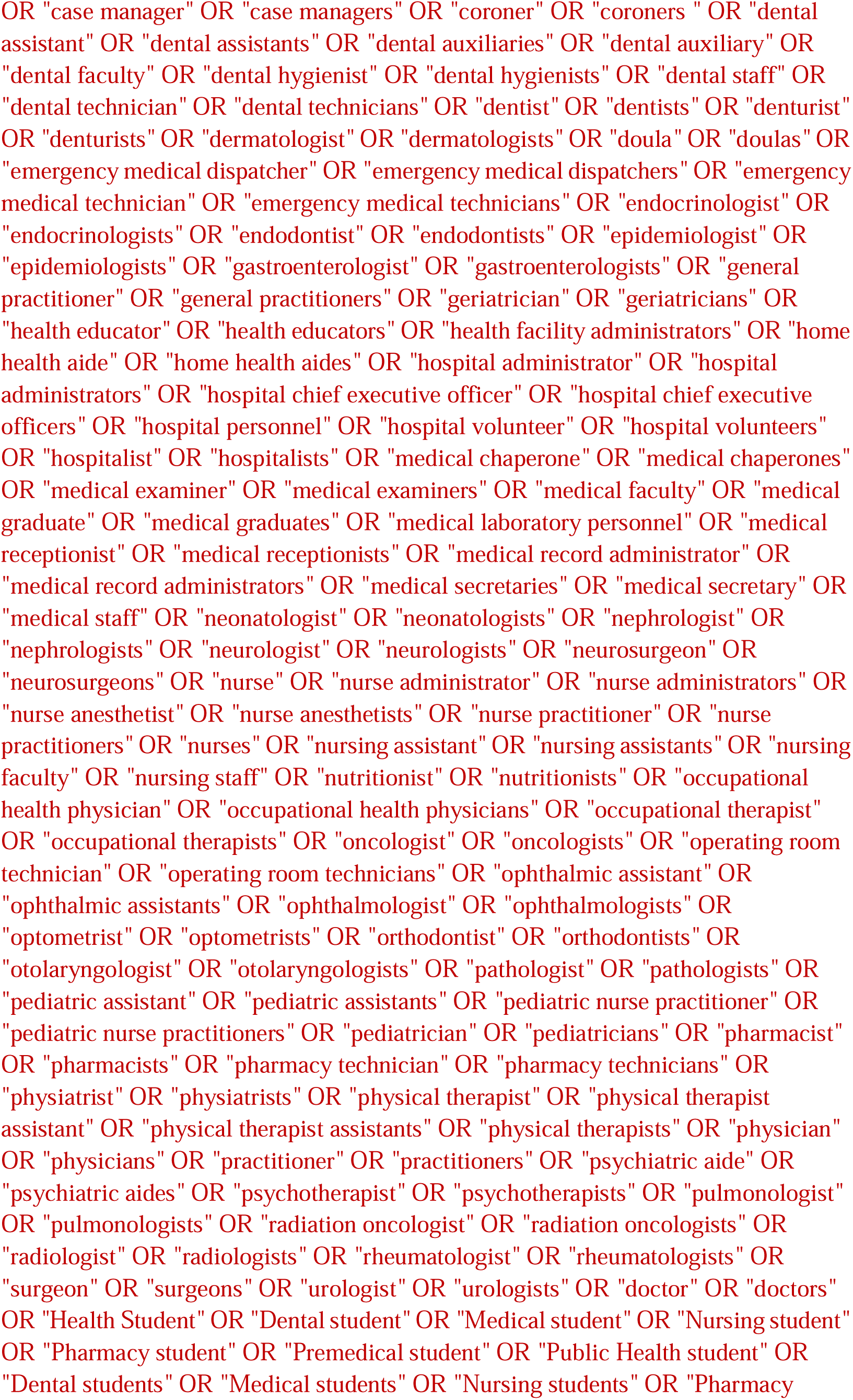
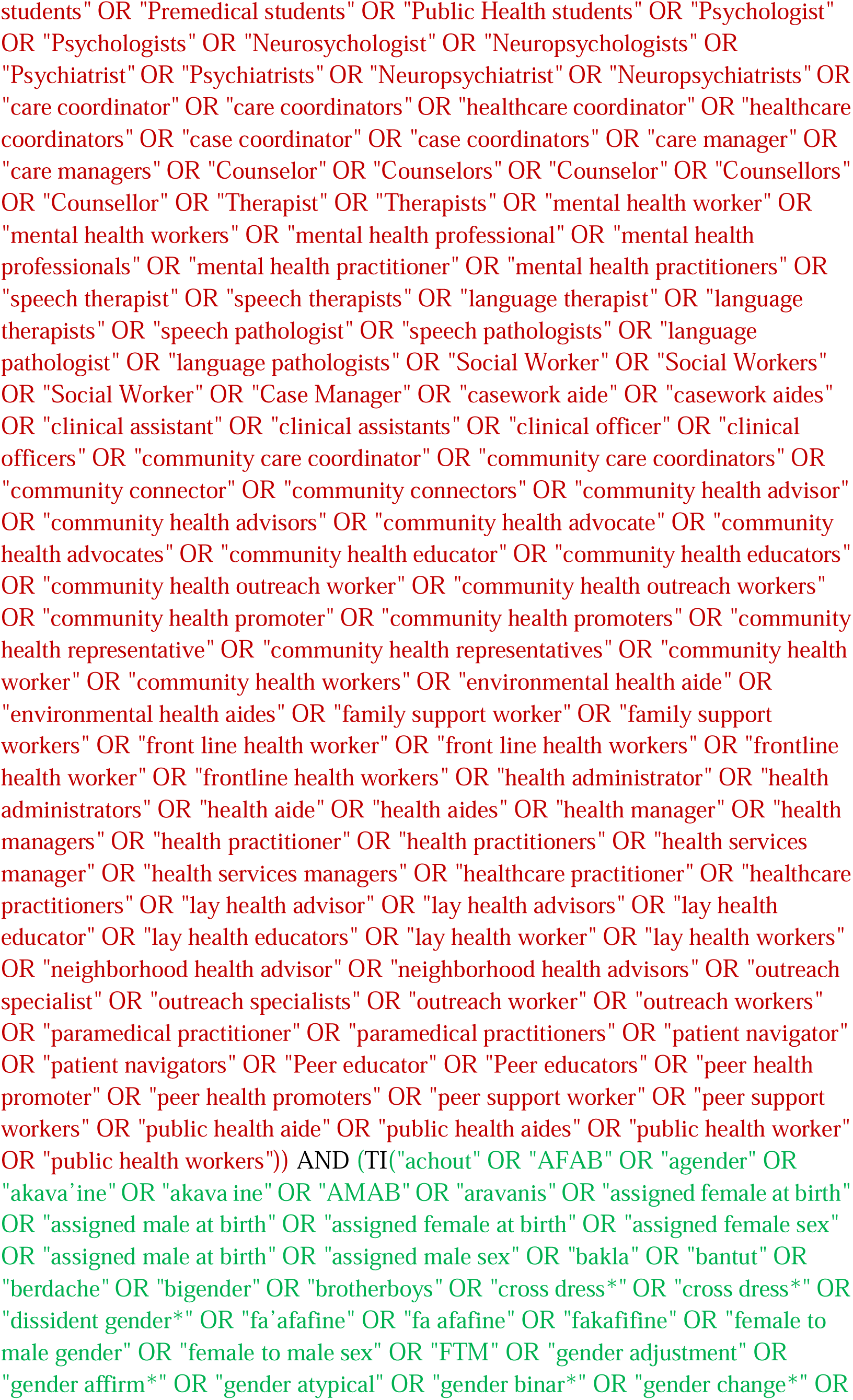
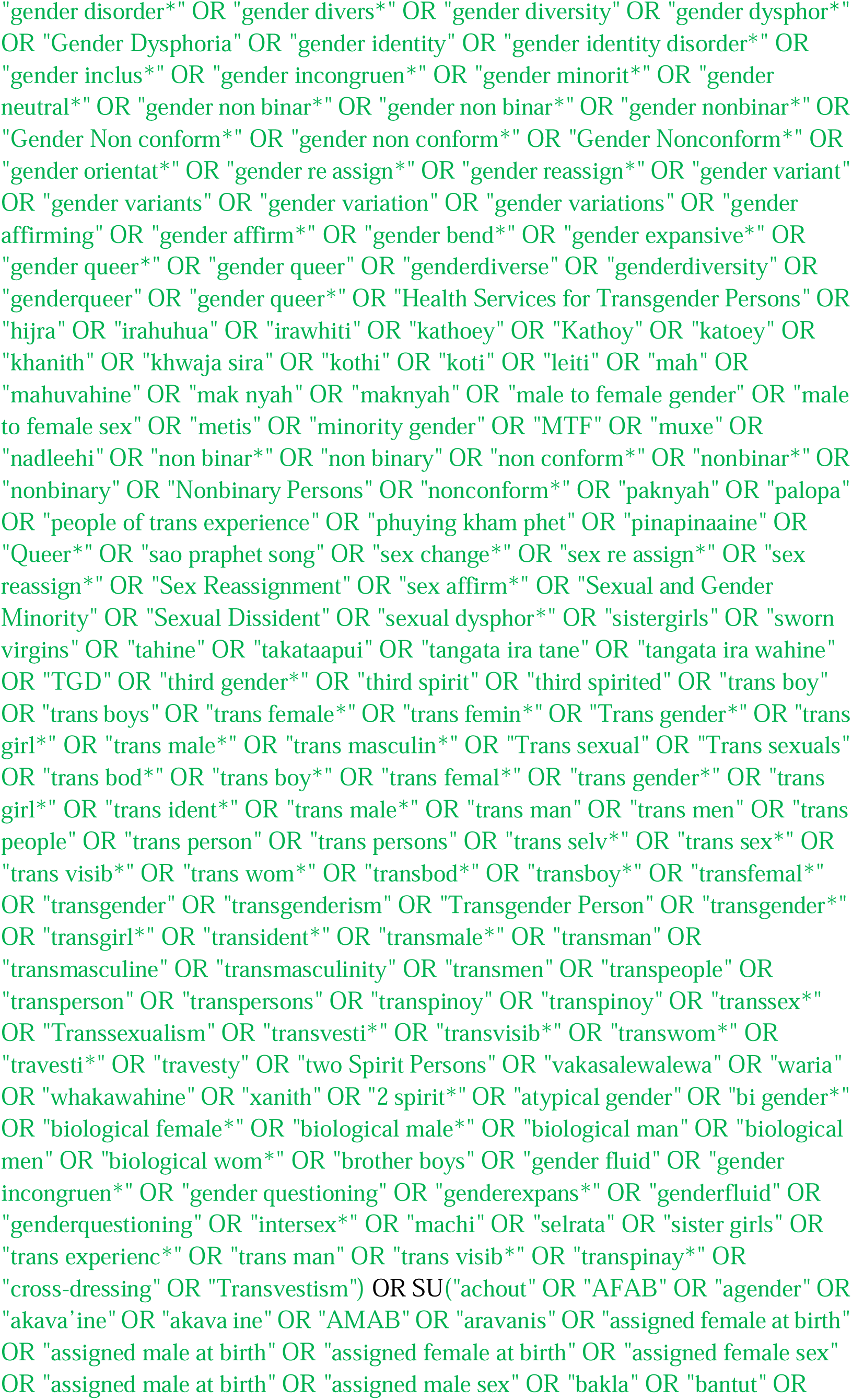
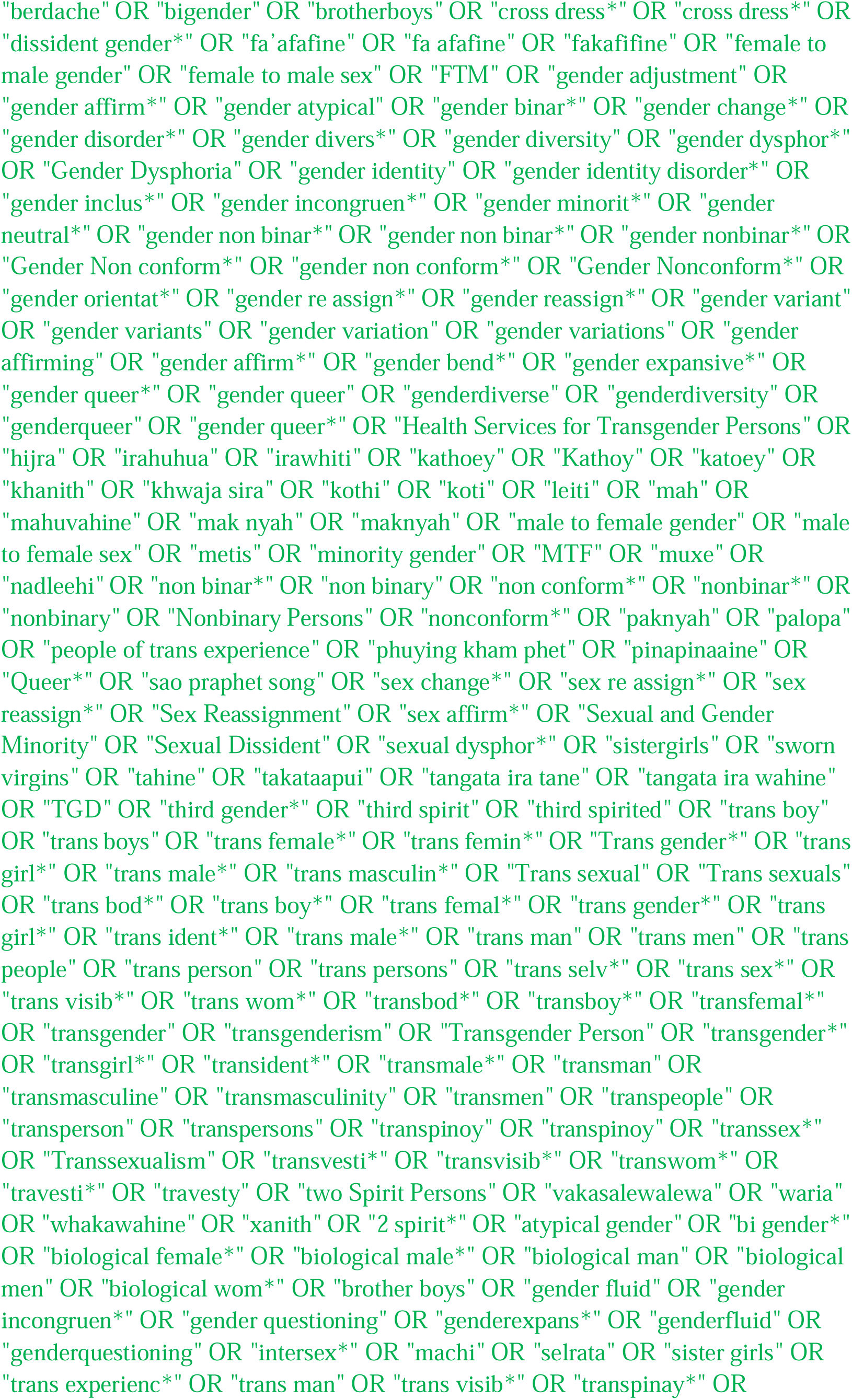
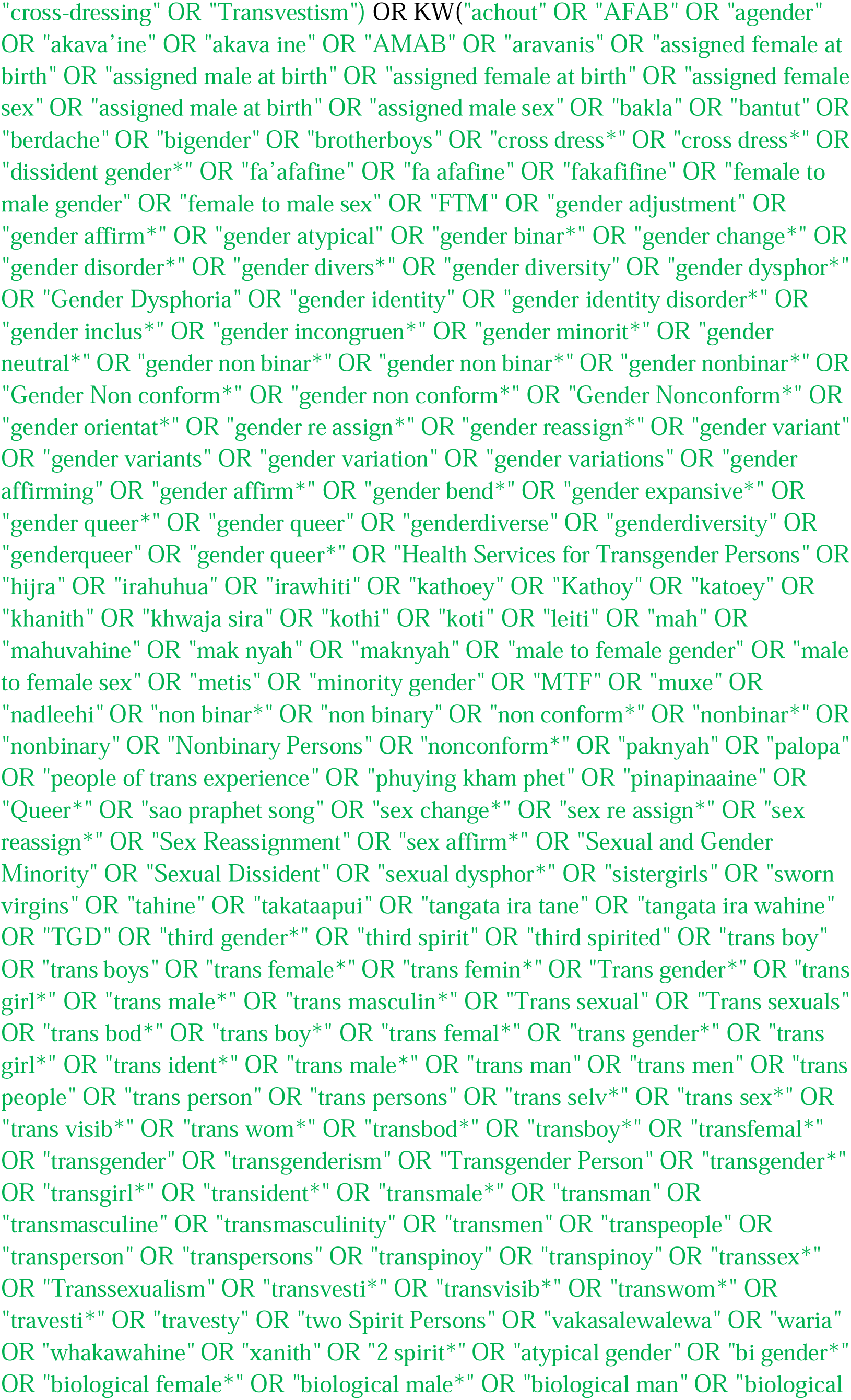
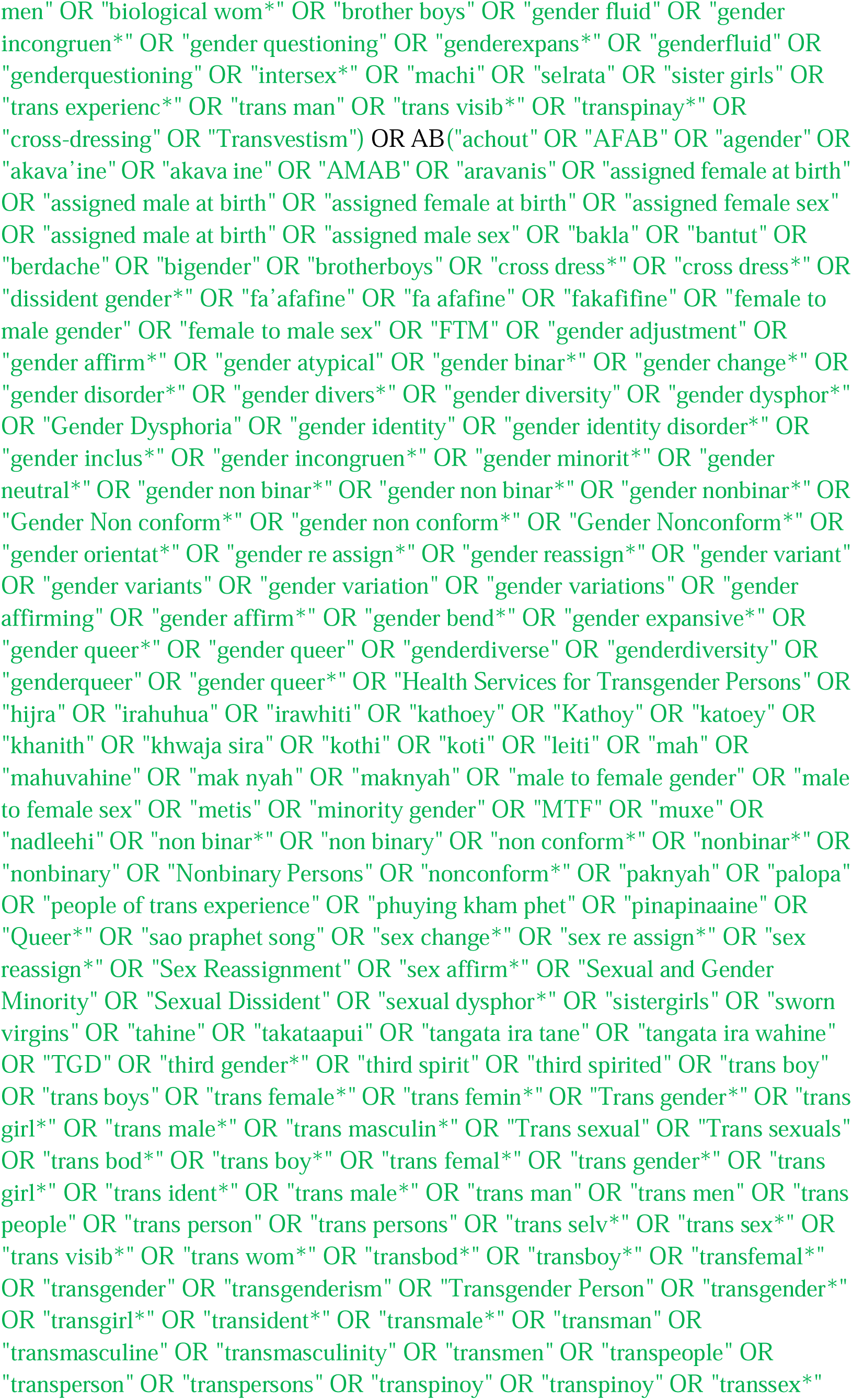
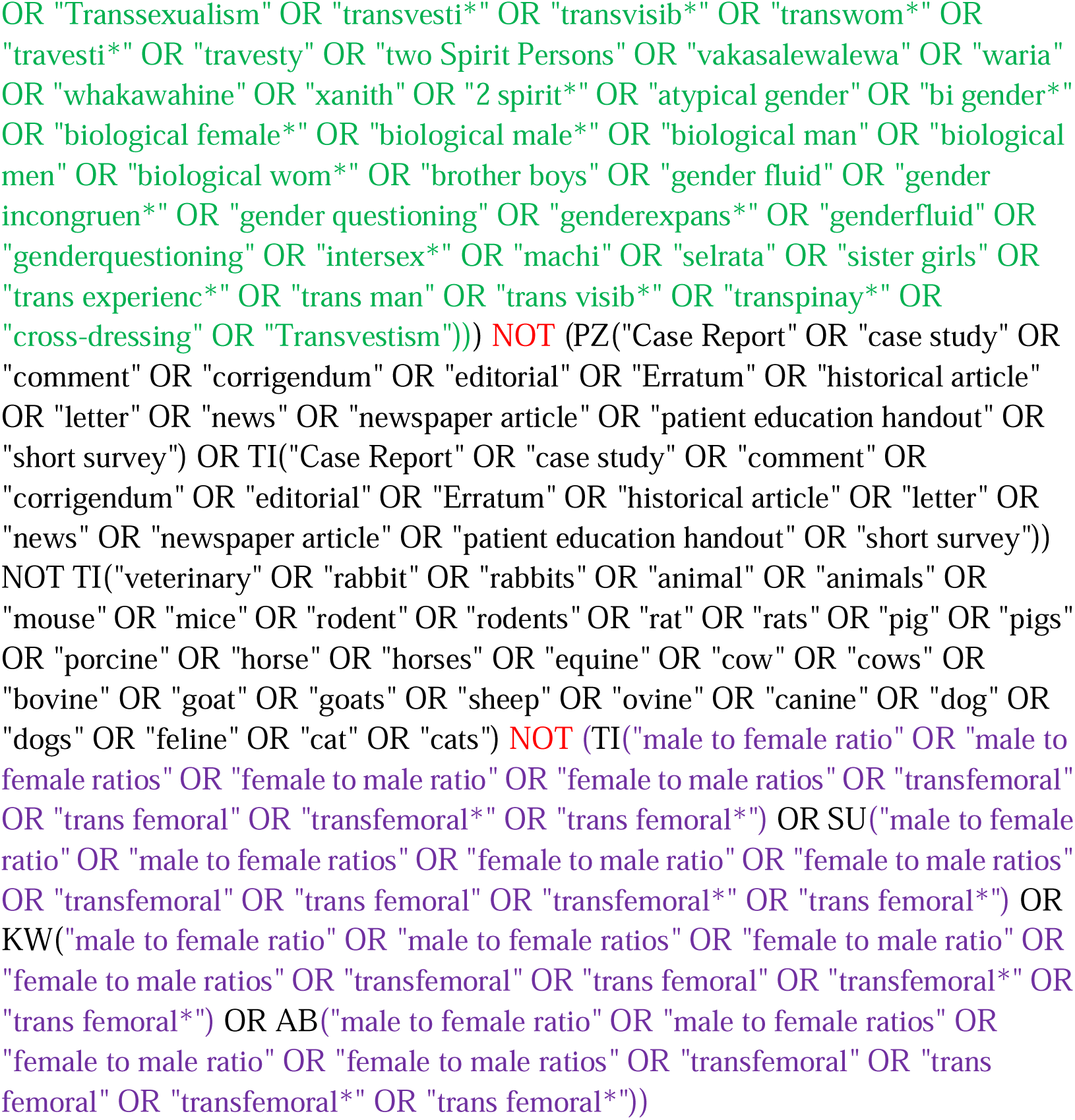

#### LILACS

https://lilacs.bvsalud.org/en/

Simplified version - three search strings

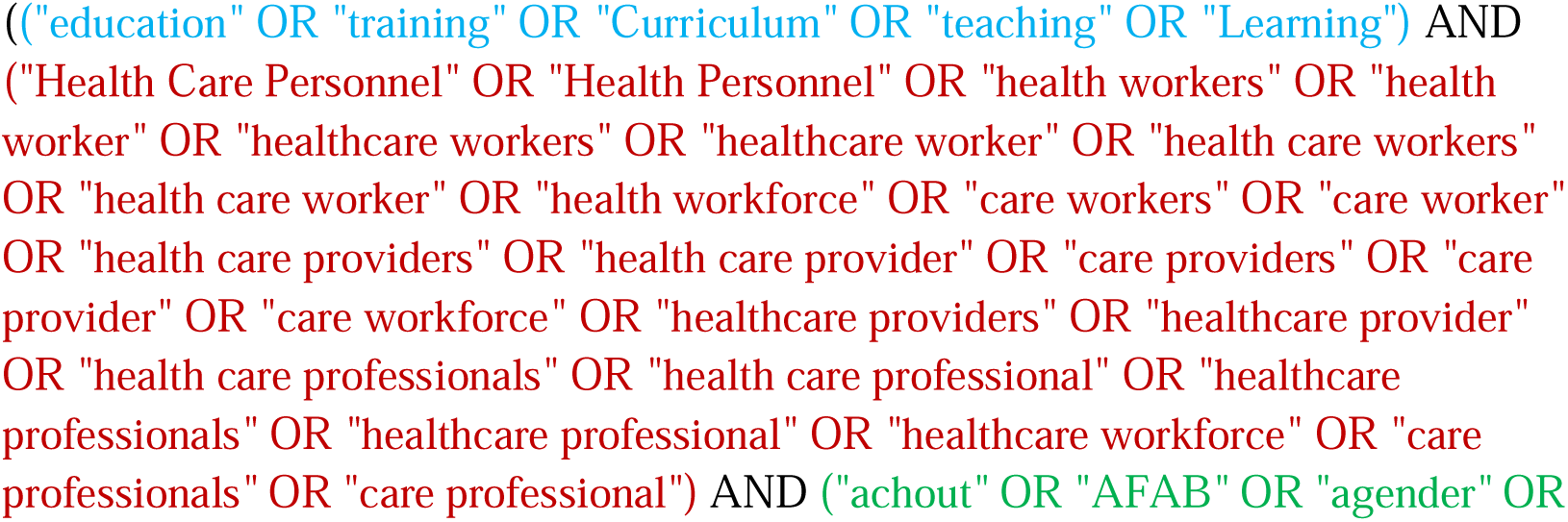
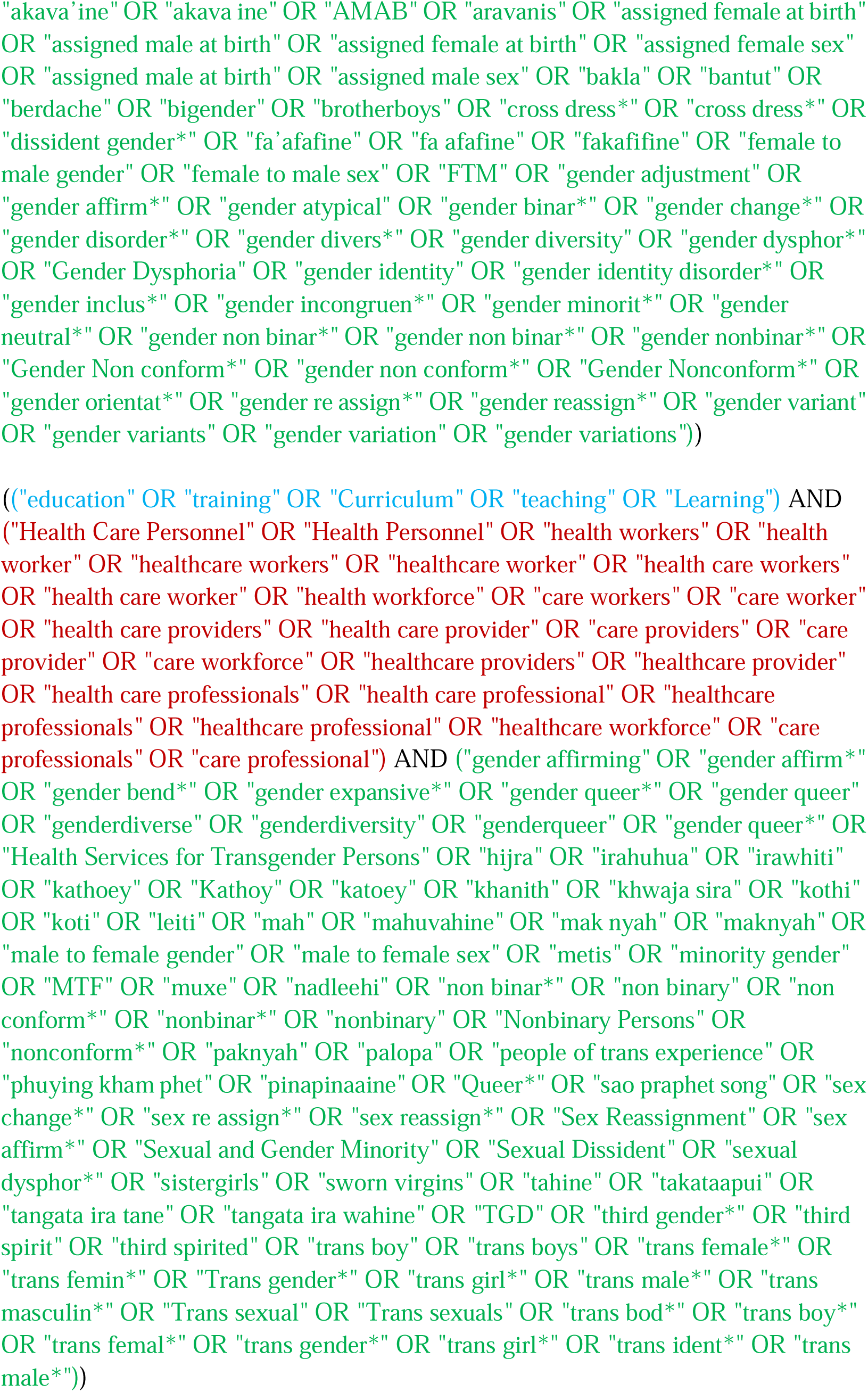
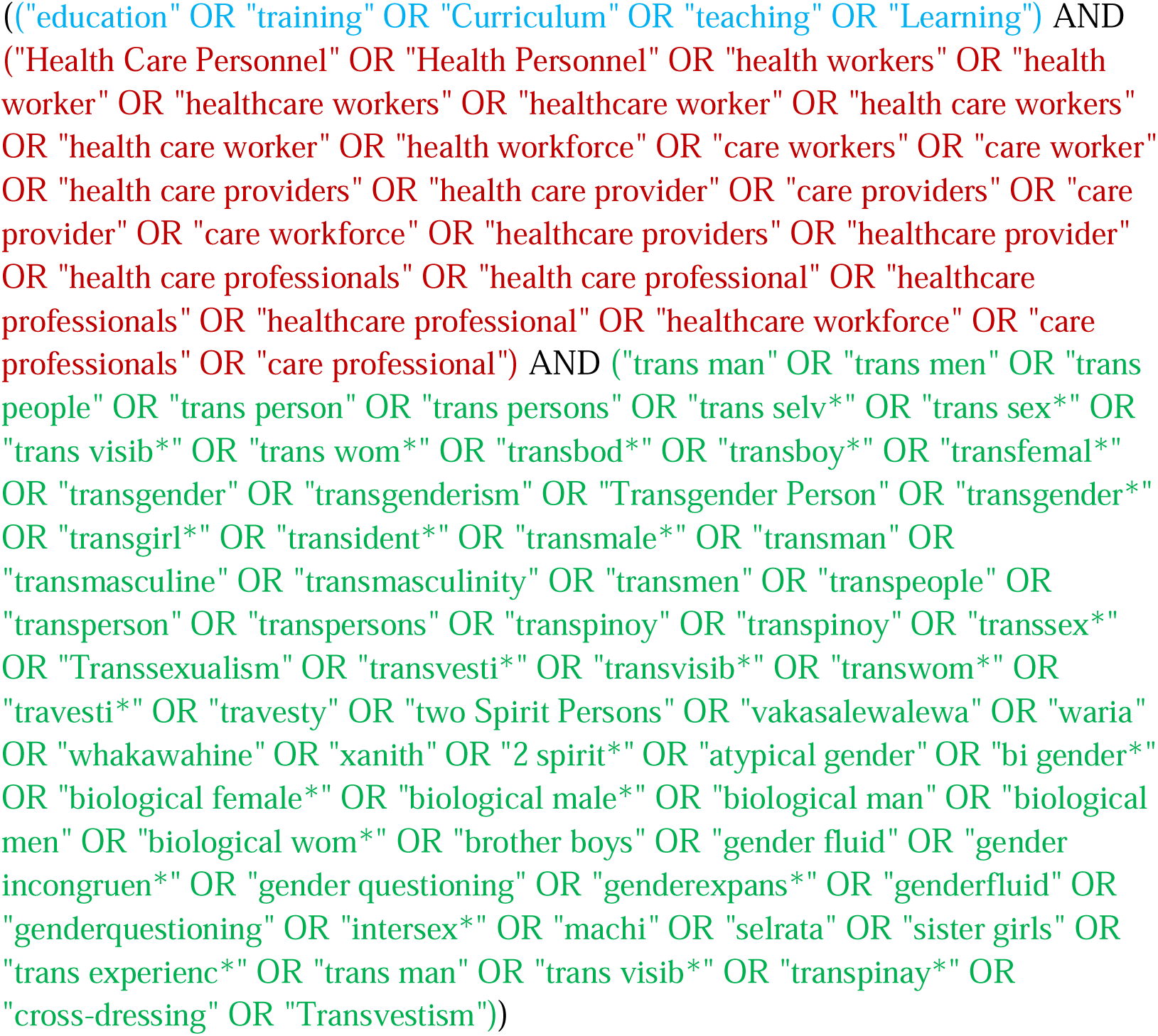

**Appendix 2:**
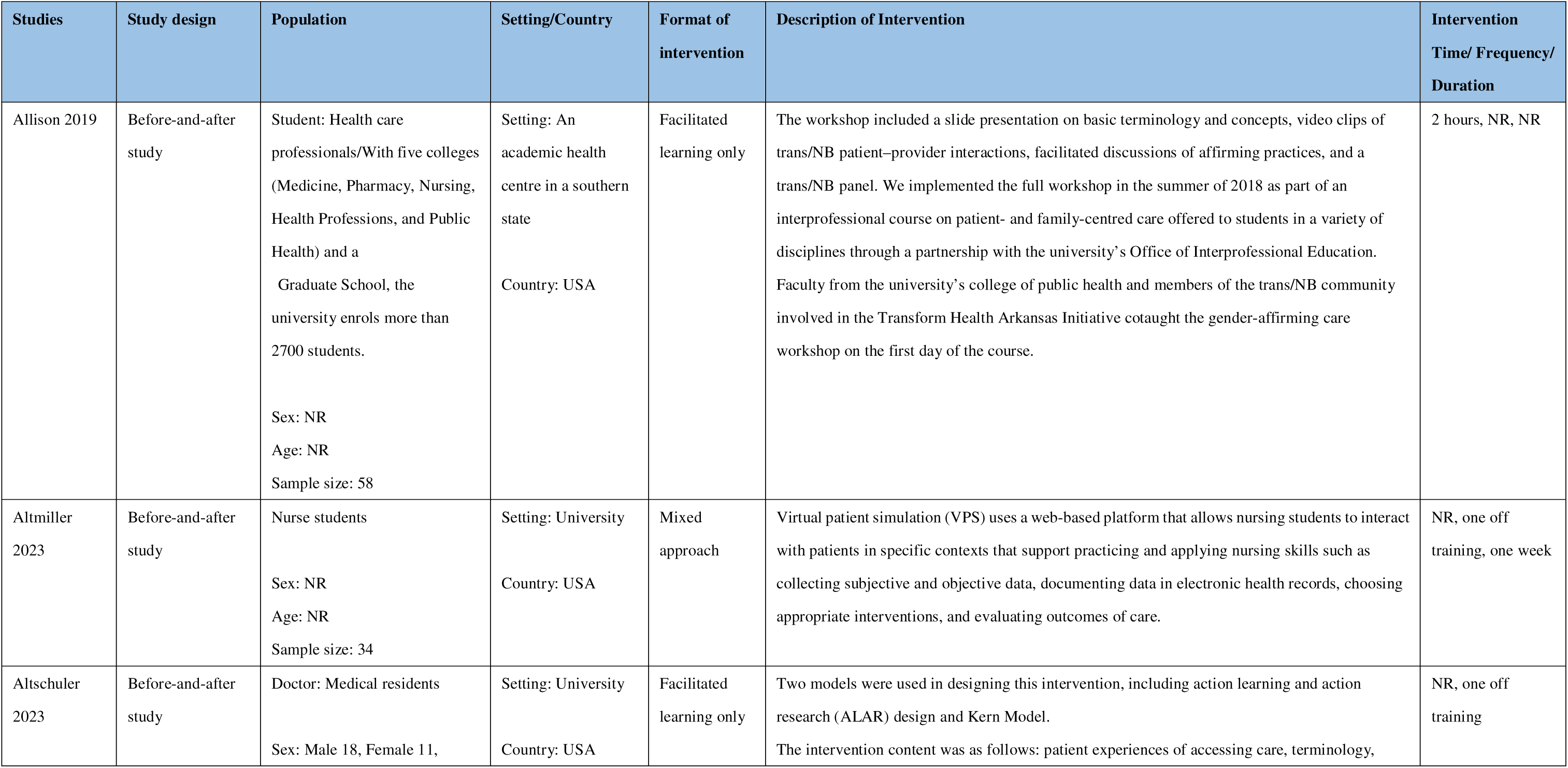
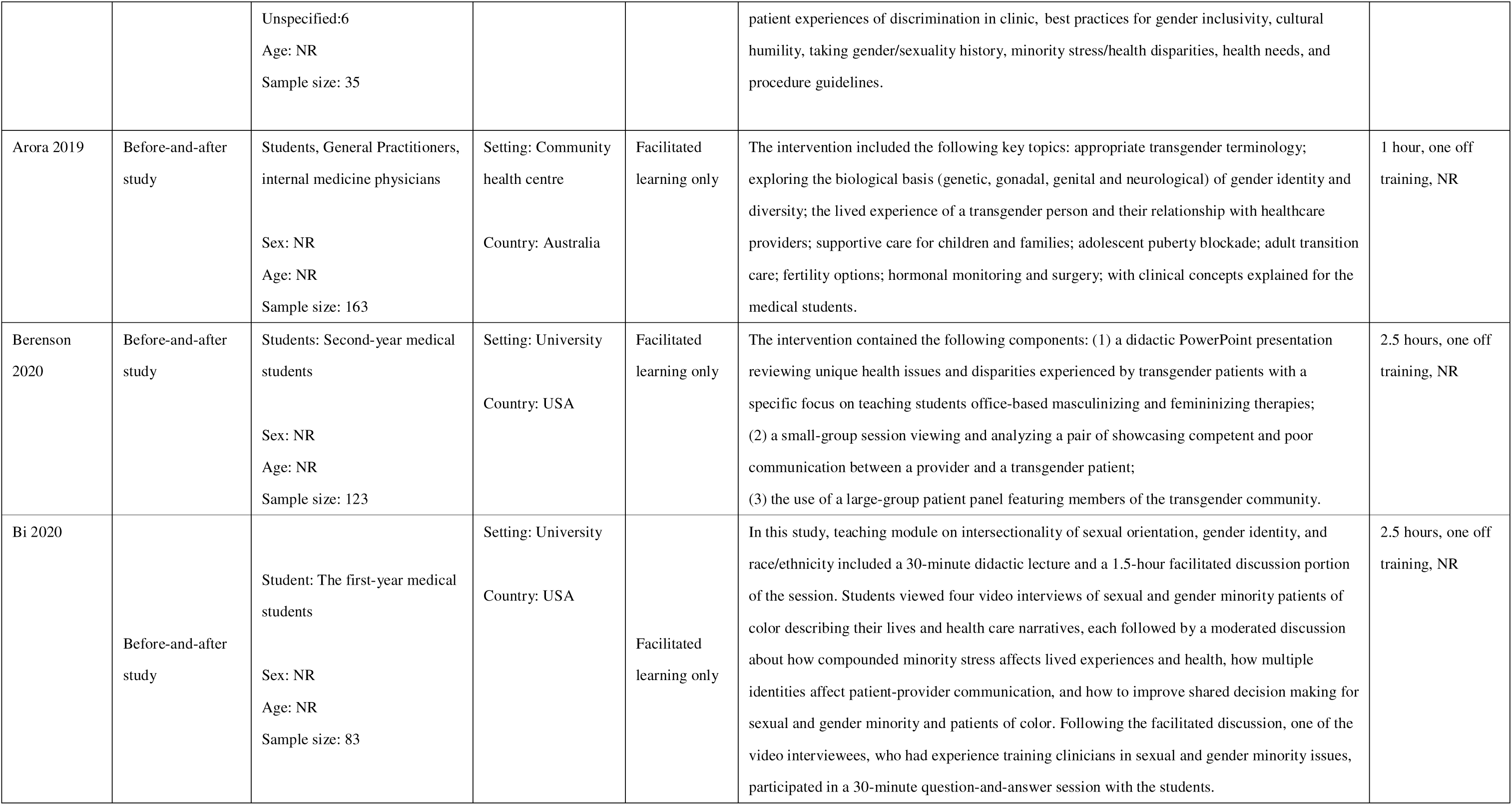
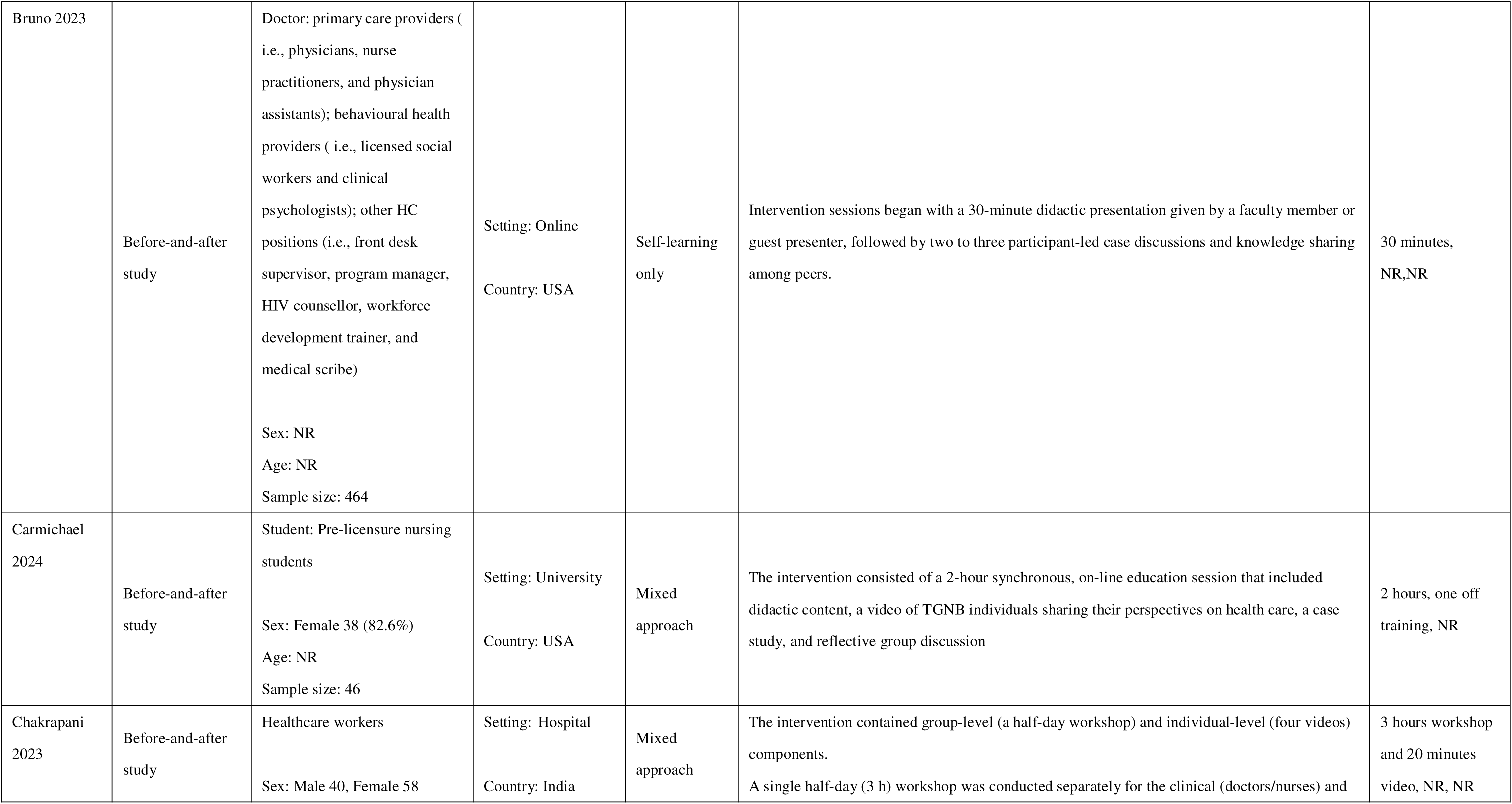
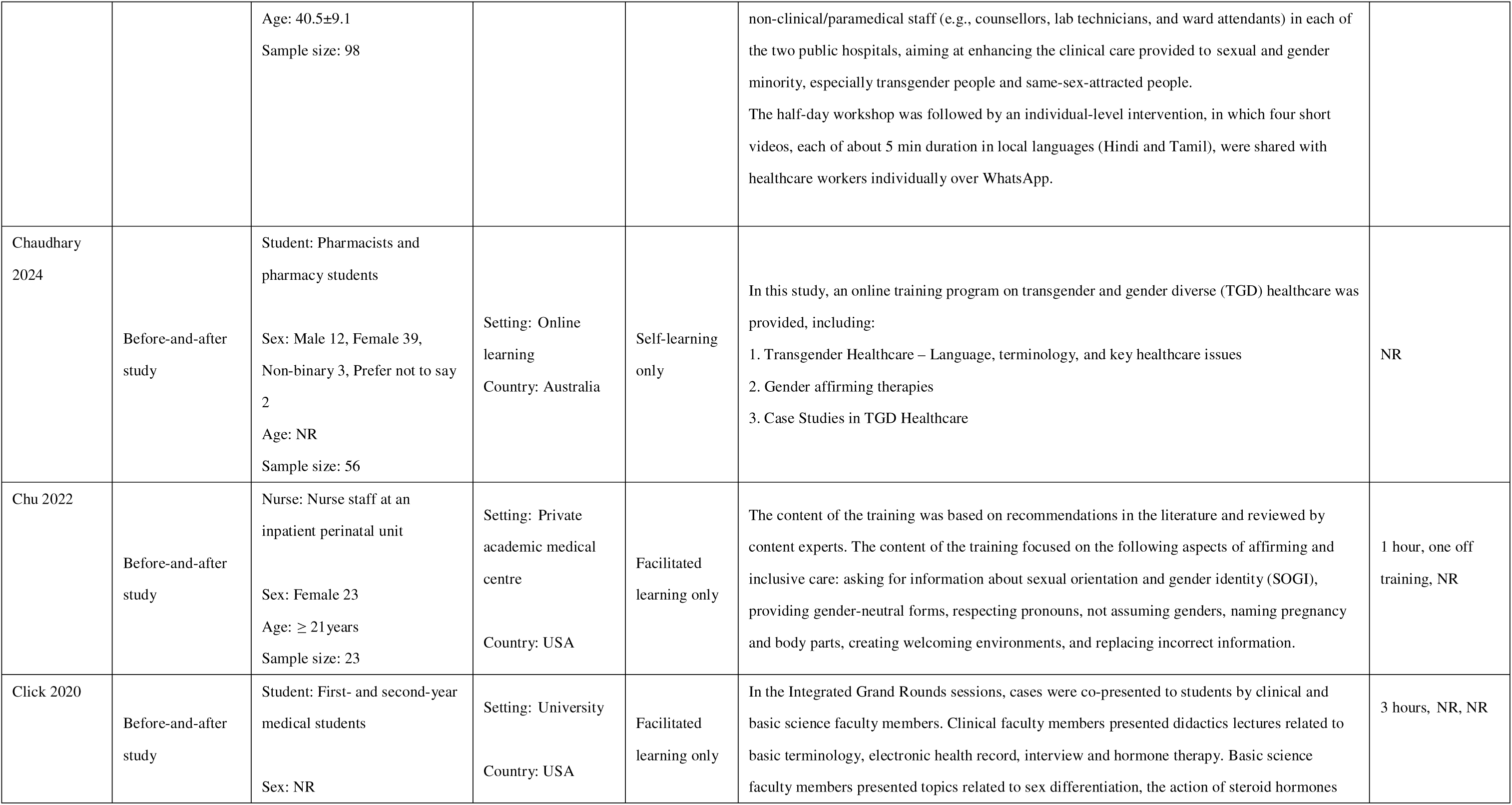
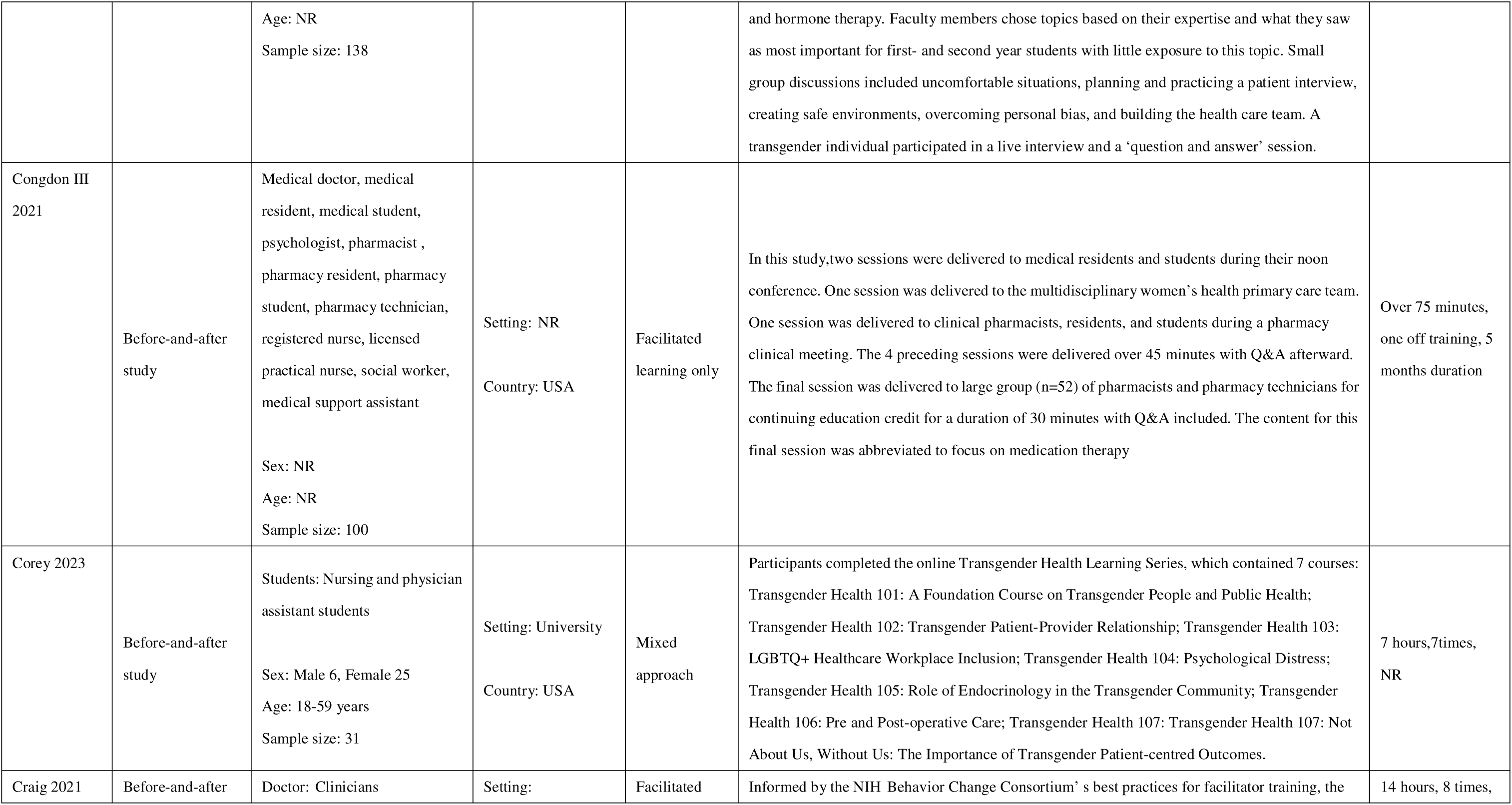
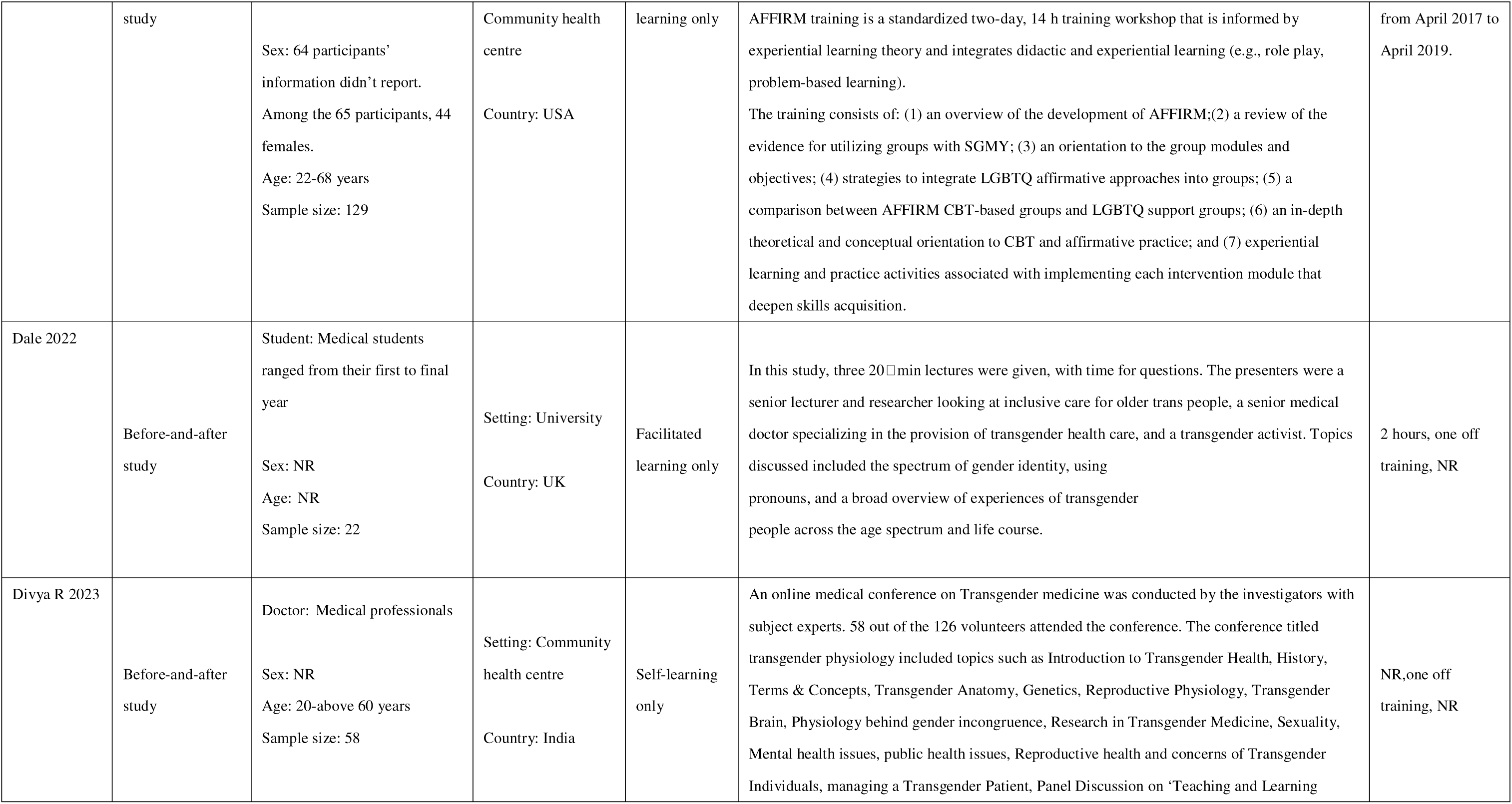
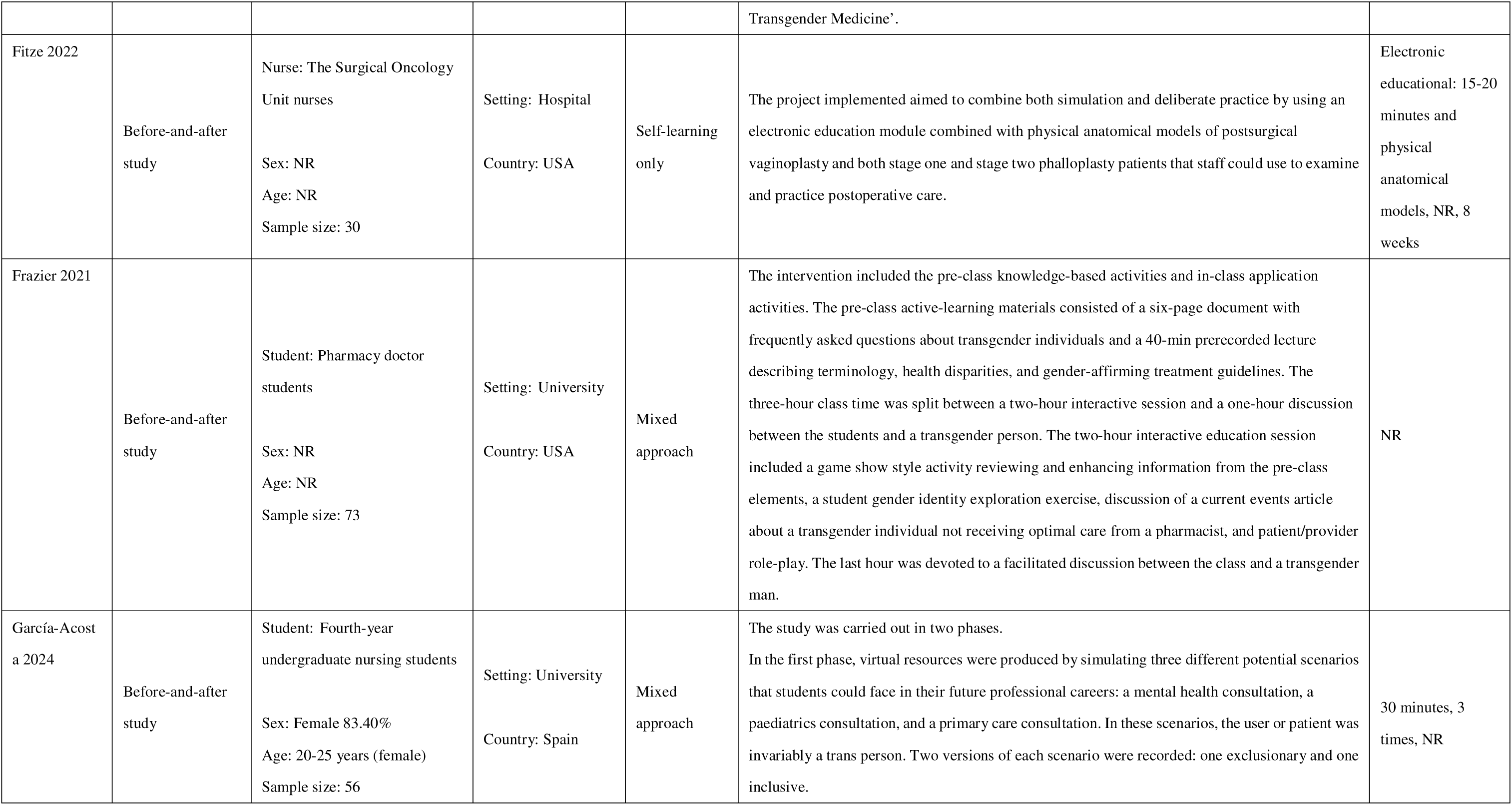
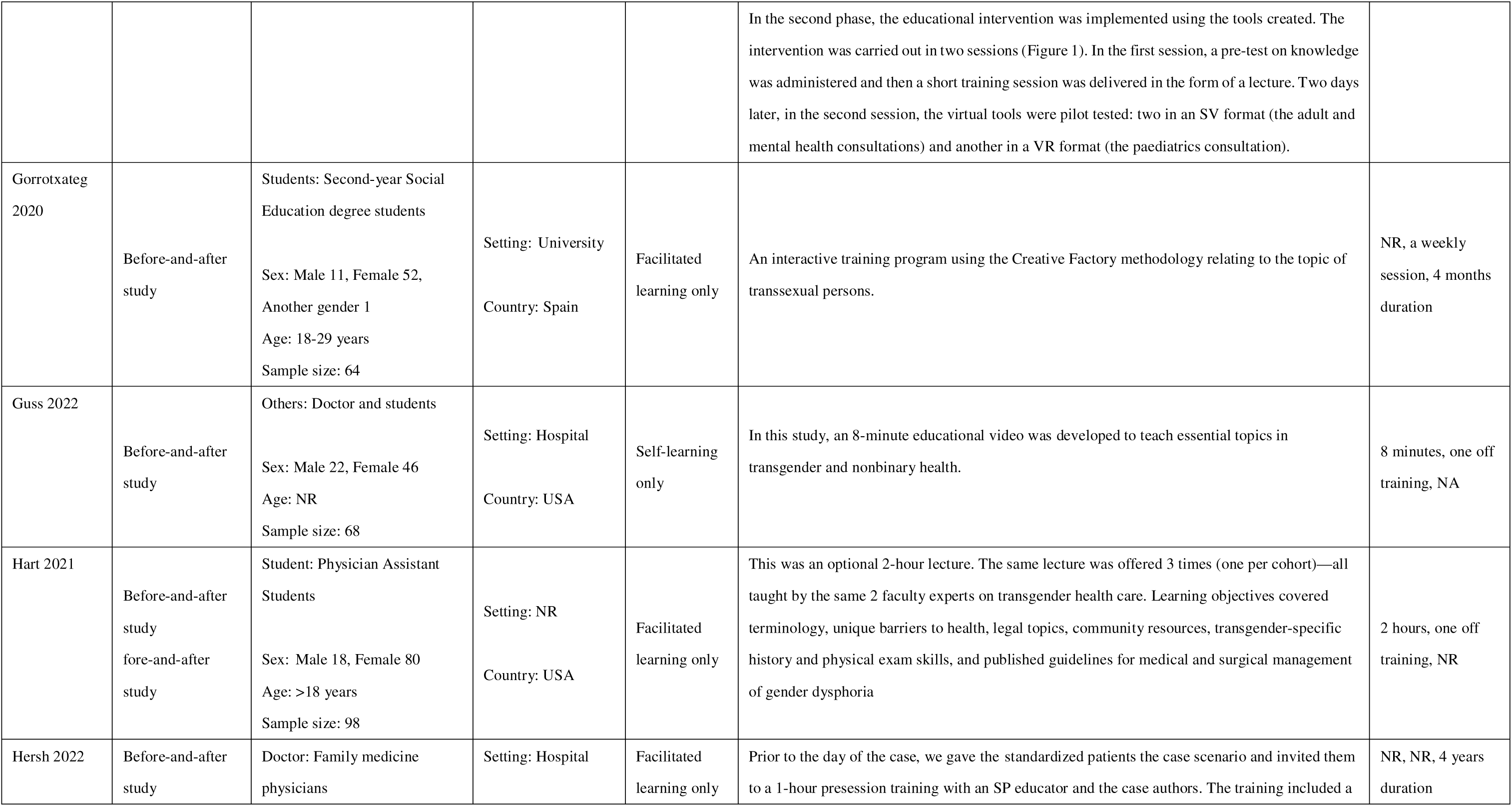
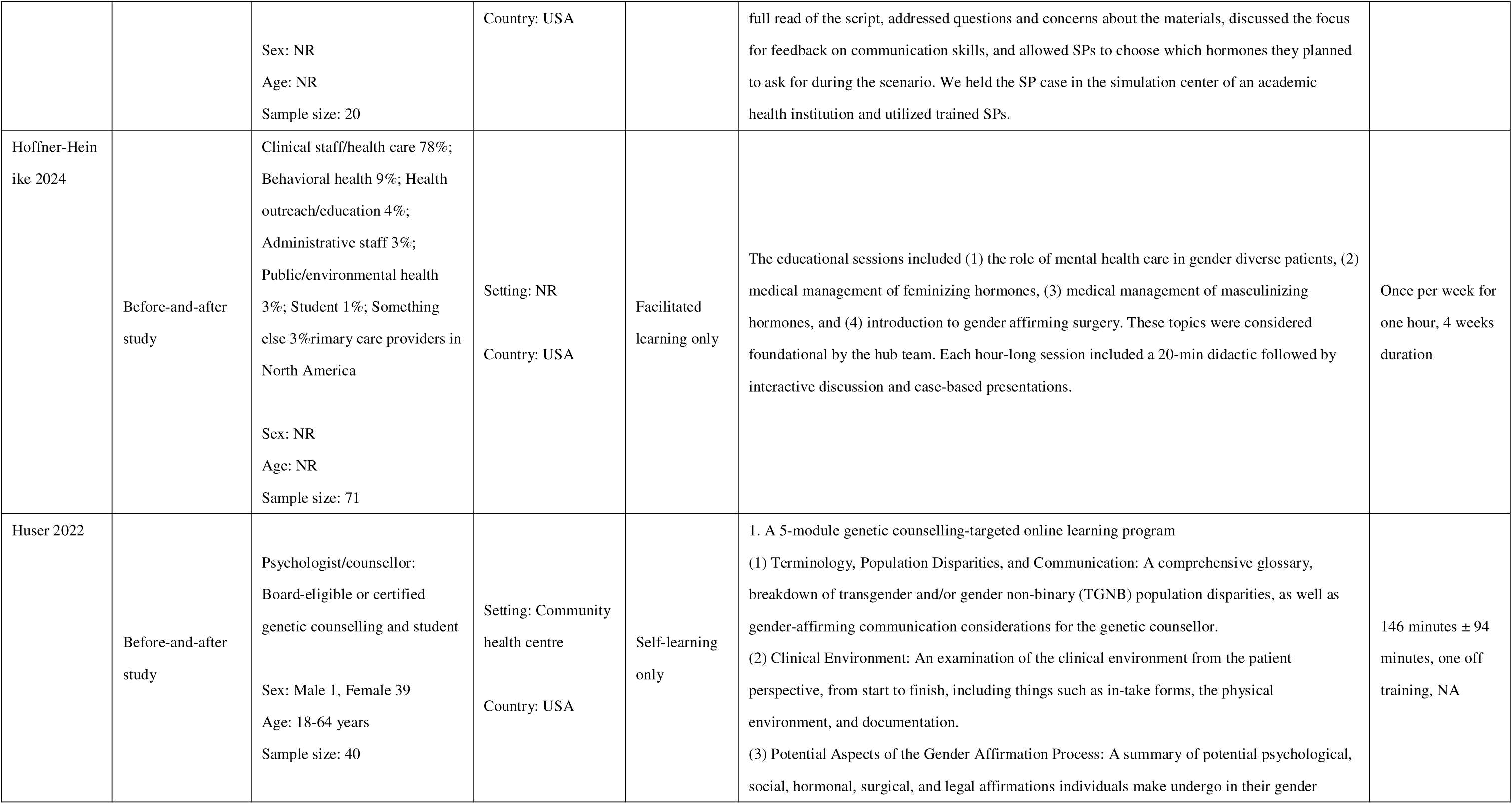
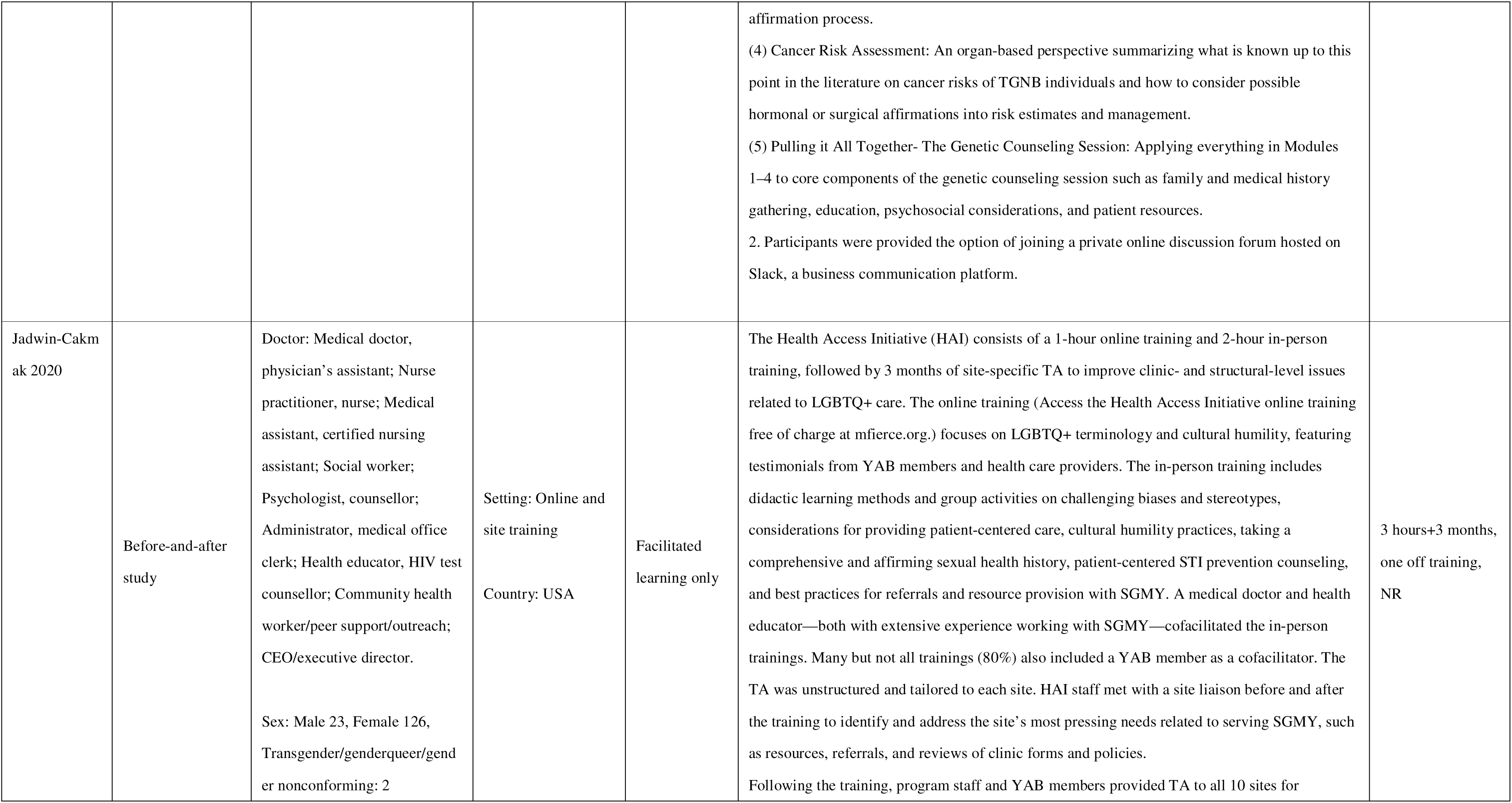
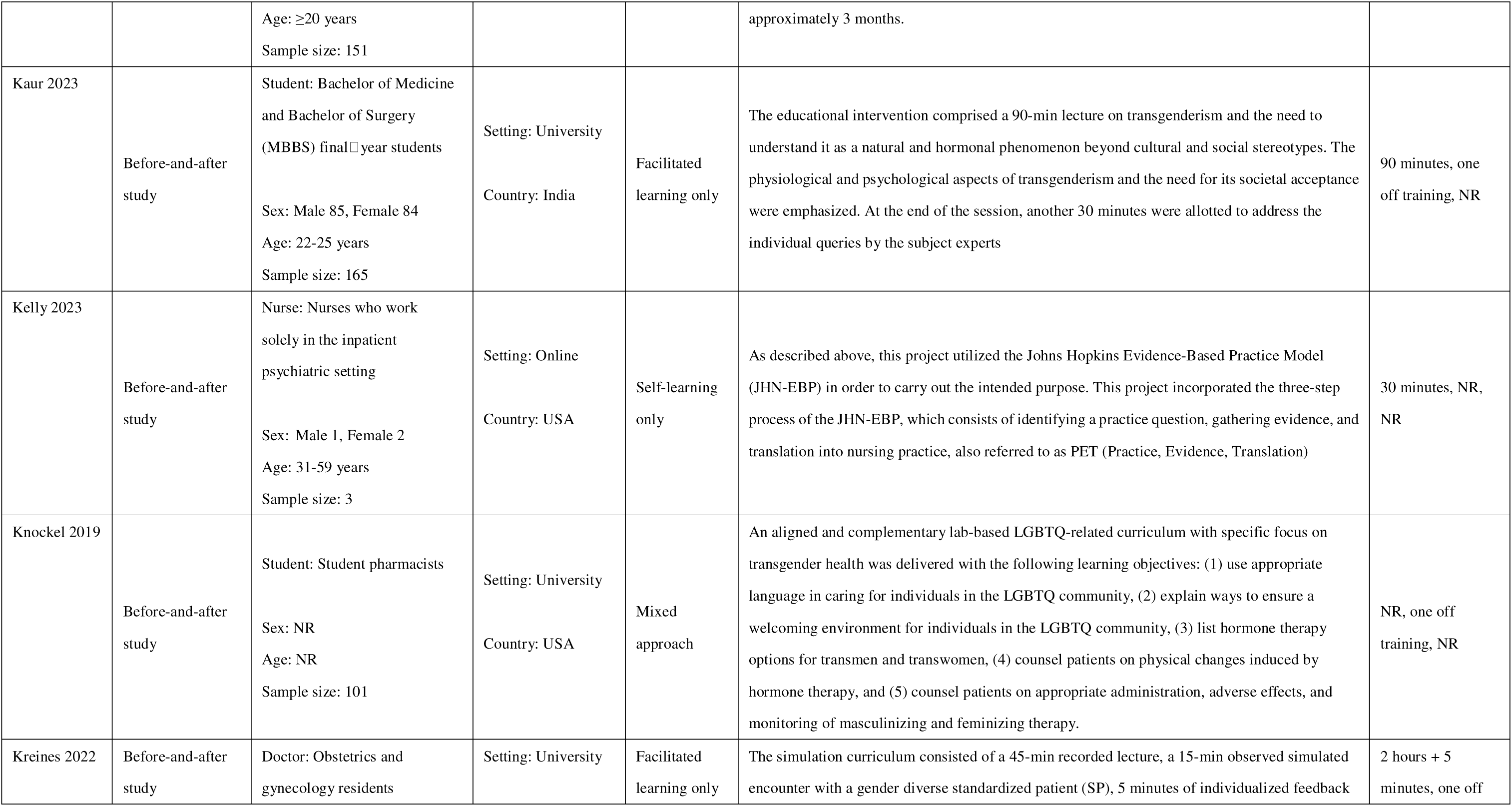
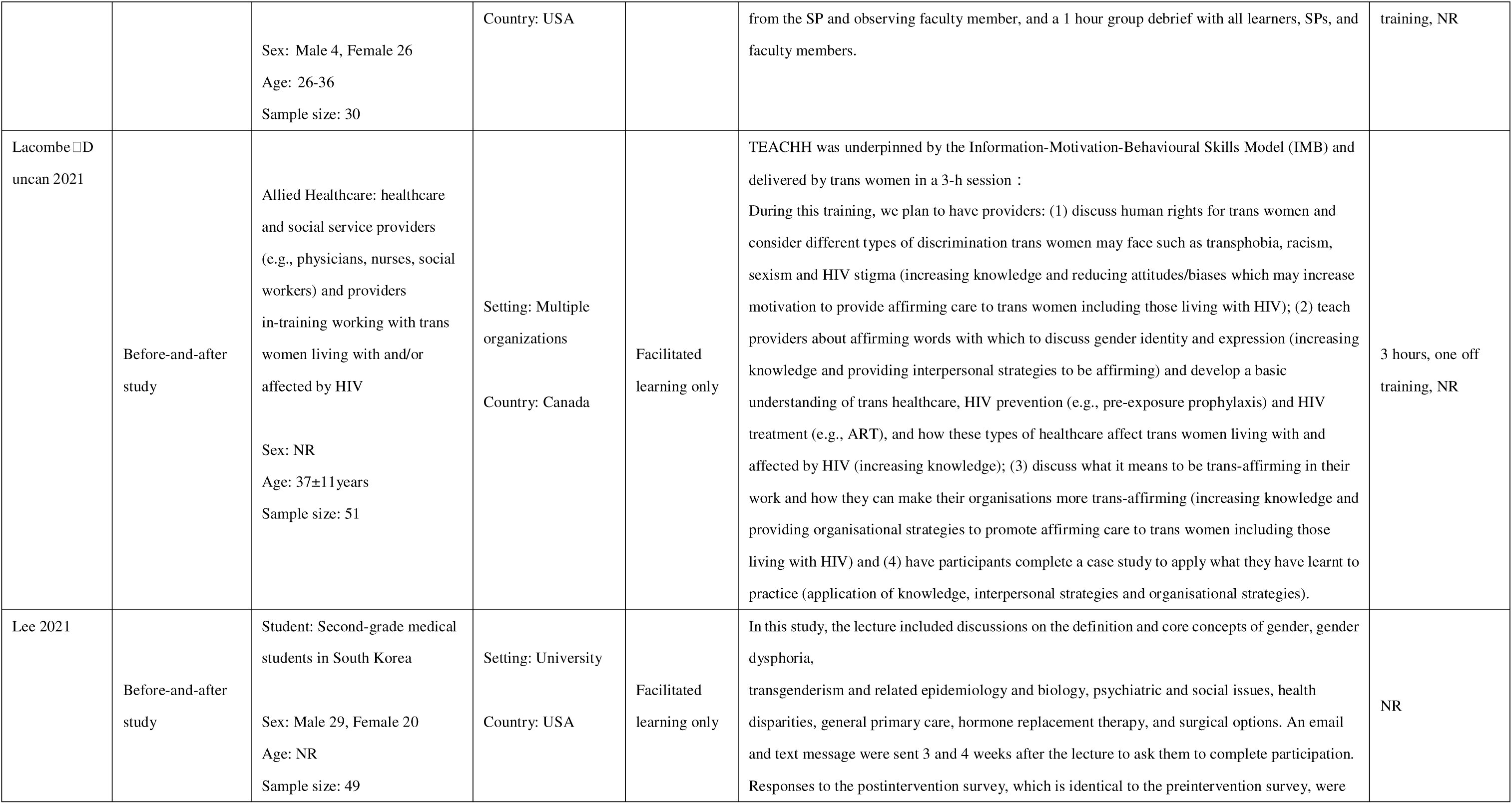
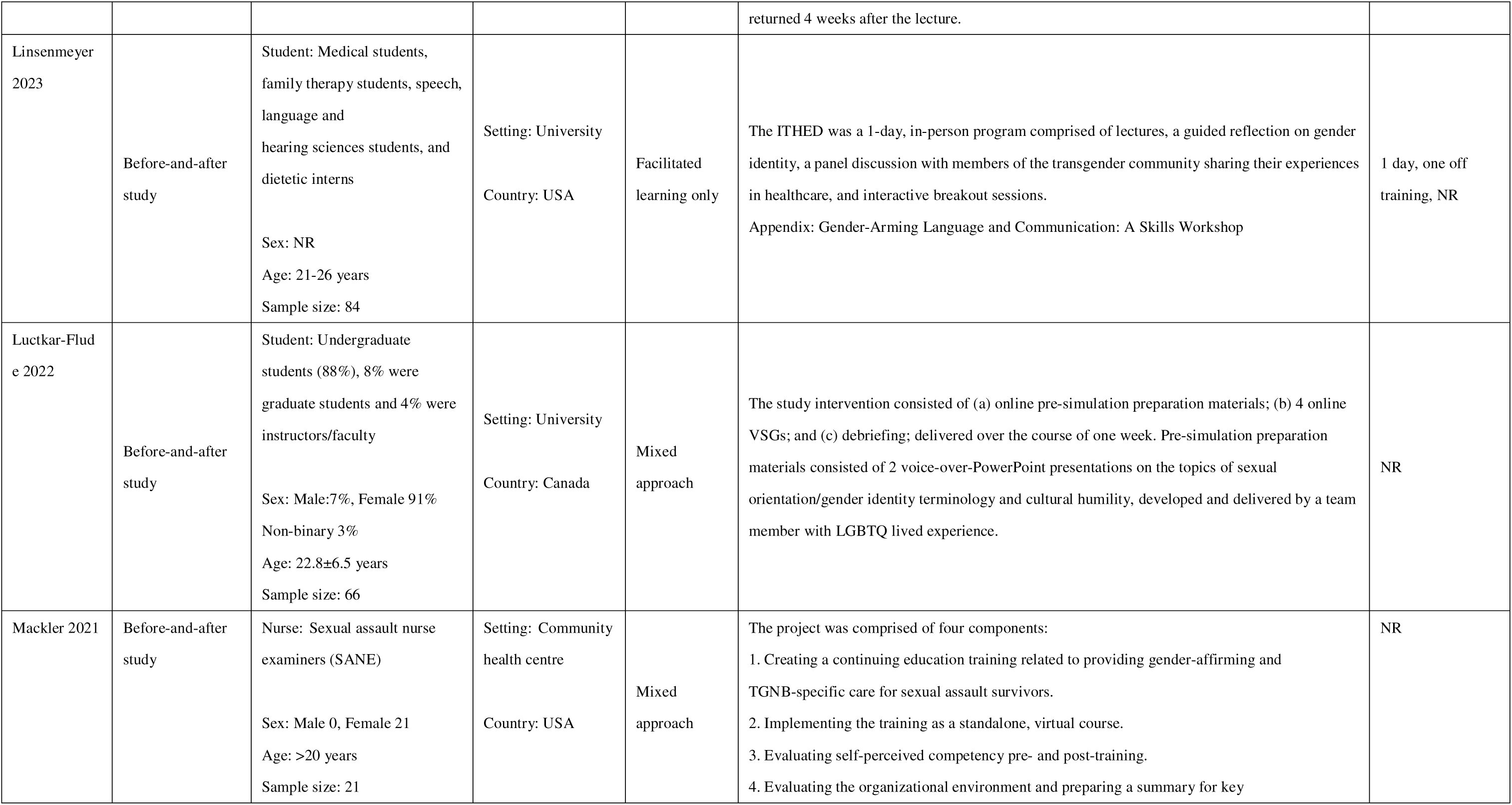
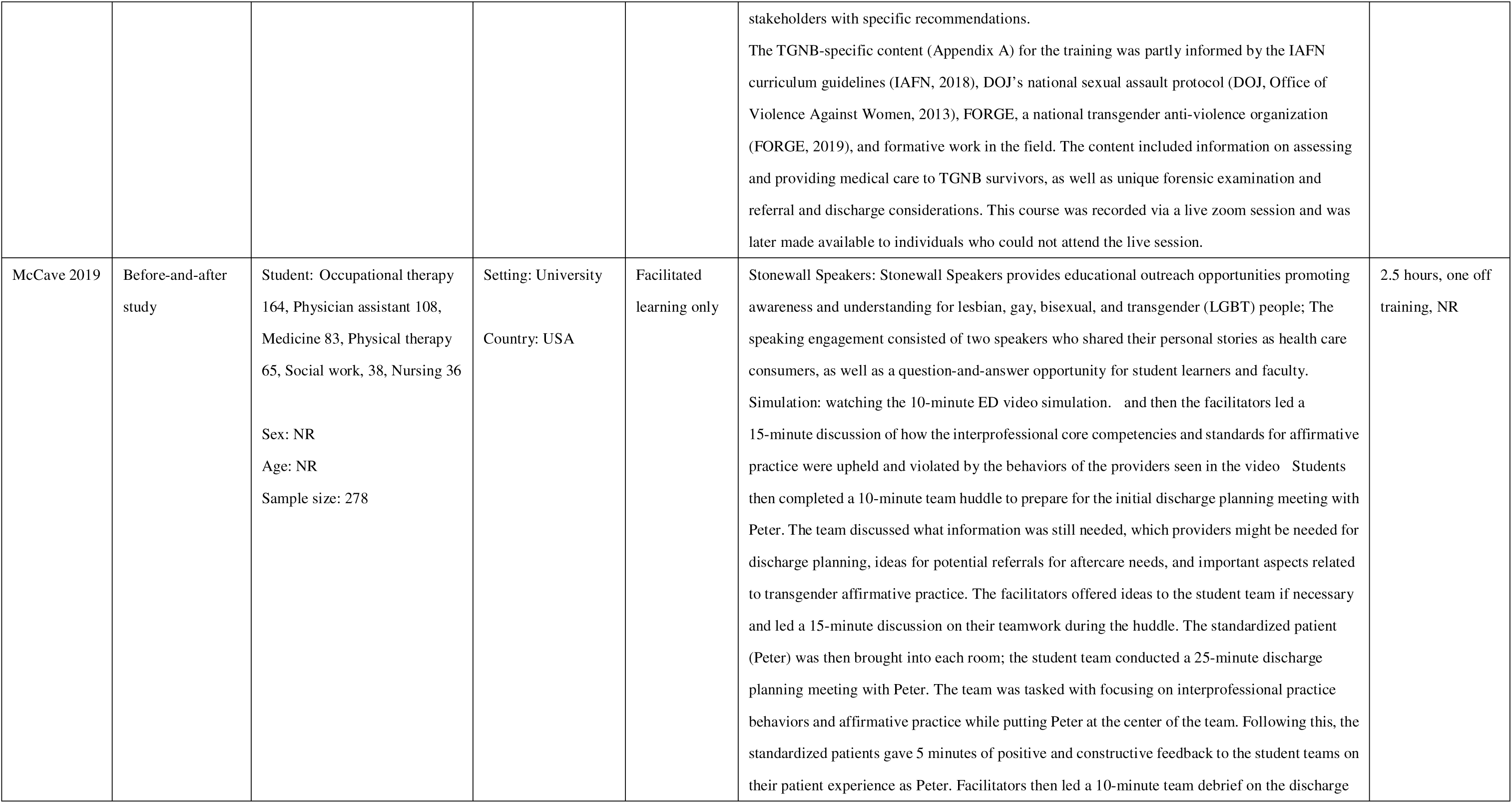
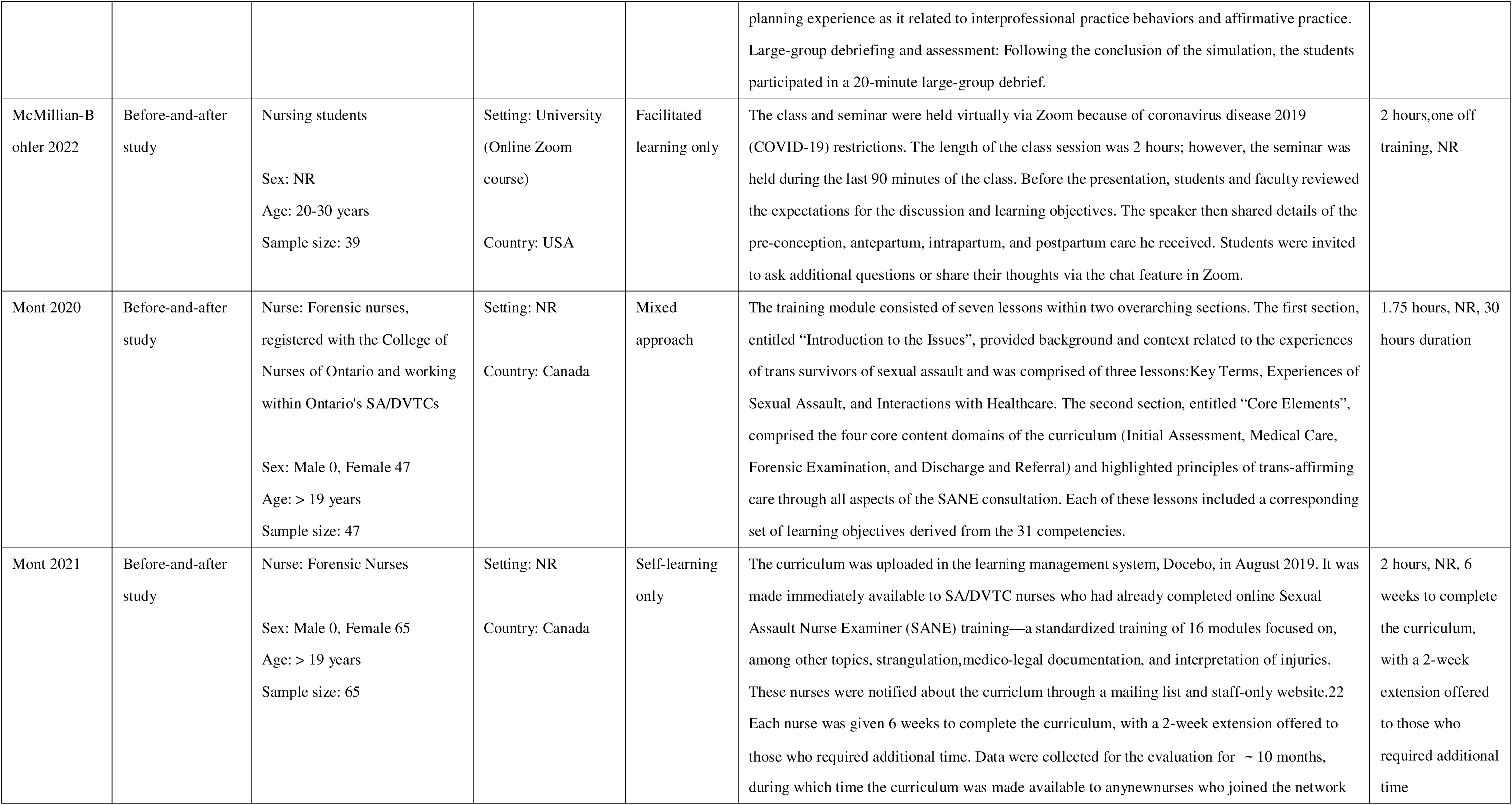
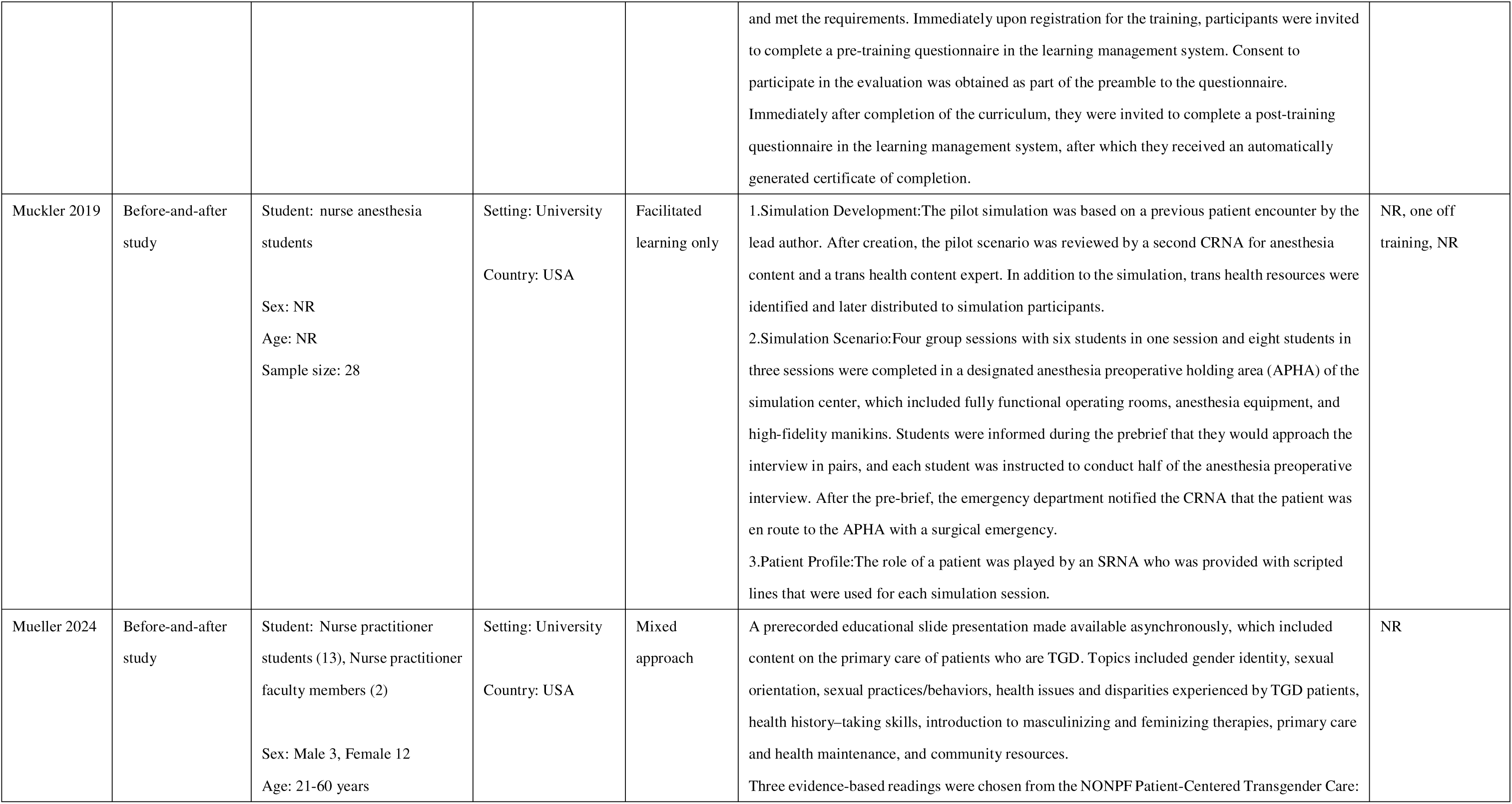
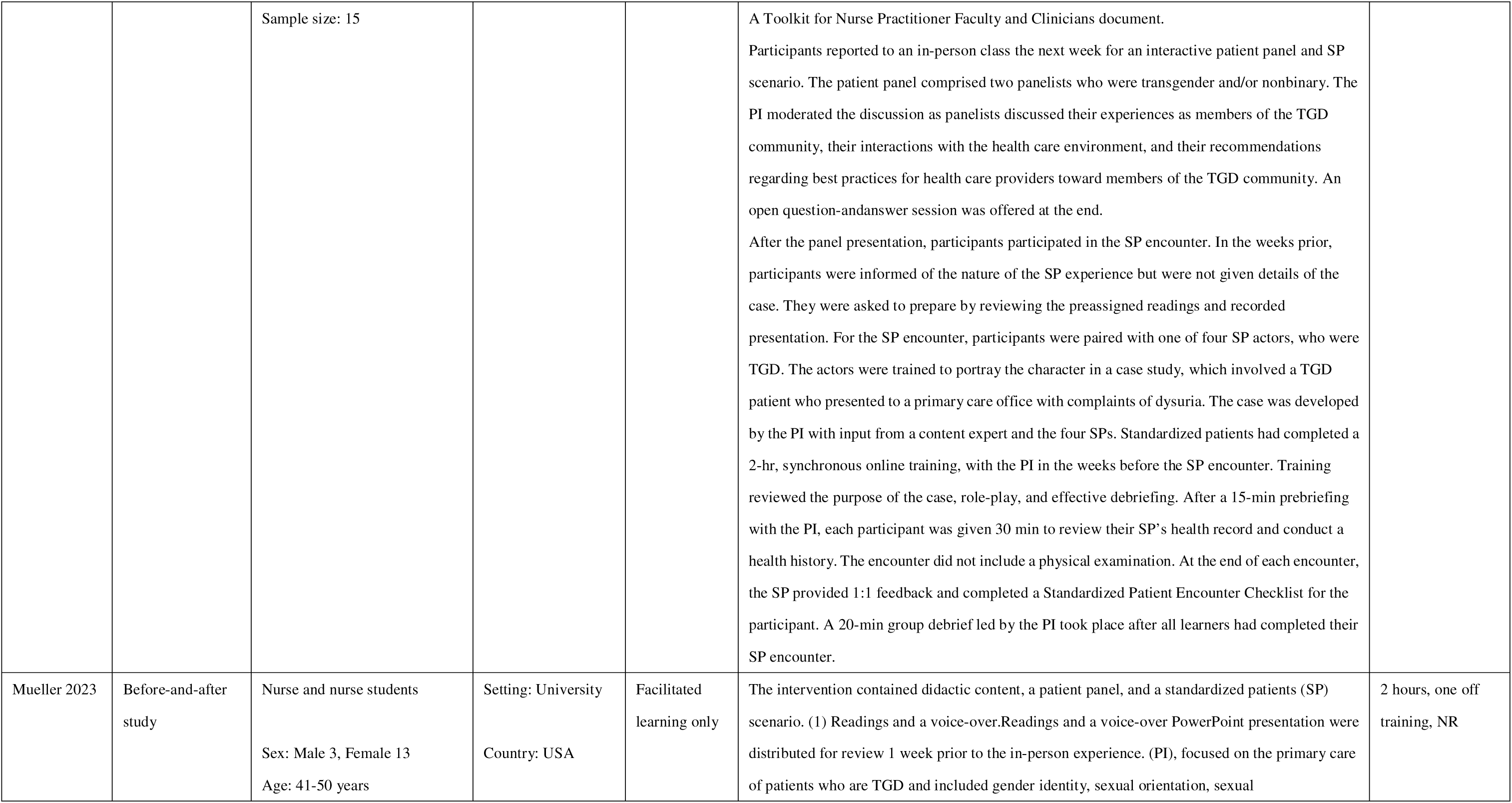
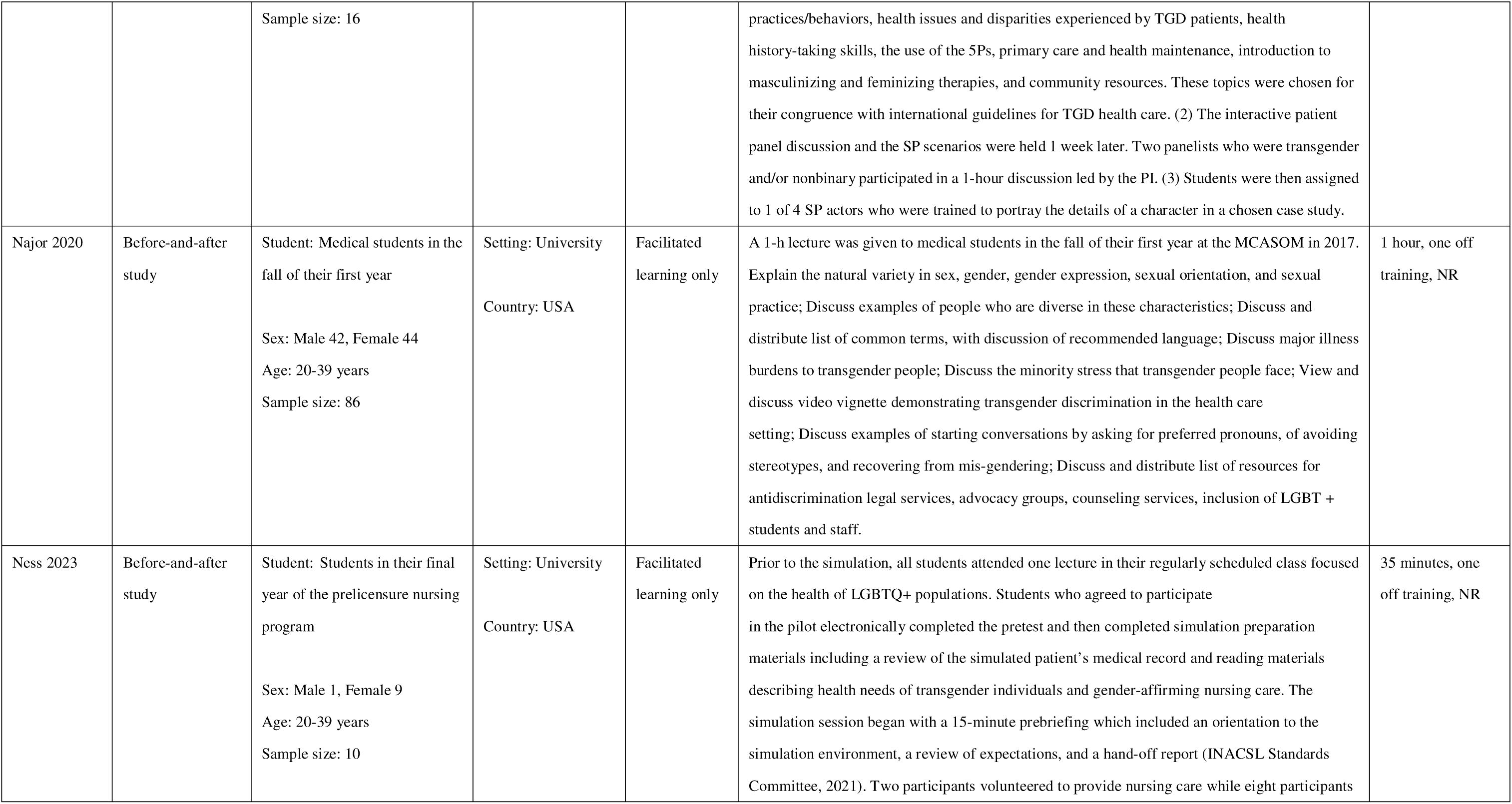
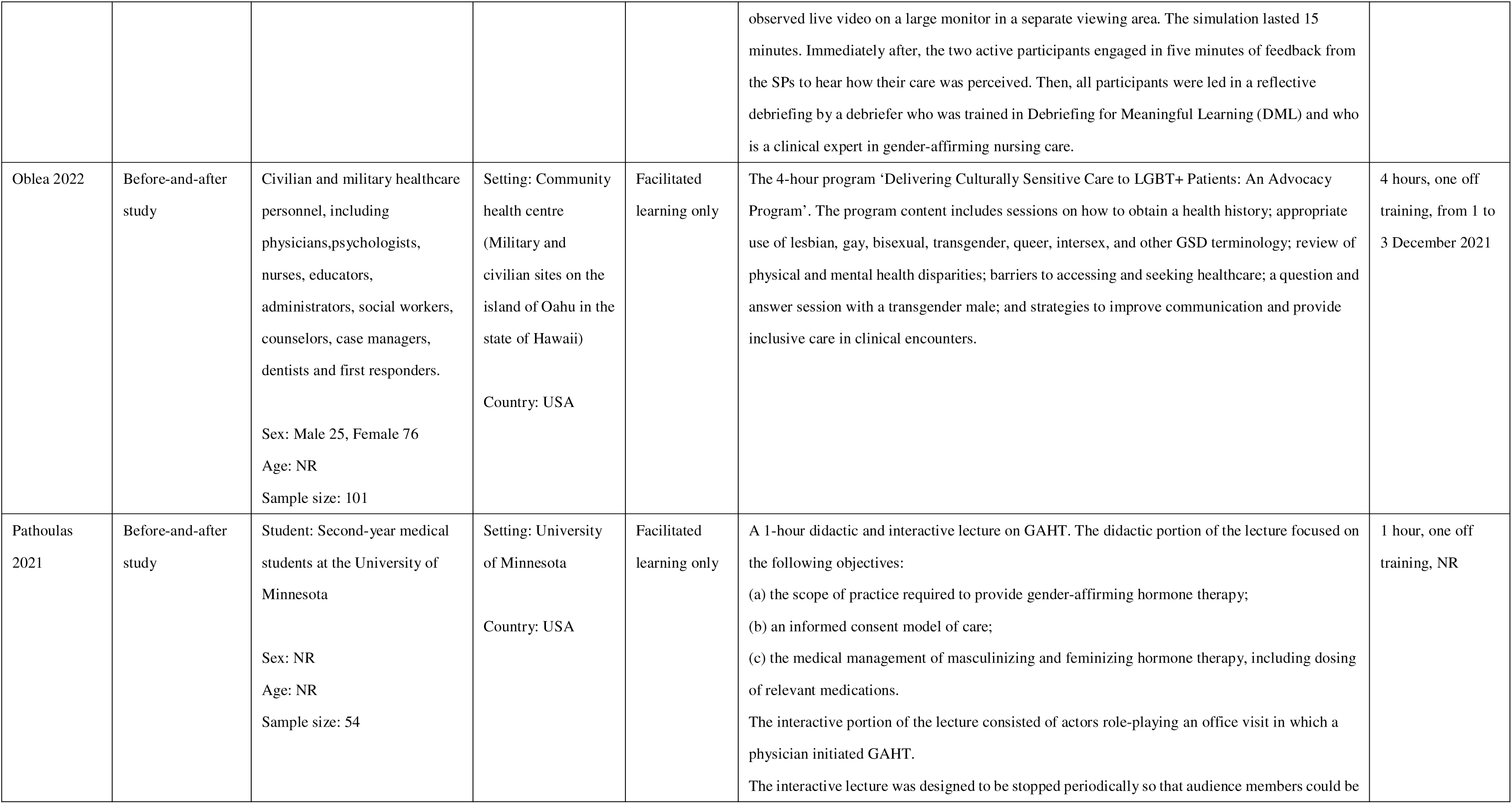
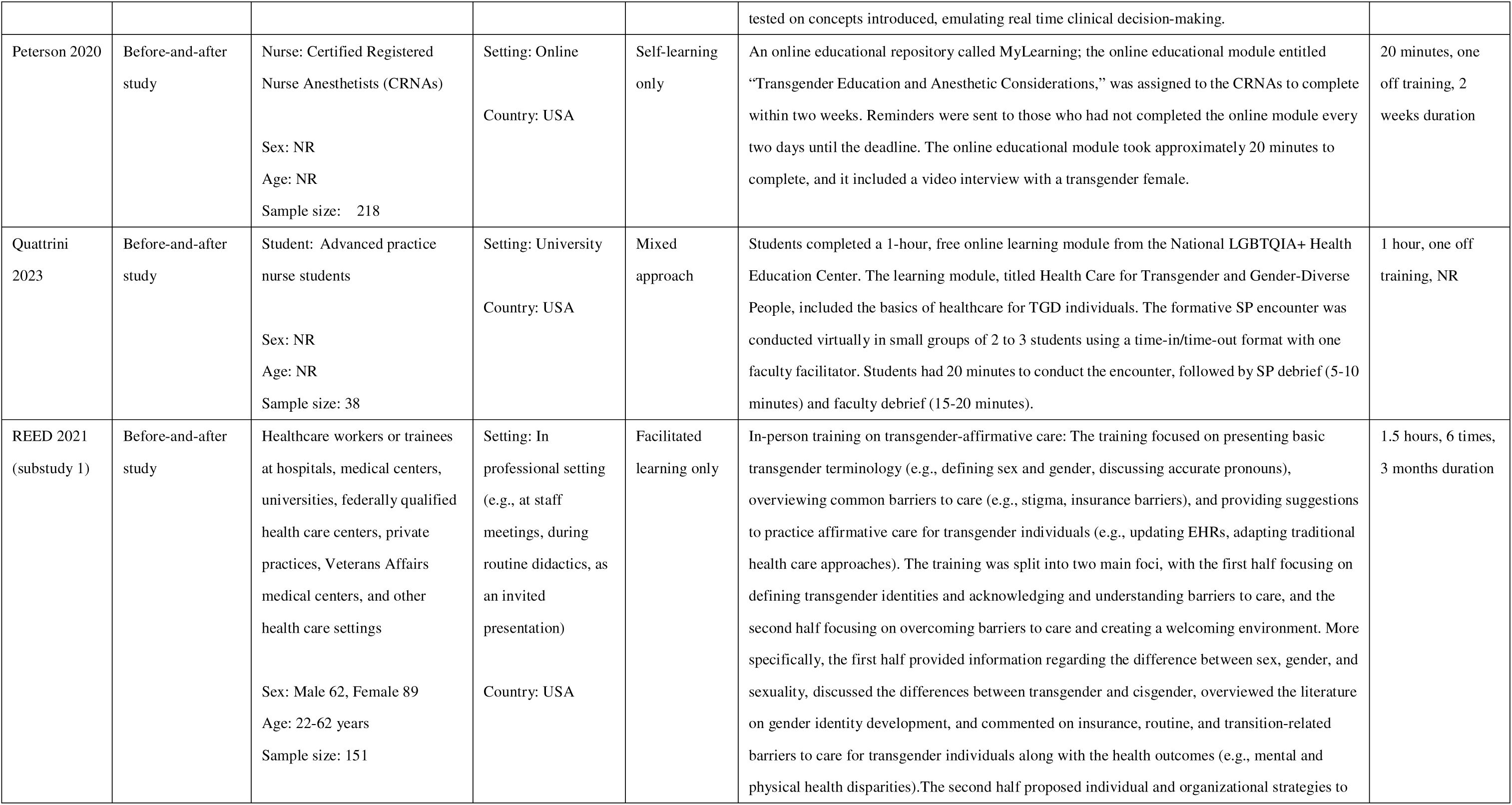
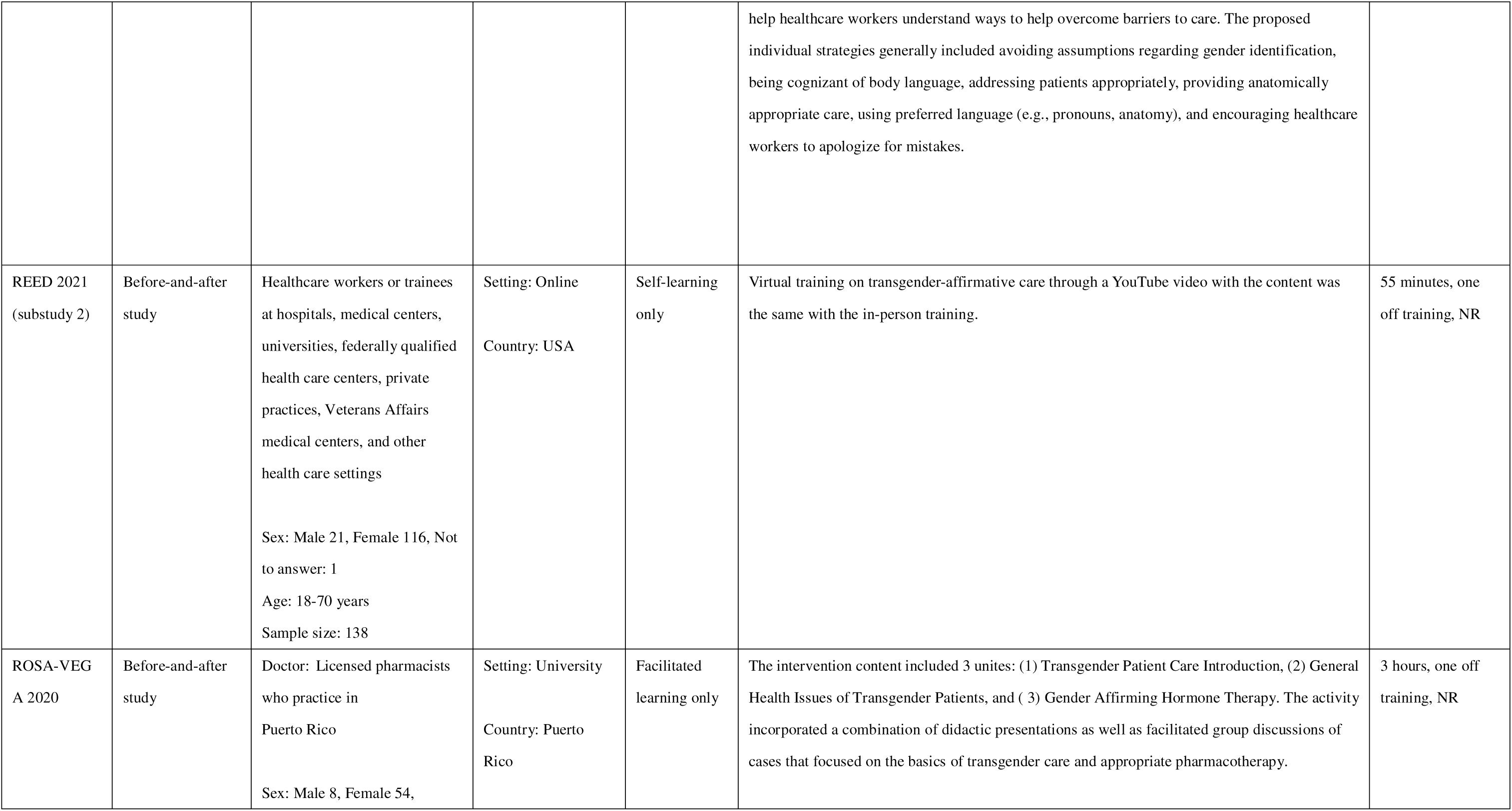
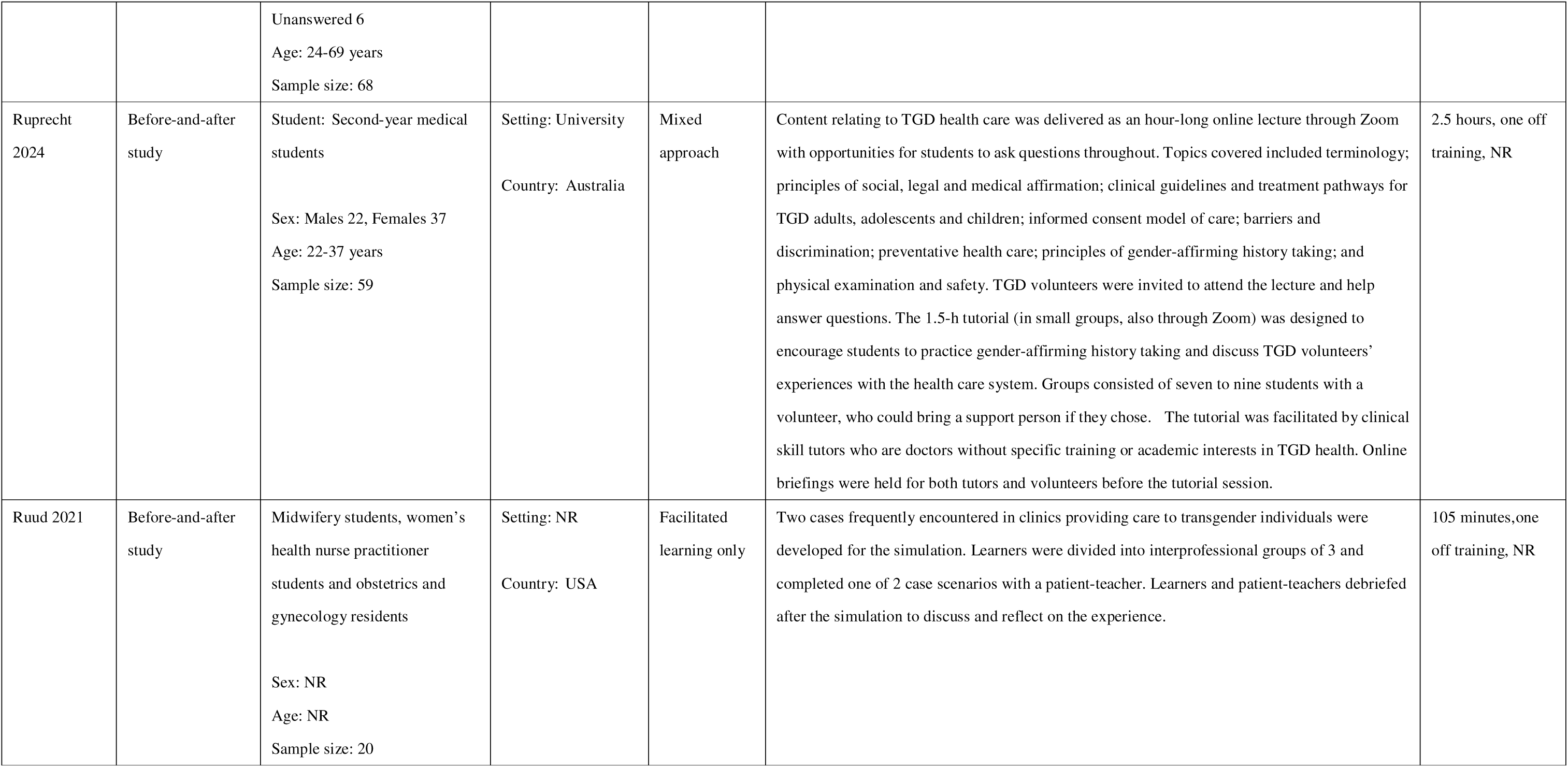
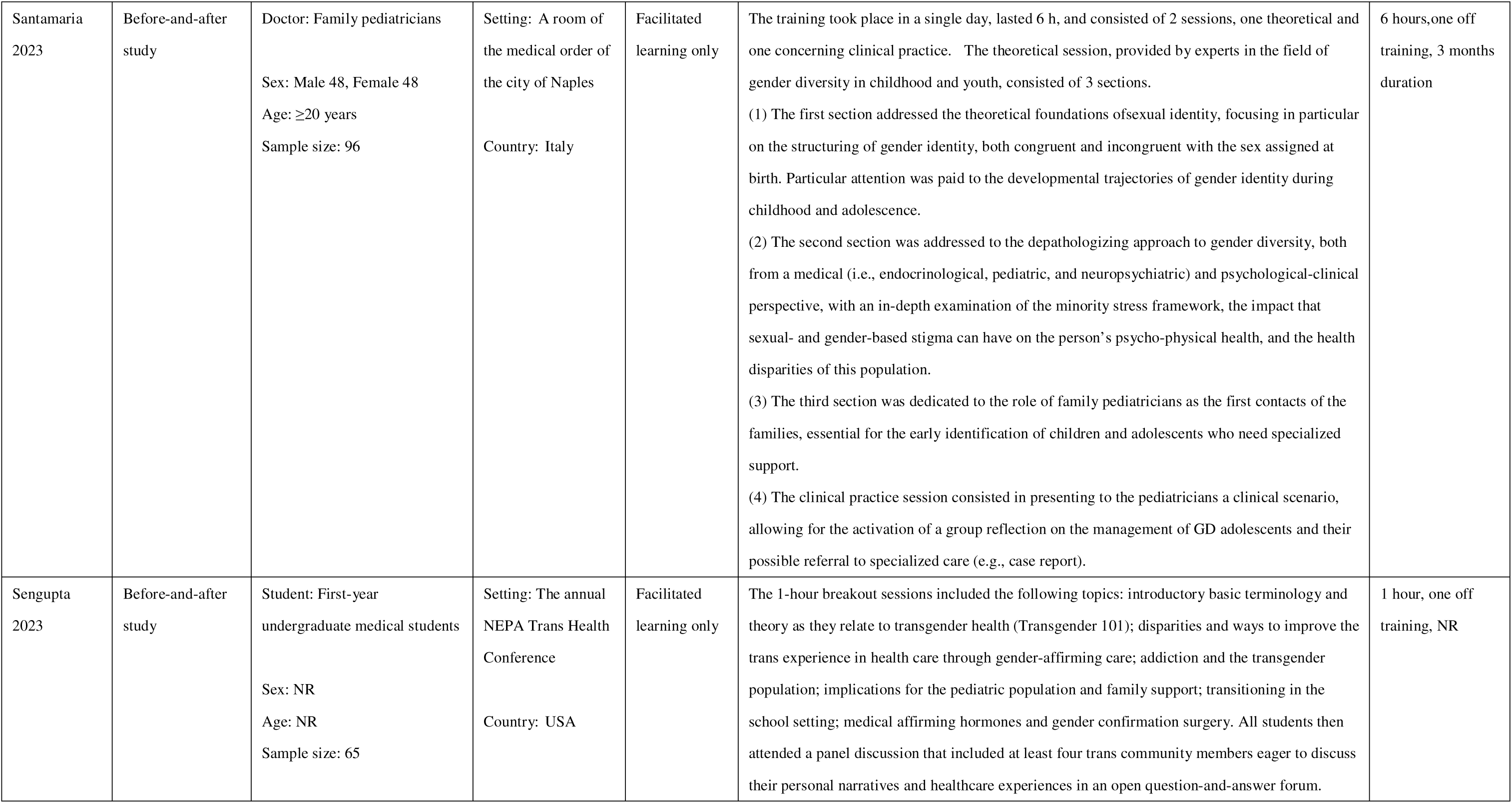
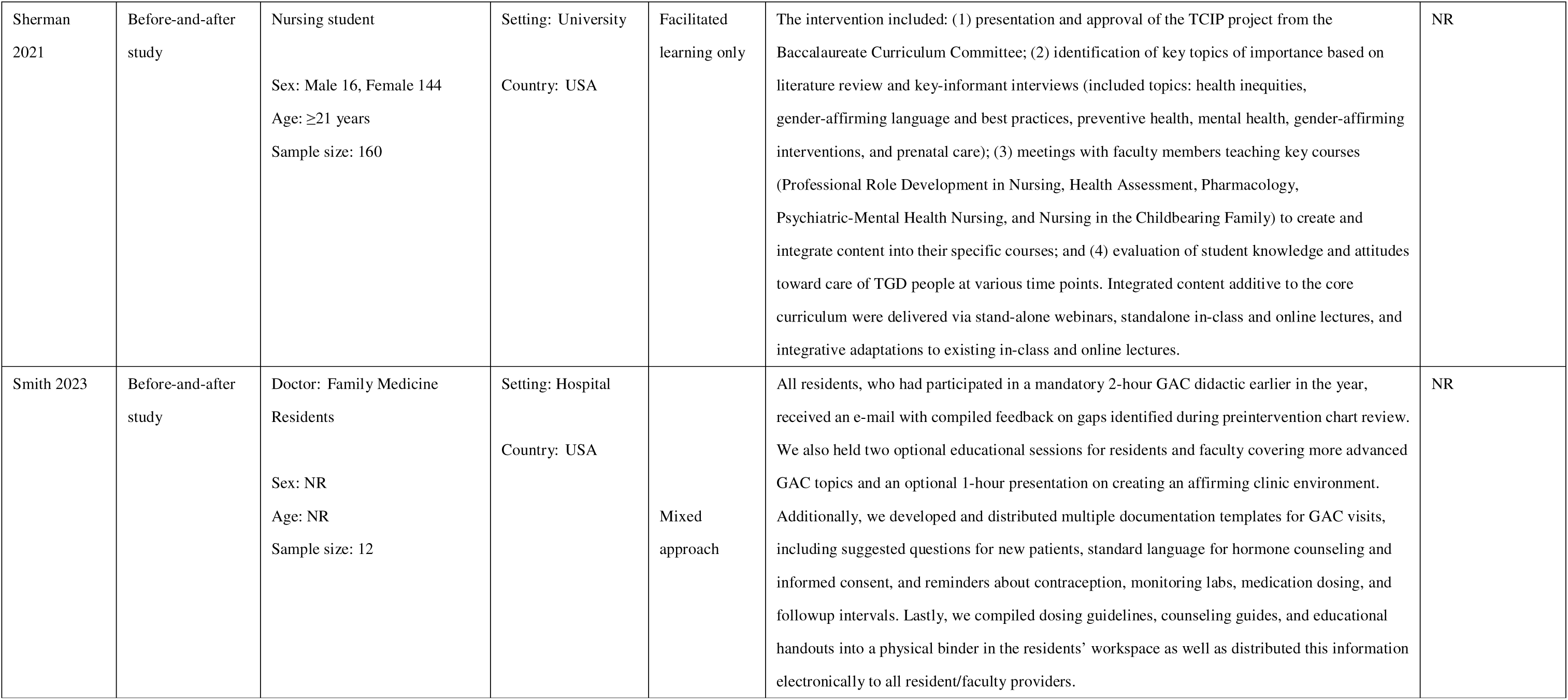
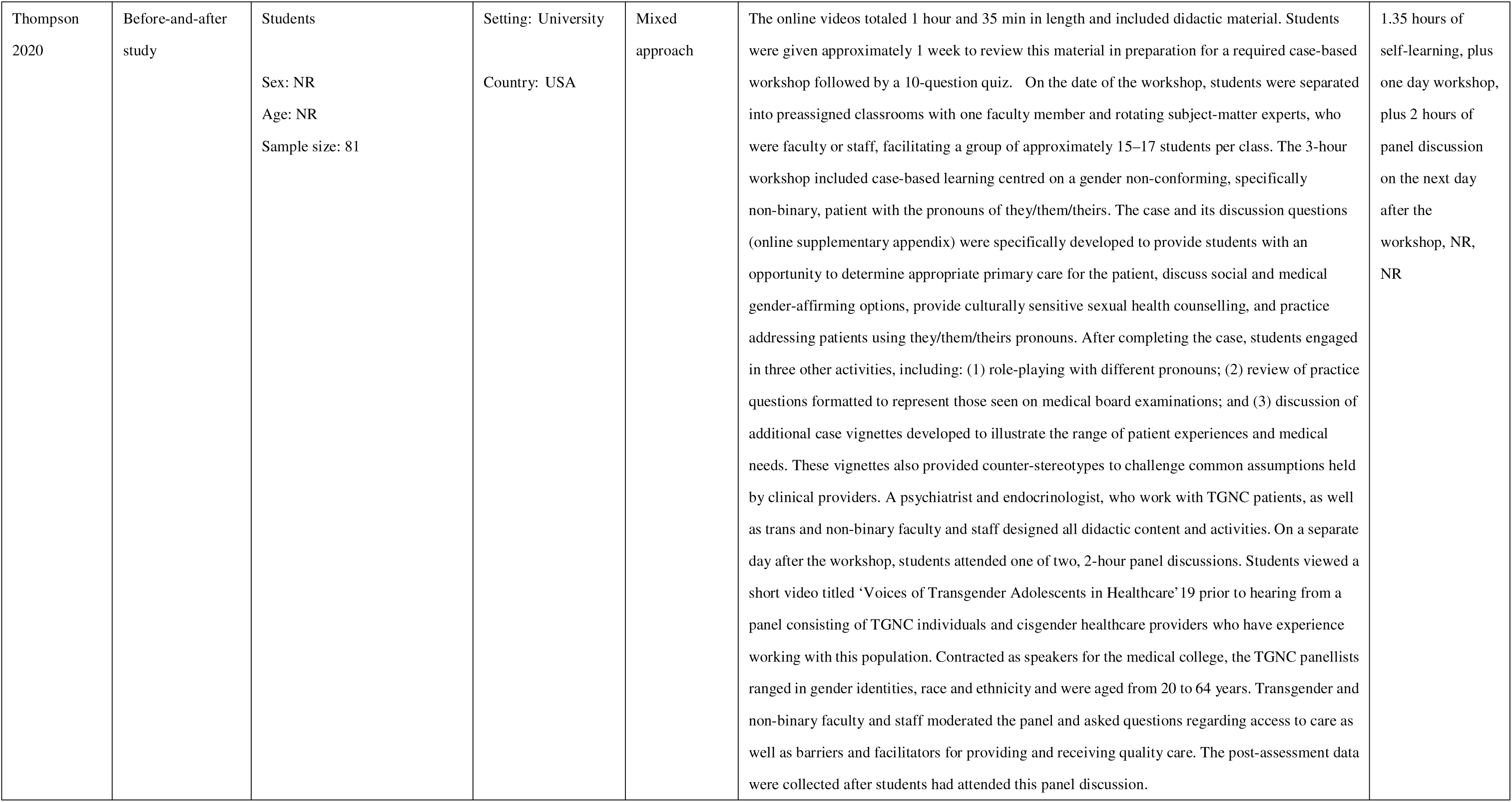
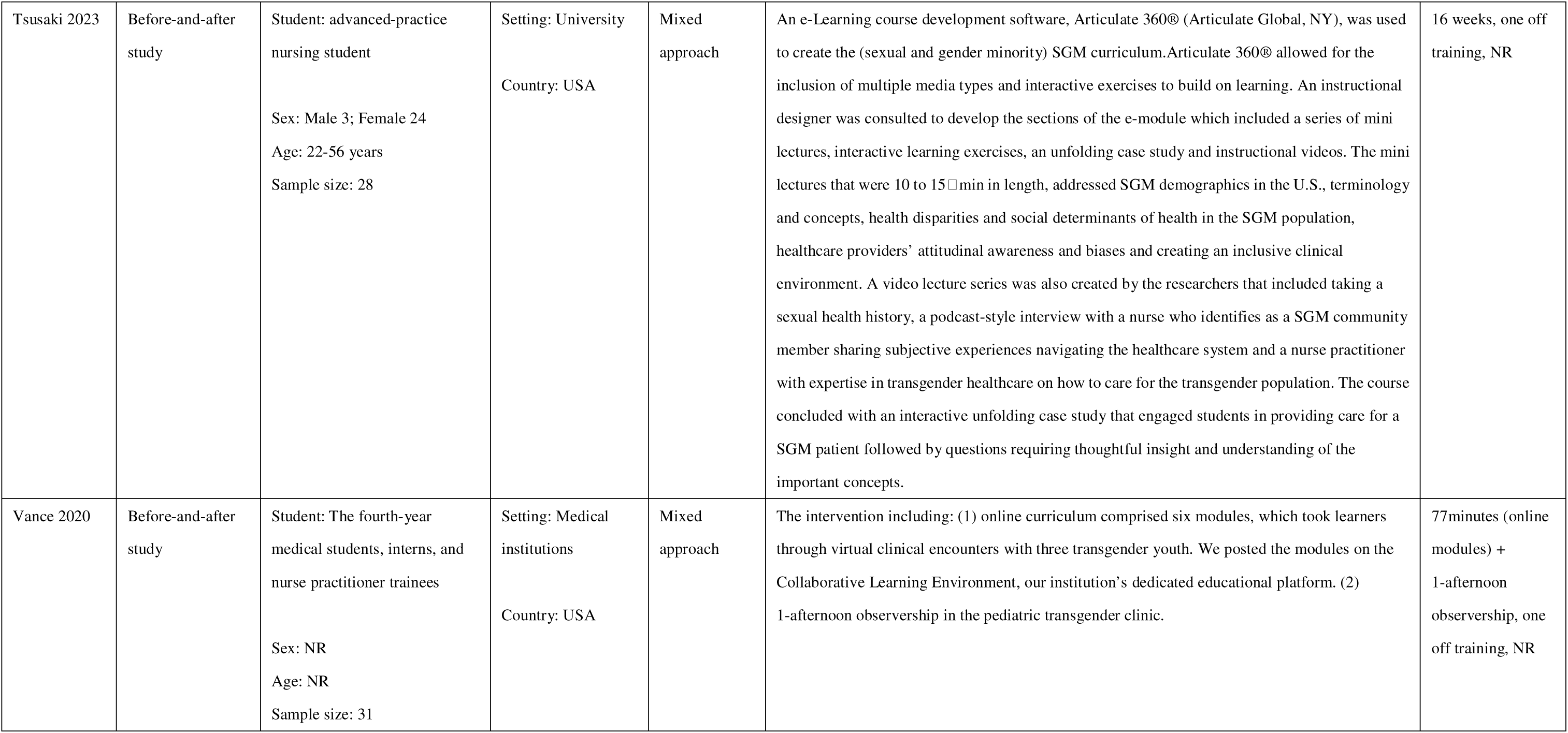
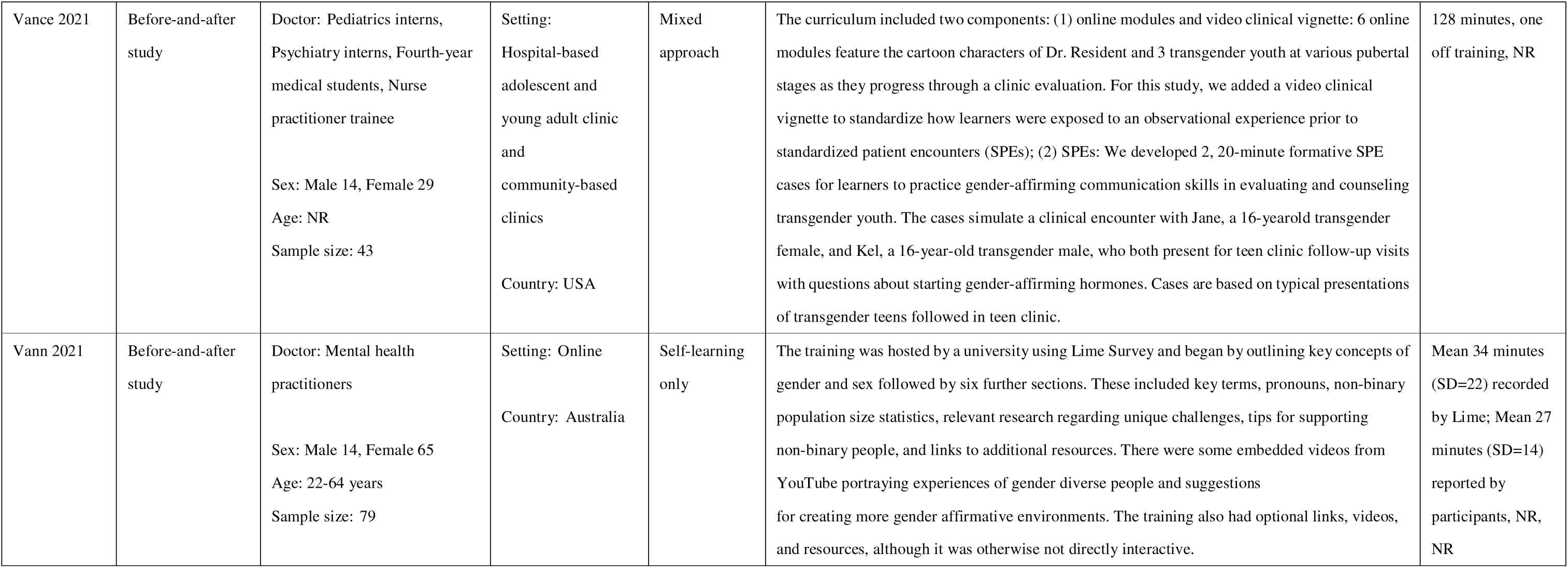
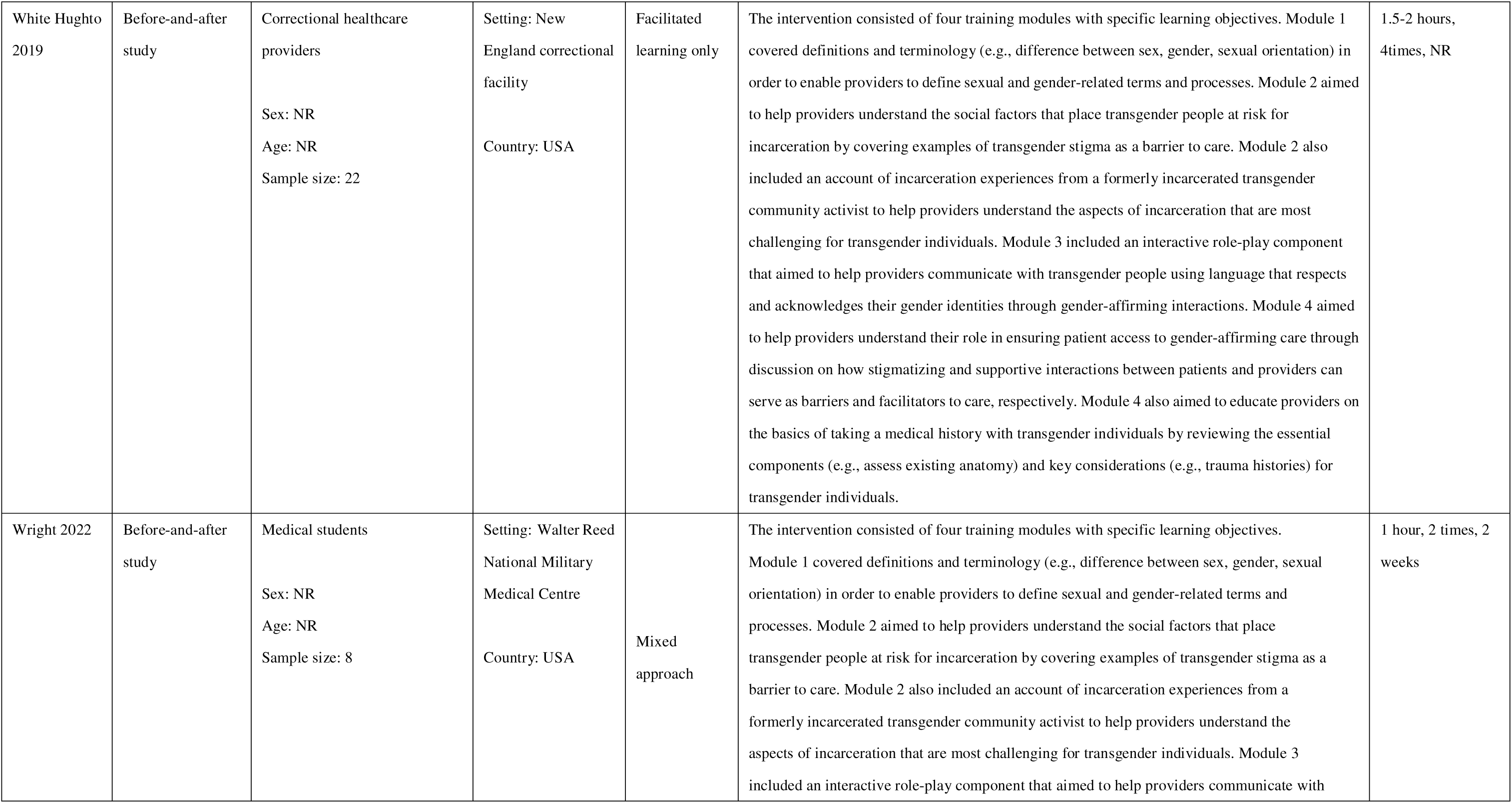
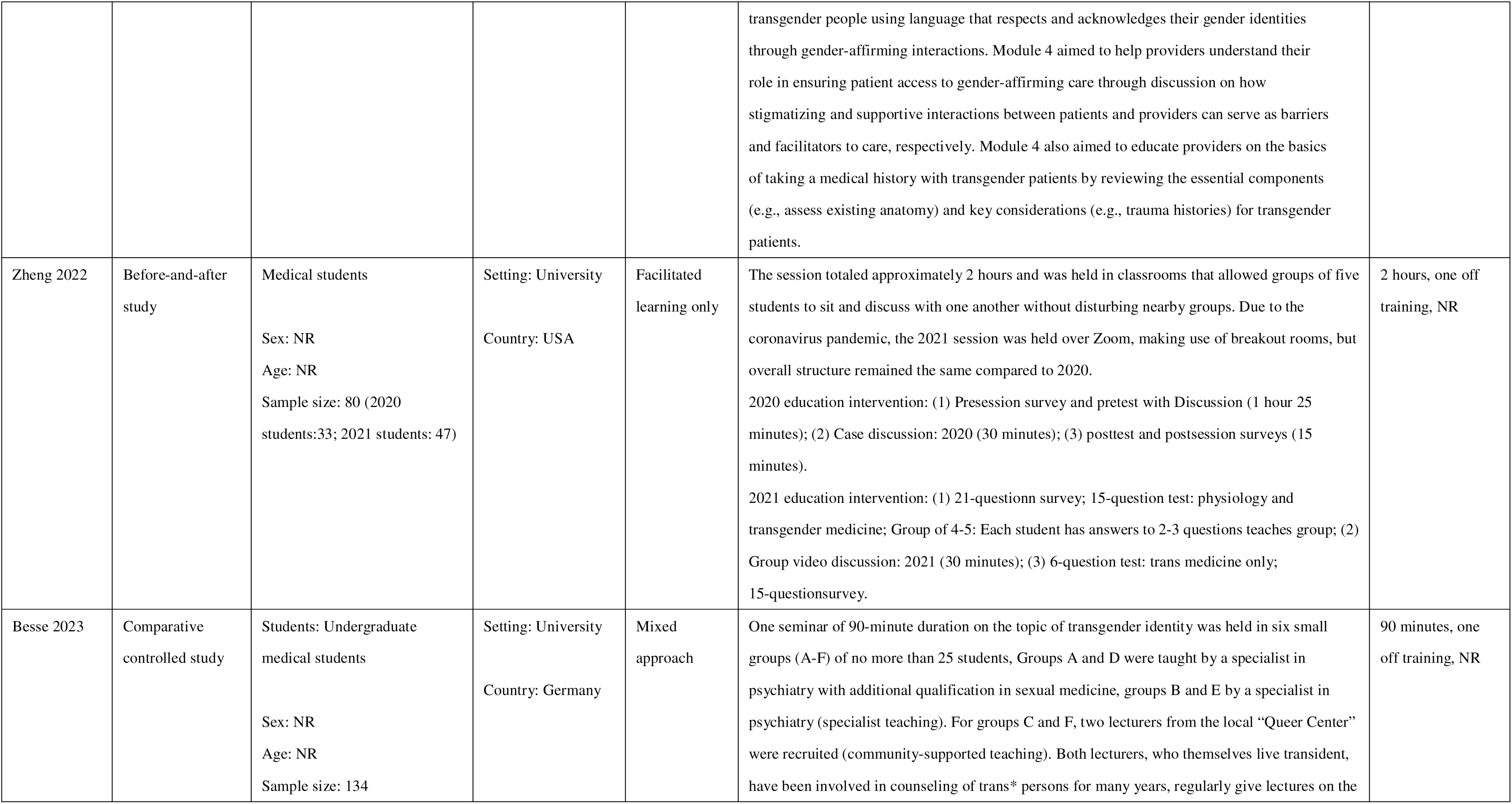
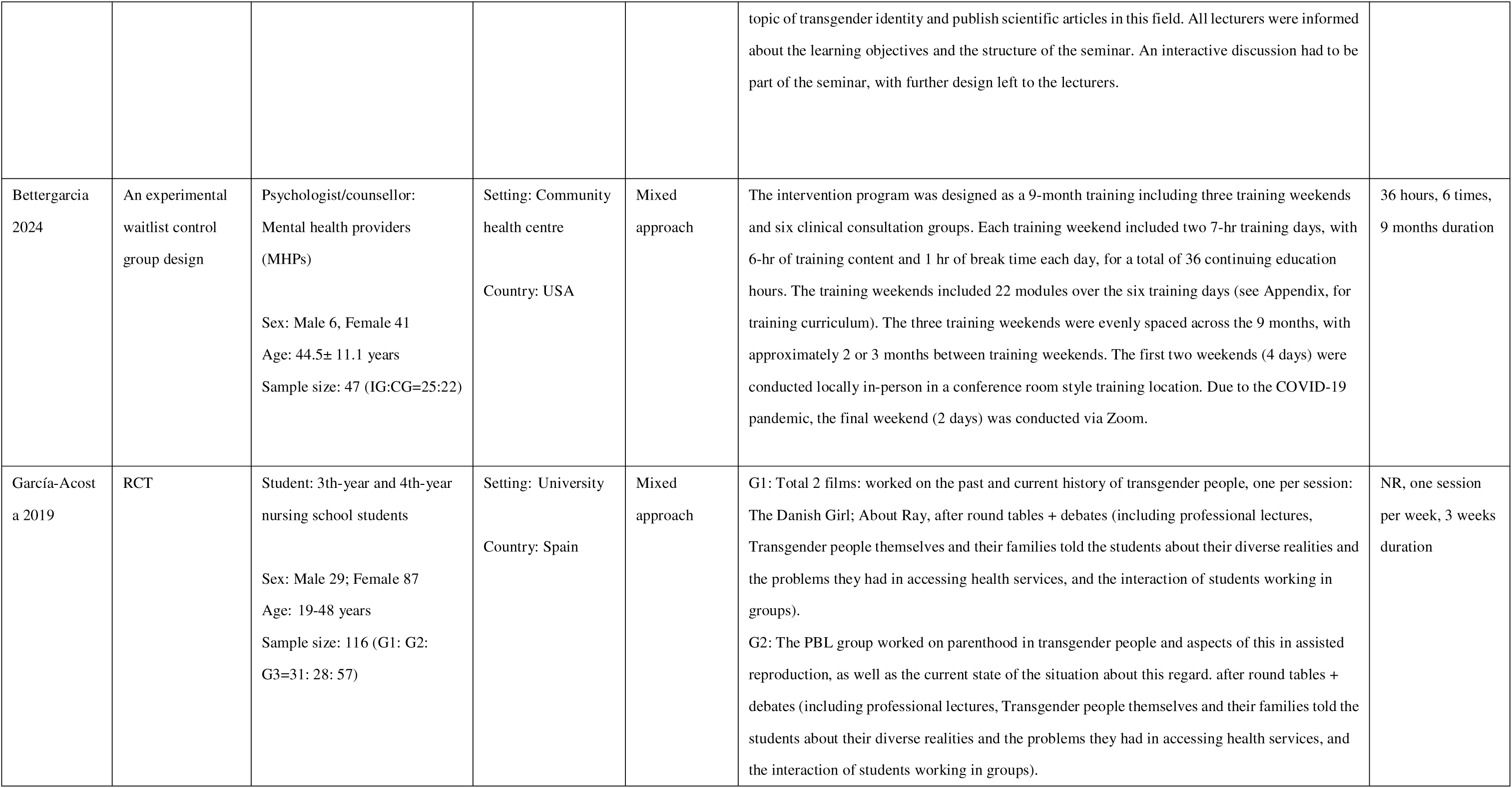
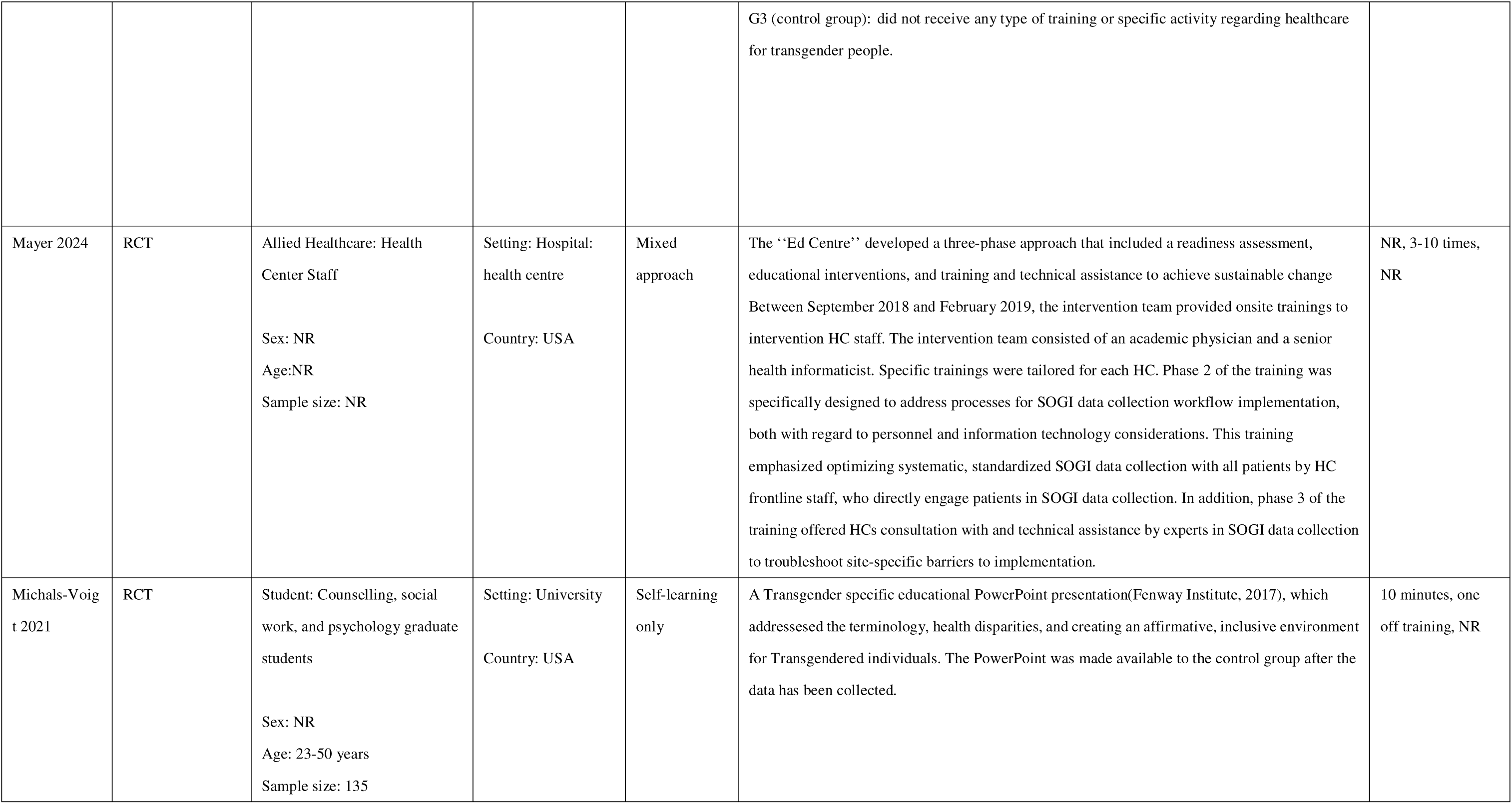
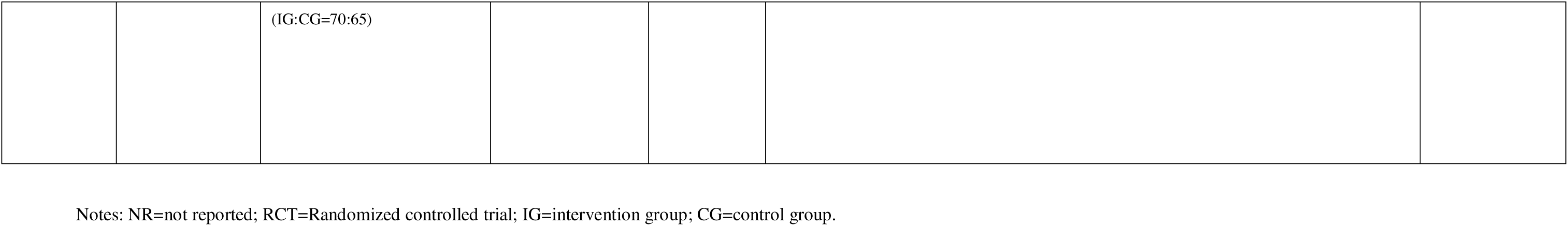
Characteristics of all included quantitative studies (n=70)

| Studies | Study design | Population | Setting/Country | Format of intervention | Description of Intervention | Intervention Time/ Frequency/ Duration |
| --- | --- | --- | --- | --- | --- | --- |
| Allison 2019 | Before-and-after study | Student: Health care professionals/With five colleges (Medicine, Pharmacy, Nursing, Health Professions, and Public Health) and a Graduate School, the university enrolls more than 2700 students.<br>Sex: NR<br>Age: NR<br>Sample size: 58 | Setting: An academic health centre in a southern state<br>Country: USA | Facilitated learning only | The workshop included a slide presentation on basic terminology and concepts, video clips of trans/NB patient–provider interactions, facilitated discussions of affirming practices, and a trans/NB panel. We implemented the full workshop in the summer of 2018 as part of an interprofessional course on patient- and family-centred care offered to students in a variety of disciplines through a partnership with the university’s Office of Interprofessional Education. Faculty from the university’s college of public health and members of the trans/NB community involved in the Transform Health Arkansas Initiative cotaught the gender-affirming care workshop on the first day of the course. | 2 hours, NR, NR |
| Altmiller 2023 | Before-and-after study | Nurse students<br>Sex: NR<br>Age: NR<br>Sample size: 34 | Setting: University<br>Country: USA | Mixed approach | Virtual patient simulation (VPS) uses a web-based platform that allows nursing students to interact with patients in specific contexts that support practicing and applying nursing skills such as collecting subjective and objective data, documenting data in electronic health records, choosing appropriate interventions, and evaluating outcomes of care. | NR, one off training, one week |
| Altschuler 2023 | Before-and-after study | Doctor: Medical residents<br>Sex: Male 18, Female 11, | Setting: University<br>Country: USA | Facilitated learning only | Two models were used in designing this intervention, including action learning and action research (ALAR) design and Kern Model.<br>The intervention content was as follows: patient experiences of accessing care, terminology, | NR, one off training |
|  |  | Unspecified:6<br>Age: NR<br>Sample size: 35 |  |  | patient experiences of discrimination in clinic, best practices for gender inclusivity, cultural humility, taking gender/sexuality history, minority stress/health disparities, health needs, and procedure guidelines. |  |
| Arora 2019 | Before-and-after study | Students, General Practitioners, internal medicine physicians<br>Sex: NR<br>Age: NR<br>Sample size: 163 | Setting: Community health centre<br>Country: Australia | Facilitated learning only | The intervention included the following key topics: appropriate transgender terminology; exploring the biological basis (genetic, gonadal, genital and neurological) of gender identity and diversity; the lived experience of a transgender person and their relationship with healthcare providers; supportive care for children and families; adolescent puberty blockade; adult transition care; fertility options; hormonal monitoring and surgery; with clinical concepts explained for the medical students. | 1 hour, one off training, NR |
| Berenson 2020 | Before-and-after study | Students: Second-year medical students<br>Sex: NR<br>Age: NR<br>Sample size: 123 | Setting: University<br>Country: USA | Facilitated learning only | The intervention contained the following components: (1) a didactic PowerPoint presentation reviewing unique health issues and disparities experienced by transgender patients with a specific focus on teaching students office-based masculinizing and femininizing therapies; (2) a small-group session viewing and analyzing a pair of showcasing competent and poor communication between a provider and a transgender patient; (3) the use of a large-group patient panel featuring members of the transgender community. | 2.5 hours, one off training, NR |
| Bi 2020 | Before-and-after study | Student: The first-year medical students<br>Sex: NR<br>Age: NR<br>Sample size: 83 | Setting: University<br>Country: USA | Facilitated learning only | In this study, teaching module on intersectionality of sexual orientation, gender identity, and race/ethnicity included a 30-minute didactic lecture and a 1.5-hour facilitated discussion portion of the session. Students viewed four video interviews of sexual and gender minority patients of color describing their lives and health care narratives, each followed by a moderated discussion about how compounded minority stress affects lived experiences and health, how multiple identities affect patient-provider communication, and how to improve shared decision making for sexual and gender minority and patients of color. Following the facilitated discussion, one of the video interviewees, who had experience training clinicians in sexual and gender minority issues, participated in a 30-minute question-and-answer session with the students. | 2.5 hours, one off training, NR |
| Bruno 2023 | Before-and-after study | Doctor: primary care providers ( i.e., physicians, nurse practitioners, and physician assistants); behavioural health providers ( i.e., licensed social workers and clinical psychologists); other HC positions (i.e., front desk supervisor, program manager, HIV counsellor, workforce development trainer, and medical scribe)<br>Sex: NR<br>Age: NR<br>Sample size: 464 | Setting: Online<br>Country: USA | Self-learning only | Intervention sessions began with a 30-minute didactic presentation given by a faculty member or guest presenter, followed by two to three participant-led case discussions and knowledge sharing among peers. | 30 minutes, NR,NR |
| Carmichael 2024 | Before-and-after study | Student: Pre-licensure nursing students<br>Sex: Female 38 (82.6%)<br>Age: NR<br>Sample size: 46 | Setting: University<br>Country: USA | Mixed approach | The intervention consisted of a 2-hour synchronous, on-line education session that included didactic content, a video of TGNB individuals sharing their perspectives on health care, a case study, and reflective group discussion | 2 hours, one off training, NR |
| Chakrapani 2023 | Before-and-after study | Healthcare workers<br>Sex: Male 40, Female 58 | Setting: Hospital<br>Country: India | Mixed approach | The intervention contained group-level (a half-day workshop) and individual-level (four videos) components.<br>A single half-day (3 h) workshop was conducted separately for the clinical (doctors/nurses) and | 3 hours workshop and 20 minutes video, NR, NR |
|  |  | Age: 40.5±9.1<br>Sample size: 98 |  |  | non-clinical/paramedical staff (e.g., counsellors, lab technicians, and ward attendants) in each of the two public hospitals, aiming at enhancing the clinical care provided to sexual and gender minority, especially transgender people and same-sex-attracted people.<br>The half-day workshop was followed by an individual-level intervention, in which four short videos, each of about 5 min duration in local languages (Hindi and Tamil), were shared with healthcare workers individually over WhatsApp. |  |
| Chaudhary 2024 | Before-and-after study | Student: Pharmacists and pharmacy students<br>Sex: Male 12, Female 39, Non-binary 3, Prefer not to say 2<br>Age: NR<br>Sample size: 56 | Setting: Online learning<br>Country: Australia | Self-learning only | In this study, an online training program on transgender and gender diverse (TGD) healthcare was provided, including:<br>1. Transgender Healthcare – Language, terminology, and key healthcare issues<br>2. Gender affirming therapies<br>3. Case Studies in TGD Healthcare | NR |
| Chu 2022 | Before-and-after study | Nurse: Nurse staff at an inpatient perinatal unit<br>Sex: Female 23<br>Age: ≥ 21years<br>Sample size: 23 | Setting: Private academic medical centre<br>Country: USA | Facilitated learning only | The content of the training was based on recommendations in the literature and reviewed by content experts. The content of the training focused on the following aspects of affirming and inclusive care: asking for information about sexual orientation and gender identity (SOGI), providing gender-neutral forms, respecting pronouns, not assuming genders, naming pregnancy and body parts, creating welcoming environments, and replacing incorrect information. | 1 hour, one off training, NR |
| Click 2020 | Before-and-after study | Student: First- and second-year medical students<br>Sex: NR | Setting: University<br>Country: USA | Facilitated learning only | In the Integrated Grand Rounds sessions, cases were co-presented to students by clinical and basic science faculty members. Clinical faculty members presented didactics lectures related to basic terminology, electronic health record, interview and hormone therapy. Basic science faculty members presented topics related to sex differentiation, the action of steroid hormones | 3 hours, NR, NR |
|  |  | Age: NR<br>Sample size: 138 |  |  | and hormone therapy. Faculty members chose topics based on their expertise and what they saw as most important for first- and second year students with little exposure to this topic. Small group discussions included uncomfortable situations, planning and practicing a patient interview, creating safe environments, overcoming personal bias, and building the health care team. A transgender individual participated in a live interview and a 'question and answer' session. |  |
| Congdon III 2021 | Before-and-after study | Medical doctor, medical resident, medical student, psychologist, pharmacist , pharmacy resident, pharmacy student, pharmacy technician, registered nurse, licensed practical nurse, social worker, medical support assistant<br>Sex: NR<br>Age: NR<br>Sample size: 100 | Setting: NR<br>Country: USA | Facilitated learning only | In this study,two sessions were delivered to medical residents and students during their noon conference. One session was delivered to the multidisciplinary women's health primary care team. One session was delivered to clinical pharmacists, residents, and students during a pharmacy clinical meeting. The 4 preceding sessions were delivered over 45 minutes with Q&A afterward. The final session was delivered to large group (n=52) of pharmacists and pharmacy technicians for continuing education credit for a duration of 30 minutes with Q&A included. The content for this final session was abbreviated to focus on medication therapy | Over 75 minutes, one off training, 5 months duration |
| Corey 2023 | Before-and-after study | Students: Nursing and physician assistant students<br>Sex: Male 6, Female 25<br>Age: 18-59 years<br>Sample size: 31 | Setting: University<br>Country: USA | Mixed approach | Participants completed the online Transgender Health Learning Series, which contained 7 courses: Transgender Health 101: A Foundation Course on Transgender People and Public Health; Transgender Health 102: Transgender Patient-Provider Relationship; Transgender Health 103: LGBTQ+ Healthcare Workplace Inclusion; Transgender Health 104: Psychological Distress; Transgender Health 105: Role of Endocrinology in the Transgender Community; Transgender Health 106: Pre and Post-operative Care; Transgender Health 107: Transgender Health 107: Not About Us, Without Us: The Importance of Transgender Patient-centred Outcomes. | 7 hours,7times, NR |
| Craig 2021 | Before-and-after | Doctor: Clinicians | Setting: | Facilitated | Informed by the NIH Behavior Change Consortium' s best practices for facilitator training, the | 14 hours, 8 times, |
|  | study | Sex: 64 participants' information didn't report.<br>Among the 65 participants, 44 females.<br>Age: 22-68 years<br>Sample size: 129 | Community health centre<br>Country: USA | learning only | AFFIRM training is a standardized two-day, 14 h training workshop that is informed by experiential learning theory and integrates didactic and experiential learning (e.g., role play, problem-based learning).<br>The training consists of: (1) an overview of the development of AFFIRM;(2) a review of the evidence for utilizing groups with SGMY; (3) an orientation to the group modules and objectives; (4) strategies to integrate LGBTQ affirmative approaches into groups; (5) a comparison between AFFIRM CBT-based groups and LGBTQ support groups; (6) an in-depth theoretical and conceptual orientation to CBT and affirmative practice; and (7) experiential learning and practice activities associated with implementing each intervention module that deepen skills acquisition. | from April 2017 to April 2019. |
| Dale 2022 | Before-and-after study | Student: Medical students ranged from their first to final year<br>Sex: NR<br>Age: NR<br>Sample size: 22 | Setting: University<br>Country: UK | Facilitated learning only | In this study, three 20 min lectures were given, with time for questions. The presenters were a senior lecturer and researcher looking at inclusive care for older trans people, a senior medical doctor specializing in the provision of transgender health care, and a transgender activist. Topics discussed included the spectrum of gender identity, using pronouns, and a broad overview of experiences of transgender people across the age spectrum and life course. | 2 hours, one off training, NR |
| Divya R 2023 | Before-and-after study | Doctor: Medical professionals<br>Sex: NR<br>Age: 20-above 60 years<br>Sample size: 58 | Setting: Community health centre<br>Country: India | Self-learning only | An online medical conference on Transgender medicine was conducted by the investigators with subject experts. 58 out of the 126 volunteers attended the conference. The conference titled transgender physiology included topics such as Introduction to Transgender Health, History, Terms & Concepts, Transgender Anatomy, Genetics, Reproductive Physiology, Transgender Brain, Physiology behind gender incongruence, Research in Transgender Medicine, Sexuality, Mental health issues, public health issues, Reproductive health and concerns of Transgender Individuals, managing a Transgender Patient, Panel Discussion on 'Teaching and Learning | NR,one off training, NR |
|  |  |  |  |  | Transgender Medicine’. |  |
| Fitze 2022 | Before-and-after study | Nurse: The Surgical Oncology Unit nurses<br>Sex: NR<br>Age: NR<br>Sample size: 30 | Setting: Hospital<br>Country: USA | Self-learning only | The project implemented aimed to combine both simulation and deliberate practice by using an electronic education module combined with physical anatomical models of postsurgical vaginoplasty and both stage one and stage two phalloplasty patients that staff could use to examine and practice postoperative care. | Electronic educational: 15-20 minutes and physical anatomical models, NR, 8 weeks |
| Frazier 2021 | Before-and-after study | Student: Pharmacy doctor students<br>Sex: NR<br>Age: NR<br>Sample size: 73 | Setting: University<br>Country: USA | Mixed approach | The intervention included the pre-class knowledge-based activities and in-class application activities. The pre-class active-learning materials consisted of a six-page document with frequently asked questions about transgender individuals and a 40-min prerecorded lecture describing terminology, health disparities, and gender-affirming treatment guidelines. The three-hour class time was split between a two-hour interactive session and a one-hour discussion between the students and a transgender person. The two-hour interactive education session included a game show style activity reviewing and enhancing information from the pre-class elements, a student gender identity exploration exercise, discussion of a current events article about a transgender individual not receiving optimal care from a pharmacist, and patient/provider role-play. The last hour was devoted to a facilitated discussion between the class and a transgender man. | NR |
| García-Acosta 2024 | Before-and-after study | Student: Fourth-year undergraduate nursing students<br>Sex: Female 83.40%<br>Age: 20-25 years (female)<br>Sample size: 56 | Setting: University<br>Country: Spain | Mixed approach | The study was carried out in two phases.<br>In the first phase, virtual resources were produced by simulating three different potential scenarios that students could face in their future professional careers: a mental health consultation, a paediatrics consultation, and a primary care consultation. In these scenarios, the user or patient was invariably a trans person. Two versions of each scenario were recorded: one exclusionary and one inclusive. | 30 minutes, 3 times, NR |
|  |  |  |  |  | In the second phase, the educational intervention was implemented using the tools created. The intervention was carried out in two sessions (Figure 1). In the first session, a pre-test on knowledge was administered and then a short training session was delivered in the form of a lecture. Two days later, in the second session, the virtual tools were pilot tested: two in an SV format (the adult and mental health consultations) and another in a VR format (the paediatrics consultation). |  |
| Gorrotxateg 2020 | Before-and-after study | Students: Second-year Social Education degree students<br>Sex: Male 11, Female 52, Another gender 1<br>Age: 18-29 years<br>Sample size: 64 | Setting: University<br>Country: Spain | Facilitated learning only | An interactive training program using the Creative Factory methodology relating to the topic of transsexual persons. | NR, a weekly session, 4 months duration |
| Guss 2022 | Before-and-after study | Others: Doctor and students<br>Sex: Male 22, Female 46<br>Age: NR<br>Sample size: 68 | Setting: Hospital<br>Country: USA | Self-learning only | In this study, an 8-minute educational video was developed to teach essential topics in transgender and nonbinary health. | 8 minutes, one off training, NA |
| Hart 2021 | Before-and-after study<br>fore-and-after study | Student: Physician Assistant<br>Students<br>Sex: Male 18, Female 80<br>Age: >18 years<br>Sample size: 98 | Setting: NR<br>Country: USA | Facilitated learning only | This was an optional 2-hour lecture. The same lecture was offered 3 times (one per cohort)—all taught by the same 2 faculty experts on transgender health care. Learning objectives covered terminology, unique barriers to health, legal topics, community resources, transgender-specific history and physical exam skills, and published guidelines for medical and surgical management of gender dysphoria | 2 hours, one off training, NR |
| Hersh 2022 | Before-and-after study | Doctor: Family medicine physicians | Setting: Hospital | Facilitated learning only | Prior to the day of the case, we gave the standardized patients the case scenario and invited them to a 1-hour presession training with an SP educator and the case authors. The training included a | NR, NR, 4 years duration |
|  |  | Sex: NR<br>Age: NR<br>Sample size: 20 | Country: USA |  | full read of the script, addressed questions and concerns about the materials, discussed the focus for feedback on communication skills, and allowed SPs to choose which hormones they planned to ask for during the scenario. We held the SP case in the simulation center of an academic health institution and utilized trained SPs. |  |
| Hoffner-Hein<br>ike 2024 | Before-and-after<br>study | Clinical staff/health care 78%;<br>Behavioral health 9%; Health<br>outreach/education 4%;<br>Administrative staff 3%;<br>Public/environmental health<br>3%; Student 1%; Something<br>else 3%primary care providers in<br>North America<br>Sex: NR<br>Age: NR<br>Sample size: 71 | Setting: NR<br>Country: USA | Facilitated<br>learning only | The educational sessions included (1) the role of mental health care in gender diverse patients, (2) medical management of feminizing hormones, (3) medical management of masculinizing hormones, and (4) introduction to gender affirming surgery. These topics were considered foundational by the hub team. Each hour-long session included a 20-min didactic followed by interactive discussion and case-based presentations. | Once per week for<br>one hour, 4 weeks<br>duration |
| Huser 2022 | Before-and-after<br>study | Psychologist/counsellor:<br>Board-eligible or certified<br>genetic counselling and student<br>Sex: Male 1, Female 39<br>Age: 18-64 years<br>Sample size: 40 | Setting: Community<br>health centre<br>Country: USA | Self-learning<br>only | 1. A 5-module genetic counselling-targeted online learning program<br>(1) Terminology, Population Disparities, and Communication: A comprehensive glossary, breakdown of transgender and/or gender non-binary (TGNB) population disparities, as well as gender-affirming communication considerations for the genetic counsellor.<br>(2) Clinical Environment: An examination of the clinical environment from the patient perspective, from start to finish, including things such as in-take forms, the physical environment, and documentation.<br>(3) Potential Aspects of the Gender Affirmation Process: A summary of potential psychological, social, hormonal, surgical, and legal affirmations individuals make undergo in their gender<br>affirmation process.<br>(4) Cancer Risk Assessment: An organ-based perspective summarizing what is known up to this point in the literature on cancer risks of TGNB individuals and how to consider possible hormonal or surgical affirmations into risk estimates and management.<br>(5) Pulling it All Together- The Genetic Counseling Session: Applying everything in Modules 1–4 to core components of the genetic counseling session such as family and medical history gathering, education, psychosocial considerations, and patient resources.<br>2. Participants were provided the option of joining a private online discussion forum hosted on Slack, a business communication platform. | 146 minutes ± 94<br>minutes, one off<br>training, NA |
| Jadwin-Cakmak 2020 | Before-and-after study | Doctor: Medical doctor, physician's assistant; Nurse practitioner, nurse; Medical assistant, certified nursing assistant; Social worker; Psychologist, counsellor; Administrator, medical office clerk; Health educator, HIV test counsellor; Community health worker/peer support/outreach; CEO/executive director.<br>Sex: Male 23, Female 126, Transgender/genderqueer/gender nonconforming; 2 | Setting: Online and site training<br>Country: USA | Facilitated learning only | The Health Access Initiative (HAI) consists of a 1-hour online training and 2-hour in-person training, followed by 3 months of site-specific TA to improve clinic- and structural-level issues related to LGBTQ+ care. The online training (Access the Health Access Initiative online training free of charge at mfierce.org.) focuses on LGBTQ+ terminology and cultural humility, featuring testimonials from YAB members and health care providers. The in-person training includes didactic learning methods and group activities on challenging biases and stereotypes, considerations for providing patient-centered care, cultural humility practices, taking a comprehensive and affirming sexual health history, patient-centered STI prevention counseling, and best practices for referrals and resource provision with SGMY. A medical doctor and health educator—both with extensive experience working with SGMY—cofacilitated the in-person trainings. Many but not all trainings (80%) also included a YAB member as a cofacilitator. The TA was unstructured and tailored to each site. HAI staff met with a site liaison before and after the training to identify and address the site's most pressing needs related to serving SGMY, such as resources, referrals, and reviews of clinic forms and policies.<br>Following the training, program staff and YAB members provided TA to all 10 sites for | 3 hours+3 months, one off training, NR |
|  |  | Age: ≥20 years<br>Sample size: 151 |  |  | approximately 3 months. |  |
| Kaur 2023 | Before-and-after study | Student: Bachelor of Medicine and Bachelor of Surgery (MBBS) final year students<br>Sex: Male 85, Female 84<br>Age: 22-25 years<br>Sample size: 165 | Setting: University<br>Country: India | Facilitated learning only | The educational intervention comprised a 90-min lecture on transgenderism and the need to understand it as a natural and hormonal phenomenon beyond cultural and social stereotypes. The physiological and psychological aspects of transgenderism and the need for its societal acceptance were emphasized. At the end of the session, another 30 minutes were allotted to address the individual queries by the subject experts | 90 minutes, one off training, NR |
| Kelly 2023 | Before-and-after study | Nurse: Nurses who work solely in the inpatient psychiatric setting<br>Sex: Male 1, Female 2<br>Age: 31-59 years<br>Sample size: 3 | Setting: Online<br>Country: USA | Self-learning only | As described above, this project utilized the Johns Hopkins Evidence-Based Practice Model (JHN-EBP) in order to carry out the intended purpose. This project incorporated the three-step process of the JHN-EBP, which consists of identifying a practice question, gathering evidence, and translation into nursing practice, also referred to as PET (Practice, Evidence, Translation) | 30 minutes, NR, NR |
| Knockel 2019 | Before-and-after study | Student: Student pharmacists<br>Sex: NR<br>Age: NR<br>Sample size: 101 | Setting: University<br>Country: USA | Mixed approach | An aligned and complementary lab-based LGBTQ-related curriculum with specific focus on transgender health was delivered with the following learning objectives: (1) use appropriate language in caring for individuals in the LGBTQ community, (2) explain ways to ensure a welcoming environment for individuals in the LGBTQ community, (3) list hormone therapy options for transmen and transwomen, (4) counsel patients on physical changes induced by hormone therapy, and (5) counsel patients on appropriate administration, adverse effects, and monitoring of masculinizing and feminizing therapy. | NR, one off training, NR |
| Kreines 2022 | Before-and-after study | Doctor: Obstetrics and gynecology residents | Setting: University | Facilitated learning only | The simulation curriculum consisted of a 45-min recorded lecture, a 15-min observed simulated encounter with a gender diverse standardized patient (SP), 5 minutes of individualized feedback | 2 hours + 5 minutes, one off |
|  |  | Sex: Male 4, Female 26<br>Age: 26-36<br>Sample size: 30 | Country: USA |  | from the SP and observing faculty member, and a 1 hour group debrief with all learners, SPs, and faculty members. | training, NR |
| Lacombe Duncan 2021 | Before-and-after study | Allied Healthcare: healthcare and social service providers (e.g., physicians, nurses, social workers) and providers in-training working with trans women living with and/or affected by HIV<br>Sex: NR<br>Age: 37±11years<br>Sample size: 51 | Setting: Multiple organizations<br>Country: Canada | Facilitated learning only | TEACHH was underpinned by the Information-Motivation-Behavioural Skills Model (IMB) and delivered by trans women in a 3-h session :<br>During this training, we plan to have providers: (1) discuss human rights for trans women and consider different types of discrimination trans women may face such as transphobia, racism, sexism and HIV stigma (increasing knowledge and reducing attitudes/biases which may increase motivation to provide affirming care to trans women including those living with HIV); (2) teach providers about affirming words with which to discuss gender identity and expression (increasing knowledge and providing interpersonal strategies to be affirming) and develop a basic understanding of trans healthcare, HIV prevention (e.g., pre-exposure prophylaxis) and HIV treatment (e.g., ART), and how these types of healthcare affect trans women living with and affected by HIV (increasing knowledge); (3) discuss what it means to be trans-affirming in their work and how they can make their organisations more trans-affirming (increasing knowledge and providing organisational strategies to promote affirming care to trans women including those living with HIV) and (4) have participants complete a case study to apply what they have learnt to practice (application of knowledge, interpersonal strategies and organisational strategies). | 3 hours, one off training, NR |
| Lee 2021 | Before-and-after study | Student: Second-grade medical students in South Korea<br>Sex: Male 29, Female 20<br>Age: NR<br>Sample size: 49 | Setting: University<br>Country: USA | Facilitated learning only | In this study, the lecture included discussions on the definition and core concepts of gender, gender dysphoria, transgenderism and related epidemiology and biology, psychiatric and social issues, health disparities, general primary care, hormone replacement therapy, and surgical options. An email and text message were sent 3 and 4 weeks after the lecture to ask them to complete participation. Responses to the postintervention survey, which is identical to the preintervention survey, were returned 4 weeks after the lecture. | NR |
| Linsenmeyer<br>2023 | Before-and-after<br>study | Student: Medical students, family therapy students, speech, language and hearing sciences students, and dietetic interns<br>Sex: NR<br>Age: 21-26 years<br>Sample size: 84 | Setting: University<br>Country: USA | Facilitated<br>learning only | The ITHED was a 1-day, in-person program comprised of lectures, a guided reflection on gender identity, a panel discussion with members of the transgender community sharing their experiences in healthcare, and interactive breakout sessions.<br>Appendix: Gender-Arming Language and Communication: A Skills Workshop | 1 day, one off training, NR |
| Luctkar-Fludde<br>2022 | Before-and-after<br>study | Student: Undergraduate students (88%), 8% were graduate students and 4% were instructors/faculty<br>Sex: Male:7%, Female 91%<br>Non-binary 3%<br>Age: 22.8±6.5 years<br>Sample size: 66 | Setting: University<br>Country: Canada | Mixed<br>approach | The study intervention consisted of (a) online pre-simulation preparation materials; (b) 4 online VSGs; and (c) debriefing; delivered over the course of one week. Pre-simulation preparation materials consisted of 2 voice-over-PowerPoint presentations on the topics of sexual orientation/gender identity terminology and cultural humility, developed and delivered by a team member with LGBTQ lived experience. | NR |
| Mackler 2021 | Before-and-after<br>study | Nurse: Sexual assault nurse examiners (SANE)<br>Sex: Male 0, Female 21<br>Age: >20 years<br>Sample size: 21 | Setting: Community health centre<br>Country: USA | Mixed<br>approach | The project was comprised of four components:<br>1. Creating a continuing education training related to providing gender-affirming and TGNB-specific care for sexual assault survivors.<br>2. Implementing the training as a standalone, virtual course.<br>3. Evaluating self-perceived competency pre- and post-training.<br>4. Evaluating the organizational environment and preparing a summary for key<br>stakeholders with specific recommendations.<br>The TGNB-specific content (Appendix A) for the training was partly informed by the IAFN curriculum guidelines (IAFN, 2018), DOJ's national sexual assault protocol (DOJ, Office of Violence Against Women, 2013), FORGE, a national transgender anti-violence organization (FORGE, 2019), and formative work in the field. The content included information on assessing and providing medical care to TGNB survivors, as well as unique forensic examination and referral and discharge considerations. This course was recorded via a live zoom session and was later made available to individuals who could not attend the live session. | NR |
| McCave 2019 | Before-and-after study | Student: Occupational therapy 164, Physician assistant 108, Medicine 83, Physical therapy 65, Social work, 38, Nursing 36<br>Sex: NR<br>Age: NR<br>Sample size: 278 | Setting: University<br>Country: USA | Facilitated learning only | Stonewall Speakers: Stonewall Speakers provides educational outreach opportunities promoting awareness and understanding for lesbian, gay, bisexual, and transgender (LGBT) people; The speaking engagement consisted of two speakers who shared their personal stories as health care consumers, as well as a question-and-answer opportunity for student learners and faculty.<br>Simulation: watching the 10-minute ED video simulation. and then the facilitators led a 15-minute discussion of how the interprofessional core competencies and standards for affirmative practice were upheld and violated by the behaviors of the providers seen in the video. Students then completed a 10-minute team huddle to prepare for the initial discharge planning meeting with Peter. The team discussed what information was still needed, which providers might be needed for discharge planning, ideas for potential referrals for aftercare needs, and important aspects related to transgender affirmative practice. The facilitators offered ideas to the student team if necessary and led a 15-minute discussion on their teamwork during the huddle. The standardized patient (Peter) was then brought into each room; the student team conducted a 25-minute discharge planning meeting with Peter. The team was tasked with focusing on interprofessional practice behaviors and affirmative practice while putting Peter at the center of the team. Following this, the standardized patients gave 5 minutes of positive and constructive feedback to the student teams on their patient experience as Peter. Facilitators then led a 10-minute team debrief on the discharge<br>planning experience as it related to interprofessional practice behaviors and affirmative practice. Large-group debriefing and assessment: Following the conclusion of the simulation, the students participated in a 20-minute large-group debrief. | 2.5 hours, one off training, NR |
| McMillian-B<br>ohler 2022 | Before-and-after<br>study | Nursing students<br>Sex: NR<br>Age: 20-30 years<br>Sample size: 39 | Setting: University<br>(Online Zoom<br>course)<br>Country: USA | Facilitated<br>learning only | The class and seminar were held virtually via Zoom because of coronavirus disease 2019 (COVID-19) restrictions. The length of the class session was 2 hours; however, the seminar was held during the last 90 minutes of the class. Before the presentation, students and faculty reviewed the expectations for the discussion and learning objectives. The speaker then shared details of the pre-conception, antepartum, intrapartum, and postpartum care he received. Students were invited to ask additional questions or share their thoughts via the chat feature in Zoom. | 2 hours,one off<br>training, NR |
| Mont 2020 | Before-and-after<br>study | Nurse: Forensic nurses,<br>registered with the College of<br>Nurses of Ontario and working<br>within Ontario's SA/DVTCs<br>Sex: Male 0, Female 47<br>Age: > 19 years<br>Sample size: 47 | Setting: NR<br>Country: Canada | Mixed<br>approach | The training module consisted of seven lessons within two overarching sections. The first section, entitled "Introduction to the Issues", provided background and context related to the experiences of trans survivors of sexual assault and was comprised of three lessons:Key Terms, Experiences of Sexual Assault, and Interactions with Healthcare. The second section, entitled "Core Elements", comprised the four core content domains of the curriculum (Initial Assessment, Medical Care, Forensic Examination, and Discharge and Referral) and highlighted principles of trans-affirming care through all aspects of the SANE consultation. Each of these lessons included a corresponding set of learning objectives derived from the 31 competencies. | 1.75 hours, NR, 30<br>hours duration |
| Mont 2021 | Before-and-after<br>study | Nurse: Forensic Nurses<br>Sex: Male 0, Female 65<br>Age: > 19 years<br>Sample size: 65 | Setting: NR<br>Country: Canada | Self-learning<br>only | The curriculum was uploaded in the learning management system, Docebo, in August 2019. It was made immediately available to SA/DVTC nurses who had already completed online Sexual Assault Nurse Examiner (SANE) training—a standardized training of 16 modules focused on, among other topics, strangulation,medico-legal documentation, and interpretation of injuries. These nurses were notified about the curriculum through a mailing list and staff-only website.22 Each nurse was given 6 weeks to complete the curriculum, with a 2-week extension offered to those who required additional time. Data were collected for the evaluation for ~ 10 months, during which time the curriculum was made available to anynewnurses who joined the network<br>and met the requirements. Immediately upon registration for the training, participants were invited to complete a pre-training questionnaire in the learning management system. Consent to participate in the evaluation was obtained as part of the preamble to the questionnaire.<br>Immediately after completion of the curriculum, they were invited to complete a post-training questionnaire in the learning management system, after which they received an automatically generated certificate of completion. | 2 hours, NR, 6<br>weeks to complete<br>the curriculum,<br>with a 2-week<br>extension offered<br>to those who<br>required additional<br>time |
| Muckler 2019 | Before-and-after study | Student: nurse anesthesia students<br>Sex: NR<br>Age: NR<br>Sample size: 28 | Setting: University<br>Country: USA | Facilitated learning only | 1.Simulation Development:The pilot simulation was based on a previous patient encounter by the lead author. After creation, the pilot scenario was reviewed by a second CRNA for anesthesia content and a trans health content expert. In addition to the simulation, trans health resources were identified and later distributed to simulation participants.<br>2.Simulation Scenario:Four group sessions with six students in one session and eight students in three sessions were completed in a designated anesthesia preoperative holding area (APHA) of the simulation center, which included fully functional operating rooms, anesthesia equipment, and high-fidelity manikins. Students were informed during the prebrief that they would approach the interview in pairs, and each student was instructed to conduct half of the anesthesia preoperative interview. After the pre-brief, the emergency department notified the CRNA that the patient was en route to the APHA with a surgical emergency.<br>3.Patient Profile:The role of a patient was played by an SRNA who was provided with scripted lines that were used for each simulation session. | NR, one off training, NR |
| Mueller 2024 | Before-and-after study | Student: Nurse practitioner students (13), Nurse practitioner faculty members (2)<br>Sex: Male 3, Female 12<br>Age: 21-60 years | Setting: University<br>Country: USA | Mixed approach | A prerecorded educational slide presentation made available asynchronously, which included content on the primary care of patients who are TGD. Topics included gender identity, sexual orientation, sexual practices/behaviors, health issues and disparities experienced by TGD patients, health history-taking skills, introduction to masculinizing and feminizing therapies, primary care and health maintenance, and community resources.<br>Three evidence-based readings were chosen from the NONPF Patient-Centered Transgender Care: | NR |
|  |  | Sample size: 15 |  |  | A Toolkit for Nurse Practitioner Faculty and Clinicians document.<br>Participants reported to an in-person class the next week for an interactive patient panel and SP scenario. The patient panel comprised two panelists who were transgender and/or nonbinary. The PI moderated the discussion as panelists discussed their experiences as members of the TGD community, their interactions with the health care environment, and their recommendations regarding best practices for health care providers toward members of the TGD community. An open question-and-answer session was offered at the end.<br>After the panel presentation, participants participated in the SP encounter. In the weeks prior, participants were informed of the nature of the SP experience but were not given details of the case. They were asked to prepare by reviewing the preassigned readings and recorded presentation. For the SP encounter, participants were paired with one of four SP actors, who were TGD. The actors were trained to portray the character in a case study, which involved a TGD patient who presented to a primary care office with complaints of dysuria. The case was developed by the PI with input from a content expert and the four SPs. Standardized patients had completed a 2-hr, synchronous online training, with the PI in the weeks before the SP encounter. Training reviewed the purpose of the case, role-play, and effective debriefing. After a 15-min prebriefing with the PI, each participant was given 30 min to review their SP's health record and conduct a health history. The encounter did not include a physical examination. At the end of each encounter, the SP provided 1:1 feedback and completed a Standardized Patient Encounter Checklist for the participant. A 20-min group debrief led by the PI took place after all learners had completed their SP encounter. |  |
| Mueller 2023 | Before-and-after study | Nurse and nurse students<br>Sex: Male 3, Female 13<br>Age: 41-50 years | Setting: University<br>Country: USA | Facilitated learning only | The intervention contained didactic content, a patient panel, and a standardized patients (SP) scenario. (1) Readings and a voice-over. Readings and a voice-over PowerPoint presentation were distributed for review 1 week prior to the in-person experience. (PI), focused on the primary care of patients who are TGD and included gender identity, sexual orientation, sexual | 2 hours, one off training, NR |
|  |  | Sample size: 16 |  |  | practices/behaviors, health issues and disparities experienced by TGD patients, health history-taking skills, the use of the 5Ps, primary care and health maintenance, introduction to masculinizing and feminizing therapies, and community resources. These topics were chosen for their congruence with international guidelines for TGD health care. (2) The interactive patient panel discussion and the SP scenarios were held 1 week later. Two panelists who were transgender and/or nonbinary participated in a 1-hour discussion led by the PI. (3) Students were then assigned to 1 of 4 SP actors who were trained to portray the details of a character in a chosen case study. |  |
| Najor 2020 | Before-and-after study | Student: Medical students in the fall of their first year<br>Sex: Male 42, Female 44<br>Age: 20-39 years<br>Sample size: 86 | Setting: University<br>Country: USA | Facilitated learning only | A 1-h lecture was given to medical students in the fall of their first year at the MCASOM in 2017. Explain the natural variety in sex, gender, gender expression, sexual orientation, and sexual practice; Discuss examples of people who are diverse in these characteristics; Discuss and distribute list of common terms, with discussion of recommended language; Discuss major illness burdens to transgender people; Discuss the minority stress that transgender people face; View and discuss video vignette demonstrating transgender discrimination in the health care setting; Discuss examples of starting conversations by asking for preferred pronouns, of avoiding stereotypes, and recovering from mis-gendering; Discuss and distribute list of resources for antidiscrimination legal services, advocacy groups, counseling services, inclusion of LGBT + students and staff. | 1 hour, one off training, NR |
| Ness 2023 | Before-and-after study | Student: Students in their final year of the prelicensure nursing program<br>Sex: Male 1, Female 9<br>Age: 20-39 years<br>Sample size: 10 | Setting: University<br>Country: USA | Facilitated learning only | Prior to the simulation, all students attended one lecture in their regularly scheduled class focused on the health of LGBTQ+ populations. Students who agreed to participate in the pilot electronically completed the pretest and then completed simulation preparation materials including a review of the simulated patient's medical record and reading materials describing health needs of transgender individuals and gender-affirming nursing care. The simulation session began with a 15-minute prebriefing which included an orientation to the simulation environment, a review of expectations, and a hand-off report (INACSL Standards Committee, 2021). Two participants volunteered to provide nursing care while eight participants<br>observed live video on a large monitor in a separate viewing area. The simulation lasted 15 minutes. Immediately after, the two active participants engaged in five minutes of feedback from the SPs to hear how their care was perceived. Then, all participants were led in a reflective debriefing by a debriefer who was trained in Debriefing for Meaningful Learning (DML) and who is a clinical expert in gender-affirming nursing care. | 35 minutes, one off training, NR |
| Oblea 2022 | Before-and-after study | Civilian and military healthcare personnel, including physicians, psychologists, nurses, educators, administrators, social workers, counselors, case managers, dentists and first responders.<br>Sex: Male 25, Female 76<br>Age: NR<br>Sample size: 101 | Setting: Community health centre (Military and civilian sites on the island of Oahu in the state of Hawaii)<br>Country: USA | Facilitated learning only | The 4-hour program 'Delivering Culturally Sensitive Care to LGBT+ Patients: An Advocacy Program'. The program content includes sessions on how to obtain a health history; appropriate use of lesbian, gay, bisexual, transgender, queer, intersex, and other GSD terminology; review of physical and mental health disparities; barriers to accessing and seeking healthcare; a question and answer session with a transgender male; and strategies to improve communication and provide inclusive care in clinical encounters. | 4 hours, one off training, from 1 to 3 December 2021 |
| Pathoulas 2021 | Before-and-after study | Student: Second-year medical students at the University of Minnesota<br>Sex: NR<br>Age: NR<br>Sample size: 54 | Setting: University of Minnesota<br>Country: USA | Facilitated learning only | A 1-hour didactic and interactive lecture on GAHT. The didactic portion of the lecture focused on the following objectives:<br>(a) the scope of practice required to provide gender-affirming hormone therapy;<br>(b) an informed consent model of care;<br>(c) the medical management of masculinizing and feminizing hormone therapy, including dosing of relevant medications.<br>The interactive portion of the lecture consisted of actors role-playing an office visit in which a physician initiated GAHT.<br>The interactive lecture was designed to be stopped periodically so that audience members could be tested on concepts introduced, emulating real time clinical decision-making. | 1 hour, one off training, NR |
| Peterson 2020 | Before-and-after study | Nurse: Certified Registered Nurse Anesthetists (CRNAs)<br>Sex: NR<br>Age: NR<br>Sample size: 218 | Setting: Online<br>Country: USA | Self-learning only | An online educational repository called MyLearning; the online educational module entitled “Transgender Education and Anesthetic Considerations,” was assigned to the CRNAs to complete within two weeks. Reminders were sent to those who had not completed the online module every two days until the deadline. The online educational module took approximately 20 minutes to complete, and it included a video interview with a transgender female. | 20 minutes, one off training, 2 weeks duration |
| Quattrini 2023 | Before-and-after study | Student: Advanced practice nurse students<br>Sex: NR<br>Age: NR<br>Sample size: 38 | Setting: University<br>Country: USA | Mixed approach | Students completed a 1-hour, free online learning module from the National LGBTQIA+ Health Education Center. The learning module, titled Health Care for Transgender and Gender-Diverse People, included the basics of healthcare for TGD individuals. The formative SP encounter was conducted virtually in small groups of 2 to 3 students using a time-in/time-out format with one faculty facilitator. Students had 20 minutes to conduct the encounter, followed by SP debrief (5-10 minutes) and faculty debrief (15-20 minutes). | 1 hour, one off training, NR |
| REED 2021 (substudy 1) | Before-and-after study | Healthcare workers or trainees at hospitals, medical centers, universities, federally qualified health care centers, private practices, Veterans Affairs medical centers, and other health care settings<br>Sex: Male 62, Female 89<br>Age: 22-62 years<br>Sample size: 151 | Setting: In professional setting (e.g., at staff meetings, during routine didactics, as an invited presentation)<br>Country: USA | Facilitated learning only | In-person training on transgender-affirmative care: The training focused on presenting basic transgender terminology (e.g., defining sex and gender, discussing accurate pronouns), overviewing common barriers to care (e.g., stigma, insurance barriers), and providing suggestions to practice affirmative care for transgender individuals (e.g., updating EHRs, adapting traditional health care approaches). The training was split into two main foci, with the first half focusing on defining transgender identities and acknowledging and understanding barriers to care, and the second half focusing on overcoming barriers to care and creating a welcoming environment. More specifically, the first half provided information regarding the difference between sex, gender, and sexuality, discussed the differences between transgender and cisgender, overviewed the literature on gender identity development, and commented on insurance, routine, and transition-related barriers to care for transgender individuals along with the health outcomes (e.g., mental and physical health disparities).The second half proposed individual and organizational strategies to help healthcare workers understand ways to help overcome barriers to care. The proposed individual strategies generally included avoiding assumptions regarding gender identification, being cognizant of body language, addressing patients appropriately, providing anatomically appropriate care, using preferred language (e.g., pronouns, anatomy), and encouraging healthcare workers to apologize for mistakes. | 1.5 hours, 6 times, 3 months duration |
| REED 2021<br>(substudy 2) | Before-and-after<br>study | Healthcare workers or trainees<br>at hospitals, medical centers,<br>universities, federally qualified<br>health care centers, private<br>practices, Veterans Affairs<br>medical centers, and other<br>health care settings<br>Sex: Male 21, Female 116, Not<br>to answer: 1<br>Age: 18-70 years<br>Sample size: 138 | Setting: Online<br>Country: USA | Self-learning<br>only | Virtual training on transgender-affirmative care through a YouTube video with the content was the same with the in-person training. | 55 minutes, one<br>off training, NR |
| ROSA-VEG<br>A 2020 | Before-and-after<br>study | Doctor: Licensed pharmacists<br>who practice in<br>Puerto Rico<br>Sex: Male 8, Female 54, | Setting: University<br>Country: Puerto<br>Rico | Facilitated<br>learning only | The intervention content included 3 unites: (1) Transgender Patient Care Introduction, (2) General Health Issues of Transgender Patients, and ( 3) Gender Affirming Hormone Therapy. The activity incorporated a combination of didactic presentations as well as facilitated group discussions of cases that focused on the basics of transgender care and appropriate pharmacotherapy. | 3 hours, one off<br>training, NR |
|  |  | Unanswered 6<br>Age: 24-69 years<br>Sample size: 68 |  |  |  |  |
| Ruprecht<br>2024 | Before-and-after<br>study | Student: Second-year medical<br>students<br>Sex: Males 22, Females 37<br>Age: 22-37 years<br>Sample size: 59 | Setting: University<br>Country: Australia | Mixed<br>approach | Content relating to TGD health care was delivered as an hour-long online lecture through Zoom with opportunities for students to ask questions throughout. Topics covered included terminology; principles of social, legal and medical affirmation; clinical guidelines and treatment pathways for TGD adults, adolescents and children; informed consent model of care; barriers and discrimination; preventative health care; principles of gender-affirming history taking; and physical examination and safety. TGD volunteers were invited to attend the lecture and help answer questions. The 1.5-h tutorial (in small groups, also through Zoom) was designed to encourage students to practice gender-affirming history taking and discuss TGD volunteers' experiences with the health care system. Groups consisted of seven to nine students with a volunteer, who could bring a support person if they chose. The tutorial was facilitated by clinical skill tutors who are doctors without specific training or academic interests in TGD health. Online briefings were held for both tutors and volunteers before the tutorial session. | 2.5 hours, one off<br>training, NR |
| Ruud 2021 | Before-and-after<br>study | Midwifery students, women's<br>health nurse practitioner<br>students and obstetrics and<br>gynecology residents<br>Sex: NR<br>Age: NR<br>Sample size: 20 | Setting: NR<br>Country: USA | Facilitated<br>learning only | Two cases frequently encountered in clinics providing care to transgender individuals were developed for the simulation. Learners were divided into interprofessional groups of 3 and completed one of 2 case scenarios with a patient-teacher. Learners and patient-teachers debriefed after the simulation to discuss and reflect on the experience. | 105 minutes, one<br>off training, NR |
| Santamaria<br>2023 | Before-and-after<br>study | Doctor: Family pediatricians<br>Sex: Male 48, Female 48<br>Age: ≥20 years<br>Sample size: 96 | Setting: A room of the medical order of the city of Naples<br>Country: Italy | Facilitated<br>learning only | The training took place in a single day, lasted 6 h, and consisted of 2 sessions, one theoretical and one concerning clinical practice. The theoretical session, provided by experts in the field of gender diversity in childhood and youth, consisted of 3 sections.<br>(1) The first section addressed the theoretical foundations of sexual identity, focusing in particular on the structuring of gender identity, both congruent and incongruent with the sex assigned at birth. Particular attention was paid to the developmental trajectories of gender identity during childhood and adolescence.<br>(2) The second section was addressed to the depathologizing approach to gender diversity, both from a medical (i.e., endocrinological, pediatric, and neuropsychiatric) and psychological-clinical perspective, with an in-depth examination of the minority stress framework, the impact that sexual- and gender-based stigma can have on the person's psycho-physical health, and the health disparities of this population.<br>(3) The third section was dedicated to the role of family pediatricians as the first contacts of the families, essential for the early identification of children and adolescents who need specialized support.<br>(4) The clinical practice session consisted in presenting to the pediatricians a clinical scenario, allowing for the activation of a group reflection on the management of GD adolescents and their possible referral to specialized care (e.g., case report). | 6 hours, one off training, 3 months duration |
| Sengupta<br>2023 | Before-and-after<br>study | Student: First-year undergraduate medical students<br>Sex: NR<br>Age: NR<br>Sample size: 65 | Setting: The annual NEPA Trans Health Conference<br>Country: USA | Facilitated<br>learning only | The 1-hour breakout sessions included the following topics: introductory basic terminology and theory as they relate to transgender health (Transgender 101); disparities and ways to improve the trans experience in health care through gender-affirming care; addiction and the transgender population; implications for the pediatric population and family support; transitioning in the school setting; medical affirming hormones and gender confirmation surgery. All students then attended a panel discussion that included at least four trans community members eager to discuss their personal narratives and healthcare experiences in an open question-and-answer forum. | 1 hour, one off training, NR |
| Sherman<br>2021 | Before-and-after<br>study | Nursing student<br>Sex: Male 16, Female 144<br>Age: ≥21 years<br>Sample size: 160 | Setting: University<br>Country: USA | Facilitated<br>learning only | The intervention included: (1) presentation and approval of the TCIP project from the Baccalaureate Curriculum Committee; (2) identification of key topics of importance based on literature review and key-informant interviews (included topics: health inequities, gender-affirming language and best practices, preventive health, mental health, gender-affirming interventions, and prenatal care); (3) meetings with faculty members teaching key courses (Professional Role Development in Nursing, Health Assessment, Pharmacology, Psychiatric-Mental Health Nursing, and Nursing in the Childbearing Family) to create and integrate content into their specific courses; and (4) evaluation of student knowledge and attitudes toward care of TGD people at various time points. Integrated content additive to the core curriculum were delivered via stand-alone webinars, standalone in-class and online lectures, and integrative adaptations to existing in-class and online lectures. | NR |
| Smith 2023 | Before-and-after<br>study | Doctor: Family Medicine<br>Residents<br>Sex: NR<br>Age: NR<br>Sample size: 12 | Setting: Hospital<br>Country: USA | Mixed<br>approach | All residents, who had participated in a mandatory 2-hour GAC didactic earlier in the year, received an e-mail with compiled feedback on gaps identified during preintervention chart review. We also held two optional educational sessions for residents and faculty covering more advanced GAC topics and an optional 1-hour presentation on creating an affirming clinic environment. Additionally, we developed and distributed multiple documentation templates for GAC visits, including suggested questions for new patients, standard language for hormone counseling and informed consent, and reminders about contraception, monitoring labs, medication dosing, and followup intervals. Lastly, we compiled dosing guidelines, counseling guides, and educational handouts into a physical binder in the residents' workspace as well as distributed this information electronically to all resident/faculty providers. | NR |
| Thompson<br>2020 | Before-and-after<br>study | Students<br>Sex: NR<br>Age: NR<br>Sample size: 81 | Setting: University<br>Country: USA | Mixed<br>approach | The online videos totaled 1 hour and 35 min in length and included didactic material. Students were given approximately 1 week to review this material in preparation for a required case-based workshop followed by a 10-question quiz. On the date of the workshop, students were separated into preassigned classrooms with one faculty member and rotating subject-matter experts, who were faculty or staff, facilitating a group of approximately 15–17 students per class. The 3-hour workshop included case-based learning centred on a gender non-conforming, specifically non-binary, patient with the pronouns of they/them/theirs. The case and its discussion questions (online supplementary appendix) were specifically developed to provide students with an opportunity to determine appropriate primary care for the patient, discuss social and medical gender-affirming options, provide culturally sensitive sexual health counselling, and practice addressing patients using they/them/theirs pronouns. After completing the case, students engaged in three other activities, including: (1) role-playing with different pronouns; (2) review of practice questions formatted to represent those seen on medical board examinations; and (3) discussion of additional case vignettes developed to illustrate the range of patient experiences and medical needs. These vignettes also provided counter-stereotypes to challenge common assumptions held by clinical providers. A psychiatrist and endocrinologist, who work with TGNC patients, as well as trans and non-binary faculty and staff designed all didactic content and activities. On a separate day after the workshop, students attended one of two, 2-hour panel discussions. Students viewed a short video titled ‘Voices of Transgender Adolescents in Healthcare’<sup>19</sup> prior to hearing from a panel consisting of TGNC individuals and cisgender healthcare providers who have experience working with this population. Contracted as speakers for the medical college, the TGNC panellists ranged in gender identities, race and ethnicity and were aged from 20 to 64 years. Transgender and non-binary faculty and staff moderated the panel and asked questions regarding access to care as well as barriers and facilitators for providing and receiving quality care. The post-assessment data were collected after students had attended this panel discussion. | 1.35 hours of self-learning, plus one day workshop, plus 2 hours of panel discussion on the next day after the workshop, NR, NR |
| Tsusaki 2023 | Before-and-after study | Student: advanced-practice nursing student<br>Sex: Male 3; Female 24<br>Age: 22-56 years<br>Sample size: 28 | Setting: University<br>Country: USA | Mixed approach | An e-Learning course development software, Articulate 360® (Articulate Global, NY), was used to create the (sexual and gender minority) SGM curriculum. Articulate 360® allowed for the inclusion of multiple media types and interactive exercises to build on learning. An instructional designer was consulted to develop the sections of the e-module which included a series of mini lectures, interactive learning exercises, an unfolding case study and instructional videos. The mini lectures that were 10 to 15 min in length, addressed SGM demographics in the U.S., terminology and concepts, health disparities and social determinants of health in the SGM population, healthcare providers' attitudinal awareness and biases and creating an inclusive clinical environment. A video lecture series was also created by the researchers that included taking a sexual health history, a podcast-style interview with a nurse who identifies as a SGM community member sharing subjective experiences navigating the healthcare system and a nurse practitioner with expertise in transgender healthcare on how to care for the transgender population. The course concluded with an interactive unfolding case study that engaged students in providing care for a SGM patient followed by questions requiring thoughtful insight and understanding of the important concepts. | 16 weeks, one off training, NR |
| Vance 2020 | Before-and-after study | Student: The fourth-year medical students, interns, and nurse practitioner trainees<br>Sex: NR<br>Age: NR<br>Sample size: 31 | Setting: Medical institutions<br>Country: USA | Mixed approach | The intervention including: (1) online curriculum comprised six modules, which took learners through virtual clinical encounters with three transgender youth. We posted the modules on the Collaborative Learning Environment, our institution's dedicated educational platform. (2) 1-afternoon observership in the pediatric transgender clinic. | 77minutes (online modules) + 1-afternoon observership, one off training, NR |
| Vance 2021 | Before-and-after study | Doctor: Pediatrics interns, Psychiatry interns, Fourth-year medical students, Nurse practitioner trainee<br>Sex: Male 14, Female 29<br>Age: NR<br>Sample size: 43 | Setting: Hospital-based adolescent and young adult clinic and community-based clinics<br>Country: USA | Mixed approach | The curriculum included two components: (1) online modules and video clinical vignette: 6 online modules feature the cartoon characters of Dr. Resident and 3 transgender youth at various pubertal stages as they progress through a clinic evaluation. For this study, we added a video clinical vignette to standardize how learners were exposed to an observational experience prior to standardized patient encounters (SPEs); (2) SPEs: We developed 2, 20-minute formative SPE cases for learners to practice gender-affirming communication skills in evaluating and counseling transgender youth. The cases simulate a clinical encounter with Jane, a 16-year-old transgender female, and Kel, a 16-year-old transgender male, who both present for teen clinic follow-up visits with questions about starting gender-affirming hormones. Cases are based on typical presentations of transgender teens followed in teen clinic. | 128 minutes, one off training, NR |
| Vann 2021 | Before-and-after study | Doctor: Mental health practitioners<br>Sex: Male 14, Female 65<br>Age: 22-64 years<br>Sample size: 79 | Setting: Online<br>Country: Australia | Self-learning only | The training was hosted by a university using Lime Survey and began by outlining key concepts of gender and sex followed by six further sections. These included key terms, pronouns, non-binary population size statistics, relevant research regarding unique challenges, tips for supporting non-binary people, and links to additional resources. There were some embedded videos from YouTube portraying experiences of gender diverse people and suggestions for creating more gender affirmative environments. The training also had optional links, videos, and resources, although it was otherwise not directly interactive. | Mean 34 minutes (SD=22) recorded by Lime; Mean 27 minutes (SD=14) reported by participants, NR, NR |
| White Hughto<br>2019 | Before-and-after<br>study | Correctional healthcare<br>providers<br>Sex: NR<br>Age: NR<br>Sample size: 22 | Setting: New<br>England correctional<br>facility<br>Country: USA | Facilitated<br>learning only | The intervention consisted of four training modules with specific learning objectives. Module 1 covered definitions and terminology (e.g., difference between sex, gender, sexual orientation) in order to enable providers to define sexual and gender-related terms and processes. Module 2 aimed to help providers understand the social factors that place transgender people at risk for incarceration by covering examples of transgender stigma as a barrier to care. Module 2 also included an account of incarceration experiences from a formerly incarcerated transgender community activist to help providers understand the aspects of incarceration that are most challenging for transgender individuals. Module 3 included an interactive role-play component that aimed to help providers communicate with transgender people using language that respects and acknowledges their gender identities through gender-affirming interactions. Module 4 aimed to help providers understand their role in ensuring patient access to gender-affirming care through discussion on how stigmatizing and supportive interactions between patients and providers can serve as barriers and facilitators to care, respectively. Module 4 also aimed to educate providers on the basics of taking a medical history with transgender individuals by reviewing the essential components (e.g., assess existing anatomy) and key considerations (e.g., trauma histories) for transgender individuals. | 1.5-2 hours,<br>4times, NR |
| Wright 2022 | Before-and-after<br>study | Medical students<br>Sex: NR<br>Age: NR<br>Sample size: 8 | Setting: Walter Reed<br>National Military<br>Medical Centre<br>Country: USA | Mixed<br>approach | The intervention consisted of four training modules with specific learning objectives. Module 1 covered definitions and terminology (e.g., difference between sex, gender, sexual orientation) in order to enable providers to define sexual and gender-related terms and processes. Module 2 aimed to help providers understand the social factors that place transgender people at risk for incarceration by covering examples of transgender stigma as a barrier to care. Module 2 also included an account of incarceration experiences from a formerly incarcerated transgender community activist to help providers understand the aspects of incarceration that are most challenging for transgender individuals. Module 3 included an interactive role-play component that aimed to help providers communicate with transgender people using language that respects and acknowledges their gender identities through gender-affirming interactions. Module 4 aimed to help providers understand their role in ensuring patient access to gender-affirming care through discussion on how stigmatizing and supportive interactions between patients and providers can serve as barriers and facilitators to care, respectively. Module 4 also aimed to educate providers on the basics of taking a medical history with transgender patients by reviewing the essential components (e.g., assess existing anatomy) and key considerations (e.g., trauma histories) for transgender patients. | 1 hour, 2 times, 2<br>weeks |
| Zheng 2022 | Before-and-after study | Medical students<br>Sex: NR<br>Age: NR<br>Sample size: 80 (2020 students:33; 2021 students: 47) | Setting: University<br>Country: USA | Facilitated learning only | The session totaled approximately 2 hours and was held in classrooms that allowed groups of five students to sit and discuss with one another without disturbing nearby groups. Due to the coronavirus pandemic, the 2021 session was held over Zoom, making use of breakout rooms, but overall structure remained the same compared to 2020.<br>2020 education intervention: (1) Presession survey and pretest with Discussion (1 hour 25 minutes); (2) Case discussion: 2020 (30 minutes); (3) posttest and postsession surveys (15 minutes).<br>2021 education intervention: (1) 21-question survey; 15-question test: physiology and transgender medicine; Group of 4-5: Each student has answers to 2-3 questions teaches group; (2) Group video discussion: 2021 (30 minutes); (3) 6-question test: trans medicine only; 15-questionsurvey. | 2 hours, one off training, NR |
| Besse 2023 | Comparative controlled study | Students: Undergraduate medical students<br>Sex: NR<br>Age: NR<br>Sample size: 134 | Setting: University<br>Country: Germany | Mixed approach | One seminar of 90-minute duration on the topic of transgender identity was held in six small groups (A-F) of no more than 25 students, Groups A and D were taught by a specialist in psychiatry with additional qualification in sexual medicine, groups B and E by a specialist in psychiatry (specialist teaching). For groups C and F, two lecturers from the local “Queer Center” were recruited (community-supported teaching). Both lecturers, who themselves live transident, have been involved in counseling of trans* persons for many years, regularly give lectures on the topic of transgender identity and publish scientific articles in this field. All lecturers were informed about the learning objectives and the structure of the seminar. An interactive discussion had to be part of the seminar, with further design left to the lecturers. | 90 minutes, one off training, NR |
| Bettergarcia<br>2024 | An experimental<br>waitlist control<br>group design | Psychologist/counsellor:<br>Mental health providers<br>(MHPs)<br>Sex: Male 6, Female 41<br>Age: 44.5± 11.1 years<br>Sample size: 47 (IG:CG=25:22) | Setting: Community<br>health centre<br>Country: USA | Mixed<br>approach | The intervention program was designed as a 9-month training including three training weekends and six clinical consultation groups. Each training weekend included two 7-hr training days, with 6-hr of training content and 1 hr of break time each day, for a total of 36 continuing education hours. The training weekends included 22 modules over the six training days (see Appendix, for training curriculum). The three training weekends were evenly spaced across the 9 months, with approximately 2 or 3 months between training weekends. The first two weekends (4 days) were conducted locally in-person in a conference room style training location. Due to the COVID-19 pandemic, the final weekend (2 days) was conducted via Zoom. | 36 hours, 6 times,<br>9 months duration |
| García-Acosta<br>2019 | RCT | Student: 3th-year and 4th-year<br>nursing school students<br>Sex: Male 29; Female 87<br>Age: 19-48 years<br>Sample size: 116 (G1: G2:<br>G3=31: 28: 57) | Setting: University<br>Country: Spain | Mixed<br>approach | G1: Total 2 films: worked on the past and current history of transgender people, one per session: The Danish Girl; About Ray, after round tables + debates (including professional lectures, Transgender people themselves and their families told the students about their diverse realities and the problems they had in accessing health services, and the interaction of students working in groups).<br>G2: The PBL group worked on parenthood in transgender people and aspects of this in assisted reproduction, as well as the current state of the situation about this regard. after round tables + debates (including professional lectures, Transgender people themselves and their families told the students about their diverse realities and the problems they had in accessing health services, and the interaction of students working in groups). | NR, one session<br>per week, 3 weeks<br>duration |
|  |  |  |  |  | G3 (control group): did not receive any type of training or specific activity regarding healthcare for transgender people. |  |
| Mayer 2024 | RCT | Allied Healthcare: Health Center Staff<br>Sex: NR<br>Age:NR<br>Sample size: NR | Setting: Hospital: health centre<br>Country: USA | Mixed approach | The “Ed Centre” developed a three-phase approach that included a readiness assessment, educational interventions, and training and technical assistance to achieve sustainable change<br>Between September 2018 and February 2019, the intervention team provided onsite trainings to intervention HC staff. The intervention team consisted of an academic physician and a senior health informaticist. Specific trainings were tailored for each HC. Phase 2 of the training was specifically designed to address processes for SOGI data collection workflow implementation, both with regard to personnel and information technology considerations. This training emphasized optimizing systematic, standardized SOGI data collection with all patients by HC frontline staff, who directly engage patients in SOGI data collection. In addition, phase 3 of the training offered HCs consultation with and technical assistance by experts in SOGI data collection to troubleshoot site-specific barriers to implementation. | NR, 3-10 times, NR |
| Michals-Voigt 2021 | RCT | Student: Counselling, social work, and psychology graduate students<br>Sex: NR<br>Age: 23-50 years<br>Sample size: 135 | Setting: University<br>Country: USA | Self-learning only | A Transgender specific educational PowerPoint presentation(Fenway Institute, 2017), which addressed the terminology, health disparities, and creating an affirmative, inclusive environment for Transgendered individuals. The PowerPoint was made available to the control group after the data has been collected. | 10 minutes, one off training, NR |
|  |  | (IG:CG=70:65) |  |  |  |  |

Notes: NR=not reported; RCT=Randomized controlled trial; IG=intervention group; CG=control group.

**Appendix 3:**
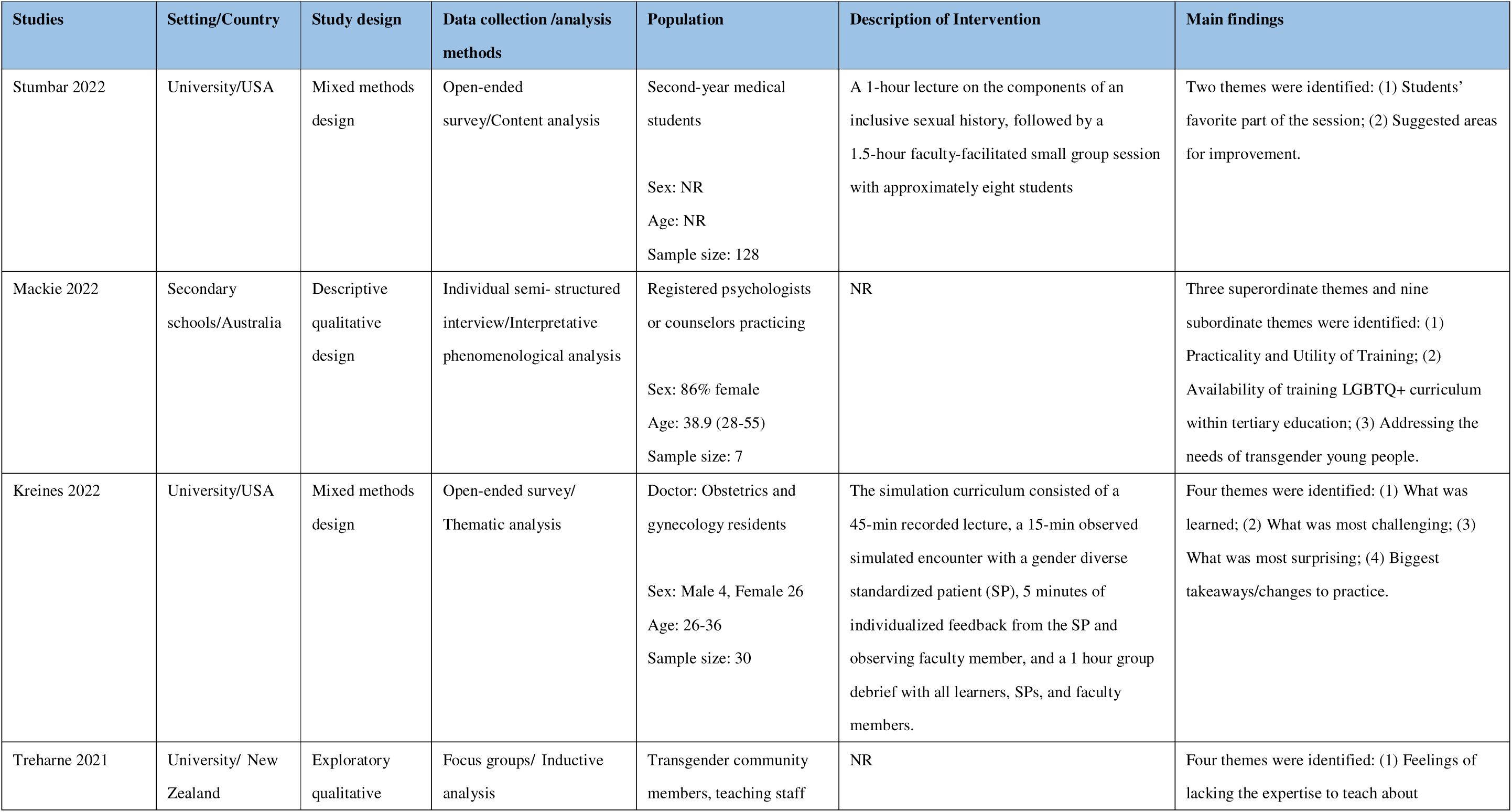
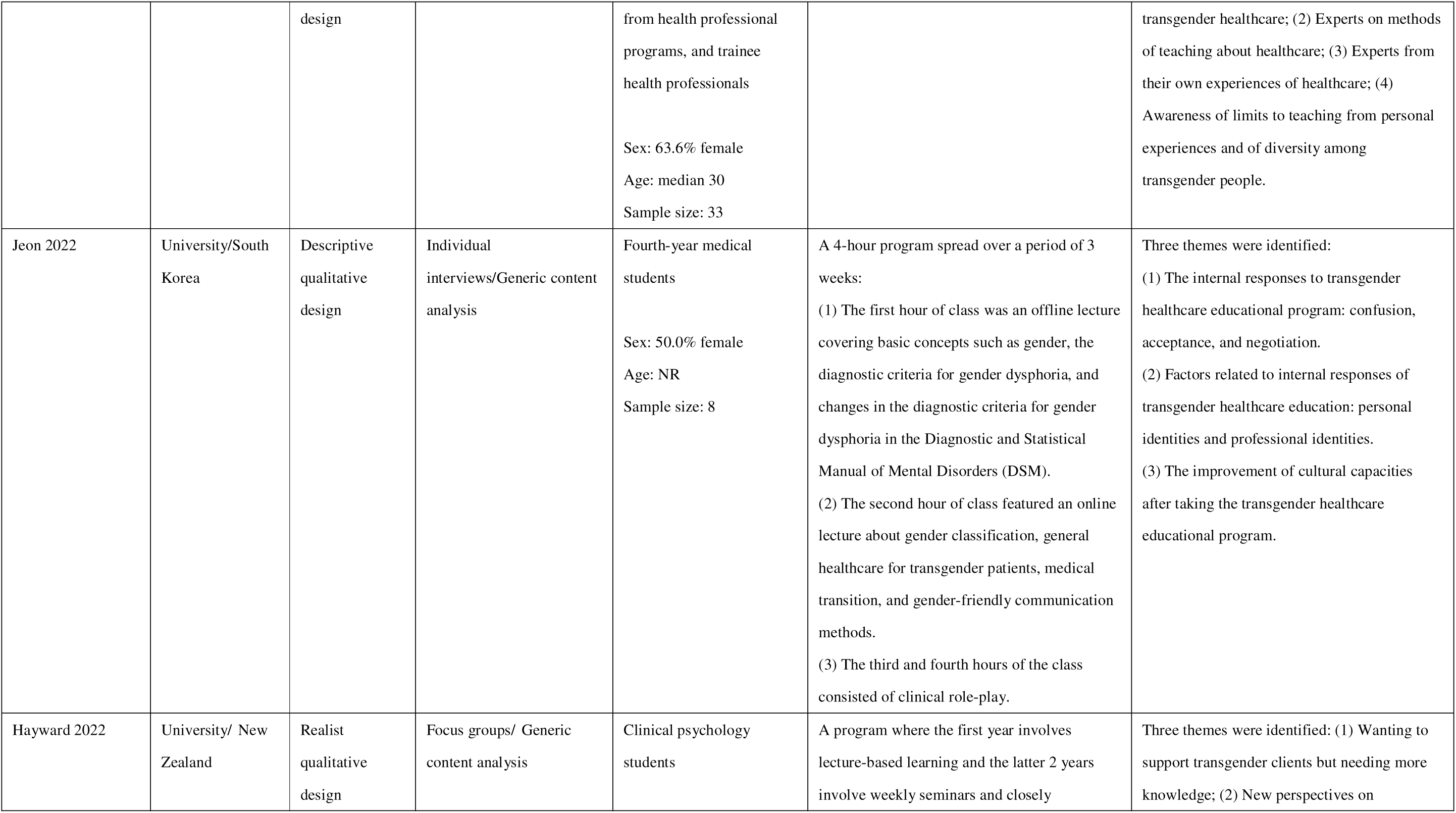
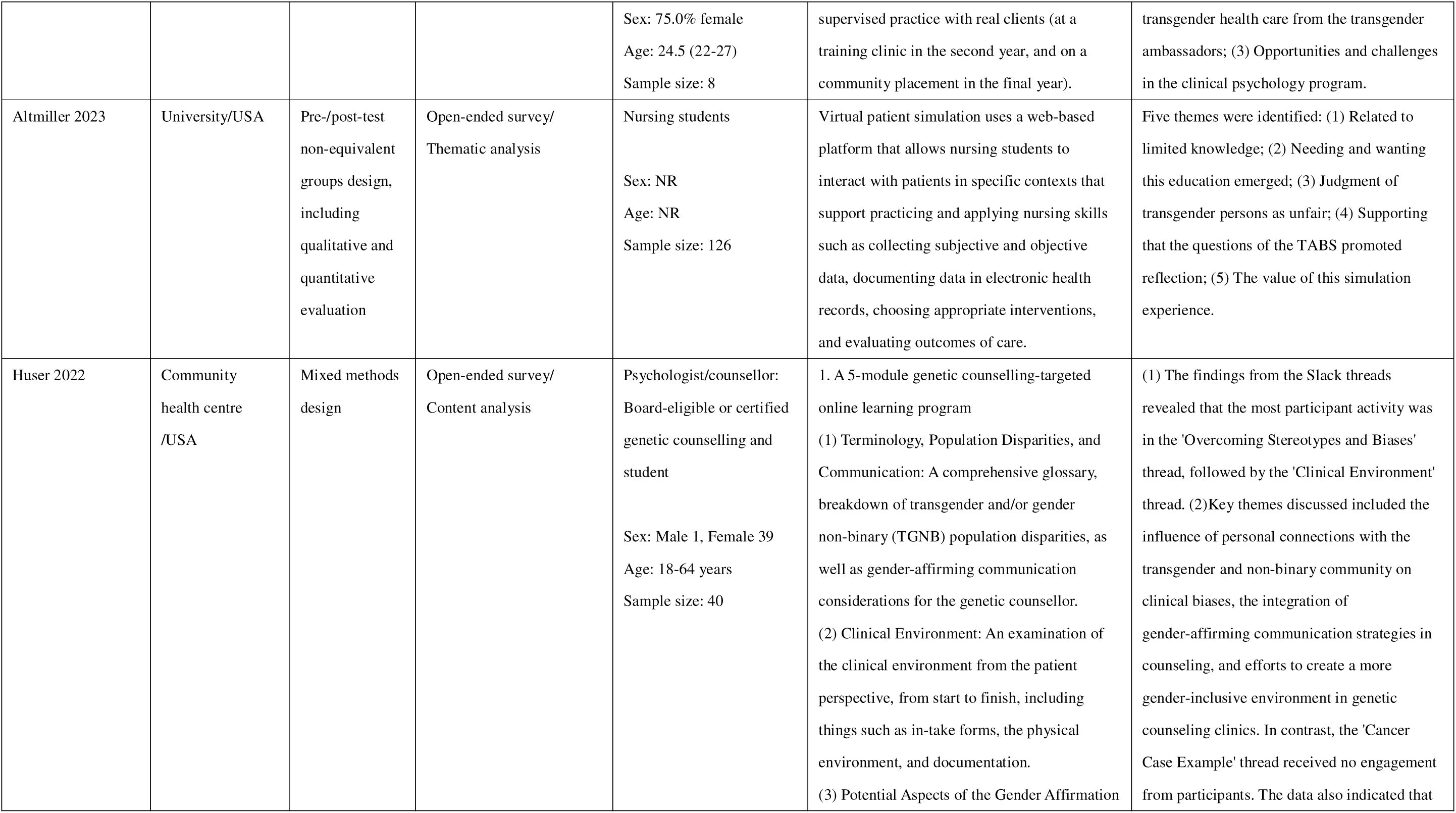
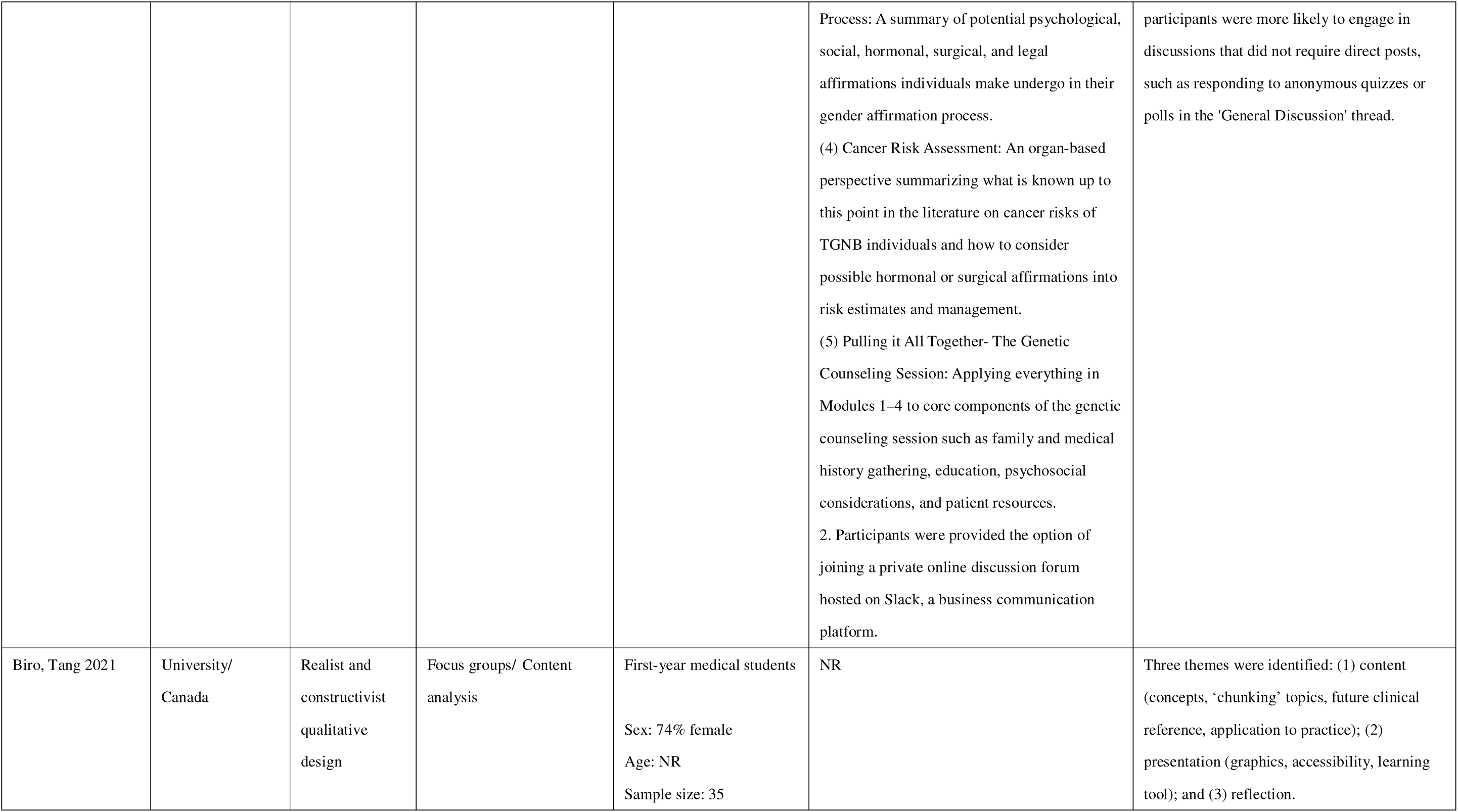
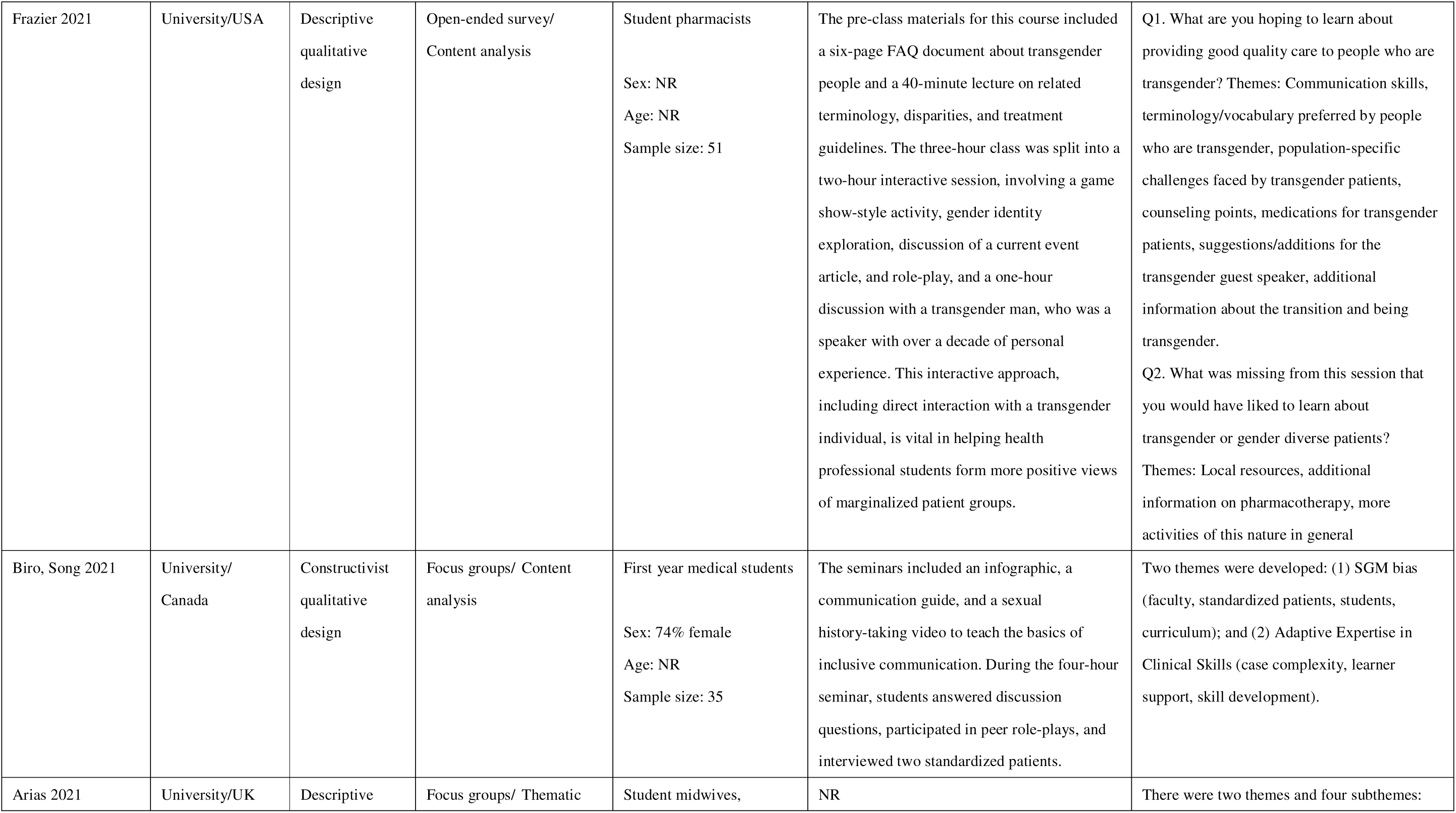
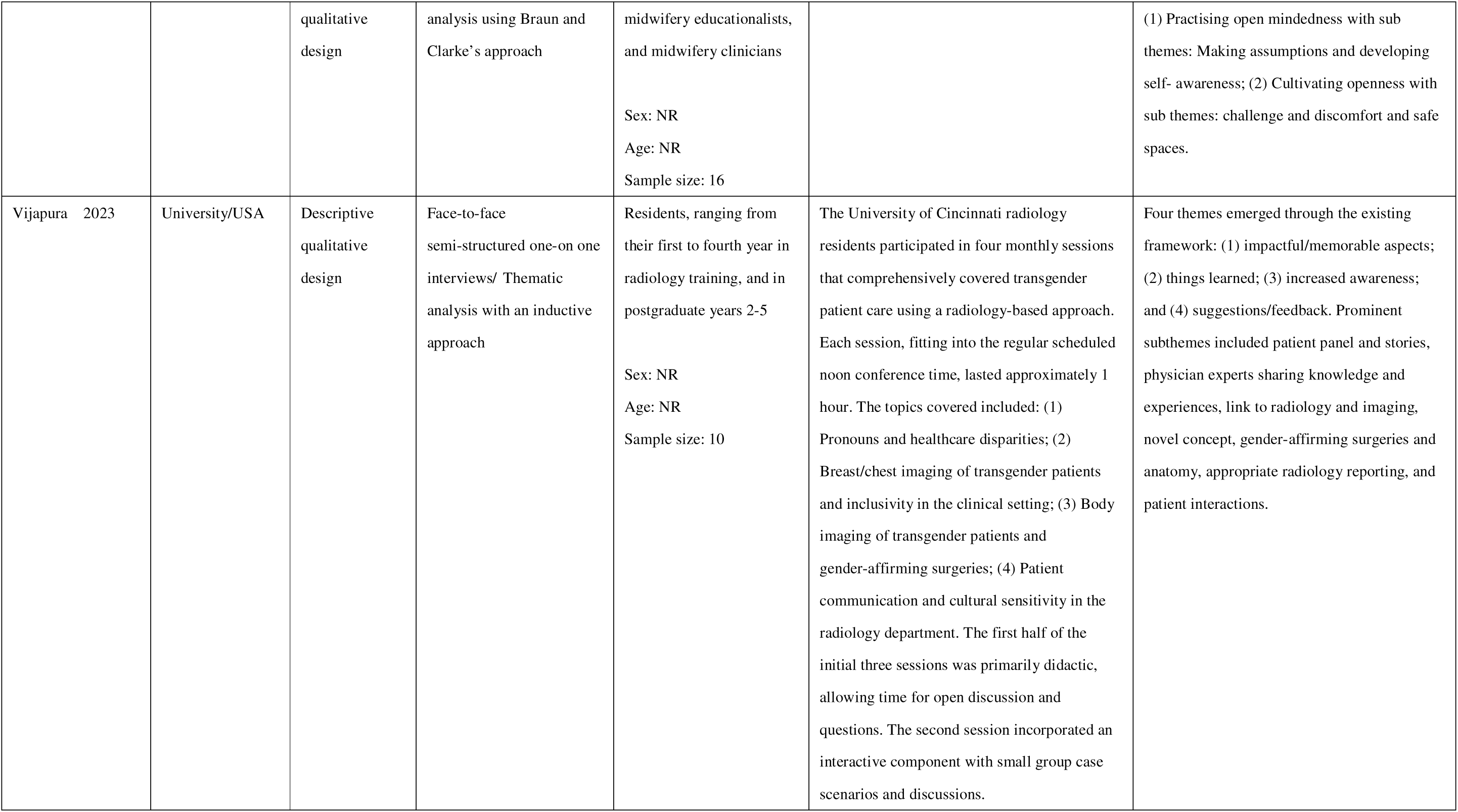
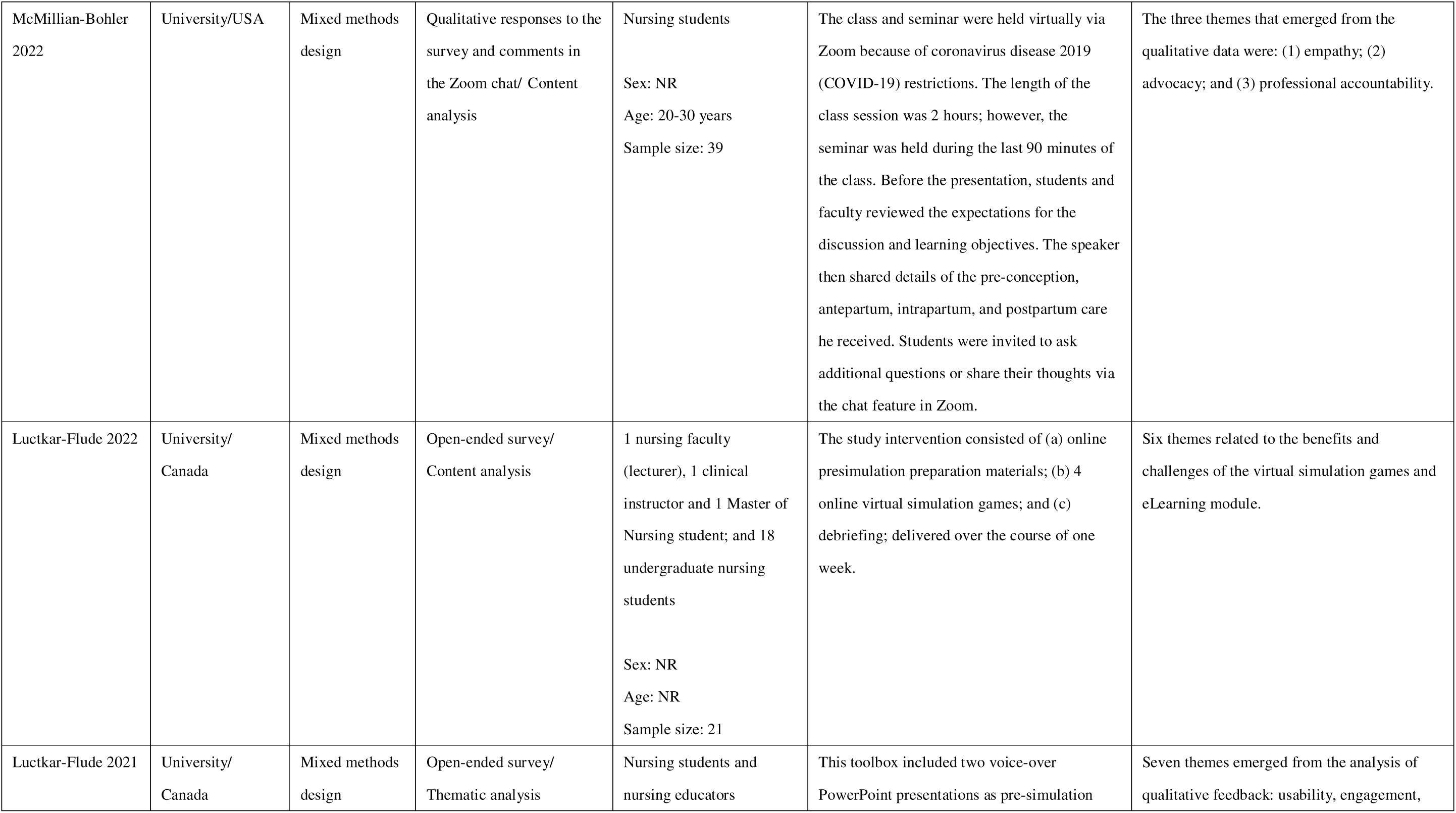
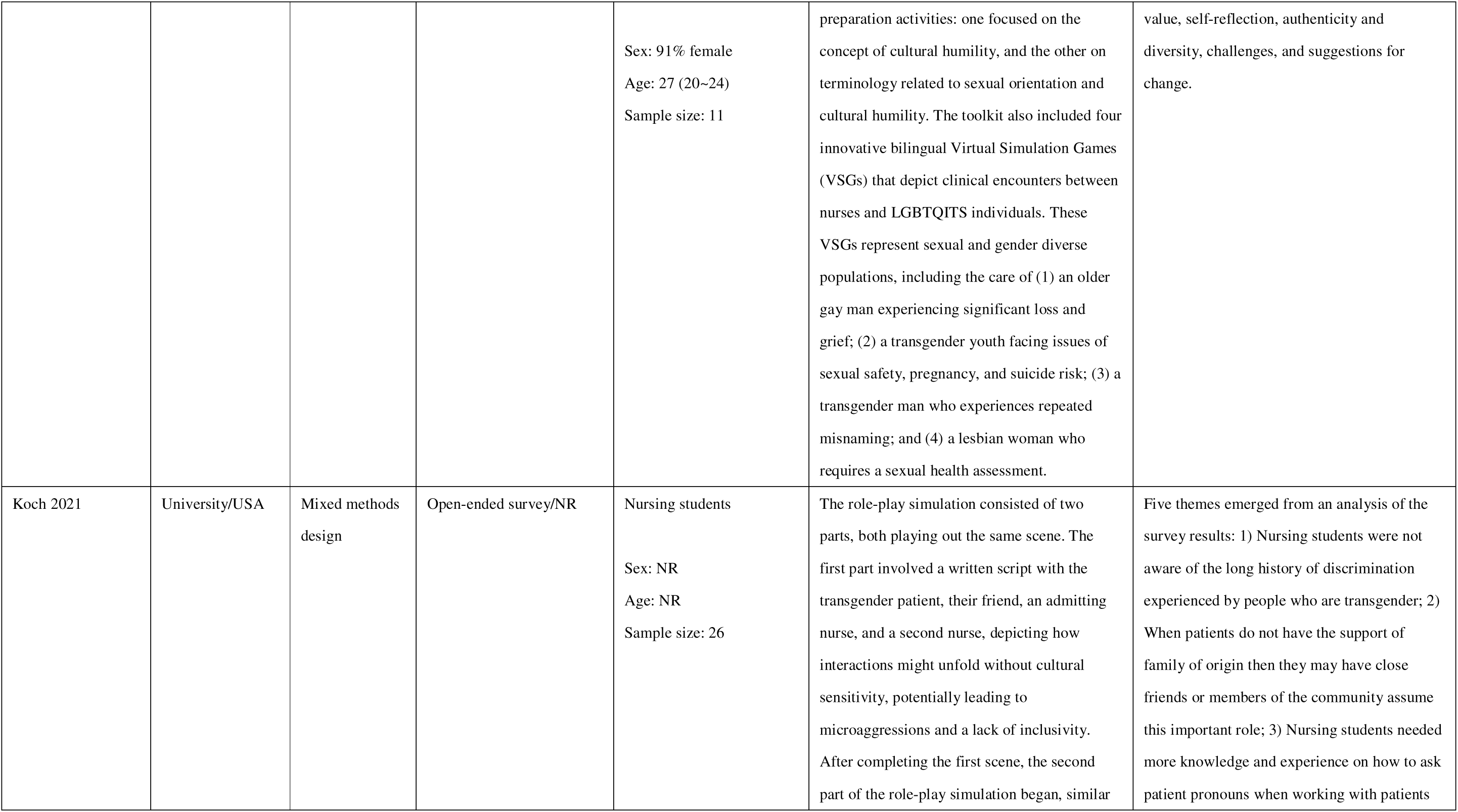
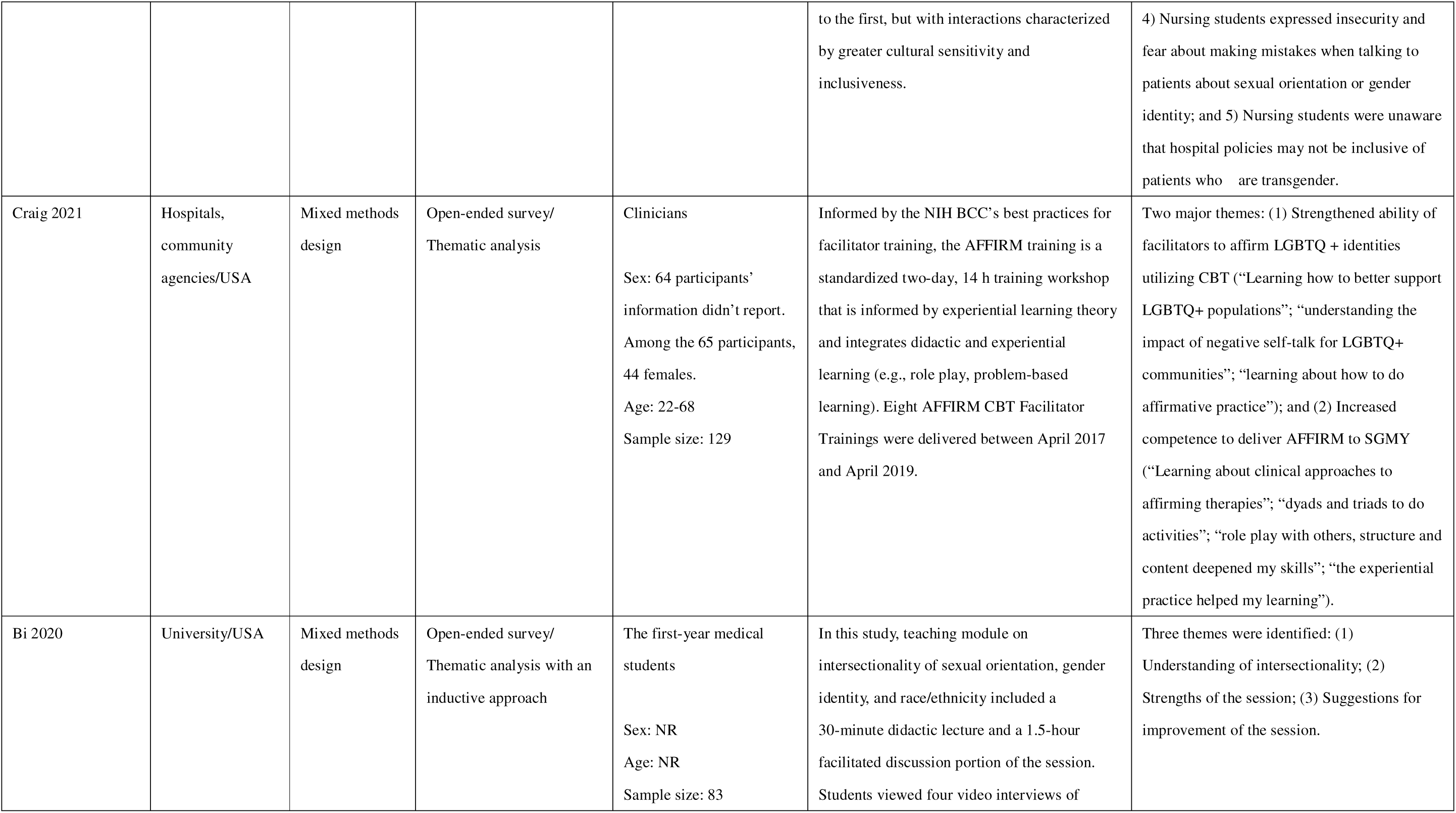
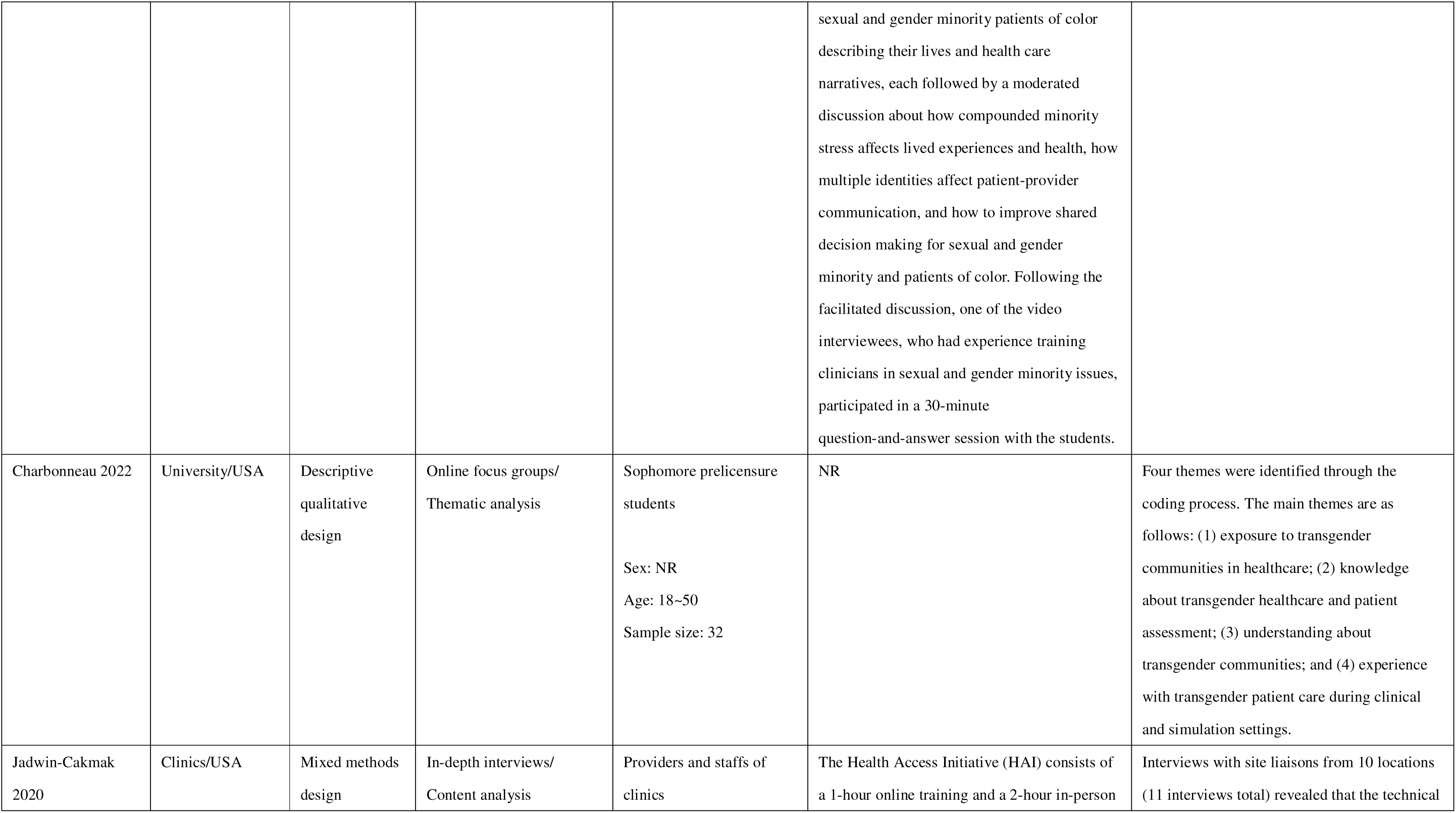
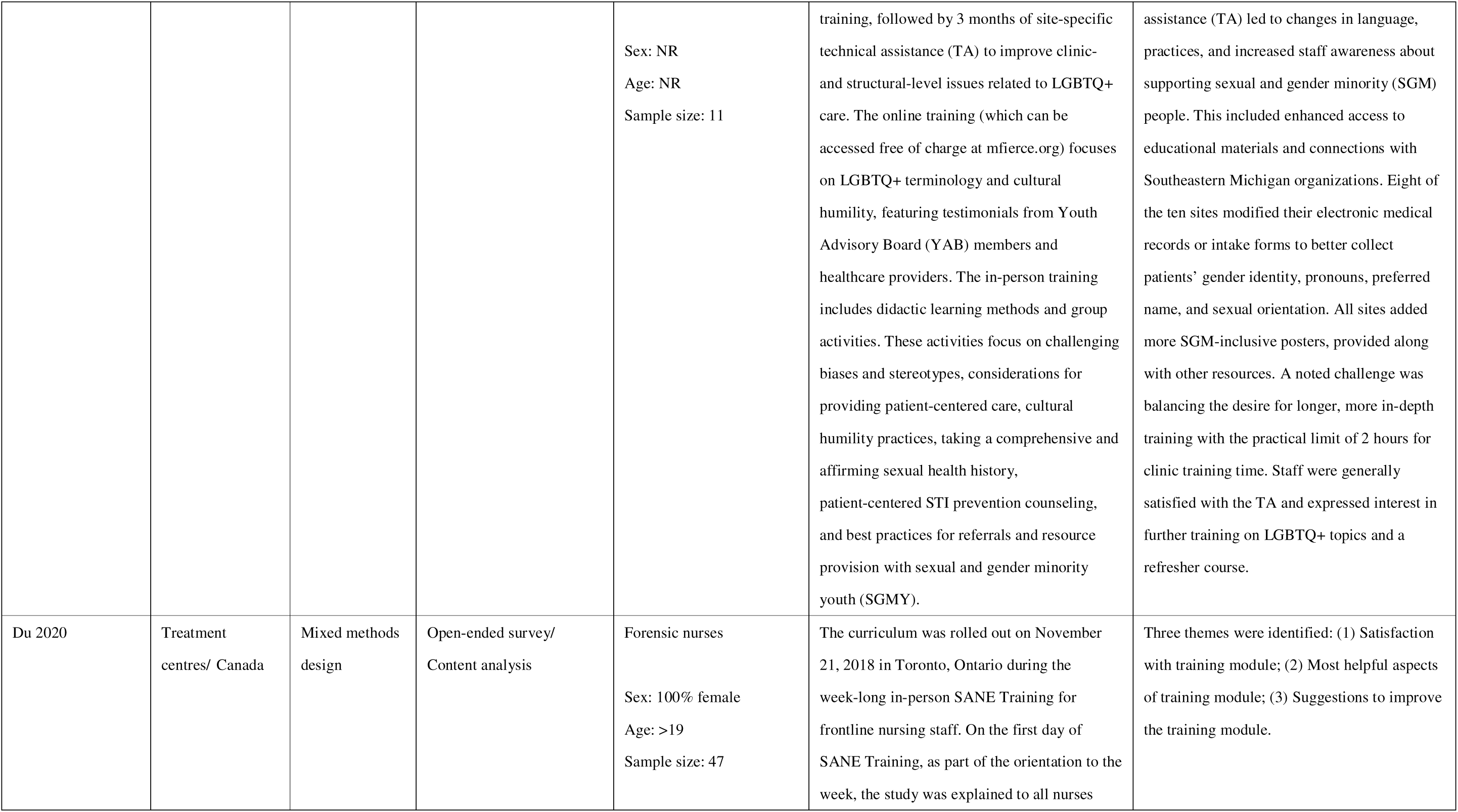
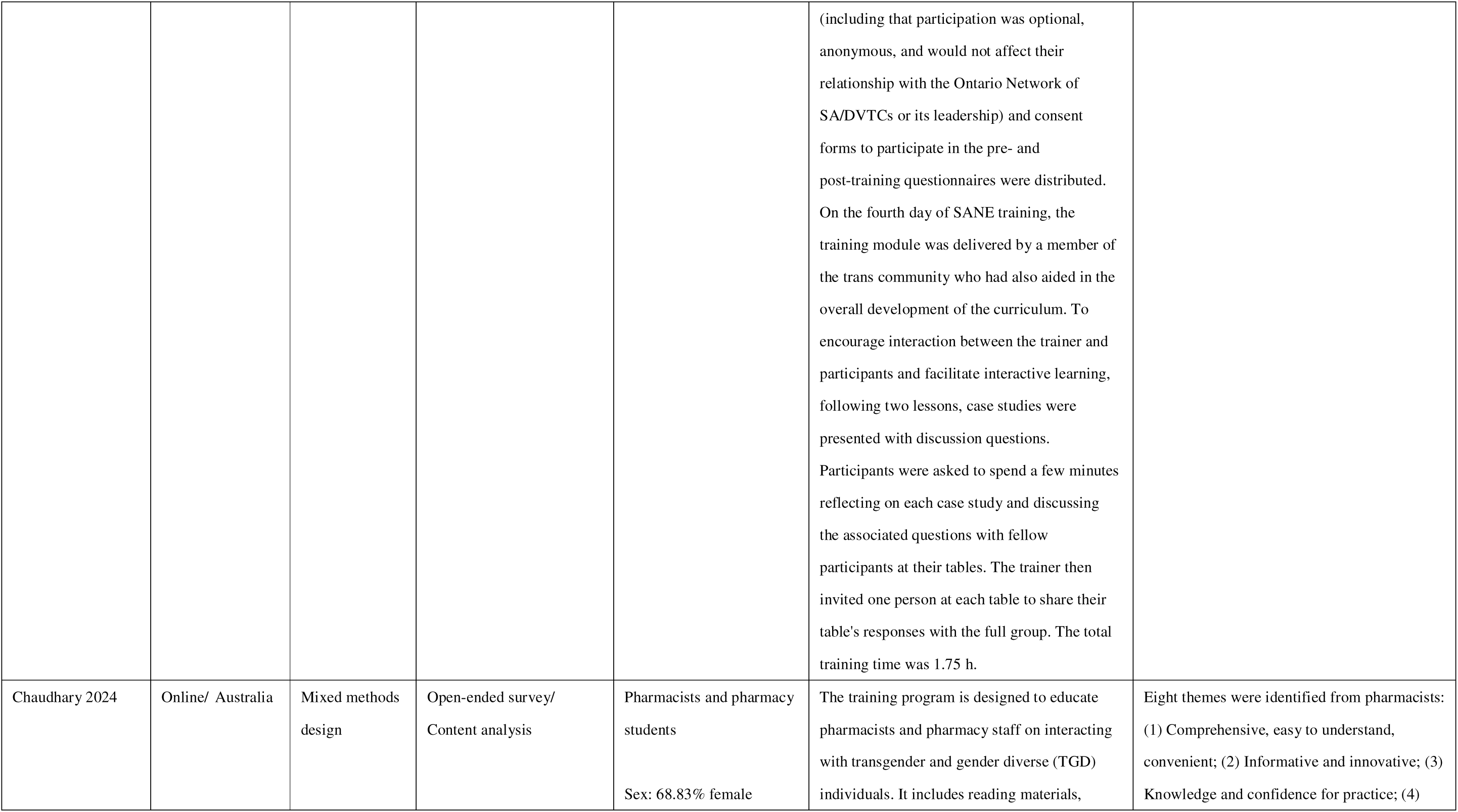
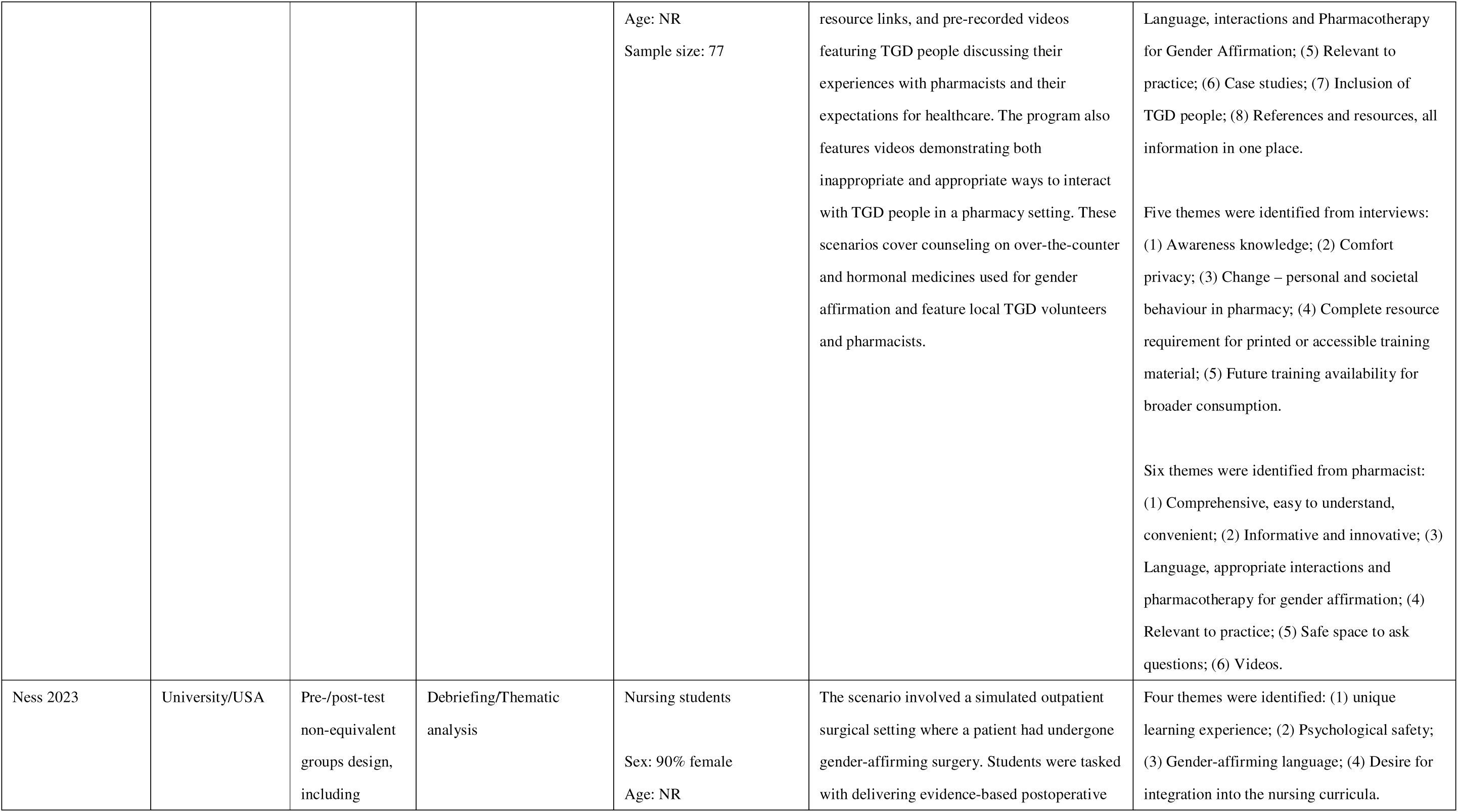
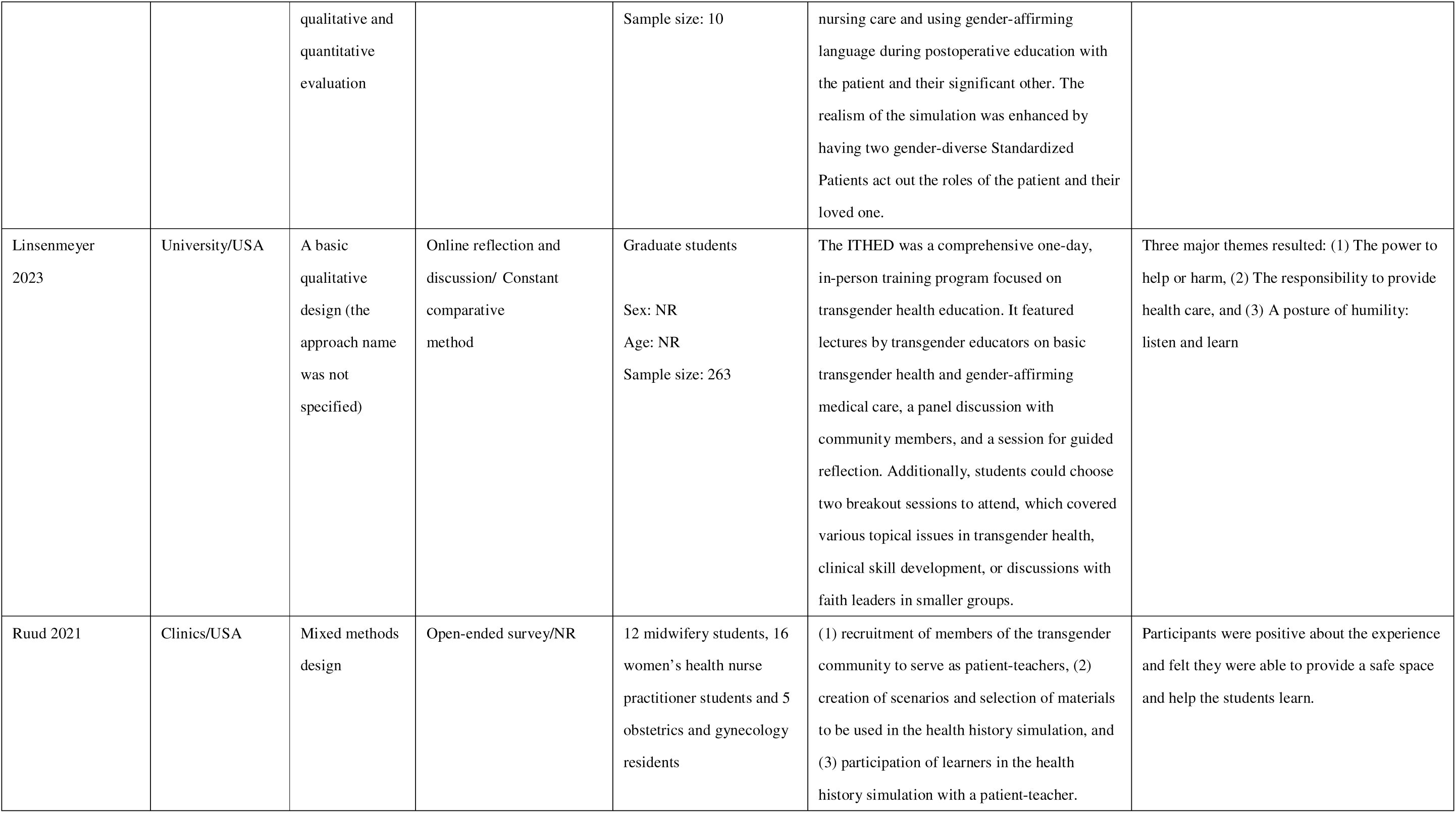
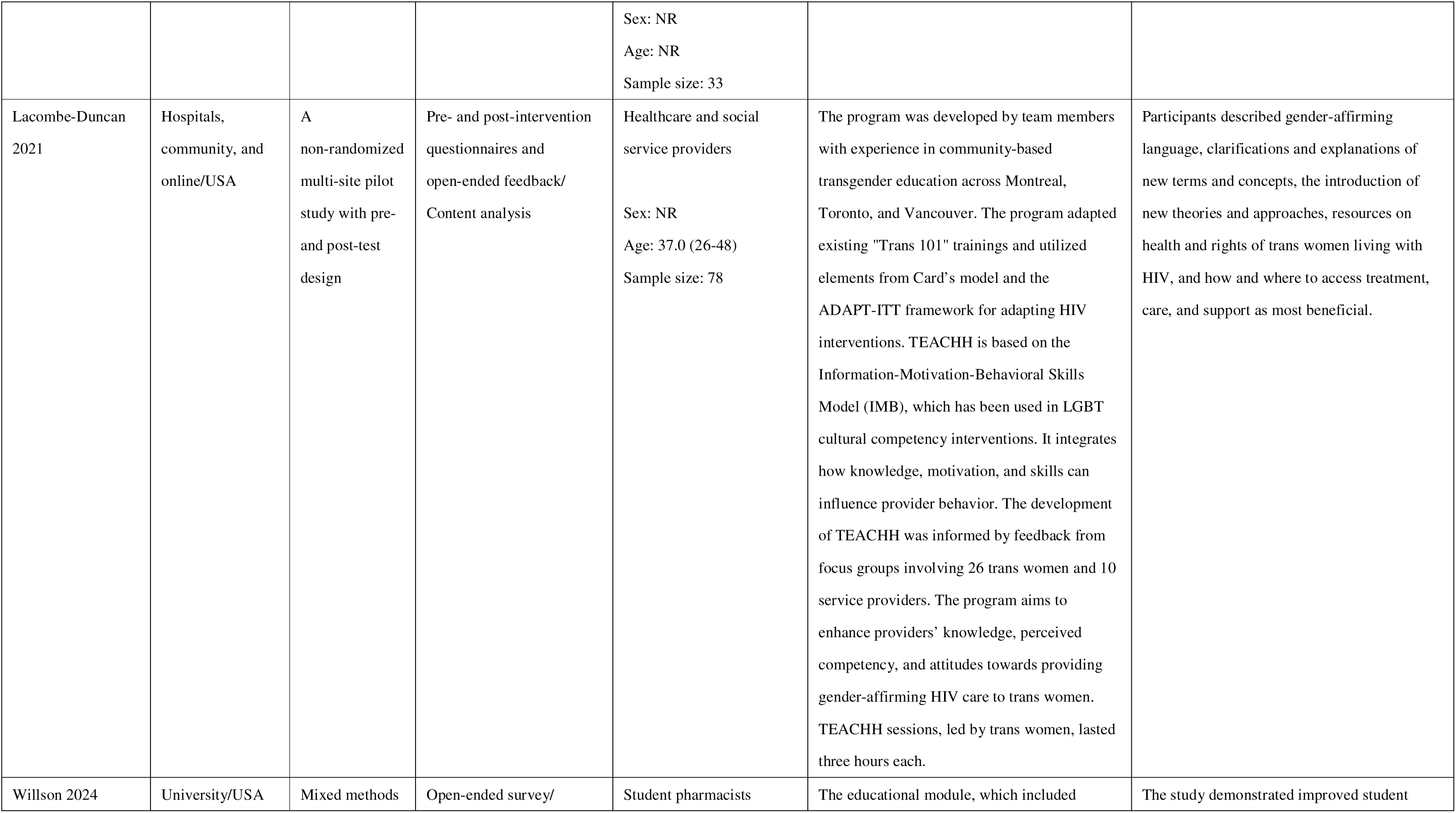
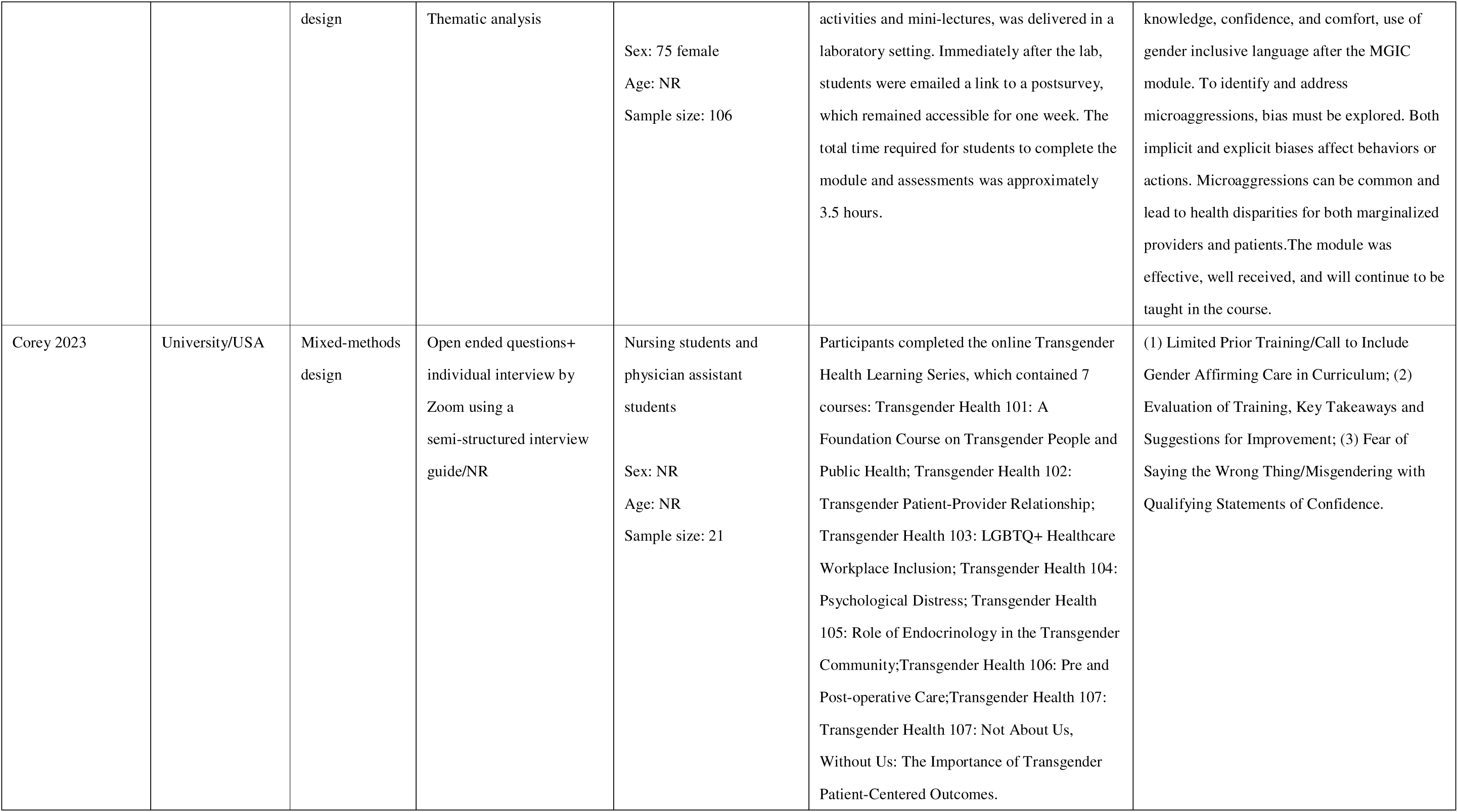
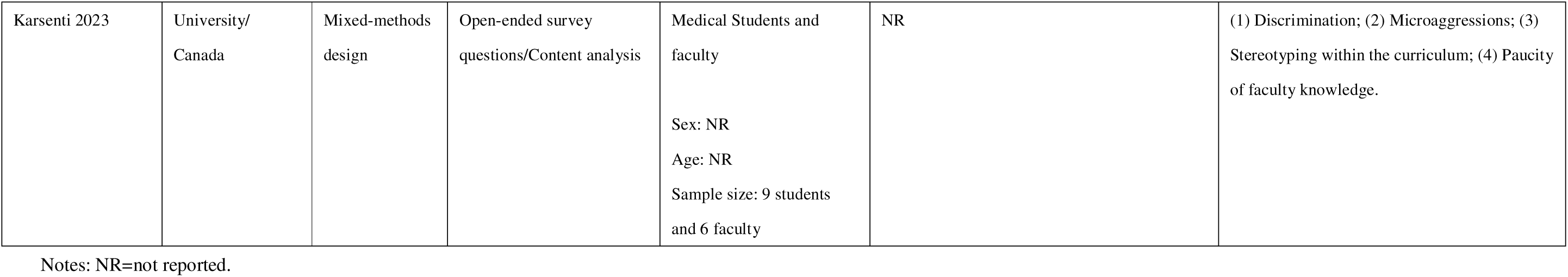
Characteristics of all included qualitative studies (n=30)

| Studies | Setting/Country | Study design | Data collection /analysis methods | Population | Description of Intervention | Main findings |
| --- | --- | --- | --- | --- | --- | --- |
| Stumbar 2022 | University/USA | Mixed methods design | Open-ended survey/Content analysis | Second-year medical students<br>Sex: NR<br>Age: NR<br>Sample size: 128 | A 1-hour lecture on the components of an inclusive sexual history, followed by a 1.5-hour faculty-facilitated small group session with approximately eight students | Two themes were identified: (1) Students' favorite part of the session; (2) Suggested areas for improvement. |
| Mackie 2022 | Secondary schools/Australia | Descriptive qualitative design | Individual semi- structured interview/Interpretative phenomenological analysis | Registered psychologists or counselors practicing<br>Sex: 86% female<br>Age: 38.9 (28-55)<br>Sample size: 7 | NR | Three superordinate themes and nine subordinate themes were identified: (1) Practicality and Utility of Training; (2) Availability of training LGBTQ+ curriculum within tertiary education; (3) Addressing the needs of transgender young people. |
| Kreines 2022 | University/USA | Mixed methods design | Open-ended survey/ Thematic analysis | Doctor: Obstetrics and gynecology residents<br>Sex: Male 4, Female 26<br>Age: 26-36<br>Sample size: 30 | The simulation curriculum consisted of a 45-min recorded lecture, a 15-min observed simulated encounter with a gender diverse standardized patient (SP), 5 minutes of individualized feedback from the SP and observing faculty member, and a 1 hour group debrief with all learners, SPs, and faculty members. | Four themes were identified: (1) What was learned; (2) What was most challenging; (3) What was most surprising; (4) Biggest takeaways/changes to practice. |
| Treharne 2021 | University/ New Zealand | Exploratory qualitative | Focus groups/ Inductive analysis | Transgender community members, teaching staff | NR | Four themes were identified: (1) Feelings of lacking the expertise to teach about |
|  |  | design |  | from health professional programs, and trainee health professionals<br>Sex: 63.6% female<br>Age: median 30<br>Sample size: 33 |  | transgender healthcare; (2) Experts on methods of teaching about healthcare; (3) Experts from their own experiences of healthcare; (4) Awareness of limits to teaching from personal experiences and of diversity among transgender people. |
| Jeon 2022 | University/South Korea | Descriptive qualitative design | Individual interviews/Generic content analysis | Fourth-year medical students<br>Sex: 50.0% female<br>Age: NR<br>Sample size: 8 | A 4-hour program spread over a period of 3 weeks:<br>(1) The first hour of class was an offline lecture covering basic concepts such as gender, the diagnostic criteria for gender dysphoria, and changes in the diagnostic criteria for gender dysphoria in the Diagnostic and Statistical Manual of Mental Disorders (DSM).<br>(2) The second hour of class featured an online lecture about gender classification, general healthcare for transgender patients, medical transition, and gender-friendly communication methods.<br>(3) The third and fourth hours of the class consisted of clinical role-play. | Three themes were identified:<br>(1) The internal responses to transgender healthcare educational program: confusion, acceptance, and negotiation.<br>(2) Factors related to internal responses of transgender healthcare education: personal identities and professional identities.<br>(3) The improvement of cultural capacities after taking the transgender healthcare educational program. |
| Hayward 2022 | University/ New Zealand | Realist qualitative design | Focus groups/ Generic content analysis | Clinical psychology students | A program where the first year involves lecture-based learning and the latter 2 years involve weekly seminars and closely | Three themes were identified: (1) Wanting to support transgender clients but needing more knowledge; (2) New perspectives on |
|  |  |  |  | Sex: 75.0% female<br>Age: 24.5 (22-27)<br>Sample size: 8 | supervised practice with real clients (at a training clinic in the second year, and on a community placement in the final year). | transgender health care from the transgender ambassadors; (3) Opportunities and challenges in the clinical psychology program. |
| Altmiller 2023 | University/USA | Pre-/post-test non-equivalent groups design, including qualitative and quantitative evaluation | Open-ended survey/<br>Thematic analysis | Nursing students<br>Sex: NR<br>Age: NR<br>Sample size: 126 | Virtual patient simulation uses a web-based platform that allows nursing students to interact with patients in specific contexts that support practicing and applying nursing skills such as collecting subjective and objective data, documenting data in electronic health records, choosing appropriate interventions, and evaluating outcomes of care. | Five themes were identified: (1) Related to limited knowledge; (2) Needing and wanting this education emerged; (3) Judgment of transgender persons as unfair; (4) Supporting that the questions of the TABS promoted reflection; (5) The value of this simulation experience. |
| Huser 2022 | Community health centre /USA | Mixed methods design | Open-ended survey/<br>Content analysis | Psychologist/counsellor:<br>Board-eligible or certified genetic counselling and student<br>Sex: Male 1, Female 39<br>Age: 18-64 years<br>Sample size: 40 | 1. A 5-module genetic counselling-targeted online learning program<br>(1) Terminology, Population Disparities, and Communication: A comprehensive glossary, breakdown of transgender and/or gender non-binary (TGNB) population disparities, as well as gender-affirming communication considerations for the genetic counsellor.<br>(2) Clinical Environment: An examination of the clinical environment from the patient perspective, from start to finish, including things such as in-take forms, the physical environment, and documentation.<br>(3) Potential Aspects of the Gender Affirmation | (1) The findings from the Slack threads revealed that the most participant activity was in the 'Overcoming Stereotypes and Biases' thread, followed by the 'Clinical Environment' thread. (2) Key themes discussed included the influence of personal connections with the transgender and non-binary community on clinical biases, the integration of gender-affirming communication strategies in counseling, and efforts to create a more gender-inclusive environment in genetic counseling clinics. In contrast, the 'Cancer Case Example' thread received no engagement from participants. The data also indicated that |
|  |  |  |  |  | Process: A summary of potential psychological, social, hormonal, surgical, and legal affirmations individuals make undergo in their gender affirmation process.<br>(4) Cancer Risk Assessment: An organ-based perspective summarizing what is known up to this point in the literature on cancer risks of TGNB individuals and how to consider possible hormonal or surgical affirmations into risk estimates and management.<br>(5) Pulling it All Together- The Genetic Counseling Session: Applying everything in Modules 1–4 to core components of the genetic counseling session such as family and medical history gathering, education, psychosocial considerations, and patient resources.<br>2. Participants were provided the option of joining a private online discussion forum hosted on Slack, a business communication platform. | participants were more likely to engage in discussions that did not require direct posts, such as responding to anonymous quizzes or polls in the 'General Discussion' thread. |
| Biro, Tang 2021 | University/<br>Canada | Realist and<br>constructivist<br>qualitative<br>design | Focus groups/ Content<br>analysis | First-year medical students<br>Sex: 74% female<br>Age: NR<br>Sample size: 35 | NR | Three themes were identified: (1) content (concepts, 'chunking' topics, future clinical reference, application to practice); (2) presentation (graphics, accessibility, learning tool); and (3) reflection. |
| Frazier 2021 | University/USA | Descriptive qualitative design | Open-ended survey/<br>Content analysis | Student pharmacists<br>Sex: NR<br>Age: NR<br>Sample size: 51 | The pre-class materials for this course included a six-page FAQ document about transgender people and a 40-minute lecture on related terminology, disparities, and treatment guidelines. The three-hour class was split into a two-hour interactive session, involving a game show-style activity, gender identity exploration, discussion of a current event article, and role-play, and a one-hour discussion with a transgender man, who was a speaker with over a decade of personal experience. This interactive approach, including direct interaction with a transgender individual, is vital in helping health professional students form more positive views of marginalized patient groups. | Q1. What are you hoping to learn about providing good quality care to people who are transgender? Themes: Communication skills, terminology/vocabulary preferred by people who are transgender, population-specific challenges faced by transgender patients, counseling points, medications for transgender patients, suggestions/additions for the transgender guest speaker, additional information about the transition and being transgender.<br>Q2. What was missing from this session that you would have liked to learn about transgender or gender diverse patients? Themes: Local resources, additional information on pharmacotherapy, more activities of this nature in general |
| Biro, Song 2021 | University/<br>Canada | Constructivist qualitative design | Focus groups/ Content analysis | First year medical students<br>Sex: 74% female<br>Age: NR<br>Sample size: 35 | The seminars included an infographic, a communication guide, and a sexual history-taking video to teach the basics of inclusive communication. During the four-hour seminar, students answered discussion questions, participated in peer role-plays, and interviewed two standardized patients. | Two themes were developed: (1) SGM bias (faculty, standardized patients, students, curriculum); and (2) Adaptive Expertise in Clinical Skills (case complexity, learner support, skill development). |
| Arias 2021 | University/UK | Descriptive | Focus groups/ Thematic | Student midwives, | NR | There were two themes and four subthemes: |
|  |  | qualitative design | analysis using Braun and Clarke's approach | midwifery educationalists, and midwifery clinicians<br>Sex: NR<br>Age: NR<br>Sample size: 16 |  | (1) Practising open mindedness with sub themes: Making assumptions and developing self- awareness; (2) Cultivating openness with sub themes: challenge and discomfort and safe spaces. |
| Vijapura 2023 | University/USA | Descriptive qualitative design | Face-to-face semi-structured one-on one interviews/ Thematic analysis with an inductive approach | Residents, ranging from their first to fourth year in radiology training, and in postgraduate years 2-5<br>Sex: NR<br>Age: NR<br>Sample size: 10 | The University of Cincinnati radiology residents participated in four monthly sessions that comprehensively covered transgender patient care using a radiology-based approach. Each session, fitting into the regular scheduled noon conference time, lasted approximately 1 hour. The topics covered included: (1) Pronouns and healthcare disparities; (2) Breast/chest imaging of transgender patients and inclusivity in the clinical setting; (3) Body imaging of transgender patients and gender-affirming surgeries; (4) Patient communication and cultural sensitivity in the radiology department. The first half of the initial three sessions was primarily didactic, allowing time for open discussion and questions. The second session incorporated an interactive component with small group case scenarios and discussions. | Four themes emerged through the existing framework: (1) impactful/memorable aspects; (2) things learned; (3) increased awareness; and (4) suggestions/feedback. Prominent subthemes included patient panel and stories, physician experts sharing knowledge and experiences, link to radiology and imaging, novel concept, gender-affirming surgeries and anatomy, appropriate radiology reporting, and patient interactions. |
| McMillian-Bohler 2022 | University/USA | Mixed methods design | Qualitative responses to the survey and comments in the Zoom chat/ Content analysis | Nursing students<br>Sex: NR<br>Age: 20-30 years<br>Sample size: 39 | The class and seminar were held virtually via Zoom because of coronavirus disease 2019 (COVID-19) restrictions. The length of the class session was 2 hours; however, the seminar was held during the last 90 minutes of the class. Before the presentation, students and faculty reviewed the expectations for the discussion and learning objectives. The speaker then shared details of the pre-conception, antepartum, intrapartum, and postpartum care he received. Students were invited to ask additional questions or share their thoughts via the chat feature in Zoom. | The three themes that emerged from the qualitative data were: (1) empathy; (2) advocacy; and (3) professional accountability. |
| Luctkar-Flude 2022 | University/Canada | Mixed methods design | Open-ended survey/ Content analysis | 1 nursing faculty (lecturer), 1 clinical instructor and 1 Master of Nursing student; and 18 undergraduate nursing students<br>Sex: NR<br>Age: NR<br>Sample size: 21 | The study intervention consisted of (a) online presimulation preparation materials; (b) 4 online virtual simulation games; and (c) debriefing; delivered over the course of one week. | Six themes related to the benefits and challenges of the virtual simulation games and eLearning module. |
| Luctkar-Flude 2021 | University/Canada | Mixed methods design | Open-ended survey/ Thematic analysis | Nursing students and nursing educators | This toolbox included two voice-over PowerPoint presentations as pre-simulation | Seven themes emerged from the analysis of qualitative feedback: usability, engagement, |
|  |  |  |  | Sex: 91% female<br>Age: 27 (20~24)<br>Sample size: 11 | preparation activities: one focused on the concept of cultural humility, and the other on terminology related to sexual orientation and cultural humility. The toolkit also included four innovative bilingual Virtual Simulation Games (VSGs) that depict clinical encounters between nurses and LGBTQITS individuals. These VSGs represent sexual and gender diverse populations, including the care of (1) an older gay man experiencing significant loss and grief; (2) a transgender youth facing issues of sexual safety, pregnancy, and suicide risk; (3) a transgender man who experiences repeated misnaming; and (4) a lesbian woman who requires a sexual health assessment. | value, self-reflection, authenticity and diversity, challenges, and suggestions for change. |
| Koch 2021 | University/USA | Mixed methods design | Open-ended survey/NR | Nursing students<br>Sex: NR<br>Age: NR<br>Sample size: 26 | The role-play simulation consisted of two parts, both playing out the same scene. The first part involved a written script with the transgender patient, their friend, an admitting nurse, and a second nurse, depicting how interactions might unfold without cultural sensitivity, potentially leading to microaggressions and a lack of inclusivity. After completing the first scene, the second part of the role-play simulation began, similar | Five themes emerged from an analysis of the survey results: 1) Nursing students were not aware of the long history of discrimination experienced by people who are transgender; 2) When patients do not have the support of family of origin then they may have close friends or members of the community assume this important role; 3) Nursing students needed more knowledge and experience on how to ask patient pronouns when working with patients |
|  |  |  |  |  | to the first, but with interactions characterized by greater cultural sensitivity and inclusiveness. | 4) Nursing students expressed insecurity and fear about making mistakes when talking to patients about sexual orientation or gender identity; and 5) Nursing students were unaware that hospital policies may not be inclusive of patients who are transgender. |
| Craig 2021 | Hospitals, community agencies/USA | Mixed methods design | Open-ended survey/<br>Thematic analysis | Clinicians<br>Sex: 64 participants' information didn't report.<br>Among the 65 participants, 44 females.<br>Age: 22-68<br>Sample size: 129 | Informed by the NIH BCC's best practices for facilitator training, the AFFIRM training is a standardized two-day, 14 h training workshop that is informed by experiential learning theory and integrates didactic and experiential learning (e.g., role play, problem-based learning). Eight AFFIRM CBT Facilitator Trainings were delivered between April 2017 and April 2019. | Two major themes: (1) Strengthened ability of facilitators to affirm LGBTQ + identities utilizing CBT ("Learning how to better support LGBTQ+ populations"; "understanding the impact of negative self-talk for LGBTQ+ communities"; "learning about how to do affirmative practice"); and (2) Increased competence to deliver AFFIRM to SGMY ("Learning about clinical approaches to affirming therapies"; "dyads and triads to do activities"; "role play with others, structure and content deepened my skills"; "the experiential practice helped my learning"). |
| Bi 2020 | University/USA | Mixed methods design | Open-ended survey/<br>Thematic analysis with an inductive approach | The first-year medical students<br>Sex: NR<br>Age: NR<br>Sample size: 83 | In this study, teaching module on intersectionality of sexual orientation, gender identity, and race/ethnicity included a 30-minute didactic lecture and a 1.5-hour facilitated discussion portion of the session. Students viewed four video interviews of sexual and gender minority patients of color describing their lives and health care narratives, each followed by a moderated discussion about how compounded minority stress affects lived experiences and health, how multiple identities affect patient-provider communication, and how to improve shared decision making for sexual and gender minority and patients of color. Following the facilitated discussion, one of the video interviewees, who had experience training clinicians in sexual and gender minority issues, participated in a 30-minute question-and-answer session with the students. | Three themes were identified: (1) Understanding of intersectionality; (2) Strengths of the session; (3) Suggestions for improvement of the session. |
| Charbonneau 2022 | University/USA | Descriptive qualitative design | Online focus groups/<br>Thematic analysis | Sophomore prelicensure students<br>Sex: NR<br>Age: 18~50<br>Sample size: 32 | NR | Four themes were identified through the coding process. The main themes are as follows: (1) exposure to transgender communities in healthcare; (2) knowledge about transgender healthcare and patient assessment; (3) understanding about transgender communities; and (4) experience with transgender patient care during clinical and simulation settings. |
| Jadwin-Cakmak 2020 | Clinics/USA | Mixed methods design | In-depth interviews/<br>Content analysis | Providers and staffs of clinics | The Health Access Initiative (HAI) consists of a 1-hour online training and a 2-hour in-person | Interviews with site liaisons from 10 locations (11 interviews total) revealed that the technical |
|  |  |  |  | Sex: NR<br>Age: NR<br>Sample size: 11 | training, followed by 3 months of site-specific technical assistance (TA) to improve clinic- and structural-level issues related to LGBTQ+ care. The online training (which can be accessed free of charge at mfierce.org) focuses on LGBTQ+ terminology and cultural humility, featuring testimonials from Youth Advisory Board (YAB) members and healthcare providers. The in-person training includes didactic learning methods and group activities. These activities focus on challenging biases and stereotypes, considerations for providing patient-centered care, cultural humility practices, taking a comprehensive and affirming sexual health history, patient-centered STI prevention counseling, and best practices for referrals and resource provision with sexual and gender minority youth (SGMY). | assistance (TA) led to changes in language, practices, and increased staff awareness about supporting sexual and gender minority (SGM) people. This included enhanced access to educational materials and connections with Southeastern Michigan organizations. Eight of the ten sites modified their electronic medical records or intake forms to better collect patients' gender identity, pronouns, preferred name, and sexual orientation. All sites added more SGM-inclusive posters, provided along with other resources. A noted challenge was balancing the desire for longer, more in-depth training with the practical limit of 2 hours for clinic training time. Staff were generally satisfied with the TA and expressed interest in further training on LGBTQ+ topics and a refresher course. |
| Du 2020 | Treatment centres/ Canada | Mixed methods design | Open-ended survey/<br>Content analysis | Forensic nurses<br>Sex: 100% female<br>Age: >19<br>Sample size: 47 | The curriculum was rolled out on November 21, 2018 in Toronto, Ontario during the week-long in-person SANE Training for frontline nursing staff. On the first day of SANE Training, as part of the orientation to the week, the study was explained to all nurses<br>(including that participation was optional, anonymous, and would not affect their relationship with the Ontario Network of SA/DVTCs or its leadership) and consent forms to participate in the pre- and post-training questionnaires were distributed. On the fourth day of SANE training, the training module was delivered by a member of the trans community who had also aided in the overall development of the curriculum. To encourage interaction between the trainer and participants and facilitate interactive learning, following two lessons, case studies were presented with discussion questions. Participants were asked to spend a few minutes reflecting on each case study and discussing the associated questions with fellow participants at their tables. The trainer then invited one person at each table to share their table's responses with the full group. The total training time was 1.75 h. | Three themes were identified: (1) Satisfaction with training module; (2) Most helpful aspects of training module; (3) Suggestions to improve the training module. |
| Chaudhary 2024 | Online/ Australia | Mixed methods design | Open-ended survey/<br>Content analysis | Pharmacists and pharmacy students<br>Sex: 68.83% female | The training program is designed to educate pharmacists and pharmacy staff on interacting with transgender and gender diverse (TGD) individuals. It includes reading materials, | Eight themes were identified from pharmacists: (1) Comprehensive, easy to understand, convenient; (2) Informative and innovative; (3) Knowledge and confidence for practice; (4) |
|  |  |  |  | Age: NR<br>Sample size: 77 | resource links, and pre-recorded videos featuring TGD people discussing their experiences with pharmacists and their expectations for healthcare. The program also features videos demonstrating both inappropriate and appropriate ways to interact with TGD people in a pharmacy setting. These scenarios cover counseling on over-the-counter and hormonal medicines used for gender affirmation and feature local TGD volunteers and pharmacists. | Language, interactions and Pharmacotherapy for Gender Affirmation; (5) Relevant to practice; (6) Case studies; (7) Inclusion of TGD people; (8) References and resources, all information in one place.<br>Five themes were identified from interviews: (1) Awareness knowledge; (2) Comfort privacy; (3) Change – personal and societal behaviour in pharmacy; (4) Complete resource requirement for printed or accessible training material; (5) Future training availability for broader consumption.<br>Six themes were identified from pharmacist: (1) Comprehensive, easy to understand, convenient; (2) Informative and innovative; (3) Language, appropriate interactions and pharmacotherapy for gender affirmation; (4) Relevant to practice; (5) Safe space to ask questions; (6) Videos. |
| Ness 2023 | University/USA | Pre-/post-test non-equivalent groups design, including | Debriefing/Thematic analysis | Nursing students<br>Sex: 90% female<br>Age: NR | The scenario involved a simulated outpatient surgical setting where a patient had undergone gender-affirming surgery. Students were tasked with delivering evidence-based postoperative | Four themes were identified: (1) unique learning experience; (2) Psychological safety; (3) Gender-affirming language; (4) Desire for integration into the nursing curricula. |
|  |  | qualitative and quantitative evaluation |  | Sample size: 10 | nursing care and using gender-affirming language during postoperative education with the patient and their significant other. The realism of the simulation was enhanced by having two gender-diverse Standardized Patients act out the roles of the patient and their loved one. |  |
| Linsenmeyer 2023 | University/USA | A basic qualitative design (the approach name was not specified) | Online reflection and discussion/ Constant comparative method | Graduate students<br>Sex: NR<br>Age: NR<br>Sample size: 263 | The ITHED was a comprehensive one-day, in-person training program focused on transgender health education. It featured lectures by transgender educators on basic transgender health and gender-affirming medical care, a panel discussion with community members, and a session for guided reflection. Additionally, students could choose two breakout sessions to attend, which covered various topical issues in transgender health, clinical skill development, or discussions with faith leaders in smaller groups. | Three major themes resulted: (1) The power to help or harm, (2) The responsibility to provide health care, and (3) A posture of humility: listen and learn |
| Ruud 2021 | Clinics/USA | Mixed methods design | Open-ended survey/NR | 12 midwifery students, 16 women's health nurse practitioner students and 5 obstetrics and gynecology residents | (1) recruitment of members of the transgender community to serve as patient-teachers, (2) creation of scenarios and selection of materials to be used in the health history simulation, and (3) participation of learners in the health history simulation with a patient-teacher. | Participants were positive about the experience and felt they were able to provide a safe space and help the students learn. |
|  |  |  |  | Sex: NR<br>Age: NR<br>Sample size: 33 |  |  |
| Lacombe-Duncan 2021 | Hospitals, community, and online/USA | A non-randomized multi-site pilot study with pre- and post-test design | Pre- and post-intervention questionnaires and open-ended feedback/<br>Content analysis | Healthcare and social service providers<br>Sex: NR<br>Age: 37.0 (26-48)<br>Sample size: 78 | The program was developed by team members with experience in community-based transgender education across Montreal, Toronto, and Vancouver. The program adapted existing "Trans 101" trainings and utilized elements from Card's model and the ADAPT-ITT framework for adapting HIV interventions. TEACHH is based on the Information-Motivation-Behavioral Skills Model (IMB), which has been used in LGBT cultural competency interventions. It integrates how knowledge, motivation, and skills can influence provider behavior. The development of TEACHH was informed by feedback from focus groups involving 26 trans women and 10 service providers. The program aims to enhance providers' knowledge, perceived competency, and attitudes towards providing gender-affirming HIV care to trans women. TEACHH sessions, led by trans women, lasted three hours each. | Participants described gender-affirming language, clarifications and explanations of new terms and concepts, the introduction of new theories and approaches, resources on health and rights of trans women living with HIV, and how and where to access treatment, care, and support as most beneficial. |
| Willson 2024 | University/USA | Mixed methods | Open-ended survey/ | Student pharmacists | The educational module, which included | The study demonstrated improved student |
|  |  | design | Thematic analysis | Sex: 75 female<br>Age: NR<br>Sample size: 106 | activities and mini-lectures, was delivered in a laboratory setting. Immediately after the lab, students were emailed a link to a postsurvey, which remained accessible for one week. The total time required for students to complete the module and assessments was approximately 3.5 hours. | knowledge, confidence, and comfort, use of gender inclusive language after the MGIC module. To identify and address microaggressions, bias must be explored. Both implicit and explicit biases affect behaviors or actions. Microaggressions can be common and lead to health disparities for both marginalized providers and patients. The module was effective, well received, and will continue to be taught in the course. |
| Corey 2023 | University/USA | Mixed-methods design | Open ended questions+ individual interview by Zoom using a semi-structured interview guide/NR | Nursing students and physician assistant students<br>Sex: NR<br>Age: NR<br>Sample size: 21 | Participants completed the online Transgender Health Learning Series, which contained 7 courses: Transgender Health 101: A Foundation Course on Transgender People and Public Health; Transgender Health 102: Transgender Patient-Provider Relationship; Transgender Health 103: LGBTQ+ Healthcare Workplace Inclusion; Transgender Health 104: Psychological Distress; Transgender Health 105: Role of Endocrinology in the Transgender Community; Transgender Health 106: Pre and Post-operative Care; Transgender Health 107: Transgender Health 107: Not About Us, Without Us: The Importance of Transgender Patient-Centered Outcomes. | (1) Limited Prior Training/Call to Include Gender Affirming Care in Curriculum; (2) Evaluation of Training, Key Takeaways and Suggestions for Improvement; (3) Fear of Saying the Wrong Thing/Misgendering with Qualifying Statements of Confidence. |
| Karsenti 2023 | University/<br>Canada | Mixed-methods<br>design | Open-ended survey<br>questions/Content analysis | Medical Students and<br>faculty<br>Sex: NR<br>Age: NR<br>Sample size: 9 students<br>and 6 faculty | NR | (1) Discrimination; (2) Microaggressions; (3)<br>Stereotyping within the curriculum; (4) Paucity<br>of faculty knowledge. |

Notes: NR=not reported.

## Appendix 4: Risk of bias assessment

### Section 1. Quantitative review: Risk of Bias summary table for RCTs

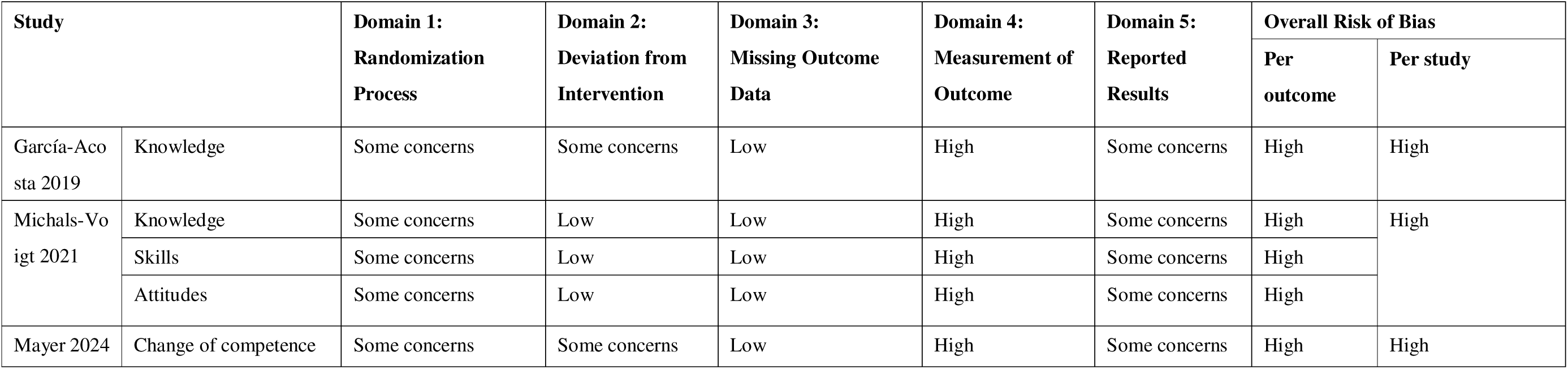

| Study |  | Domain 1:<br>Randomization<br>Process | Domain 2:<br>Deviation from<br>Intervention | Domain 3:<br>Missing Outcome<br>Data | Domain 4:<br>Measurement of<br>Outcome | Domain 5:<br>Reported<br>Results | Overall Risk of Bias |  |
| --- | --- | --- | --- | --- | --- | --- | --- | --- |
|  |  |  |  |  |  |  | Per<br>outcome | Per study |
| García-Acosta 2019 | Knowledge | Some concerns | Some concerns | Low | High | Some concerns | High | High |
| Michals-Voigt 2021 | Knowledge | Some concerns | Low | Low | High | Some concerns | High | High |
|  | Skills | Some concerns | Low | Low | High | Some concerns | High |  |
|  | Attitudes | Some concerns | Low | Low | High | Some concerns | High |  |
| Mayer 2024 | Change of competence | Some concerns | Some concerns | Low | High | Some concerns | High | High |

### Section 2. Quantitative review: Risk of Bias summary table for non-randomized comparative studies*

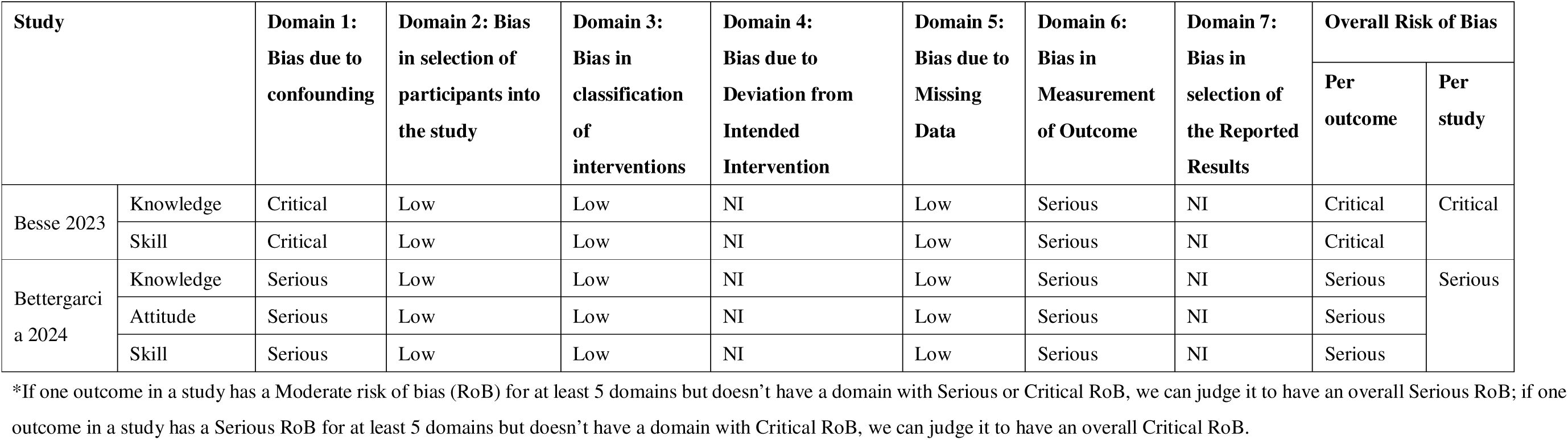

### Section 3. Quantitative review: Risk of Bias summary table for before-and-after studies*

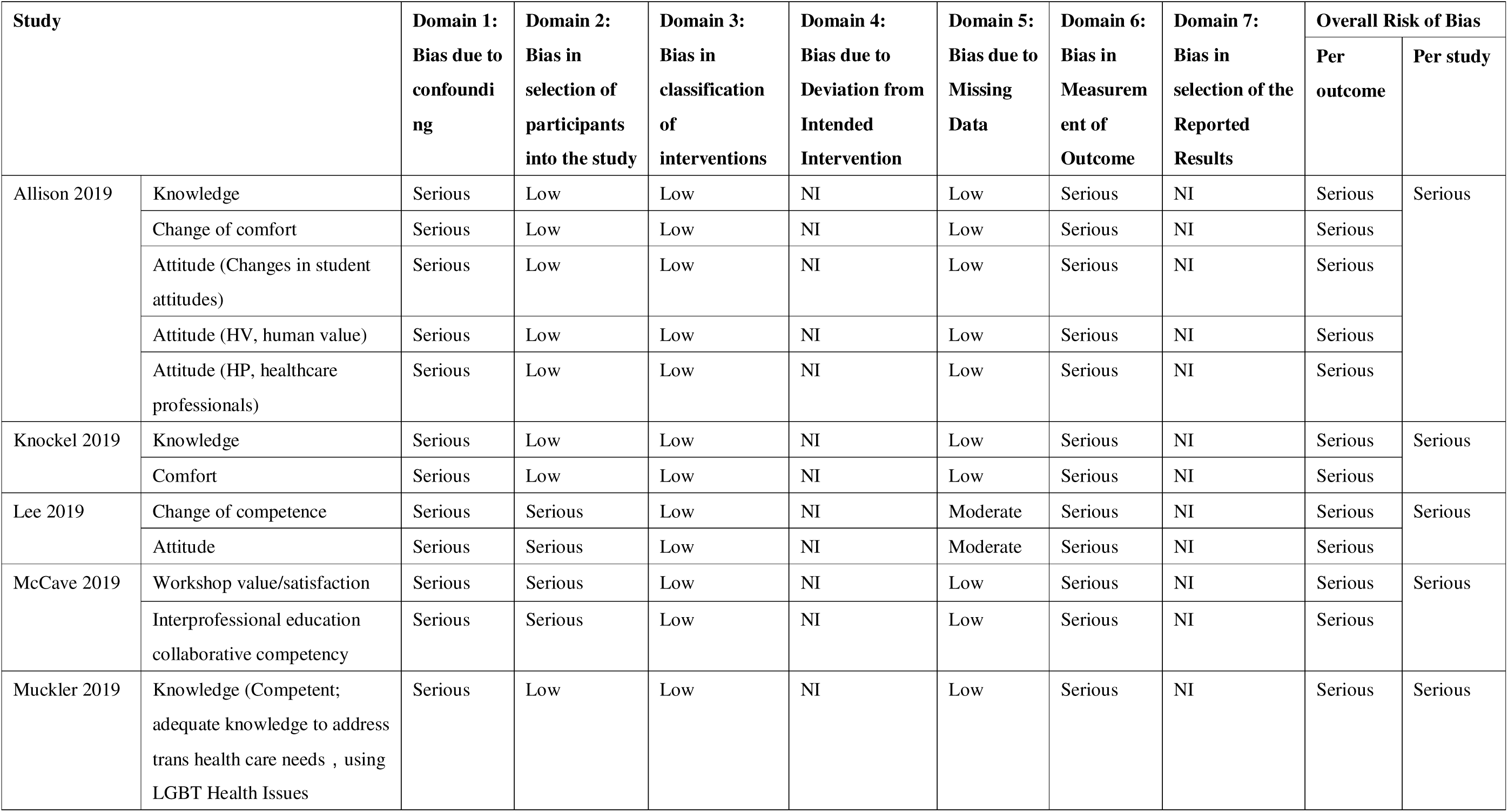
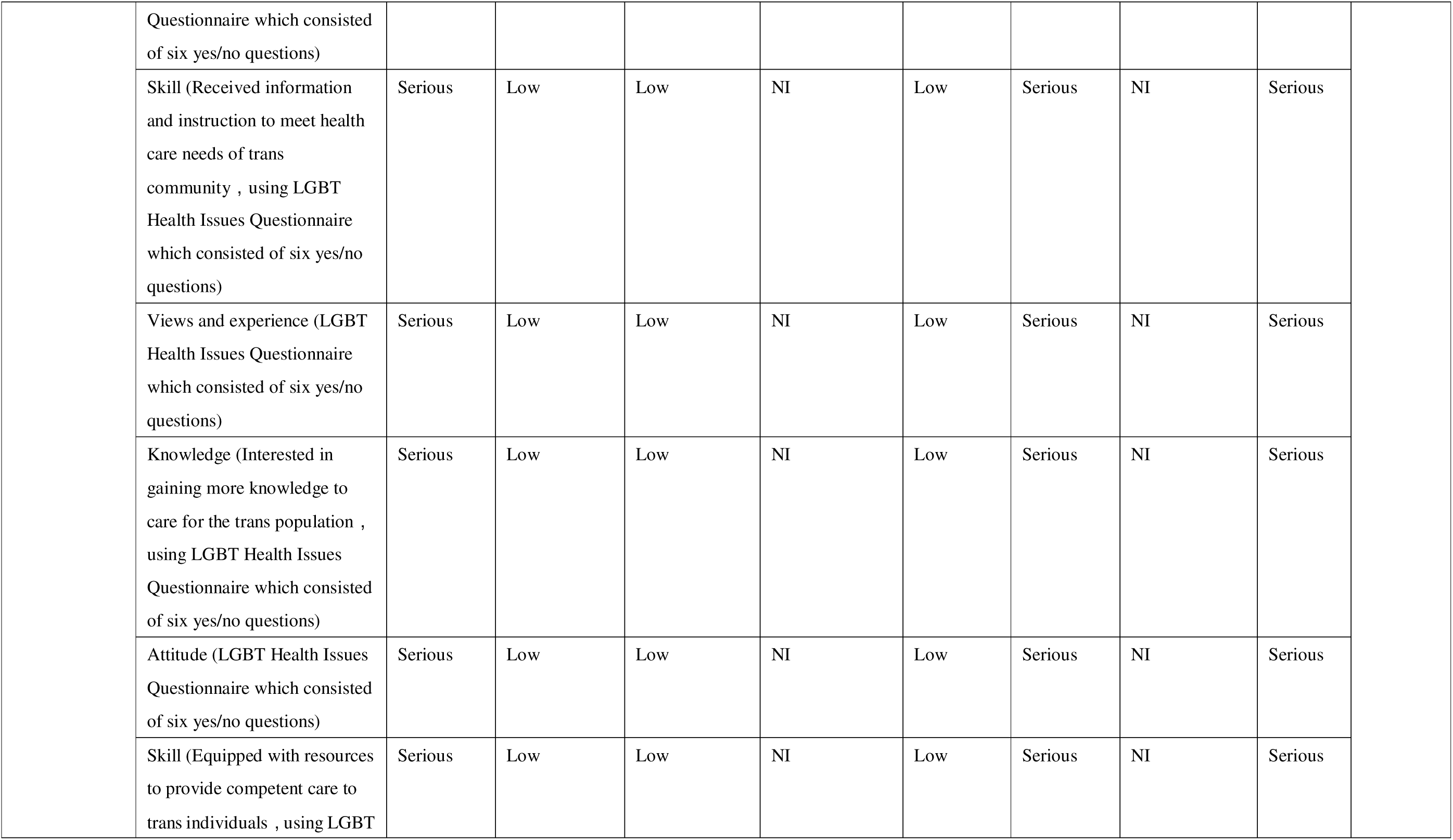
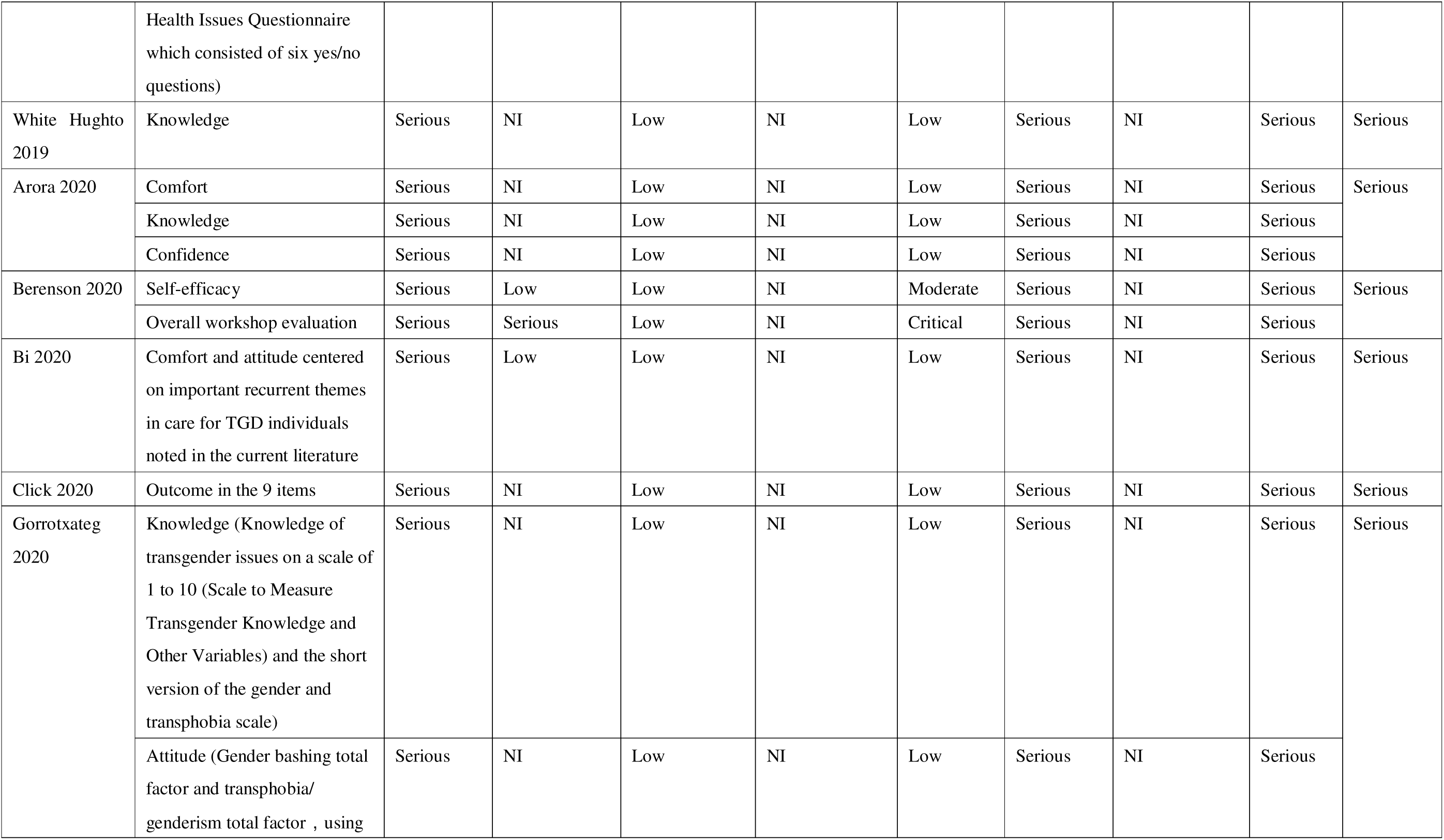
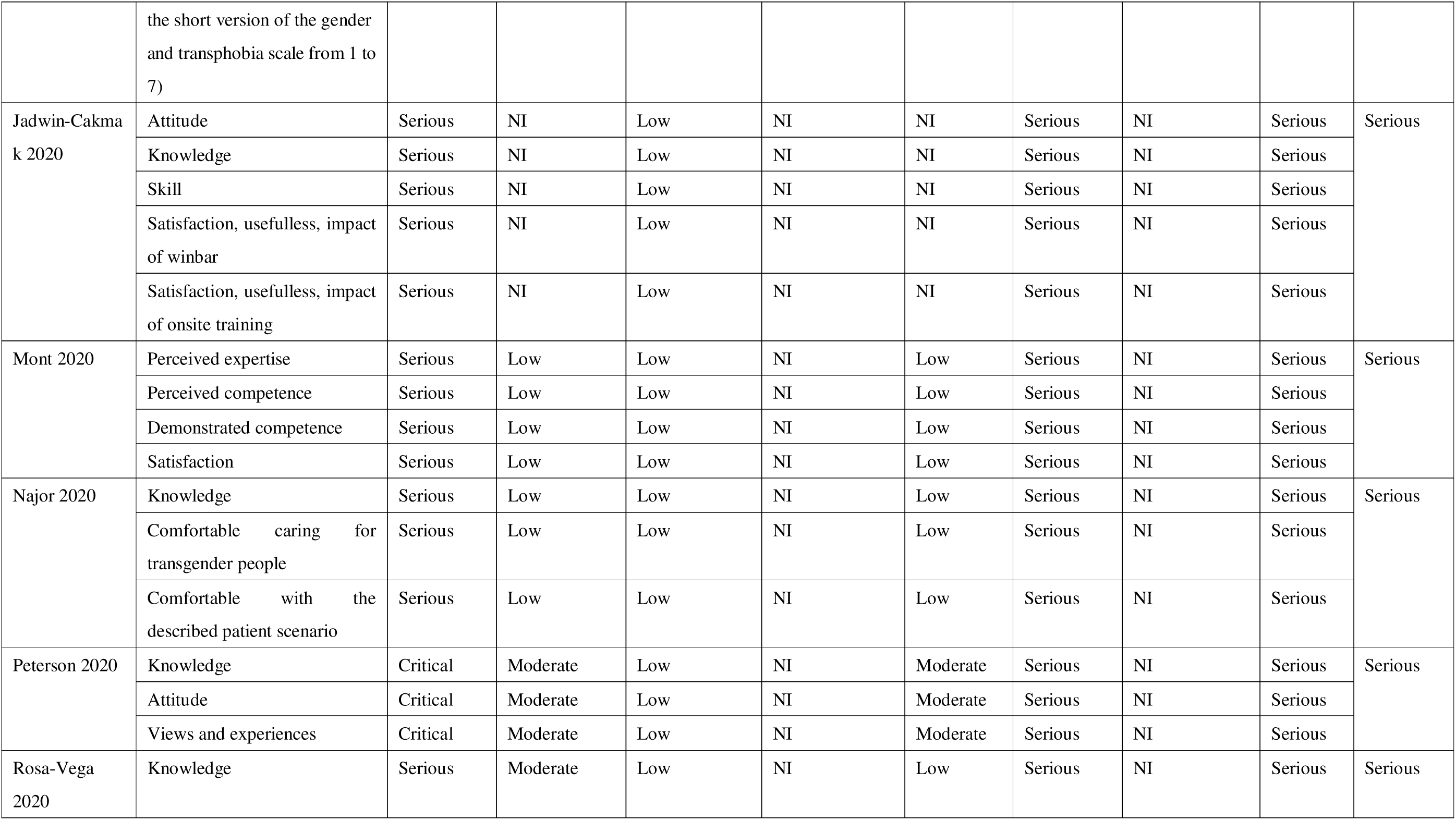
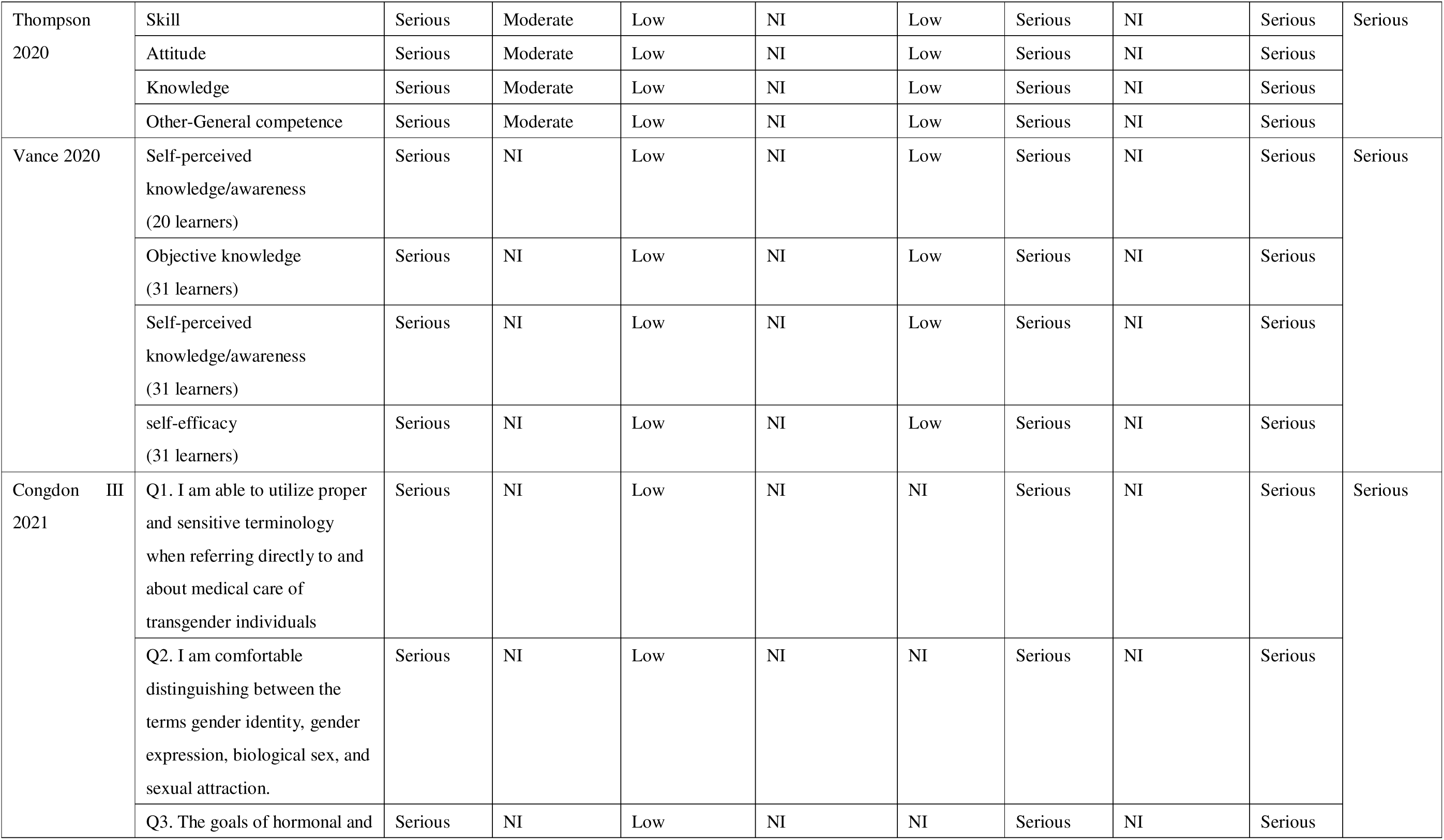
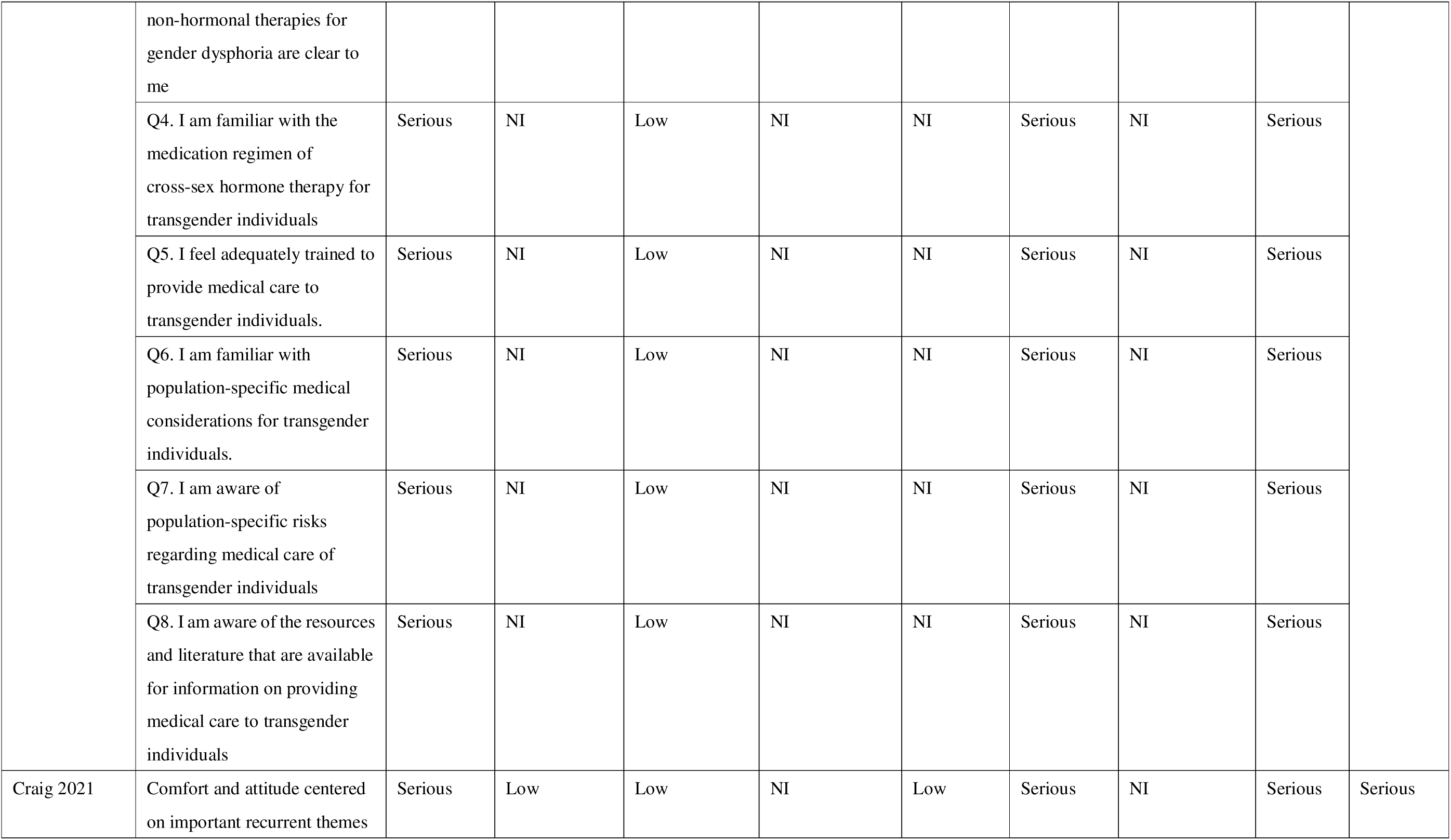
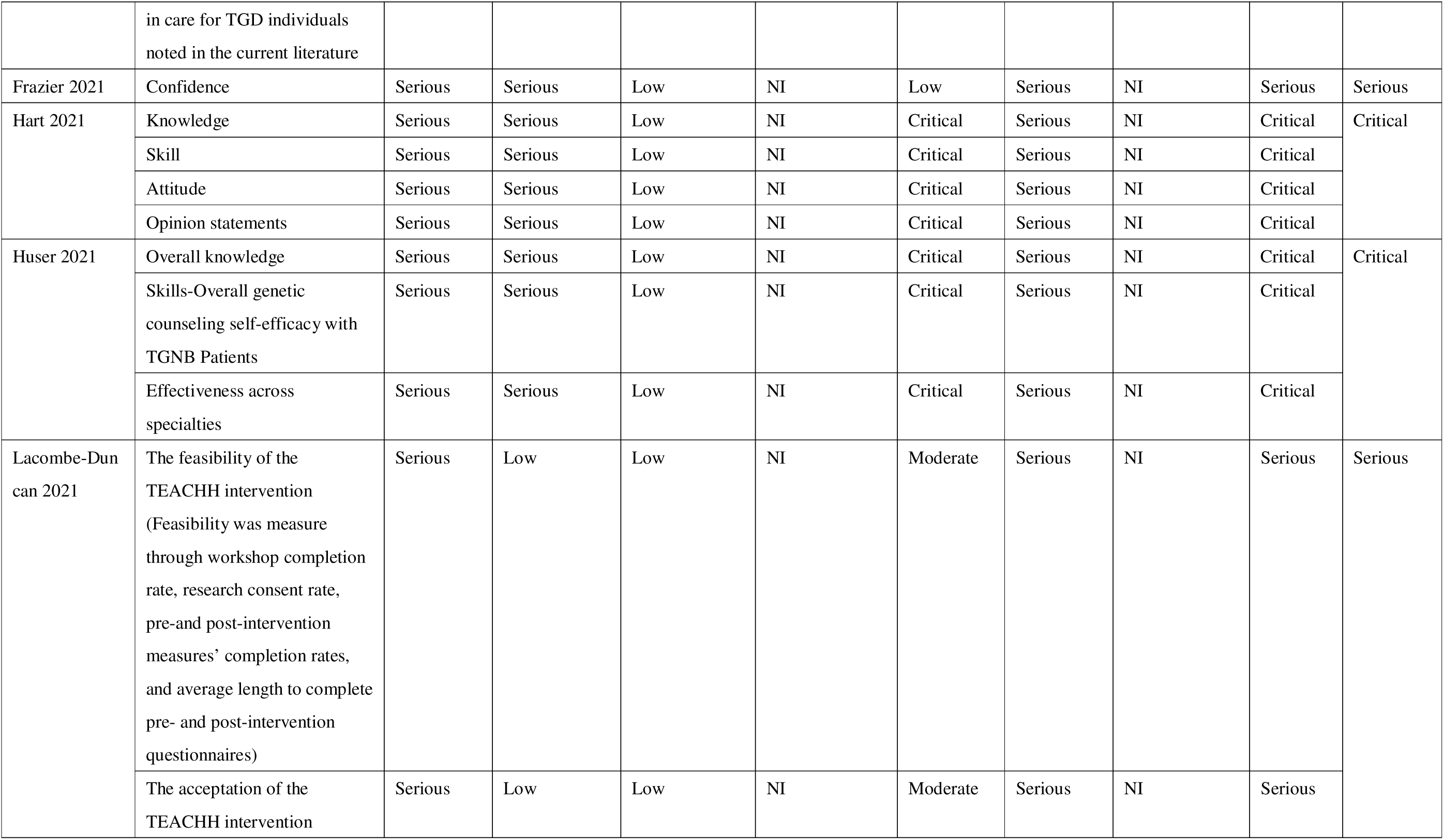
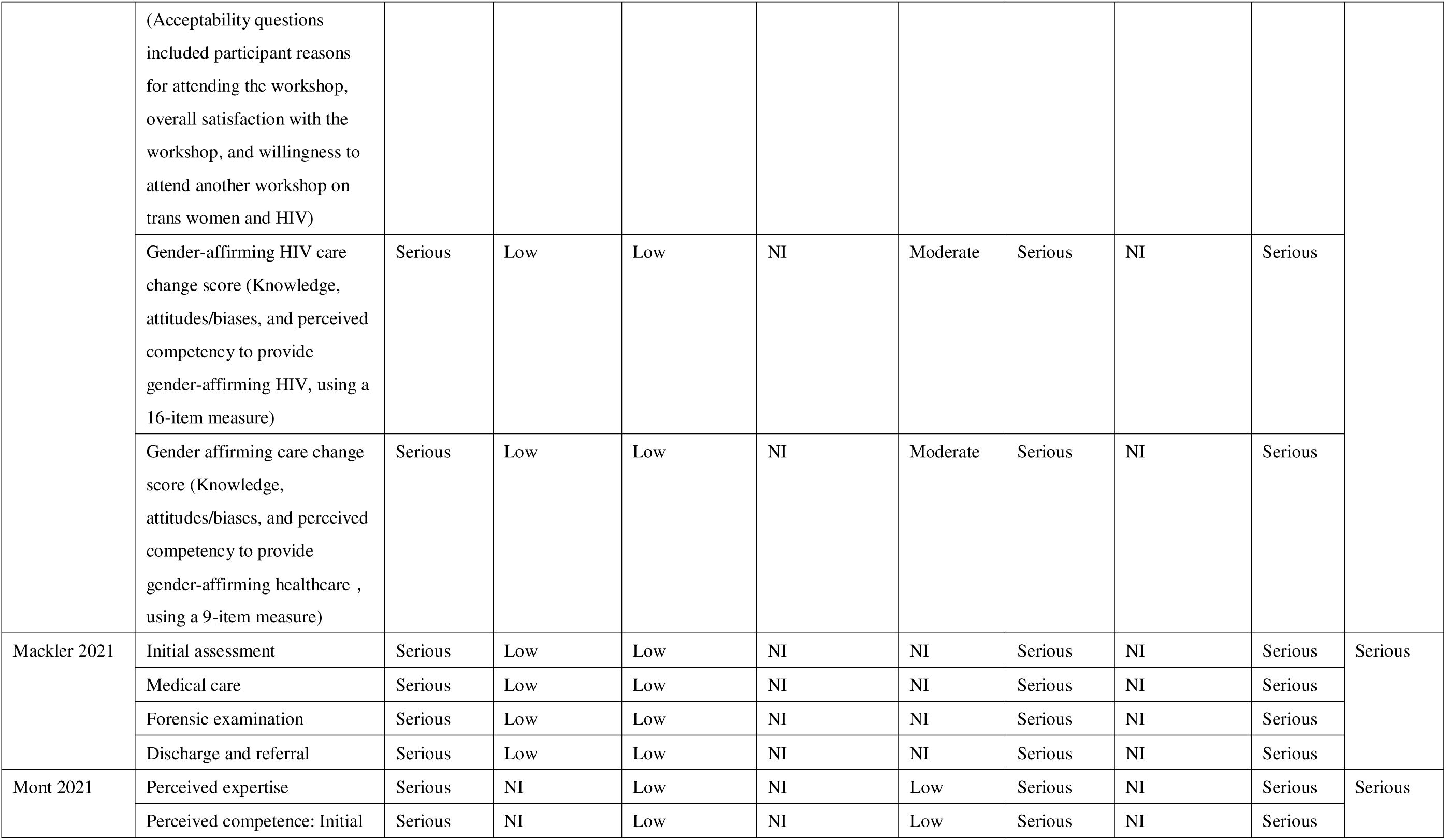
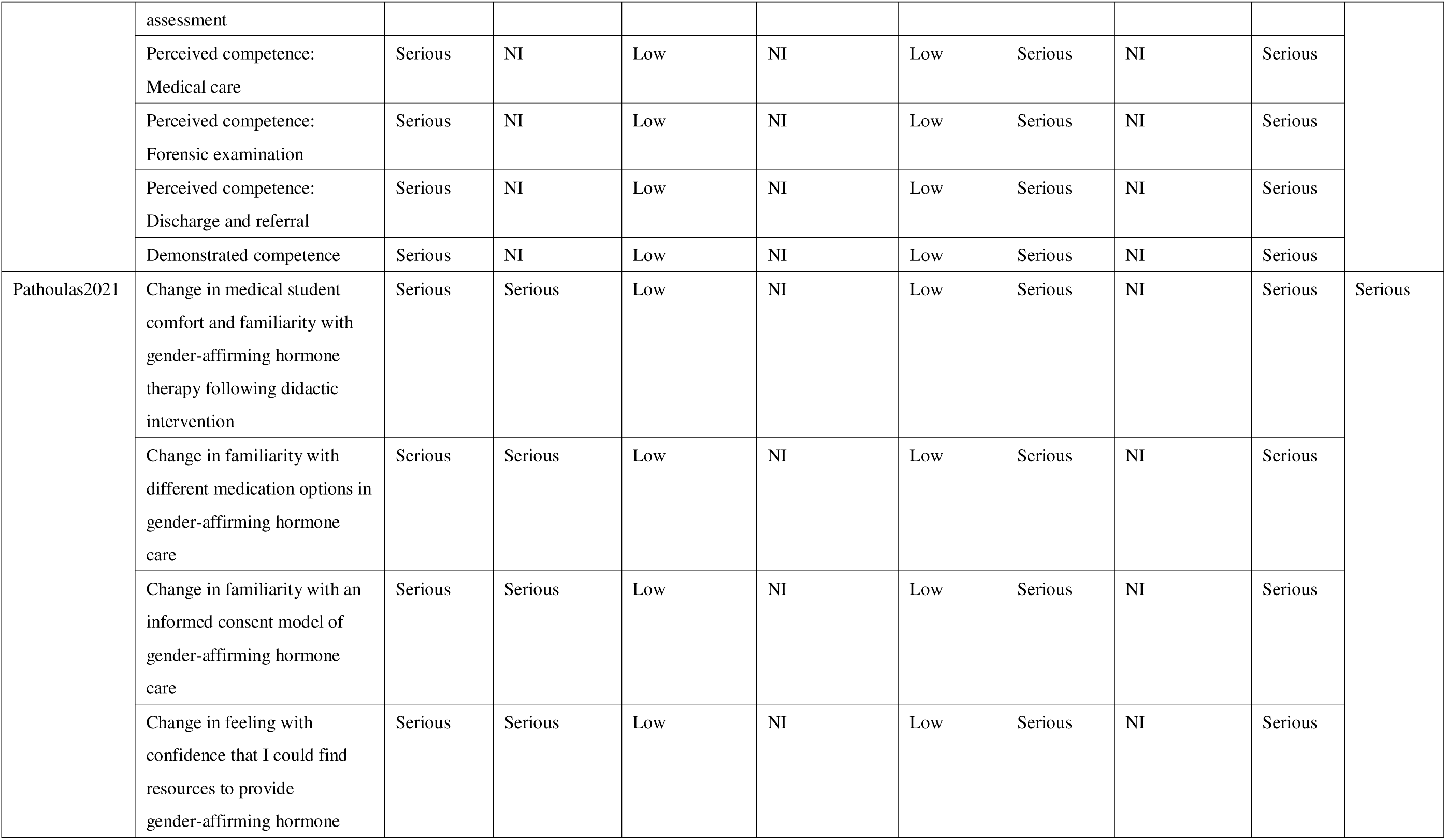
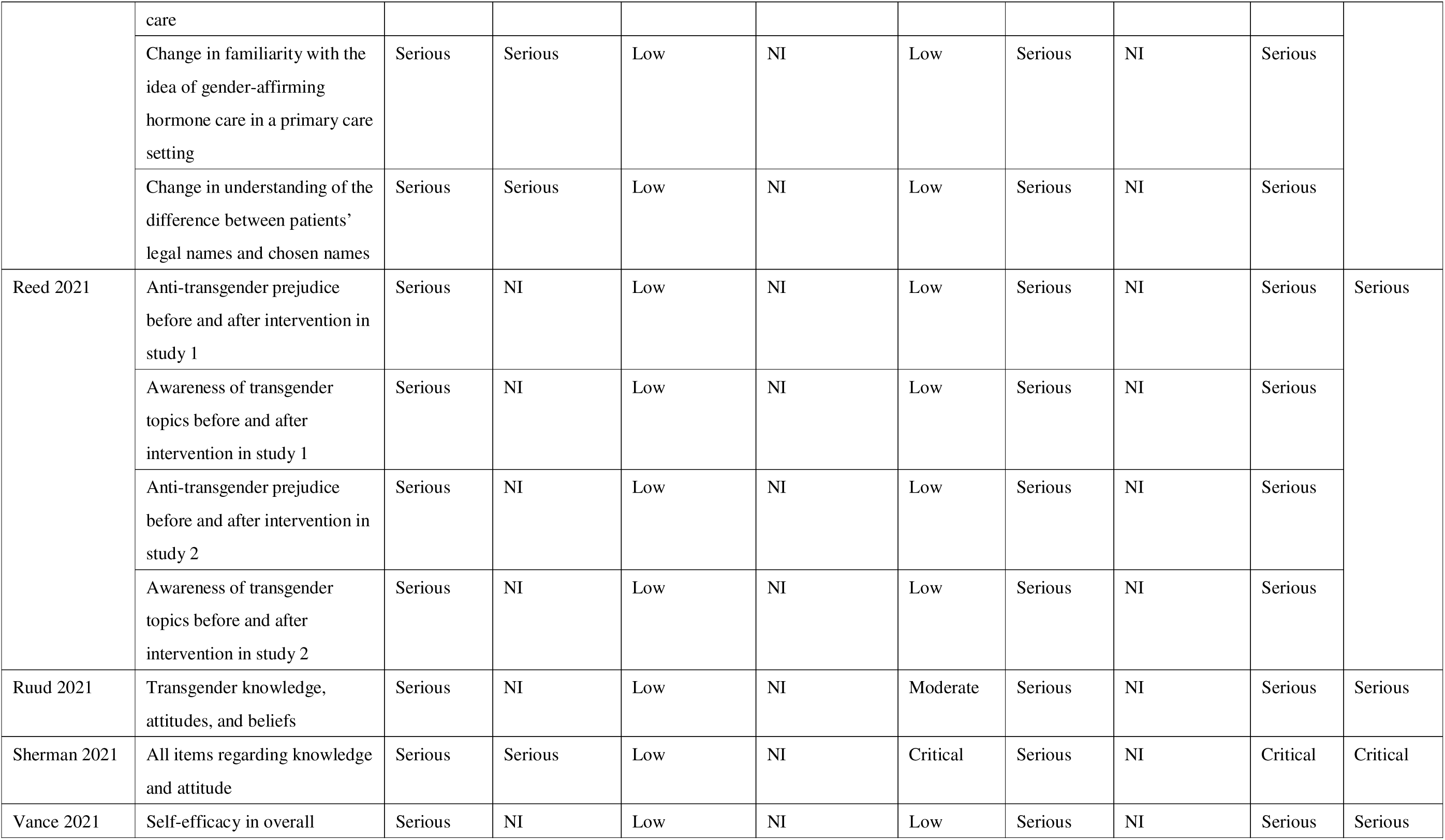
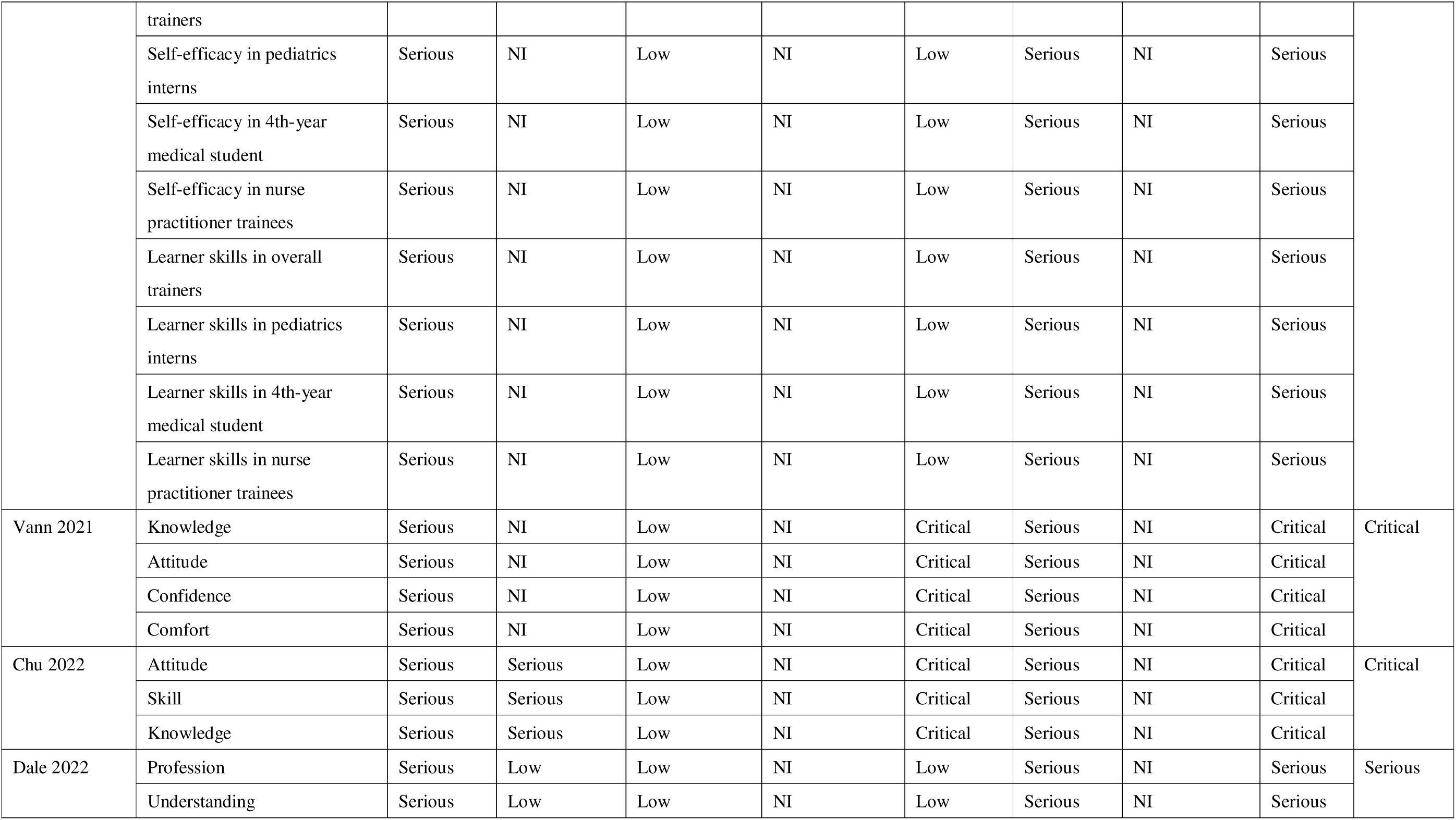
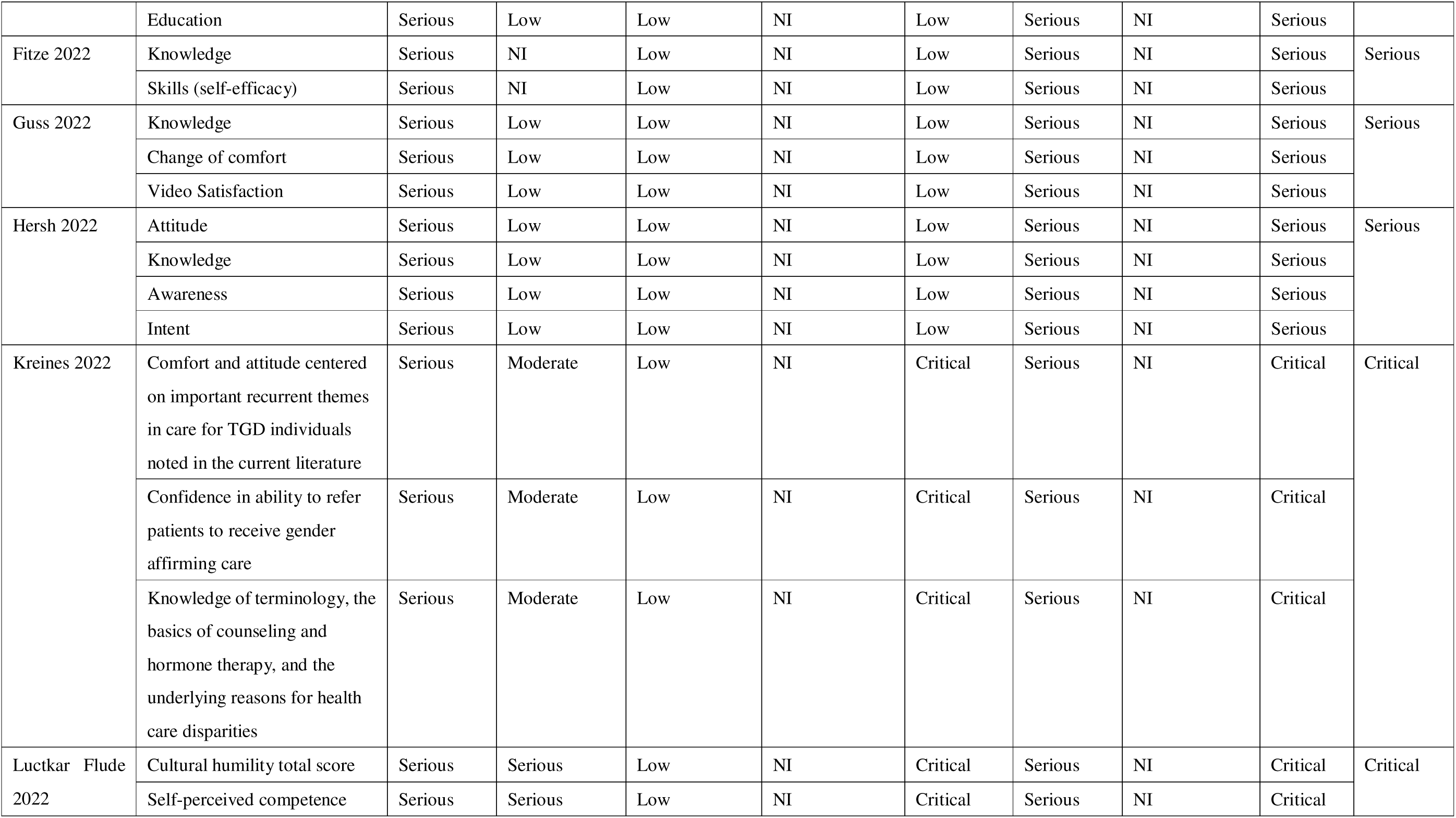
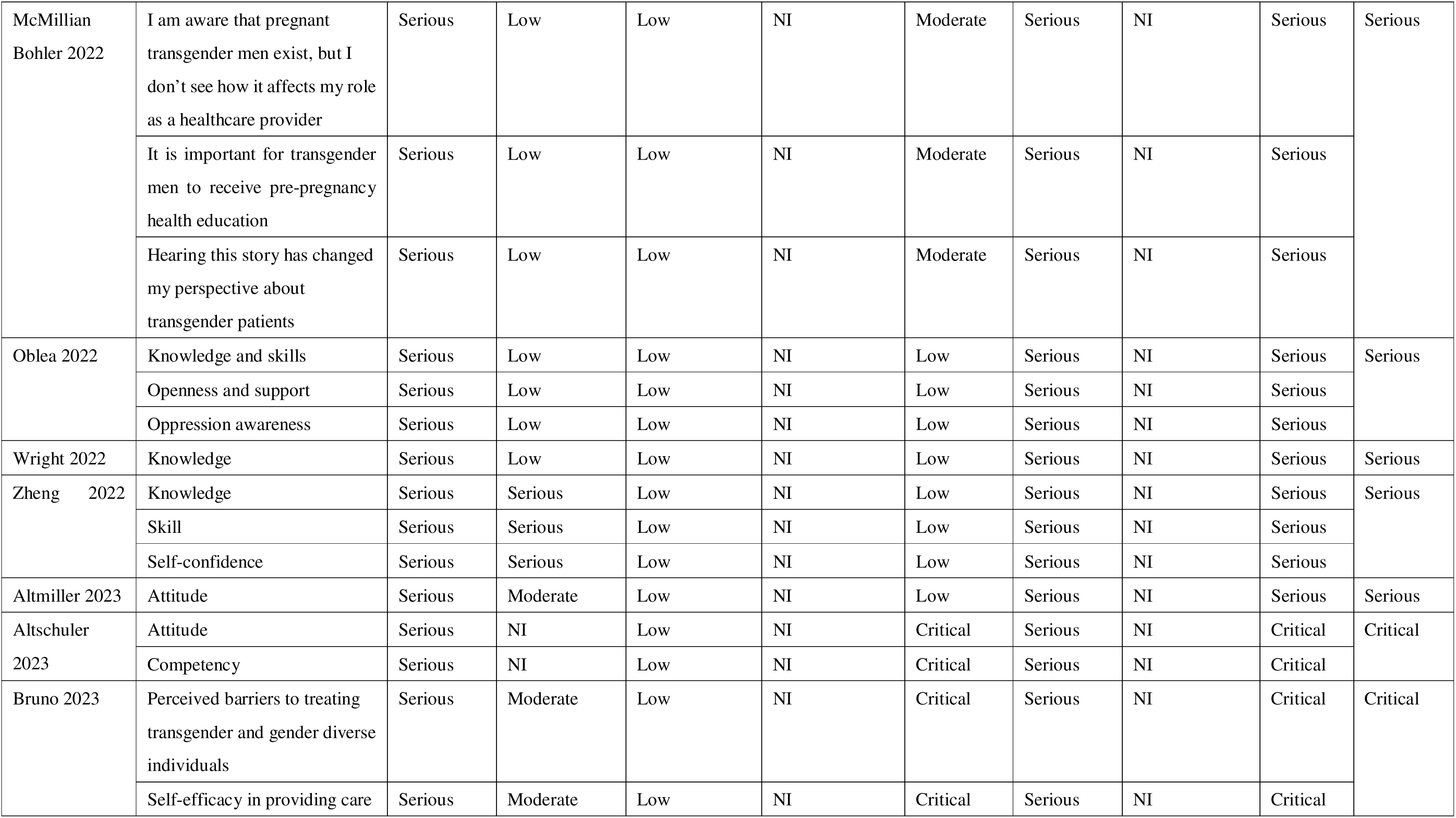
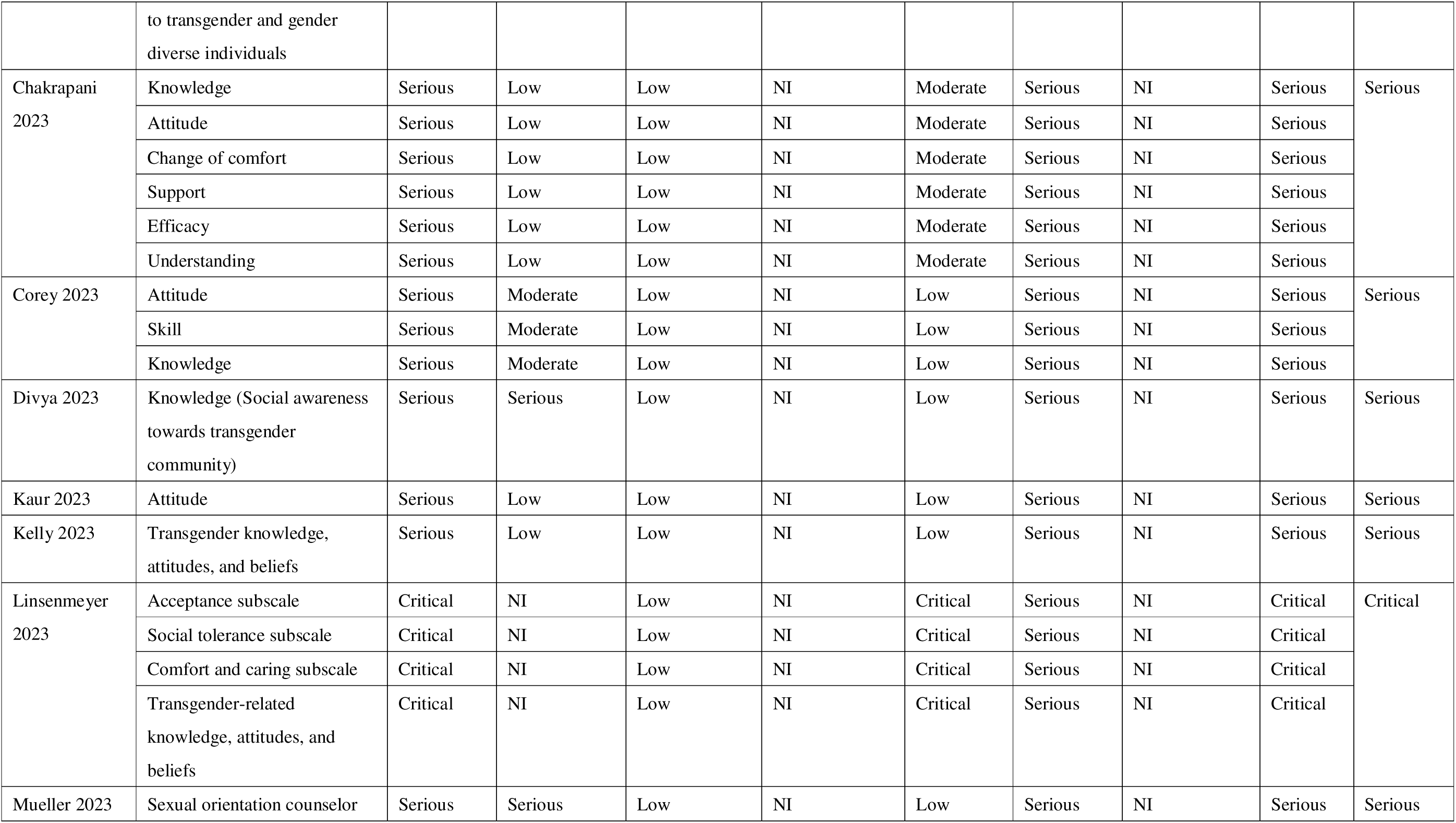
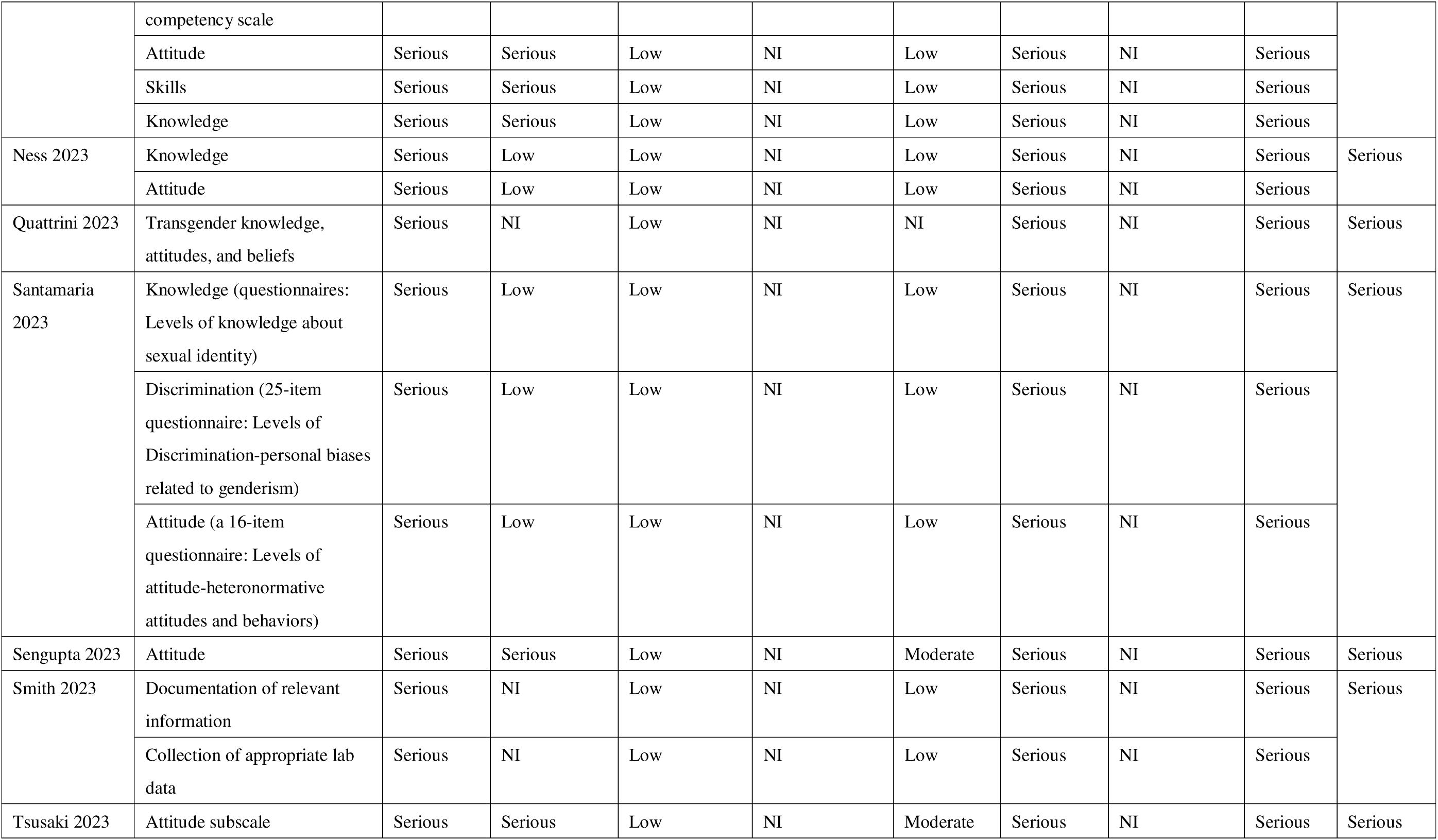
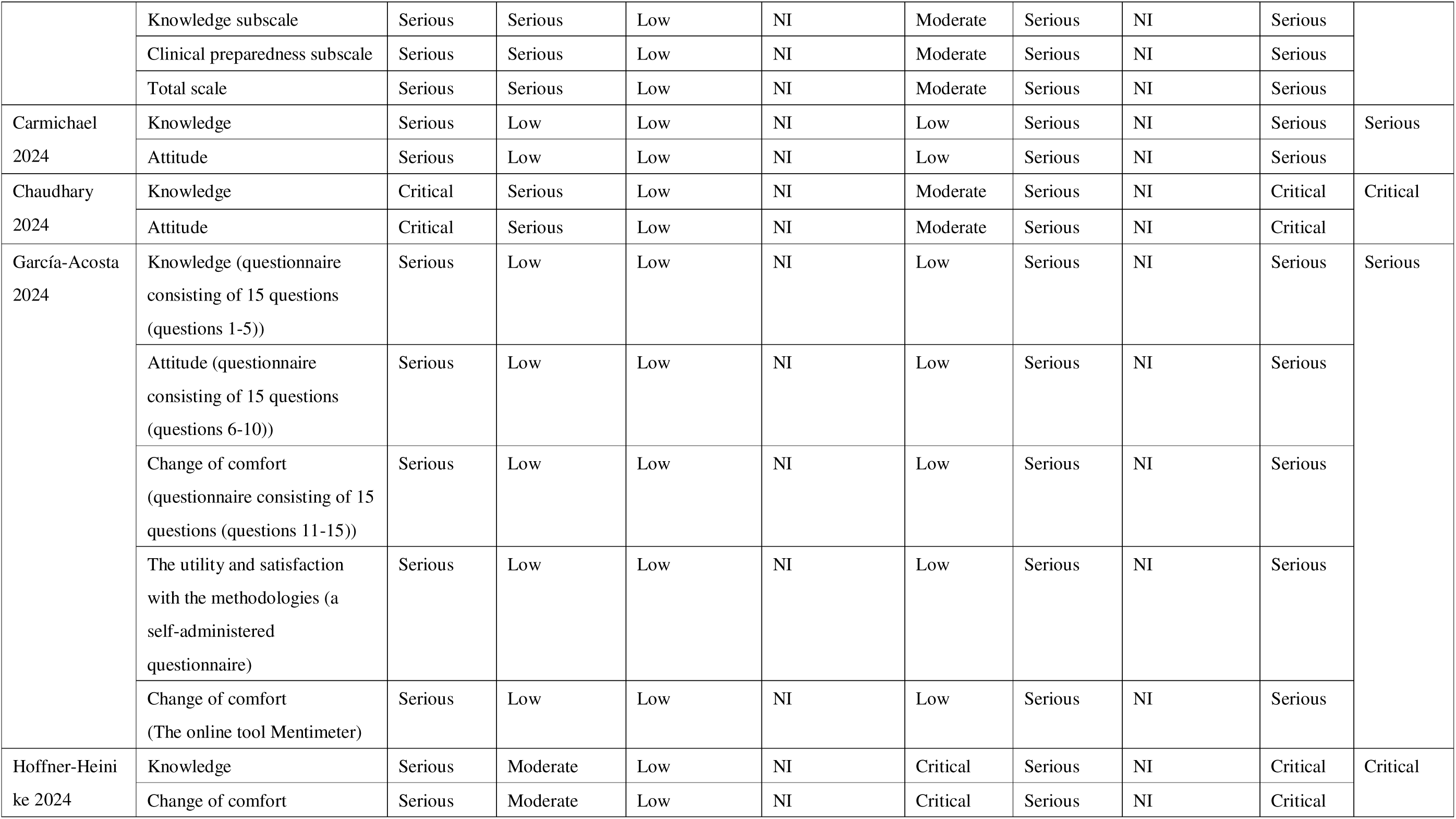
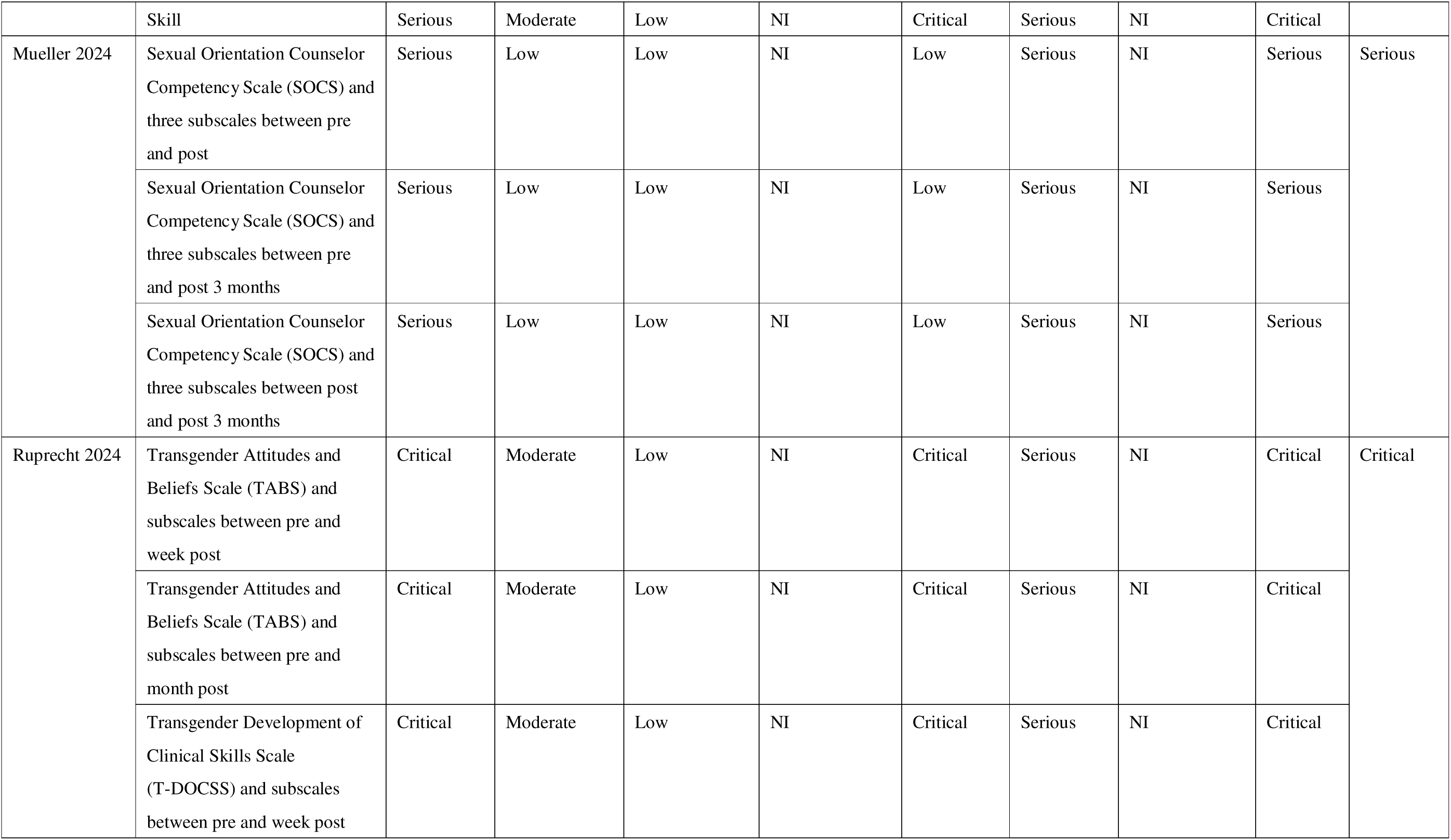
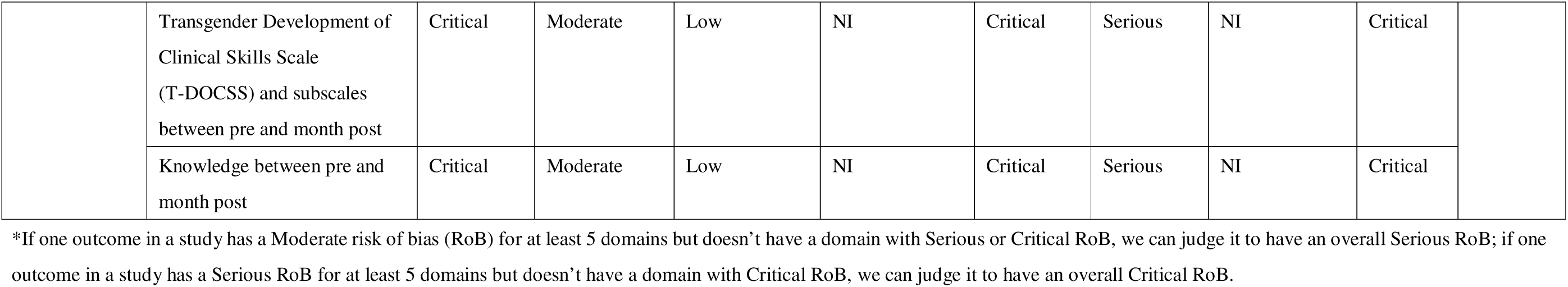

### Section 4.1 Qualitative review: Critical appraisal of included studies using the CASP checklist

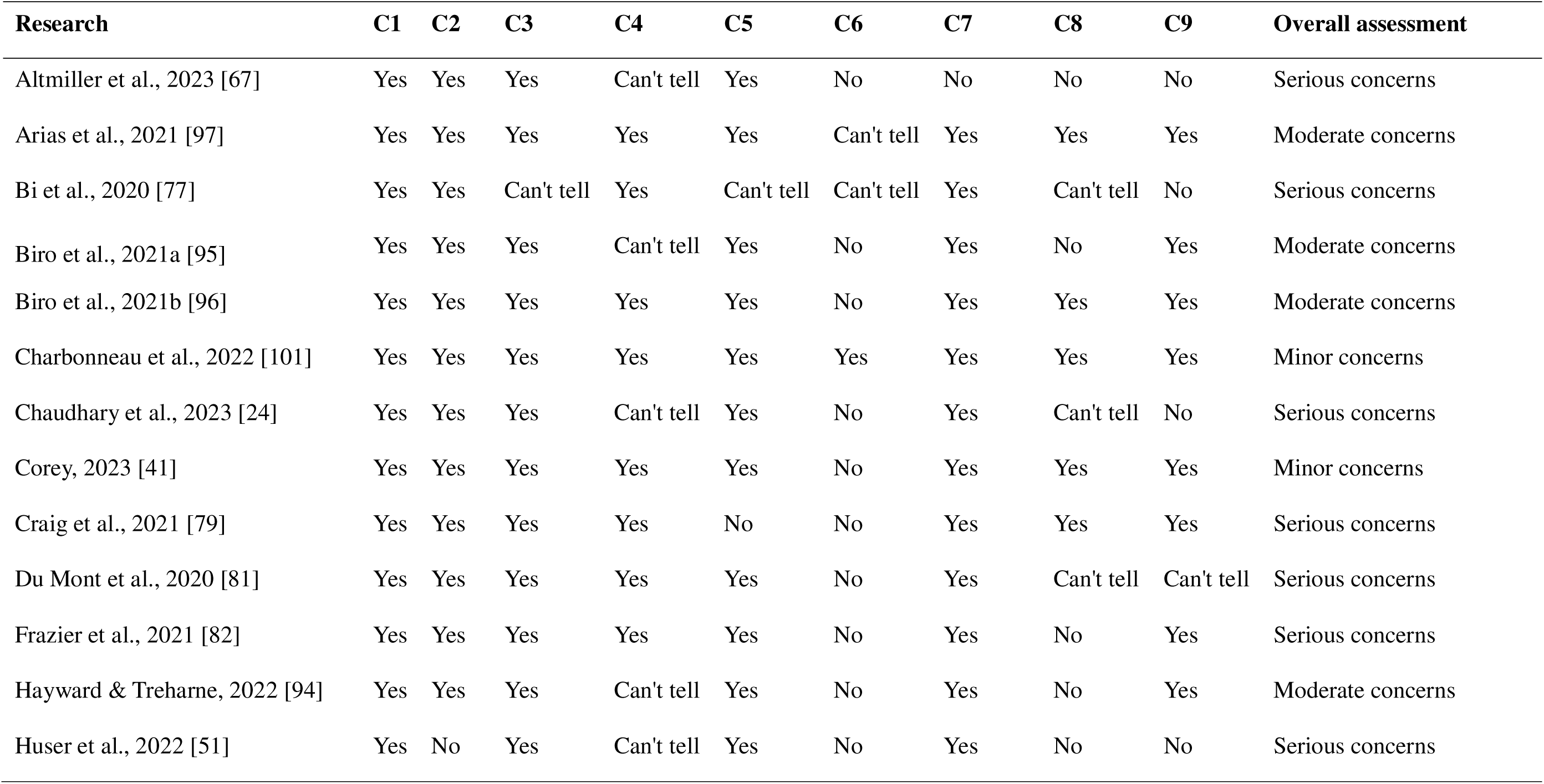
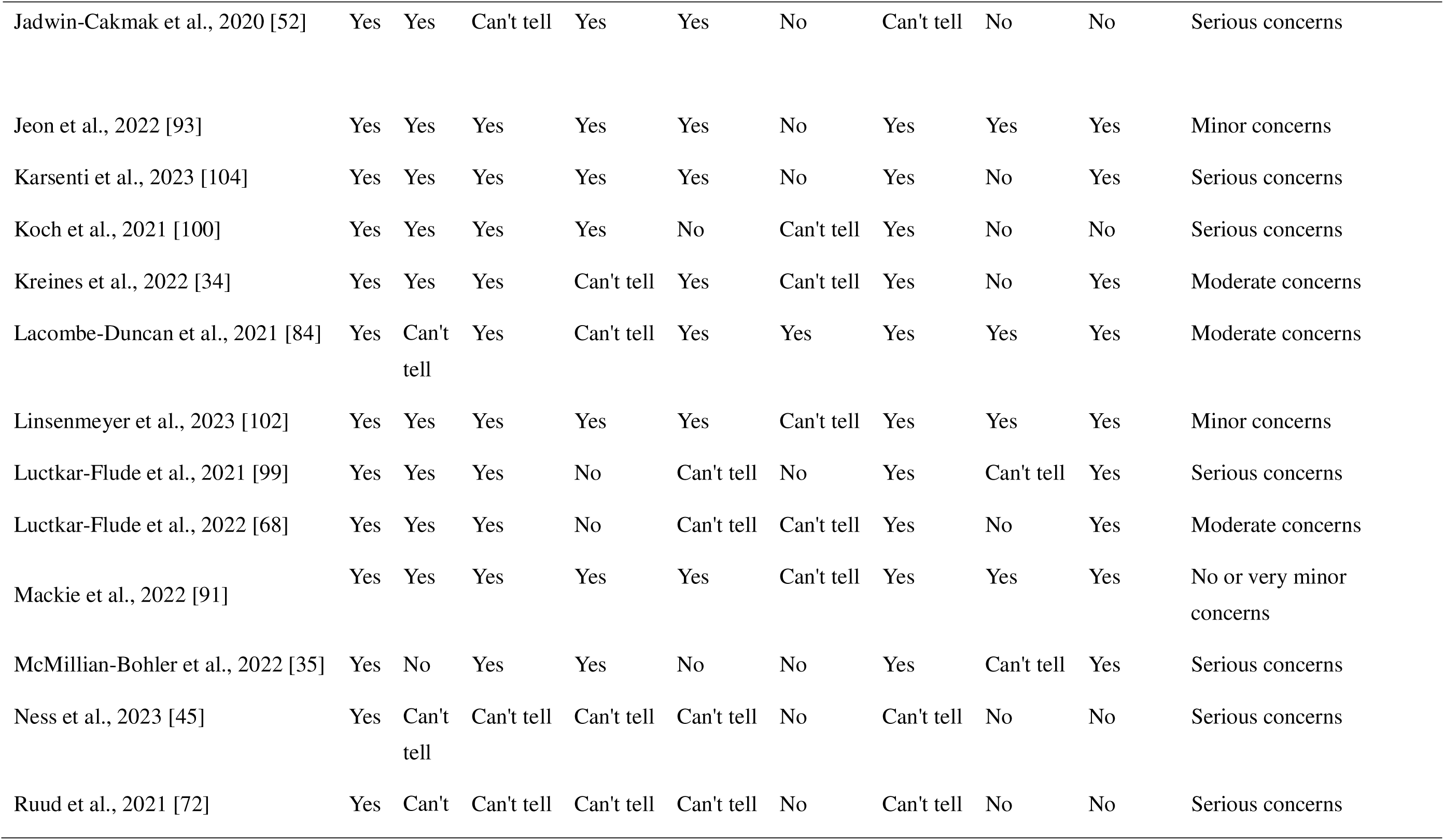
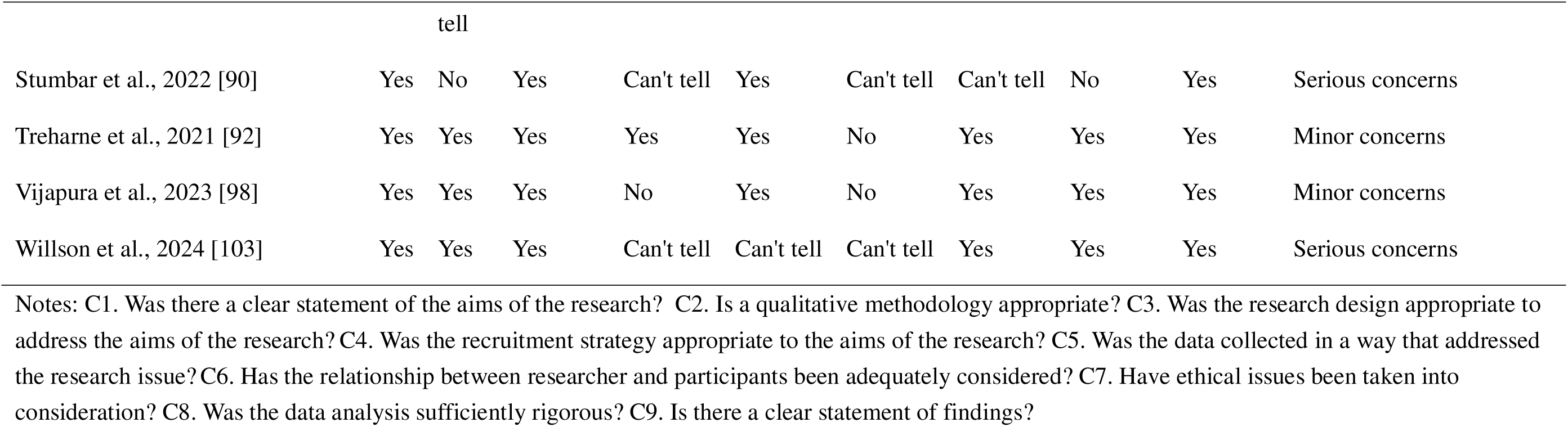

### Section 4.2 Qualitative review: Overall evaluations of methodological limitations of the studies

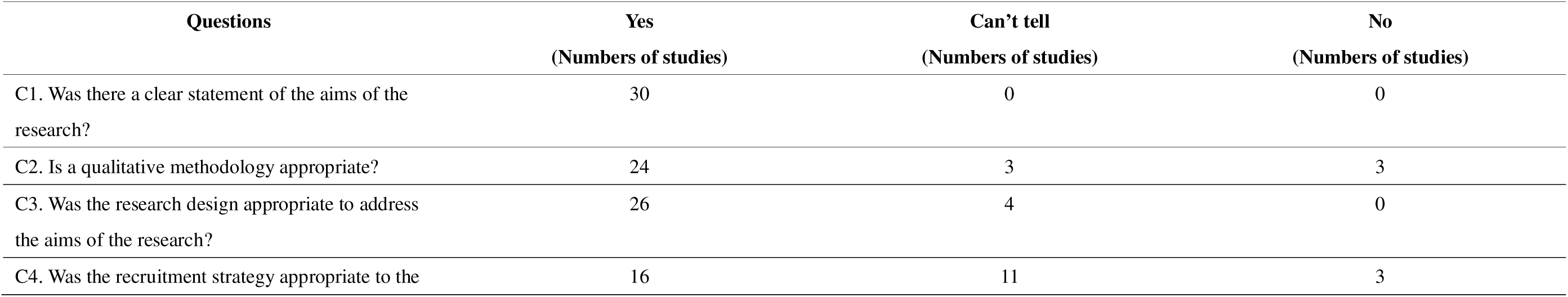
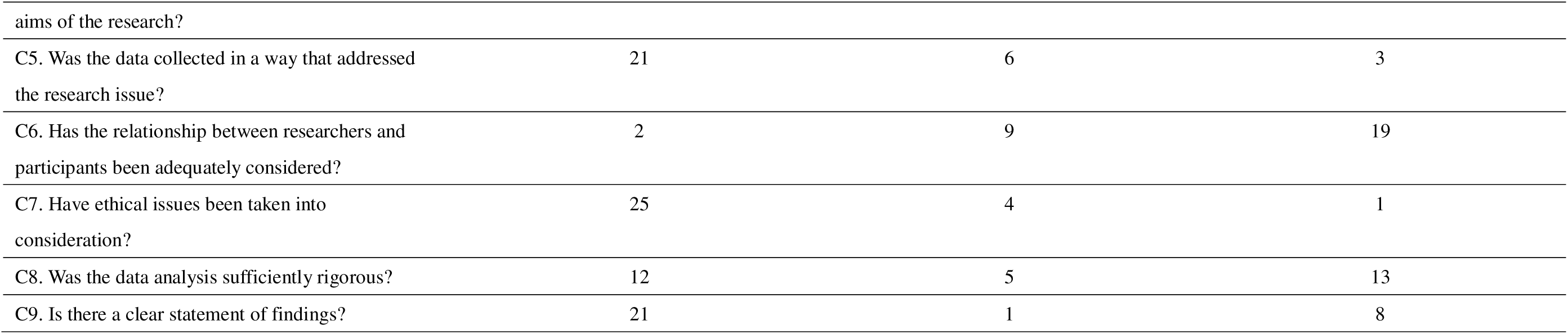

## Appendix 5. Summary of Findings Table

### Section 1: Knowledge outcome

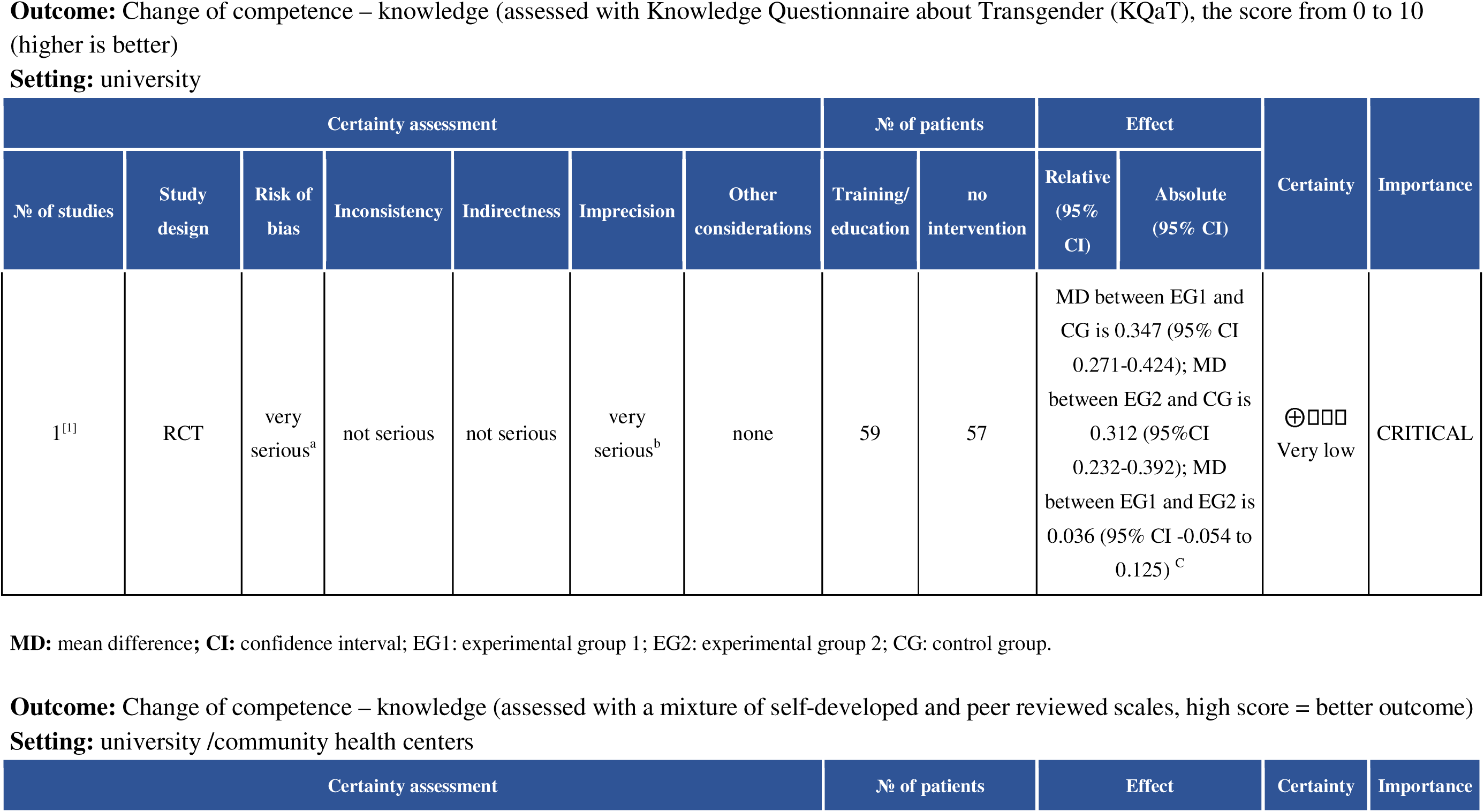
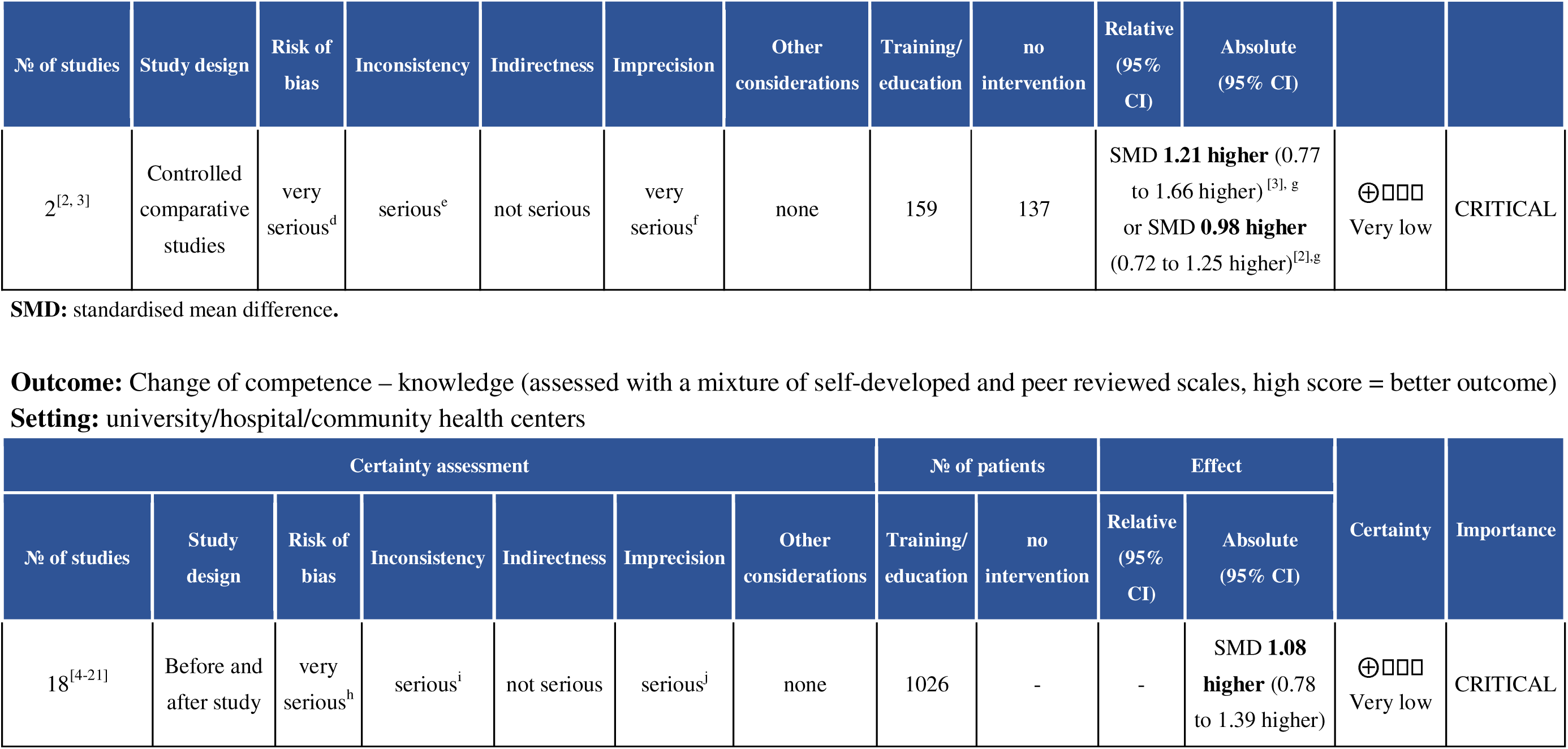

| Certainty assessment |  |  |  |  |  |  | № of patients |  | Effect |  | Certainty | Importance |
| --- | --- | --- | --- | --- | --- | --- | --- | --- | --- | --- | --- | --- |
| № of studies | Study design | Risk of bias | Inconsistency | Indirectness | Imprecision | Other considerations | Training/education | no intervention | Relative (95% CI) | Absolute (95% CI) |  |  |
| 1<sup>[1]</sup> | RCT | very serious<sup>a</sup> | not serious | not serious | very serious<sup>b</sup> | none | 59 | 57 | MD between EG1 and CG is 0.347 (95% CI 0.271-0.424); MD between EG2 and CG is 0.312 (95%CI 0.232-0.392); MD between EG1 and EG2 is 0.036 (95% CI -0.054 to 0.125)<sup>c</sup> |  | ⊕ □ □ □<br>Very low | CRITICAL |

| Certainty assessment |  |  |  |  |  |  | № of patients |  | Effect | Certainty | Importance |
| --- | --- | --- | --- | --- | --- | --- | --- | --- | --- | --- | --- |

| Nº of studies | Study design | Risk of bias | Inconsistency | Indirectness | Imprecision | Other considerations | Training/ education | no intervention | Relative (95% CI) | Absolute (95% CI) |  |  |
| --- | --- | --- | --- | --- | --- | --- | --- | --- | --- | --- | --- | --- |
| 2<sup>[2, 3]</sup> | Controlled comparative studies | very serious<sup>d</sup> | serious<sup>e</sup> | not serious | very serious<sup>f</sup> | none | 159 | 137 | SMD **1.21 higher** (0.77 to 1.66 higher)<sup>[3], g</sup><br>or SMD **0.98 higher** (0.72 to 1.25 higher)<sup>[2],g</sup> |  | ⊕ □□□<br>Very low | CRITICAL |

SMD: standardised mean difference.

| Certainty assessment |  |  |  |  |  |  | Nº of patients |  | Effect |  | Certainty | Importance |
| --- | --- | --- | --- | --- | --- | --- | --- | --- | --- | --- | --- | --- |
| Nº of studies | Study design | Risk of bias | Inconsistency | Indirectness | Imprecision | Other considerations | Training/ education | no intervention | Relative (95% CI) | Absolute (95% CI) |  |  |
| 18<sup>[4-21]</sup> | Before and after study | very serious<sup>h</sup> | serious<sup>i</sup> | not serious | serious<sup>j</sup> | none | 1026 | - | - | SMD **1.08 higher** (0.78 to 1.39 higher) | ⊕ □□□<br>Very low | CRITICAL |

#### Explanations

a. Risk of bias: Downgraded two levels because the study was judged to have some concerns in randomisation process, deviation from the intended interventions, measurements of the outcome, and selected reporting.
b. Imprecision: Downgraded two levels, because GRADE guidance suggests the sample size should be >/=800 for a continuous variable if the assumed small effect is 0.2 SMD; and if the overall sample size is between 240 to 400, downgrading by two levels is appropriate.
c. EG1= film forum intervention group; EG2=the problem-based learning group; CG=control group.
d. Risk of bias: Downgraded two levels because one study has serious risk of bias, and the other has critical risk of bias across multiple domains of ROBINS-I.
e. Inconsistency: Downgraded one level because the results of the two controlled comparative studies are contradictory.
f. Imprecision: Downgraded two levels because the estimated effect of one of the study^[3]^ crossed one decision thresholds, and the sample size is small. GRADE guidance suggests the sample size should be >/=800 for a continuous variable if the assumed small effect is 0.2 SMD; and if the overall sample size is between 240 to 400, downgrading by two levels is appropriate.
g. The two comparative studies measured knowledge concerning the care of trans people. One study ^[3]^ reported the post-training AIM score (intervention group 4.31±0.47, N=25; control group 3.42±0.76, N=22; p<0.001), and the LGBT-DOCCS-Knowledge subscale score (intervention group 6.42±0.59, N=25; control group 5.67±0.82, N=22; p<0.05). We pooled the above data using SMD to produce an average effect on change of knowledge SMD 1.21 higher (0.77 to 1.66 higher). The other study ^[2]^ reported intervention effect on three aspects of knowledge using a self-developed Likert scale (ranged 1 to 6) and found no statistical difference between the intervention and comparison group. These are, education in the course of study regarding gender identity disorders (pre and post training means were reported, and we calculated SMD 0.98 higher (0.72 to 1.25 higher), relevance of the topic for later professional life (no post training data reported), suitability of the module for teaching the topic (no post training data reported).
h. Risk of Bias: Downgraded two levels for the studies ^[4, 5, 7–10, 12–21]^have serious risk of bias across multiple ROBINS-I assessment domains, for example, high level of non-response rate (>20%) leads to bias due to selection of participants, all studies ultilized self-reported outcome measures leading to bias in measurement of outcomes, no information on the selection of the reported results; and 2 studies ^[6,11]^ were of critical risk of bias due to missing data.
i. Inconsistency: Downgraded one level. Although the direction of effect is all towards beneficial, there is heterogeneity (I^2^=90.7%) between the studies on the magnitude of effects. We explored the inconsistency through subgroup analysis by sample size, by category of health worker, by setting and by intervention (e.g. self-learning, facilitated learning, mixed approach), but the inconsistency in magnitude of effect remains unexplained. This potentially reduces our certainty on the observed magnitude of effect.
j. Imprecision: Downgraded one level. 95% CI of the estimated effect across two decision thresholds.

#### References

1. García-Acosta, J.M., et al., *Impact of a Formative Program on Transgender Healthcare for Nursing Students and Health Professionals. Quasi-Experimental Intervention Study.* Int J Environ Res Public Health, 2019. **16**(17).
2. Besse, M., et al., *Community-supported teaching on the topic of transgender identity in undergraduate medical education - a pilot project.* GMS J Med Educ, 2023. **40**(5): p. Doc58.
3. Bettergarcia, J., et al., *Effectiveness of a 9-month queer- and trans-affirming training for mental health providers: A waitlist control group design.* Professional Psychology: Research and Practice, 2024. **55**(1): p. 18-27.
4. Allison, M.K., et al., *Community-Engaged Development, Implementation, and Evaluation of an Interprofessional Education Workshop on Gender-Affirming Care.* Transgend Health, 2019. **4**(1): p. 280-286.
5. Carmichael, T.N., L.C. Copel, and R. McDermott-Levy, *Effect of an Education Intervention on Nursing Students’ Knowledge of and Attitudes Toward Caring for Transgender and Nonbinary People.* Nurse Educ, 2024.
6. Chu, H., et al., *Providing gender affirming and inclusive care to transgender men experiencing pregnancy.* Midwifery, 2023. **116**: p. 103550.
7. Corey, D., *Effectiveness of transgender health training on healthcare student’s knowledge, attitudes, and perceived competency providing gender-affiming healthcare*. 2023, Northern Arizona University. p. 160.
8. Dale, V.H. and R. Philomin, *Educating for Quality Transgender Health Care: A Survey Study of Medical Students.* Educ Health (Abingdon), 2022. **35**(1): p. 31-34.
9. Fitze, A.K. and B.K. Goodroad, *Implementation of low-fidelity simulation education to improve knowledge and self-efficacy for nurses caring for Post-genital affirming surgical patients.* Clinical Simulation in Nursing, 2022. **71**: p. 97-104.
10. Gorrotxategi, M.P., et al., *Improvement in Gender and Transgender Knowledge in University Students Through the Creative Factory Methodology.* Front Psychol, 2020. **11**: p. 367.
11. Hart, B.G., et al., *Developing a Curriculum on Transgender Health Care for Physician Assistant Students.* J Physician Assist Educ, 2021. **32**(1): p. 48-53.
12. Huser, N., et al., *Improving gender-affirming care in genetic counseling: Using educational tools that amplify transgender and/or gender non-binary community voices.* J Genet Couns, 2022. **31**(5): p. 1102-1112.
13. Jadwin-Cakmak, L., et al., *The Health Access Initiative: A Training and Technical Assistance Program to Improve Health Care for Sexual and Gender Minority Youth.* J Adolesc Health, 2020. **67**(1): p. 115-122.
14. Mueller, R.C., *Measuring the longitudinal impact of a transgender and gender diverse curriculum on nurse practitioner students’ and nurse practitioners’ cultural competence, knowledge, skills, and attitudes.* J Am Assoc Nurse Pract, 2024. **36**(2): p. 94-99.
15. Peterson, M.I., *Increasing certified registered nurse anesthetist (CRNA) knowledge and comfort in the care of transgender patients*. 2020, ProQuest Information & Learning.
16. Reed, O.M., *Evaluating the effectiveness of a transgender-affirmative care training on healthcare workers’ and trainees’ attitudes toward and knowledge of routine care and transition support for transgender individuals*. 2021, ProQuest Information & Learning.
17. Rosa-Vega, J., et al., *Educational intervention to improve pharmacist knowledge to provide care for transgender patients.* Pharm Pract (Granada), 2020. **18**(4): p. 2061.
18. Santamaria, F., et al., *Unmet Needs of Pediatricians in Transgender-Specific Care: Results of a Short-Term Training.* Horm Res Paediatr, 2023: p. 1-7.
19. Thompson, H., et al., *Evaluation of a gender-affirming healthcare curriculum for second-year medical students.* Postgrad Med J, 2020. **96**(1139): p. 515-519.
20. White Hughto, J.M. and K.A. Clark, *Designing a Transgender Health Training for Correctional Health Care Providers: A Feasibility Study.* Prison J, 2019. **99**(3): p. 329-342.
21. Wright, D. and A. Campedelli, *Pilot Study: Increasing Medical Student Comfort in Transgender Gynecology.* MedEdPublish (2016), 2022. **12**: p. 8.

### Section 2: Attitude outcome

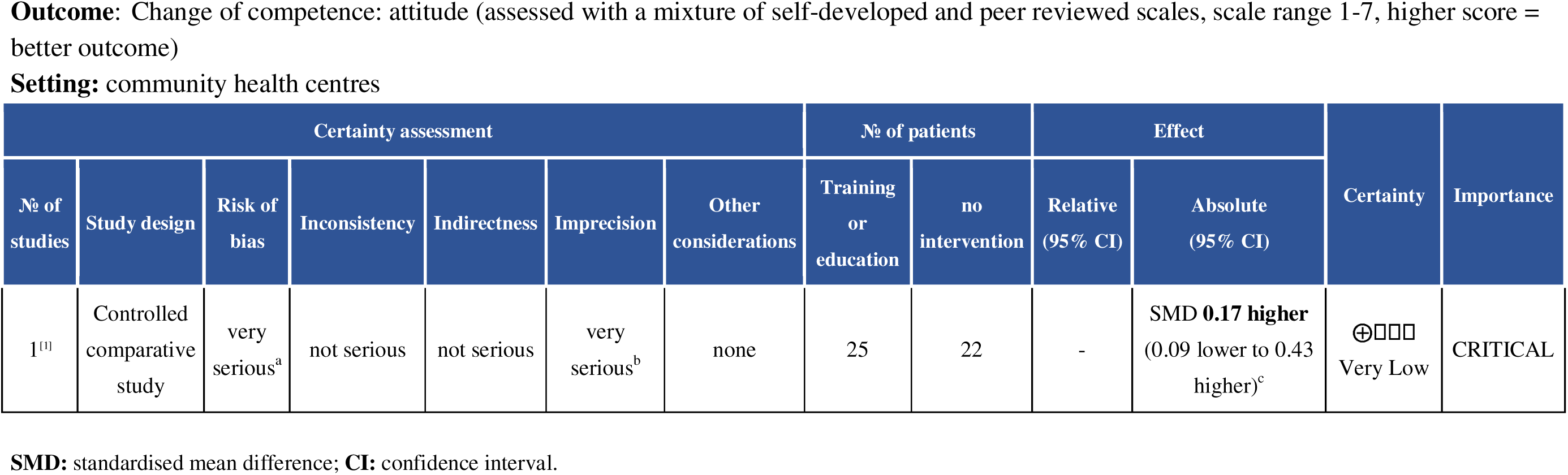
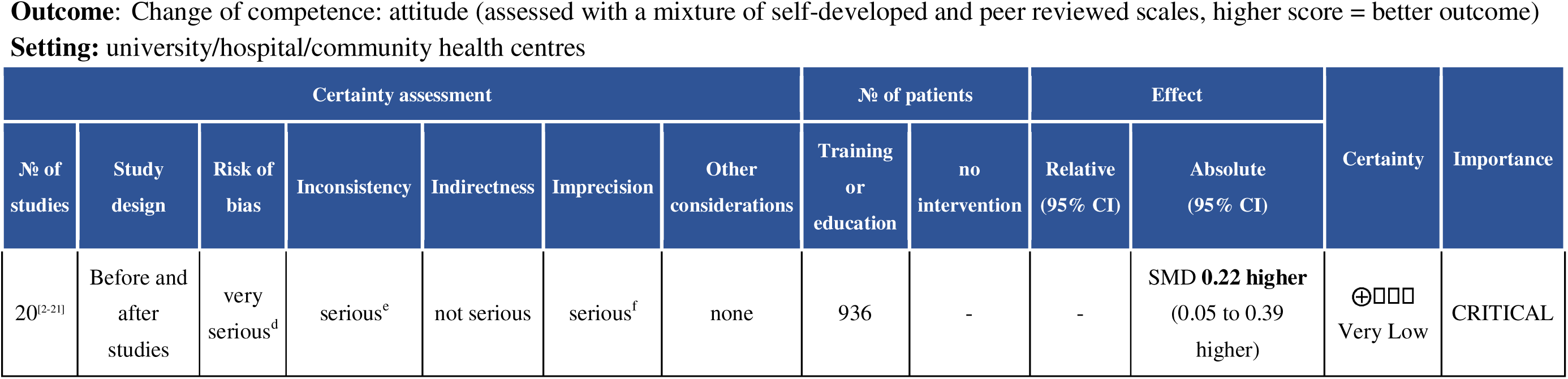

| Certainty assessment |  |  |  |  |  |  | Nº of patients |  | Effect |  | Certainty | Importance |
| --- | --- | --- | --- | --- | --- | --- | --- | --- | --- | --- | --- | --- |
| Nº of studies | Study design | Risk of bias | Inconsistency | Indirectness | Imprecision | Other considerations | Training or education | no intervention | Relative (95% CI) | Absolute (95% CI) |  |  |
| 1<sup>[1]</sup> | Controlled comparative study | very serious<sup>a</sup> | not serious | not serious | very serious<sup>b</sup> | none | 25 | 22 | - | SMD **0.17 higher** (0.09 lower to 0.43 higher)<sup>c</sup> | ⊕□□□<br>Very Low | CRITICAL |
| Nº of studies | Study design | Risk of bias | Inconsistency | Indirectness | Imprecision | Other considerations | Training or education | no intervention | Relative (95% CI) | Absolute (95% CI) |  |  |
| 20<sup>[2-21]</sup> | Before and after studies | very serious<sup>d</sup> | serious<sup>e</sup> | not serious | serious<sup>f</sup> | none | 936 | - | - | SMD **0.22 higher**<br>(0.05 to 0.39 higher) | ⊕□□□<br>Very Low | CRITICAL |

#### Explanations

a. Risk of bias: Downgraded by two level for the study utilized self-reported outcome measure, and lack of blinding introduces bias to outcome assessment.
b. Imprecision: Downgraded by two levels because 95%CI of the estimated effect crossed two decision thresholds, and the sample size is Small. GRADE guidance suggests the sample size should be >/=800 for a continuous variable if the assumed small effect is 0.2 SMD; and if the overall sample size is between 240 to 400, downgrading by two levels is appropriate.
c. The single controlled comparative study measured change in attitudes using several scales, and found no obvious difference between groups. The scales data includes the following (higher score indicates better outcome): ARBS-stability (4.25±0.45 versus 3.97±0.52, N=47, p=0.62), ARBS-tolerance (4.4±0.22 versus 4.43±0.17, N=47, p=0.42), LGBT-DOCCS-A (6.89±0.25 versus 6.88±0.54, N=47, p=0.36), Transphobia Scale (6.47±0.54 versus 6.44±0.37, N=47, p=0.79), and feelings themometers (92.13±10.71 versus 87.48±15.12, N=47, p=0.46). We pooled the above scale scores using SMD to produce an average effect SMD 0.17 higher (0.09 lower to 0.43 higher).
d. Risk of bias: Downgraded by two levels for majority ^[2, 4, 6-8, 10, 11, 14–21]^ of the studies were assessed as high risk of bias across multiple domains with the ROBINS-I tool. The overall risk of bias was very serious for this outcome.
e. Inconsistency: The I^2^ value is high (I^2^=73%).. We explored the inconsistency through subgroup analysis by sample size, by category of health worker, by setting and by intervention (e.g. self-learning, facilitated learning, mixed approach), but the inconsistency in magnitude of effect remains unexplained. This potentially reduces our certainty on the observed magnitude of effect.
f. Imprecision: Downgraded by one level for impression: the 95%CI of the estimated effect crossed the threshold of small and medium effect.

#### References

1. Bettergarcia, J., et al., *Effectiveness of a 9-month queer- and trans-affirming training for mental health providers: A waitlist control group design.* Professional Psychology: Research and Practice, 2024. **55**(1): p. 18-27.
2. Allison, M.K., et al., *Community-Engaged Development, Implementation, and Evaluation of an Interprofessional Education Workshop on Gender-Affirming Care.* Transgend Health, 2019. **4**(1): p. 280-286.
3. Altschuler, R., *Approaching trans healthcare competency: The implementation of trans health education for medical providers in Appalachia*. 2023, ProQuest Information & Learning.
4. Carmichael, T.N., L.C. Copel, and R. McDermott-Levy, *Effect of an Education Intervention on Nursing Students’ Knowledge of and Attitudes Toward Caring for Transgender and Nonbinary People.* Nurse Educ, 2024.
5. Chu, H., et al., *Providing gender affirming and inclusive care to transgender men experiencing pregnancy.* Midwifery, 2023. **116**: p. 103550.
6. Click, I.A., et al., *Transgender health education for medical students.* Clin Teach, 2020. **17**(2): p. 190-194.
7. Corey, D., *Effectiveness of transgender health training on healthcare student’s knowledge, attitudes, and perceived competency providing gender-affiming healthcare*. 2023, Northern Arizona University. p. 160.
8. Gorrotxategi, M.P., et al., *Improvement in Gender and Transgender Knowledge in University Students Through the Creative Factory Methodology.* Front Psychol, 2020. **11**: p. 367.
9. Hart, B.G., et al., *Developing a Curriculum on Transgender Health Care for Physician Assistant Students.* J Physician Assist Educ, 2021. **32**(1): p. 48-53.
10. Jadwin-Cakmak, L., et al., *The Health Access Initiative: A Training and Technical Assistance Program to Improve Health Care for Sexual and Gender Minority Youth.* J Adolesc Health, 2020. **67**(1): p. 115-122.
11. Lee, S.R., et al., *Attitudes Toward Transgender People Among Medical Students in South Korea.* Sex Med, 2021. **9**(1): p. 100278.
12. Linsenmeyer, W., et al., *Advancing Inclusion of Transgender Identities in Health Professional Education Programs: The Interprofessional Transgender Health Education Day.* J Allied Health, 2023. **52**(1): p. 24-31.
13. Luctkar-Flude, M., et al., *Impact of Virtual Simulation Games to Promote Cultural Humility Regarding the Care of Sexual and Gender Diverse Persons: A Multi-Site Pilot Study.* Clinical Simulation in Nursing, 2022. **71**: p. 146-158.
14. McMillian-Bohler, J., et al., *The Power of a Story: Enhancing Students’ Empathy for Transgender Pregnant Men.* J Nurs Educ, 2022. **61**(8): p. 489-492.
15. Mueller, R.C., *Measuring the longitudinal impact of a transgender and gender diverse curriculum on nurse practitioner students’ and nurse practitioners’ cultural competence, knowledge, skills, and attitudes.* J Am Assoc Nurse Pract, 2024. **36**(2): p. 94-99.
16. Mueller, R.C. and M.E. DeSimone, *Bringing Gender-Affirming Care to Primary Care: Use of a Multimodal Curriculum to Educate Nurse Practitioners and Nurse Practitioner Students.* Nurse Educ, 2023.
17. Ruud, M.N., et al., *Health History Skills for Interprofessional Learners in Transgender and Nonbinary Populations.* J Midwifery Womens Health, 2021. **66**(6): p. 778-786.
18. Santamaria, F., et al., *Unmet Needs of Pediatricians in Transgender-Specific Care: Results of a Short-Term Training.* Horm Res Paediatr, 2023: p. 1-7.
19. Sengupta, T., et al., *Knowledge, Attitudes, and Beliefs of Medical Students Toward Transgender Healthcare: A Community-Driven Initiative.* Cureus, 2023. **15**(12): p. e49992.
20. Sherman, A.D.F., et al., *Transgender and gender diverse health education for future nurses: Students’ knowledge and attitudes.* Nurse Educ Today, 2021. **97**: p. 104690.
21. Thompson, H., et al., *Evaluation of a gender-affirming healthcare curriculum for second-year medical students.* Postgrad Med J, 2020. **96**(1139): p. 515-519.

### Section 3: Discrimination outcome

| Certainty assessment | Nº of patients | Effect | Certainty | Importance |
| --- | --- | --- | --- | --- |

| № of studies | Study design | Risk of bias | Inconsistency | Indirectness | Imprecision | Other considerations | Training or education | no intervention | Relative (95% CI) | Absolute (95% CI) |  |  |
| --- | --- | --- | --- | --- | --- | --- | --- | --- | --- | --- | --- | --- |
| 2<sup>[1, 2]</sup> | Before and after studies | serious<sup>a</sup> | not serious | not serious | very serious<sup>b</sup> | none | 247 |  | - | SMD **0.31 lower**<br>(0.68 lower to 0.06 higher) | ⊕□□□<br>Very low | CRITICAL |

**SMD:** standardised mean difference; **CI:** confidence interval; **ATPS:** Anti-Transgender Prejudice Scale; **ATTM:** Awareness of Transgender Topics Measure

#### Explanations

a. a. One study^[1]^ recruited participants from a variety of settings, including hospitals, medical centers, universities, federally qualified health care centers, private practices, Veterans Affairs medical centers, and other health care settings.
b. b. The study was downgraded one level for serious risk of bias across multiple domains of ROBINS-I.
c. c. Imprecission: Downgraded by two levels because the 95%CI of the estimated effect crossed two decision thresholds, and the sample size is small. GRADE guidance suggests the sample size should be >/=800 for a continuous variable if the assumed small effect is 0.2 SMD; and if the overall sample size is between 240 to 400, downgrading by two levels is appropriate.

#### References

1. Reed, O.M., *Evaluating the effectiveness of a transgender-affirmative care training on healthcare workers’ and trainees’ attitudes toward and knowledge of routine care and transition support for transgender individuals*. 2021, ProQuest Information & Learning.
2. Santamaria, F., et al., *Unmet Needs of Pediatricians in Transgender-Specific Care: Results of a Short-Term Training.* Horm Res Paediatr, 2023: p. 1-7.

### Section 4: Skill outcome

| Certainty assessment |  |  |  |  |  |  | Nº of patients |  | Effect |  | Certainty | Importance |
| --- | --- | --- | --- | --- | --- | --- | --- | --- | --- | --- | --- | --- |
| Nº of studies | Study design | Risk of bias | Inconsistency | Indirectness | Imprecision | Other considerations | Training or education | no intervention | Relative (95% CI) | Absolute (95% CI) |  |  |
| 1<sup>[1]</sup> | Non-randomised comparative study | very serious<sup>a</sup> | not serious | not serious | serious<sup>b</sup> | none | 134 | 115 |  | Learning gain in diagnostic criteria (MD -1.61), and in treatment and care (MD -1.74)<sup>c</sup> | ⊕□□□<br>Very low | CRITICAL |
| 1<sup>[2]</sup> | Non-randomised comparative study | very serious<sup>a</sup> | not serious | not serious | serious<sup>b</sup> | none | 25 | 22 | - | SMD 1.29 **higher** (0.84 to 1.74 higher)<sup>[2],d</sup> | ⊕□□□<br>Very low | CRITICAL |

| Certainty assessment |  |  |  |  |  |  | № of patients |  | Effect |  | Certainty | Importance |
| --- | --- | --- | --- | --- | --- | --- | --- | --- | --- | --- | --- | --- |
| № of studies | Study design | Risk of bias | Inconsistency | Indirectness | Imprecision | Other considerations | Training or education | no intervention | Relative (95% CI) | Absolute (95% CI) |  |  |
| 9<sup>[3-11]</sup> | Before and after study | very serious<sup>e</sup> | serious<sup>f</sup> | not serious | serious<sup>g</sup> | none | 423 |  | - | SMD **0.96 higher**<br>(0.56 to 1.37 higher) | ⊕ □ □ □<br>Very low | CRITICAL |

#### Explanations

a. Risk of bias: Downgraded by two levels because the studies had moderate^[2]^ to serious^[1]^ risk of bias due to confounding, and serous risk of bias due to the utilisaition of self-reported outcome measures, and the outcome assessors were aware of the intervention received by the study participants.
b. Imprecision: Downgraded by two levels GRADE guidance suggests the sample size should be >/=800 for a continuous variable if the assumed small effect is 0.2 SMD; and if the overall sample size is between 240 to 400, downgrading by two levels is appropriate.
c. The study reported skill related learning gain in diagnostic criteria (pre-mean 4.55, post-mean 2.94, MD -1.61), and in treatment and care (pre-mean 4.57, post-mean 2.83, MD -1.74). A numerically larger learning gain was found in the intervention group (negative MD value indicates learning gain). Authors used a self-developed 6-point Likert scale, lower score indicates better outcome.
d. Assessed using Lesbian, Gay, and Bisexual Affirmative Counseling Self-Efficacy Inventory (LGB-CSI), 6-point Likert scale, higher score means better skill; and Lesbian, Gay, Bisexual, and Transgender Development of Clinical Skills Scale (LGBT-DOCSS subscale), 7-point Likert scale, higher score indicates better skill. The observed between group difference as measured by LGB-CSI is 116.28±25.59 versus 115.91±29.53, and by LGBT-DOCCS-clinical preparedness 4.45±1.13 versus 4.49±1.13. We were able to pool the above scale data to produce an average change in skill, SMD 1.29 (95%CI 0.84 to 1.74).
e. Risk of bias: Downgraded by two level because majority of the studies were assessed as high risk of bias across multiple domains with the ROBINS-I tool. The overall risk of bias was very serious for this outcome.
f. Inconsistency: Downgraded by one level because the heterogeneity (I^2^=89.6%) between the studies was apparent both in terms of direction and magnitude of effect. We explored the inconsistency through subgroup analysis by sample size, by category of health worker, by setting and by intervention (e.g. self-learning, facilitated learning, mixed approach), but the inconsistency in magnitude of effect remains unexplained. This potentially reduces our certainty on the observed magnitude of effect.
g. Imprecision: Downgraded one level: the 95%CI of the estimated effect crossed the threshold of large effect.

#### References

1. Besse, M., et al., *Community-supported teaching on the topic of transgender identity in undergraduate medical education - a pilot project.* GMS J Med Educ, 2023. **40**(5): p. Doc58.
2. Bettergarcia, J., et al., *Effectiveness of a 9-month queer- and trans-affirming training for mental health providers: A waitlist control group design.* Professional Psychology: Research and Practice, 2024. **55**(1): p. 18-27.
3. Bruno, J., et al., *TransECHO: A National Tele-Education Program for Expanding Transgender and Gender Diverse Health Care.* LGBT Health, 2023. **10**(6): p. 456-462.
4. Chu, H., et al., *Providing gender affirming and inclusive care to transgender men experiencing pregnancy.* Midwifery, 2023. **116**: p. 103550.
5. Corey, D., *Effectiveness of transgender health training on healthcare student’s knowledge, attitudes, and perceived competency providing gender-affiming healthcare*. 2023, Northern Arizona University. p. 160.
6. Fitze, A.K. and B.K. Goodroad, *Implementation of low-fidelity simulation education to improve knowledge and self-efficacy for nurses caring for Post-genital affirming surgical patients.* Clinical Simulation in Nursing, 2022. **71**: p. 97-104.
7. Jadwin-Cakmak, L., et al., *The Health Access Initiative: A Training and Technical Assistance Program to Improve Health Care for Sexual and Gender Minority Youth.* J Adolesc Health, 2020. **67**(1): p. 115-122.
8. Mueller, R.C., *Measuring the longitudinal impact of a transgender and gender diverse curriculum on nurse practitioner students’ and nurse practitioners’ cultural competence, knowledge, skills, and attitudes.* J Am Assoc Nurse Pract, 2024. **36**(2): p. 94-99.
9. Mueller, R.C. and M.E. DeSimone, *Bringing Gender-Affirming Care to Primary Care: Use of a Multimodal Curriculum to Educate Nurse Practitioners and Nurse Practitioner Students.* Nurse Educ, 2023.
10. Ruud, M.N., et al., *Health History Skills for Interprofessional Learners in Transgender and Nonbinary Populations.* J Midwifery Womens Health, 2021. **66**(6): p. 778-786.
11. Thompson, H., et al., *Evaluation of a gender-affirming healthcare curriculum for second-year medical students.* Postgrad Med J, 2020. **96**(1139): p. 515-519.

### Section 5: Competence outcome

| Certainty assessment | Nº of patients | Effect | Certainty | Importance |
| --- | --- | --- | --- | --- |

| Nº of studies | Study design | Risk of bias | Inconsistency | Indirectness | Imprecision | Other considerations | Training or education | no intervention | Relative (95% CI) | Absolute (95% CI) |  |  |
| --- | --- | --- | --- | --- | --- | --- | --- | --- | --- | --- | --- | --- |
| 2<sup>[1, 2]</sup> | RCT | very serious<sup>a</sup> | not serious | not serious | very serious<sup>b</sup> | none | 70 | 65 | - | SMD 0.66 higher (0.31 to 1 higher)<sup>c</sup> | ⊕□□□<br>Very low | CRITICAL |

| Certainty assessment |  |  |  |  |  |  | Nº of patients |  | Effect |  | Certainty | Importance |
| --- | --- | --- | --- | --- | --- | --- | --- | --- | --- | --- | --- | --- |
| Nº of studies | Study design | Risk of bias | Inconsistency | Indirectness | Imprecision | Other considerations | Training or education | no intervention | Relative (95% CI) | Absolute (95% CI) |  |  |
| 16<sup>[3-18]</sup> | Before and after studies | very serious<sup>d</sup> | serious<sup>e</sup> | not serious | serious<sup>f</sup> | none | 909 | - | - | SMD **0.55 higher** (0.29 to 0.81 higher) | ⊕□□□<br>Very low | CRITICAL |

#### Explanations

a. Risk of bias: Downgraded by two levels due to the following reasons: 1) concerns over the lack of information about the allocation sequence and concealment process,
b. high risk of bias in the measurement of outcomes, due to the outcomes were self-reported; 3) lack of study protocol caused some concerns over selective reporting.
c. Imprecision: Downgraded by two levels for the 95%CI of the estimated effect crossed two decision thresholds, and the sample size is small. GRADE guidance suggests the sample size should be >/=800 for a continuous variable if the assumed small effect is 0.2 SMD; and if the overall sample size is between 240 to 400, downgrading by two levels is appropriate.
d. One study^[2]^ measured general competence using SOCCS-3, and reported post-training mean as 4.45±0.39 versus 4.13±0.57. We were able to calculate the MD between groups (SMD=0.66, 95%CI 0.31 to 1.00). The other study^[1]^ did not report any numerical outcome data, but provided the following statement “*Changes in the documentation of gender identity were more with five of the intervention health centers increasing documentation, although generally by less than 10%, compared to two of the control health centers” which means Intervention health centers tended to increase documentation of preventive services more than control health centers, but the changes were not significant*”.
e. Risk of bias: Downgraded two levels for majority^[4, 6-8, 10–12, 14–16, 18]^ of the studies were assessed as high risk of bias across multiple domains with the ROBINS-I tool. The overall risk of bias was rated as very serious for this outcome. Response rate falling below 60% is a major issue in these studies, which leads to potential selection bias. Missing data, and bias in measurement of outcomes are the other common risk of bias among this body of evidence.
f. Inconsistency: Downgraded one level because the unexplained heterogeneity between the studies was apparent both in terms of direction and magnitude of effect (I^2^=85.7%), We explored the inconsistency through subgroup analysis by sample size, by category of health worker, by setting and by intervention (e.g. self-learning, facilitated learning, mixed approach), but the inconsistency in magnitude of effect remains unexplained. This potentially reduces our certainty on the observed magnitude of effect.
g. Imprecision: Downgraded one level because the 95%CI of the estimated effect crossed threshold of medium to large effect.

#### References

1. Mayer, K.H., et al., *Training Health Center Staff in the Provision of Culturally Responsive Care for Sexual and Gender Minority Patients: Results of a Randomized Controlled Trial.* LGBT Health, 2024. **11**(2): p. 131-142.
2. Michals-Voigt, M., *The effect of a transgender PowerPoint on knowledge, skills, and attitudes of mental health graduate students*. 2021, ProQuest Information & Learning.
3. Altschuler, R., *Approaching trans healthcare competency: The implementation of trans health education for medical providers in Appalachia*. 2023, ProQuest Information & Learning.
4. Bi, S., et al., *Teaching Intersectionality of Sexual Orientation, Gender Identity, and Race/Ethnicity in a Health Disparities Course.* MedEdPORTAL, 2020. **16**: p. 10970.
5. Bruno, J., et al., *TransECHO: A National Tele-Education Program for Expanding Transgender and Gender Diverse Health Care.* LGBT Health, 2023. **10**(6): p. 456-462.
6. Craig, S.L., et al., *The role of facilitator training in intervention delivery: Preparing clinicians to deliver AFFIRMative group cognitive behavioral therapy to sexual and gender minority youth.* Journal of Gay & Lesbian Social Services, 2021. **33**(1): p. 56-77.
7. Du Mont, J., S.D. Kosa, and S. Macdonald, *Evaluation of an e-Learning Curriculum for Forensic Nurses on Trans-Affirming Postsexual Assault Care.* Transgend Health, 2021. **6**(5): p. 284-289.
8. Du Mont, J., et al., *Providing trans-affirming care for sexual assault survivors: An evaluation of a novel curriculum for forensic nurses.* Nurse Educ Today, 2020. **93**: p. 104541.
9. Hart, B.G., et al., *Developing a Curriculum on Transgender Health Care for Physician Assistant Students.* J Physician Assist Educ, 2021. **32**(1): p. 48-53.
10. Kelly, A., *Implementing a gender identity assessment in the inpatient psychiatric setting: a quality improvement project*. 2022, Regis College. p. 66.
11. Lacombe-Duncan, A., et al., *Implementation and evaluation of the ’Transgender Education for Affirmative and Competent HIV and Healthcare (TEACHH)’ provider education pilot.* BMC Med Educ, 2021. **21**(1): p. 561.
12. Lee, S.R., et al., *Attitudes Toward Transgender People Among Medical Students in South Korea.* Sex Med, 2021. **9**(1): p. 100278.
13. Linsenmeyer, W., et al., *Advancing Inclusion of Transgender Identities in Health Professional Education Programs: The Interprofessional Transgender Health Education Day.* J Allied Health, 2023. **52**(1): p. 24-31.
14. Mackler, C.D., *Gender affirming sexual assault nurse examiner care: A program evaluation and quality improvement project at a community-based rape crisis center*. 2021, ProQuest Information & Learning.
15. Mueller, R.C. and M.E. DeSimone, *Bringing Gender-Affirming Care to Primary Care: Use of a Multimodal Curriculum to Educate Nurse Practitioners and Nurse Practitioner Students.* Nurse Educ, 2023.
16. Oblea, P.N., et al., *Outcomes of LGBTQ culturally sensitive training among civilian and military healthcare personnel.* Journal of public health (Oxford, England), 2022.
17. Sherman, A.D.F., et al., *Transgender and gender diverse health education for future nurses: Students’ knowledge and attitudes.* Nurse Educ Today, 2021. **97**: p. 104690.
18. Thompson, H., et al., *Evaluation of a gender-affirming healthcare curriculum for second-year medical students.* Postgrad Med J, 2020. **96**(1139): p. 515-519.

### Section 6: Comfort outcome

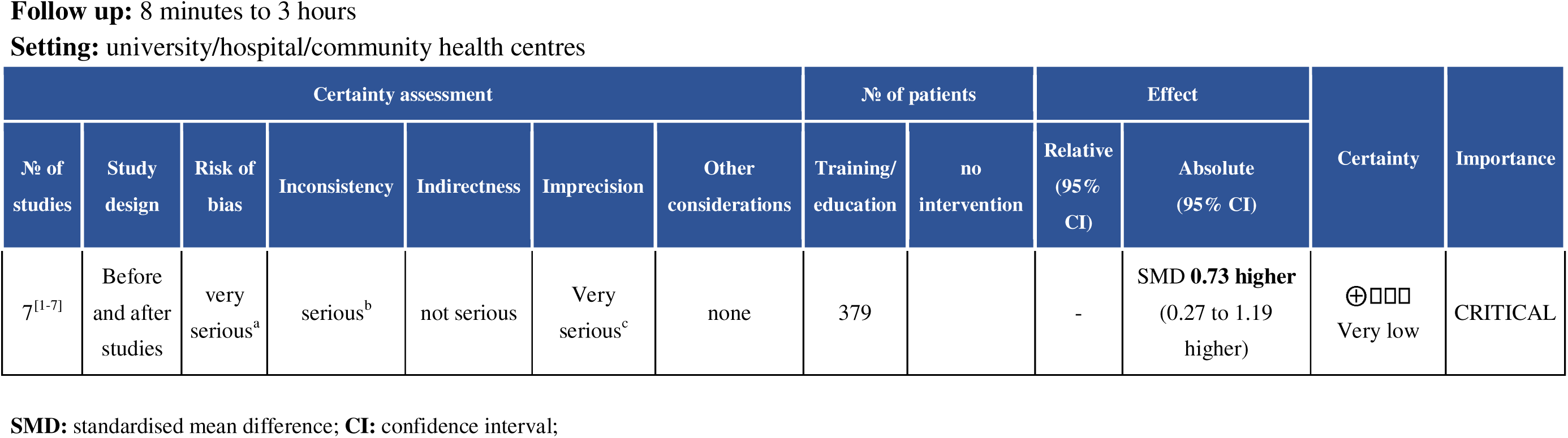

#### Explanations

a. Risk of bias: Downgraded by two levels because 3 ^[4, 6, 7]^ out of the 7 studies were assessed as high or critical risk of bias across multiple domains with the ROBINS-I tool, including bias due to confounding, bias due to selection of participants, bias due to missing data. The overall risk of bias was rated as very serious for this outcome.
b. b. Inconsistency: Downgraded by one level because the heterogeneity (I^2^=88.8%) between the studies was apparent both in terms of direction and magnitude of effect. We explored the inconsistency through subgroup analysis by sample size, by category of health worker, by setting and by intervention (e.g. self-learning, facilitated learning, mixed approach), but the inconsistency in magnitude of effect remains unexplained through subgroup analysis. This potentially reduces our certainty on the observed magnitude of effect.
c. Imprecision: Downgraded by two levels because the 95%CI of the estimated effect crossed the threshold of medium and large effect. The sample size is small. GRADE guidance suggests the sample size should be >/=800 for a continuous variable if the assumed small effect is 0.2 SMD; and if the overall sample size is between 240 to 400, downgrading by two levels is appropriate.

#### References

1. Allison, M.K., et al., *Community-Engaged Development, Implementation, and Evaluation of an Interprofessional Education Workshop on Gender-Affirming Care.* Transgend Health, 2019. **4**(1): p. 280-286.
2. Click, I.A., et al., *Transgender health education for medical students.* Clin Teach, 2020. **17**(2): p. 190-194.
3. Dale, V.H. and R. Philomin, *Educating for Quality Transgender Health Care: A Survey Study of Medical Students.* Educ Health (Abingdon), 2022. **35**(1): p. 31-34.
4. Linsenmeyer, W., et al., *Advancing Inclusion of Transgender Identities in Health Professional Education Programs: The Interprofessional Transgender Health Education Day.* J Allied Health, 2023. **52**(1): p. 24-31.
5. Pathoulas, J.T., et al., *Effectiveness of an Educational Intervention to Improve Medical Student Comfort and Familiarity With Providing Gender-Affirming Hormone Therapy.* Fam Med, 2021. **53**(1): p. 61-64.
6. Ruud, M.N., et al., *Health History Skills for Interprofessional Learners in Transgender and Nonbinary Populations.* J Midwifery Womens Health, 2021. **66**(6): p. 778-786.
7. Sherman, A.D.F., et al., *Transgender and gender diverse health education for future nurses: Students’ knowledge and attitudes.* Nurse Educ Today, 2021. **97**: p. 104690.

## Appendix 6: GRADECERQual Evidence Profile

| Num. | Summary of review finding | Study contributing to the review finding | GRADE-CERQual assessment | Explanation of GRADE-CERQual assessment |
| --- | --- | --- | --- | --- |
| Synthesized finding 1: Training structure and content. Pedagogical preferences of healthcare professionals and students regarding training on inclusive and gender-affirming care. |  |  |  |  |
| 1 | **ENGAGEMENT WITH TGD INDIVIDUALS**<br>Study participants appreciated training programs that included an element of engagement and interaction with transgender and gender-diverse individuals, including strategies such as interactions with standardised patients, involvement of the transgender community, use of complex case studies, patient panels, expert lectures, or the ability to draw upon faculty experiences. These approaches were felt to enhance communication skills, insight, and empathy and to challenge personal assumptions. | N=11 [1, 4, 6, 7, 8, 10, 11, 13, 15, 20, 23] | **Very Low** | Due to serious concerns regarding methodological limitation, minor concerns regarding coherence, serious concerns regarding relevance, and moderate concerns regarding adequacy |
| 2 | **INTERACTIVE TRAINING**<br>Participants expressed an appreciation for training approaches that were interactive, engaging, well-structured, easy to access and that offered opportunities for discussion. This included, for example, the use of standardised patient interactions and small group sessions. | N=9 [1, 2, 4, 6, 12, 13, 19, 23, 26] | **Low** | Due to minor concerns regarding methodological limitation, minor concerns regarding coherence, serious concerns regarding relevance, and minor concerns regarding adequacy |
| 3 | **CASE STUDIES**<br>Participants appreciated the use of real-life case studies as key tools for learning and discussion. Utilisation of case studies was considered to be more engaging and offered greater opportunities for learning than the didactic presentation of facts and statistics. | N=9 [5, 6, 13, 14, 19, 22, 23, 25, 30] | **Low** | Due to moderate concerns regarding methodological limitation, minor concerns regarding coherence, moderate concerns regarding relevance, and minor concerns regarding adequacy |
| 4 | **INTERSECTIONAL AND HOLISTIC**<br>Participants appreciated training content that adopted a holistic and intersectional approach to the topic that would also help to link theory to practice, including, for example, clarification of terminology, a focus on social issues, and illustrations of the complexity of issues. | N=7 [5, 9, 10-13, 29] | **Low** | Due to moderate concerns regarding methodological limitation, minor concerns regarding coherence, moderate concerns regarding relevance, and no concerns regarding adequacy |
| Synthesized finding 2: Impacts of training: Healthcare professionals and students highlighted the multifaceted values and gains of attending training programs. |  |  |  |  |
| 5 | CONSIDERATION OF PERSONAL POSITIONALITY<br>Participants felt that training programs could enhance and deepen their understanding, in particular, by helping them to consider their own positionality and developing a more empathic approach. | N=7 [4, 6, 7, 11-13, 19] | **Very Low** | Due to moderate concerns regarding methodological limitation, minor concerns regarding coherence, serious concerns regarding relevance, and minor concerns regarding adequacy |
| 6 | SELF-CONFIDENCE<br>Following training, participants reported feeling more confident in interacting with transgender and gender-diverse individuals, and thereby felt more able to adopt inclusive care approaches. | N=8 [3-7, 12, 14, 27] | **Low** | Due to moderate concerns regarding methodological limitation, minor concerns regarding coherence, moderate concerns regarding relevance, and minor concerns regarding adequacy |
| 7 | INCREASE IN KNOWLEDGE AND AWARENESS<br>Following training, participants reported having more knowledge and awareness of issues related to the care of transgender individuals and were able to identify areas where further education was still required. | N=11 [2, 4, 5, 7, 9, 12, 14, 23, 24, 26, 29] | **Low** | Due to moderate concerns regarding methodological limitation, minor concerns regarding coherence, moderate concerns regarding relevance, and minor concerns regarding adequacy |
| 8 | PROFESSIONAL GROWTH<br>Participants reported that training had promoted professional growth, in particular, by prompting them to reflect upon their own practices (including the need for further specialist education) and to reflect more consciously on gender diversity and health equity. | N=8 [2, 5, 6, 7, 11, 12, 16, 25] | **Low** | Due to moderate concerns regarding methodological limitation, minor concerns regarding coherence, moderate concerns regarding relevance, and minor concerns regarding adequacy |
| 9 | EMPATHY DEVELOPMENT<br>Participants reported that the training program enhanced their empathy and sensitivity to the specific situations and health conditions of transgender individuals, thereby building confidence in their ability to offer appropriate and personalised care. | N=9 [5, 6, 10, 16, 23-25, 27, 28] | **Very Low** | Due to serious concerns regarding methodological limitation, minor concerns regarding coherence, moderate concerns regarding relevance, and minor concerns regarding adequacy |
| 10 | COMFORT WITH TERMINOLOGY AND LANGUAGE<br>Following training, participants reported feeling more confident in their communication skills and more knowledgeable about the language and terminology specific to transgender issues within clinical practice. | N=15 [1, 3, 5, 11, 13, 14, 16, 18, 19, 21, 23, 24, 27-29] | **Very Low** | Due to serious concerns regarding methodological limitation, minor concerns regarding coherence, moderate concerns regarding relevance, and moderate concerns regarding adequacy |
| 11 | SELF EFFICACY<br>Participants who had undertaken training reported feeling more able to apply their knowledge to real-world clinical practice. | N=5<sup>[3, 15-18]</sup> | **Very Low** | Due to serious concerns regarding methodological limitation, minor concerns regarding coherence, serious concerns regarding relevance, and moderate concerns regarding adequacy |
| Synthesized finding 3: Factors that influenced implementation of training. |  |  |  |  |
| 12 | ANXIETY AND DISCOMFORT<br>Participants reported a range of issues that posed challenges for them. These related to personal beliefs, lack of knowledge or a social fear of 'saying the wrong thing' which were reported to create anxiety, discomfort or resistance among the trainees. | N=13<sup>[2-7, 11, 12, 15, 17, 19, 20, 21]</sup> | **Low** | Due to moderate concerns regarding methodological limitation, minor concerns regarding coherence, moderate concerns regarding relevance, and minor concerns regarding adequacy |
| 13 | SAFE SPACE<br>Participants reported the need for the implementation of training programs to be based around strategies that would create safe spaces for learning and discussion. | N=4<sup>[6, 11, 12, 17]</sup> | **Very Low** | Due to moderate concerns regarding methodological limitation, minor concerns regarding coherence, serious concerns regarding relevance, and minor concerns regarding adequacy |
| Synthesized finding 4: Suggestions made by healthcare professionals on appropriate training interventions on inclusive and/or gender-affirming care. |  |  |  |  |
| 14 | LONGER, INTERACTIVE, AND PRACTICAL TRAINING<br>In evaluations of training, participants called for the structure of training programs to be longer and to include more frequent sessions, more interactive sessions (such as standardised patient interactions), and practical skill exercises. | N=5<sup>[1, 8, 10, 13, 14]</sup> | **Very Low** | Due to serious concerns regarding methodological limitation, minor concerns regarding coherence, serious concerns regarding relevance, and moderate concerns regarding adequacy |
| 15 | DIVERSITY IN TRAINING DELIVERY<br>In evaluations of training, participants suggested that there was a need for training delivery to incorporate diverse expertise in training programs, such as transgender community members, clinical psychologists, and representation of gender-diverse people of colour. | N=3<sup>[6, 11, 19]</sup> | **Low** | Due to minor concerns regarding methodological limitation, minor concerns regarding coherence, serious concerns regarding relevance, and minor concerns regarding adequacy |
| 16 | CONTEXT-SPECIFIC AND HOLISTIC<br>In evaluations of training, participants suggested a need for context-specific and holistic training content. | N=12<sup>[1, 2, 6, 11-14, 16, 19, 21, 22, 29]</sup> | **Very Low** | Due to serious concerns regarding methodological limitation, minor concerns regarding coherence, serious concerns regarding relevance, and moderate concerns regarding adequacy |
| 17 | INTEGRATE INTO STANDARD TRAINING<br>In evaluations of training, participants suggested that it may be advisable to integrate the training into ongoing faculty development and the broader clinical skills curriculum for continuous learning and application. | N=3<sup>[2, 4, 11]</sup> | **Low** | Due to minor concerns regarding methodological limitation, minor concerns regarding coherence, serious concerns regarding relevance, and minor concerns regarding adequacy |
| 18 | ONGOING ACCESSIBLE LEARNING<br>In evaluations of training, participants stressed the importance of continuous and accessible learning by expanding course availability, nurturing safe and inclusive environments for open discussions, and emphasizing the importance of ongoing education. | N=6<sup>[8, 11, 14, 20, 25, 29]</sup> | **Very Low** | Due to moderate concerns regarding methodological limitation, minor concerns regarding coherence, serious concerns regarding relevance, and moderate concerns regarding adequacy |

### Reference

1. Stumbar SE, Garba NA, de la Cruz M, Bhoite P, Uchiyama E. Integration of Sex and Gender Minority Standardized Patients into a Workshop on Gender-Inclusive Patient Care: Exploring Medical Student Perspectives. South Med J. 2022;115(9):722-726.
2. Mackie G, Patlamazoglou L, Lambert K. School psychologists’ perceptions of transgender training and education: An Australian qualitative investigation. Psychol Sex Orientat Gend Divers. 2022; 10(4):699-710.
3. Kreines FM, Quinn GP, Cardamone S, Pi GE, Cook T, Salas-Humara C, Fino E, Shaw J. Training clinicians in culturally relevant care: a curriculum to improve knowledge and comfort with the transgender and gender diverse population. J Assist Reprod Genet. 2022;39(12):2755-2766.
4. Treharne GJ, Blakey AG, Graham K, Carrington SD, McLachlan LA, Withey-Rila C, Pearman-Beres L, Anderson L. Perspectives on expertise in teaching about transgender healthcare: A focus group study with health professional programme teaching staff and transgender community members. Int J Transgend Health. 2021;23(3):334-354.
5. Jeon SY, Yoon HB, Park JE, Lee SY, Yoon JW. A qualitative study on the internal response of medical students during the transgender healthcare education: a focus on professional identity. Korean J Med Educ. 2022;34(4):281-297.
6. Hayward M, Treharne GJ. Clinical psychology students’ perspectives on involving transgender community members in teaching activities within their training in Aotearoa New Zealand. Psychol Sex Orientat Gend Divers. 2022;9(3):309.
7. Altmiller G, Wilson C, Jimenez FA, Perron T. Impact of a Virtual Patient Simulation on Nursing Students’ Attitudes of Transgender Care. Nurse Educ. 2023;48(3):131-136.
8. Huser N, Hulswit BB, Koeller DR, Yashar BM. Improving gender-affirming care in genetic counseling: Using educational tools that amplify transgender and/or gender non-binary community voices. J Genet Couns. 2022;31(5):1102-1112.
9. Biro L, Tang H, Tang G, Song K, Nyhof-Young J. Introducing sexual and gender minority health: Medical students develop and evaluate an LGBT+ infographic. Am J Sex Educ. 2021;16(2):181-98.
10. Frazier CC, Nguyen TL, Gates BJ, McKeirnan KC. Teaching transgender patient care to student pharmacists. Curr Pharm Teach Learn. 2021;13(12):1611-1618.
11. Biro L, Song K, Nyhof-Young J. First year medical student experiences with a clinical skills seminar emphasizing sexual and gender minority population complexity. Can Med Educ J. 2021;12(2):e11-e20.
12. Arias T, Greaves B, McArdle J, Rayment H, Walker S. Cultivating awareness of sexual and gender diversity in a midwifery curriculum. Midwifery. 2021;101:103050.
13. Vijapura C, Tobler J, Wahab RA, Smith ML, Brown AL, Pickle S, Stryker SD, Spalluto LB, England E, Kanfi A. Resident Attitudes and Experiences with a Novel Radiology-based Transgender Curriculum: A Qualitative Study. Acad Radiol. 2023:S1076-6332(23)00068-5.
14. McMillian-Bohler J, Gedzyk-Neiman S, Hepler B, May JT, Koch A. The Power of a Story: Enhancing Students’ Empathy for Transgender Pregnant Men. J Nurs Educ. 2022;61(8):489-492.
15. Luctkar-Flude M, Ziegler E, Foronda C, Walker S, Tyerman J. Impact of virtual simulation games to promote cultural humility regarding the care of sexual and gender diverse persons: A multi-site pilot study. Clin Simul Nurs. 2022;71:146-58.
16. Luctkar-Flude M, Tyerman J, Ziegler E, Walker S, Carroll B. Usability testing of the sexual orientation and gender identity nursing education eLearning toolkit and virtual simulation games. Teach Learn Nurs. 2021;16(4):321-5.
17. Koch A, Ritz M, Morrow A, Grier K, McMillian-Bohler JM. Role-play simulation to teach nursing students how to provide culturally sensitive care to transgender patients. Nurse Educ Pract. 2021;54:103123.
18. Craig SL, Iacono G, Austin A, et al. The role of facilitator training in intervention delivery: Preparing clinicians to deliver AFFIRMative group cognitive behavioral therapy to sexual and gender minority youth. J Gay Les Social Serv. 2021;33(1): 56-77.
19. Bi S, Vela M B, Nathan A G, et al. Teaching intersectionality of sexual orientation, gender identity, and race/ethnicity in a health disparities course. MedEdPORTAL, 2020;16: 10970.
20. Charbonneau G. The Use of Simulation with the School of Nursing and Health Professions (SONHP) Prelicensure Students to Support Affirming Practice with Transgender Communities. University of San Francisco, 2022.
21. Jadwin-Cakmak L, Bauermeister JA, Cutler JM, Loveluck J, Kazaleh Sirdenis T, Fessler KB, Popoff EE, Benton A, Pomerantz NF, Gotts Atkins SL, Springer T, Harper GW. The Health Access Initiative: A Training and Technical Assistance Program to Improve Health Care for Sexual and Gender Minority Youth. J Adolesc Health. 2020;67(1):115-122.
22. Du Mont J, Saad M, Kosa SD, Kia H, Macdonald S. Providing trans-affirming care for sexual assault survivors: An evaluation of a novel curriculum for forensic nurses. Nurse Educ Today. 2020;93:104541.
23. Chaudhary S, Lindsay D, Ray R, Glass BD. Evaluation of a transgender health training program for pharmacists and pharmacy students in Australia: A pre-post study. Explor Res Clin Soc Pharm. 2023;13:100394.
24. Ness M, Bradley CS, Flaten CB. Gender Affirming Postop Care Simulation for Prelicensure Nursing Students: A Pilot Project. Clinical Simulation in Nursing. 2023;81:101432.
25. Linsenmeyer W, Heiden-Rootes K, Drallmeier T, Rahman R, Buxbaum E, Walcott K, Rosen W, Gombos B. The power to help or harm: student perceptions of transgender health education using a qualitative approach. BMC Medical Education. 2023;23(1):836.
26. Ruud MN, Demma JM, Woll A, Miller JM, Hoffman S, Avery MD. Health history skills for interprofessional learners in transgender and nonbinary populations. Journal of Midwifery & Women’s Health. 2021;66(6):778-86.
27. Lacombe-Duncan A, Logie CH, Persad Y, et al. Implementation and evaluation of the ‘Transgender Education for Affirmative and Competent HIV and Healthcare (TEACHH)’ provider education pilot. BMC Med Educ. 2021;21(1):561.
28. Willson MN, Frazier CC, McKeirnan KC. Training Student Pharmacists in Microaggressions and Gender Inclusive Communication. Am J Pharm Educ. 2024;88(3):100676.
29. Corey D. Effectiveness of Transgender Health Training on Healthcare Students’ Knowledge, Attitudes, and Perceived Competency Providing Gender-Affirming Healthcare. Northern Arizona University, 2023.
30. Karsenti N, Chambers J, Espinosa A. Effects of SGM Education for Undergraduate Medical Students in a Canadian Context. Med Sci Educ. 2023;33(4):813-824.

**Table 2.**
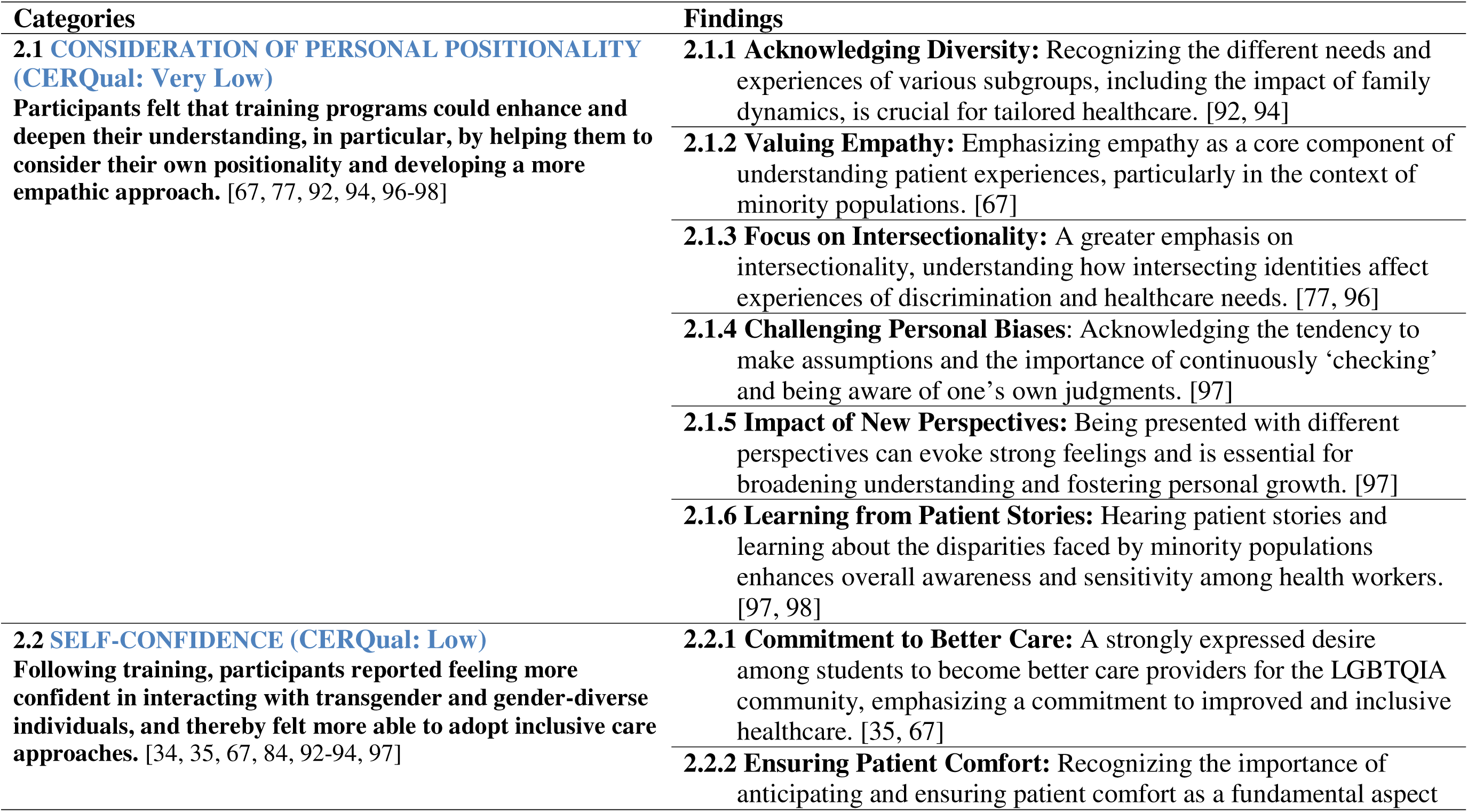
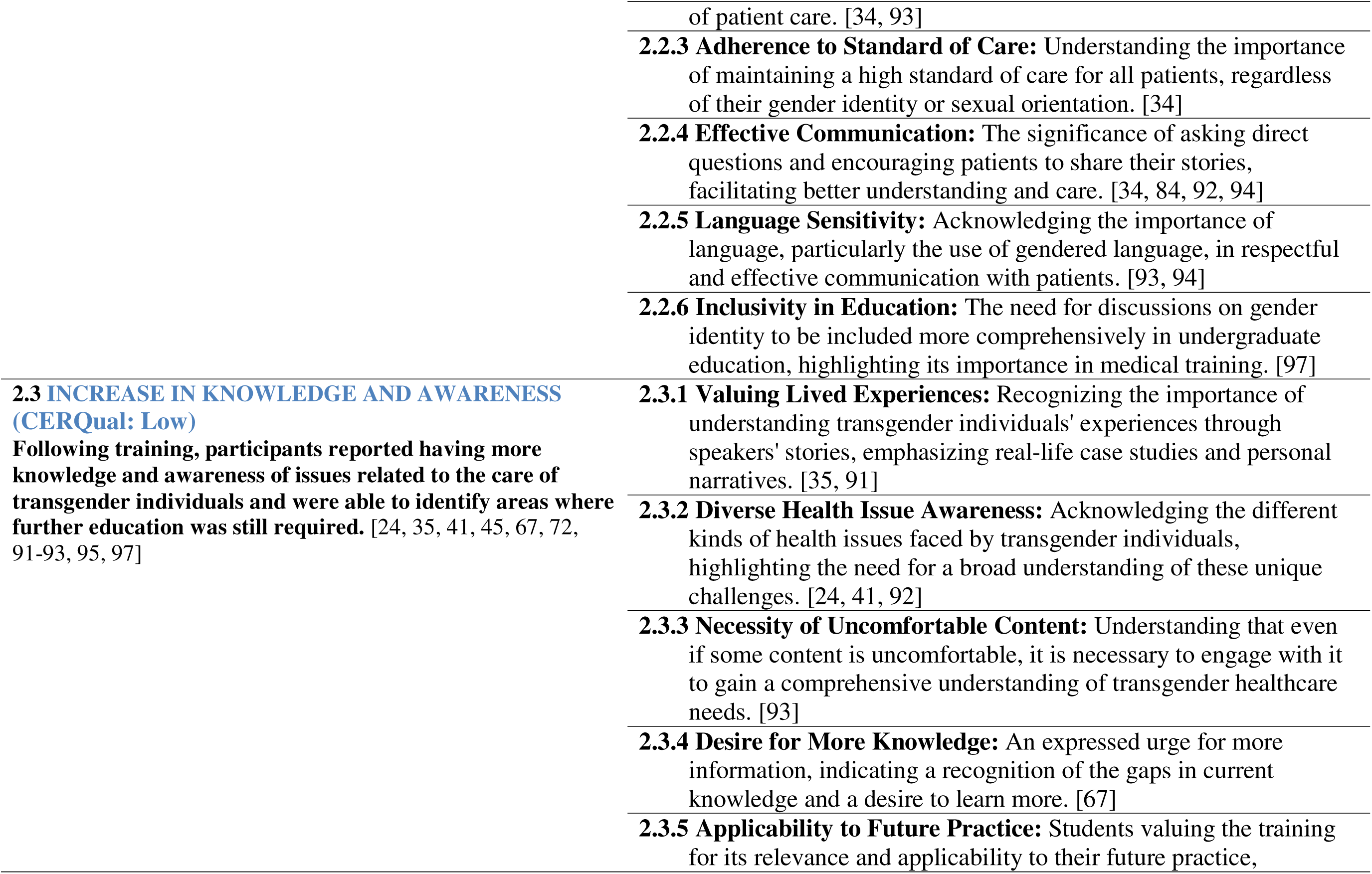
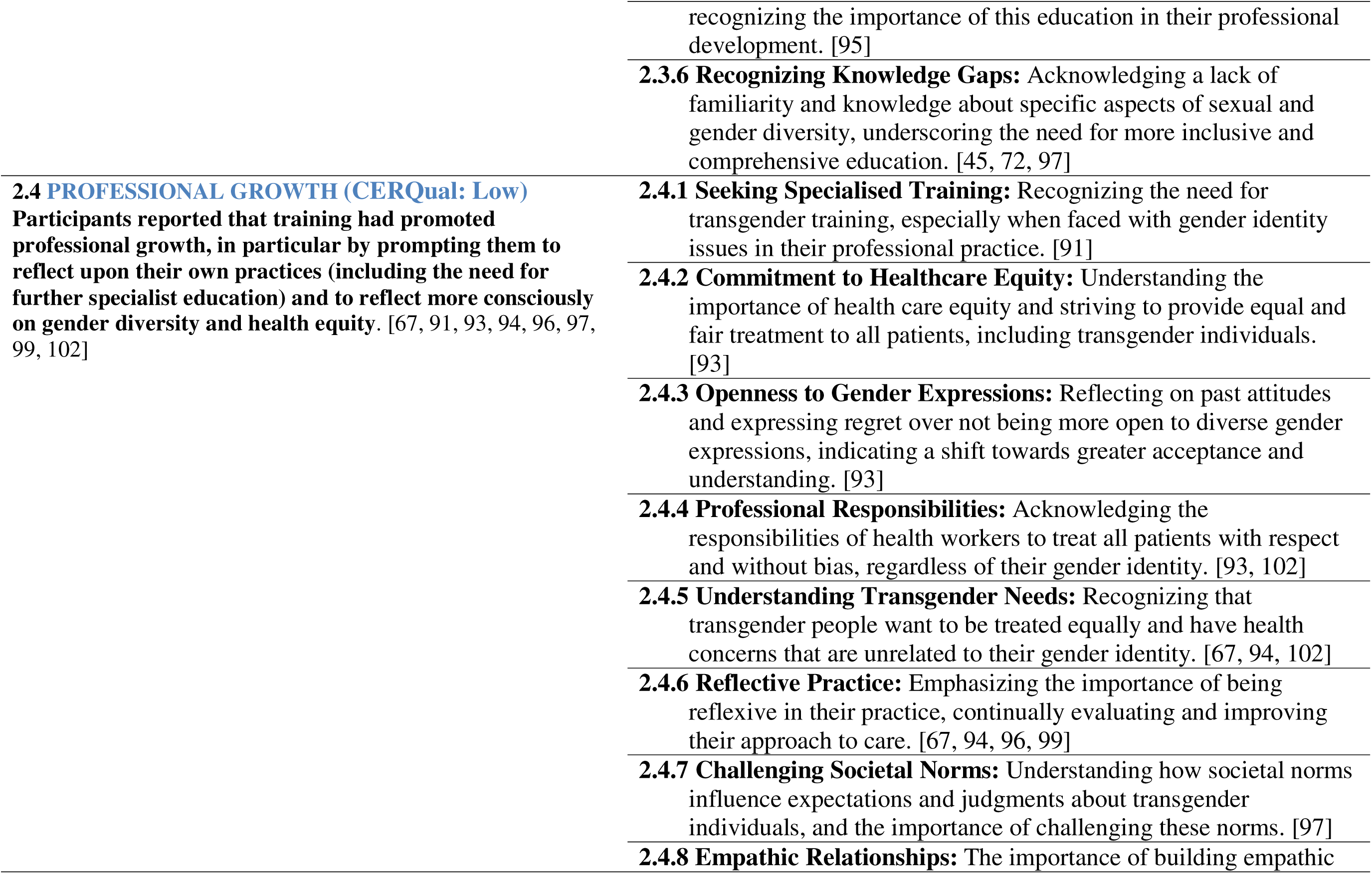
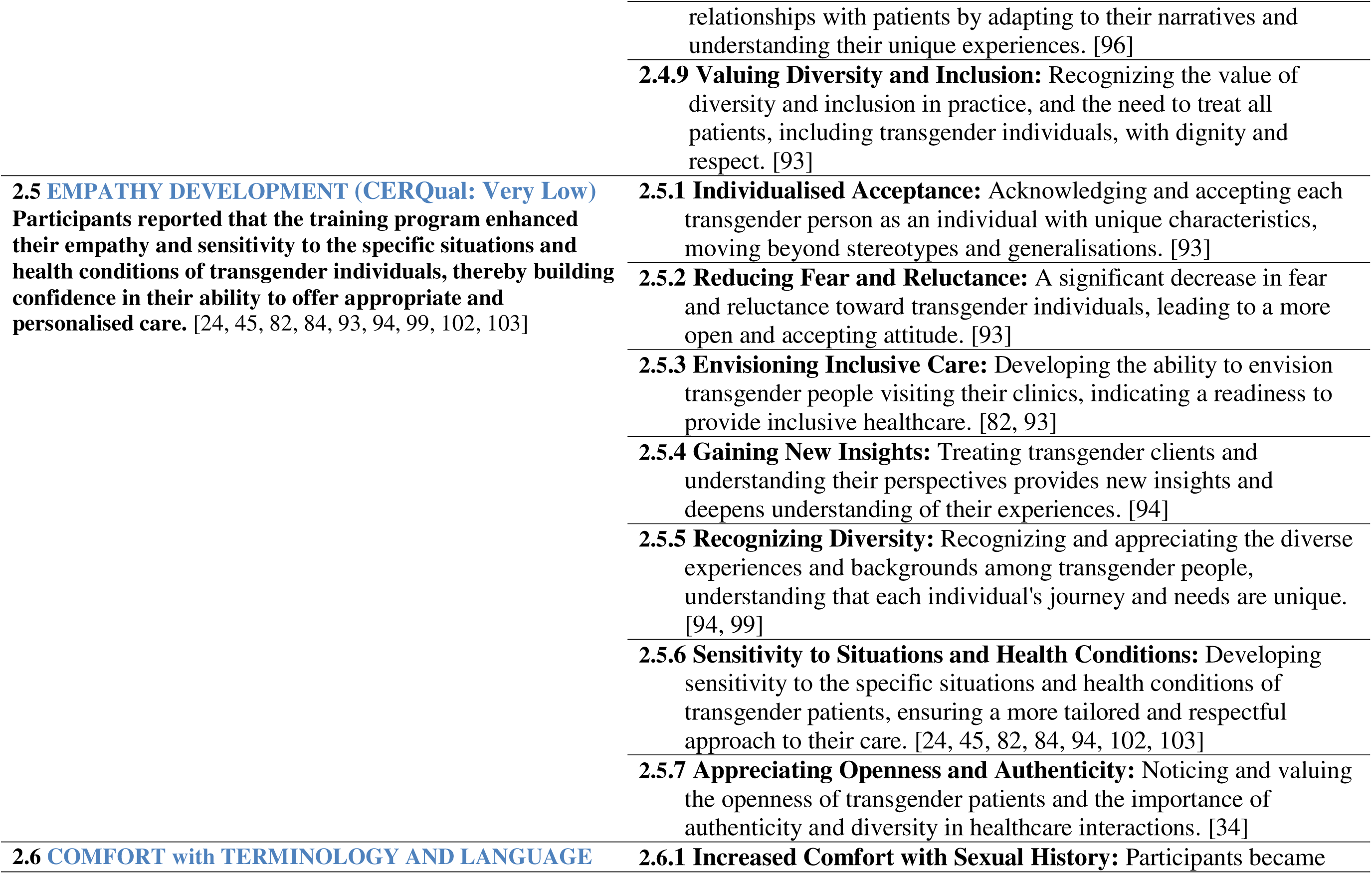
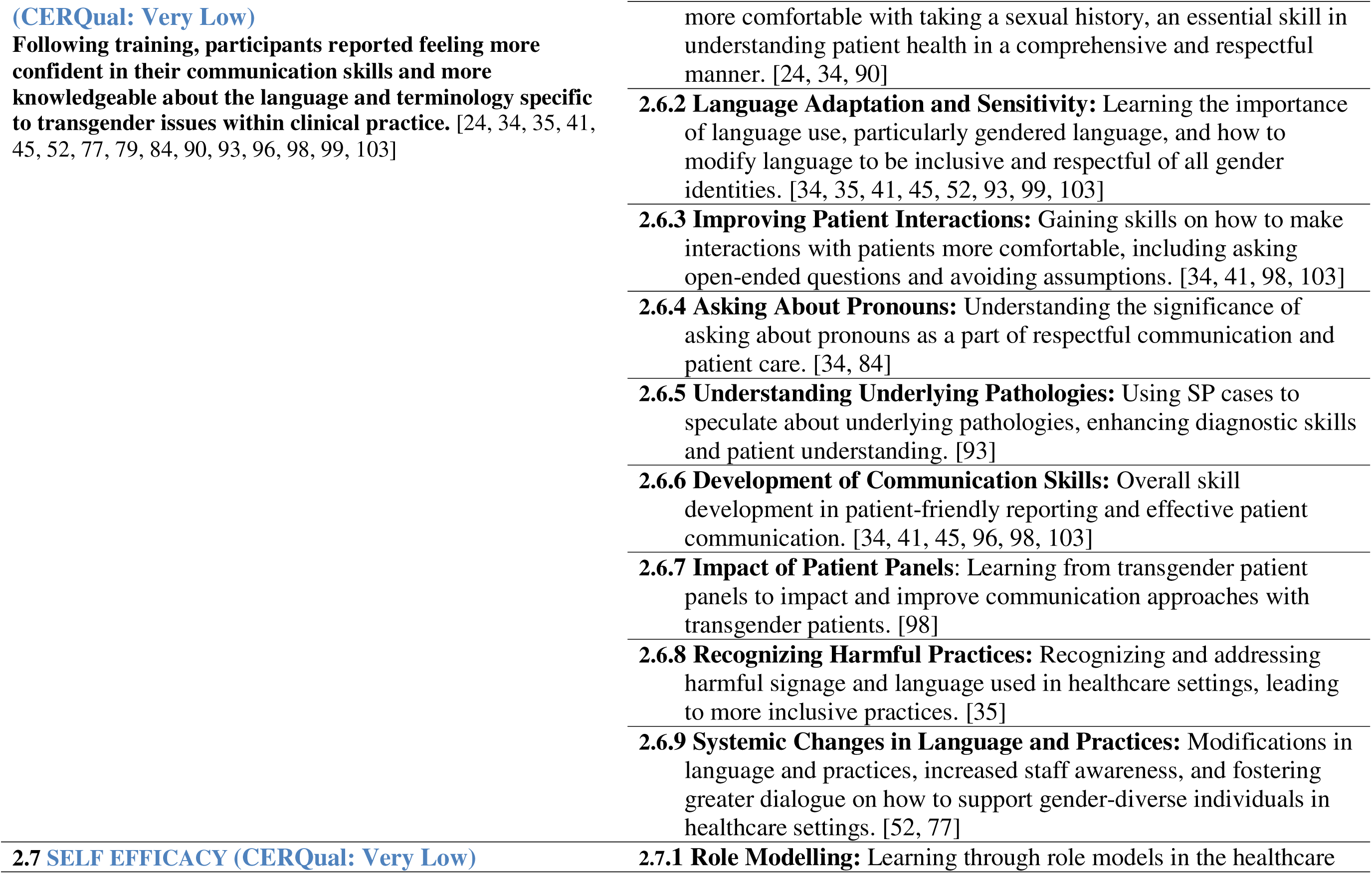
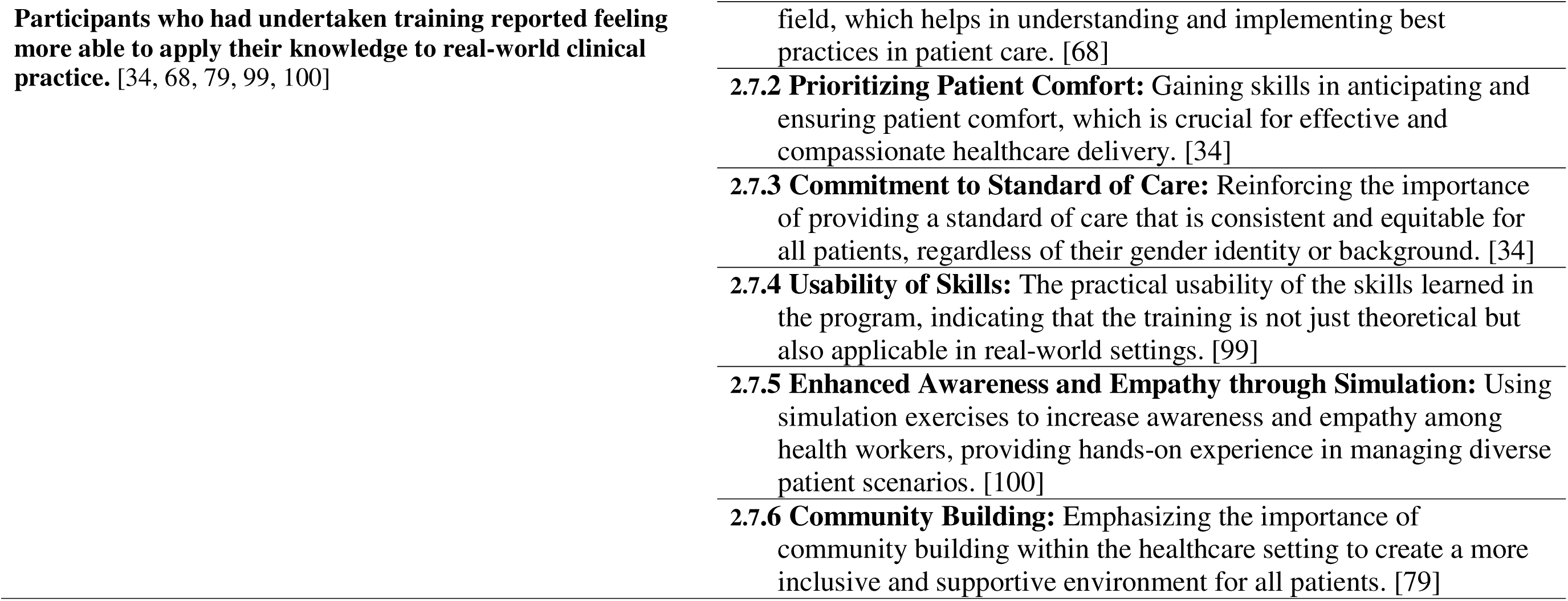
Categories and findings for synthesised finding 2.

**Table 3.** Categories and findings for synthesised finding 3.

| Categories | Findings |
| --- | --- |
| **3.1 ANXIETY AND DISCOMFORT (CERQual: Low)**<br>**Participants reported a range of issues that posed challenges for them. These related to personal beliefs, lack of knowledge, or a social fear of ‘saying the wrong thing’ which were reported to create anxiety, discomfort, or resistance among the trainees. [34, 52, 67, 68, 77, 91-94, 96, 97, 100, 101]** | **3.1.1 Anxiety and Discomfort:** Participants felt anxiety and discomfort, often stemming from personal experiences, religious beliefs, or a general lack of exposure to transgender issues. [34, 93, 96, 101]<br>**3.1.2 Knowledge Gaps:** There were significant gaps in knowledge about transgender care among participants. [77, 91, 92, 94, 100, 101]<br>**3.1.3 Logistical Challenges:** Practical and logistical challenges in the training process, including limited experience and intentional avoidance of the subject. [94]<br>**3.1.4 Risk Aversion:** Less supported participants were more risk-averse, potentially due to insecurity and fear of making mistakes. [96, 97]<br>**3.1.5 Lack of Awareness:** A general lack of awareness about the long history of discrimination against transgender individuals and non-inclusive hospital policies. [100]<br>**3.1.6 Addressing Specific Needs:** Recognizing and addressing the specific needs of transgender individuals, which can be challenging due to varying and complex requirements. [92]<br>**3.1.7 Difficulty in Changing Mindsets:** Acknowledging the challenge in altering pre-existing mindsets and biases, both personally and among peers. [93]<br>**3.1.8 Anticipating Peer Resistance:** Expecting and dealing with potential refusal or resistance from fellow students towards transgender education and issues. [93] |
|  | **3.1.9 Media Influence:** The impact of media exposure on perceptions and understanding of transgender issues, which can sometimes lead to confusion or misinformation. [93] |
|  | **3.1.10 Complexity of Biological Education:** Understanding the biological aspects of transgender identity can be confusing, requiring clear and comprehensive educational approaches. [67] |
|  | **3.1.11 Concerns Over Topic Sensitivity:** Worries about inappropriately combining topics like sexually transmitted infections (STIs) with being a gender-diverse individual, and the need for sensitive handling of these subjects. [96] |
|  | **3.1.12 Navigating Complex Cases:** The process of working through complex transgender cases is likened to solving puzzles, indicating both the challenge and the intellectual engagement involved. [96] |
|  | **3.1.13 Feedback and Communication Issues:** Addressing feedback and the tension between the desires of site liaisons and some training participants, highlighting communication challenges in the training environment. [52, 68] |
| **3.2 SAFE SPACE (CERQual: Very Low)**<br>**Participants reported the need for the implementation of training programs to be based around strategies that would create safe spaces for learning and discussion. [94, 96, 97, 100]** | **3.2.1 Motivation for Learning:** Developing clinical competence and societal responsibility for the care of gender-diverse individuals served as strong motivators for learning. [96] |
|  | **3.2.2 Open-mindedness:** Participants who were more open-minded tended to engage more effectively with the training. [96] |
|  | **3.2.3 Support Systems:** The support of family of origin and other support systems played a crucial role in facilitating learning. [97] |
|  | **3.2.4 Increased Knowledge and Experience:** Gaining more knowledge |
|  | and experience, particularly in areas like asking for patient pronouns, helped in overcoming barriers. [94, 100] |
|  | **3.2.5 Addressing Insecurities:** Recognizing and addressing insecurities and fears about making mistakes was key to facilitating learning and confidence. [100] |

**Table 4.**
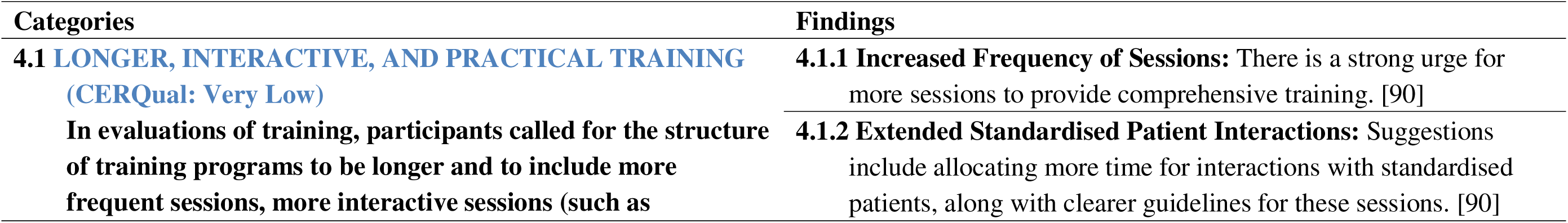
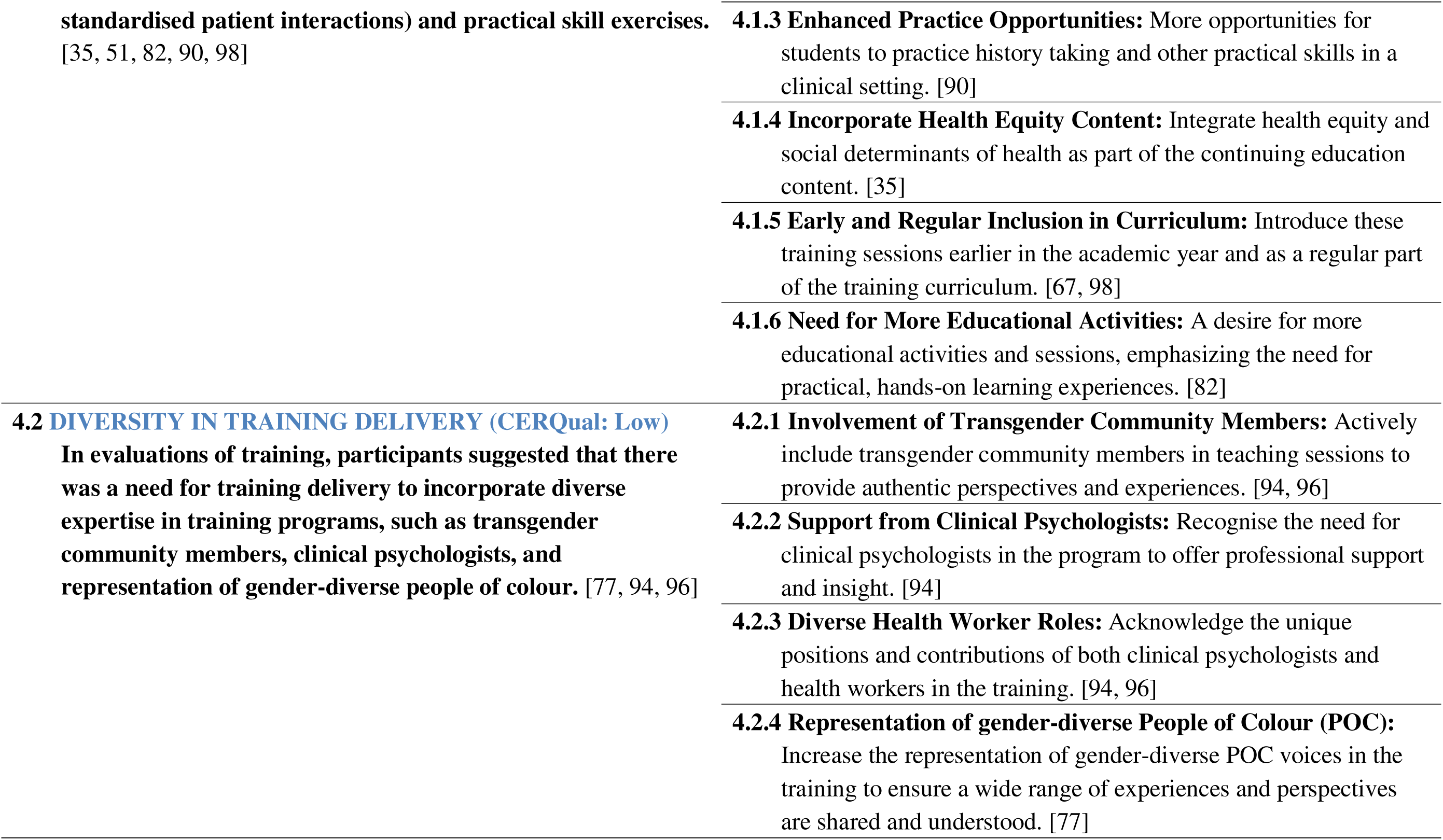
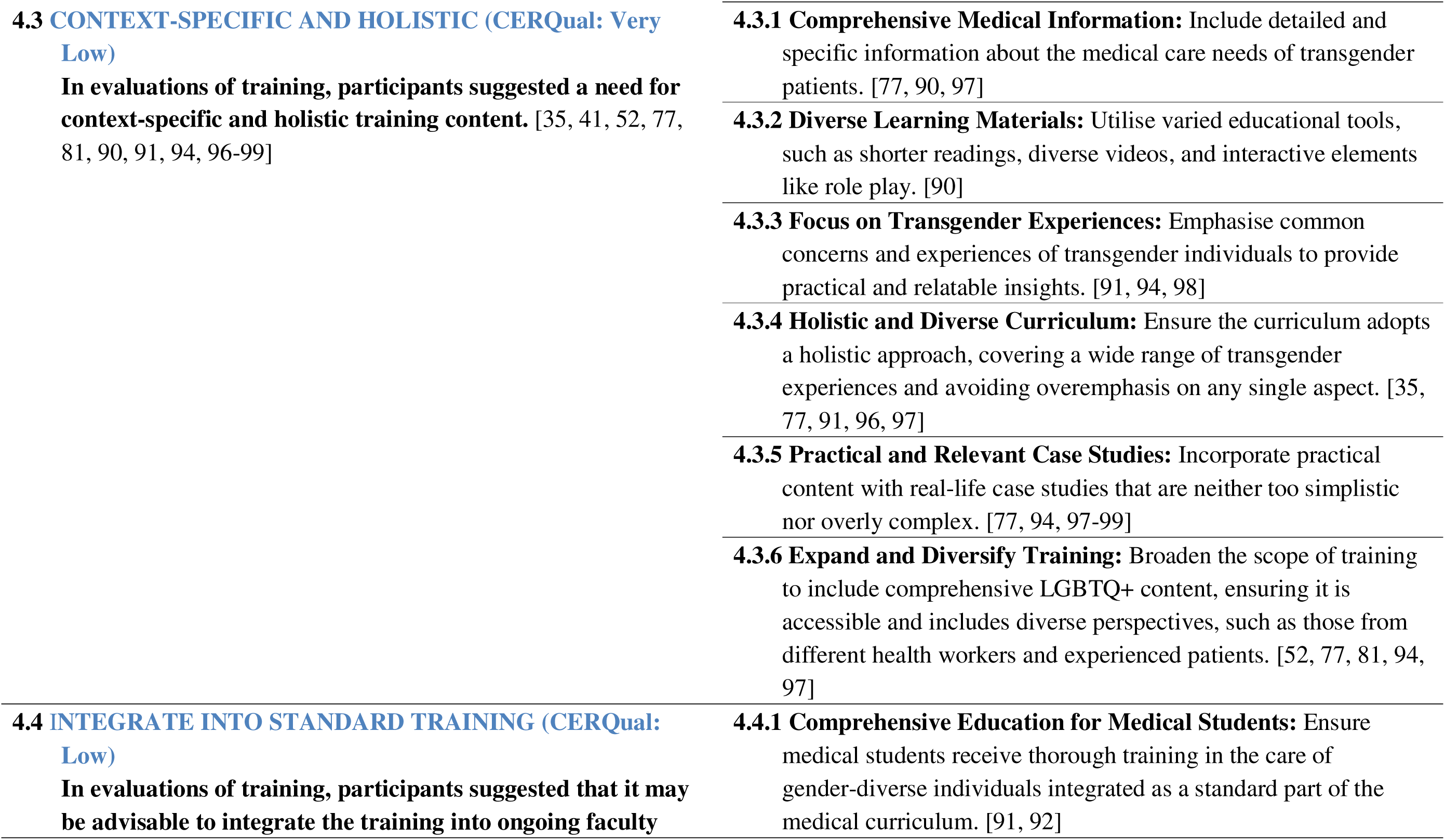
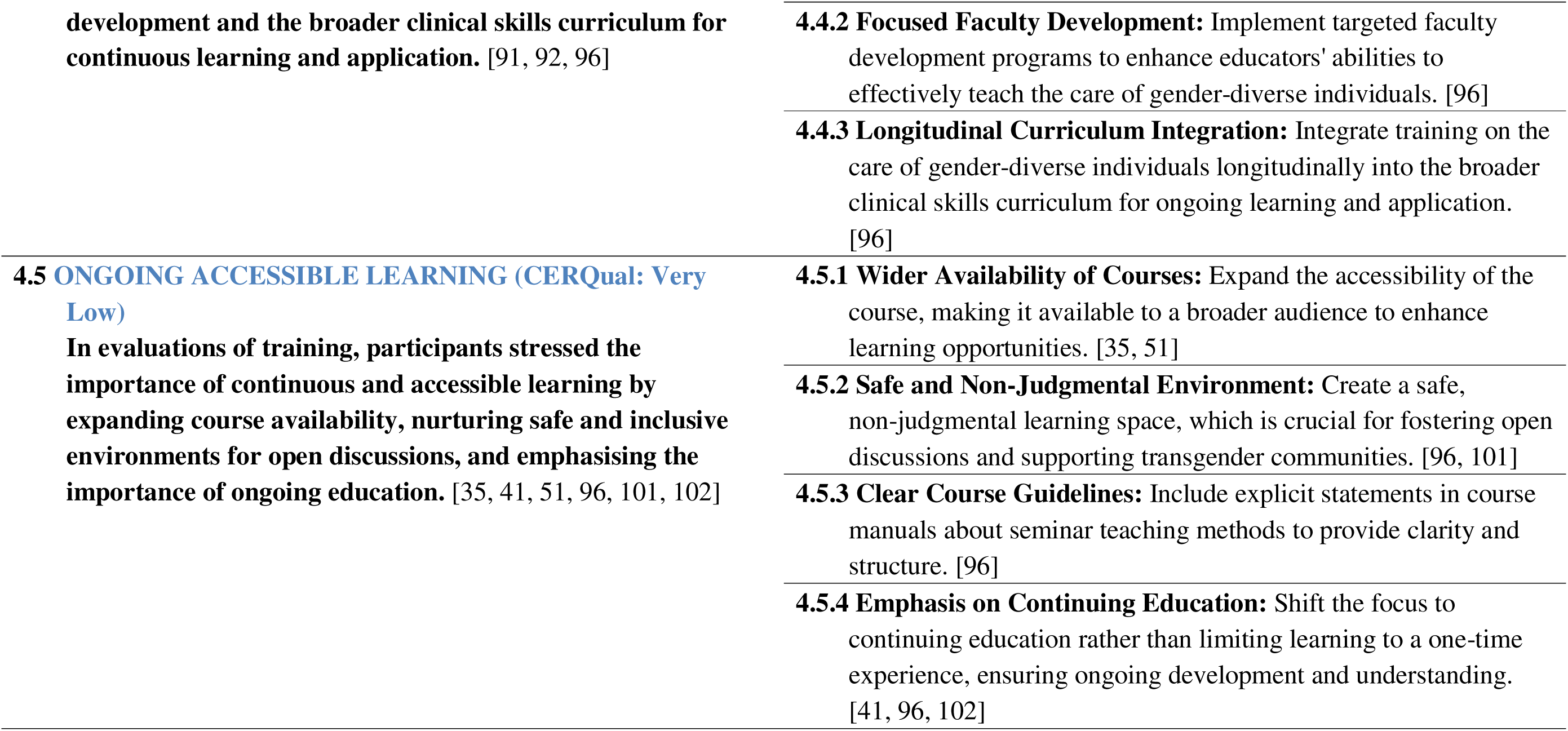
Categories and findings for synthesised finding 4.

## Appendix 7: Outcomes Figures

**Figure 3.**
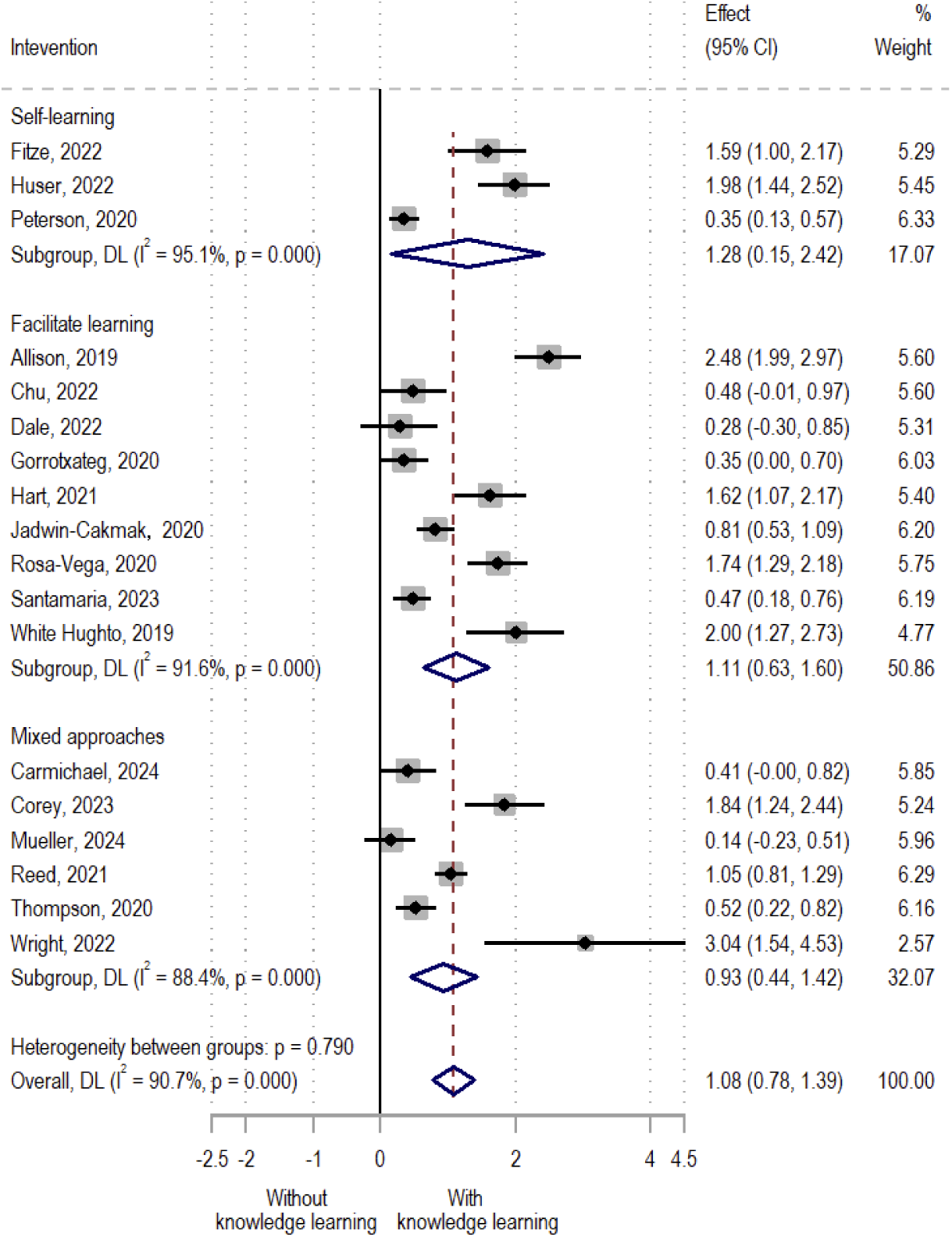
Change in competence – knowledge (subgroup analysis by intervention)

**Figure 4:**
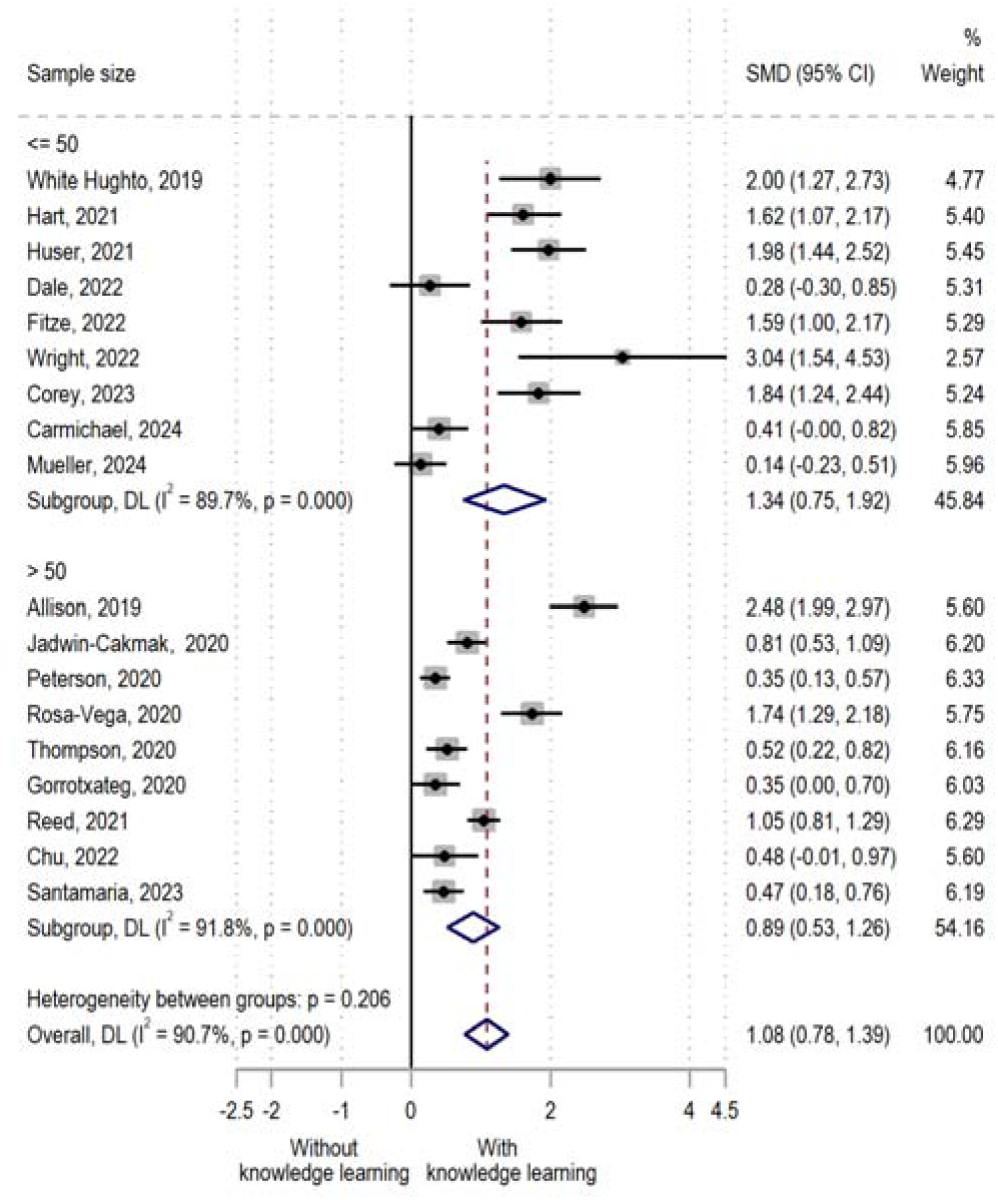
Change in competence – knowledge (subgroup analysis by sample size)

**Figure 5:**
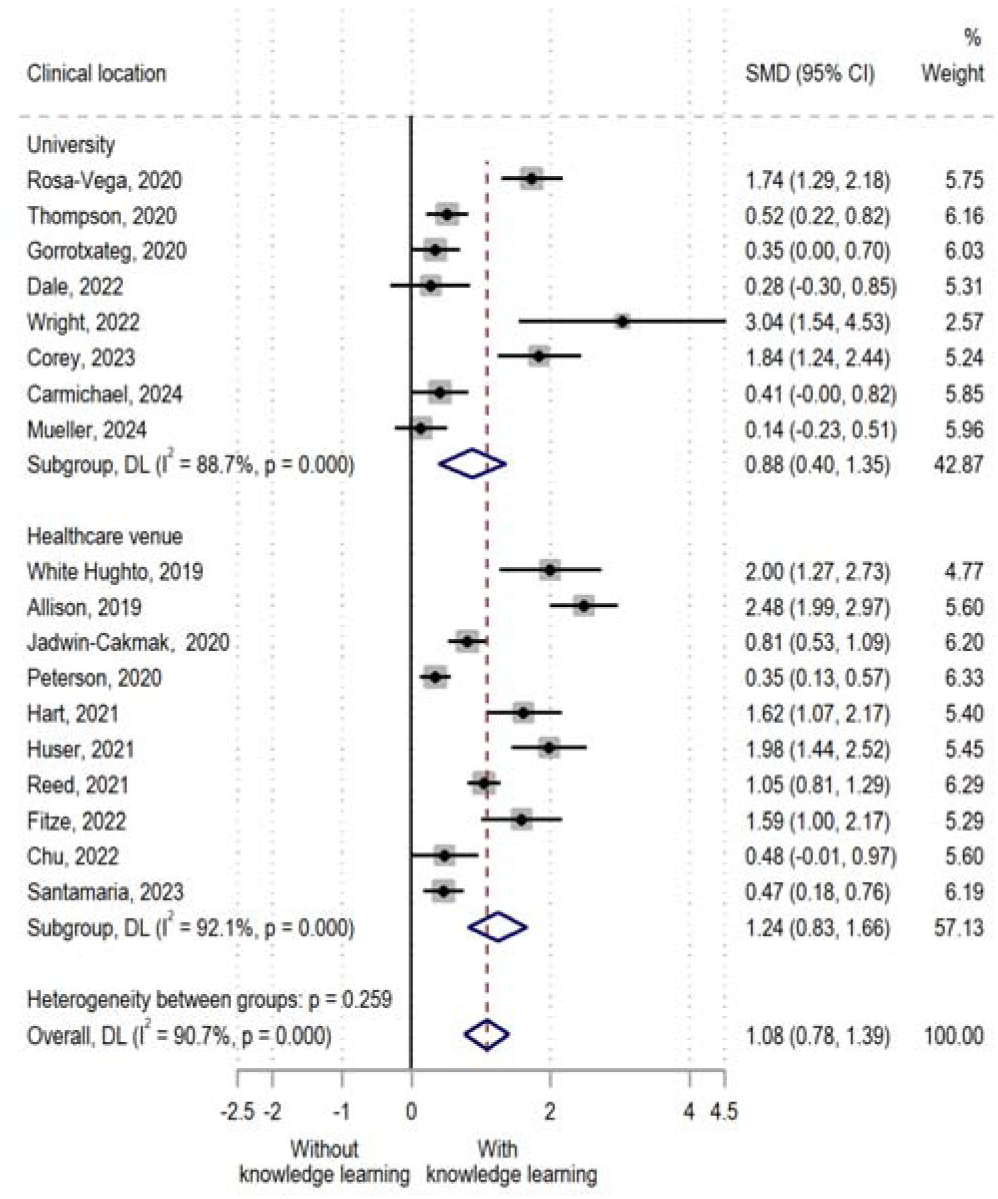
Change in competence – knowledge (Subgroup analysis by setting)

**Figure 6:**
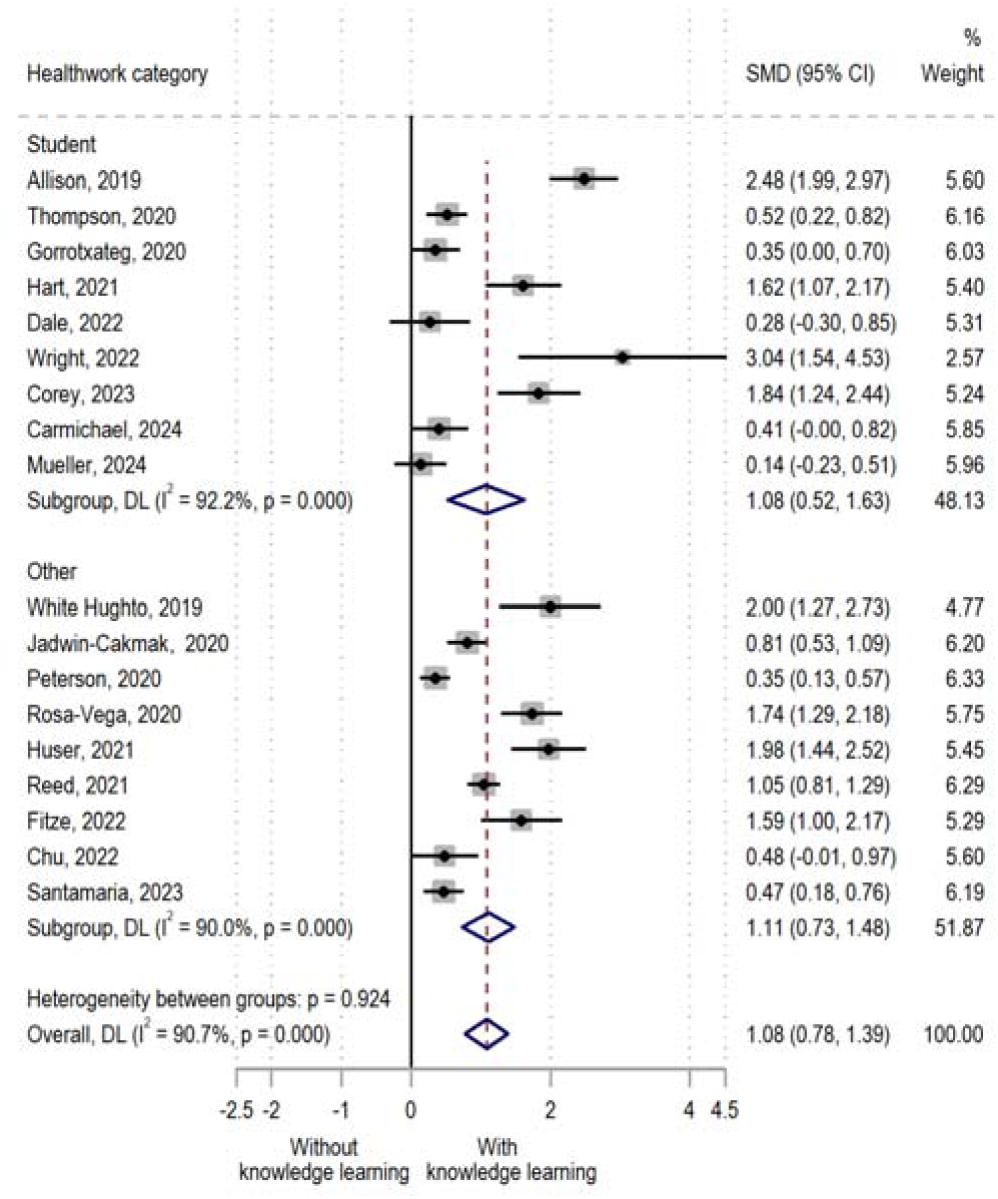
Change in competence – knowledge (Subgroup analysis by health worker category)

**Figure 7:**
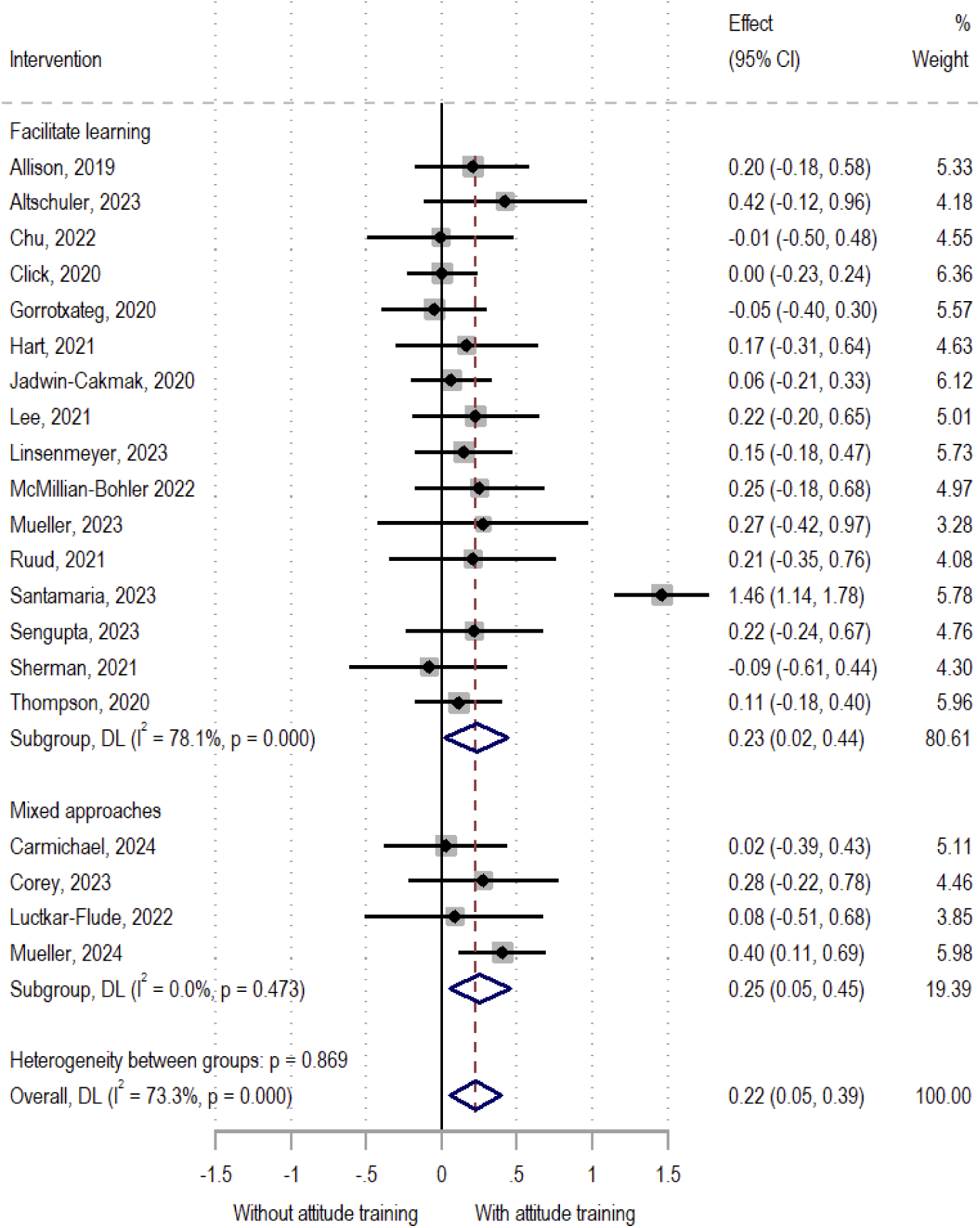
Change of competence – attitude (subgroup analysis by intervention)

**Figure 8:**
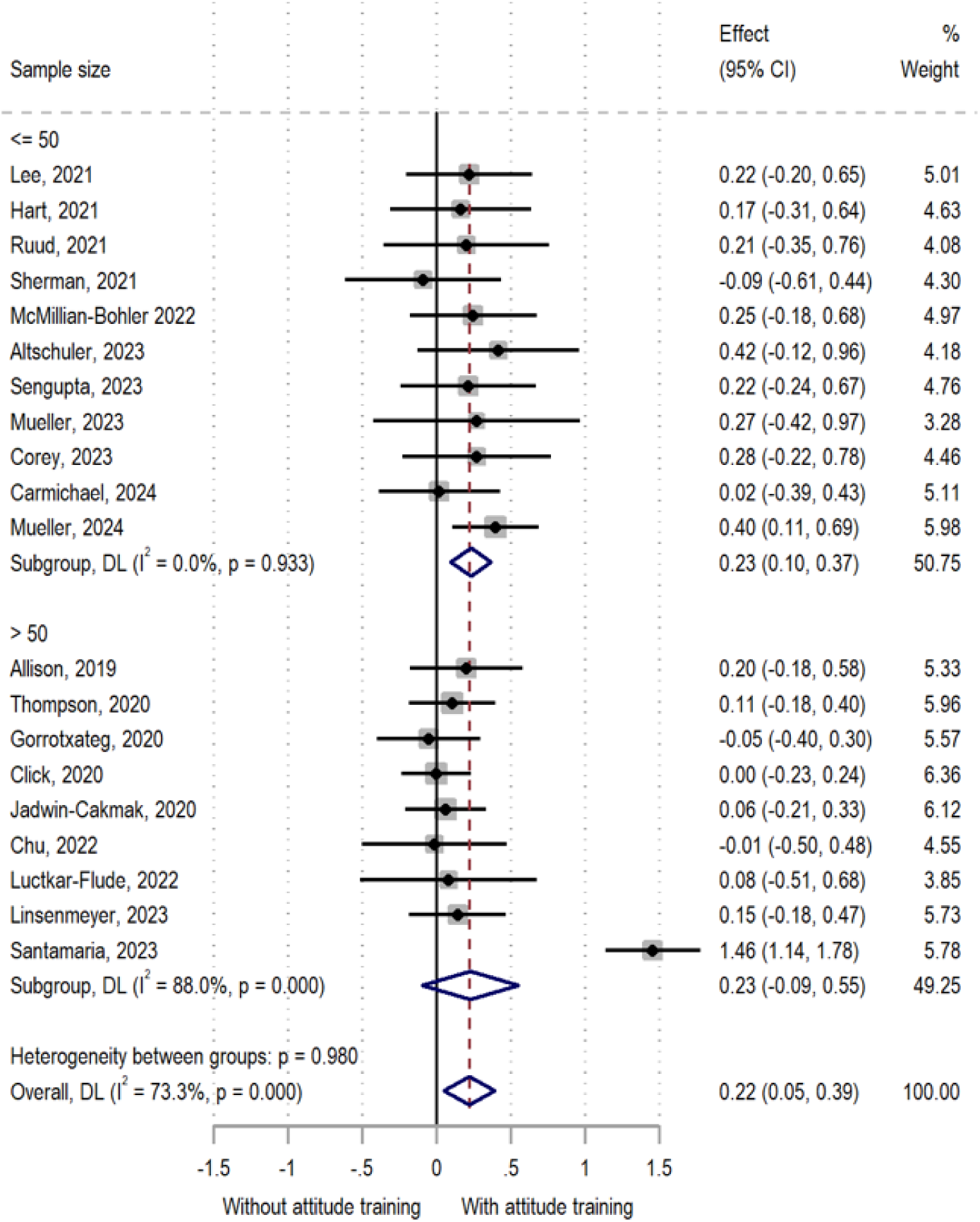
Change of competence – attitude (subgroup analysis by sample size)

**Figure 9:**
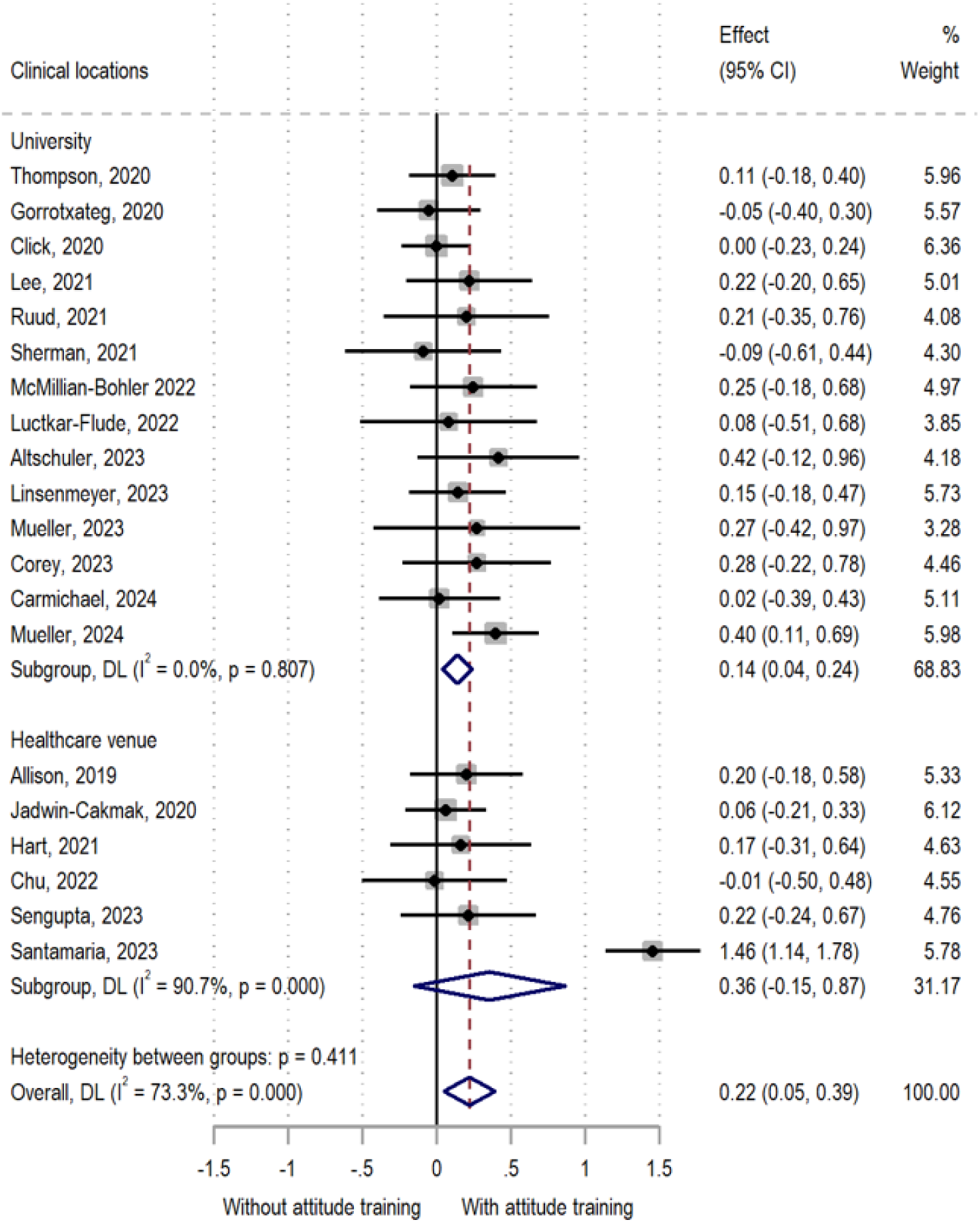
Change of competence – attitude (subgroup analysis by setting)

**Figure 10:**
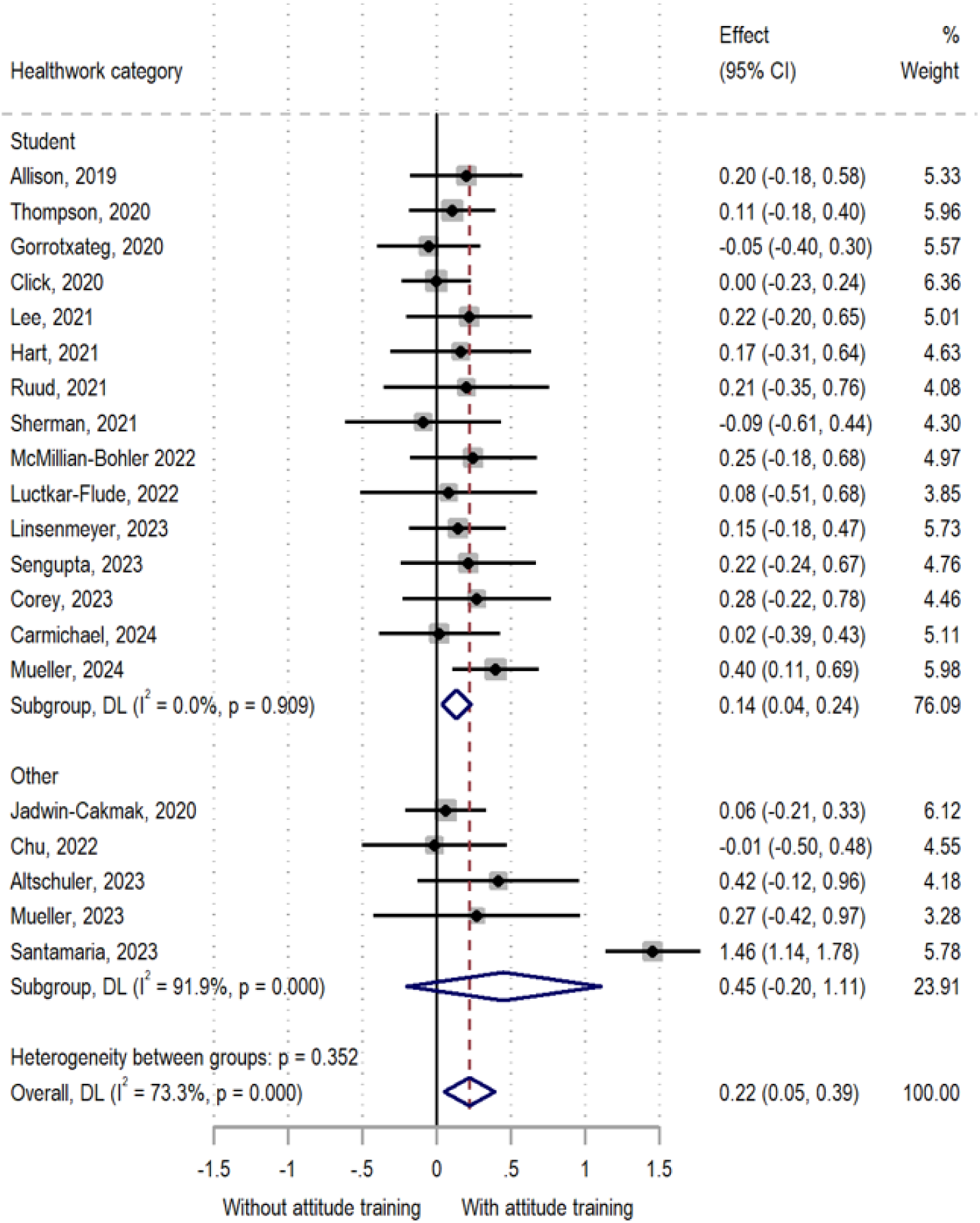
Change of competence – attitude (subgroup analysis by heath worker category)

**Figure 11:**
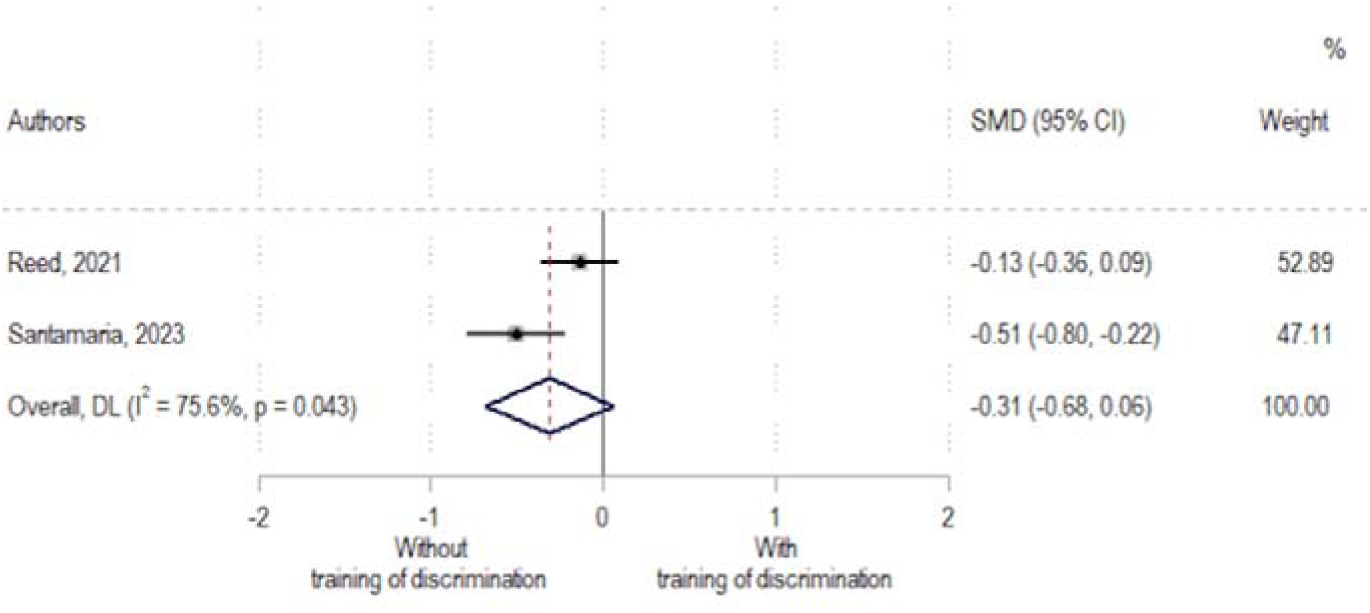
Discrimination.

**Figure 12:**
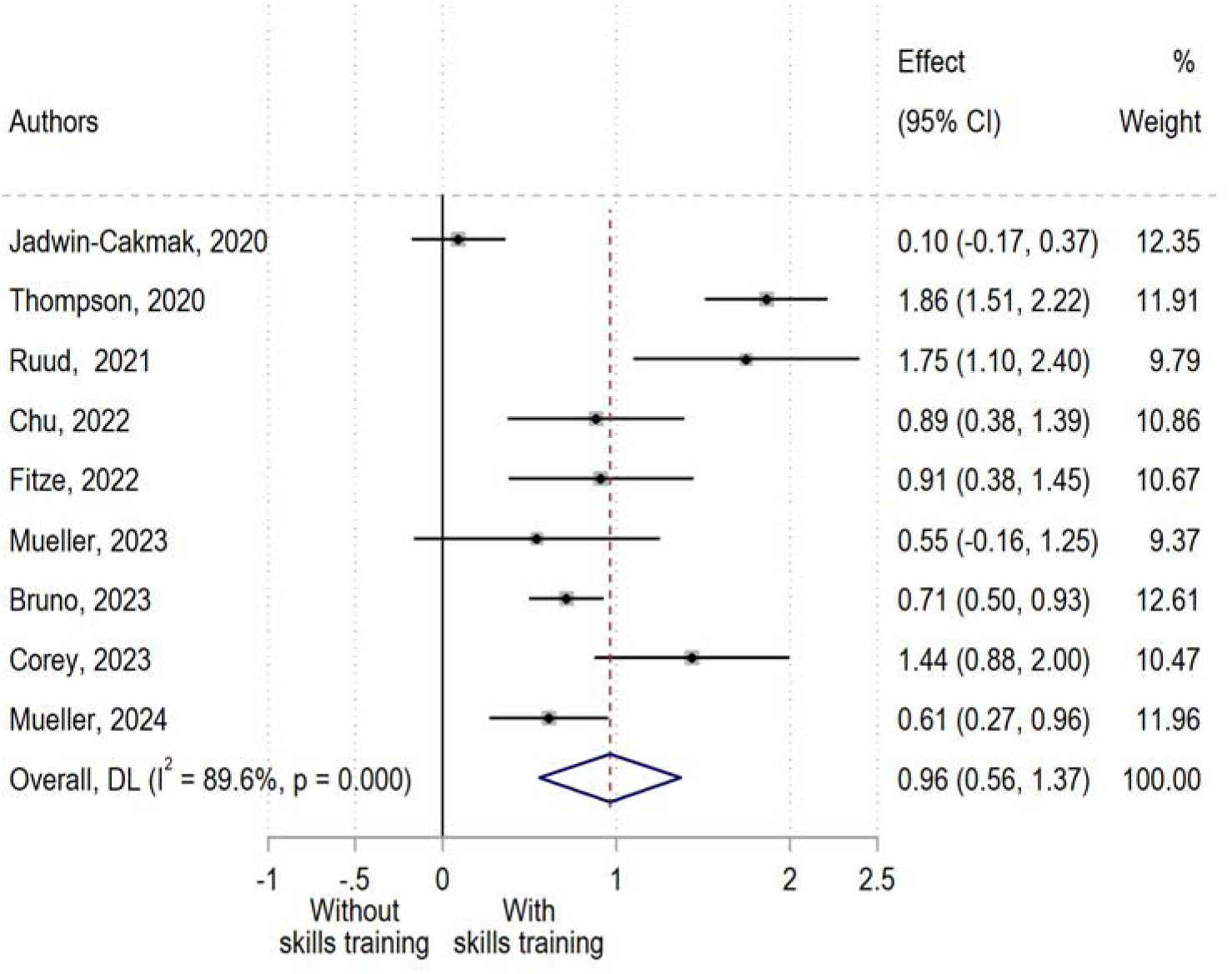
Change in competency – skills (high score=better outcome)

**Figure 13:**
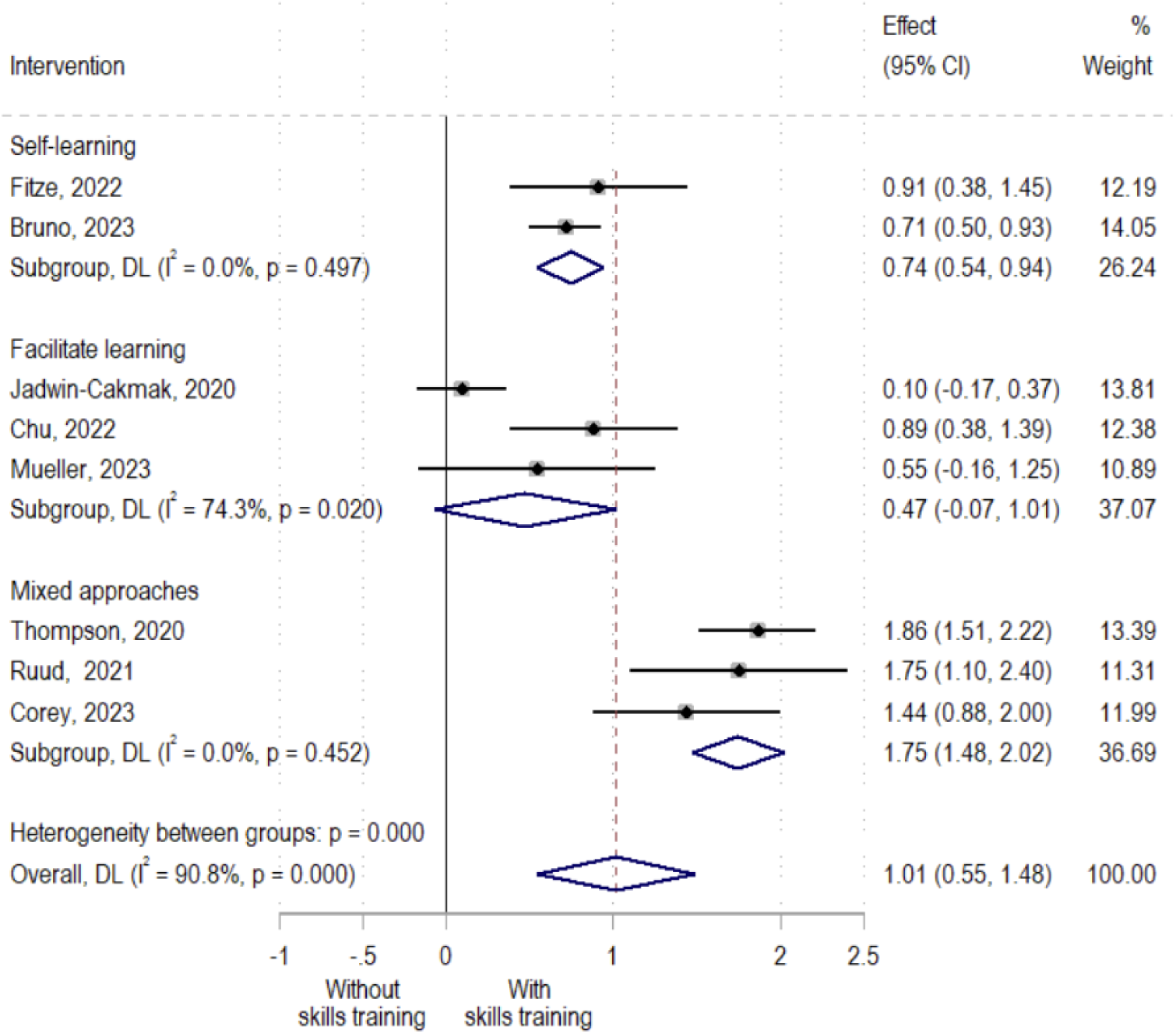
Change in competency – skills (subgroup analysis by type of intervention)

**Figure 14:**
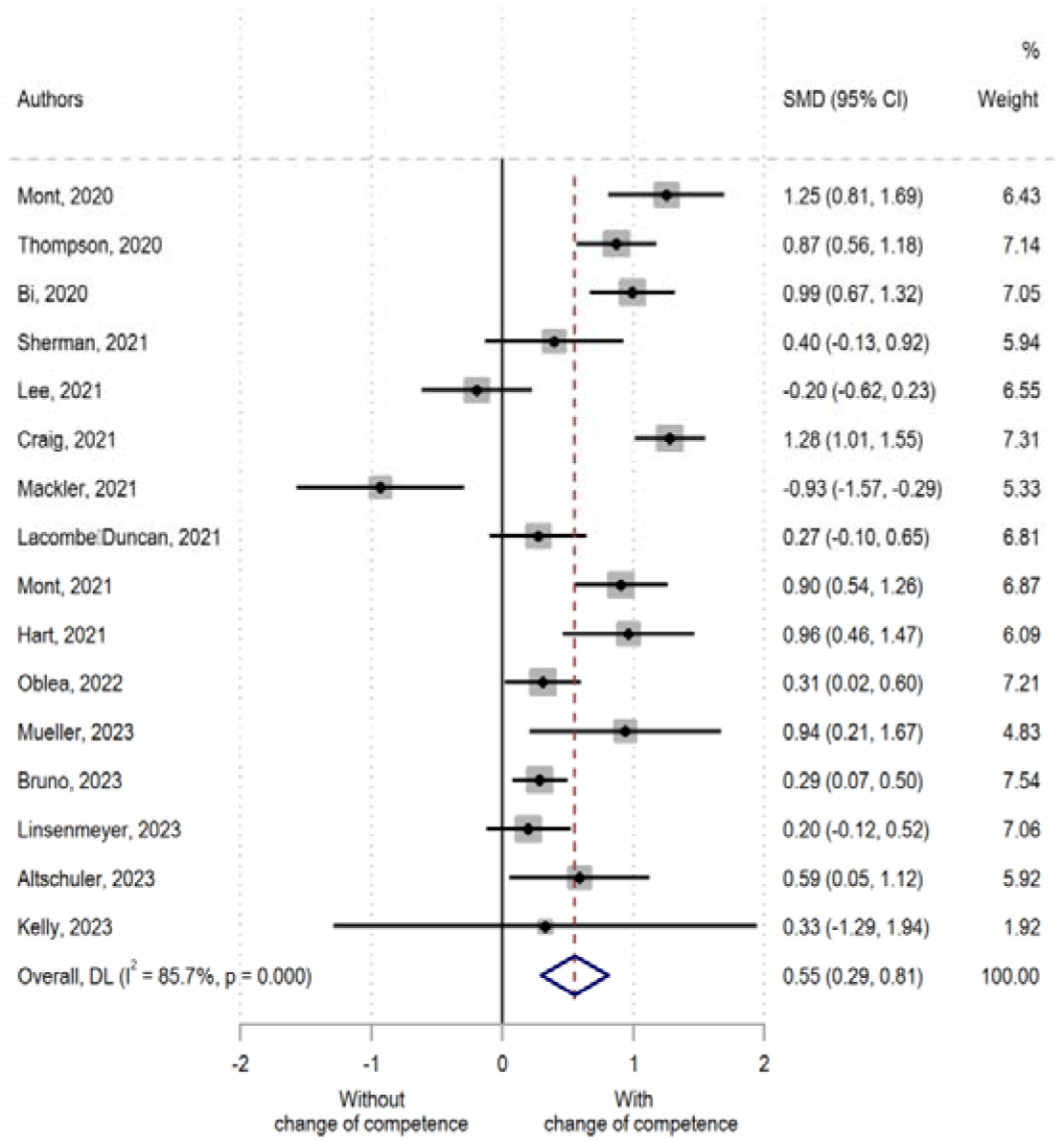
Change in competence: general (main analysis)

**Figure 15:**
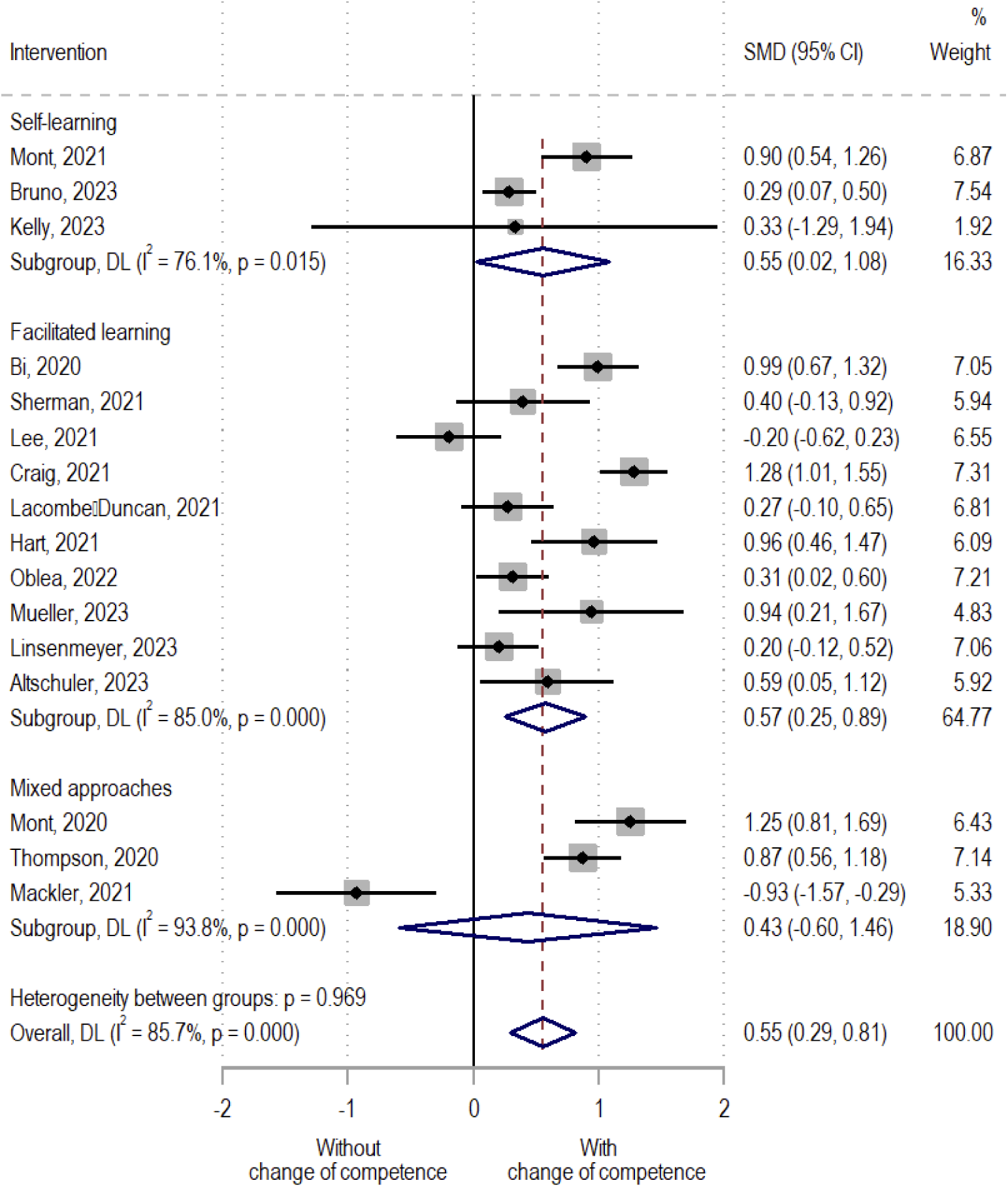
Change of competence: general (subgroup analysis by intervention)

**Figure 16:**
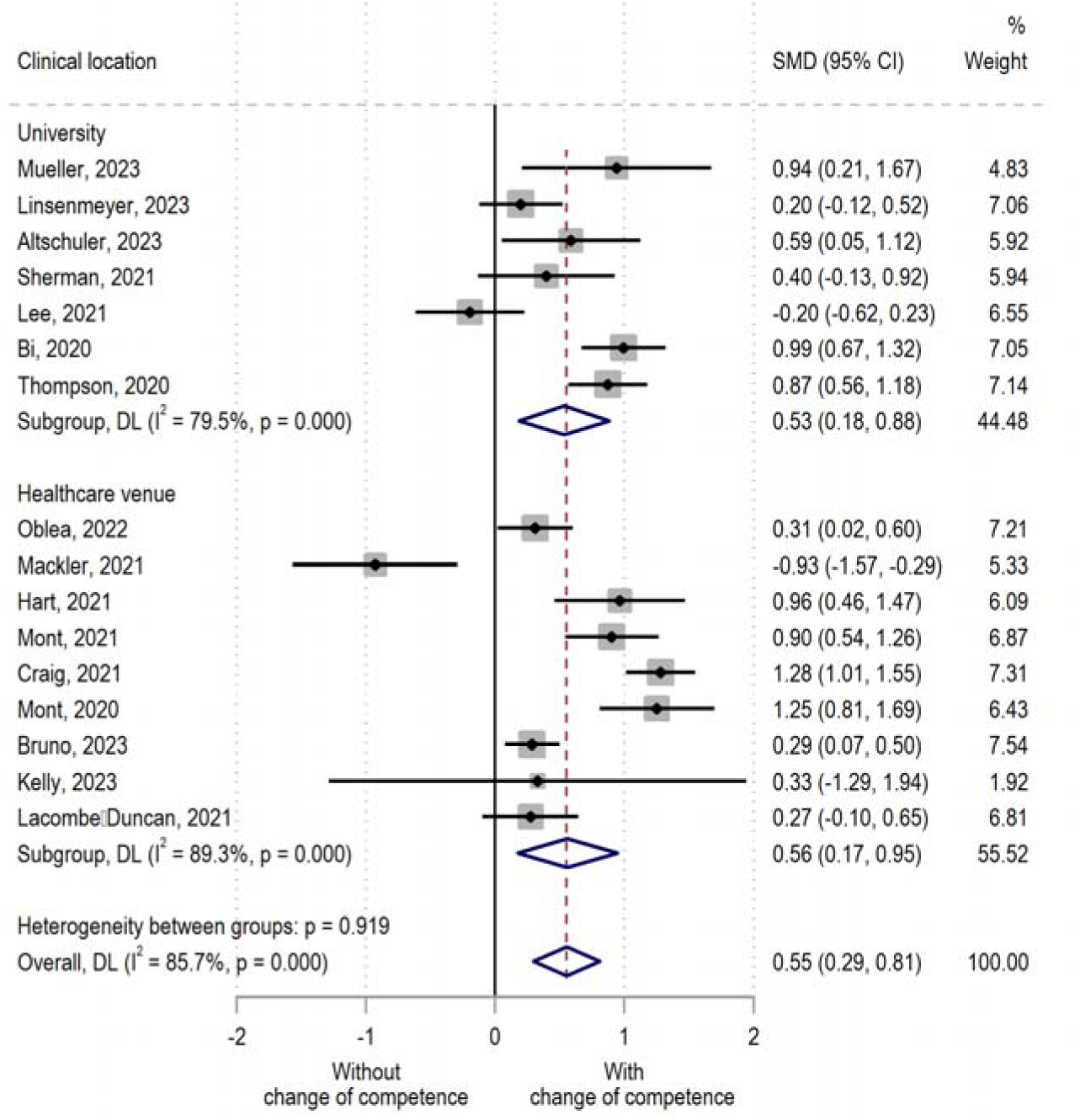
Change of competence: general (subgroup analysis by setting)

**Figure 17:**
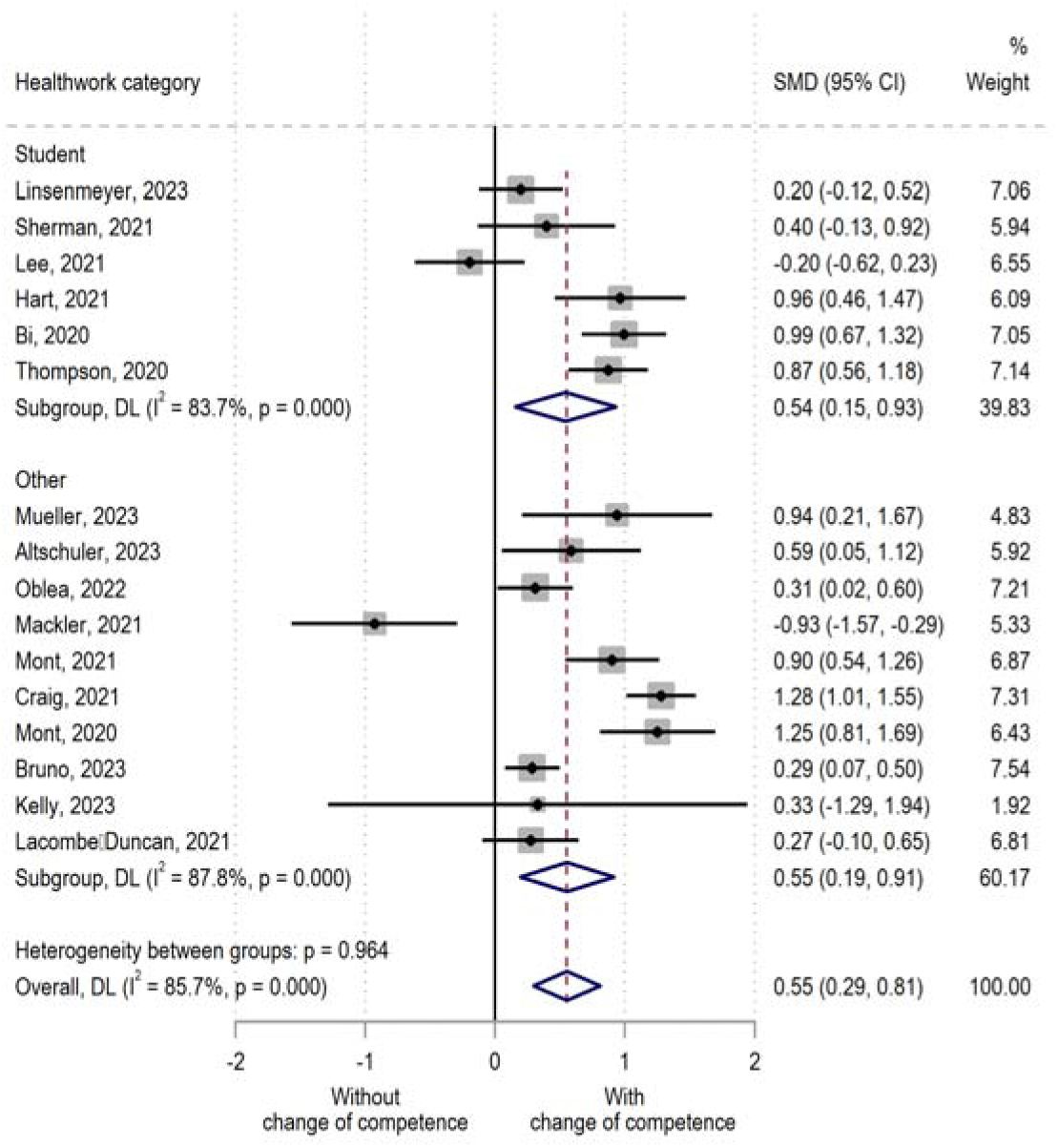
Change of competence: general (subgroup analysis by category of health workers)

**Figure 18:**
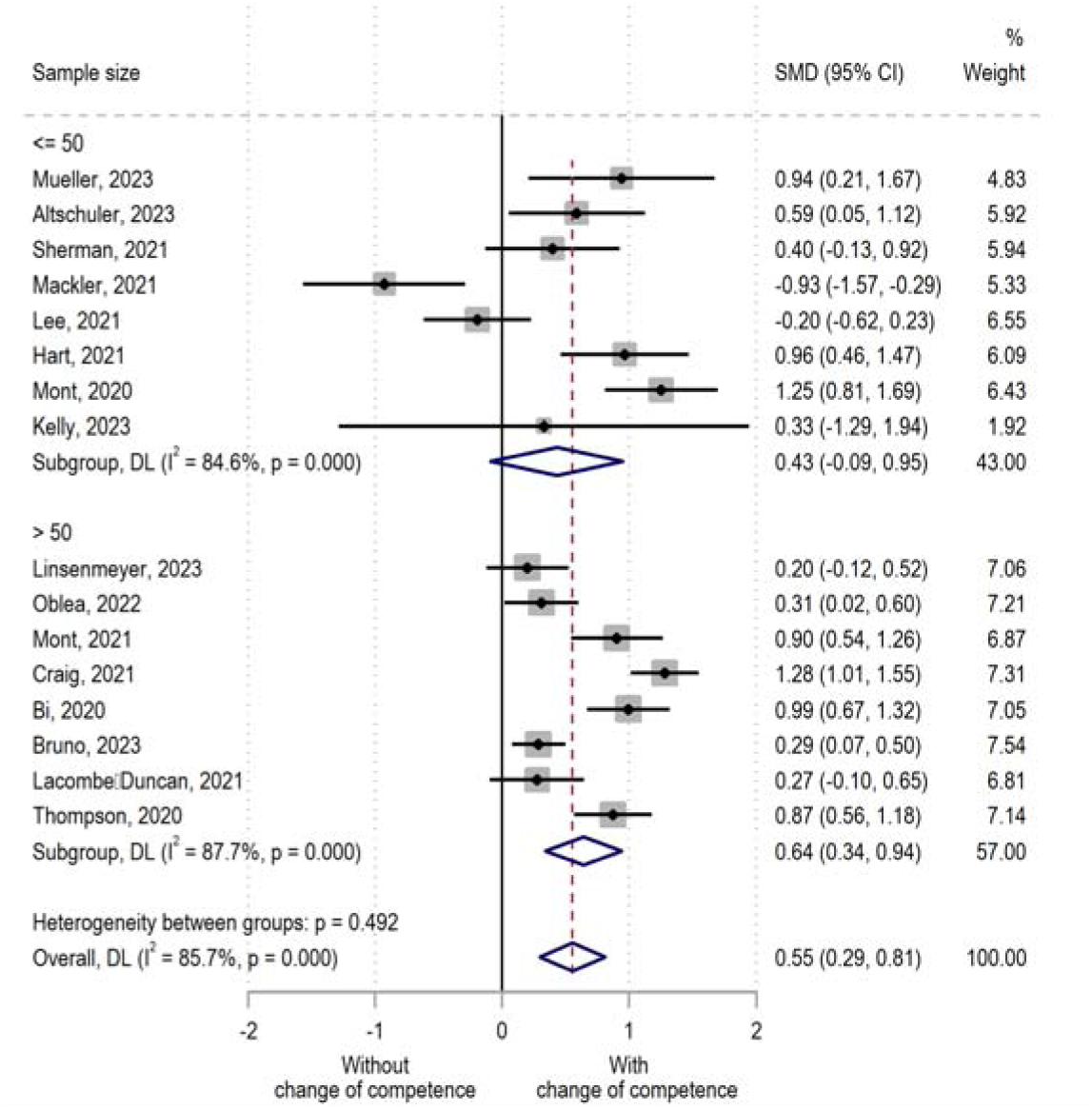
Change of competence: general (subgroup analysis by sample size)

**Figure 19:**
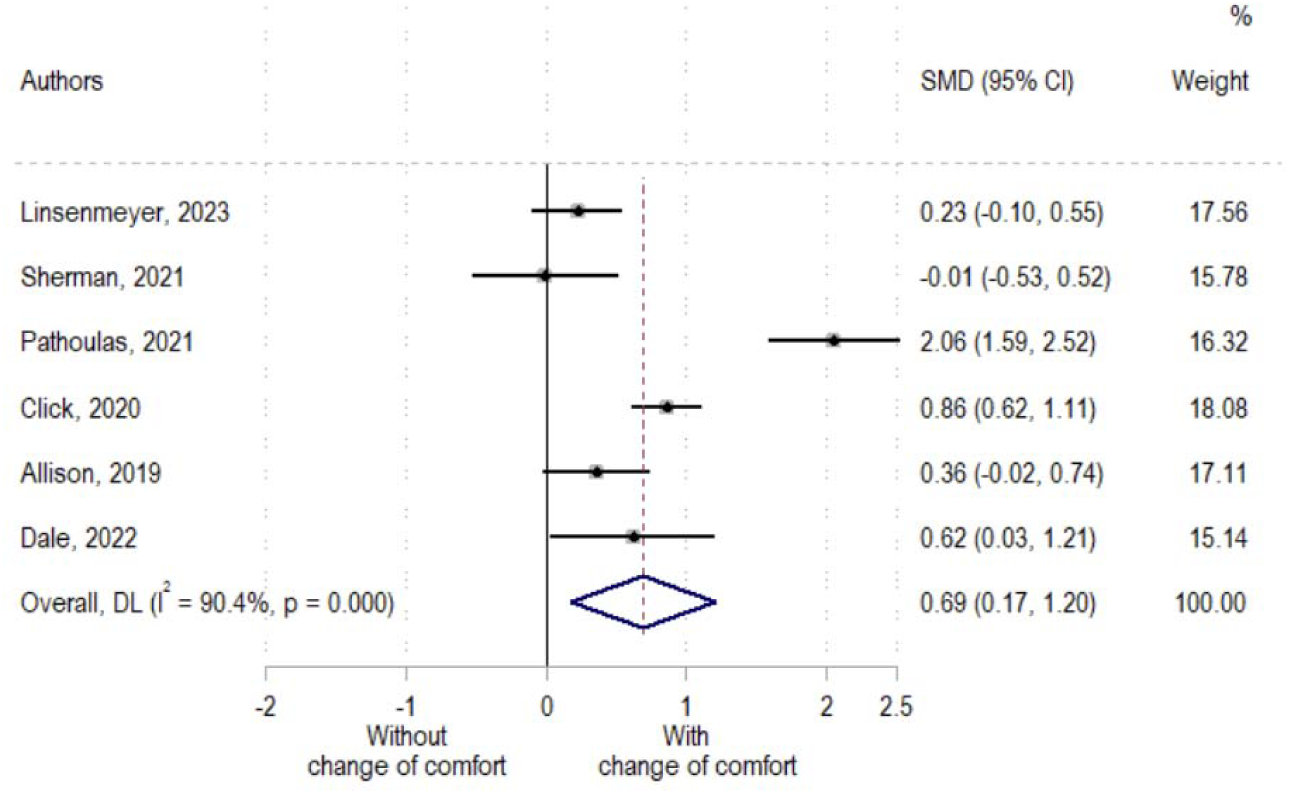
Change in Comfort.

